# N-terminal-targeted anti-amyloid monoclonal antibodies illuminate the therapy for Alzheimer’s disease: a systematic review and comprehensive meta-analysis

**DOI:** 10.1101/2023.11.20.23298794

**Authors:** Yu-Hui Qiu, Ming Liu, Jie Zhan, Ling-Ling Liu, Jia-Yi Zheng, Dan Wu, Guang-Liang Wu, Ye-Feng Cai, Shi-Jie Zhang

## Abstract

**Background:** Recent clinical trials of anti-amyloid-beta (Aβ) monoclonal antibodies (mAbs) have demonstrated that the removal of Aβ in symptomatic patients can slow down the progression of Alzheimer’s disease (AD) and reinforce the “amyloid cascade” hypothesis. However, further investigation and analysis of integrated clinical data is needed to interpret the clinical efficacy of these mAbs. In this study, we aimed to estimate the effectiveness of mAbs for AD and firstly assessed the therapeutic efficacy from a perspective of mAbs targeting specific Aβ domains (N-terminal, C-terminal, central domain, and N-terminal+central domain) in pre-specified subgroups.

**Methods:** In this systematic review and meta-analysis, we searched on Pubmed, Embase, the Cochrane Library, and ClinicalTrials.gov from their inception until 31 August, 2023, and updated our search on 15 October, 2023, to identify all published randomised controlled trials (RCTs) on various clinical outcomes of anti-Aβ mAbs in AD. The primary outcomes of interest included Alzheimer’s Disease Assessment Scale-cognitive subscale (ADAS-cog), Mini Mental State Examination (MMSE), and Clinical Dementia Rating scale-Sum of Boxes (CDR-SB), as well as amyloid positron emission tomography (PET), the main biomarker. Additionally, we collected the data of volumetric Magnetic Resonance Imaging (vMRI), cerebrospinal fluid (CSF), plasma AD biomarkers, and the risks of amyloid-related imaging abnormalities (ARIA). Random-effects models to calculate pooled risk ratios (RRs) and standardized mean differences (SMDs) were employed to analyze the data across multiple studies of all mAbs. Furthermore, we also examined the interrelationships among changes in alterations of cognitive performance, Aβ deposition, variations in AD biomarkers, and the risks of ARIA both in all mAbs and N-terminal-targeted mAbs by calculating Pearson’s correlation coefficients. This study is registered with PROSPERO, No. CRD42023430637.

**Findings:** We identified a total of 37 eligible studies on quality assessment, of which 34 were included in the meta-analysis. The analysis revealed that eight monoclonal antibodies (aducanumab, lecanemab, donanemab, bapineuzumab, gantenerumab, crenezumab, solanezumab, and ponezumab) demonstrated statistical improvements in cognitive outcomes. Specifically, the ADAS-cog [(total mAbs: SMD: -0.08, 95% CI: -0.11 to -0.04); (N-terminal-targeted mAbs: SMD: -0.08, 95% CI: -0.13 to -0.03)], CDR-SB [(total mAbs: SMD: -0.06, 95% CI: -0.12 to -0.00); (N-terminal-targeted mAbs: SMD: -0.08, 95% CI: -0.16 to -0.01)], and MMSE [(total mAbs: SMD: 0.06, 95% CI: 0.02 to 0.10); (N-terminal-targeted mAbs: SMD: 0.05, 95% CI: -0.00 to 0.10)] demonstrated significant improvement. In addition, the meta-analyses indicated that mAbs also had a statistically significant impact on reducing amyloid PET [(total mAbs: SMD: -1.13, 95% CI: -1.66 to -0.61); (N-terminal-targeted mAbs: SMD: -1.64, 95% CI: -2.27 to -1.02)], accelerating ventricle enlargement [(total mAbs: SMD: 0.22, 95% CI: 0.06 to 0.38); (N-terminal-targeted mAbs: SMD: 0.44, 95% CI: 0.30 to 0.57)], and causing whole-brain atrophy [(total mAbs: SMD: -0.14, 95% CI: -0.26 to -0.03); (N-terminal-targeted mAbs: SMD: -0.24, 95% CI: -0.41 to -0.07)], while increased the risk ARIA-E (total mAbs: RR: 9.79, 95% CI 7.83 to 12.26); (N-terminal-targeted mAbs: RR: 10.79, 95% CI: 8.61 to 13.52)] and ARIA-H [(total mAbs: RR: 1.28, 95% CI 0.98 to 1.67); (N-terminal-targeted mAbs: RR: 1.94, 95% CI: 1.64 to 2.29)]. However, no significant hippocampal volume atrophy was observed [(total mAbs: SMD: 0.00, 95% CI: -0.07 to 0.07); (N-terminal-targeted mAbs: SMD: -0.03, 95% CI: -0.13 to 0.07)]. It should be noted that all above effects were more significant in AD patients treated with N-terminal-targeted mAbs, as observed in subgroup analyses. Additionally, we observed a negative association between ventricular enlargement and Aβ clearance (Pearson’s r: -0.76), especially with the administration of N-terminal-targeted mAbs (Pearson’s r: -0.79), indicating a stronger effect on Aβ clearance. Moreover, there was a strong negative correlation between the improvement in cognitive function and the preservation of hippocampal volume (Pearson’s r: -0.70), particularly in the case of N-terminal-targeted mAbs. Lastly, a strong correlation was also found between the risk of ARIA and Aβ reduction in amyloid PET (Pearson’s r: -0.60), brain atrophy (Pearson’s r: -0.83), and ventricle enlargement (Pearson’s r: 0.92).

**Interpretation:** The administration of mAbs that specifically target the N-terminus of Aβ showed promising results in reducing Aβ burden and ameliorating cognitive decline. Furthermore, our preliminary findings shed light on the occurrence of brain atrophy, ventricular enlargement, and ARIA, might be attributed to the well clearance of Aβ deposits caused by mAb administration. In future anti-Aβ mAb development, our systematic review and meta-analysis indicated that N-terminal-targeted mAbs is an optimizing approach.

**Funding:** This work was supported by National Natural Science Foundation of China (No. 82004430, 82174310).

**Research in context:** *Evidence before this study:* The recent trials of lecanemab and donanemab have provided initial conclusive evidences that removal of Aβ from symptomatic patients’ brains can decelerate the progression of Alzheimer’s disease (AD). These findings offer clinical substantiation for the significance of aberrant Aβ in AD pathogenesis, thereby reinforcing the validity of the "amyloid cascade" hypothesis. While, the clinical benefit of the monoclonal antibodies (mAbs) is still limited and it is important to note that the treated subjects are still experienced disease progression, albeit at a slower rate. Targeting various forms of Aβ (monomers, oligomers, fibrils) is considered as the key mechanism of these mAbs’ efficacy. However, the results indicated that it is not crucial direction to explain the ideal antibody efficacy. In order to discover underlying mechanisms and formulate an enhanced immunotherapeutic regimen, it is essential to further analyze the integrated data of clinical trials.

*Added value of this study:* This comprehensive systematic review and meta-analysis not only encompassed all reported RCTs investigating the effects of anti-Aβ mAbs on various clinical outcomes in AD, but also firstly assessed the therapeutic efficacy of targeting specific Aβ domains (N-terminal, N-terminal+central-domain, central-domain, and C-terminal) by subgroup analyses. Enhanced data syntheses of all included 34 studies demonstrated significant enhancements in cognitive outcomes (ADAS-cog, CDR-SB and MMSE) with the utilization of mAbs. The meta-analysis also revealed that mAbs significantly reduced amyloid burden and certain AD biomarkers, expedited ventricle enlargement and whole-brain atrophy, concurrently increased the risk of ARIA. In addition, a notable efficacy was observed in AD patients by using the mAbs targeting the N-terminus of Aβ, as evidenced by subgroup analyses by employing different epitopes of Aβ. Association analysis identified that there was a positive correlation between the extent of reduction in Aβ deposition after mAbs therapy and the degree of improvement in cognitive function, thereby supporting Aβ plaques as a pivotal driver of cognitive decline in AD and emphasizing the clinical advantages associated with Aβ elimination from the brain. Further, we observed a possible association between brain atrophy or ventricular enlargement and Aβ clearance, especially with the administration of N-terminal-targeted mAbs, which demonstrated a stronger Aβ clearance. Improvement in cognitive function seemed to be related to both Aβ clearance and preservation of hippocampal volume. Moreover, the risk of ARIA was strongly correlated with reductions in amyloid PET and brain atrophy, as well as ventricle enlargement. Hence, it is essential for us to recognize that the clinical efficacy of N-terminal-targeted mAbs in clearing Aβ is crucial. Nevertheless, the exacerbation of cerebral atrophy and the occurrence of ARIA of higher severity are both caused by the great abilities of Aβ clearance.

*Implications of all the available evidence:* The findings of this comprehensive meta-analysis provided a strong support for the efficacy of N-terminal-targeted Aβ antibodies in significantly reducing Aβ burden and ameliorating cognitive decline in AD patients, which represented a potentially groundbreaking therapeutic strategy. The principle of “structure dictates function” is a guiding tenet that targeting N-terminal region of Aβ to design superior mAbs is a promising direction for the future.

## Introduction

Alzheimer’s disease (AD) is the most prevalent cause of dementia, accounting for 60-70% of cases ^1^. AD diminishes quality of life, amplifies the burden on patients and their families, and incurs substantial costs for caregiving ^2^. The main pathophysiology of AD is amyloid-beta (Aβ) peptide deposition, which results in neurodegeneration with cognitive disorder ^3,4^. Targeting Aβ clearance is considered to be a key way to slow the clinical progression of AD, such as active and passive immunotherapy against Aβ, and decrease Aβ production (β-secretase inhibitors or γ-secretase inhibitors) ^5^.

Various strategies have been explored to combat AD, and among them, passive immunotherapy by using monoclonal antibodies (mAbs) against Aβ is emerged as a prominent option, due to its well-tolerated and selective targeting capabilities ^6^. Despite the impressive preclinical efficacy and specificity of these mAbs, numerous clinical trials have yielded negative outcomes earlily, igniting debates on whether Aβ is a significant therapeutic target ^7^. Bapineuzumab ^8^ and solanezumab ^9,10^ showed no significant improvements in AD patients. Two phase III trials of crenezumab ^11^ and a phase III trial of gantenerumab ^12^ were terminated due to a lack of efficacy or futility. However, recent studies have shown promising results in Aβ clearance with aducanumab, lecanemab, and donanemab, which have benefit for AD patients ^13-16^. Aducanumab, the first potentially disease-modifying medication, received accelerated approval from the US Food and Drug Administration (FDA) ^14,17^. However, the clinical efficacy of aducanumab remains inconclusive, leading to ongoing controversies among experts ^17-20^. Lecanemab therapy demonstrated a 27% reduction in cognitive decline, as measured by Clinical Dementia Rating scale-Sum of Boxes (CDR-SB), compared to placebo in a multinational phase III study for early AD ^15^. Thus, lecanemab is considered as the first breakthrough in AD therapy, which had received the full FDA approval ^21-23^. In addition, the topline results of the TRAILBLAZER-ALZ 2 study demonstrated that a 35.1% deceleration in disease progression among patients with early symptomatic AD in the low/medium tau group who received donanemab compared to those received placebo over an 18-month period ^16^. It is introducing a new era of therapy for AD, and FDA is expected to grant a comparable standard approval for a third antibody ^18^.

The preliminary success of the current generation of anti-Aβ mAbs incentivizes scientists to elucidate the underlying principles and patterns. Scholars proposed that the effectiveness of the mAbs in AD is attributed to their abilities to target various forms of Aβ (monomers, oligomers, fibrils) ^24-27^, with soluble Aβ oligomers being more neurotoxic and clinically relevant ^28-30^. Significantly, these previous meta-analyses indicated that targeting various forms of Aβ was not crucial for the ideal antibody efficacy ^31^. The antibodies exhibit differential recognition of distinct antigenic sites on Aβ, and they vary in terms of their apparent clinical efficacy and ability to reduce Aβ accumulation ^26^. It is noteworthy that the mAbs with the greatest potential, namely aducanumab, lecanemab and donanemab, exhibit specific binding to the N-terminal pyroglutamate Aβ epitope ^14-16^. Conversely, mAbs targeting other regions, such as N-terminal+central-domain (gantenerumab), central-domain (crenezumab and solanezumab), and C-terminus (ponezumab), have been largely reported to lack efficacy ^10-12,32^. The effectiveness of N-terminal-targeted mAbs in clearing aggregated Aβ may also be attributed to the fact that N-terminal is exposed in Aβ40 or Aβ42 monomer aggregates, including oligomers and fibrils ^13,16,33^, which subsequently induce microglial activation and phagocytosis ^34^. Thus, we hypothesized that mAbs targeting the N-terminus of Aβ were likely to be the most effective in slowing AD progression.

In this systematic review and meta-analysis, we comprehensively included all reported randomised controlled trials (RCTs) on various clinical outcomes of anti-Aβ mAbs in AD, exploring potential relationships among clinical effects, biomarkers associated with Aβ and tau pathologies, volumetric Magnetic Resonance Imaging (vMRI), and the risk for specific adverse events (amyloid-related imaging abnormalities, ARIA). Subgroup analyses based on different mAbs recognizing Aβ epitopes (including those targeting the N-terminal, C-terminal, central domain, and N-terminal+central domain ^31^) were conducted, aiming to identify specific domains of Aβ that could be targeted for improved therapeutic effects. Our findings revealed that mAbs targeting the N-terminus of Aβ showed a remarkable efficacy for AD patients, surpassing other types of mAbs.

## Methods

### Search strategy and selection criteria

The study was registered with PROSPERO (No. CRD42023430637). In accordance with the Preferred Reporting Items for Systematic Reviews and Meta-analyses (PRISMA) guidelines ^35^, this study involved a systematic review and meta-analysis by comprehensively searching the relative literatures in PubMed, Embase, the Cochrane Library, and ClinicalTrials.gov from their inception until 31 August, 2023, and updated our search on 15 October, 2023. To ascertain a comprehensive coverage, we implemented search strategies that combined database-specific subject headings (including Medical Subject Headings terms) and relevant free text terms (such as AD, monoclonal antibody, Drug Therapy, and RCTs) in order to identify potentially eligible studies. The search strategy details were presented in the Appendix 1.

Inclusion criteria:

1. Study populations: Patients with mild cognitive impairment (MCI) or any stage of AD;
2. Intervention and comparison: anti-Aβ mAbs with placebo or alternative doses of the drugs;
3. Study design: RCTs.

On the other hand, studies meeting any of the following criteria were excluded:

1. Narrative/systematic reviews, meta-analyses, open-label trials, single-arm trials, prospective cohort studies, case-control studies, cross-sectional studies, case series studies, case report studies, opinion/editorial pieces.
2. Post-hoc or secondary analyses of main studies.
3. Studies involving non-human subjects.
4. Studies including participants diagnosed with dementia other than AD (familial AD and other types of dementias were excluded).
5. Studies that did not report any cognitive/functional outcomes or AD biomarkers.

By applying these inclusion and exclusion criteria, we ensured that the selected studies were appropriate for this systematic review and meta-analysis.

### Study selection and data extraction

Two reviewers (YHQ and LLL) independently conducted title and abstract screening to identify eligible full-text studies, and extracted data from these studies using a pre-defined data extraction form. Any discrepancies regarding in the eligibility of full-text articles were resolved through discussion or arbitration with third-party reviewers (ML, JZ, JYZ, DW, GLW, YFC and SJZ).

The collected information from the included studies consisted of various aspects:

1. Details about the studies: eg, the author’s name, year of publication, study design, and sample size;
2. Baseline participant characteristics: eg, the mean age, gender distribution, percentage of Apolipoprotein ε4 (APOE4) carriers, baseline Mini Mental State Examination (MMSE), baseline CDR-SB, AD medications used and degree of AD;
3. The intervention and comparison characteristics: eg, the mode of treatment, type of drug, dose, duration of study treatment, as well as the type of control, dose, and duration of treatment;
4. Outcomes: the primary outcomes: the main clinical outcomes of interest [Alzheimer’s Disease Assessment Scale - cognitive subscale (ADAS-cog), MMSE, and CDR-SB], the main biomarkers [amyloid positron emission tomography (PET)]; the secondary outcomes: vMRI (volumes of the whole brain, hippocampus, and ventricle), cerebrospinal fluid (CSF) biomarkers [Aβ40, Aβ42, phosphorylated tau (p-tau), total tau, phosphorylated tau protein at position 181 (p-tau181), Neurofilament Light (Nfl), Neurogranin], plasma biomarkers (Aβ40, Nfl, p-tau181), tau PET, the risks for adverse reactions[Amyloid-Related Imaging Abnormalities-Edema (ARIA-E) and Amyloid-Related Imaging Abnormalities-Hemorrhage (ARIA-H)].

The outcome data was collected at the longest follow-up for trials that included multiple follow-up points, data, or reports. In cases where precise values were unavailable, approximations were made based on graphical representations ^36^.

The extraction of volumetric data involved calculating mean volume changes using baseline volumes of the whole brain, hippocampus, and ventricle. In cases where these values were not reported in the publication or on Clinical Trials.gov, a representative study ^37^ was used to impute the baseline ^36^. Baseline values were not reported in four studies of crenezumab (NCT01397578 ^38^, NCT01343966 ^39^, NCT03114657 ^11^ and NCT02670083 ^11^). In the bapineuzumab study (NCT01254773 ^40^), left and right hippocampus volumes were reported separately. To obtain a single mean value for the hippocampus and a single value for the standard deviation (SD), we combined both left and right using the formula 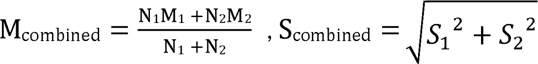^36^.

### Quality assessment

The revised Cochrane RoB tool for randomized trials, version 2 (RoB2) ^41^ was used to evaluate the methodological quality of eligible studies with risk of bias. The Excel-based version officially released by the Cochrane Collaboration was employed. Potential biases arising from the randomisation procedure, variations in planned interventions, incomplete outcome data, measurement of outcomes, and choice of reported findings were assessed. The risk of bias for individual domains and studies was classified as “low”, “some concerns”, or “high”. Risk of bias assessments were done independently by four of the investigators (YHQ, LLL, JZ, and GLW), with disagreements resolved through discussion.

### Data analysis

The pooled risk ratios (RRs) with 95% confidence intervals (CIs) were estimated for binary outcomes, while the standardized mean differences (SMDs) with 95% CIs were calculated for continuous outcomes. In studies involving multiple intervention groups with varying doses, we consolidated all relevant experimental intervention groups into a single group (with the comparison being made between all active interventions and placebo) ^42^. The means and SDs were then combined by using appropriate formulas for continuous outcomes as outlined in the Cochrane Handbook^42^. To accommodate the heterogeneity among studies in our statistical analysis, we adhered to the recommendation of employing a random-effects model ^43^.

The assessment of publication bias involved the examination of funnel plots and Egger’s regression intercept, with a minimum requirement of having an adequate number of studies (≥10) ^44^.

In this research, we implemented subgroup analyses to investigate the impact of individual drugs and specific Aβ epitopes targeted by mAbs, namely those directed towards the N-terminal, C-terminal, central-domain, and N-terminal+central-domain regions. Furthermore, meta-regressions were employed to assess the influence of baseline MMSE, baseline CDR-SB, the mean age, percentage of APOE4 carriers, percentage of AD medications used, and sex on treatment efficacy.

To ensure the robustness of our findings, we also performed a sensitivity analysis by systematically removing each study to evaluate its individual impact on the overall effect size. Furthermore, in addition to the random-effects model, analyses were conducted by using different modeling approaches, such as fixed-effects models.

To examine the interrelationships among alterations in cognitive performance, changes in Aβ deposition, variations in AD biomarkers, and the risks of ARIA-E and ARIA-H across multiple studies, we computed Pearson’s correlation coefficients. All statistical analyses were done by using R (version 4.3.1) and MetaXL (version 5.3).

### Role of the funding source

The funders of the study had no role in study design, data collection, data analysis, data interpretation, or writing of the report. The corresponding author had full access to all the data in the study and had final responsibility for the decision to submit for publication.

## Results

After identifying 4160 records from the electronic databases, a total of 4041 were excluded based on the evaluation of their titles and abstracts. Out of the 119 records that underwent a comprehensive review, 66 were subsequently excluded due to failure in meeting the eligible criteria. Finally, 37 RCTs with 21,609 participants were included in this review (Figure 1). Of the eligible RCTs, 34 studies provided sufficient data for quantitative synthesis, and their outcomes were included in the meta-analysis. However, 3 RCTs ^45-47^ without available data were only included in the qualitative analysis.

**Figure 1.**
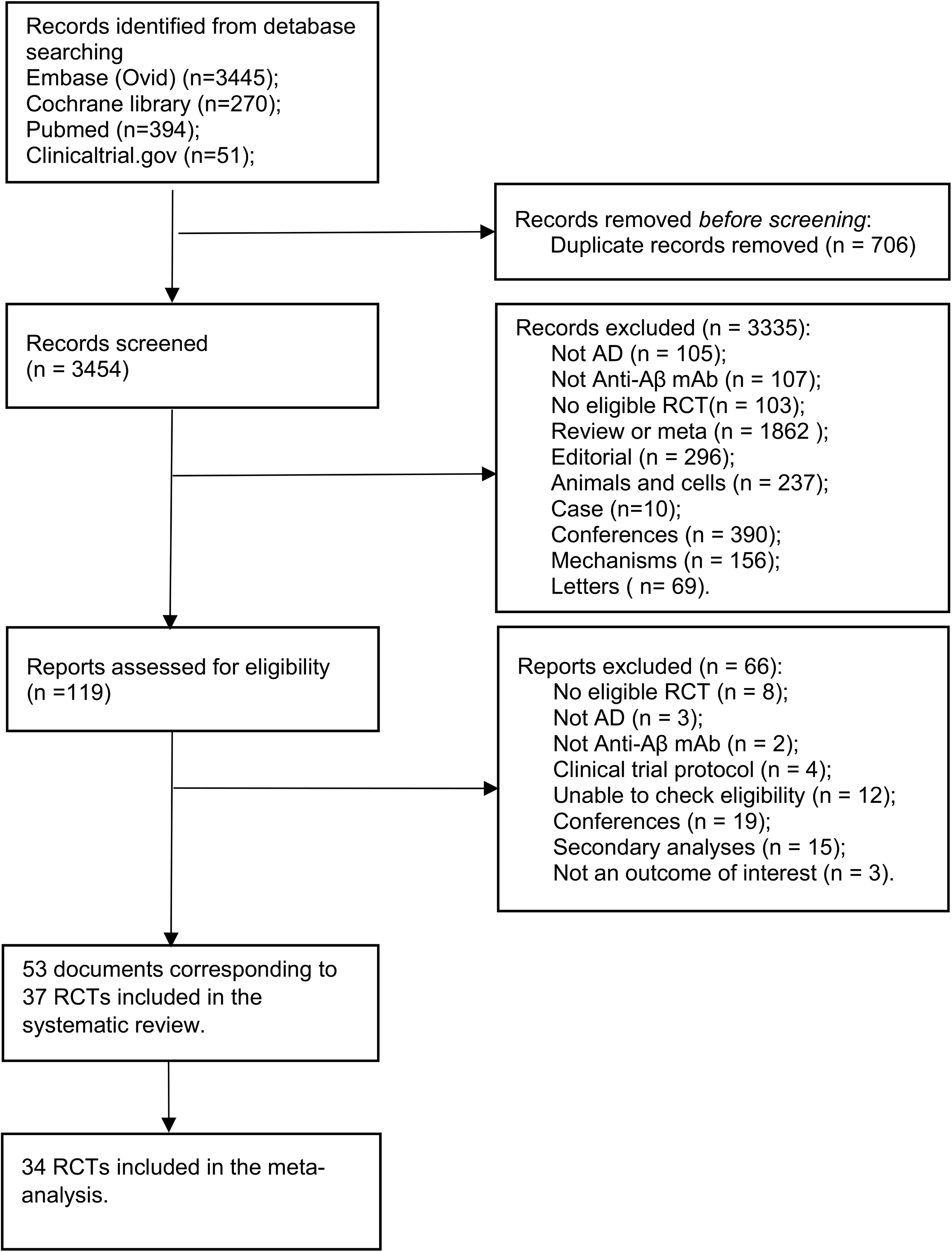
Study selection

Appendix 2 provided a summary of the baseline characteristics of the 37 eligible RCTs: four studies tested aducanumab ^14,34,48^, three tested donanemab ^13,16,49^, three tested lecanemab ^15,50,51^, eleven tested bapineuzumab ^8,40,45,52-56^, six tested solanezumab ^9,10,46,57,58^, five tested crenezumab ^11,38,39,59^, one tested gantenerumab ^12^ and four tested ponezumab ^32,47,60,61^. Additionally, twenty-one (56.76%) studies examined N-terminal-targeted anti-Aβ mAbs (aducanumab, donanemab, lecanemab and bapineuzumab), eleven (29.73%) studies focused on mAbs targeting the central domain (solanezumab and crenezumab), one (2.70%) study investigated the efficacy of N-terminal+central-domain-targeted mAb (gantenerumab), four (10.81%) studies evaluated C-terminal-targeted mAbs (ponezumab). Out of the total number of studies analyzed, eight (21.62%) were categorized as phase I studies, ten (27.03%) were classified as phase II studies, fifteen (40.54%) were identified as phase III studies, and four (10.81%) were unclear.

Regarding risk of bias, fifteen (40.54%) studies were at “low risk” and twenty-two (59.46%) studies were deemed as “some concerns” (Appendix 4).

Compared with placebo, the mAbs statistically improved performance on the cognitive measurements, ADAS-cog score (SMD: -0.08, 95% CI: -0.11 to -0.04; I^2^ = 0%) (negative change was indicated improvement) (Figure 2A), CDR-SB (SMD: -0.06, 95% CI: -0.12 to -0.00; I^2^ = 63%) (negative change was indicated improvement) (Figure 2B) and MMSE (SMD: 0.06, 95% CI: 0.02 to 0.10; I^2^ = 0%) (positive change was indicated improvement) (Appendix 6). Funnel plots were symmetric for all three primary outcomes (Appendix 5.1-5.3) and Egger’s test p-values were 0.088, 0.392 and 0.192, respectively; thus, there was no evidence for publication bias. Subgroup analyses revealed that ADAS-cog was statistically improved by aducanumab, lecanemab, donanemab and solanezumab separately (Figure 2A); CDR-SB was statistically improved only by aducanumab, lecanemab and donanemab (Figure 2B); and MMSE was statistically improved by donanemab and solanezumab (Appendix 6). Among them, donanemab, the change on CDR-SB was of moderate effect size (SMD: -0.28, 95% CI: -0.39 to -0.18; I^2^ = 0%) (Figure 2B), ADAS-cog and MMSE were of small effect size [(ADAS-cog: SMD: -0.19, 95% CI: -0.29 to -0.09; I^2^ = 0%) (Figure 2A), (MMSE: SMD: 0.13, 95% CI: 0.03 to 0.23; I^2^ = 0%)(Appendix 6)]; The pooled effect for all lecanemab studies reached significance for both ADAS-cog and CDR-SB [(ADAS-cog: SMD: -0.15, 95% CI: -0.23 to -0.06; I^2^ = 0%)(Figure 2A), (CDR-SB: SMD: -0.17, 95% CI: -0.26 to -0.09; I^2^ = 0%) (Figure 2B)], and corresponding effect sizes were similar to each other. The results of additional subgroup analyses did not show any significant differences in the mAbs type of targeting different Aβ epitopes. There were some evidences that mAbs targeting N-terminal and central-domain had some favorable effects on cognitive outcomes [ADAS-cog: (N-terminal: SMD: -0.08, 95% CI: -0.13 to -0.03), (central-domain: SMD: -0.08, 95% CI: -0.14 to -0.02) (Figure 2A); CDR-SB: (N-terminal: SMD: -0.08, 95% CI: -0.16 to -0.01), (central-domain: SMD: -0.03, 95% CI: -0.11 to 0.05;) (Figure 2B); MMSE: (N-terminal: SMD: 0.05, 95% CI: -0.00 to 0.10), (central-domain: SMD: 0.09, 95% CI: 0.02 to 0.15) (Appendix 6)], whereas mAbs targeting C-terminal and N-terminal+central-domain did not produce any effects on cognitive outcomes [ADAS-cog: (C-terminal: SMD: 0.22, 95% CI: -0.08 to 0.52), (N-terminal+central-domain: SMD: -0.07, 95% CI: -0.30 to 0.17) (Figure 2A); CDR-SB: (N-terminal+central-domain: SMD: 0.07, 95% CI: -0.17 to 0.30) (Figure 2B); MMSE: (C-terminal: SMD: 0.38, 95% CI: -0.19 to 0.95), (N-terminal+central-domain: SMD: 0.02, 95% CI: -0.22 to 0.25) (Appendix 6)].

**Figure 2.**
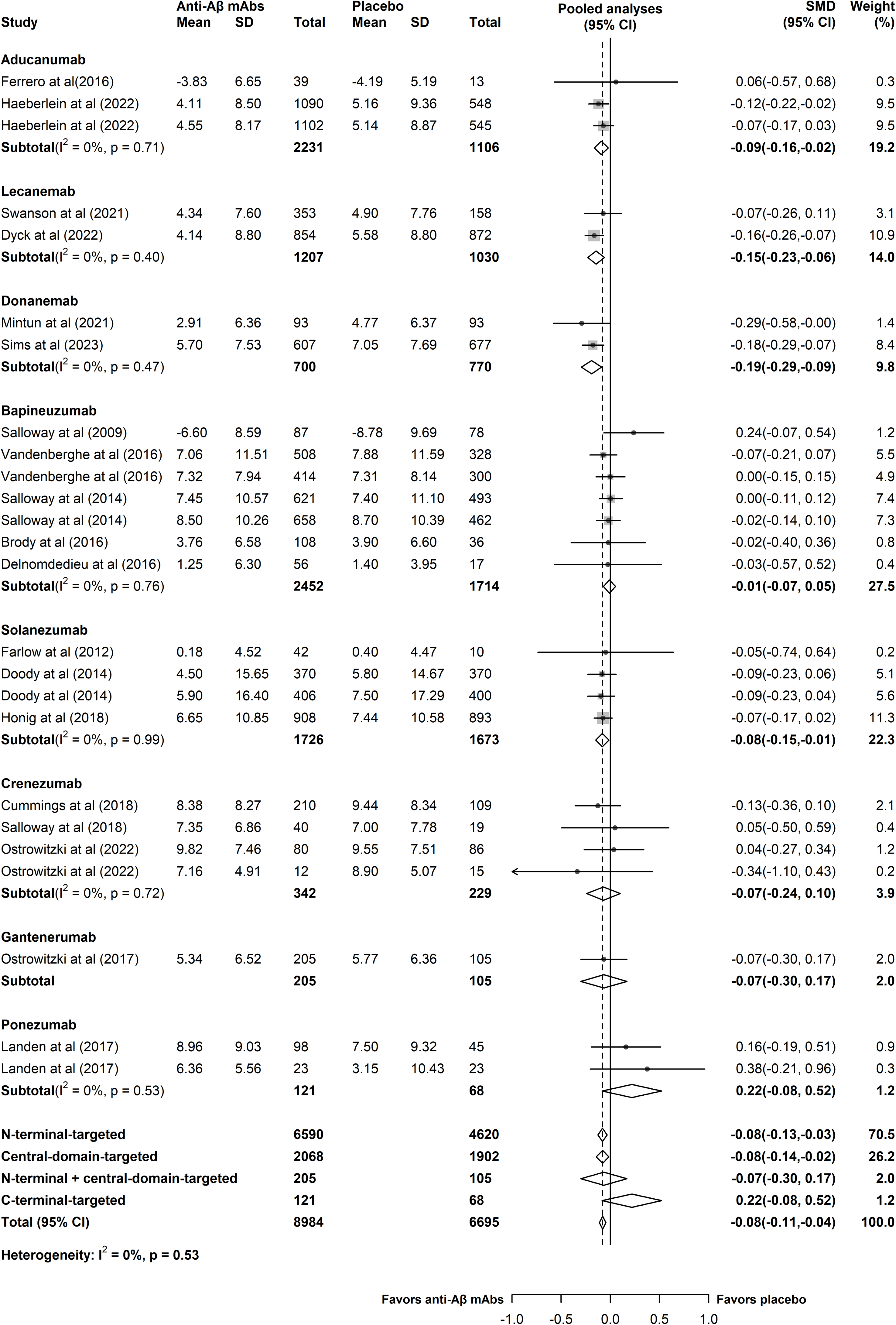
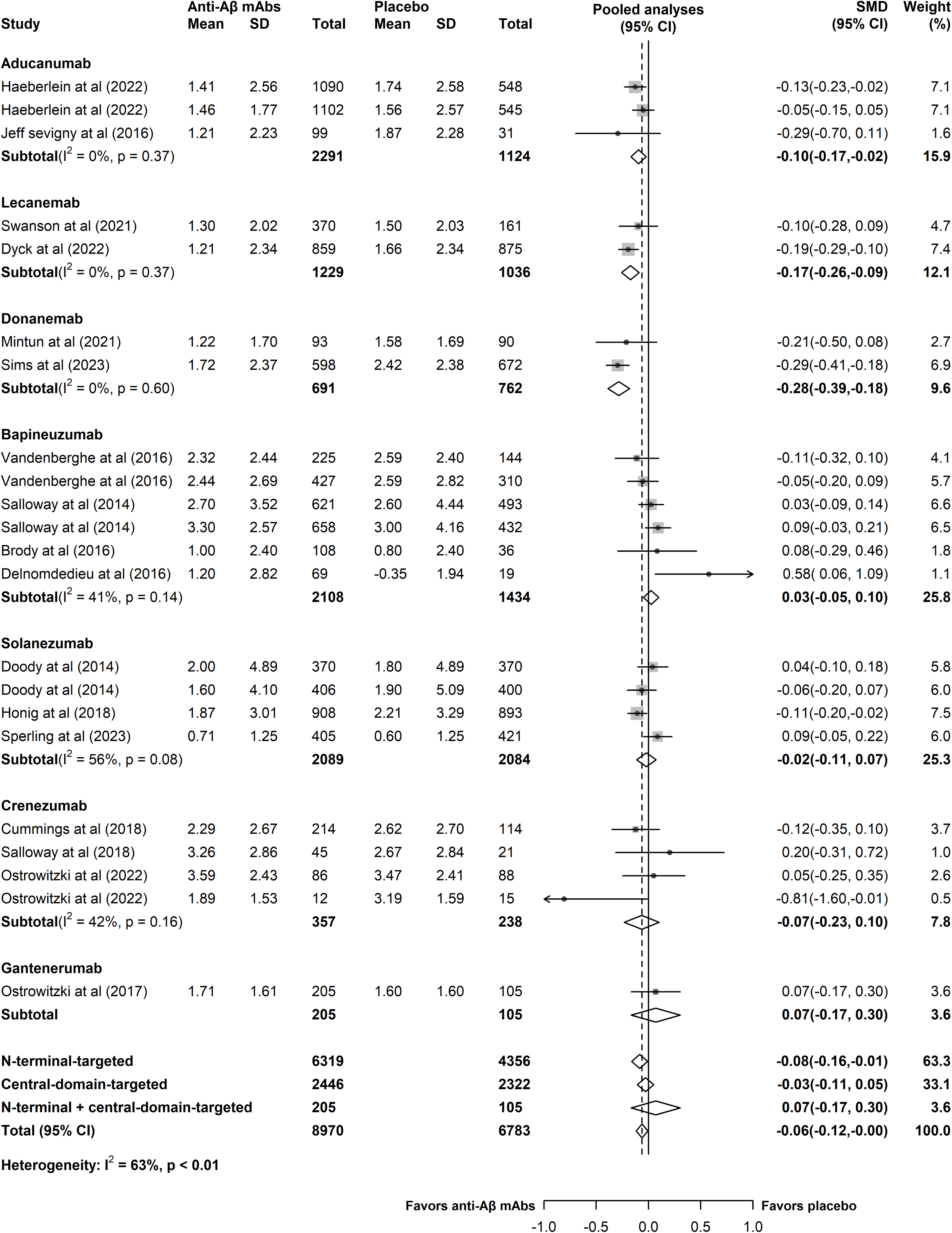
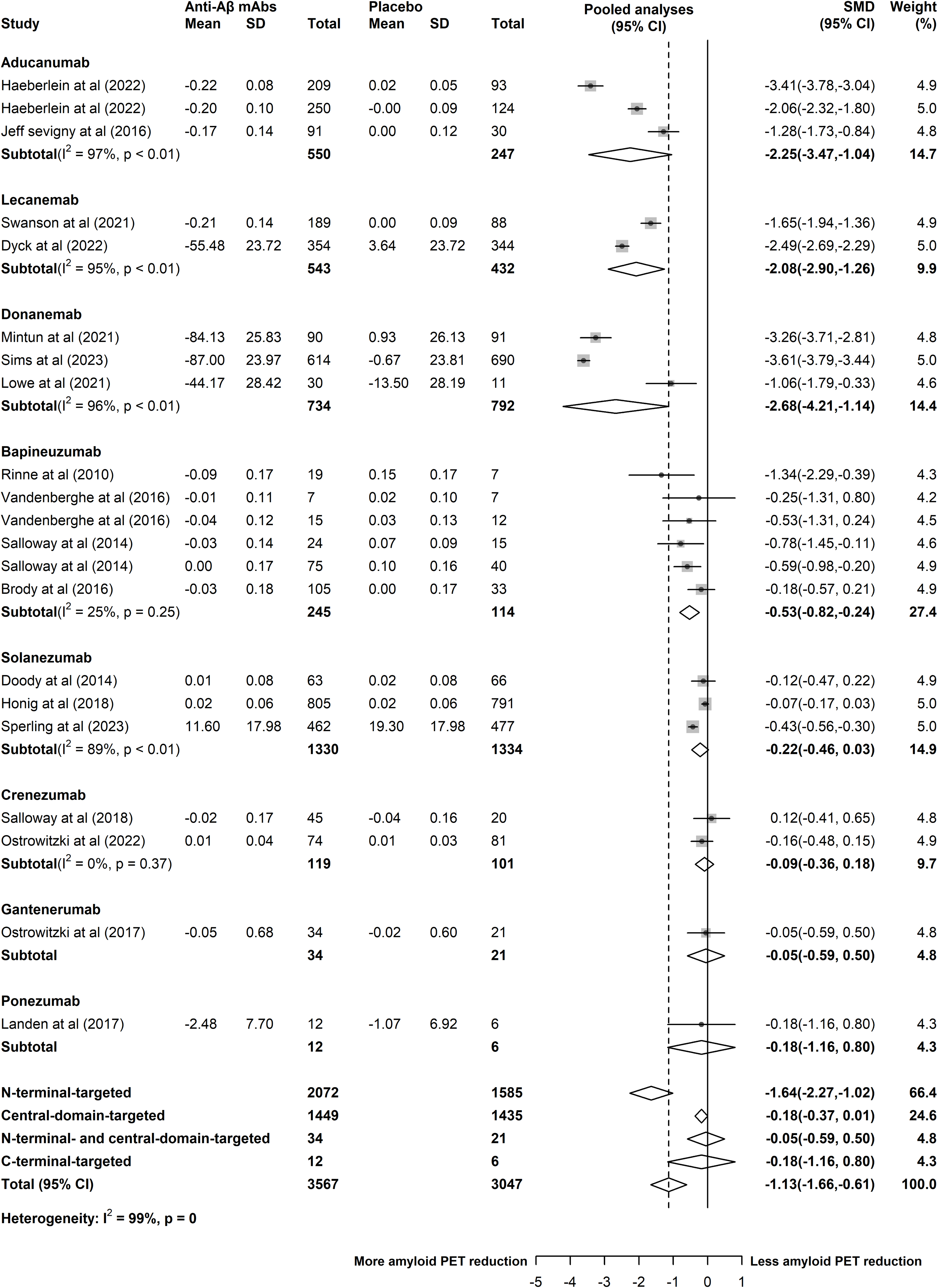
Forest plot of standardized mean differences for clinical assessments. Data were presented for Alzheimer’s Disease Assessment Scale-cognitive subscale (ADAS-cog) (A), and Clinical Dementia Rating scale-Sum of Boxes (CDR-SB) (B). Weights were assigned by random effects analysis. SMD: standardized mean difference.

The mAbs treatment induced a significant reduction in amyloid PET (SMD: -1.13, 95% CI: -1.66 to -0.61; I^2^ = 99%) (negative change was indicated more amyloid PET reduction), compared to the placebo group (Figure 3). A trend towards a reduction on tau PET (SMD: -0.89, 95% CI: -1.84 to 0.05; I^2^ = 99%) (negative change was indicated more tau PET reduction) was also observed, however, this finding did not reach statistical significance (Appendix 7.11). No publication bias was observed for amyloid PET (Egger test, p = 0.465) (Appendix 5.4). In the subgroup analyses specifically focusing on mAbs, the findings revealed that aducanumab (SMD: -2.25, 95% CI: -3.47 to -1.04; I^2^ = 97%), lecanemab (SMD: -2.08, 95% CI: -2.90 to -1.26; I^2^ = 95%), and donanemab (SMD: -2.68, 95% CI: -4.21 to -1.14; I^2^ = 96%) exhibited a very strong effect in reducing amyloid PET levels (Figure 3). Likewise, bapineuzumab demonstrated a large effect size in decreasing amyloid PET (SMD: -0.53, 95% CI: -0.82 to -0.24; I^2^ = 25%) (Figure 3). An interesting observation was that all the drugs targeting the N-terminal region of Aβ resulted in the most significant reduction of brain amyloid (SMD: -1.64, 95% CI: -2.27 to -1.02) (Figure 3). On the contrary, there was no noticeable change in terms of reducing amyloid PET levels in the mAbs targeting the central-domain and the others [(central-domain: SMD: -0.18, 95% CI: -0.37 to 0.01), (N-terminal+central-domain: SMD: -0.05, 95% CI: -0.59 to 0.50), (C-terminal: SMD: -0.18, 95% CI: -1.16 to 0.80)] (Figure 3).

**Figure 3.**
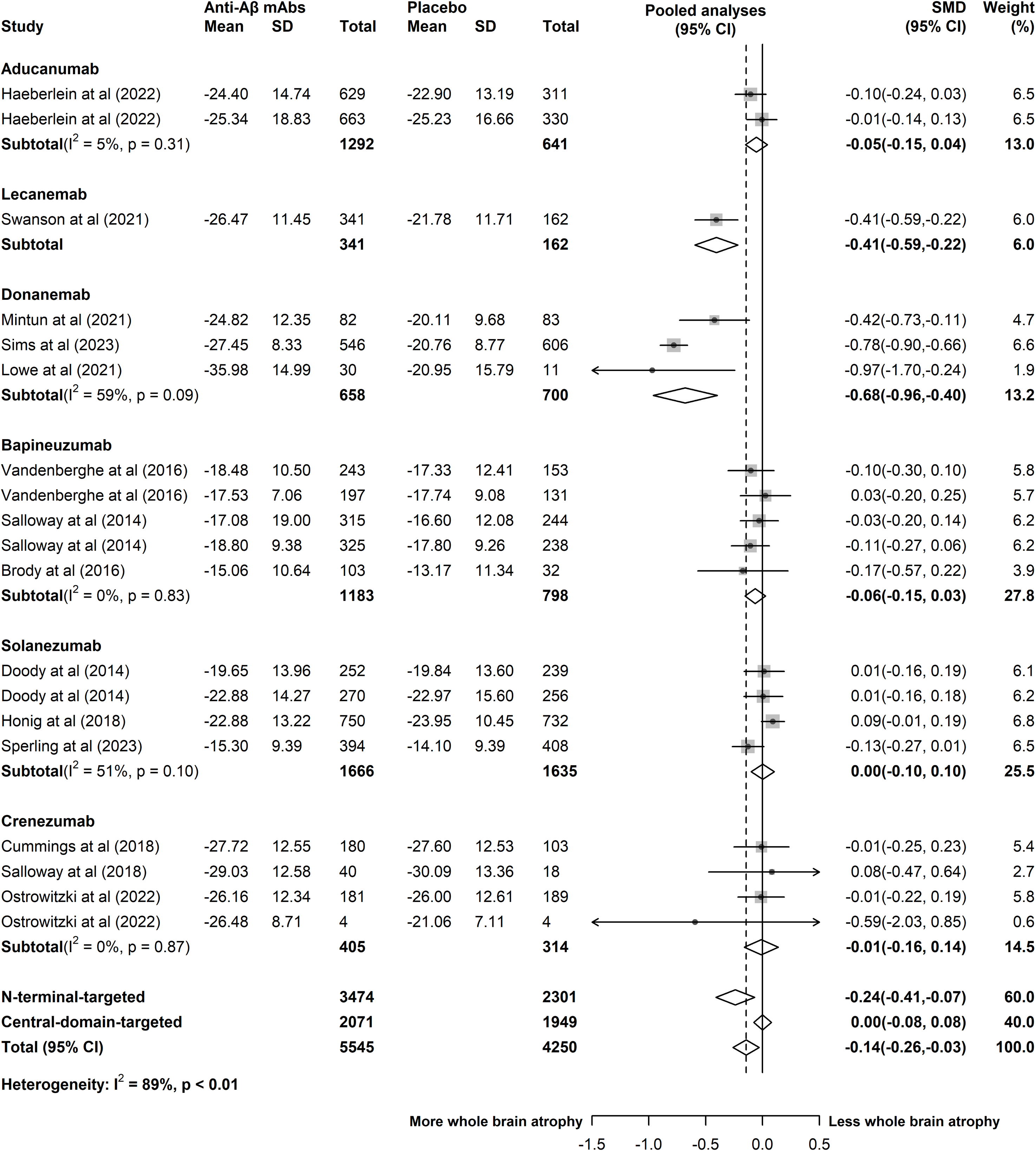
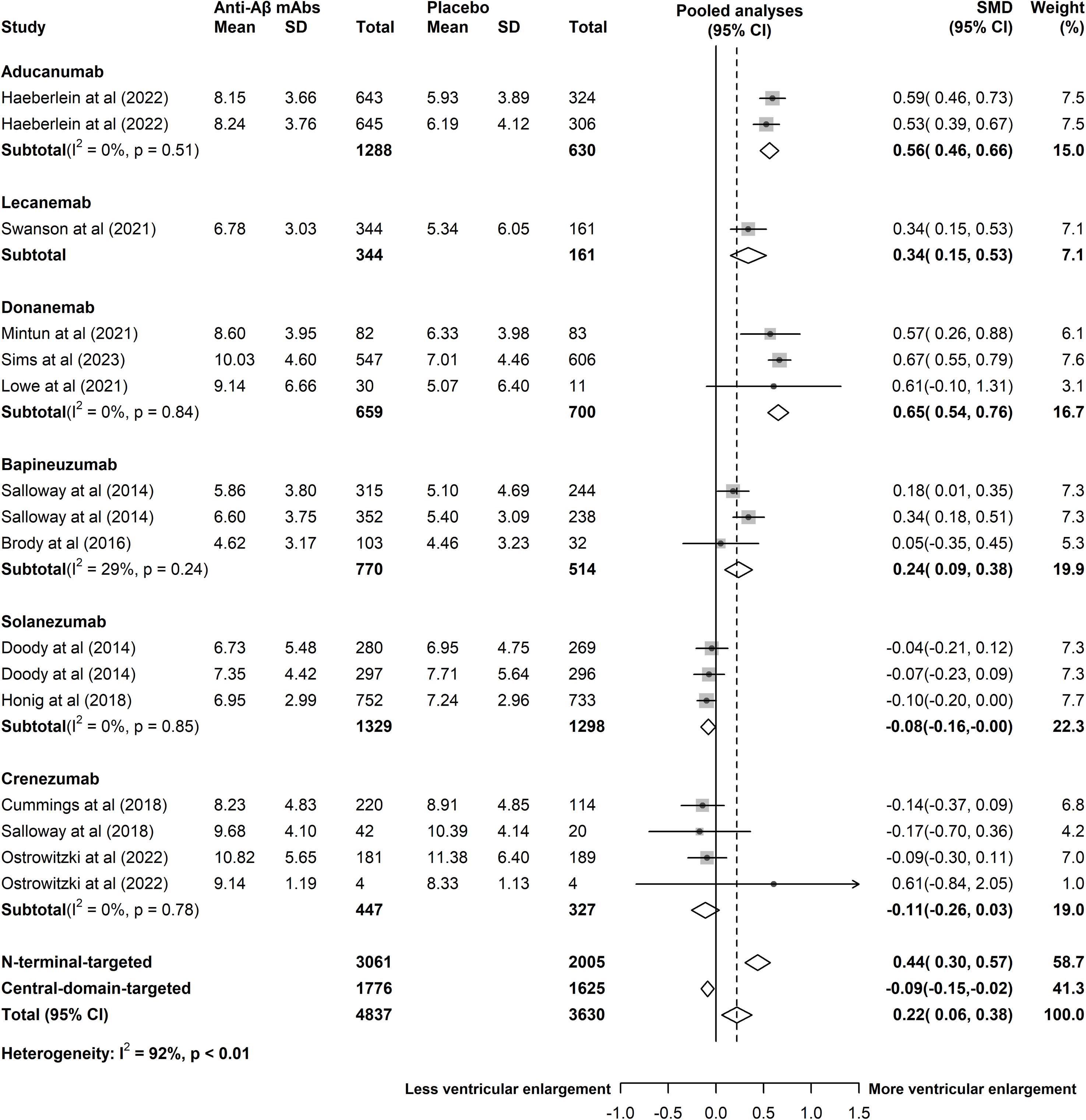
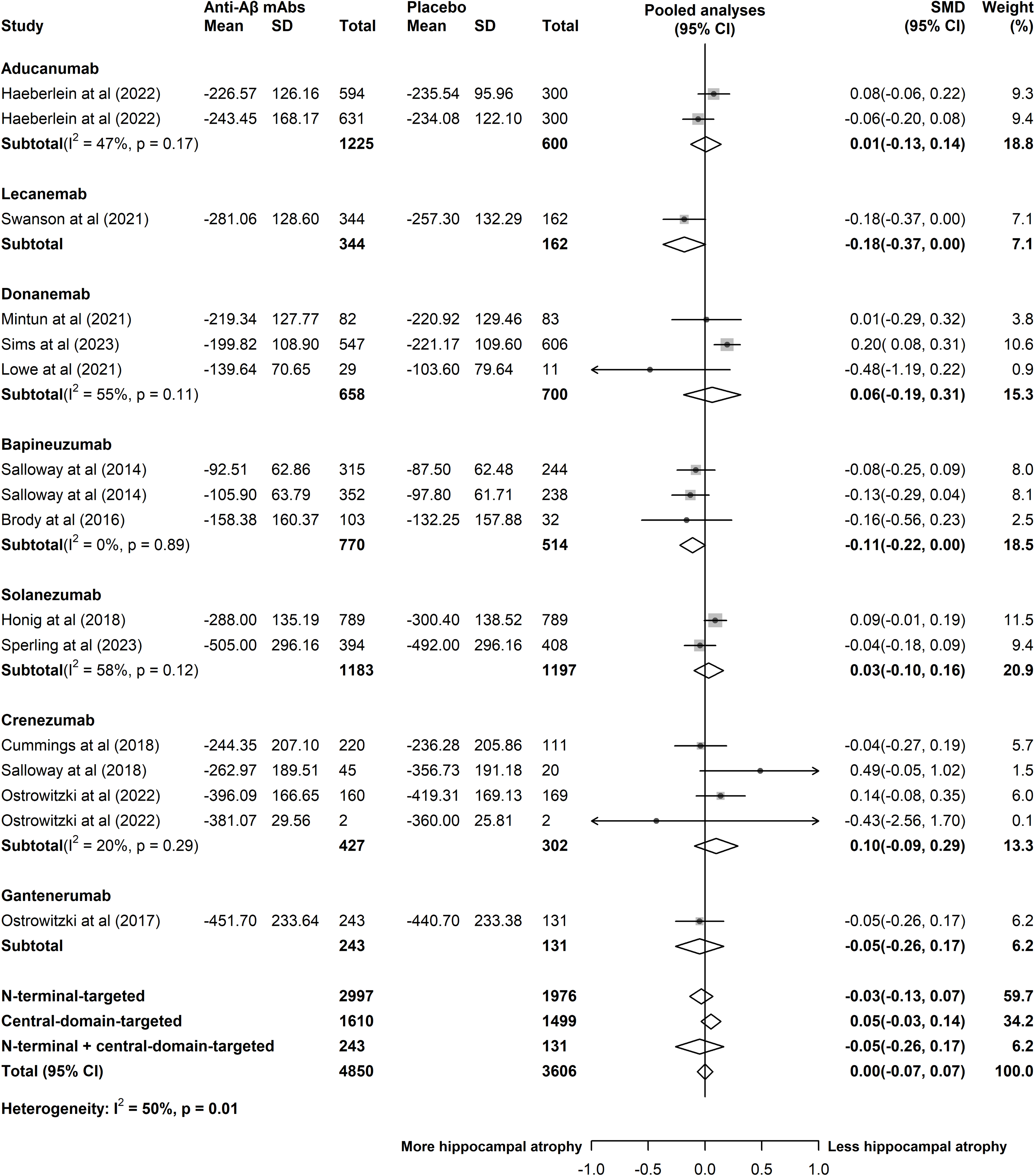
Forest plot of standardized mean differences for amyloid positron emission tomography (PET). Weights were assigned by random effects analysis. SMD: standardized mean difference.

These anti-Aβ mAbs significantly accelerated whole-brain atrophy (SMD: -0.14, 95% CI: -0.26 to -0.03; I^2^ = 89%) (negative change was indicated more whole-brain atrophy) (Figure 4A), and ventricle enlargement (SMD: 0.22, 95% CI: 0.06 to 0.38; I^2^ = 92%) (positive change was indicated more ventricle enlargement) (Figure 4B). However, there was no significant effect of the mAbs on hippocampal volumes compared to placebo treatment (SMD: 0.00, 95% CI: -0.07 to 0.07; I^2^ = 50%) (negative change was indicated more hippocampus atrophy) (Figure 4C). The funnel plots, which assess publication bias, demonstrated symmetrical distribution for the all three primary outcomes (Appendix 5.5-5.7). Additionally, the p-values from Egger’s test were 0.828, 0.845, and 0.276 respectively, further supporting the absence of publication bias. Lecanemab and donanemab obviously accelerated whole-brain atrophy [(lecanemab: SMD: -0.41, 95% CI: -0.59 to -0.22), (donanemab: SMD: -0.68, 95% CI: -0.96 to -0.40; I^2^ = 59%)] (Figure 4A) and ventricular enlargement [(lecanemab: SMD: 0.34, 95% CI: 0.15 to 0.53), (donanemab: SMD: 0.65, 95% CI: 0.54 to 0.76; I^2^ = 0%)] (Figure 4B). However, neither lecanemab nor donanemab had a significant impact on hippocampal volumes [(lecanemab: SMD: -0.18, 95% CI: -0.37 to 0.00), (donanemab: SMD: 0.06, 95% CI: -0.19 to 0.31; I^2^ = 55%)] (Figure 4C). On the other hand, aducanumab and bapineuzumab were found to only accelerate ventricular enlargement [(aducanumab: SMD: 0.56, 95% CI: 0.46 to 0.66; I^2^ = 0%), (bapineuzumab: SMD: 0.24, 95% CI: 0.09 to 0.38; I^2^ = 29%)] (Figure 4A-C). The subgroup tests showed that N-terminal-targeted mAbs had a stronger efficacy than central-domain-targeted in whole-brain atrophy [(N-terminal-targeted: SMD: -0.24, 95% CI: -0.41 to -0.07), (central-domain-targeted: SMD: 0.00, 95% CI: -0.08 to 0.08)] (Figure 4A) and ventricular enlargement [(N-terminal-targeted: SMD: 0.44, 95% CI: 0.30 to 0.57), (central-domain-targeted: SMD: -0.09, 95% CI: -0.15 to -0.02)] (Figure 4B). However, there was no clear impact of preferential Aβ epitope targeting on hippocampal volumes [(N-terminal-targeted: SMD: -0.03, 95% CI: -0.13 to 0.07), (central-domain-targeted: SMD: 0.05, 95% CI: -0.03 to 0.14), (N-terminal + central-domain-targeted: SMD: -0.05, 95% CI: -0.26 to 0.17)] (Figure 4C).

**Figure 4.**
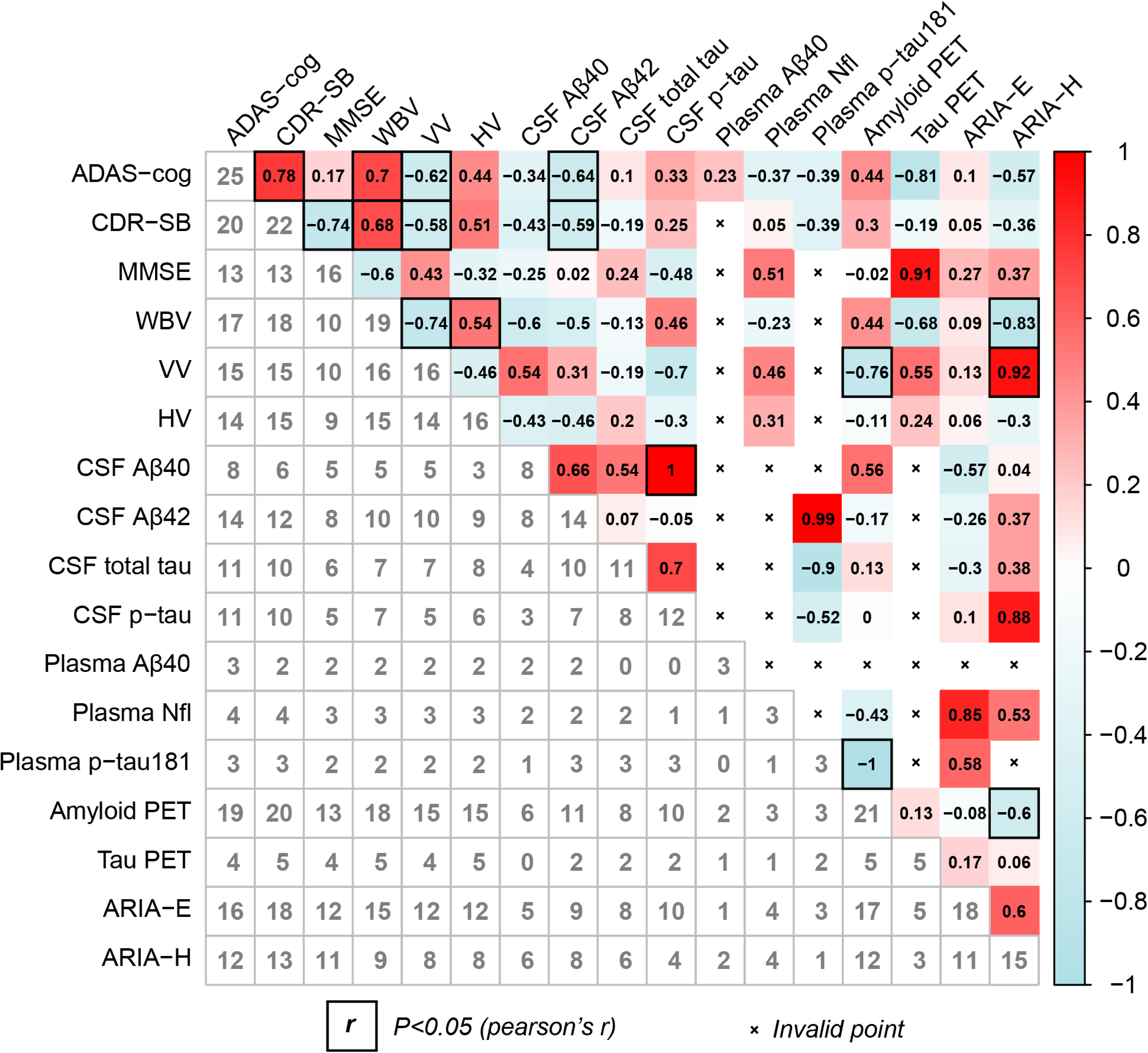
Forest plot of standardized mean differences for volumetric Magnetic Resonance Imaging (vMRI). Data were presented for volumes of the whole brain (A), volumes of ventricle (B), and volumes of hippocampus (C). Weights were assigned by random effects analysis. SMD: standardized mean difference.

Compared to placebo, the mAbs substantially increased CSF Aβ40 levels (SMD: 0.54, 95% CI: 0.11 to 0.98; I^2^ = 82%), and CSF Aβ42 levels (SMD: 0.78, 95% CI: 0.44 to 1.12; I^2^ = 85%) (Appendix 7.1-7.2). In terms of other CSF biomarkers, the meta-analysis showed a decrease in the levels of total tau, p-tau, and neurogranin in CSF when comparing the mAbs to placebo [(CSF total tau: SMD: -0.44, 95%CI: -0.73 to -0.16; I^2^ = 68%), (CSF p-tau: SMD: -0.59, 95% CI: -0.94 to -0.25; I^2^ = 88%), (CSF neurogranin: SMD: -0.52, 95% CI: -0.71 to -0.34; I^2^ = 0%)] (Appendix 7.3-7.4, 7.7). For CSF p-tau181 and CSF Nfl, there was no significant difference between the mAbs and placebo, regarding the mean change from baseline [(CSF p-tau181: SMD: -0.39, 95% CI: -0.94 to 0.16; I^2^ = 82%), (CSF Nfl: SMD: -0.15, 95% CI: -0.40 to 0.11; I^2^ = 0%)] (Appendix 7.5-7.6). The funnel plot analysis suggested that there was no evidence of publication bias or other small study effects for CSF Aβ42, total tau, and p-tau (Egger test, p = 0.418, 0.640, and 0.541, respectively) (Appendix 5.8-5.10). In the subgroup analysis, it was observed that N-terminal-targeted mAbs exhibited a superior efficacy in terms of CSF levels of total tau (SMD: -0.48, 95% CI: -0.68 to -0.28) and p-tau (SMD: -0.49, 95% CI: -0.72 to -0.26) (Appendix 7.3-7.4). While, central-domain-targeted mAbs showed a significant difference in CSF levels of Aβ40 (SMD: 0.83, 95% CI: 0.12 to 1.55) and Aβ42 (SMD: 0.98, 95% CI: 0.43 to 1.53) (Appendix 7.1-7.2). In addition, the anti-Aβ mAbs had no significant effect on plasma Aβ40 (SMD: 1.61, 95% CI -0.53 to 3.75; I^2^ = 100%) and Nfl levels (SMD: -0.06, 95% CI -0.18 to 0.06; I^2^ = 0%) (Appendix 7.8-7.9). However, they showed a reduction in plasma p-tau181 levels (SMD: -0.63, 95% CI: -0.72 to -0.54; I^2^ = 27%) (Appendix 7.10).

Compared with placebo, the mAbs substantially increased the risk of developing ARIA-E (RR: 9.79, 95% CI 7.83 to 12.26; I^2^ = 39%) and ARIA-H (RR: 1.28, 95% CI 0.98 to 1.67; I^2^ = 78%) (Appendix 7.12-7.13). The funnel plots, which assess publication bias, demonstrated symmetrical distribution for outcomes (Appendix 5.11-5.12). The symmetrical distribution of funnel plots and non-significant p-values from Egger’s test (p-values of 0.475 and 0.647) suggested the absence of publication bias in the assessed outcomes. Upon subgroup analysis, it was found that all mAbs, except solanezumab (RR: 1.59, 95% CI 0.47 to 5.39; I^2^ = 14%) and crenezumab (RR: 1.59, 95% CI 0.20 to 12.92; I^2^ = 0%) were responsible for the increased risk of ARIA-E (Appendix 7.12). Additionally, lecanemab (RR: 1.86, 95% CI 1.48 to 2.34; I^2^ = 57%) and donanemab (RR: 1.99, 95% CI 1.56 to 2.54; I^2^ = 0%) significantly increased the risk of ARIA-H (Appendix 7.13). The incidences of ARIA-E and ARIA-H were significantly higher in patients treated with N-terminal-targeted mAbs, compared to the other agents based on different mAbs recognition of Aβ epitopes [ARIA-E: (N-terminal-targeted: RR: 10.79, 95% CI 8.61 to 13.52), (central-domain-targeted: RR: 1.67, 95% CI 0.65 to 4.28), (N-terminal+central-domain-targeted: RR: 13.27, 95% CI 3.26 to 54.05) (Appendix 7.12); ARIA-H: (N-terminal-targeted: RR: 1.94, 95% CI 1.64 to 2.29), (central-domain-targeted: RR: 0.92, 95% CI 0.81 to 1.06), (N-terminal+central-domain-targeted: RR: 0.89, 95% CI 0.60 to 1.31), (C-terminal-targeted: RR: 1.00, 95% CI 0.10 to 9.96)] (Appendix 7.13).

The meta-regression analysis revealed a statistically significant positive association between the percentage of participants on symptomatic medication for AD and the ADAS-cog score (β = 0.190, p = 0.021) (Appendix 8.1). Additionally, the analysis showed that baseline CDR-SB had a significant negative impact on CDR-SB (β = -0.058, p = 0.012) (Appendix 8.2). In further meta-regression analysis, it was found that there was a significant positive relationship between the percentage of participants on symptomatic medication for AD (β = 25.943, p = 0.002), baseline participant mean age (β = 0.422, p = 0.024), and tau PET (Appendix 8.5). Furthermore, significant heterogeneity was observed in the relationship between baseline MMSE (β = 0.074, p = 0.041) and CDR-SB (β = -0.283, p = 0.013), and volumetric changes in the ventricles (Appendix 8.8). A statistically significant association was found in the meta-regression analysis, which examined the association between baseline MMSE score and CSF p-tau (β = -0.141, p = 0.018) (Appendix 8.11). Moreover, CDR-SB (β = 0.578, p = 0.043) as well as MMSE (β = -0.219, p = 0.023), the mean age (β = -0.705, p < 0.001), and the percentage of APOE4 carrier (β = 8.251, p < 0.001) at baseline were found to have a significant impact on CSF p-tau181 in the meta-regression analysis (Appendix 8.13). Furthermore, significant heterogeneity was observed in the relationship between the percentage of female participants at baseline and plasma Aβ40 in the meta-regression analysis (β = 49.200, p < 0.001) (Appendix 8.16). However, additional meta-regression analysis based on mean baseline MMSE, CDR-SB, mean age, percentage of APOE4 carriers, percentage of females, and percentage of participants on symptomatic medication for AD did not suggest that these factors were effect modifiers of MMSE, amyloid PET, volumetric changes in the whole brain and hippocampus, CSF Aβ40, CSF Aβ42, CSF total tau, CSF Nfl, CSF Neurogranin, Plasma Nfl, Plasma p-tau181, ARIA-E, and ARIA-H (Appendix 8.1-8.20).

Upon sequentially removing each study from the sensitivity analysis, the p-value showed variation from 0.05 to 0.08 when excluding phase III studies NCT02484547 of aducanumab ^14^, NCT03887455 of lecanemab ^15^, NCT04437511 of donanemab ^16^, and NCT01900665 of solanezumab ^10^, which indicates that these studies had a substantial impact on the overall result on CDR-SB (Appendix 9.2). The sensitivity analysis utilizing the one-study-removed procedure in the meta-analysis revealed a notable change in the p-value of the results for CSF Aβ40 following the exclusion of NCT01900665 of solanezumab ^10^ (Appendix 9.9). Similarly, the application of the one-study-removed method exposed a qualitative shift in the p-value of the results for CSF p-tau181 when NCT01397578 of crenezumab ^38^ was removed (Appendix 9.13). Upon sequentially removing each study from the analysis, the p-value was less than 0.05 when excluding NCT01230853 of lecanemab ^51^, NCT02008357 of solanezumab ^58^, and NCT02670083 of crenezumab ^11^, indicating that these studies had a substantial impact on the overall result of ARIA-H (Appendix 9.20). Despite the influence of individual studies, the overall effect size remained significant and consistent, suggesting the robustness of the meta-analysis results on ADAS-cog, MMSE, amyloid PET, tau PET, volumes of the whole brain, hippocampus, as well as ventricle, amyloid PET, other CSF and plasma biomarkers, and ARIA-E (Appendix 9.1-9.20). Additionally, the results of sensitivity analysis also indicated that substituting the effect model did not have a meaningful impact on the stability of the results (Appendix 9.1-9.20).

Additionally, Pearson’s correlation coefficients were calculated to assess the interrelationships between alterations in cognitive performance, Aβ deposition, changes in AD biomarkers, and the risks of ARIA for both all mAbs and N-terminal-targeted mAbs. The results of this analysis are presented below:

The effects of all mAbs on volumetric changes of the whole-brain were found to be positively correlated with their effects on decreasing (improving) ADAS-cog scores (Pearson’s r = +0.70, p = 0.002) and CDR-SB scores (Pearson’s r = +0.68, p = 0.002). Furthermore, the effects of all mAbs on volumetric changes of the ventricles were found to be negatively correlated with their effects on decreasing (improving) ADAS-cog scores (Pearson’s r = -0.62, p = 0.014) or CDR-SB scores (Pearson’s r = -0.58, p = 0.022). The association between cognition and vMRI of mAbs targeting N-terminal yielded similar results to all mAbs. Interestingly, although no correlation between volumetric changes of the hippocampus and cognitive outcome effect sizes was found across all drugs, there was a significant association between volumetric changes of the hippocampus and CDR-SB scores specifically in mAbs targeting the N-terminal (Pearson’s r = -0.80, p = 0.017) (Figure 5A, B).

**Figure 5.**
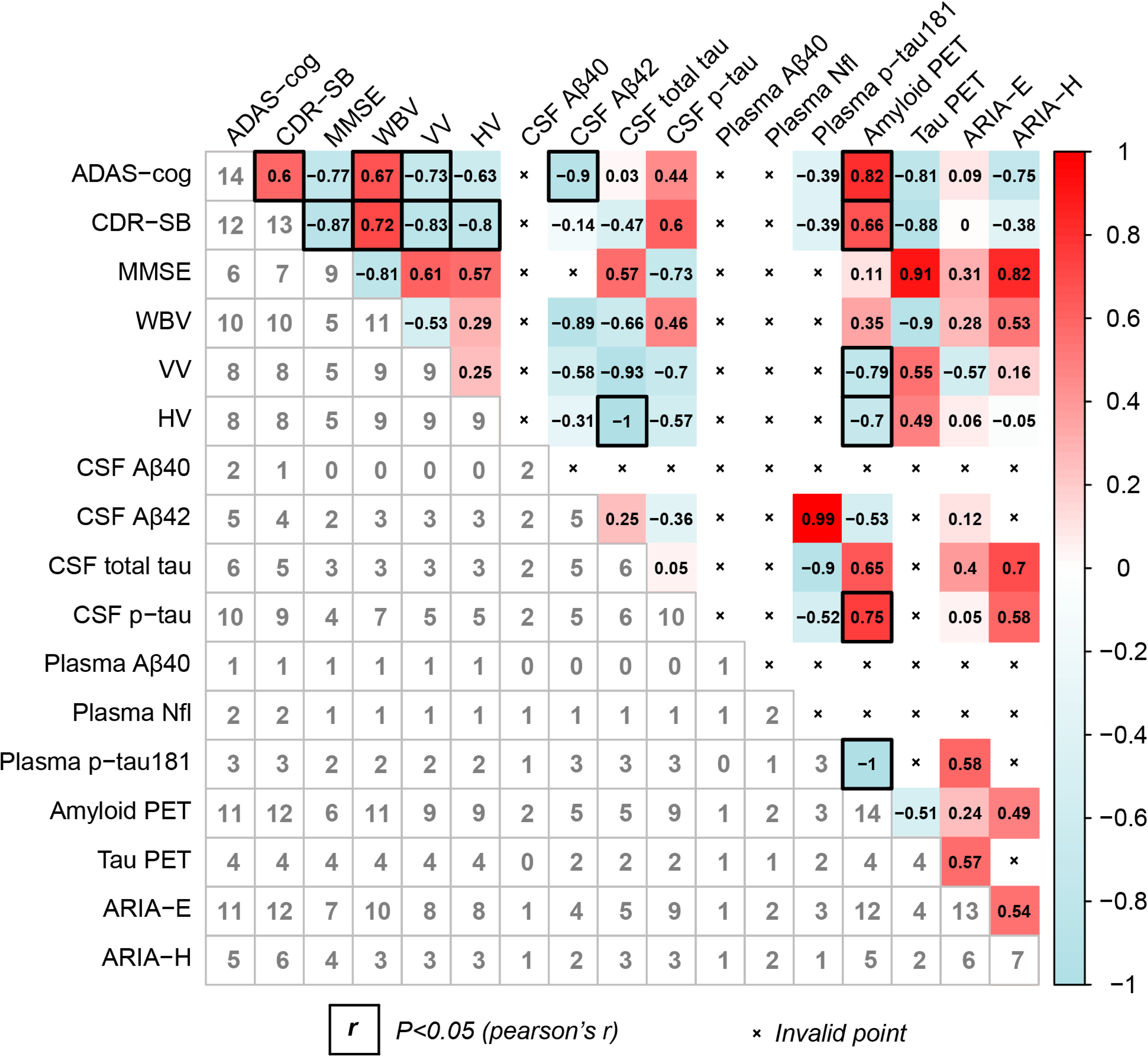
Heat map of Pearson’s correlation coefficients among Aβ deposition, variations in AD biomarkers, alterations in cognitive performance, and the risks of ARIA-E as well as ARIA-H across multiple studies. Data were presented for Pearson’s correlation coefficients among studies of all monoclonal antibodies (A), and Pearson’s correlation coefficients among studies of N-terminal-targeted monoclonal antibodies (B), with the r value. Results of Pearson’s correlation coefficients were presented in the upper right half and the number of studies from Pearman’s correlation coefficients were presented in the left lower half. Correlations with statistical significance were emphasized with black frames. ADAS-cog: Alzheimer’s Disease Assessment Scale-cognitive subscale, CDR-SB: Clinical Dementia Rating scale-Sum of Boxes, MMSE: Mini Mental State Examination, WBV: whole brain volumes, VV: ventricle volumes, HV: hippocampal volumes, CSF: cerebrospinal fluid, p-tau: phosphorylated tau, Nfl: Neurofilament Light, p-tau 181: phosphorylated tau protein at position 181, PET: positron emission tomography, ARIA-E: Amyloid-Related Imaging Abnormalities-Edema, ARIA-H: Amyloid-Related Imaging Abnormalities-Hemorrhag.

The levels of CSF Aβ42 of all mAbs exhibited a significant negative correlation with ADAS-cog scores (Pearson’s r = -0.64, p = 0.013) and CDR-SB scores (Pearson’s r = -0.59, p = 0.044). Specifically, the levels of CSF Aβ42 for mAbs targeting the N-terminal were found to have a strong negative association with ADAS-cog scores (Pearson’s r = -0.90, p = 0.040). (Figure 5A, B).

No significant correlation was observed between amyloid PET of all mAbs and either the effect sizes of CDR-SB (Pearson’s r = +0.30, p = 0.197) or ADAS-cog (Pearson’s r = +0.44, p = 0.060). While, the reduction in amyloid PET resulting from mAbs targeting the N-terminal region showed a positive correlation with their improvement in ADAS-cog scores (Pearson’s r = +0.82, p = 0.002) and CDR-SB scores (Pearson’s r = +0.66, p = 0.019) (Figure 5A, B).

The impact of both all antibodies (Pearson’s r = -0.76, p = 0.001) and N-terminal-targeted mAbs (Pearson’s r = -0.79, p = 0.011) on reducing amyloid PET was inversely correlated with their effects on volumetric changes of the ventricle. Specifically, there was a significant association between reducing amyloid PET and volumetric changes of the hippocampus, specifically in N-terminal-targeted mAbs (Pearson’s r = -0.70, p = 0.035) (Figure 5A, B).

The effects of all antibodies on the incidence of ARIA-H were found negatively associated with their impact on volumetric changes of the whole-brain (Pearson’s r = -0.83, p = 0.005) and ventricular volume (Pearson’s r = 0.92, p = 0.001). No significant correlation was observed between ARIA and vMRI of N-terminal-targeted mAbs (Figure 5A, B).

The effects of mAbs on reducing amyloid PET were found negatively associated with their effects on the incidence of ARIA-H (Pearson’s r = -0.60, p = 0.037). No significant correlation was observed between reducing amyloid PET of mAbs targeting the N-terminal and ARIA (Figure 5A, 5B).

## Discussion

The present meta-analysis study was strengthened by including the latest findings from donanemab ^16^, lecanemab ^15^, and solanezumab ^58^ phase III results, providing statistically significant effects. Robust data syntheses of all included 34 studies showed statistical improvements in cognitive outcomes (ADAS-cog, CDR-SB and MMSE) with the use of anti-Aβ mAbs. The meta-analysis also showed that the mAbs statistically reduced amyloid burden (amyloid PET) and some AD biomarkers (the levels of total tau, p-tau, and neurogranin in CSF, as well as GFAP and p-tau181 levels in plasma), improved Aβ biomarkers (CSF Aβ40 levels and CSF Aβ42 level), accelerated whole-brain atrophy and ventricle enlargement, but also increased the risk of the hallmark adverse events, ARIA-E and ARIA-H. Moreover, remarkable effectiveness was observed in AD patients treated with the mAbs that specifically targeting N-terminus of Aβ, as reflected in subgroup analyses of the mAbs targeting different epitopes of Aβ. Notably, our findings suggested a significant and interconnected relationship among cognition, Aβ deposition, brain atrophy, and the risk of ARIA after anti-Aβ mAbs treatment, especially N-terminal-targeted mAbs.

However, the meta-analysis also had some limitations. It was based on study-level data, which might lack detailed information for certain patients, potentially affecting the internal or external credibility of the findings. The inclusion of phase I, II, and single-dose studies might limit the robustness of the results. Furthermore, missing or unpublished data of CSF or plasma biomarkers might cause potential bias in assessing the correlation between mAbs treatment and these biomarkers.

The robust data syntheses of all included studies showed statistically improvements in cognitive outcomes, as measured by ADAS-cog, MMSE, and CDR-SB. However, the clinical benefits of mAbs had rather small effect sizes (-0.08 for ADAS-cog, -0.06 for CDR-SB and 0.06 for MMSE), despite their statistical significance, which might have limited clinical relevance ^62^ and the observed benefit was not superior to that of palliative acetylcholinesterase inhibitor drugs ^24,63^. Importantly, two mAbs targeting N-terminal domain of Aβ (donanemab and lecanemab) exhibited a moderate effect size on CDR-SB (SMD = -0.28 to -0.17), a crucial measure for establishing efficacy in clinical trials for AD ^64^. The treatment effect resulted in an approximate 25% reduction in disease progression observed over a period of 1.5 years in CLARITY-AD ^15^. This reduction would be equivalent to a delay of 4-5 months in clinical deterioration related to the disease ^65^.

The mAbs demonstrated a robust target engagement, with substantial effect sizes observed for reductions in amyloid PET (SMD: -1.13). Our analysis revealed no statistically significant correlation between the reduction of amyloid deposition measured by PET and cognitive improvements on ADAS-cog or CDR-SB across all mAbs, whereas N-terminal-targeted mAbs exhibited such a correlation. The implication suggests that agents with greater reductions of Aβ deposition, such as N-terminal-targeted mAbs, might lead to more significant improvements in cognitive and functional abilities. These findings further support the pivotal role of Aβ plaques in driving cognitive decline in AD and underscore the clinical benefits associated with Aβ removal from the brain ^66-68^.

The subgroup analysis results pertaining to the targeting of Aβ epitopes by mAbs yielded noteworthy findings. Specifically, mAbs targeting N-terminus of Aβ exhibited a remarkable capacity of removing Aβ, surpassing other antibody types. This outcome was not serendipitous, as the prior evidence had found the ability of N-terminal-targeted antibodies to impede Aβ aggregation, neutralize Aβ neurotoxicity, and disassemble pre-existing Aβ fibrils ^69^. Moreover, these mAbs have showed a superior efficacy in reducing cerebral amyloid burden ^69,70^. Aβ40 peptide monomers undergo aggregation and form oligomers with a three-unit multiplicity through seeded fibril growth ^33^. Characterized by solid-state nuclear magnetic resonance and electron microscopy, this aggregation process leaves the N-termini of Aβ exposed to the solvent, while the hydrophobic C-termini remains sequestered within the trimer core^24^. Additionally, the tightly packed, hydrophobic arrangement of these trimers facilitates the assembly of amyloid fibrils, ensuring all N-termini within the fibril remain solvent-exposed ^33^. Although not yet documented for the Aβ42 peptide, this structural arrangement provides a rationale for directing mAbs designed to bind the N-terminus of Aβ ^25^. It underlies the observed efficacy of N-terminal antibodies in eliminating Aβ oligomers. Moreover, the effectiveness of N-terminal mAbs in fostering the clearance of aggregated Aβ may also be attributed to their potential to activate microglia and stimulate phagocytosis ^25^. These shared characteristics have been postulated as a contributing factor of the favorable outcomes observed with N-terminal mAbs ^34,70-72^

It is widely proposed that the effectiveness of the mAbs therapies may be attributed to their capability to target various forms of Aβ, including monomers, oligomers, and fibrils ^24,25,27^. Additionally, this notion is supported by a growing body of evidences, which indicating that soluble Aβ oligomers exhibit a greater neurotoxicity compared to fibrillar aggregates and better correlate with the clinical manifestations of AD ^28-30^. Previous meta-analysis, however, did not yield positive results when subgrouping was performed based on targeting Aβ forms ^31^. This suggests that while mAbs with widely different characteristics may have some positive effects on AD, the lack of Aβ target specificity is not a desirable feature of an “ideal” antibody ^31^. After conducting a comprehensive analysis of these three mAbs with remarkable efficacy, aducanumab, lecanemab, and donanemab, it becomes apparent that there was an absence of consistent patterns in terms of their specific targeting towards diverse forms of Aβ aggregates ^14-16^. Each drug tackles different aspects of Aβ aggregates, such as soluble oligomers, insoluble fibrils, and deposited Aβ ^73^, without exhibiting a clear or uniform trend. It is worth noting that all of these drugs fall under the category of N-terminal-targeted mAbs, which potentially be a critical aspect of their mechanism of action ^74,75^.

Aβ immunotherapies are more likely to have beneficial effects at the onset or at the early stage of AD with minimal clinical symptoms ^25,73,76^. Nonetheless, it is crucial for us to acknowledge that the successful eradication of Aβ plaques plays a pivotal role in mitigating the advancement of the disease, rather than confining our attention to the exclusive treatment of AD’s initial phase ^24,77^. The A4 trial of solanezumab, a mAb that binds to the central-domain of the Aβ monomer, marked the first clinical trial to assess a disease-modifying therapy in cognitively normal individuals displaying abnormal Aβ PET scan results ^58^. Throughout a 4.5-year follow-up period, solanezumab failed to exhibit efficacy in terms of clearing Aβ plaques, and no statistically significant disparities were noted between solanezumab and placebo with respect to the primary outcome measure, the Preclinical Alzheimer Cognitive Composite (PACC) utilized for evaluating cognitive function ^58^. In contrast, the phase III trial of lecanemab and donanemab, both N-terminal-targeted mAbs, not only showcased the successful attenuation of cognitive decline, but also underscored their potency in eliminating Aβ ^15,16^. Additionally, the TRAILBLAZER-ALZ 2 trial employed an innovative approach to mitigate heterogeneity in disease progression by incorporating tau PET as an inclusion criterion alongside Aβ positivity ^16^. This strategic inclusion demonstrated that treatment effects were most pronounced in participants with mild-to-moderate tau burden, as evidenced by stratified analyses of tau PET burden ^16^. In summary, the essence of our argument is that the premise for earlier intervention should be centered around the effective elimination of Aβ. It should firstly identify appropriate targets in the development of disease-modifying interventions for AD, specifically focusing on N-terminal of Aβ, then followed by the subsequent step of identifying individuals suitable for treatment through biomarkers^78^.

In addition to meeting all outcomes of cognition and function, mAbs treatment also yielded differences in AD biomarkers in CSF and plasma, supporting a downstream pathophysiologic effect of amyloid-β plaque removal. The mAbs exhibited robust biomarker responses, with moderate effect sizes observed for CSF Aβ40, CSF Aβ42, CSF p-Tau, CSF total Tau, plasma GFAP and plasma pTau181. These findings provide further evidences for the potential benefits of targeting Aβ plaques in the treatment of AD, particularly in terms of reducing the burden of tau pathology ^79,80^.

The findings suggested a potential acceleration of brain atrophy and ventricle enlargement but not hippocampal atrophy with mAbs therapies, particularly N-terminal-targeted mAbs. It was likely that reduction in Aβ plaque volume could account for the degree of brain volume change, which reported in clinical trials of anti-Aβ drugs ^36,81^. It is noteworthy that while brain atrophy is commonly associated with cognitive decline in AD ^36,82^, this meta-analysis indicated that brain atrophy might not contribute to cognitive decline and might even decelerate disease progression. Moreover, the preservation of hippocampal volume was observed through correlation analysis between the impact of N-terminal-targeted mAbs on hippocampal volumetric changes with its effect on reducing CDR-SB and amyloid PET reduction. It was evident that Aβ clearance did not result in hippocampal atrophy, ultimately preserved cognitive functions in AD patients. The hippocampus plays a crucial role in spatial and declarative memory formation, encompassing the acquisition, consolidation, and retrieval of information ^83^, further supporting a disease modifying effect of mAbs therapy. The findings of our study further hypothesized that mAbs therapy had the potential efficacy to effectively mitigate cognitive decline by reducing amyloid levels and preserving hippocampal volume. The removal of Aβ, however, has been associated with an expansion in ventricular volume and a greater reduction in brain volume. The functional significance is still uncertain.

The ARIA (ARIA-E and ARIA-H) risk was high for all mAbs, except for the central-domain-targeted drugs, crenezumab and solanezumab. The higher ARIA risk might mainly be attributed to the significant reduction of brain amyloid level by N-terminal-targeted antibodies. Additionally, the risk of ARIA was strongly correlated with reductions in amyloid PET (Pearson’s r = -0.60) and brain atrophy, as well as ventricle enlargement (Pearson’s |r| = 0.83-0.92). The aforementioned findings also provided further support for the previous hypothesis that the mobilization of brain Aβ through anti-amyloid immunotherapy might be causally associated with both treatment effectiveness and the risk of ARIA ^31,84,85^. The exact mechanism is thought that clearance of Aβ by mAbs may cause inflammation and disruption of the blood-brain barrier, leading to increased vascular permeability ^86^. This concept is supported by empirical evidence that the crucial role of perivascular macrophages in modulating vascular permeability is associated with cerebral amyloid angiopathy (CAA) and the occurrence of microhemorrhages is related to amyloid immunotherapy^87^.

ARIA is shown to be a dose-dependent hallmark side effect of anti-Aβ mAbs treatment ^88,89^. It cannot fail to make us notice of the fact that venetoclax, a targeted therapy drug, works by inhibiting the function of the BCL-2 protein, effectively kills a large number of cancer cells ^90^. However, in the meantime, tumour lysis syndrome (TLS), one of the common side effects, is happened. To reduce the risk of TLS, researcher used a process known as dose titration to allow the body to adjust to the drug’s effects and helps to prevent a sudden and massive breakdown of cancer cells ^91^. As a result of the successful implementation of the dose titration, the researchers completed the Phase III clinical trial and obtained approval from the FDA ^92^. The research and development of venetoclax can serve as a tangible example to help us understand how to reduce the risk of ARIA caused by the anti-amyloid immunotherapy. According to this result, it is also believed that administering Aβ immunotherapies during the early stages of AD with low Aβ deposition may potentially reduce the risk of ARIA caused by the extensive clearance of Aβ in the later stages of the disease.

In conclusion, in our meta-analysis, it has not escaped our notice that N-terminal-targeted Aβ antibodies (not C-terminal-targeted, central-domain-targeted, nor N-terminal+central-domain-targeted antibodies) substantially alleviate cognitive deterioration and noticeably reduce plaque burden in AD patients. Structure determines function. This approach may be regarded as a potentially groundbreaking strategy and should not be underestimated in AD immunotherapy development. However, the underlying mechanisms of this approach is still need to be elucidated. Further investigations are needed to explore whether a better efficacy can be achieved through stratification with biomarkers and to explore the potential benefits of combination therapies to enhance therapeutic efficacy.

## Data Availability

All data produced in the present study are available upon reasonable request to the authors

## Contributors

SJZ conceived the meta-analysis and developed its design with YHQ. YHQ performed the systematic literature search with LLL. YHQ and DW extracted data. YHQ and ML performed the statistical analyses. YHQ wrote the first draft of the manuscript with JZ and SJZ. All other authors contributed to data interpretation and manuscript review. All authors had access to all the data, and YHQ, LLL and ML accessed and verified the data. YFC and SJZ take final responsibility for the decision to submit for publication.

## Declaration of interests

We declared no competing interests.

## Data sharing

All analysed summary data were extracted from published sources that are publicly available or were requested from individual trials (and are provided in the presented tables and figures). For the purpose of open access, the authors have applied a Creative Commons Attribution (CC BY) licence to any Author-Accepted Manuscript version arising.

## Acknowledgments

This work was supported by National Natural Science Foundation of China (No. 82004430, 82174310).

## Notes

### Competing Interest Statement

The authors have declared no competing interest.

### Author Declarations

All data produced in the present study are available upon reasonable request to the authors

