## Supplementary material for "N-terminal-targeted anti-amyloid monoclonal antibodies illuminate the therapy for Alzheimer’s disease: a systematic review and comprehensive meta-analysis": //

### Supplementary appendix

### Appendix

8.5.5 Meta-regressions of tau PET by the percentage of female participants at the baseline ... 96
8.5.6 Meta-regressions of tau PET by the percentage of participants on anti-AD medication at the baseline ... 97
8.6 Volumes of the whole brain ... 98
8.6.1 Meta-regressions of volumes of the whole brain by baseline CDR-SB score 98
8.6.2 Meta-regressions of volumes of the whole brain by baseline MMSE score ... 99
8.6.3 Meta-regressions of volumes of the whole brain by the mean age at the baseline ... 100
8.6.4 Meta-regressions of volumes of the whole brain by the percentage of APOE4 carrier at the baseline ... 101
8.6.5 Meta-regressions of volumes of the whole brain by the percentage of female participants at the baseline ... 102
8.6.6 Meta-regressions of volumes of the whole brain by the percentage of participants on anti-AD medication at the baseline ... 103
8.7 Volumes of hippocampus ... 104
8.7.1 Meta-regressions of volumes of hippocampus by baseline CDR-SB score . 104
8.7.2 Meta-regressions of volumes of hippocampus by baseline MMSE score ... 105
8.7.3 Meta-regressions of volumes of hippocampus by the mean age at the baseline ... 106
8.7.4 Meta-regressions of volumes of hippocampus by the percentage of APOE4 carrier at the baseline ... 107
8.7.5 Meta-regressions of volumes of hippocampus by the percentage of female participants at the baseline ... 108
8.7.6 Meta-regressions of volumes of hippocampus volumes of hippocampus by the percentage of participants on anti-AD medication at the baseline ... 109
8.8 Volumes of ventricle ... 110
8.8.1 Meta-regressions of volumes of ventricle by baseline CDR-SB score ... 110
8.8.2 Meta-regressions of volumes of ventricle by baseline MMSE score ... 111
8.8.3 Meta-regressions of volumes of ventricle by the mean age at the baseline 112
8.8.4 Meta-regressions of volumes of ventricle by the percentage of APOE4 carrier at the baseline ... 113
8.8.5 Meta-regressions of volumes of ventricle by the percentage of female participants at the baseline ... 114
8.8.6 Meta-regressions of volumes of ventricle by the percentage of participants on anti-AD medication at the baseline ... 115
8.9 CSF Aβ40 ... 116
8.9.1 Meta-regressions of CSF Aβ40 by baseline CDR-SB score ... 116
8.9.2 Meta-regressions of CSF Aβ40 by baseline MMSE score ... 117
8.9.3 Meta-regressions of CSF Aβ40 by the mean age at the baseline ... 118
8.9.4 Meta-regressions of CSF Aβ40 by the percentage of APOE4 carrier at the baseline ... 119
8.9.5 Meta-regressions of CSF Aβ40 by the percentage of female participants at the baseline ... 120

8.9.6 Meta-regressions of CSF Aβ40 by the percentage of participants on anti-AD medication at the baseline ........ 121
8.10 CSF Aβ42 ........ 122
8.10.1 Meta-regressions of CSF Aβ42 by baseline CDR-SB score ........ 122
8.10.2 Meta-regressions of CSF Aβ42 by baseline MMSE score ........ 123
8.10.3 Meta-regressions of CSF Aβ42 by the mean age at the baseline ........ 124
8.10.4 Meta-regressions of CSF Aβ42 by the percentage of APOE4 carrier at the baseline ........ 125
8.10.5 Meta-regressions of CSF Aβ42 by the percentage of female participants at the baseline ........ 126
8.10.6 Meta-regressions of CSF Aβ42 by the percentage of participants on anti-AD medication at the baseline ........ 127
8.11 CSF p-tau ........ 128
8.11.1 Meta-regressions of CSF p-tau by baseline CDR-SB score ........ 128
8.11.2 Meta-regressions of CSF p-tau by baseline MMSE score ........ 129
8.11.3 Meta-regressions of CSF p-tau by the mean age at the baseline ........ 130
8.11.4 Meta-regressions of CSF p-tau by the percentage of APOE4 carrier at the baseline ........ 131
8.11.5 Meta-regressions of CSF p-tau by the percentage of female participants at the baseline ........ 132
8.11.6 Meta-regressions of CSF p-tau by the percentage of participants on anti-AD medication at the baseline ........ 133
8.12 CSF total tau ........ 134
8.12.1 Meta-regressions of CSF total tau by baseline CDR-SB score ........ 134
8.12.2 Meta-regressions of CSF total tau by baseline MMSE score ........ 135
8.12.3 Meta-regressions of CSF total tau by the mean age at the baseline ........ 136
8.12.4 Meta-regressions of CSF total tau by the percentage of APOE4 carrier at the baseline ........ 137
8.12.5 Meta-regressions of CSF total tau by the percentage of female participants at the baseline ........ 138
8.12.6 Meta-regressions of CSF total tau by the percentage of participants on anti-AD medication at the baseline ........ 139
8.13 CSF p-tau181 ........ 140
8.13.1 Meta-regressions of CSF p-tau181 by baseline CDR-SB score ........ 140
8.13.2 Meta-regressions of CSF p-tau181 by baseline MMSE score ........ 141
8.13.3 Meta-regressions of CSF p-tau181 by the mean age at the baseline ........ 142
8.13.4 Meta-regressions of CSF p-tau181 by the percentage of APOE4 carrier at the baseline ........ 143
8.13.5 Meta-regressions of CSF p-tau181 by the percentage of female participants at the baseline ........ 144
8.13.6 Meta-regressions of CSF p-tau181 by the percentage of participants on anti-AD medication at the baseline ........ 145
8.14 CSF Nfl ........ 146
8.14.1 Meta-regressions of CSF Nfl by baseline CDR-SB score ........ 146

8.14.2 Meta-regressions of CSF Nfl by baseline MMSE score 147
8.14.3 Meta-regressions of CSF Nfl by the mean age at the baseline 148
8.14.4 Meta-regressions of CSF Nfl by the percentage of APOE4 carrier at the baseline 149
8.15 CSF Neurogranin 150
8.15.1 Meta-regressions of CSF Neurogranin by baseline CDR-SB score 150
8.15.2 Meta-regressions of CSF Neurogranin by baseline MMSE score 151
8.15.3 Meta-regressions of CSF Neurogranin by the mean age at the baseline 152
8.15.4 Meta-regressions of CSF Neurogranin by the percentage of APOE4 carrier at the baseline 153
8.15.5 Meta-regressions of CSF Neurogranin by the percentage of female participants at the baseline 154
8.15.6 Meta-regressions of CSF Neurogranin by the percentage of participants on anti-AD medication at the baseline 155
8.16 Plasma Aβ40 156
8.16.1 Meta-regressions of Plasma Aβ40 by baseline MMSE score 156
8.16.2 Meta-regressions of Plasma Aβ40 by the mean age at the baseline 157
8.16.3 Meta-regressions of Plasma Aβ40 by the percentage of female participants at the baseline 158
8.17 Plasma Nfl 159
8.17.1 Meta-regressions of Plasma Nfl by baseline CDR-SB score 159
8.17.2 Meta-regressions of Plasma Nfl by baseline MMSE score 160
8.17.3 Meta-regressions of Plasma Nfl by the mean age at the baseline 161
8.17.4 Meta-regressions of Plasma Nfl by the percentage of APOE4 carrier at the baseline 162
8.17.5 Meta-regressions of Plasma Nfl by the percentage of female participants at the baseline 163
8.17.6 Meta-regressions of Plasma Nfl by the percentage of participants on anti-AD medication at the baseline 164
8.18 Plasma p-tau181 165
8.18.1 Meta-regressions of plasma p-tau181 by baseline CDR-SB score 165
8.18.2 Meta-regressions of plasma p-tau181 by baseline MMSE score 166
8.18.3 Meta-regressions of plasma p-tau181 by the mean age at the baseline 167
8.18.4 Meta-regressions of plasma p-tau181 by the percentage of APOE4 carrier at the baseline 168
8.18.5 Meta-regressions of plasma p-tau181 by the percentage of female participants at the baseline 169
8.18.6 Meta-regressions of plasma p-tau181 by the percentage of participants on anti-AD medication at the baseline 170
8.19 ARIA-E 171
8.19.1 Meta-regressions of ARIA-E by baseline CDR-SB score 171
8.19.2 Meta-regressions of ARIA-E by baseline MMSE score 172
8.19.3 Meta-regressions of ARIA-E by the mean age at the baseline 173
8.19.4 Meta-regressions of ARIA-E by the percentage of APOE4 carrier at the

baseline 174
8.19.5 Meta-regressions of ARIA-E by the percentage of female participants at the baseline 175
8.19.6 Meta-regressions of ARIA-E by the percentage of participants on anti-AD medication at the baseline 176
8.20 ARIA-H 177
8.20.1 Meta-regressions of ARIA-H by baseline CDR-SB score 177
8.20.2 Meta-regressions of ARIA-H by baseline MMSE score 178
8.20.3 Meta-regressions of ARIA-H by the mean age at the baseline 179
8.20.4 Meta-regressions of ARIA-H by the percentage of APOE4 carrier at the baseline 180
8.20.5 Meta-regressions of ARIA-H by the percentage of female participants at the baseline 181
8.20.6 Meta-regressions of ARIA-H by the percentage of participants on anti-AD medication at the baseline 182
Appendix 9: Sensitivity Analysis 183
9.1 ADAS-cog 183
9.1.1 Leave-one-out method 183
9.1.2 Comparing the results between the fixed-effect and random-effects models 184
9.2 CDR-SB 185
9.2.1 Leave-one-out method 185
9.2.2 Comparing the results between the fixed-effect and random-effects model 186
9.3 MMSE 187
9.3.1 Leave-one-out method 187
9.3.2 Comparing the results between the fixed-effect and random-effects model 188
9.4 Amyloid PET 189
9.4.1 Leave-one-out method 189
9.4.2 Comparing the results between the fixed-effect and random-effects model 190
9.5 Tau PET 191
9.5.1 Leave-one-out method 191
9.5.2 Comparing the results between the fixed-effect and random-effects model 192
9.6 Volumes of the whole brain 193
9.6.1 Leave-one-out method 193
9.6.2 Comparing the results between the fixed-effect and random-effects model 194
9.7 Volumes of hippocampus 195
9.7.1 Leave-one-out method 195
9.7.2 Comparing the results between the fixed-effect and random-effects model 196

9.8 Volumes of ventricle 197
9.8.1 Leave-one-out method 197
9.8.2 Comparing the results between the fixed-effect and random-effects model 198
9.9 CSF Aβ40 199
9.9.1 Leave-one-out method 199
9.9.2 Comparing the results between the fixed-effect and random-effects model 200
9.10 CSF Aβ42 201
9.10.1 Leave-one-out method 201
9.10.2 Comparing the results between the fixed-effect and random-effects model 202
9.11 CSF p-tau 203
9.11.1 Leave-one-out method 203
9.11.2 Comparing the results between the fixed-effect and random-effects model 204
9.12 CSF total tau 205
9.12.1 Leave-one-out method 205
9.12.2 Comparing the results between the fixed-effect and random-effects model 206
9.13 CSF p-tau181 207
9.13.1 Leave-one-out method 207
9.13.2 Comparing the results between the fixed-effect and random-effects model 208
9.14 CSF Nfl 209
9.14.1 Leave-one-out method 209
9.14.2 Comparing the results between the fixed-effect and random-effects model 210
9.15 CSF Neurogranin 211
9.15.1 Leave-one-out method 211
9.15.2 Comparing the results between the fixed-effect and random-effects model 212
9.16 Plasma Aβ40 213
9.16.1 Leave-one-out method 213
9.16.2 Comparing the results between the fixed-effect and random-effects model 214
9.17 Plasma Nfl 215
9.17.1 Leave-one-out method 215
9.17.2 Comparing the results between the fixed-effect and random-effects model 216
9.18 Plasma p-tau181 217
9.18.1 Leave-one-out method 217
9.18.2 Comparing the results between the fixed-effect and random-effects model 218

### Appendix 1: Search Strategy

#### 1.1 Pubmed search strategy

#1. (((((((((((((((((((((((((((((((((((Alzheimer disease[Title/Abstract]) OR (Alzheimer Syndrome[Title/Abstract])) OR (Alzheimer-Type Dementia (ATD[Title/Abstract]))) OR (Alzheimer Type Dementia (ATD[Title/Abstract]))) OR (Dementia, Alzheimer-Type (ATD[Title/Abstract]))) OR (Primary Senile Degenerative Dementia[Title/Abstract])) OR (Dementia, Primary Senile Degenerative[Title/Abstract])) OR (Alzheimer Type Senile Dementia[Title/Abstract])) OR (Alzheimer Dementia[Title/Abstract])) OR (Alzheimer Dementias[Title/Abstract])) OR (Dementia, Alzheimer[Title/Abstract])) OR (Alzheimer's Disease[Title/Abstract])) OR (Dementia, Senile[Title/Abstract])) OR (Senile Dementia[Title/Abstract])) OR (Dementia, Alzheimer Type[Title/Abstract])) OR (Alzheimer Type Dementia[Title/Abstract])) OR (Senile Dementia, Alzheimer Type[Title/Abstract])) OR (Alzheimer Sclerosis[Title/Abstract])) OR (Sclerosis, Alzheimer[Title/Abstract])) OR (Alzheimer's Diseases[Title/Abstract])) OR (Alzheimer Diseases[Title/Abstract])) OR (Alzheimers Diseases[Title/Abstract])) OR (Acute Confusional Senile Dementia[Title/Abstract])) OR (Senile Dementia, Acute Confusional[Title/Abstract])) OR (Dementia, Presenile[Title/Abstract])) OR (Presenile Dementia[Title/Abstract])) OR (Alzheimer Disease, Late Onset[Title/Abstract])) OR (Late Onset Alzheimer Disease[Title/Abstract])) OR (Alzheimer's Disease, Focal Onset[Title/Abstract])) OR (Focal Onset Alzheimer's

Disease[Title/Abstract])) OR (Familial Alzheimer Disease (FAD[Title/Abstract]))) OR (Alzheimer Disease, Familial (FAD[Title/Abstract]))) OR (Familial Alzheimer Diseases (FAD[Title/Abstract]))) OR (Alzheimer Disease, Early Onset[Title/Abstract])) OR (Early Onset Alzheimer Disease[Title/Abstract])) OR (Presenile Alzheimer Dementia[Title/Abstract])

#2. (((Antibodies, Monoclonal [Title/Abstract]) OR (Monoclonal Antibodies [Title/Abstract])) OR (Monoclonal Antibody [Title/Abstract])) OR (Antibody, Monoclonal [Title/Abstract])

#3. "randomized controlled trial"[pt] OR "controlled clinical trial"[pt] OR randomized[tiab] OR placebo[tiab] OR "drug therapy"[sh] OR randomly[tiab] OR trial[tiab] OR groups[tiab]

#4. (((((((((((((((((((((((((((((((((((((((((((Alzheimer disease[Title/Abstract]) OR (Alzheimer Syndrome[Title/Abstract])) OR (Alzheimer-Type Dementia (ATD[Title/Abstract]))) OR (Alzheimer Type Dementia (ATD[Title/Abstract]))) OR (Dementia, Alzheimer-Type (ATD[Title/Abstract]))) OR (Primary Senile Degenerative Dementia[Title/Abstract])) OR (Dementia, Primary Senile Degenerative[Title/Abstract])) OR (Alzheimer Type Senile Dementia[Title/Abstract])) OR (Alzheimer Dementia[Title/Abstract])) OR (Alzheimer Dementias[Title/Abstract])) OR (Dementia, Alzheimer[Title/Abstract])) OR (Alzheimer's Disease[Title/Abstract])) OR (Dementia, Senile[Title/Abstract])) OR (Senile Dementia[Title/Abstract])) OR (Dementia, Alzheimer Type[Title/Abstract])) OR (Alzheimer Type Dementia[Title/Abstract])) OR (Senile Dementia, Alzheimer

Type[Title/Abstract])) OR (Alzheimer Sclerosis[Title/Abstract])) OR (Sclerosis, Alzheimer[Title/Abstract])) OR (Alzheimer's Diseases[Title/Abstract])) OR (Alzheimer Diseases[Title/Abstract])) OR (Alzheimers Diseases[Title/Abstract])) OR (Acute Confusional Senile Dementia[Title/Abstract])) OR (Senile Dementia, Acute Confusional[Title/Abstract])) OR (Dementia, Presenile[Title/Abstract])) OR (Presenile Dementia[Title/Abstract])) OR (Alzheimer Disease, Late Onset[Title/Abstract])) OR (Late Onset Alzheimer Disease[Title/Abstract])) OR (Alzheimer's Disease, Focal Onset[Title/Abstract])) OR (Focal Onset Alzheimer's Disease[Title/Abstract])) OR (Familial Alzheimer Disease (FAD[Title/Abstract]))) OR (Alzheimer Disease, Familial (FAD[Title/Abstract]))) OR (Familial Alzheimer Diseases (FAD[Title/Abstract]))) OR (Alzheimer Disease, Early Onset[Title/Abstract])) OR (Early Onset Alzheimer Disease[Title/Abstract])) OR (Presenile Alzheimer Dementia[Title/Abstract])) AND ((((Antibodies, Monoclonal[Title/Abstract]) OR (Monoclonal Antibodies[Title/Abstract])) OR (Monoclonal Antibody[Title/Abstract])) OR (Antibody, Monoclonal[Title/Abstract]))) AND ("randomized controlled trial"[pt] OR "controlled clinical trial"[pt] OR randomized[tiab] OR placebo[tiab] OR "drug therapy"[sh] OR randomly[tiab] OR trial[tiab] OR groups[tiab])

**1.2 Embase search strategy**

('alzheimer disease'/exp OR 'alzheimer-type dementia (atd)':ab,ti OR 'alzheimer type dementia (atd)':ab,ti OR 'dementia, alzheimer-type (atd)':ab,ti OR 'primary senile degenerative dementia':ab,ti OR 'dementia, primary senile degenerative':ab,ti OR 'alzheimer type senile dementia':ab,ti OR 'alzheimer dementia':ab,ti OR 'alzheimer dementias':ab,ti OR 'dementia, alzheimer':ab,ti OR 'alzheimers disease':ab,ti OR 'dementia, senile':ab,ti OR 'senile dementia':ab,ti OR 'dementia, alzheimer type':ab,ti OR 'alzheimer type dementia':ab,ti OR 'senile dementia, alzheimer type':ab,ti OR 'alzheimer sclerosis':ab,ti OR 'sclerosis, alzheimer':ab,ti OR 'alzheimer diseases':ab,ti OR 'alzheimers diseases':ab,ti OR 'acute confusional senile dementia':ab,ti OR 'senile dementia, acute confusional':ab,ti OR 'dementia, presenile':ab,ti OR 'presenile dementia':ab,ti OR 'alzheimer disease, late onset':ab,ti OR 'late onset alzheimer disease':ab,ti OR 'alzheimers disease, focal onset':ab,ti OR 'focal onset alzheimers disease':ab,ti OR 'familial alzheimer disease (fad)':ab,ti OR 'alzheimer disease, familial (fad)':ab,ti OR 'familial alzheimer diseases (fad)':ab,ti OR 'alzheimer disease, early onset':ab,ti OR 'early onset alzheimer disease':ab,ti OR 'presenile alzheimer dementia':ab,ti) AND ('antibodies, monoclonal'/exp OR 'monoclonal antibodies':ab,ti OR 'monoclonal antibody':ab,ti OR 'antibody, monoclonal':ab,ti) AND ('randomized controlled trial'/exp OR 'controlled clinical trial'/exp OR randomized:ti,ab OR placebo:ti,ab OR 'drug therapy':lnk OR randomly:ti,ab OR trial:ti,ab OR groups:ti,ab)

##### 1.3 Cochrane search strategy

#1. MeSH descriptor: [Alzheimer Disease] explode all trees

#2. MeSH descriptor: [Antibodies, Monoclonal] explode all trees

#3. (Alzheimer Syndrome OR Alzheimer-Type Dementia (ATD) OR Alzheimer Type Dementia (ATD) OR Dementia, Alzheimer-Type (ATD) OR Primary Senile Degenerative Dementia OR Dementia, Primary Senile Degenerative OR Alzheimer Type Senile Dementia OR Alzheimer Dementia OR Alzheimer Dementias OR Dementia, Alzheimer OR Alzheimer's Disease OR Dementia, Senile OR Senile Dementia OR Dementia, Alzheimer Type OR Alzheimer Type Dementia OR Senile Dementia, Alzheimer Type OR Alzheimer Sclerosis OR Sclerosis, Alzheimer OR Alzheimer's Diseases OR Alzheimer Diseases OR Alzheimers Diseases OR Acute Confusional Senile Dementia OR Senile Dementia, Acute Confusional OR Dementia, Presenile OR Presenile Dementia OR Alzheimer Disease, Late Onset OR Late Onset Alzheimer Disease OR Alzheimer's Disease, Focal Onset OR Focal Onset Alzheimer's Disease OR Familial Alzheimer Disease (FAD) OR Alzheimer Disease, Familial (FAD) OR Familial Alzheimer Diseases (FAD) OR Alzheimer Disease, Early Onset OR Early Onset Alzheimer Disease OR Presenile Alzheimer Dementia):ti,ab,kw

#4. (Monoclonal Antibodies Monoclonal Antibody Antibody, Monoclonal):ti,ab,kw

#5. #1 or #3

#6. #2 or #4

#7. #5 and #6

##### 1.4 Clinicaltrials.gov search strategy

Intervention/Treatment aducanumab /Condition or disease Alzheimer Disease / Other terms Random = 8

Intervention/Treatment donanemab /Condition or disease Alzheimer Disease / Other terms Random = 0

Intervention/Treatment lecanemab /Condition or disease Alzheimer Disease / Other terms Random = 4

Intervention/Treatment solanezumab /Condition or disease Alzheimer Disease / Other terms Random = 2

Intervention/Treatment crenezumab /Condition or disease Alzheimer Disease / Other terms Random = 5

Intervention/Treatment ponezumab /Condition or disease Alzheimer Disease / Other terms Random = 6

Intervention/Treatment gantenerumab /Condition or disease Alzheimer Disease / Other terms Random = 4

Intervention/Treatment bapineuzumab /Condition or disease Alzheimer Disease / Other terms Random = 10

Intervention/Treatment monoclonal antibodies /Condition or disease Alzheimer Disease / Other terms Random = 12

Total = 51

### Appendix 2: Table of study characteristics: substance use

| Soure | Trial phrase | Degree of AD (MMSE score) | Number of participants | | APOE4 Carrier, n (%) | | Mean age, years (SD) | | Women, n (%) | | AD medications used, n (%) | | CDR-SB score at Baseline, mean (SD) | | MMSE score at Baseline, mean (SD) | | Treatment duration | Intervention | | Outcomes |
|---|---|---|---|---|---|---|---|---|---|---|---|---|---|---|---|---|---|---|---|---|
| | | | experimental | control | experiment | control | experiment | control | experiment | control | experiment | control | experiment | control | experiment | control | | experiment | control | |
| Sperling at al. [1] (2023) Asymptomatic Alzheimer's Disease (A4) NCT02008357 | III | Preclinical (25 to 30) | 564 | 583 | 333 (59.0) | 342 (58.7) | 72.0 (4.7) | 71.9 (5.0) | 329 (58.3) | 352 (60.4) | Na | Na | 0.1 (0.2) | 0.0 (0.2) | 28.8 (1.3) | 28.8 (1.2) | 240 weeks | Solanezumab (Central-domain), 1600 mg, IV, Q4W | Placebo | 1. PACC, CFI, ADL, CDR-SB; 2. Amyloid PET; 3. vMRI. |
| Sims at al. [2] (2023) TRAILBLAZER-ALZ 2 NCT04437511 | III | mci-mild (20 to 28) | 860 | 876 | 598 (69.8) | 621 (71.2) | 73.0 (6.2) | 73.0 (6.2) | 493 (57.3) | 503 (57.4) | 521 (60.6) | 538 (61.4) | 4.0 (2.1) | 3.9 (2.1) | 22.4 (3.8) | 22.2 (3.9) | 76 weeks | Donanemab (N-terminal), 700 mg for the first 3 doses and 1400 mg thereafter, IV, Q4W | Placebo | 1. iADRS, ADAS-cog13, ADCS-iADL, MWPC, CDR-SB, MMSE, CDR-G; 2. Amyloid PET; 3. vMRI; 4. Plasma biomarkers (p-tau217). |
| Dyck at al. [3] (2022) Clarity AD | III | mci-mild (Na) | 859 | 875 | 468 (53.5) | 600 (68.6) | 71.(7.9) | 71.0 (7.8) | 443 (51.6) | 464 (53.0) | 447 (52.0) | 468 (53.5) | 3.17 (1.3) | 3.22 (1.3) | 25.5 (2.2) | 25.6 (2.2) | 72 weeks | Lecanemab (N-terminal), 10 mg/kg IV, Q2W | Placebo | 1.CDR-SB, ADAS-cog14, ADCOMS, |

| | | | | | | | | | | | | | | | | | | | | |
|---|---|---|---|---|---|---|---|---|---|---|---|---|---|---|---|---|---|---|---|---|
| NCT03887455 | | | | | | | | | | | | | | | | | | | | ADCS-MCI-ADL, CDR-G;<br>2. Amyloid PET and tau PET;<br>3.vMRI;<br>4.CSF biomarkers (Aβ1-40, Aβ1-42, total tau, p-tau181, neurogranin, and NfL) and plasma biomarkers (Aβ42/40ratio, p-tau181, GFAP, and NfL). |
| Doody at al. [4] (2014) EXPEDITION NCT00905372 | III | mild-moderate (16 to 26) | 506 | 506 | 266 (57.3) | 288 (61.3) | 75.0 (7.9) | 74.4 (8.0) | 299 (59.1) | 287 (56.7) | 196 (38.7) | 196 (38.7) | Na | Na | 21 (4.0) | 21 (3.0) | 76 weeks | Solanezumab (Central-domain), 400 mg, IV, Q4W | Placebo | 1. ADAS-cog14, ADAS-cog11, ADCS-ADL, CDR-SB, MMSE, NPI, RUD-Lite, EQ-5D, QOL-AD;<br>2. Amyloid PET;<br>3. vMRI;<br>4. CSF biomarkers (free Aβ40, free Aβ42, total Aβ40, total Aβ42) and |
| Doody at al. [4] (2014) EXPEDITION2 NCT00904683 | III | | 521 | 519 | 263 (56.8) | 281 (59.5) | 72.5 (8.0) | 72.4 (7.8) | 283 (54.3) | 286 (55.1) | 90 (17.3) | 91 (17.5) | Na | Na | 21 (3.0) | 21 (3.0) | 76 weeks | Solanezumab (Central-domain), 400 mg, IV, Q4W | Placebo | |

| | | | | | | | | | | | | | | | | | | | | |
|---|---|---|---|---|---|---|---|---|---|---|---|---|---|---|---|---|---|---|---|---|
| | | | | | | | | | | | | | | | | | | | | plasma biomarkers (Aβ40, Aβ42). |
| Honig at al. [5] (2018) EXPEDITION3 NCT01900665 | III | mild (20 to 26) | 1057 | 1072 | 712 (69.3) | 685 (66.3) | 72.7 (7.8) | 73.3 (8.0) | 631 (58.9) | 631 (58.9) | 856 (79.9) | 856 (79.9) | 3.9 (1.9) | 3.9 (2.0) | 22.8 (2.8) | 22.6 (2.9) | 76 weeks | Solanezumab (Central-domain), 400 mg, IV, Q4W | Placebo | 1. ADAS-cog14, MMSE, ADCS, ADCS-ADL, ADCS-iADL, CDR-SB, FAQ, EQ-5D, QoL-AD, iADRS, RUD-Lite, NPI; 2 Amyloid PET; 3. vMRI; 4.CSF biomarkers (Aβ40, Aβ42, total tau). |
| Haeberlein at al. [6] (2022) EMERGE NCT02484547 | III | mci-mild (24 to 30) | 543 | 548 | 362 (66.7) | 368 (67.1) | 70.6 (7.4) | 70.8 (7.4) | 269 (49.5) | 290 (52.9) | 282 (51.9) | 285 (52.0) | 2.47 (1.0) | 2.51 (1.1) | 26.4 (1.8) | 26.3 (1.7) | 76 weeks | Aducanumab (N-terminal), 3 mg/kg or 6 mg/kg, IV, Q4W | Placebo | 1. CDR-SB, MMSE, ADAS-cog13, ADCS-ADL-MCI, NPI-10; 2. Amyloid PET, and tau PET; 3. CSF biomarkers (Aβ42, p-tau, total tau) and plasma (p-tau181). |
| | | | 547 | | 365 (66.7) | | 70.6 (7.5) | | 284 (51.9) | | 281 (51.4) | | 2.46 (1.0) | | 26.3 (1.7) | | | Aducanumab (N-terminal), 6 mg/kg or 10 mg/kg, IV, Q4W | | |
| Haeberlein at al. [6] (2022) ENGAGE NCT02477800 | III | mci-mild (24 to 30) | 547 | 545 | 391 (71.5) | 365 (67.0) | 70.4 (7.0) | 69.8 (7.7) | 284 (51.9) | 287 (52.7) | 299 (54.7) | 313 (57.4) | 2.40 (1.0) | 2.40 (1.0) | 26.4 (1.7) | 26.4 (1.8) | | Aducanumab (N-terminal), 3 mg/kg or 6 mg/kg, IV, Q4W | | |
| | | | 555 | | 378 (68.1) | | 70.0 (7.7) | | 292 (52.6) | | 317 (57.1) | | 2.43 (1.0) | | 26.4 (1.8) | | | Aducanumab (N-terminal), 6 mg/kg or | | |

| | | | | | | | | | | | | | | | | | | | | |
|---|---|---|---|---|---|---|---|---|---|---|---|---|---|---|---|---|---|---|---|---|
| | | | | | | | | | | | | | | | | | | 10 mg/kg, IV, Q4W | | |
| Ostrowitzki at al. [7] (2022) CREAD NCT02670083 | III | prodromal or mild (22 or higher) | 409 | 404 | 293 (72.7) | 292(71. 7) | 71.0 (7.9) | 70.3 (8.4) | 236 (58.4) | 247 (60.4) | 291 (71.1) | 284 (70.3) | 3.88 (1.7) | 3.79 (1.6) | 23.7 (3.0) | 23.4 (2.9) | 100 weeks | Crenezumab (Central-domainl), 60 mg/kg, IV, Q4W | Placebo | 1. CDR-SB, MMSE, ADAS-cog13, ADAS-cog11, CDR-GS, ADCS-ADL, ADCS-iADL, NPI-Q, QOL-AD, ZCI-AD, EQ-5D; 2. Amyloid PET, and tau PET; 3. vMRI; 4. CSF biomarkers (Aβ40, Aβ42, total tau, p-tau181 and Aβ oligomers) and plasma biomarkers (Aβ42 and Aβ40). |
| Ostrowitzki at al. [7] (2022) CREAD2 NCT03114657 | III | | 399 | 407 | 271 (66.9) | 263 (65.9) | 71.1 (7.5) | 70.7 (7.9) | 231 (56.8) | 225 (56.4) | 289 (72.4) | 279 (68.6) | 3.68 (1.6) | 3.76 (1.6) | 23.6 (2.8) | 23.5 (2.9) | | | | |
| Swanson at al. [8] (2021) NCT01767311 | IIb | mci-mild (na) | 52 | 238 | 38 (73.1) | 169 (71.0) | 70.49 (8.0) | 71.85 (7.0) | 26 (50.0) | 137 (57.6) | 28 (53.8) | 128 (53.8) | 3.0 (1.6) | 2.9 (1.5) | 25.7 (2.5) | 26.0 (2.3) | 18 months | Lecanemab (N-terminal), 2.5mg/kg, IV, Q2W | Placebo | 1. ADCOMS, CDR-SB, ADAS-Cog14; 2. Amyloid PET; 3. vMRI; 4. CSF biomarkers (Aβ42, p-tau, total |
| | | | 48 | | 37 (77.1) | | 70.73 (6.5) | | 24 (50.0) | | 25 (52.1) | | 2.9 (1.4) | | 25.3 (2.6) | | | Lecanemab (N-terminal), 5mg/kg, IV, Q4W | | |
| | | | 89 | | 81 (91.0) | | 71.70 | | 48 (53.9) | | 56 (62.9) | | 3.0 (1.3) | | 25.6 (2.3) | | | Lecanemab | | |

| | | | | | | | | | | | | | | | | | | | | |
|---|---|---|---|---|---|---|---|---|---|---|---|---|---|---|---|---|---|---|---|---|
| | | | | | | | (7.1) | | | | | | | | | | | (N-terminal), 5mg/kg, IV, Q2W | | tau, NfL). |
| | | | 246 | | 218 (88.6) | | 71.03 (6.6) | | 110 (44.7) | | 131 (53.2) | | 2.9 (1.3) | | 25.7 (2.4) | | | Lecanemab (N-terminal), 10mg/kg, IV, Q4W | | |
| | | | 152 | | 46 (30.3) | | 72.704 (7.0) | | 64 (42.1) | | 79 (52.0) | | 3.0 (1.4) | | 25.6 (2.4) | | | Lecanemab (N-terminal), 10mg/kg, IV, Q2W | | |
| Mintun at al. [9] (2021) NCT03367403 | II | mci-mild (20 to 28) | 131 | 126 | 95 (72.5) | 92 (74.2) | 75.0 (5.6) | 75.4 (5.4) | 68 (51.9) | 65 (51.6) | 78 (59.5) | 74 (58.7) | 3.6 (2.1) | 3. 4(1.7) | 23.6 (3.1) | 23.7 (2.9) | 72 weeks | Donanemab (N-terminal)，700 mg for the first three doses and 1400 mg thereafter，Q4W | Placebo | 1. iADRS, CDR-SB, ADAS-cog13, ADCS-ADL, ADCS-iADL, MMSE; 2. Amyloid PET, and tau PET; 3. vMRI. |
| Lowe at al. [10](2021) | Ib | mci-mild (16 to 30) | 7 | 15 | 5 (71.4) | 10 (66.7) | 78.3 (8.4) | 74.5 (10.1) | 4 (57.1) | 11 (73.3) | Na | Na | Na | Na | 21.71 (4.2) | 21.67 (4.8) | single-dose | Donanemab (N-terminal), 10mg/kg, IV, SD | Placebo | 1. ADAS-Cog14, CDR-SB, MMSE, FCSRT- IR, ADCS- MCI- ADL - 24, NTB; 2. Amyloid PET; 3. vMRI; |
| | | | 7 | | 5 (71.4) | | 75.6 (6.4) | | 3 (42.9) | | Na | | Na | | 23.57 (4.5) | | single-dose | Donanemab (N-terminal), 20mg/kg, IV, SD | | |
| | | | 4 | | 4 (100.0) | | 75.3 (6.7) | | 2 (50.0) | | Na | | Na | | 23.25 (3.9) | | single-dose | Donanemab (N-terminal), 40mg/kg, IV, SD | | |

| | | | | | | | | | | | | | | | | | | | | |
|---|---|---|---|---|---|---|---|---|---|---|---|---|---|---|---|---|---|---|---|---|
| | | | 10 | | 8 (80.0) | | 66.8 (8.6) | | 4 (40.0) | | Na | | Na | | 20.00<br>(3.4) | | 24 weeks | Donanemab<br>(N-terminal), 10mg/kg,<br>IV, Q2W | | |
| | | | 8 | | 6 (75.0) | | 73.0 (7.8) | | 4 (50.0) | | Na | | Na | | 18.86<br>(2.4) | | 72 weeks | Donanemab<br>(N-terminal), 10mg/kg,<br>IV, Q4W | | |
| | | | 10 | | 9 (90.0) | | 71.7 (9.2) | | 6 (60.0) | | Na | | Na | | 19.50<br>(2.9) | | 72 weeks | Donanemab<br>(N-terminal), 20mg/kg,<br>IV, Q4W | | |
| Cummings at al. [11]<br>(2018)<br>ABBY<br>NCT01343966 | II | mild-moderate<br>(18 to 26) | 122 | 62 | 78 (63.9) | 40<br>(64.5) | 71.2 (6.3) | 70.3<br>(7.2) | 66 (54.1) | 34<br>(48.4) | 103<br>(84.4) | 52<br>(87.1) | 4.7 (1.9) | 4.6<br>(2.2) | 21.7 (2.8) | 21.5<br>(2.6) | 68 weeks | Crenezumab<br>(Central-domainl), 300<br>mg, SC, Q2W | Placeb<br>o | 1. ADAS-Cog12,<br>CDR-SB,<br>ADCS-ADL;<br>2.vMRI;<br>3. CSF biomarkers<br>(Aβ42, CSF total<br>tau and p-tau181). |
| | | | 165 | 84 | 117<br>(70.9) | 60<br>(71.4) | 70.9 (6.9) | 69.9<br>(7.1) | 89 (50.9) | 45<br>(57.1) | 139<br>(88.5) | 71<br>(86.9) | 4.5 (2.2) | 4.5<br>(2.1) | 21.9 (2.7) | 21.6<br>(2.5) | | Crenezumab<br>(Central-domainl), 15<br>mg/kg, IV, Q4W | Placeb<br>o | |
| Salloway at al. [12]<br>(2018)<br>BLAZE<br>NCT01397578 | II | mild-moderate<br>(18 to 26) | 26 | 13 | 22 (84.6) | 12<br>(92.3) | 66.7 (9.5) | 68.9<br>(8.3) | 14 (53.8) | 7<br>(61.5) | 22 (84.6) | 11<br>(92.3) | 4.2 (2.1) | 4.3<br>(1.5) | 21.5 (2.4) | 22.3<br>(2.4) | 68 weeks | Crenezumab<br>(Central-domainl), 300<br>mg, SC, Q2W | Placeb<br>o | 1. ADAS-cog12、<br>CDR-SB;<br>2. Amyloid PET;<br>3.CSF biomarkers<br>(Aβ42, p-tau and<br>total tau) and<br>plasma biomarkers<br>(Aβ42 and Aβ40);<br>4.<br>Fluorodeoxyglucos |
| | | | 35 | 17 | 24 (68.6) | 12<br>(70.6) | 71.4 (7.1) | 69.8<br>(7.7) | 19 (68.6) | 9<br>(35.3) | 30 (91.4) | 14(82.<br>4) | 4.9 (2.0) | 5.9<br>(1.9) | 20.8(2.3) | 20.5(2.<br>2) | | Crenezumab<br>(Central-domainl), 15<br>mg/kg,IV ,Q4W | Placeb<br>o | |

| | | | | | | | | | | | | | | | | | | | | |
|---|---|---|---|---|---|---|---|---|---|---|---|---|---|---|---|---|---|---|---|---|
| | | | | | | | | | | | | | | | | | | | | e PET. |
| Guthrie at al. [13] (2020) GN29632 NCT02353598 | Ib | mild-moderate (18 to 28) | 10 | 14 | 6 (60.0) | 10 (71.4) | 71.16 (9.05) | 71.08 (7.9) | 28 (45.9) | 10 (71.4) | Na | Na | Na | Na | 23.3 (3.8) | 20.9 (2.8) | 13 weeks | Crenezumab (Central-domainl), 30mg/kg, IV, Q4W | Placebo | 1. MMSE;<br>2. Amyloid PET;<br>3. vMRI;<br>4. Plasma biomarkers (Aβ40, Aβ1-42). |
| | | | 11 | | 10 (90.9) | | 71.72 (8.1) | | | | Na | | Na | | 22.2 (2.7) | | | Crenezumab (Central-domainl), 45mg/kg, IV, Q4W | Placebo | |
| | | | 21 | | 17 (80.9) | | 71.77 (9.5) | | | | Na | | Na | | 23.2 (3.3) | | | Crenezumab (Central-domainl), 60mg/kg, IV, Q4W | Placebo | |
| | | | 19 | | 13 (68.4) | | 69.89 (9.2) | | | | Na | | Na | | 22.9 (3.1) | | | Crenezumab (Central-domainl), 120mg/kg, IV, Q4W | Placebo | |
| Lu at al. [14] (2018) | I | mild-moderate (14 to 26) | 6 | 10 | Na | Na | 65.8 (11.6) | 70.3 (12.2) | 3 (50.0) | 4 (40.0) | Na | Na | Na | Na | 19.9 (5.2) | 19.9 (5.2) | single-dose | Bapineuzumab (N-terminal), 5mg, SC, SD | Placebo | 1. MMSE;<br>2. PK/PD assessments. |
| | | | 6 | | Na | | 74.2 (10.7) | | 3 (50.0) | | Na | | Na | | 19.7 (4.32) | | single-dose | Bapineuzumab (N-terminal), 10mg, SC, SD | | |
| | | | 6 | | Na | | 69.0 (9.5) | | 3 (50.0) | | Na | | Na | | 20.3 (4.3) | | single-dose | Bapineuzumab (N-terminal), 20mg, SC, SD | | |
| | | | 6 | | Na | | 79.3 (4.0) | | 3 (50.0) | | Na | | Na | | 20.3 (4.3) | | single-dose | Bapineuzumab (N-terminal), 40mg, SC, SD | | |
| | | | 6 | | Na | | 66.8 (9.5) | | 3 (50.0) | | Na | | Na | | 19.5 | | single-do | Bapineuzumab | | |

| | | | | | | | | | | | | | | | | | | | | |
|---|---|---|---|---|---|---|---|---|---|---|---|---|---|---|---|---|---|---|---|---|
| | | | | | | | | | | | | | | | (2.6) | | se | (N-terminal), 80mg, SC, SD | | |
| Ostrowitzki at al. [15] (2017) SCarlet RoAD NCT01224106 | III | prodromal-moderate (24 or higher) | 271 | 266 | 214 (79.0) | 187 (70.3) | 70.3 (7.0) | 69.5 (7.5) | Na | Na | Na | Na | 2.2 (1.0) | 2.1 (1.0) | 25.7 (2.3) | 25.7 (2.1) | 96weeks | Gantenerumab (N-terminal+central-domain), 105 mg, SC, Q4W | Placebo | 1. CDR-SB, ADAS-Cog 13, MMSE, CANTAB, FCSRT, NPI-Q, FAQ; 2. Amyloid PET; 3. vMRI; 4. CSF biomarkers (Aβ42, p-tau181, total tau, and neurogranin). |
| | | | 260 | | 160 (61.5) | | 71.3 (7.1) | | Na | | Na | | 2.0 (0.9) | | 25.7 (2.2) | | | Gantenerumab (N-terminal+central-domain), 225 mg, SC, Q4W | | |
| Salloway at al. [16] (2014) 301: ApoE ε4 noncarrier study NCT00574132 | III | mild-moderate (Na) | 314 | 493 | 0 (0.0) | 0 (0.0) | 73.1 (9.3) | 71.9 (10.1) | 165 (52.5) | 248 (50.3) | 281 (89.5) | 442 (89.7) | Na | Na | 21.2 (3.4) | 21.2 (3.2) | 78 weeks | Bapineuzumab (N-terminal), 0.5 mg/kg，IV, Q13W | Placebo | 1. ADAS-cog11, DAD, NTB, CDR-SB, MMSE; 2. Amyloid PET; 3. vMRI; 4. CSF biomarkers (p-tau). |
| | | | 307 | | 0 (0.0) | | 73.5 (9.1) | | 175 (57.0) | | 278 (90.6) | | Na | | 21.2 (3.4) | | | Bapineuzumab (N-terminal), 1.0mg/kg，IV, Q13W | Placebo | |
| Salloway at al. [16] (2014) 302: ApoE ε4 carrier study NCT00575055 | III | mild-moderate(Na) | 658 | 432 | 658 (100.0) | 432 (100.0) | 72.0 (8.0) | 72.3 (8.4) | 358 (54.4) | 242 (56.0) | 606 (92.1) | 400 (92.6) | Na | Na | 20.7 (3.2) | 20.7 (3.2) | | Bapineuzumab (N-terminal), 0.5 mg/kg，IV, Q13W | Placebo | |
| Landen at al. [17](2017) | II | mild-moderate (16 to 26) | 25 | 24 | Na | Na | 70.8 (8.2) | 70.0 (7.8) | 12 (48.0) | 13 (54.2) | Na | Na | Na | Na | 21.5 (2.9) | 21.0 (3.4) | 72 weeks | Ponezumab(C-terminal)，0.1mg/kg，IV, Q60D | Placebo | 1. ADAS-Cog, DAD, MMSE; |

| | | | | | | | | | | | | | | | | | | | | |
|---|---|---|---|---|---|---|---|---|---|---|---|---|---|---|---|---|---|---|---|---|
| NCT00722046 | | | 25 | | Na | Na | 71.9 (9.4) | | 15 (60.0) | | Na | | Na | | 21.4 (3.6) | | | Ponezumab(C-terminal), 0.5mg/kg, IV, Q60D | | 2. Amyloid PET;<br>3. vMRI;<br>4. PK/PD assessments. |
| | | | 25 | | Na | Na | 72.2 (8.4) | | 11 (44.0) | | Na | | Na | | 20.8 (3.0) | | | Ponezumab(C-terminal), 1.0 mg/kg，IV, Q60D | | |
| | | | 32 | 32 | Na | Na | 70.5 (8.9) | 70.4 (10.3) | 20 (62.5) | 17(53.1) | Na | Na | Na | Na | 22.5 (2.5) | 21.9 (3.4) | | Ponezumab(C-terminal)，3.0 mg/kg，IV, Q60D | Placebo | |
| | | | 31 | | Na | Na | 71.8 (7.3) | | 17 (54.8) | | Na | | Na | | 20.9 (3.1) | | | Ponezumab (C-terminal), 8.5 mg/kg, IV, Q60D | | |
| Landen at al [18] (2017) NCT00945672 | II | mild-moderate (16 to 26) | 12 | 6 | Na | Na | 65.1 (7.4) | 71.3 (8.5) | 4 (33.3) | 3 (50.0) | Na | Na | Na | Na | 22.5 (2.8) | 20.8 (3.0) | 52weeks | Ponezumab(C-terminal), 10 mg/kg, IV, quarterly | Placebo | 1. ADAS-Cog, DAD, CogState, NTB, NPI-12, CDR-SOB, EQ-5D;<br>2. vMRI;<br>3. PK/PD assessments. |
| | | | 12 | 6 | Na | Na | 69.8 (7.5) | 65.8 (8.3) | 3 (25.0) | 5 (83.3) | Na | Na | Na | Na | 21.2 (3.0) | 22.5 (4.0) | | single loading dose of ponezumab(C-terminal) 10 mg/kg, quarterly, followed by monthly ponezumab (C-terminal) 7.5 mg/kg，IV | Placebo | |
| Vandenberghe at al. [19] (2016) 3001: ApoE ε4 carrier study NCT00676143 | III | mild-moderate (16 to 26) | 650 | 431 | 650 (100.0) | 431 (100.0) | 70.9 (Na) | 70.2 (Na) | 419 (64.5) | 278 (60.1) | 578 (88.9) | 386 (89.6) | Na | Na | 20.9 (3.1) | 21.0 (3.0) | 72 weeks | Bapineuzumab (N-terminal), 0.5 mg/kg, IV，Q13W | Placebo | 1. ADAS-cog11, DAD, CDR-SB, NTB;<br>2. Amyloid PET;<br>3. vMRI;<br>4. CSF biomarkers (p-tau) and plasma (Aβx–40). |
| Vandenberghe at al. [19] (2016) 3000: ApoE ε4 noncarrier study | III | mild-moderate (16 to 26) | 255 | 328 | 0 (0.0) | 0 (0.0) | 71.1 (Na) | 69.7 (Na) | 164 (55.7) | 212 (57.9) | 204 (80.0) | 274 (83.5) | Na | Na | 20.8 (3.2) | 20.8 (3.1) | | Bapineuzumab (N-terminal), 0.5 mg/kg, IV, Q13W | Placebo | |
| | | | 253 | | 0 (0.0) | 0 (0.0) | 70.7 (Na) | | 163 | | 209 | | Na | | 20.8 (3.1) | | | Bapineuzumab | Placeb | |

| | | | | | | | | | | | | | | | | | | | | |
|---|---|---|---|---|---|---|---|---|---|---|---|---|---|---|---|---|---|---|---|---|
| NCT00667810 | | | | | | | | | (57.3) | | (82.6) | | | | | | | (N-terminal)，1.0mg/kg，IV，Q13W | o | |
| Sevigny at al. [20] (2016) PRIME NCT01677572 | Ib | prodromal-mild (NA) | 31 | 40 | 19 (61.3) | 26 (65.0) | 72.6 (7.8) | 72.8 (7.2) | 13 (41.9) | 23 (57.5) | 19 (61.3) | 24 (60.0) | 3.40 (1.8) | 2.66 (1.5) | 23.6 (3.3) | 24.7 (3.6) | 52weeks | Aducanumab (N-terminal)，1 mg/kg，IV，Q4W | Placebo | 1. CDR-SB，MMSE, NTB, FCSRT, ADAS-cog11, DAD, NTB; 2. Amyloid PET. |
| | | | 32 | | 19 (59.3) | | 70.5 (8.2) | | 17 (53.1) | | 28 (87.5) | | 3.50 (2.1) | | 23.2 (4.2) | | | Aducanumab (N-terminal)，3 mg/kg，IV，Q4W | | |
| | | | 30 | | 21 (70.0) | | 73.3 (9.3) | | 15 (50.0) | | 20 (66.7) | | 3.32 (1.5) | | 24.4 (2.9) | | | Aducanumab (N-terminal)，6 mg/kg，IV，Q4W | | |
| | | | 32 | | 20 (62.5) | | 73.7 (8.3) | | 15 (46.9) | | 17 (53.1) | | 3.14 (1.7) | | 24.8 (3.1) | | | Aducanumab (N-terminal)，10 mg/kg，IV，Q4W | | |
| Logovinsky at al. [21] (2016) NCT01230853 | Na | mild-moderate (16 to 28) | 6 | 12 | Na | Na | 70.0 (12.0) | 72.1 (9.2) | 5 (41.7) | 5 (41.7) | Na | Na | Na | Na | 25.2 (Na) | 23.5 (Na) | single-dose | Lecanemab (N-terminal)，0.1 mg/kg，IV, SD | Placebo | 1. CSF biomarkers (Aβ42, total tau, p-tau), and plasma (Aβ40); 2. PK/PD assessments. |
| | | | 6 | | Na | | 72.7 (6.5) | | 2 (33.3) | | Na | | Na | | 25. 2 (Na) | | | Lecanemab (N-terminal)，0.3 mg/kg，IV, SD | | |
| | | | 6 | | Na | | 75.0 (14.0) | | 2 (33.3) | | Na | | Na | | 23.5 (Na) | | | Lecanemab (N-terminal)，1 mg/kg，IV, SD | | |
| | | | 6 | | Na | | 68.2 (8.4) | | 3 (50.0) | | Na | | Na | | 24 (Na) | | | Lecanemab，3mg/kg，IV，SD | | |
| | | | 6 | | Na | | 68.7 (9.1) | | 2 (33.3) | | Na | | Na | | 24.2 (Na) | | | Lecanemab | | |

| | | | | | | | | | | | | | | | | | | | | |
|---|---|---|---|---|---|---|---|---|---|---|---|---|---|---|---|---|---|---|---|---|
| | | | | | | | | | | | | | | | | | | (N-terminal)，10mg/kg，IV, SD | | |
| | | | 6 | | Na | | 70.8 (11.5) | | 5 (83.3) | | Na | | Na | | 21.5 (Na) | | | Lecanemab (N-terminal)，15mg/kg，IV, SD | | |
| | | | 6 | 8 | Na | Na | 69.0 (13.1) | 70.0 (11.7) | 2 (33.3) | 2 (25.0) | Na | Na | Na | Na | 24 (Na) | 24.1 (Na) | 16 weeks | Lecanemab (N-terminal)，0.3mg/kg，IV，Q2W | Placebo | |
| | | | 6 | | Na | Na | 69.0 (7.0) | | 5 (83.3) | | Na | | Na | | 21.7 (Na) | | | Lecanemab (N-terminal)，1mg/kg，IV，Q2W | | |
| | | | 6 | | Na | Na | 71.8 (11.5) | | 4 (66.7) | | Na | | Na | | 23.7 (Na) | | | Lecanemab (N-terminal)，3mg/kg，IV，Q2W | | |
| | | | 6 | | Na | Na | 70.3 (9.8) | | 2 (33.3) | | Na | | Na | | 23.8 (Na) | | | Lecanemab (N-terminal)，10mg/kg，IV，Q2W | | |
| Ferrero at al. [22] (2016) NCT01397539 | I | mild-moderate (14 to 26) | 6 | 14 | 2 (33.3) | 4 (28.6) | 72.0 (8.4) | 66.9 (8.7) | 2 (33.3) | 9(64.3) | Na | Na | Na | Na | 23.0 (1.9) | 22.1 (2.4) | single-dose | Aducanumab (N-terminal)，0.3mg/kg，IV， SD | Placebo | 1. ADAS-Co13g; 2. CSF biomarkers (Aβ40, Aβ42); 3. PK/PD assessments. |
| | | | 6 | | 2 (33.3) | | 67.0 (8.8) | | 4 (66.7) | | Na | | Na | | 22.0 (3.4) | | | Aducanumab (N-terminal)，1mg/kg，IV， SD | Placebo | |
| | | | 6 | | 1 (16.7) | | 63.0 (5.0) | | 3 (50.0) | | Na | | Na | | 18.3 (2.7) | | | Aducanumab (N-terminal)，10mg/kg，IV, SD | Placebo | |

| | | | | | | | | | | | | | | | | | | | | |
|---|---|---|---|---|---|---|---|---|---|---|---|---|---|---|---|---|---|---|---|---|
| | | | 6 | | 4 (66.7) | | 72.7 (4.5) | | 5 (83.3) | | Na | | Na | | 18.3 (4.9) | | | Aducanumab (N-terminal)，20mg/kg，IV， SD | Placebo | |
| | | | 6 | | 2 (33.3) | | 66.8 (8.9) | | 5 (83.3) | | Na | | Na | | 23.0 (3.1) | | | Aducanumab (N-terminal)，20mg/kg，IV， SD | Placebo | |
| | | | 6 | | 2 (33.3) | | 63.3 (9.0) | | 5(83.3) | | Na | | Na | | 19.8 (4.6) | | | Aducanumab (N-terminal)，30mg/kg，IV, SD | Placebo | |
| | | | 3 | | 1 (33.3) | | 73.7 (9.5) | | 3 (100) | | Na | | Na | | 24.7 (1.5) | | | Aducanumab (N-terminal)，60 mg/kg，IV, SD | Placebo | |
| Delnomdedieu at al.[23] (2016) NCT01193608 | Na | mild-moderate (16 to 26) | 6 | 19 | Na | Na | 64.5 (6.4) | 71.1 (7.6) | 4 (66.7) | 7 (36.8) | Na | Na | Na | Na | 22.7 (1.4) | 21.2 (3.8) | 26 weeks | Bapineuzumab (AAB-003) (N-terminal), 0.5 mg/kg, IV, Q13W | Placebo | 1. ADAS-Cog, DAD, CDR-SB, MMSE; 2. CSF biomarkers (AβX-40, AβX-42, total tau, p-tau); 3. PK/PD assessments. |
| | | | 6 | | Na | Na | 67.2 (7.4) | | 3 (50.0) | | Na | | Na | | 22.5 (2.4) | | | Bapineuzumab (AAB-003) (N-terminal), 1 mg/kg, IV, Q13W | Placebo | |
| | | | 16 | | Na | Na | 70.7 (10.8) | | 14 (87.5) | | Na | | Na | | 20.8 (3.7) | | | Bapineuzumab (AAB-003) (N-terminal), 2 mg/kg, IV, Q13W | Placebo | |
| | | | 17 | | Na | Na | 71.5 (8.8) | | 9 (52.9) | | Na | | Na | | 20.1 (3.0) | | | Bapineuzumab (AAB-003) | Placebo | |

| | | | | | | | | | | | | | | | | | | | | |
|---|---|---|---|---|---|---|---|---|---|---|---|---|---|---|---|---|---|---|---|---|
| | | | | | | | | | | | | | | | | | | (N-terminal), 4 mg/kg, IV, Q13W | | |
| | | | 24 | | Na | Na | 64.5 (7.6) | | 14 (58.3) | | Na | | Na | | 21.2 (2.8) | | | Bapineuzumab (AAB-003) (N-terminal), 8 mg/kg, IV, Q13W | Placeb o | |
| Brody at al. [24] (2016) NCT01254773 | II | mild-moderate (16 to 26) | 37 | 36 | 22 (59.5) | 22 (61.1) | 73.5 (8.3) | 73.3 (8.8) | 19 (51.4) | 23 (63.9) | 30 (81.1) | 29 (80.6) | 4.5 (2.5) | 4.5 (2.4) | 22.3 (2.8) | 22.3 (2.8) | 12 months | Bapineuzumab (N-terminal), 2 mg, SC, Q4W | Placeb o | 1. ADAS-Cog, DAD, CDR-SB; 2. Amyloid PET; 3. vMRI; 4. PK/PD assessments. |
| | | | 36 | | 22 (59.5) | | 74.1 (9.3) | | 17 (47.2) | | 34 (94.4) | | 4.6 (2.1) | | 21.5 (2.5) | | | Bapineuzumab (N-terminal), 7 mg, SC, Q4W | | |
| | | | 37 | | 22 (59.5) | | 70.5 (8.7) | | 25 (67.6) | | 30 (81.1) | | 4.2 (2.2) | | 21.9 (3.00) | | | Bapineuzumab (N-terminal), 20 mg, SC, Q4W | | |
| Salloway at al. [25] (2009) Study 201 NCT00112073 | II | mild-moderate (16 to 26) | 122 | 107 | 72 (60.5) | 74 (69.8) | 70.1 (9.1) | 67.9 (8.8) | 61 (50.0) | 72 (60.5) | 116 (95.1) | 103 (96.3) | Na | Na | 20.9 (3.2) | 20.7 (3.1) | 78 weeks | Bapineuzumab (N-terminal), 0.15mg/kg, IV, Q13W | Placeb o | 1. ADAS-cog 12, DAD, MMSE, NTB, CDR-SB; 2. vMRI; 3. CSF; biomarkers (Aβ42, AβX-42, total tau, p-tau, p-tau181). |
| | | | | | | | | | | | | | Na | | | | | Bapineuzumab (N-terminal), 0.5mg/kg, IV, Q13W | | |
| | | | | | | | | | | | | | Na | | | | | Bapineuzumab (N-terminal), 1.0mg/kg, IV, Q13W | | |
| | | | | | | | | | | | | | Na | | | | | Bapineuzumab (N-terminal), 2.0mg/kg, | | |

| | | | | | | | | | | | | | | | | | | | | |
|---|---|---|---|---|---|---|---|---|---|---|---|---|---|---|---|---|---|---|---|---|
| | | | | | | | | | | | | | | | | | | IV, Q13W | | |
| Rinne at al.[26] (2010) Study 202 EudraCT number 2004-004120-12 ISRCTN17517446 | II | mild-moderate (16 to 26) | 19 | 7 | 12 (63.2) | 5 (71.4) | 67.26 (8.6) | 70.00 (8.8) | 8 (42.1) | 4 (57.1) | 19 (100.0) | 7 (100.0 ) | 5.61 (1.64) | 3.50 (1.50) | 21.00 (2.3) | 22.29 (2.7) | 78 weeks | Bapineuzumab (N-terminal),0.5,1.0, or2.0 mg/kg, IV, Q13W | Placebo | 1. ADAS-cog 11, ADAS-cog 12, DAD, CDR-SB, NTB、MMSE; 2. Amyloid PET; 3. vMRI; 4. CSF biomarkers (AβX-42, total tau, p-tau); 5. Fluorodeoxyglucose PET. |
| Arai at al.[27] (2016) NCT00397891 | I | mild-moderate (14 to 26) | 6 | 8 | Na | Na | 60.7 (5.2) | 68.8 (8.9) | 3 (50.0) | 4 (50.0) | Na | Na | Na | Na | 16.8 (2.9) | 20.6 (3.0) | single-dose | Bapineuzumab (N-terminal), 0.15 mg/kg, IV, SD | Placebo | 1. MMSE; 2. PK/PD assessments. |
| | | | 6 | | Na | | 72.2 (8.4) | | 1 (16.7) | | Na | | Na | | 21.0 (3.6) | | | Bapineuzumab (N-terminal), 0.5 mg/kg,IV, SD | | |
| | | | 6 | | Na | | 72.2 (10.9) | | 3 (50.0) | | Na | | Na | | 21.0 (4.6) | | | Bapineuzumab (N-terminal), 1.0 mg/kg, IV, SD | | |
| | | | 6 | | Na | | 64.8 (5.2) | | 3 (50.0) | | Na | | Na | | 20.2 (2.8) | | | Bapineuzumab (N-terminal), 2.0 mg/kg, IV, SD | | |
| Landen at al.[28] | I | mild-moderate | 4 | 11 | 1 (25.0) | 1 (9.1) | 69.0 (8.1) | 71.8(7. | 2 (50.0) | 3 | Na | Na | Na | Na | 22.5 (2.5) | 21.5 | single-do | Ponezumab, 0.1 mg/kg, | Placeb | 1. MMSE, |

| | | | | | | | | | | | | | | | | | | | | |
|---|---|---|---|---|---|---|---|---|---|---|---|---|---|---|---|---|---|---|---|---|
| (2013) | | (16 to 26) | | | | | | 0) | | (27.3) | | | | | | (3.4) | se | IV, SD | o | ADAS-Cog, CogState; 2. PK/PD assessments. |
| | | | 4 | | 2 (50.0) | | 70.0 (3.7) | | 1 (25.0) | | Na | | Na | | 22.5 (2.5) | | | Ponezumab, 0.3 mg/kg, IV, SD | | |
| | | | 4 | | 3 (75.0) | | 70.0 (3.7) | | 2 (50.0) | | Na | | Na | | 21.3 (3.6) | | | Ponezumab, 1mg/kg, IV, SD | | |
| | | | 6 | | 2 (33.3) | | 66.5 (11.8) | | 2 (33.3) | | Na | | Na | | 22.7 (3.3) | | | Ponezumab, 3mg/kg, IV, SD | | |
| | | | 8 | | 1 (12.5) | | 72.1 (8.0) | | 4 (50.0) | | Na | | Na | | 21.9 (2.8) | | | Ponezumab, 10mg/kg, IV, SD | | |
| Farlow at al. [29] (2012) NCT00329082 | II | mild-moderate (15 to 26) | 10 | 10 | Na | Na | Na | Na | Na | Na | Na | Na | Na | Na | Na | Na | 12 weeks | Solanezumab (Central-domainl), 100 mg, IV, Q4W | Placebo | 1. ADAS-Cog11, ADAS-Cog14; 2. CSF biomarkers (Aβ42, Aβ40, total tau, and p-tau) and plasma (Aβ40, and Aβ42); 2. PK/PD assessments. |
| | | | 11 | | Na | | Na | | Na | | Na | | Na | | Na | | | Solanezumab (Central-domainl), 100 mg, IV, QW | | |
| | | | 10 | | Na | | Na | | Na | | Na | | Na | | Na | | | Solanezumab, 400 mg, IV, Q4W | | |
| | | | 11 | | Na | | Na | | Na | | Na | | Na | | Na | | | Solanezumab (Central-domainl), 400 mg, IV, QW | Placebo | |
| Miyoshi at al. [30] (2013) NCT00607308 | I | mild-moderate (16 to 26) | 3 | 5 | 2 (66.7) | 4 (80.0) | 65.7 (6.4) | 72.2(7.1) | 2 (66.7) | 3 (60.0) | Na | Na | Na | Na | 16.33 (3.2) | 21.20 (3.6) | single-dose | Ponezumab (C-terminal), 0.1 mg/kg, IV, SD | Placebo | 1. ADAS-Cog11, ADAS-Cog14; 2. CSF biomarkers (Aβ1-x, Aβ42, Aβ40, total tau, and p-tau) and plasma |
| | | | 3 | | 1 (33.3) | | 67.7 (8.1) | | 2 (66.7) | | Na | | Na | | 19.67 (1.5) | | | Ponezumab(C-terminal), 0.5 mg/kg, IV, SD | | |
| | | | 3 | | 0 (0.0) | | 67.0 | | 0 (0.0) | | Na | | Na | | 18.33 | | | Ponezumab(C-terminal), | | |

| | | | | | | | | | | | | | | | | | | | | |
|---|---|---|---|---|---|---|---|---|---|---|---|---|---|---|---|---|---|---|---|---|
| | | | | | | | (10.1) | | | | | | | | (1.5) | | | 1 mg/kg, IV, SD | | (Aβ40, and Aβ1-40); 2. PK/PD assessments. |
| | | | 3 | | 1 (33.3) | | 68.3 (5.1) | | 1 (33.3) | | Na | | Na | | 20.67 (6.1) | | | Ponezumab(C-terminal), 5 mg/kg, IV, SD | | |
| | | | 3 | | 3 (100.0) | | 71.7 (7.2) | | 3 (100.0) | | Na | | Na | | 24.67 (1.5) | | | Ponezumab(C-terminal), 10 mg/kg, IV, SD | | |
| Siemers at al. [31] (2010) H8A-LC-LZAH | Na | mild-moderate (14 to 26) | 4 | 3 | 4 (100.0) | 2 (66.7) | 61.0 (6.2) | 70.3 (5.5) | 1 (25.0) | 3 (100.0) | Na | Na | Na | Na | 18.6 (3.9) | 19.3 (2.5) | single-dose | Solanezumab (Central-domainl), 0.5mg/kg, IV, SD | Placebo | 1. ADAS-Cog; 2. PK/PD assessments. |
| | | | 4 | | 2 (50.0) | | 71.5 (12.3) | | 1 (25.0) | | Na | | Na | | 19.0 (2.4) | | | Solanezumab (Central-domainl), 1.5mg/kg, IV, SD | | |
| | | | 4 | | 3 (75.0) | | 67.5 (7.6) | | 2 (50.0) | | Na | | Na | | 20.2 (3.5) | | | Solanezumab (Central-domainl), 4mg/kg, IV, SD | | |
| | | | 4 | | 4 (100.0) | | 75.3 (3.9) | | 1 (25.0) | | Na | | Na | | 21.0 (4.8) | | | Solanezumab (Central-domainl), 10mg/kg, IV, SD | | |
| Black at al. [26] (2010) | Na | mild-moderate (14 to 26) | 6 | 8 | Na | Na | 74.67 (5.7) | 69.88 (10.7) | 3 (50.0) | 7 (87.5) | Na | Na | Na | Na | 21.8 (Na) | 20.8 (Na) | single-dose | Bapineuzumab (N-terminal), 0.5mg/kg, IV, SD | Placebo | 1. MMSE; 2. PK assessments. |
| | | | 6 | | Na | | 72.33 (9.9) | | 1 (16.7) | | Na | | Na | | 21.3 (Na) | | | Bapineuzumab (N-terminal), 1.5mg/kg, IV, SD | | |
| | | | 10 | | Na | | 74.70 (7.4) | | 3 (30.0) | | Na | | Na | | 22.3 (Na) | | | Bapineuzumab (N-terminal), 5mg/kg, IV, SD | | |

#### Appendix 3:  Table of excluded studies

Exclusion codes:

1. Post-hoc or secondary analyses of main studies

2. Studies involving non-human subjects.

3. Study is not a randomised controlled trial

4. Studies including participants diagnosed with dementia other than AD (familial AD and other types of dementias were excluded)

5. Intervention no mAbs against amyloid-β

6. Reviews

7. Cannot locate full text

8. Conference proceedings

9. Protocol for a study not meeting the inclusion criteria

10. Not an outcome of interest

11. Other – please specify

| Study ID | Title | DOI | Reason for exclusion |
|---|---|---|---|
| Hu at al. [32] (2015) | Confirmatory Population Pharmacokinetic Analysis for Bapineuzumab Phase 3 Studies in Patients with Mild to Moderate Alzheimer's Disease | 10.1002/jcph.393 | Exclude 1 |
| Tariot at al. [33] (2018) | The Alzheimer's Prevention Initiative Autosomal-Dominant Alzheimer's Disease Trial: A study of crenezumab versus placebo in preclinical PSEN1 E280A mutation carriers to evaluate efficacy and safety in the treatment of autosomal-dominant Alzheimer's disease, including a placebo-treated noncarrier cohort | 10.1016/j.trci.2018.02.002 | Exclude 9 |
| Liu at al. [34] (2015) | Delayed-start analysis: Mild Alzheimer's disease patients in solanezumab trials, 3.5 years | 10.1016/j.trci.2015.06.006 | Exclude 1 |
| Salloway at al. [35] (2018) | Long-Term Follow Up of Patients with Mild-to-Moderate Alzheimer's Disease Treated with Bapineuzumab in a Phase III, Open-Label, Extension Study | 10.3233/jad-171157 | Exclude 3 |
| Henley at al. [36] (2012) | Safety profile of Alzheimer's disease populations in Alzheimer's Disease Neuroimaging Initiative and other 18-month studies | 10.1016/j.jalz.2011.05.2413 | Exclude 1 |
| Liu at al. [37] (2018) | Biomarker pattern of ARIA-E participants in phase 3 randomized clinical trials with bapineuzumab | 10.1212/wnl.0000000000005060 | Exclude 1 |
| Liu at al. [38] (2015) | Cognitive and functional decline and their relationship in patients with mild Alzheimer's dementia | 10.3233/JAD-140792 | Exclude 1 |
| Siemers at al. [39] (2023) | ACU193, a Monoclonal Antibody that Selectively Binds Soluble Aβ Oligomers: Development Rationale, Phase 1 Trial Design, and Clinical Development Plan | 10.14283/jpad.2022.93 | Exclude 9 |
| Carlson at al. [40] (2016) | Amyloid-related imaging abnormalities from trials of solanezumab for Alzheimer's disease | 10.1016/j.dadm.2016.02.004 | Exclude 3 |
| Barakos al. | MR Imaging Features of Amyloid-Related Imaging Abnormalities | 10.3174/ajnr.A3500 | Exclude 1 |

| | | | |
|---|---|---|---|
| [41] (2013) | | | |
| Lacey at al. [42] (2015) | Quality of Life and Utility Measurement in a Large Clinical Trial Sample of Patients with Mild to Moderate Alzheimer's Disease: Determinants and Level of Changes Observed | 10.1016/j.jval.2015.03.1787 | Exclude 5 |
| Ezzati1 at al. [43] (2021) | Application of predictive models in boosting power of Alzheimer's disease clinical trials: A post hoc analysis of phase 3 solanezumab trials | 10.1002/trc2.12223 | Exclude 1 |
| Portron at al.[44] (2020) | A Phase I Study to Assess the Effect of Speed of Injection on Pain, Tolerability, and Pharmacokinetics After High-volume Subcutaneous Administration of Gantenerumab in Healthy Volunteers | 10.1016/j.clinthera.2019.11.015 | Exclude 4 |
| Euctr at al. [45] (2020) | A Clinical Trial to evaluate AL002 in Participants with early Alzheimer's Disease | https://trialsearch.who.int/Trial2.aspx?TrialID=EUCTR2019-001476-11-NL | Exclude 9 |
| Romenets at al. [46] (2020) | Baseline demographic, clinical, and cognitive characteristics of the Alzheimer's Prevention Initiative (API) Autosomal-Dominant Alzheimer's Disease Colombia Trial | 10.1002/alz.12109 | Exclude 10 |
| Maslyar at al. [47] (2022) | A Phase 1 Study of AL003 in Healthy Volunteers and Participants with Alzheimer's disease | https://www.cochranelibrary.com/central/doi/10.1002/central/CN-02421827/full | Exclude 5 |
| Toth at al. [48] (2017) | Pooled safety analysis of evolocumab in over 6000 patients from double-blind and open-label extension studies | 10.1161/CIRCULATIONAHA.116.025233 | Exclude 3 |
| Sevigny at al. [49] (2015) | ADUCANUMAB (BIIB037), AN ANTI-AMYLOID BETA MONOCLONAL ANTIBODY, IN PATIENTS WITH PRODROMAL OR MILD ALZHEIMER'S DISEASE: INTERIM RESULTS OFA RANDOMIZED, DOUBLE-BLIND, PLACEBOCONTROLLED, PHASE 1B STUDY | https://www.cochranelibrary.com/central/doi/10.1002/central/CN-01163350/full | Exclude 8 |
| Galvin at al. [50] (2021) | ICARE AD-US: design of a prospective, single-arm, multicenter, noninterventional real-world study of aducanumab in the United States | 10.1002/alz.057522 | Exclude 8 |
| Klein at al. [51] (2020) | Thirty-Six-Month Amyloid Positron Emission Tomography Results Show Continued Reduction in Amyloid Burden with Subcutaneous Gantenerumab | 10.14283/jpad.2020.68 | Exclude 3 |
| Blennow at al. [52] (2016) | Gantenerumab treatment reduces biomarkers of neuronal and synaptic degeneration in Alzheimer's disease | https://www.cochranelibrary.com/central/doi/10.1002/central/CN-01251227/full | Exclude 8 |
| Lynch at al. | A stepwise tier-based approach for determining patient eligibility in clarity AD: a phase 3 placebo-controlled, | 10.14283/jpad.2021.57 | Exclude 8 |

| | | | |
|---|---|---|---|
| [53] (2021) | double-blind study to confirm the safety and efficacy of lecanemab (BAN2401) 10 mg/kg biweekly in patients with early alzheimer's disease | | |
| Burkett at al. [54] (2021) | Considerations for the real-worldmanagement of ARIA from the aducanumab phase 3 studies EMERGE and ENGAGE | 10.1002/alz.057498 | Exclude 8 |
| Lin at al. [55] (2018) | BASELINE CHARACTERICS FROM A PHASE 3 TRIAL OF CRENEZUMAB IN PRODROMAL TO MILD ALZHEIMER'S DISEASE (CREAD) | 10.1016/j.jalz.2018.06.2339 | Exclude 8 |
| Haeberlein at al. [56] (2018) | Aducanumab titration dosing regimen: 36-month analyses from prime, a phase 1b study in patients with early Alzheimer's disease | 10.14283/jpad.2018.39 | Exclude 8 |
| Klein at al. [57] (2019) | Thirty-six-month amyloid pet results show continued reduction in amyloid burden with gantenerumab | 10.14283/jpad.2019.47 | Exclude 8 |
| Carlson at al. [58] (2011) | Prevalence of asymptomatic vasogenic edema in pretreatment Alzheimer's disease study cohorts from phase 3 trials of semagacestat and solanezumab | 10.1016/j.jalz.2011.05.2353 | Exclude 8 |
| Molinuevo at al. [59] (2019) | BAN2401 IN EARLY ALZHEIMER'S DISEASE: NEURODEGENERATION BIOMARKER ANALYSIS FROM A RANDOMIZED PHASE 2 STUDY | 10.1016/j.jalz.2019.08.004 | Exclude 8 |
| Rosenstiel at al. [60] (2018) | 24-MONTH ANALYSIS OF APOE ε4 CARRIERS IN PRIME: a RANDOMIZED PHASE 1B STUDY OF THE ANTI–AMYLOID BETA MONOCLONAL ANTIBODY ADUCANUMAB | 10.1016/j.jalz.2018.06.042 | Exclude 8 |
| Hehn at al. [61] (2019) | Baseline characteristics from ENGAGE and EMERGE: two phase 3 studies to evaluate aducanumab in patients with early Alzheimer's disease | https://www.cochranelibrary.com/central/doi/10.1002/central/CN-01999385/full | Exclude 8 |
| Voyle at al. [62] (2018) | THE EFFECT OF LOW DOSES OF GANTENERUMAB ON AMYLOID AND TAU BIOMARKERS IN CEREBROSPINAL FLUID (CSF) IN THE MARGUERITE ROAD STUDY | 10.1016/j.jalz.2018.06.2379 | Exclude 8 |
| Viglietta at al. [63] (2016) | Randomized, Placebo-Controlled, Phase 1b study of the anti-Beta-amyloid antibody Aducanumab (BIIB037) in patients with prodromal or Mild Alzheimer's Disease: interim results | 10.3233/JAD-169002 | Exclude 8 |
| Schwarz at | Effect of solanezumab on biomarkers of neurodegeneration in the expedition3 trial in mild Alzheimer | https://www.cochranelibrary.com/central/ | Exclude 8 |

| | | | |
|---|---|---|---|
| al. [64] (2017) | disease | doi/10.1002/central/CN-01457704/full | |
| Benzinger at al. [65] (2021) | Defining a standardized mri acquisition protocol to be proposed to icare ad-us sites for baseline and aria monitoring | 10.14283/jpad.2021.57 | Exclude 8 |
| Lane at al. [66] 2022 | Post Graduate Open-label Rollover Study: evaluation of Subcutaneous Gantenerumab Long-term Safety, Tolerability, and Efficacy in Participants with Alzheimer's Disease | https://www.cochranelibrary.com/central/doi/10.1002/central/CN-02421856/full | Exclude 8 |
| Ratti at al. [67] (2018) | RANDOMIZED, DOUBLE-BLIND, PLACEBO-CONTROLLED STUDY TO ASSESS TREATMENT OF BIIB092 IN SUBJECTS WITH EARLY ALZHEIMER'S DISEASE: TANGO PHASE 2 STUDY DESIGN | 10.1016/j.jalz.2018.06.1388 | Exclude 8 |
| Uenaka at al. [68] (2012) | Comparison of pharmacokinetics, pharmacodynamics, safety, and tolerability of the amyloid β monoclonal antibody solanezumab in japanese and white patients with mild to moderate Alzheimer disease | 10.1097/WNF.0b013e31823a13d3 | Exclude 10 |
| Burstein at al. [69] (2013) | Safety and pharmacology of ponezumab (PF-04360365) after a single 10-minute intravenous infusion in subjects with mild to moderate Alzheimer disease | 10.1097/WNF.0b013e318279bcfa | Exclude 3 |
| Liu at al. [70]. (2013) | Relationship between cognitive and functional progression in patients with mild Alzheimer's disease | https://www.cochranelibrary.com/central/doi/10.1002/central/CN-01064098/full | Exclude 7 |
| Logovinsky at al. [71] (2013) | First-in-human study of BAN2401, a novel monoclonal antibody against amyloid-beta protofibrils | https://www.cochranelibrary.com/central/doi/10.1002/central/CN-01064108/full | Exclude 7 |
| Lucas at al. [72] (2013) | Intravenous bapineuzumab in mild to moderate Alzheimer's disease: results from two double-blind, placebo-controlled phase 3 trials | https://www.cochranelibrary.com/central/doi/10.1002/central/CN-01064099/full | Exclude 7 |
| Cummings at al. [73] (2014) | A randomized, double-blind, placebocontrolled phase 2 study to evaluate the efficacyand safety of crenezumab in patients with mild to moderate Alzheimer's disease | https://www.cochranelibrary.com/central/doi/10.1002/central/CN-01056778/full | Exclude 7 |
| Liu at al. [74] (2014) | Cognitive and functional decline and their relationship in patients with mild alzheimer's disease | https://www.cochranelibrary.com/central/doi/10.1002/central/CN-01062879/full | Exclude 7 |
| Novak at al. | Rates of change in brain volume with subcutaneous bapineuzumab | https://www.cochranelibrary.com/central/ | Exclude 7 |

| | | | |
|---|---|---|---|
| [75] (2014) | | doi/10.1002/central/CN-01757223/full | |
| Andreasen at al. [76] (2015) | First administration of the Fc-attenuated anti-beta amyloid antibody GSK933776 to patients with mild Alzheimer's disease: a randomized, placebo-controlled study | 10.1371/journal.pone.0098153 | Exclude 10 |
| Carlson at al. [77] (2015) | Safety of solanezumab in the EXPEDITION-EXT study up to 2 years in a mild to moderate Alzheimer's disease population | https://www.cochranelibrary.com/central/doi/10.1002/central/CN-01163311/full | Exclude 7 |
| Samtani at al. [78] (2015) | Alzheimer's disease assessment scale-cognitive 11-item progression model in mild-to-moderate Alzheimer's disease trials of bapineuzumab | 10.1016/j.trci.2015.09.001 | Exclude 1 |
| Sevigny at al. [49] (2015) | Aducanumab (BIIB037), an anti-amyloid beta monoclonal antibody, in patients with prodromal or mild Alzheimer's disease: interim results of a randomized, double-blind, placebo controlled, phase 1b study | https://www.cochranelibrary.com/central/doi/10.1002/central/CN-01163350/full | Exclude 8 |
| Xu at al. [79] (2015) | Alzheimer's disease progression model using disability assessment for dementia scores from bapineuzumab trials | 10.1016/j.trci.2015.06.005 | Exclude 1 |
| Blaettler at al. [80] (2016) | Clinical trial design of cread: a randomized, double-blind, placebo controlled, parallel-group phase 3 study to evaluate crenezumab treatment in patients with prodromalto-mild Alzheimer's disease | https://www.cochranelibrary.com/central/doi/10.1002/central/CN-01251145/full | Exclude 9 |
| Chen at al. [81] (2016) | Quantile regression to characterize solanezumab effects in Alzheimer's disease trials | 10.1016/j.trci.2016.07.005 | Exclude 1 |
| Degenhardt at al. [82] (2016) | Florbetapir F18 PET Amyloid Neuroimaging and Characteristics in Patients With Mild and Moderate Alzheimer Dementia | 10.1016/j.psym.2015.12.002 | Exclude 1 |
| Ivanoiu at al. [83] (2016) | Long-term safety and tolerability of bapineuzumab in patients with Alzheimer's disease in two phase 3 extension studies | 10.1186/s13195-016-0193-y | Exclude 3 |
| JapicCti at al. [84] (2016) | Single-dose and multiple-Dose, dose-escalation study with LY2599666 to evaluate the safety, pharmacokinetics, and tolerability in healthy subjects and patients with mild cognitive impairment due to Alzheimer's disease and mild to moderate Alzheimer's disease | https://www.cochranelibrary.com/central/doi/10.1002/central/CN-01308190/full | Exclude 7 |
| JapicCti at | A single- and multiple-dose study to assess the safety, tolerability, pharmacokinetics, and | https://www.cochranelibrary.com/central/ | Exclude 7 |

| | | | |
|---|---|---|---|
| al. [85] (2016) | pharmacodynamics of single and multiple intravenous doses of LY3002813 | doi/10.1002/central/CN-01308191/full | |
| Roher at al. [86] (2016) | Chemical and neuropathological analyses of an alzheimer's disease patient treated with solanezumab | https://www.embase.com/search/results?subaction=viewrecord&id=L612651154&from=export | Exclude 7 |
| Haeberlein at al. [87] (2017) | Clinical Development of Aducanumab, an Anti-Aβ Human Monoclonal Antibody Being Investigated for the Treatment of Early Alzheimer's Disease | 10.14283/jpad.2017.39 | Exclude 7 |
| Euctr at al. [88] (2017) | To evaluate the efficacy and safety of CNP520 in participants at risk for the onset of clinical symptoms of Alzheimer's Disease (AD) | https://www.cochranelibrary.com/central/doi/10.1002/central/CN-01457021/full | Exclude 7 |
| Fleisher at al. [89] (2017) | Use of white matter reference regions for detection of change in florbetapir positron emission tomography from completed phase 3 solanezumab trials | 10.1016/j.jalz.2017.02.009 | Exclude 1 |
| Liu at al. [90] (2018) | Delayed-Start Analyses in the Phase 3 Solanezumab EXPEDITION3 Study in Mild Alzheimer's Disease | 10.14283/jpad.2018.1 | Exclude 1 |
| Salloway at al. [91] (2018) | Long-Term Safety and Efficacy of Bapineuzumab in Patients with Mild-to-Moderate Alzheimer's Disease: a Phase 2, Open-Label Extension Study | 10.2174/1567205015666180821114813 | Exclude 3 |
| Salloway at al. [35] (2018) | Long-Term Follow Up of Patients with Mild-to-Moderate Alzheimer's Disease Treated with Bapineuzumab in a Phase III, Open-Label, Extension Study | 10.3233/jad-171157 | Exclude 3 |
| Salloway at al. [92] (2021) | A trial of gantenerumab or solanezumab in dominantly inherited Alzheimer's disease | 10.1038/s41591-021-01369-8 | Exclude 4 |
| Mathurin at al. [93] (2022) | Amyloid-Related Imaging Abnormalities in the DIAN-TU-001 Trial of Gantenerumab and Solanezumab: Lessons from a Trial in Dominantly Inherited Alzheimer Disease | 10.1002/ana.26511 | Exclude 4 |
| Shcherbinin at al. [94] (2022) | Association of Amyloid Reduction after Donanemab Treatment with Tau Pathology and Clinical Outcomes: The TRAILBLAZER-ALZ Randomized Clinical Trial | 10.1001/jamaneurol.2022.2793 | Exclude 1 |
| Svaldi at al. [95] (2022) | Magnetic resonance imaging measures of brain volumes across the EXPEDITION trials in mild and moderate Alzheimer's disease dementia | 10.1002/trc2.12313 | Exclude 1 |

### Appendix 4: Rick of Bias

The following table shows the risk of bias assessment for the individual domains.

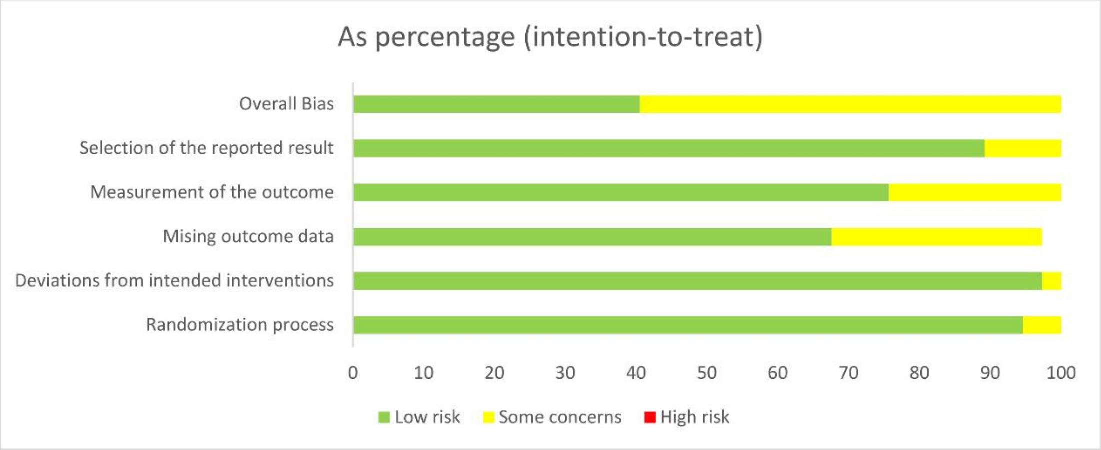

Risk of bias assessment for the individual studies:

| Unique ID | D1 | D2 | D3 | D4 | D5 | Overall |
|---|---|---|---|---|---|---|
| Cummings at al (2018) | + | + | ! | + | + | ! |
| Salloway at al (2018) | + | + | + | + | + | + |
| Ostrowitzki at al (2022)* | + | + | + | ! | + | ! |
| Doody at al (2014)* | + | + | + | + | + | + |
| Haeberlein at al (2022)* | + | + | + | ! | ! | ! |
| Rinne at al (2010) | + | + | + | + | + | + |
| Doody at al (2014)** | + | + | + | + | ! | ! |
| Honig at al (2018) | + | + | + | + | ! | ! |
| Ostrowitzki at al (2022)** | + | + | + | ! | + | ! |
| Landen at al (2013) | + | + | + | + | + | + |
| Lu at al (2018) | + | + | + | + | + | + |
| Salloway at al (2009) | + | + | ! | + | + | ! |
| Farlow at al (2012) | + | + | + | + | + | + |
| Arai at al (2016) | ! | + | + | + | + | ! |
| Salloway at al (2014)* | + | + | + | ! | + | ! |
| Vandenberghe at al (2016)* | + | + | + | ! | + | ! |
| Landen at al (2017)* | + | + | ! | + | + | ! |
| Delnomdedieu at al (2016) | + | + | ! | + | + | + |
| Ostrowitzki at al (2017) | + | + | ! | ! | + | ! |
| Logovinsky at al (2016) | + | + | + | + | + | + |
| Brody at al (2016) | + | + | ! | + | + | + |
| Ferrero at al(2016) | + | + | + | + | + | + |
| Swanson at al (2021) | + | + | ! | + | + | ! |
| Mintun at al (2021) | + | + | ! | + | + | ! |
| Dyck at al (2022) | + | + | + | + | + | + |
| Sevigny at al (2016) | + | + | ! | + | + | ! |
| Lowe at al (2021) | + | + | ! | + | + | ! |
| Haeberlein at al (2022)** | + | + | + | ! | ! | ! |
| Vandenberghe at al (2016)** | + | + | + | ! | + | ! |
| Salloway at al (2014)** | + | + | + | ! | + | ! |
| Sims at al (2023) | + | + | ! | + | + | ! |
| Sperling at al (2023) | + | + | + | + | + | + |
| Guthrie at al (2020) | + | + | + | + | + | + |
| Landen at al (2017)** | + | + | + | + | + | + |
| Siemers at al.(2010) | ! | + | | + | + | ! |
| Black at al.(2010) | + | ! | + | + | + | ! |
| Miyoshi at al.(2013) | + | + | + | + | + | + |

+ Low risk
! Some concerns
- High risk

D1 Randomisation process
D2 Deviations from the intended interventions
D3 Missing outcome data
D4 Measurement of the outcome
D5 Selection of the reported result

### Appendix 5: Funnel plots of meta-analyses

#### 5.1 ADAS-cog

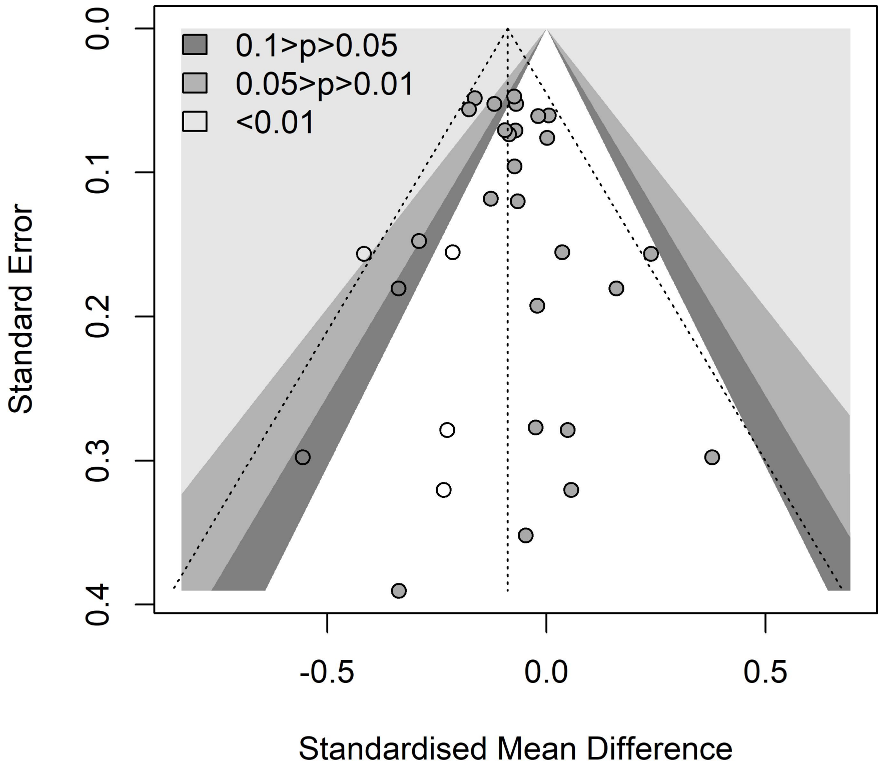

#### 5.2 CDR-SB

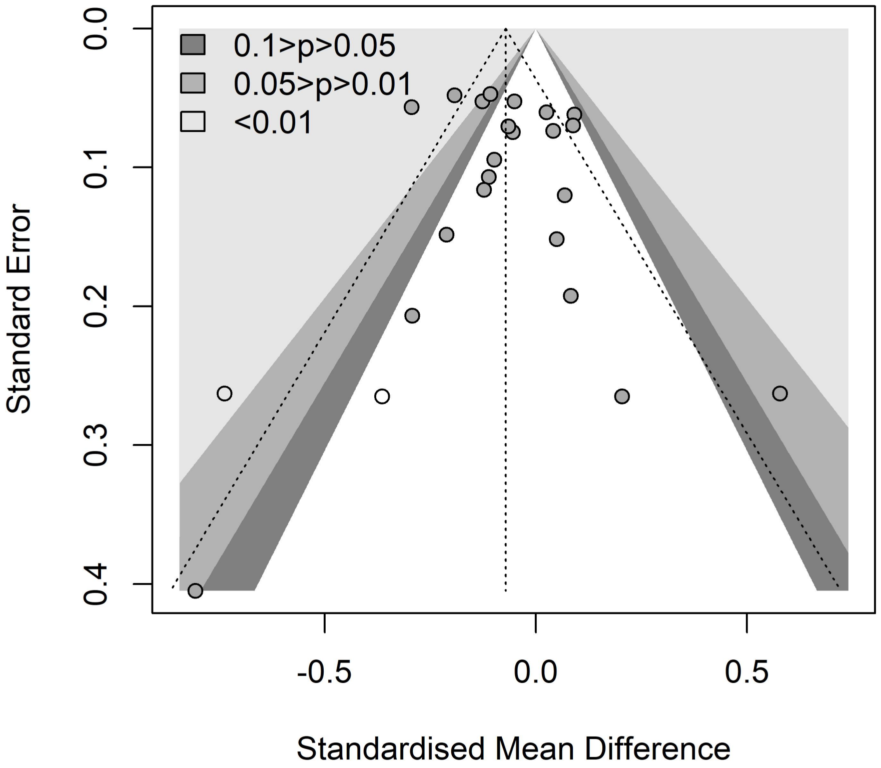
0.1>p>0.05
0.05>p>0.01
<0.01
Standard Error
0.0
0.1
0.2
0.3
0.4
-0.5
0.0
0.5
Standardised Mean Difference

**5.3 MMSE**

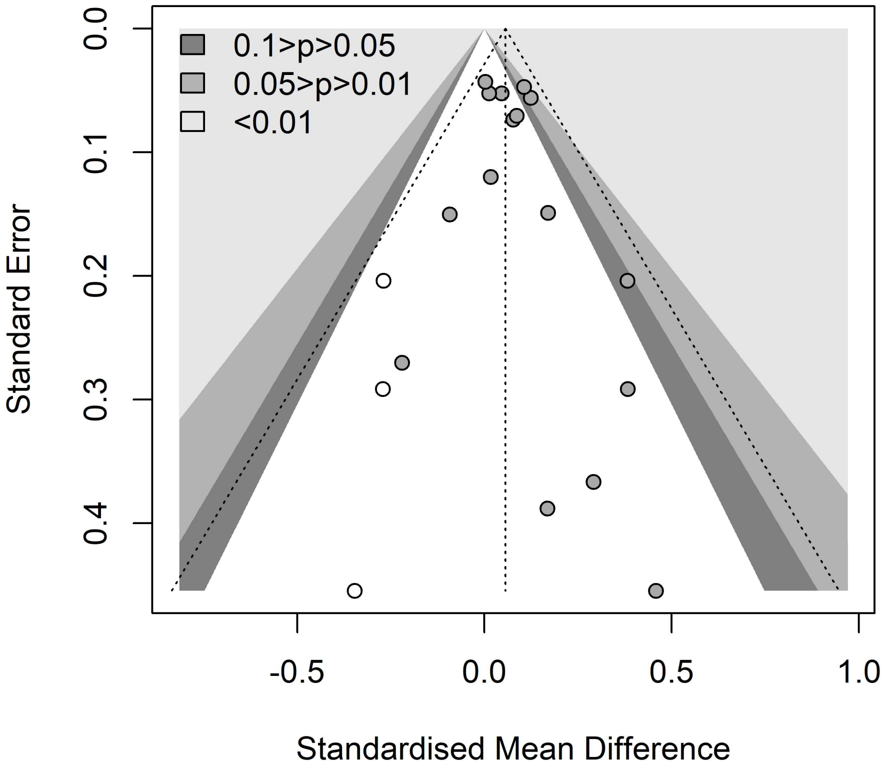
0.1>p>0.05
0.05>p>0.01
<0.01
Standard Error
0.0
0.1
0.2
0.3
0.4
-0.5
0.0
0.5
1.0
Standardised Mean Difference

#### 5.4 Amyloid PET

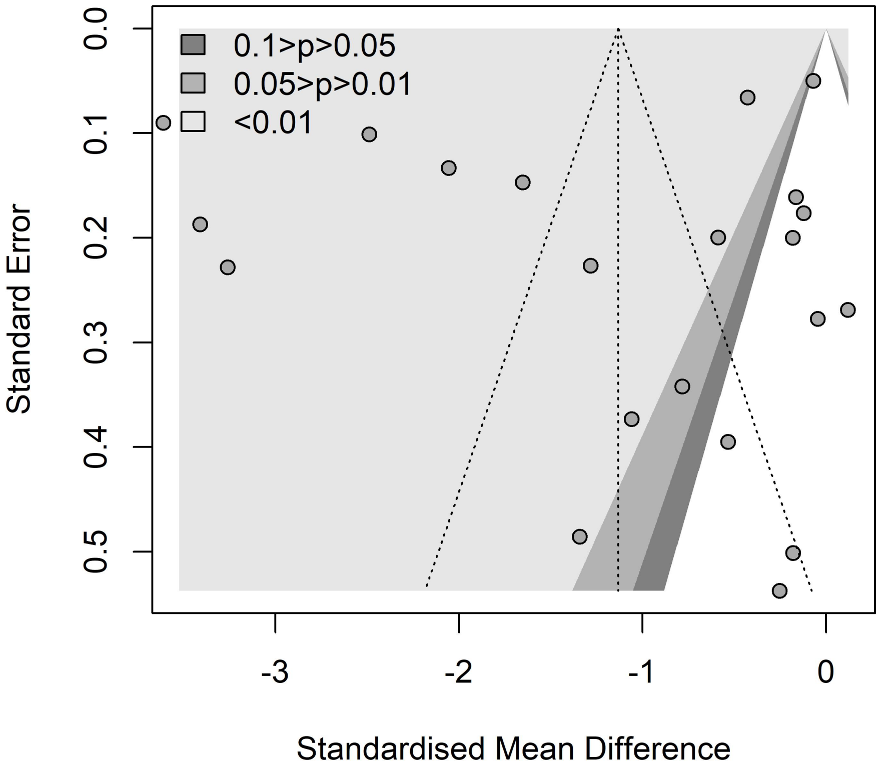

##### 5.5 Volumes of the whole brain

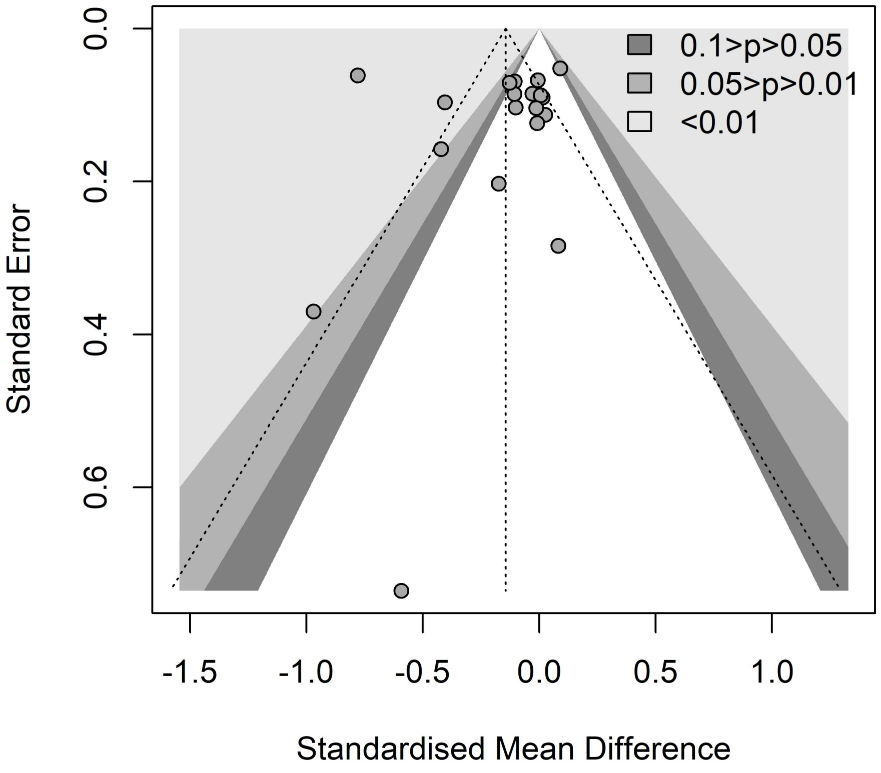

#### 5.6 Volumes of hippocampus

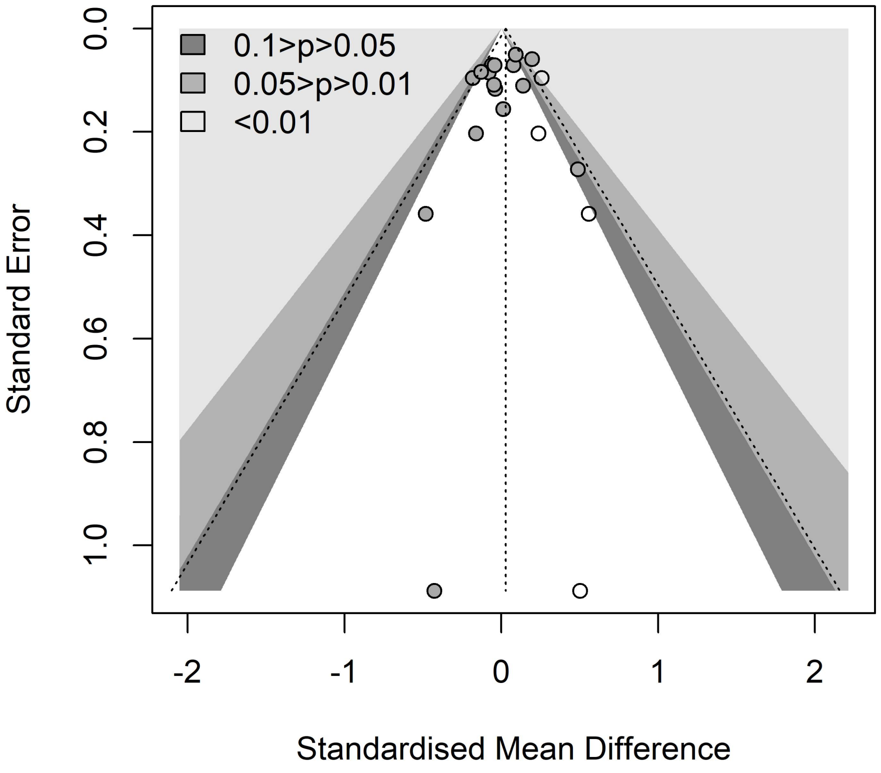

**5.7 Volumes of ventricle**

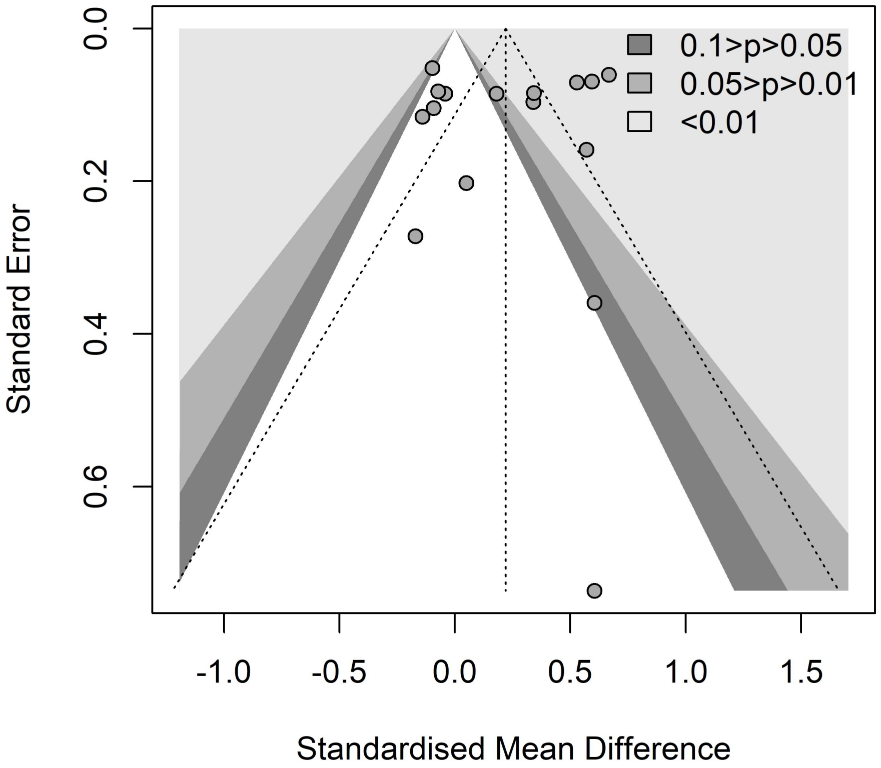

**5.8 CSF Aβ42**

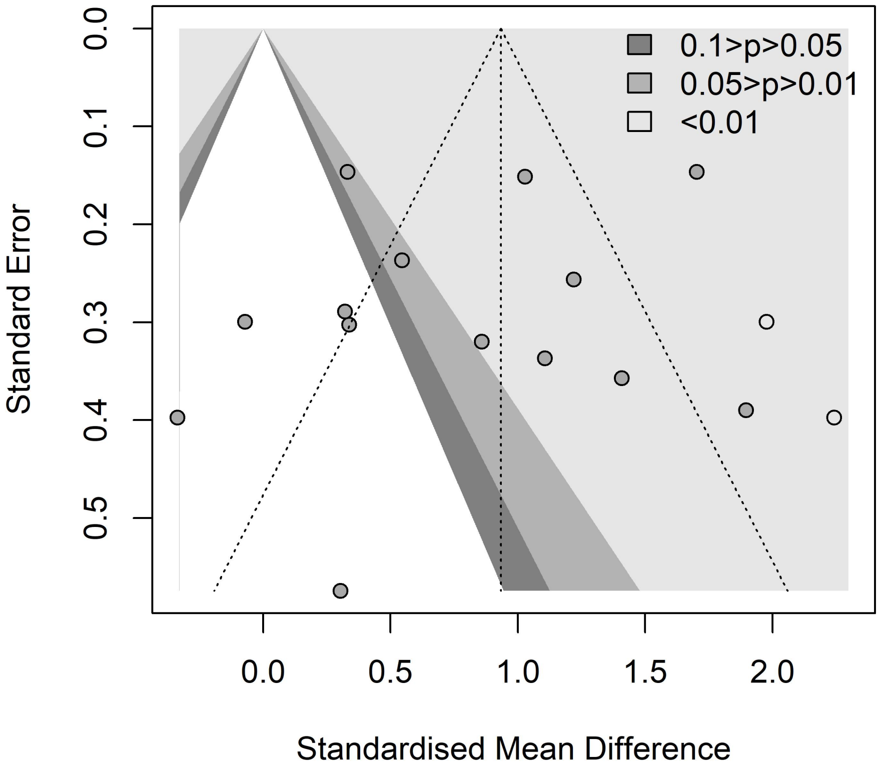

0.1>p>0.05
0.05>p>0.01
<0.01
Standard Error
0.0
0.1
0.2
0.3
0.4
0.5
0.0
0.5
1.0
1.5
2.0
Standardised Mean Difference

#### 5.9 CSF p-tau

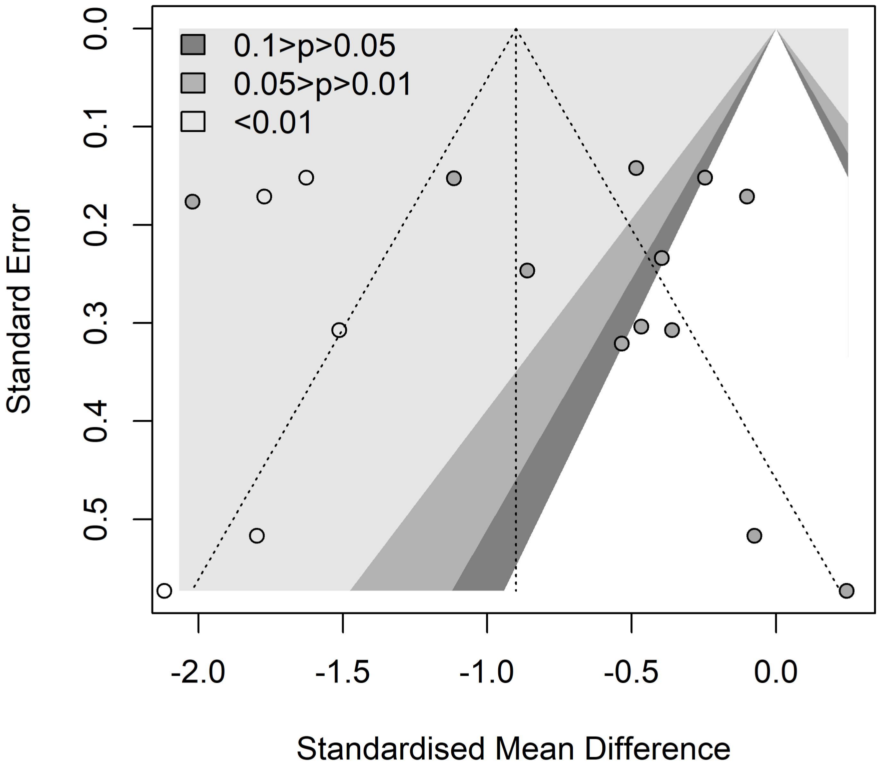
0.1>p>0.05
0.05>p>0.01
<0.01
Standard Error
0.0
0.1
0.2
0.3
0.4
0.5
-2.0
-1.5
-1.0
-0.5
0.0
Standardised Mean Difference

##### 5.10 CSF total tau

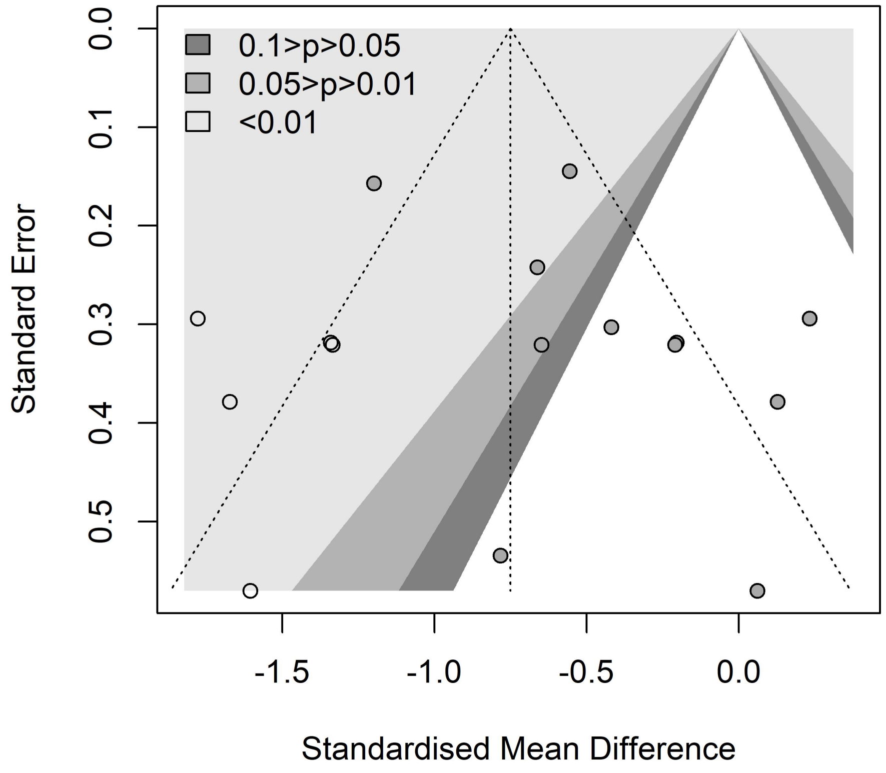

##### 5.11 ARIA-E

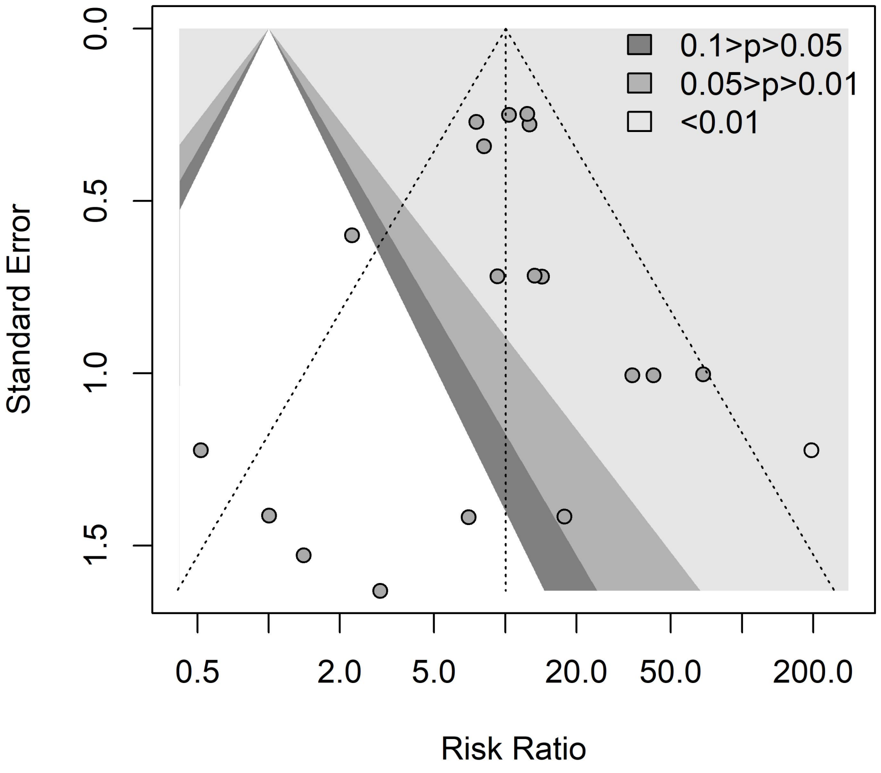
0.1>p>0.05
0.05>p>0.01
<0.01
Standard Error
0.0
0.5
1.0
1.5
0.5
2.0
5.0
20.0
50.0
200.0
Risk Ratio

**5.12 ARIA-H**

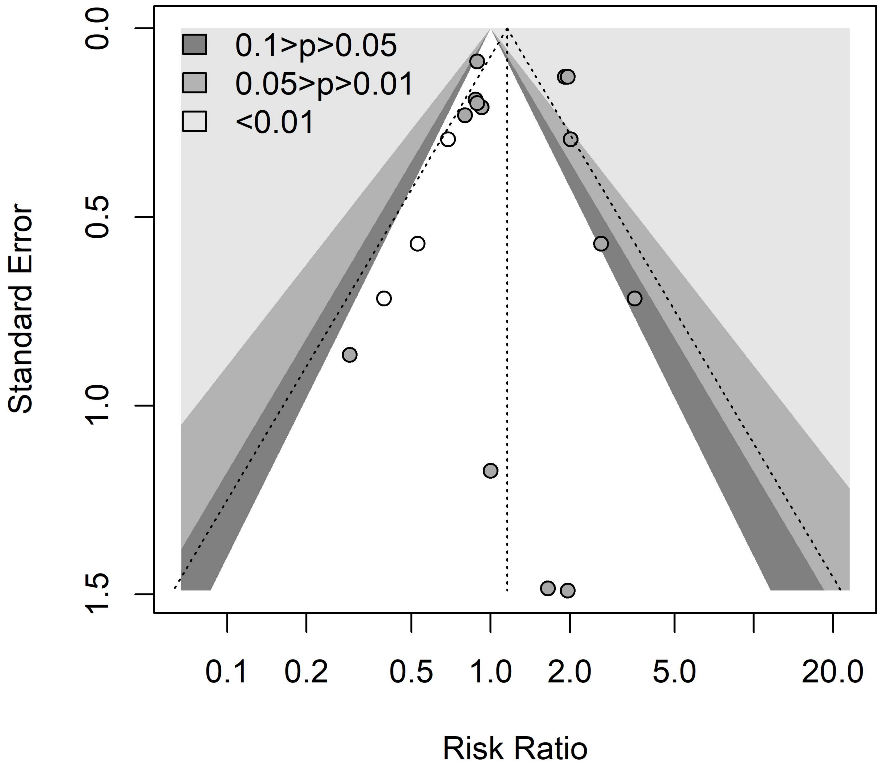
0.1>p>0.05
0.05>p>0.01
<0.01
Standard Error
0.0
0.5
1.0
1.5
0.1
0.2
0.5
1.0
2.0
5.0
20.0
Risk Ratio

### Appendix 6: Forest plots for the primary outcomes of MMSE

| Study | Anti-Aβ mAbs Mean | SD | Total | Placebo Mean | SD | Total | SMD (95% CI) | Weight (%) |
|---|---|---|---|---|---|---|---|---|
| **Aducanumab** | | | | | | | | |
| Haeberlein at al (2022) | -3.05 | 5.81 | 1090 | -3.30 | 4.68 | 548 | 0.05(-0.06, 0.15) | 13.8 |
| Haeberlein at al (2022) | -3.45 | 3.72 | 1102 | -3.50 | 4.67 | 545 | 0.01(-0.09, 0.11) | 13.8 |
| Jeff sevigny at al (2016) | -1.35 | 3.81 | 102 | -2.81 | 3.79 | 32 | 0.38(-0.02, 0.78) | 0.9 |
| **Subtotal**($I^2$ = 35%, p = 0.21) | | | **2294** | | | **1125** | **0.04(-0.03, 0.11)** | **28.5** |
| **Donanemab** | | | | | | | | |
| Mintun at al (2021) | -2.35 | 3.68 | 91 | -2.98 | 3.70 | 90 | 0.17(-0.12, 0.46) | 1.7 |
| Sims at al (2023) | -2.75 | 3.74 | 600 | -3.22 | 3.85 | 679 | 0.12( 0.01, 0.23) | 12.0 |
| **Subtotal**($I^2$ = 0%, p = 0.77) | | | **691** | | | **769** | **0.13( 0.03, 0.23)** | **13.8** |
| **Bapineuzumab** | | | | | | | | |
| Salloway at al (2014) | -4.17 | 5.23 | 1279 | -4.18 | 4.32 | 925 | 0.00(-0.08, 0.09) | 20.1 |
| Lu at al (2018) | -1.40 | 1.92 | 30 | -2.10 | 3.40 | 10 | 0.29(-0.43, 1.01) | 0.3 |
| Arai at al (2016) | -1.38 | 3.67 | 17 | -2.90 | 1.30 | 7 | 0.46(-0.43, 1.35) | 0.2 |
| Delnomdedieu at al (2016) | -0.24 | 3.39 | 72 | 0.50 | 3.06 | 17 | -0.22(-0.75, 0.31) | 0.5 |
| **Subtotal**($I^2$ = 0%, p = 0.51) | | | **1398** | | | **959** | **0.00(-0.08, 0.09)** | **21.1** |
| **Solanezumab** | | | | | | | | |
| Doody at al (2014) | -1.40 | 7.83 | 370 | -2.00 | 7.83 | 370 | 0.08(-0.07, 0.22) | 7.0 |
| Doody at al (2014) | -2.10 | 8.20 | 406 | -2.80 | 8.14 | 400 | 0.09(-0.05, 0.22) | 7.7 |
| Honig at al (2018) | -3.17 | 4.52 | 908 | -3.66 | 4.78 | 893 | 0.11( 0.01, 0.20) | 16.9 |
| **Subtotal**($I^2$ = 0%, p = 0.94) | | | **1684** | | | **1663** | **0.09( 0.03, 0.16)** | **31.6** |
| **Crenezumab** | | | | | | | | |
| Ostrowitzki at al (2022) | -4.96 | 3.57 | 87 | -4.63 | 3.53 | 90 | -0.09(-0.39, 0.20) | 1.7 |
| Ostrowitzki at al (2022) | -3.21 | 2.38 | 12 | -3.63 | 2.46 | 15 | 0.17(-0.59, 0.93) | 0.3 |
| **Subtotal**($I^2$ = 0%, p = 0.53) | | | **99** | | | **105** | **-0.06(-0.33, 0.22)** | **2.0** |
| **Gantenerumab** | | | | | | | | |
| Ostrowitzki at al (2017) | -2.88 | 3.01 | 205 | -2.93 | 2.95 | 105 | 0.02(-0.22, 0.25) | 2.7 |
| **Subtotal** | | | **205** | | | **105** | **0.02(-0.22, 0.25)** | **2.7** |
| **Ponezumab** | | | | | | | | |
| Landen at al (2017) | -1.42 | 2.80 | 24 | -2.88 | 4.50 | 24 | 0.38(-0.19, 0.95) | 0.5 |
| **Subtotal** | | | **24** | | | **24** | **0.38(-0.19, 0.95)** | **0.5** |
| **N-terminal-targeted** | | | **4383** | | | **2853** | **0.05(-0.00, 0.10)** | **63.3** |
| **Central-domain-targeted** | | | **1783** | | | **1768** | **0.09( 0.02, 0.15)** | **33.6** |
| **N-terminal + central-domain-targeted** | | | **205** | | | **105** | **0.02(-0.22, 0.25)** | **2.7** |
| **C-terminal-targeted** | | | **24** | | | **24** | **0.38(-0.19, 0.95)** | **0.5** |
| **Total (95% CI)** | | | **6395** | | | **4750** | **0.06( 0.02, 0.10)** | **100.0** |

**Heterogeneity: $I^2$ = 0%, p = 0.62**

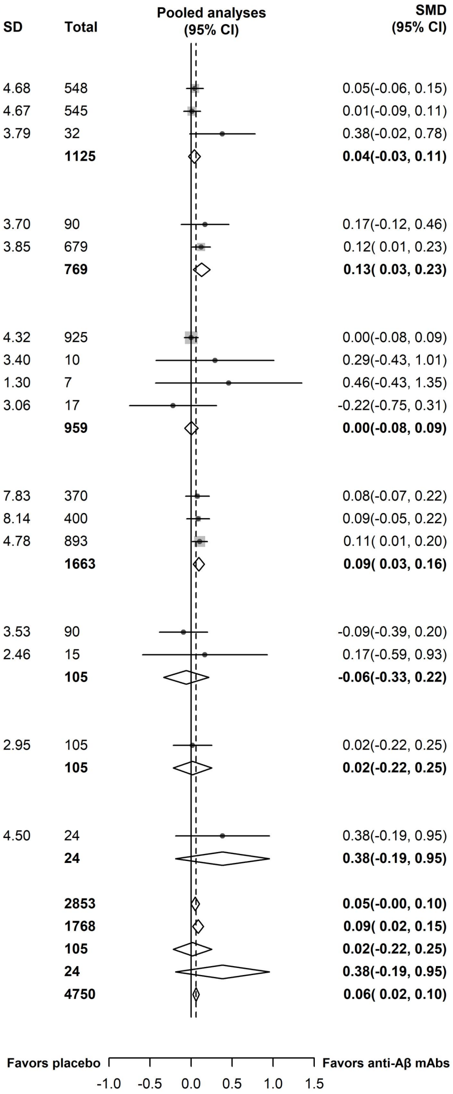

### Appendix 7: Forest plots for the secondary outcomes

#### 7.1 CSF Aβ40

| Study | Anti-Aβ mAbs Mean | SD | Total | Placebo Mean | SD | Total | SMD (95% CI) | Weight (%) |
|---|---|---|---|---|---|---|---|---|
| **Lecanemab** | | | | | | | | |
| Dyck at al (2022) | -437.56 | 2885.20 | 71 | -95.15 | 2890.44 | 71 | -0.12(-0.45, 0.21) | 15.3 |
| **Subtotal** | | | **71** | | | **71** | **-0.12(-0.45, 0.21)** | **15.3** |
| **Bapineuzumab** | | | | | | | | |
| Blennow at al (2012) | -188.50 | 1803.38 | 26 | -700.40 | 1891.55 | 19 | 0.27(-0.32, 0.87) | 12.7 |
| **Subtotal** | | | **26** | | | **19** | **0.27(-0.32, 0.87)** | **12.7** |
| **Solanezumab** | | | | | | | | |
| Doody at al (2014) | 1325.40 | 9588.17 | 20 | -1902.10 | 11526.73 | 25 | 0.30(-0.30, 0.89) | 12.8 |
| Doody at al (2014) | 2156.80 | 11080.24 | 44 | -876.40 | 9613.68 | 32 | 0.29(-0.17, 0.74) | 14.1 |
| Honig at al (2018) | 3313.35 | 5108.77 | 127 | -1231.57 | 4693.37 | 121 | 0.92( 0.66, 1.18) | 15.8 |
| **Subtotal**($I^2$ = 74%, p = 0.02) | | | **191** | | | **178** | **0.55( 0.10, 1.01)** | **42.6** |
| **Crenezumab** | | | | | | | | |
| Ostrowitzki at al (2022) | 4917.10 | 2248.40 | 17 | -355.00 | 2846.10 | 22 | 1.98( 1.20, 2.77) | 10.8 |
| **Subtotal** | | | **17** | | | **22** | **1.98( 1.20, 2.77)** | **10.8** |
| **Ponezumab** | | | | | | | | |
| Landen at al (2017) | -4830.63 | 5034.70 | 21 | -6332.71 | 5395.40 | 10 | 0.28(-0.47, 1.04) | 11.1 |
| Landen at al (2013) | 26.39 | 35.55 | 8 | -1.23 | 29.86 | 5 | 0.76(-0.41, 1.93) | 7.5 |
| **Subtotal**($I^2$ = 0%, p = 0.50) | | | **29** | | | **15** | **0.43(-0.21, 1.06)** | **18.6** |
| **N-terminal-targeted** | | | **97** | | | **90** | **-0.00(-0.35, 0.34)** | **28.0** |
| **Central-domain-targeted** | | | **208** | | | **200** | **0.83( 0.12, 1.55)** | **53.4** |
| **C-terminal-targeted** | | | **29** | | | **15** | **0.43(-0.21, 1.06)** | **18.6** |
| **Total (95% CI)** | | | **334** | | | **305** | **0.54( 0.11, 0.98)** | **100.0** |

**Heterogeneity: $I^2$ = 82%, p < 0.01**

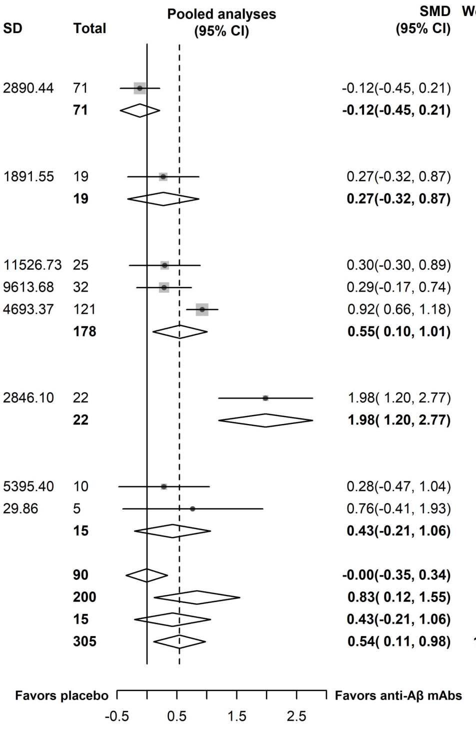

#### 7.2 CSF Aβ42

| Study | Anti-Aβ mAbs Mean | SD | Total | Placebo Mean | SD | Total | SMD (95% CI) | Weight (%) |
|---|---|---|---|---|---|---|---|---|
| **Aducanumab** | | | | | | | | |
| Haeberlein at al (2022) | 193.06 | 206.10 | 50 | -31.44 | 128.39 | 28 | 1.22( 0.72, 1.72) | 7.6 |
| Haeberlein at al (2022) | 147.34 | 167.11 | 36 | 5.12 | 153.03 | 15 | 0.86( 0.23, 1.48) | 7.0 |
| **Subtotal**($I^2$ = 0%, p = 0.38) | | | **86** | | | **43** | **1.08( 0.69, 1.47)** | **14.6** |
| **Lecanemab** | | | | | | | | |
| Swanson at al (2021) | 242.92 | 175.39 | 21 | -2.87 | 166.30 | 19 | 1.41( 0.71, 2.11) | 6.6 |
| Dyck at al (2022) | 282.11 | 287.79 | 101 | -6.38 | 270.86 | 97 | 1.03( 0.73, 1.32) | 8.5 |
| **Subtotal**($I^2$ = 0%, p = 0.33) | | | **122** | | | **116** | **1.09( 0.81, 1.36)** | **15.1** |
| **Bapineuzumab** | | | | | | | | |
| Blennow at al (2012) | -10.50 | 51.06 | 27 | -6.10 | 71.79 | 19 | -0.07(-0.66, 0.52) | 7.2 |
| **Subtotal** | | | **27** | | | **19** | **-0.07(-0.66, 0.52)** | **7.2** |
| **Solanezumab** | | | | | | | | |
| Doody at al (2014) | 471.40 | 1938.83 | 20 | -242.30 | 2185.43 | 25 | 0.34(-0.26, 0.93) | 7.1 |
| Doody at al (2014) | 726.60 | 780.19 | 44 | 323.80 | 658.93 | 32 | 0.54( 0.08, 1.01) | 7.8 |
| Honig at al (2018) | 462.61 | 313.61 | 131 | -65.59 | 305.11 | 124 | 1.70( 1.41, 1.99) | 8.6 |
| **Subtotal**($I^2$ = 93%, p < 0.01) | | | **195** | | | **181** | **0.89( 0.04, 1.74)** | **23.5** |
| **Crenezumab** | | | | | | | | |
| Cummings at al (2018) | 109.55 | 132.68 | 35 | -37.42 | 125.87 | 14 | 1.11( 0.45, 1.77) | 6.8 |
| Salloway at al (2018) | -36.88 | 96.82 | 37 | -67.46 | 87.72 | 18 | 0.32(-0.25, 0.89) | 7.3 |
| Ostrowitzki at al (2022) | 232.38 | 103.83 | 17 | -23.40 | 149.60 | 23 | 1.90( 1.13, 2.66) | 6.2 |
| **Subtotal**($I^2$ = 82%, p < 0.01) | | | **89** | | | **55** | **1.08( 0.19, 1.97)** | **20.3** |
| **Gantenerumab** | | | | | | | | |
| Ostrowitzki at al (2017) | 16.00 | 62.02 | 137 | -2.83 | 44.34 | 72 | 0.33( 0.04, 0.62) | 8.6 |
| **Subtotal** | | | **137** | | | **72** | **0.33( 0.04, 0.62)** | **8.6** |
| **Ponezumab** | | | | | | | | |
| Landen at al (2017) | -305.99 | 433.95 | 18 | -166.07 | 334.75 | 10 | -0.34(-1.12, 0.44) | 6.2 |
| Landen at al (2013) | 9.48 | 8.60 | 8 | 7.11 | 3.92 | 5 | 0.30(-0.82, 1.43) | 4.6 |
| **Subtotal**($I^2$ = 0%, p = 0.36) | | | **26** | | | **15** | **-0.13(-0.77, 0.51)** | **10.7** |
| **N-terminal-targeted** | | | **235** | | | **178** | **0.89( 0.41, 1.37)** | **36.9** |
| **Central-domain-targeted** | | | **284** | | | **236** | **0.98( 0.43, 1.53)** | **43.8** |
| **N-terminal + central-domain-targeted** | | | **137** | | | **72** | **0.33( 0.04, 0.62)** | **8.6** |
| **C-terminal-targeted** | | | **26** | | | **15** | **-0.13(-0.77, 0.51)** | **10.7** |
| **Total (95% CI)** | | | **682** | | | **501** | **0.78( 0.44, 1.12)** | **100.0** |

**Heterogeneity: $I^2$ = 85%, p < 0.01**

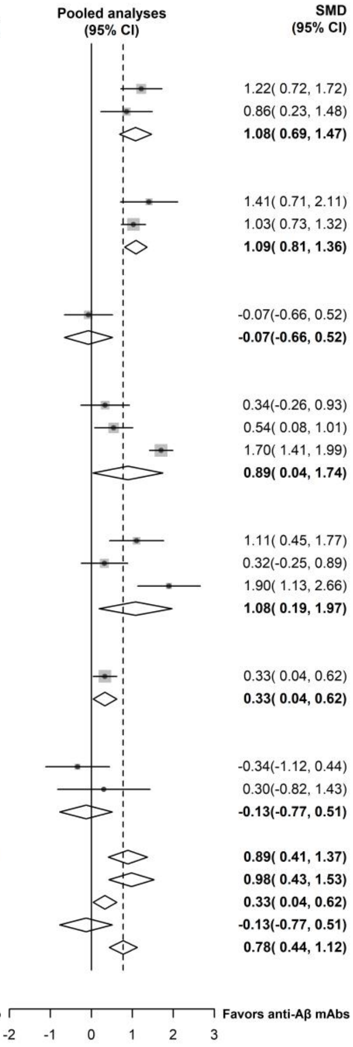

### 7.3 CSF p-tau

| Study | Anti-Aβ mAbs Mean | SD | Total | Placebo Mean | SD | Total | SMD (95% CI) | Weight (%) |
|---|---|---|---|---|---|---|---|---|
| **Aducanumab** | | | | | | | | |
| Haeberlein at al (2022) | -18.59 | 20.21 | 50 | -0.51 | 21.77 | 28 | -0.86(-1.34,-0.38) | 8.7 |
| Haeberlein at al (2022) | -13.43 | 29.94 | 38 | -2.49 | 29.81 | 15 | -0.36(-0.96, 0.24) | 7.9 |
| **Subtotal**($I^2$ = 38%, p = 0.20) | | | **88** | | | **43** | **-0.64(-1.13,-0.16)** | **16.6** |
| **Lecanemab** | | | | | | | | |
| Swanson at al (2021) | -12.40 | 23.19 | 23 | -0.05 | 21.92 | 18 | -0.53(-1.16, 0.09) | 7.8 |
| Dyck at al (2022) | -15.88 | 26.63 | 101 | 15.35 | 29.10 | 98 | -1.12(-1.42,-0.82) | 9.7 |
| **Subtotal**($I^2$ = 63%, p = 0.10) | | | **124** | | | **116** | **-0.89(-1.45,-0.34)** | **17.5** |
| **Bapineuzumab** | | | | | | | | |
| Blennow at al (2012) | -9.90 | 15.93 | 27 | -2.60 | 14.52 | 19 | -0.47(-1.06, 0.13) | 8.0 |
| Vandenberghe at al (2016) | -6.45 | 17.26 | 43 | 0.66 | 18.43 | 33 | -0.40(-0.85, 0.06) | 8.8 |
| Vandenberghe at al (2016) | -0.72 | 14.89 | 76 | 0.83 | 15.90 | 62 | -0.10(-0.44, 0.23) | 9.5 |
| Salloway at al (2014) | -5.26 | 13.48 | 101 | -1.98 | 12.97 | 77 | -0.25(-0.54, 0.05) | 9.7 |
| Salloway at al (2014) | -5.80 | 16.81 | 127 | 0.95 | 7.63 | 85 | -0.48(-0.76,-0.21) | 9.8 |
| Delnomdedieu at al (2016) | 1.23 | 8.99 | 15 | 1.92 | 7.76 | 5 | -0.08(-1.09, 0.94) | 5.5 |
| **Subtotal**($I^2$ = 0%, p = 0.58) | | | **389** | | | **281** | **-0.32(-0.48,-0.15)** | **51.4** |
| **Gantenerumab** | | | | | | | | |
| Ostrowitzki at al (2017) | -4.52 | 2.26 | 137 | 0.01 | 2.18 | 72 | -2.02(-2.37,-1.68) | 9.5 |
| **Subtotal** | | | **137** | | | **72** | **-2.02(-2.37,-1.68)** | **9.5** |
| **Ponezumab** | | | | | | | | |
| Landen at al (2013) | -0.43 | 25.20 | 8 | -6.29 | 15.94 | 5 | 0.24(-0.88, 1.37) | 5.0 |
| **Subtotal** | | | **8** | | | **5** | **0.24(-0.88, 1.37)** | **5.0** |
| **N-terminal-targeted** | | | **601** | | | **440** | **-0.49(-0.72,-0.26)** | **85.6** |
| **N-terminal + central-domain-targeted** | | | **137** | | | **72** | **-2.02(-2.37,-1.68)** | **9.5** |
| **C-terminal-targeted** | | | **8** | | | **5** | **0.24(-0.88, 1.37)** | **5.0** |
| **Total (95% CI)** | | | **746** | | | **517** | **-0.59(-0.94,-0.25)** | **100.0** |

**Heterogeneity: $I^2$ = 88%, p < 0.01**

**Pooled analyses (95% CI)**

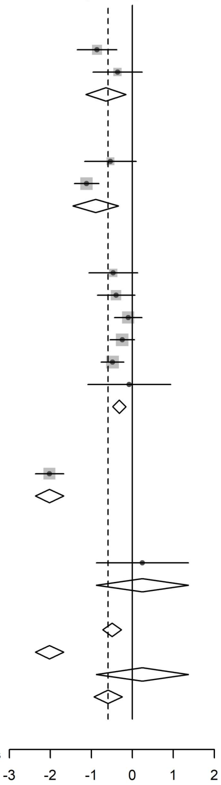

**Favors anti-Aβ mAbs** **Favors placebo**

#### 7.4 CSF total tau

| Study | Anti-Aβ mAbs Mean | SD | Total | Placebo Mean | SD | Total | SMD (95% CI) | Weight (%) |
|---|---|---|---|---|---|---|---|---|
| **Aducanumab** | | | | | | | | |
| Haeberlein at al (2022) | -96.29 | 139.50 | 50 | -2.39 | 142.48 | 28 | -0.66(-1.14,-0.19) | 10.9 |
| Haeberlein at al (2022) | -72.41 | 189.59 | 34 | -33.45 | 184.40 | 14 | -0.20(-0.83, 0.42) | 8.9 |
| **Subtotal**($I^2$ = 24%, p = 0.25) | | | **84** | | | **42** | **-0.48(-0.92,-0.04)** | **19.8** |
| **Lecanemab** | | | | | | | | |
| Swanson at al (2021) | -18.65 | 142.03 | 14 | -37.94 | 150.38 | 14 | 0.13(-0.61, 0.87) | 7.5 |
| Dyck at al (2022) | -31.35 | 242.81 | 101 | 95.18 | 209.17 | 98 | -0.56(-0.84,-0.27) | 13.5 |
| **Subtotal**($I^2$ = 65%, p = 0.09) | | | **115** | | | **112** | **-0.30(-0.95, 0.34)** | **21.1** |
| **Bapineuzumab** | | | | | | | | |
| Blennow at al (2012) | -72.30 | 175.94 | 27 | -5.60 | 123.86 | 19 | -0.42(-1.01, 0.18) | 9.3 |
| Delnomdedieu at al (2016) | -15.26 | 110.23 | 15 | 67.94 | 63.51 | 5 | -0.78(-1.83, 0.26) | 5.0 |
| **Subtotal**($I^2$ = 0%, p = 0.55) | | | **42** | | | **24** | **-0.51(-1.02, 0.01)** | **14.2** |
| **Crenezumab** | | | | | | | | |
| Cummings at al (2018) | 33.49 | 151.27 | 37 | 133.55 | 154.45 | 14 | -0.65(-1.28,-0.02) | 8.8 |
| Salloway at al (2018) | 10.77 | 180.19 | 37 | -31.20 | 170.14 | 17 | 0.23(-0.34, 0.81) | 9.5 |
| Ostrowitzki at al (2022) | -14.92 | 186.08 | 17 | 26.38 | 198.26 | 23 | -0.21(-0.84, 0.42) | 8.8 |
| **Subtotal**($I^2$ = 51%, p = 0.13) | | | **91** | | | **54** | **-0.20(-0.70, 0.31)** | **27.2** |
| **Gantenerumab** | | | | | | | | |
| Ostrowitzki at al (2017) | -10.78 | 18.36 | 137 | 11.35 | 18.47 | 72 | -1.20(-1.51,-0.89) | 13.2 |
| **Subtotal** | | | **137** | | | **72** | **-1.20(-1.51,-0.89)** | **13.2** |
| **Ponezumab** | | | | | | | | |
| Landen at al (2013) | 2.77 | 33.13 | 8 | 0.56 | 34.13 | 5 | 0.06(-1.06, 1.18) | 4.5 |
| **Subtotal** | | | **8** | | | **5** | **0.06(-1.06, 1.18)** | **4.5** |
| **N-terminal-targeted** | | | **241** | | | **178** | **-0.48(-0.68,-0.28)** | **55.1** |
| **Central-domain-targeted** | | | **91** | | | **54** | **-0.20(-0.70, 0.31)** | **27.2** |
| **N-terminal + central-domain-targeted** | | | **137** | | | **72** | **-1.20(-1.51,-0.89)** | **13.2** |
| **C-terminal-targeted** | | | **8** | | | **5** | **0.06(-1.06, 1.18)** | **4.5** |
| **Total (95% CI)** | | | **477** | | | **309** | **-0.44(-0.73,-0.16)** | **100.0** |

**Heterogeneity: $I^2$ = 68%, p < 0.01**

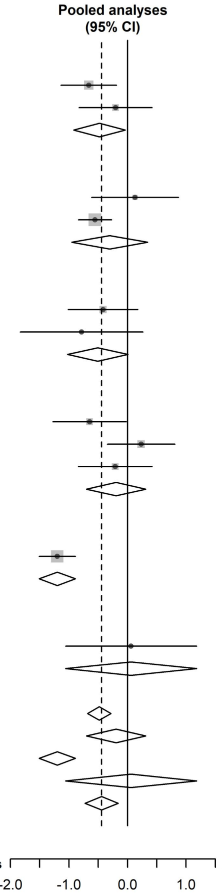

#### 7.5 CSF p-tau181

| Study | Anti-Aβ mAbs Mean | SD | Total | Placebo Mean | SD | Total | SMD (95% CI) | Weight (%) |
|---|---|---|---|---|---|---|---|---|
| **Lecanemab** | | | | | | | | |
| Dyck at al (2022) | -16.14 | 30.68 | 101 | 12.51 | 28.19 | 98 | -0.97(-1.26,-0.67) | 30.0 |
| **Subtotal** | | | **101** | | | **98** | **-0.97(-1.26,-0.67)** | **30.0** |
| **Crenezumab** | | | | | | | | |
| Cummings at al (2018) | 0.30 | 15.07 | 37 | 8.48 | 16.00 | 14 | -0.53(-1.15, 0.10) | 23.0 |
| Salloway at al (2018) | -2.46 | 13.95 | 37 | -6.33 | 13.06 | 17 | 0.28(-0.30, 0.86) | 24.1 |
| Ostrowitzki at al (2022) | -2.09 | 15.30 | 17 | 1.60 | 18.85 | 23 | -0.21(-0.84, 0.42) | 22.9 |
| **Subtotal**($I^2$ = 43%, p = 0.17) | | | **91** | | | **54** | **-0.14(-0.61, 0.33)** | **70.0** |
| **N-terminal-targeted** | | | **101** | | | **98** | **-0.97(-1.26,-0.67)** | **30.0** |
| **Central-domain-targeted** | | | **91** | | | **54** | **-0.14(-0.61, 0.33)** | **70.0** |
| **Total (95% CI)** | | | **192** | | | **152** | **-0.39(-0.94, 0.16)** | **100.0** |

**Heterogeneity: $I^2$ = 82%, p < 0.01**

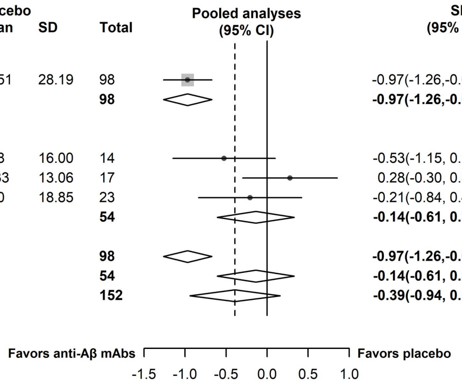

### 7.6 CSF Nfl

| Study | Anti-Aβ mAbs Mean | SD | Total | Placebo Mean | SD | Total | SMD (95% CI) | Weight (%) |
|---|---|---|---|---|---|---|---|---|
| **Lecanemab** | | | | | | | | |
| Swanson at al (2021) | 9.77 | 46.83 | 23 | 69.35 | 247.69 | 16 | -0.36(-1.00, 0.28) | 15.2 |
| Dyck at al (2022) | 51.73 | 756.62 | 104 | 79.16 | 718.58 | 97 | -0.04(-0.31, 0.24) | 68.3 |
| **Subtotal**($I^2$ = 0%, p = 0.37) | | | **127** | | | **113** | **-0.09(-0.34, 0.17)** | **83.5** |
| **Gantenerumab** | | | | | | | | |
| Ostrowitzki at al (2017) | -32.06 | 277.28 | 18 | 46.63 | 92.41 | 24 | -0.40(-1.02, 0.22) | 16.5 |
| **Subtotal** | | | **18** | | | **24** | **-0.40(-1.02, 0.22)** | **16.5** |
| **N-terminal-targeted** | | | **127** | | | **113** | **-0.09(-0.34, 0.17)** | **83.5** |
| **N-terminal + central-domain-targeted** | | | **18** | | | **24** | **-0.40(-1.02, 0.22)** | **16.5** |
| **Total (95% CI)** | | | **145** | | | **137** | **-0.15(-0.40, 0.11)** | **100.0** |

**Heterogeneity: $I^2$ = 0%, p = 0.44**

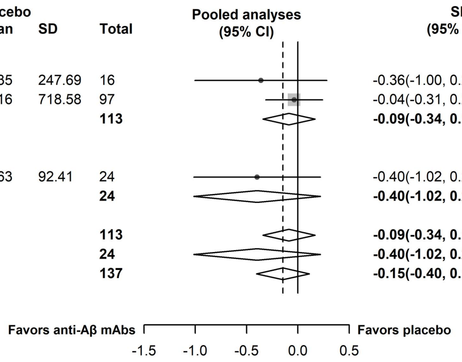

### 7.7 CSF Neurogranin

| Study | Anti-Aβ mAbs Mean | SD | Total | Placebo Mean | SD | Total | SMD (95% CI) | Weight (%) |
|---|---|---|---|---|---|---|---|---|
| **Lecanemab** | | | | | | | | |
| Swanson at al (2021) | -37.05 | 129.96 | 23 | 6.80 | 115.58 | 16 | -0.35(-0.99, 0.30) | 8.5 |
| Dyck at al (2022) | -71.67 | 180.38 | 104 | 18.31 | 170.42 | 97 | -0.51(-0.79,-0.23) | 44.6 |
| **Subtotal**($I^2$ = 0%, p = 0.65) | | | **127** | | | **113** | **-0.48(-0.74,-0.23)** | **53.1** |
| **Crenezumab** | | | | | | | | |
| Ostrowitzki at al (2022) | -84.50 | 143.30 | 18 | 67.10 | 141.80 | 24 | -1.04(-1.70,-0.39) | 8.2 |
| **Subtotal** | | | **18** | | | **24** | **-1.04(-1.70,-0.39)** | **8.2** |
| **Gantenerumab** | | | | | | | | |
| Ostrowitzki at al (2017) | -36.47 | 44.16 | 129 | -16.22 | 41.19 | 65 | -0.47(-0.77,-0.17) | 38.7 |
| **Subtotal** | | | **129** | | | **65** | **-0.47(-0.77,-0.17)** | **38.7** |
| **N-terminal-targeted** | | | **127** | | | **113** | **-0.48(-0.74,-0.23)** | **53.1** |
| **Central-domain-targeted** | | | **18** | | | **24** | **-1.04(-1.70,-0.39)** | **8.2** |
| **N-terminal + central-domain-targeted** | | | **129** | | | **65** | **-0.47(-0.77,-0.17)** | **38.7** |
| **Total (95% CI)** | | | **274** | | | **202** | **-0.52(-0.71,-0.34)** | **100.0** |

**Heterogeneity: $I^2$ = 0%, p = 0.41**

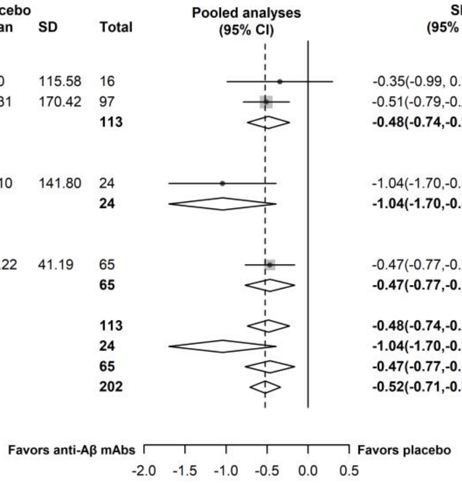

#### 7.8 Plasma Aβ40

| Study | Anti-Aβ mAbs Mean | SD | Total | Placebo Mean | SD | Total | SMD (95% CI) | Weight (%) |
|---|---|---|---|---|---|---|---|---|
| **Donanemab** | | | | | | | | |
| Mintun at al (2021) | 18.07 | 40.32 | 83 | 14.58 | 41.01 | 78 | 0.09(-0.22, 0.39) | 33.3 |
| **Subtotal** | | | **83** | | | **78** | **0.09(-0.22, 0.39)** | **33.3** |
| **Solanezumab** | | | | | | | | |
| Honig at al (2018) | 172528.4[illegible] | [illegible]5098.72 | 841 | 836.19 | 4585.48 | 842 | 3.72( 3.56, 3.88) | 33.5 |
| **Subtotal** | | | **841** | | | **842** | **3.72( 3.56, 3.88)** | **33.5** |
| **Ponezumab** | | | | | | | | |
| Landen at al (2017) | 9372.45 | 10828.00 | 101 | 163.75 | 133.22 | 43 | 1.01( 0.63, 1.38) | 33.2 |
| **Subtotal** | | | **101** | | | **43** | **1.01( 0.63, 1.38)** | **33.2** |
| **N-terminal-targeted** | | | **83** | | | **78** | **0.09(-0.22, 0.39)** | **33.3** |
| **Central-domain-targeted** | | | **841** | | | **842** | **3.72( 3.56, 3.88)** | **33.5** |
| **C-terminal-targeted** | | | **101** | | | **43** | **1.01( 0.63, 1.38)** | **33.2** |
| **Total (95% CI)** | | | **1025** | | | **963** | **1.61(-0.53, 3.75)** | **100.0** |

**Heterogeneity: $I^2$ = 100%, $p < 0.01$**

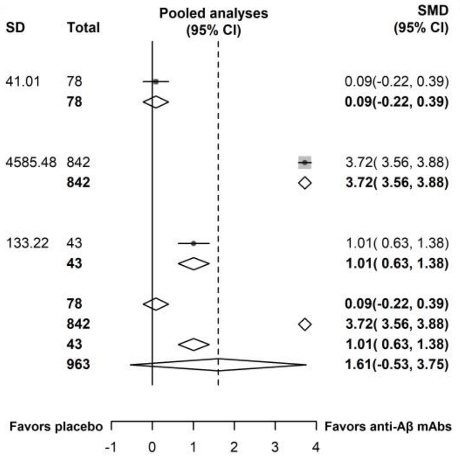

### 7.9 Plasma Nfl

| Study | Anti-Aβ mAbs Mean | SD | Total | Placebo Mean | SD | Total | SMD (95% CI) | Weight (%) |
|---|---|---|---|---|---|---|---|---|
| **Lecanemab** | | | | | | | | |
| Dyck at al (2022) | 1.82 | 10.73 | 529 | 2.94 | 10.62 | 574 | -0.10(-0.22, 0.01) | 73.8 |
| **Subtotal** | | | **529** | | | **574** | **-0.10(-0.22, 0.01)** | **73.8** |
| **Donanemab** | | | | | | | | |
| Mintun at al (2021) | 1.95 | 10.75 | 69 | -0.77 | 23.15 | 57 | 0.15(-0.20, 0.51) | 11.0 |
| **Subtotal** | | | **69** | | | **57** | **0.15(-0.20, 0.51)** | **11.0** |
| **Crenezumab** | | | | | | | | |
| Ostrowitzki at al (2022) | 0.74 | 2.20 | 91 | 0.78 | 1.82 | 85 | -0.02(-0.32, 0.28) | 15.2 |
| **Subtotal** | | | **91** | | | **85** | **-0.02(-0.32, 0.28)** | **15.2** |
| **N-terminal-targeted** | | | **598** | | | **631** | **-0.03(-0.26, 0.20)** | **84.8** |
| **Central-domain-targeted** | | | **91** | | | **85** | **-0.02(-0.32, 0.28)** | **15.2** |
| **Total (95% CI)** | | | **689** | | | **716** | **-0.06(-0.18, 0.06)** | **100.0** |

**Heterogeneity: $I^2$ = 0%, p = 0.37**

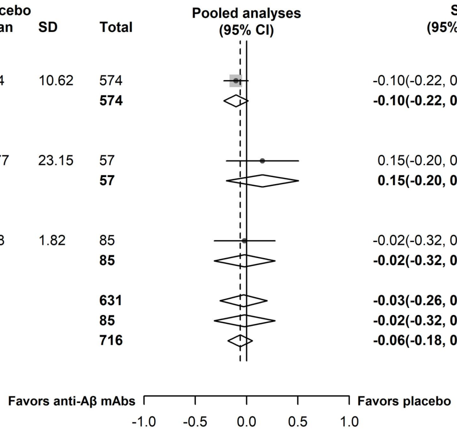

### 7.10 Plasma p-tau181

| Study | Anti-Aβ mAbs Mean | SD | Total | Placebo Mean | SD | Total | SMD (95% CI) | Weight (%) |
|---|---|---|---|---|---|---|---|---|
| **Aducanumab** | | | | | | | | |
| Haeberlein at al (2022) | -0.39 | 1.19 | 540 | 0.24 | 1.20 | 273 | -0.53(-0.68,-0.38) | 28.7 |
| Haeberlein at al (2022) | -0.40 | 1.03 | 647 | 0.29 | 0.89 | 271 | -0.70(-0.85,-0.56) | 29.5 |
| **Subtotal**($I^2$ = 62%, p = 0.10) | | | **1187** | | | **544** | **-0.62(-0.79,-0.45)** | **58.2** |
| **Lecanemab** | | | | | | | | |
| Dyck at al (2022) | -0.58 | 1.30 | 590 | 0.20 | 1.14 | 609 | -0.64(-0.76,-0.53) | 41.8 |
| **Subtotal** | | | **590** | | | **609** | **-0.64(-0.76,-0.53)** | **41.8** |
| **N-terminal-targeted** | | | **1777** | | | **1153** | **-0.63(-0.72,-0.54)** | **100.0** |
| **Total (95% CI)** | | | **1777** | | | **1153** | **-0.63(-0.72,-0.54)** | **100.0** |

**Heterogeneity: $I^2$ = 27%, p = 0.25**

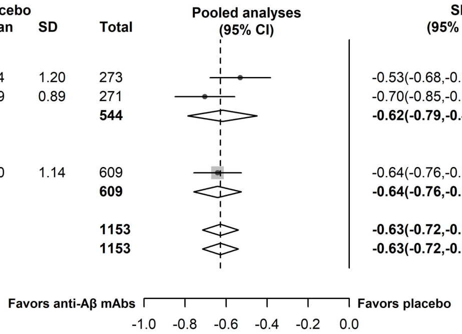

### 7.11 Tau PET

| Study | Anti-Aβ mAbs Mean | SD | Total | Placebo Mean | SD | Total | SMD (95% CI) | Weight (%) |
|---|---|---|---|---|---|---|---|---|
| **Aducanumab** | | | | | | | | |
| Haeberlein at al (2022) | -0.22 | 0.11 | 209 | 0.01 | 0.10 | 93 | -2.20(-2.50,-1.90) | 19.9 |
| Haeberlein at al (2022) | -0.20 | 0.10 | 250 | -0.00 | 0.11 | 124 | -1.95(-2.20,-1.69) | 20.0 |
| **Subtotal**($I^2$ = 36%, p = 0.21) | | | **459** | | | **217** | **-2.06(-2.30,-1.81)** | **39.8** |
| **Donanemab** | | | | | | | | |
| Mintun at al (2021) | 0.09 | 0.08 | 90 | 0.10 | 0.07 | 87 | -0.15(-0.44, 0.15) | 19.9 |
| Sims at al (2023) | 0.04 | 0.09 | 578 | 0.04 | 0.10 | 654 | -0.04(-0.16, 0.07) | 20.2 |
| **Subtotal**($I^2$ = 0%, p = 0.53) | | | **668** | | | **741** | **-0.06(-0.16, 0.05)** | **40.1** |
| **Solanezumab** | | | | | | | | |
| Sperling at al (2023) | 0.05 | 0.09 | 172 | 0.07 | 0.09 | 168 | -0.14(-0.35, 0.08) | 20.1 |
| **Subtotal** | | | **172** | | | **168** | **-0.14(-0.35, 0.08)** | **20.1** |
| **N-terminal-targeted** | | | **1127** | | | **958** | **-1.08(-2.20, 0.04)** | **79.9** |
| **Central-domain-targeted** | | | **172** | | | **168** | **-0.14(-0.35, 0.08)** | **20.1** |
| **Total (95% CI)** | | | **1299** | | | **1126** | **-0.89(-1.84, 0.05)** | **100.0** |

**Heterogeneity: $I^2$ = 99%, p < 0.01**

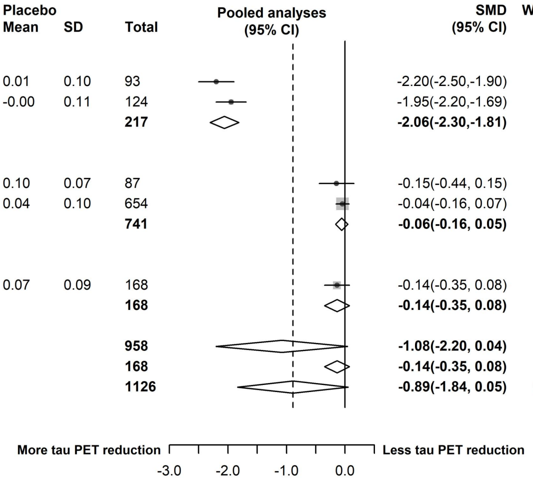

### 7.12 ARIA-E

| Study | Anti-Aβ mAbs Events | Anti-Aβ mAbs Total | Placebo Events | Placebo Total | RR (95% CI) | Weight (%) |
|---|---|---|---|---|---|---|
| **Aducanumab** | | | | | | |
| Haeberlein at al (2022) | 328.00 | 1091 | 13.00 | 547 | 12.65( 7.34, 21.81) | 15.7 |
| Haeberlein at al (2022) | 340.00 | 1107 | 16.00 | 540 | 10.37( 6.35, 16.93) | 19.0 |
| Jeff sevigny at al (2016) | 27.00 | 125 | 0.00 | 40 | 17.75( 1.11, 284.49) | 0.7 |
| **Subtotal**($I^2$ = 0%, p = 0.83) | | **2323** | | **1127** | **11.42( 7.96, 16.39)** | **35.3** |
| **Lecanemab** | | | | | | |
| Swanson at al (2021) | 46.00 | 609 | 2.00 | 245 | 9.25( 2.26, 37.82) | 2.5 |
| Dyck at al (2022) | 113.00 | 898 | 15.00 | 897 | 7.52( 4.43, 12.79) | 16.5 |
| Logovinsky at al (2016) | 0.00 | 72 | 0.00 | 24 | | 0.0 |
| **Subtotal**($I^2$ = 0%, p = 0.79) | | **1579** | | **1166** | **7.72( 4.70, 12.69)** | **19.0** |
| **Donanemab** | | | | | | |
| Mintun at al (2021) | 36.00 | 131 | 1.00 | 125 | 34.35( 4.78, 246.77) | 1.3 |
| Sims at al (2023) | 205.00 | 853 | 17.00 | 874 | 12.36( 7.60, 20.08) | 19.3 |
| Lowe at al (2021) | 10.00 | 46 | 0.00 | 15 | 7.00( 0.44, 112.63) | 0.7 |
| **Subtotal**($I^2$ = 0%, p = 0.56) | | **1030** | | **1014** | **12.87( 8.09, 20.49)** | **21.3** |
| **Bapineuzumab** | | | | | | |
| Vandenberghe at al (2016) | 44.00 | 530 | 2.00 | 344 | 14.28( 3.48, 58.52) | 2.5 |
| Vandenberghe at al (2016) | 109.00 | 654 | 9.00 | 439 | 8.13( 4.16, 15.87) | 10.7 |
| Salloway at al (2014) | 65.00 | 807 | 1.00 | 524 | 42.21( 5.87, 303.22) | 1.3 |
| Salloway at al (2014) | 103.00 | 673 | 1.00 | 448 | 68.56( 9.60, 489.61) | 1.3 |
| Delnomdedieu at al (2016) | 2.00 | 69 | 0.00 | 19 | 1.40( 0.07, 28.02) | 0.6 |
| **Subtotal**($I^2$ = 49%, p = 0.10) | | **2733** | | **1774** | **14.27( 5.66, 36.01)** | **16.3** |
| **Solanezumab** | | | | | | |
| Farlow at al (2012) | 0.00 | 42 | 0.00 | 10 | | 0.0 |
| Doody at al (2014） | 9.00 | 1027 | 4.00 | 1025 | 2.25( 0.69, 7.27) | 3.6 |
| Sperling at al (2023) | 1.00 | 572 | 2.00 | 591 | 0.52( 0.05, 5.68) | 0.9 |
| **Subtotal**($I^2$ = 14%, p = 0.28) | | **1641** | | **1626** | **1.59( 0.47, 5.39)** | **4.5** |
| **Crenezumab** | | | | | | |
| Salloway at al (2018) | 0.00 | 62 | 0.00 | 29 | | 0.0 |
| Ostrowitzki at al (2022) | 1.00 | 404 | 1.00 | 405 | 1.00( 0.06, 15.97) | 0.7 |
| Ostrowitzki at al (2022) | 1.00 | 404 | 0.00 | 398 | 2.96( 0.12, 72.33) | 0.5 |
| Guthrie at al (2020) | 0.00 | 61 | 0.00 | 14 | | 0.0 |
| **Subtotal**($I^2$ = 0%, p = 0.62) | | **931** | | **846** | **1.59( 0.20, 12.92)** | **1.1** |
| **Gantenerumab** | | | | | | |
| Ostrowitzki at al (2017) | 53.00 | 531 | 2.00 | 266 | 13.27( 3.26, 54.05) | 2.5 |
| **Subtotal** | | **531** | | **266** | **13.27( 3.26, 54.05)** | **2.5** |
| **N-terminal-targeted** | | **7665** | | **5081** | **10.79( 8.61, 13.52)** | **91.9** |
| **Central-domain-targeted** | | **2572** | | **2472** | **1.67( 0.65, 4.28)** | **5.6** |
| **N-terminal + central-domain-targeted** | | **531** | | **266** | **13.27( 3.26, 54.05)** | **2.5** |
| **Total (95% CI)** | | **10768** | | **7819** | **9.79( 7.83, 12.26)** | **100.0** |

**Heterogeneity: $I^2$ = 39%, p = 0.05**

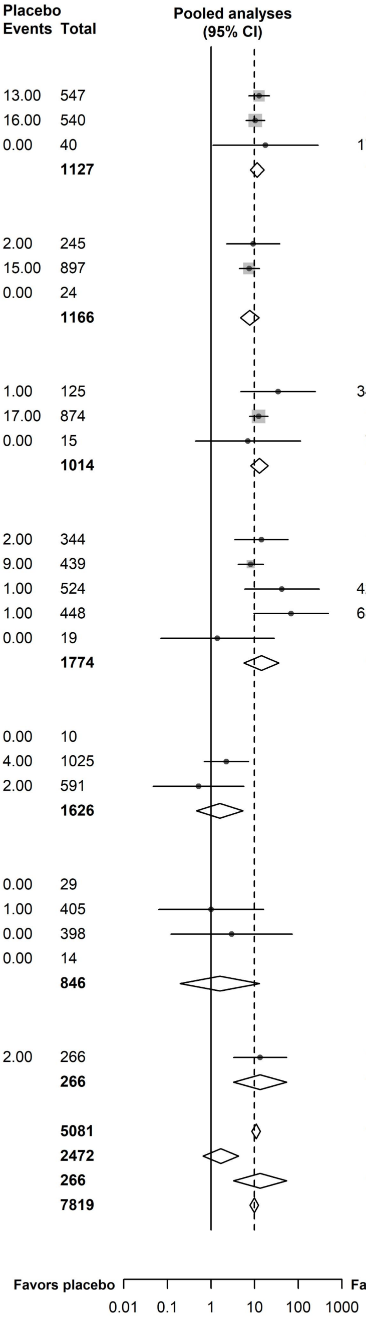

### 7.13 ARIA-H

| Study | Anti-Aβ mAbs Events | Anti-Aβ mAbs Total | Placebo Events | Placebo Total | RR (95% CI) | Weight (%) |
|---|---|---|---|---|---|---|
| **Aducanumab** | | | | | | |
| Jeff sevigny at al (2016) | 22.00 | 125 | 2.00 | 40 | 3.52( 0.87, 14.32) | 2.8 |
| **Subtotal** | | **125** | | **40** | **3.52( 0.87, 14.32)** | **2.8** |
| **Lecanemab** | | | | | | |
| Swanson at al (2021) | 65.00 | 609 | 13.00 | 245 | 2.01( 1.13, 3.58) | 7.8 |
| Dyck at al (2022) | 155.00 | 898 | 81.00 | 897 | 1.91( 1.49, 2.46) | 11.2 |
| Logovinsky at al (2016) | 3.00 | 72 | 2.00 | 14 | 0.29( 0.05, 1.59) | 2.1 |
| **Subtotal**($I^2$ = 57%, p = 0.10) | | **1579** | | **1156** | **1.86( 1.48, 2.34)** | **21.1** |
| **Donanemab** | | | | | | |
| Mintun at al (2021) | 11.00 | 131 | 4.00 | 125 | 2.62( 0.86, 8.02) | 3.9 |
| Sims at al (2023) | 155.00 | 853 | 81.00 | 874 | 1.96( 1.52, 2.52) | 11.2 |
| **Subtotal**($I^2$ = 0%, p = 0.62) | | **984** | | **999** | **1.99( 1.56, 2.54)** | **15.1** |
| **Bapineuzumab** | | | | | | |
| Salloway at al (2009) | 0.00 | 122 | 0.00 | 107 | | |
| Rinne at al (2010) | 0.00 | 19 | 0.00 | 7 | | |
| Delnomdedieu at al (2016) | 3.00 | 69 | 0.00 | 19 | 1.96( 0.11, 36.43) | 0.8 |
| **Subtotal** | | **210** | | **133** | **1.96( 0.11, 36.43)** | **0.8** |
| **Solanezumab** | | | | | | |
| Doody at al (2014) | 50.00 | 1027 | 57.00 | 1025 | 0.88( 0.60, 1.27) | 10.0 |
| Honig at al (2018) | 42.00 | 1054 | 46.00 | 1067 | 0.92( 0.61, 1.39) | 9.6 |
| Sperling at al (2023) | 167.00 | 572 | 194.00 | 591 | 0.89( 0.75, 1.06) | 11.8 |
| **Subtotal**($I^2$ = 0%, p = 0.98) | | **2653** | | **2683** | **0.89( 0.77, 1.03)** | **31.4** |
| **Crenezumab** | | | | | | |
| Ostrowitzki at al (2022) | 31.00 | 397 | 39.00 | 399 | 0.80( 0.51, 1.25) | 9.2 |
| Ostrowitzki at al (2022) | 65.00 | 609 | 13.00 | 245 | 2.01( 1.13, 3.58) | 7.8 |
| Guthrie at al (2020) | 3.00 | 61 | 0.00 | 14 | 1.65( 0.09, 30.22) | 0.8 |
| **Subtotal**($I^2$ = 68%, p = 0.05) | | **1067** | | **658** | **1.26( 0.57, 2.81)** | **17.8** |
| **Gantenerumab** | | | | | | |
| Ostrowitzki at al (2017) | 62.00 | 531 | 35.00 | 266 | 0.89( 0.60, 1.31) | 9.8 |
| **Subtotal** | | **531** | | **266** | **0.89( 0.60, 1.31)** | **9.8** |
| **Ponezumab** | | | | | | |
| Landen at al (2017) | 0.00 | 138 | 0.00 | 56 | | |
| Landen at al (2017) | 2.00 | 24 | 1.00 | 12 | 1.00( 0.10, 9.96) | 1.2 |
| **Subtotal** | | **162** | | **68** | **1.00( 0.10, 9.96)** | **1.2** |
| **N-terminal-targeted** | | **2898** | | **2328** | **1.94( 1.64, 2.29)** | **39.7** |
| **Central-domain-targeted** | | **3720** | | **3341** | **0.92( 0.81, 1.06)** | **49.2** |
| **N-terminal + central-domain-targeted** | | **531** | | **266** | **0.89( 0.60, 1.31)** | **9.8** |
| **C-terminal-targeted** | | **162** | | **68** | **1.00( 0.10, 9.96)** | **1.2** |
| **Total (95% CI)** | | **7311** | | **6003** | **1.28( 0.98, 1.67)** | **100.0** |

**Heterogeneity: $I^2$ = 78%, p < 0.01**

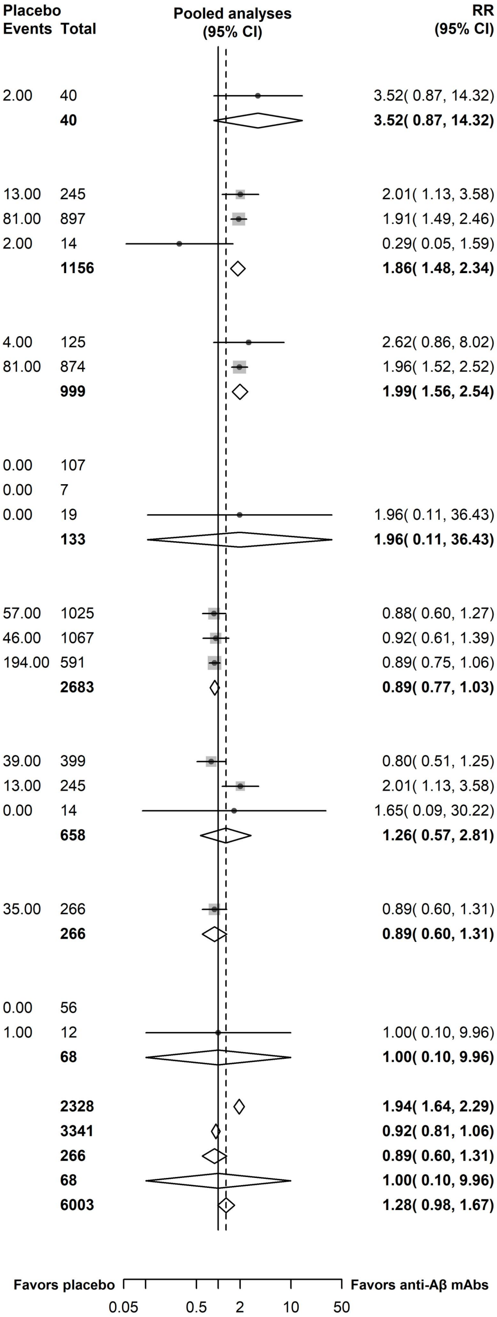

### Appendix 8: Meta-regressions

(baseline CDR-SB score, baseline MMSE score, the mean age, % APOE4 carrier, % female participants, % on anti-AD medication)

#### 8.1 ADAS-cog

##### 8.1.1 Meta-regressions of ADAS-Cog by baseline CDR-SB score

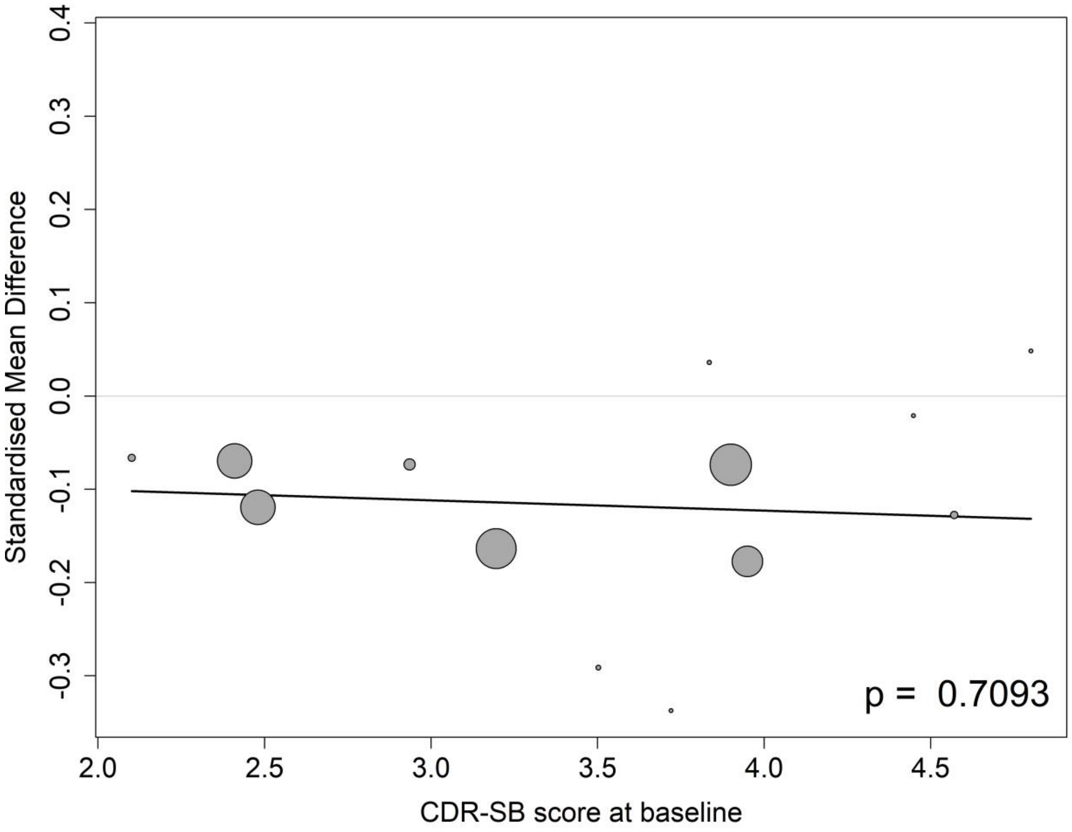

##### 8.1.2 Meta-regressions of ADAS-Cog by baseline MMSE score

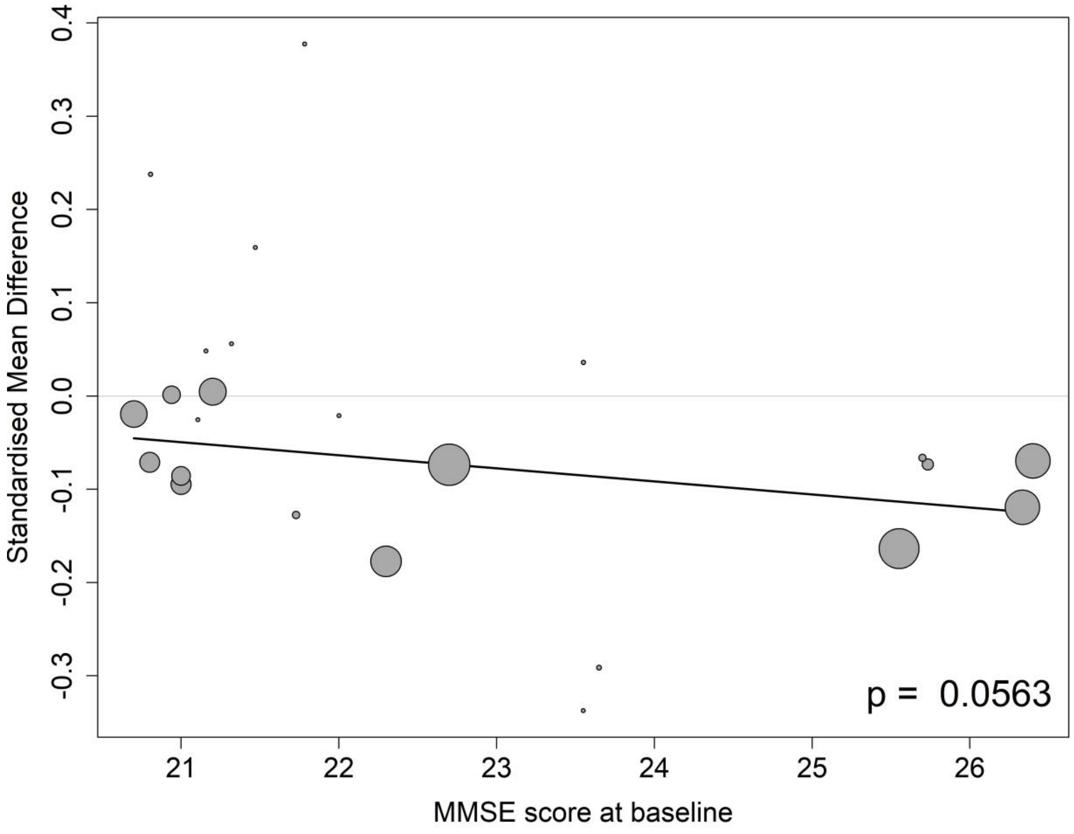

##### 8.1.3 Meta-regressions of ADAS-Cog by the mean age at the baseline

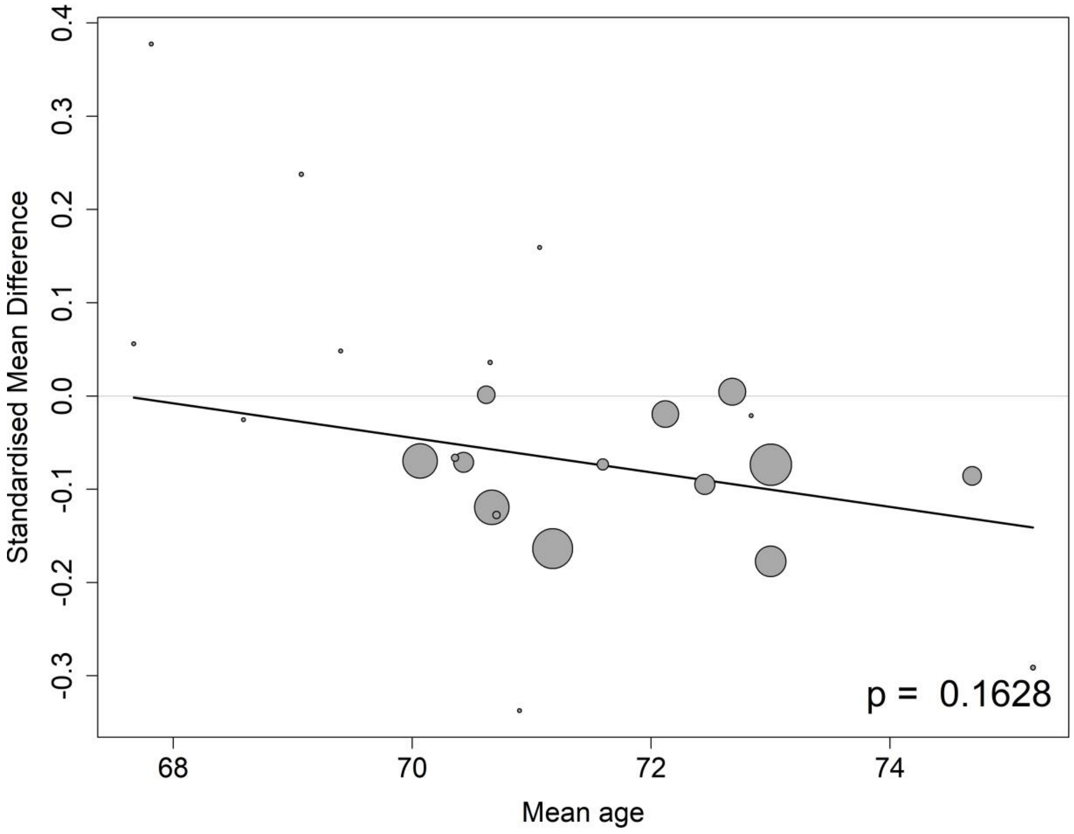

##### 8.1.4 Meta-regressions of ADAS-Cog by the percentage of APOE4 carrier at the baseline

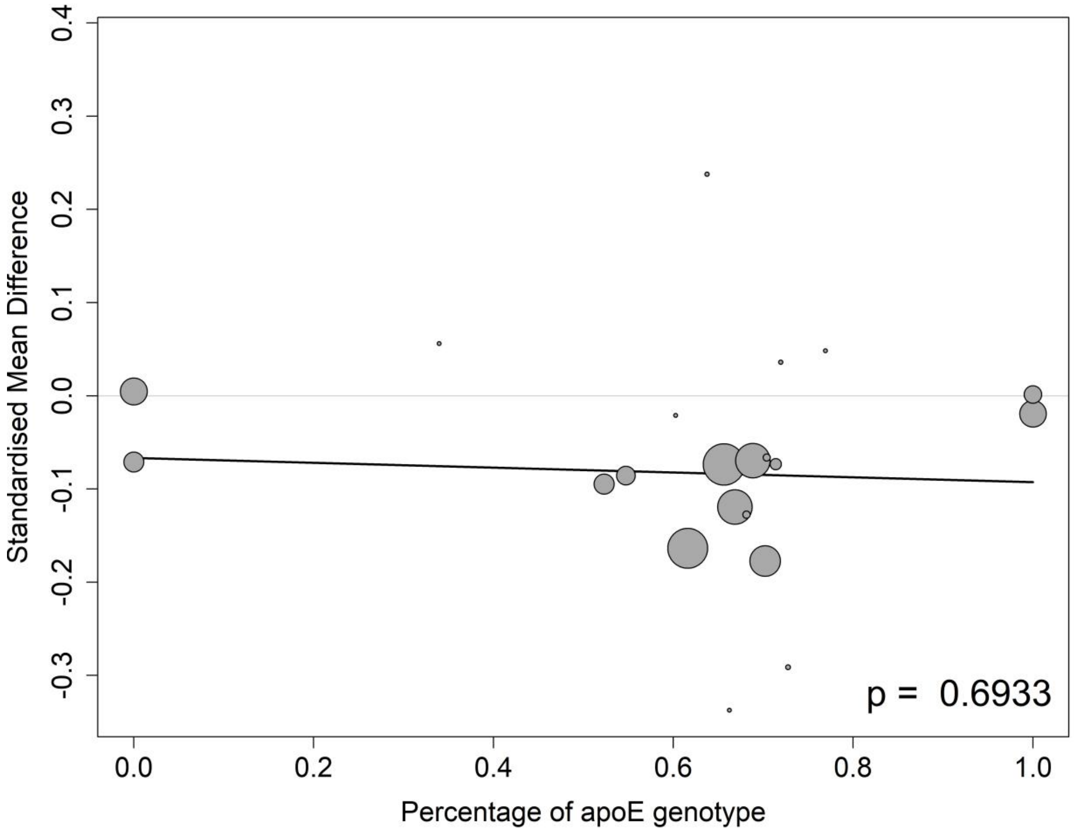

##### 8.1.5 Meta-regressions of ADAS-Cog by the percentage of female participants at the baseline

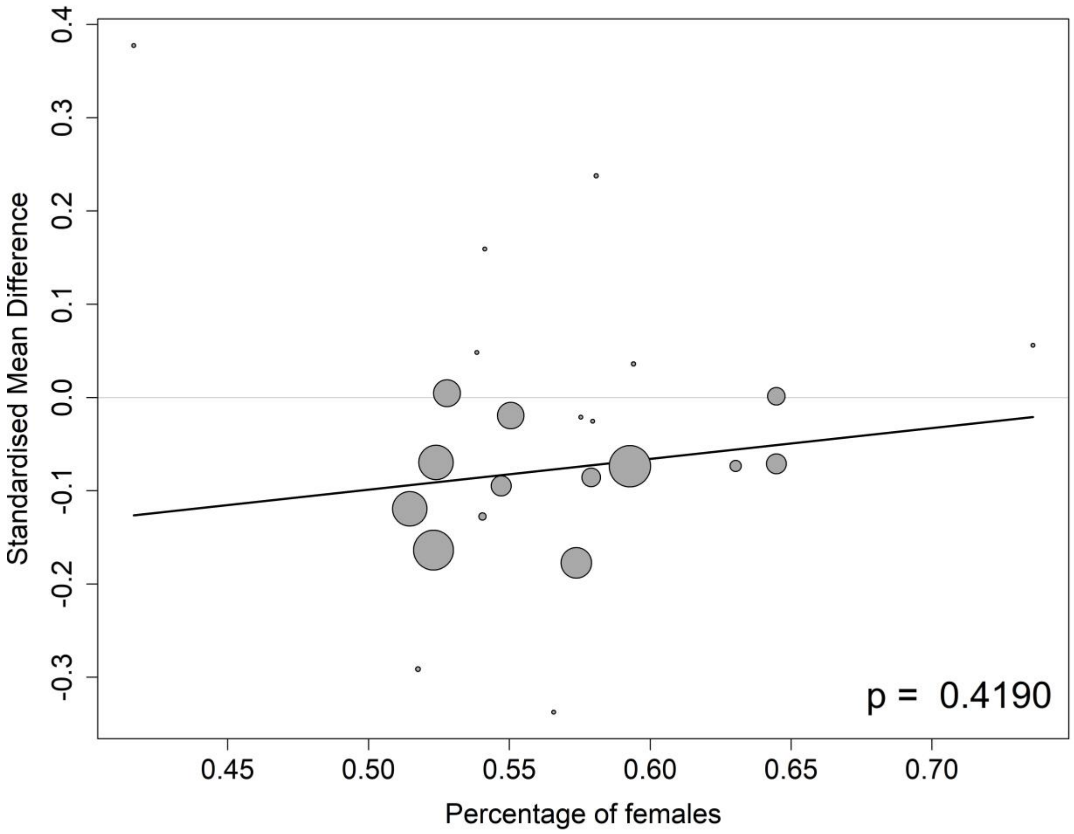

##### 8.1.6 Meta-regressions of ADAS-Cog by the percentage of participants on anti-AD medication at the baseline

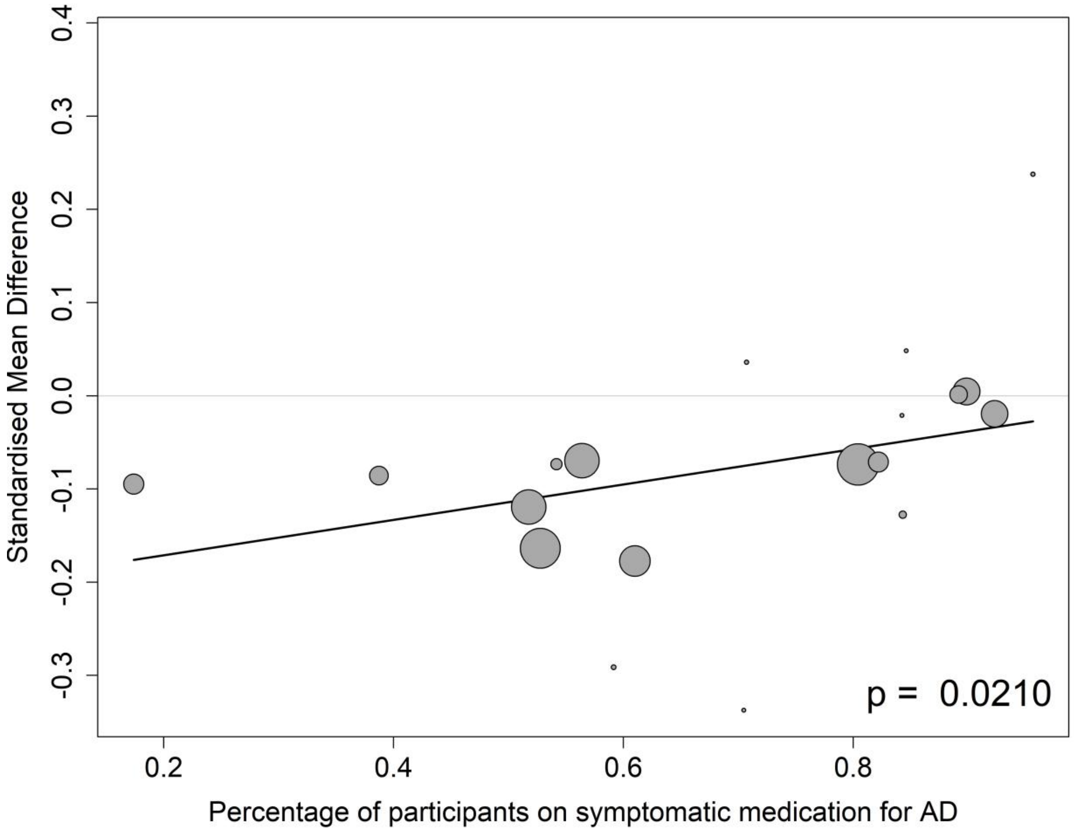

#### 8.2 CDR-SB

##### 8.2.1 Meta-regressions of CDR-SB by baseline CDR-SB score

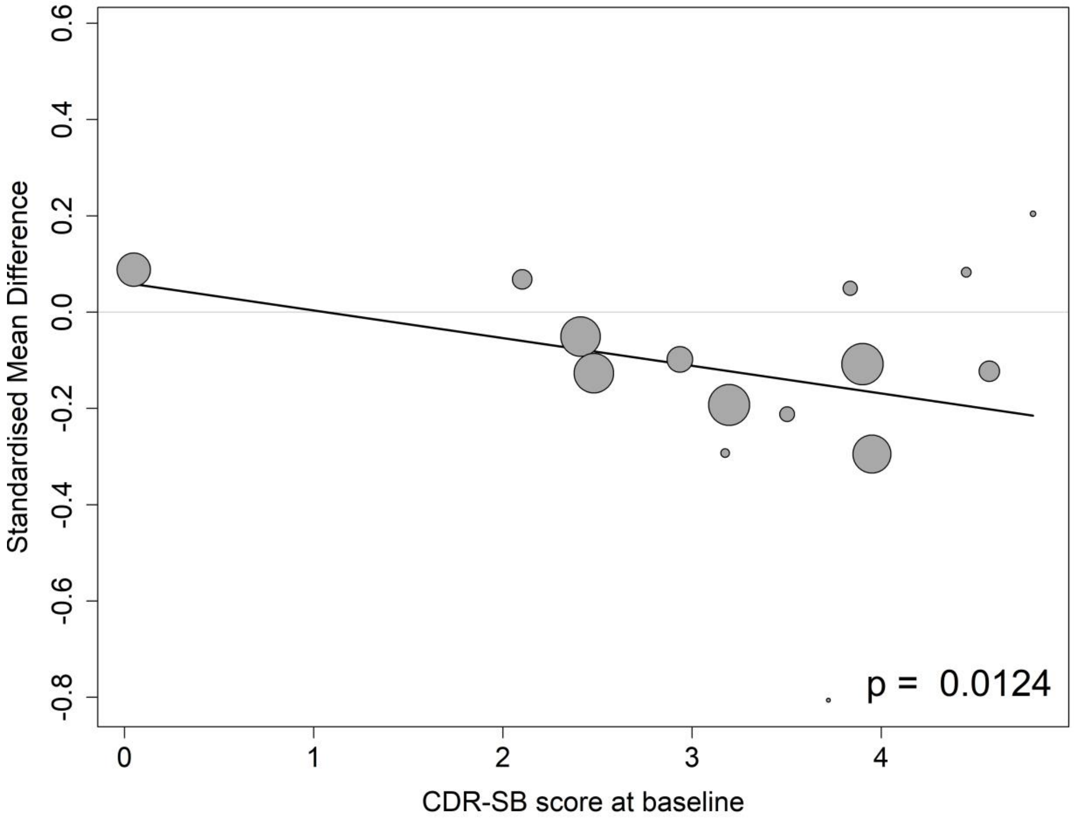

##### 8.2.2 Meta-regressions of CDR-SB by baseline MMSE score

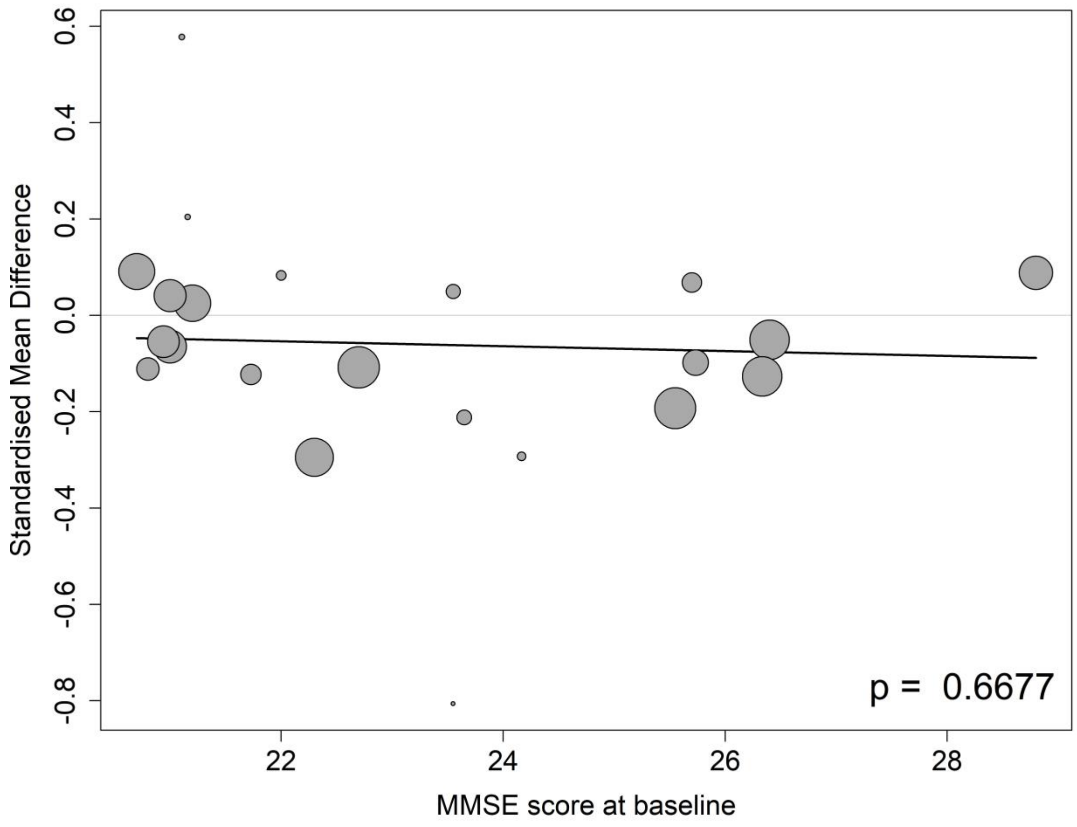

##### 8.2.3 Meta-regressions of CDR-SB by the mean age at the baseline

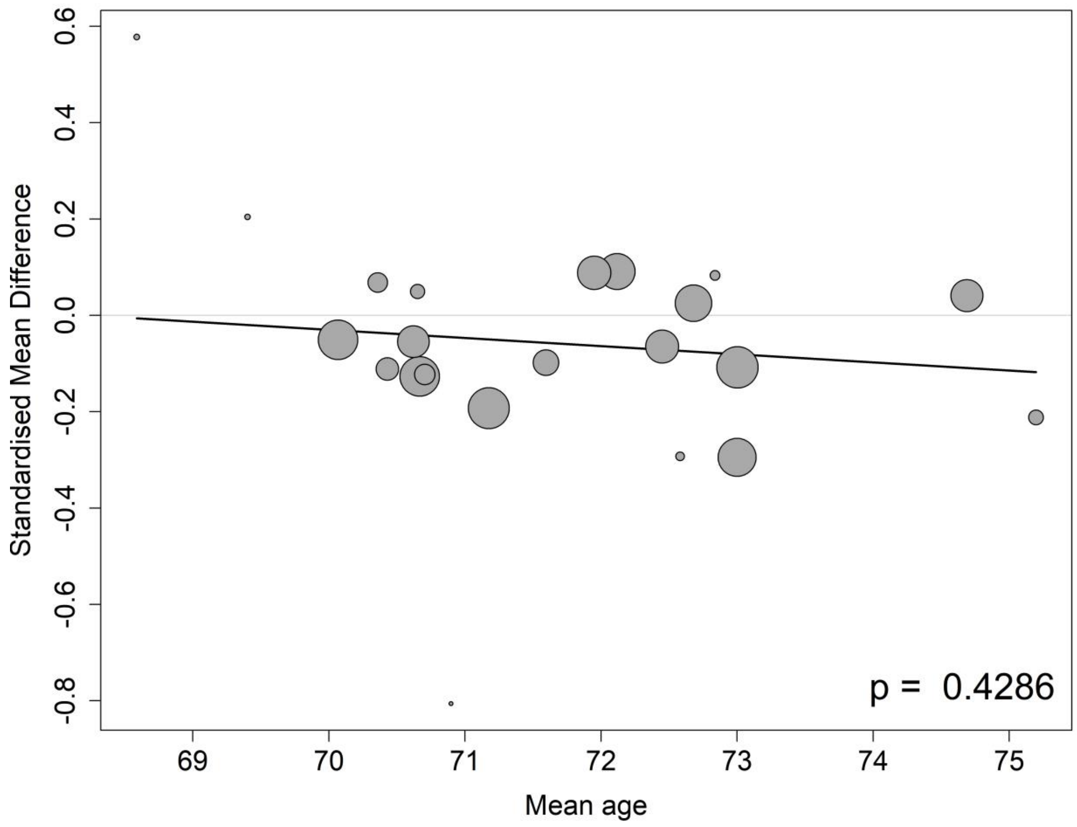

##### 8.2.4 Meta-regressions of CDR-SB by the percentage of APOE4 carrier at the baseline

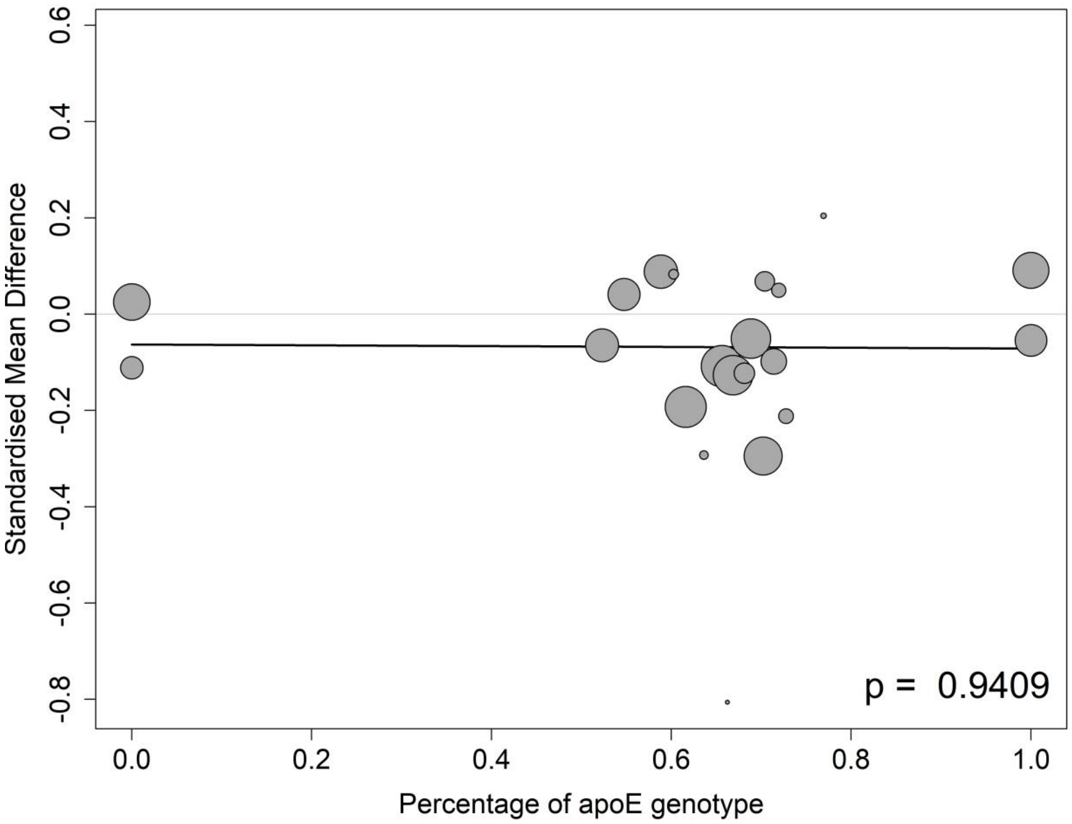

##### 8.2.5 Meta-regressions of CDR-SB by the percentage of female participants at the baseline

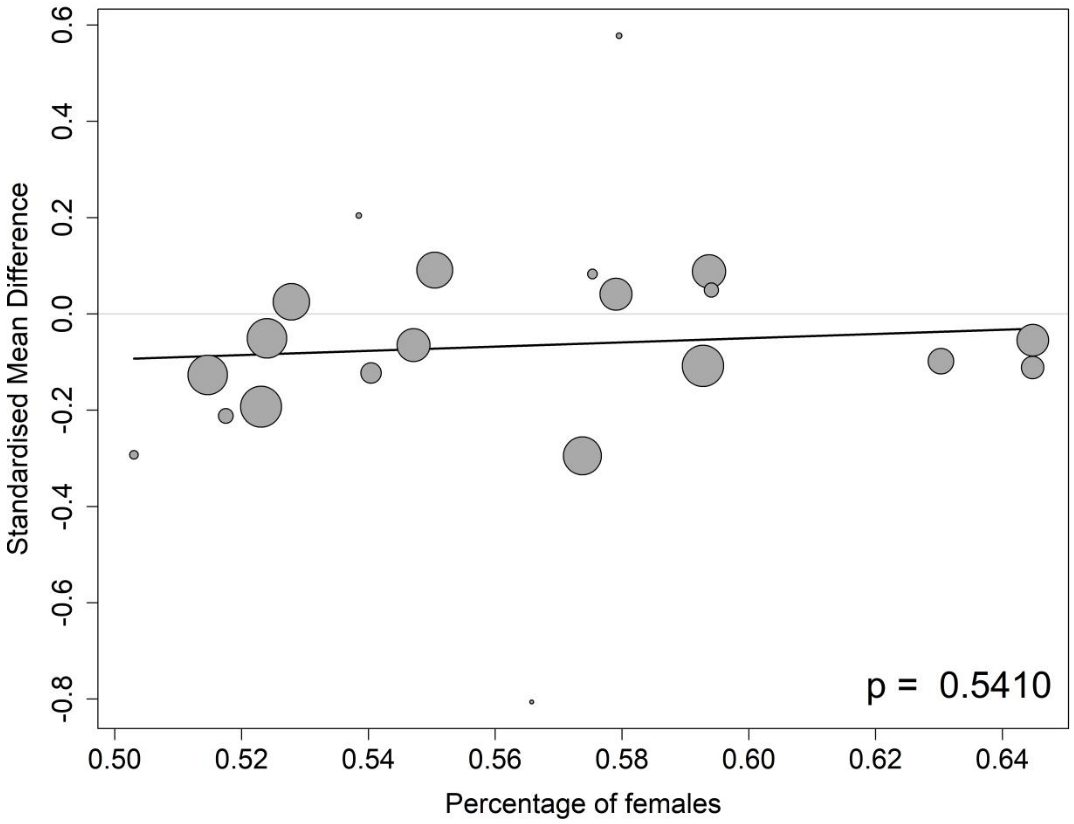

##### 8.2.6 Meta-regressions of CDR-SB by the percentage of participants on anti-AD medication at the baseline

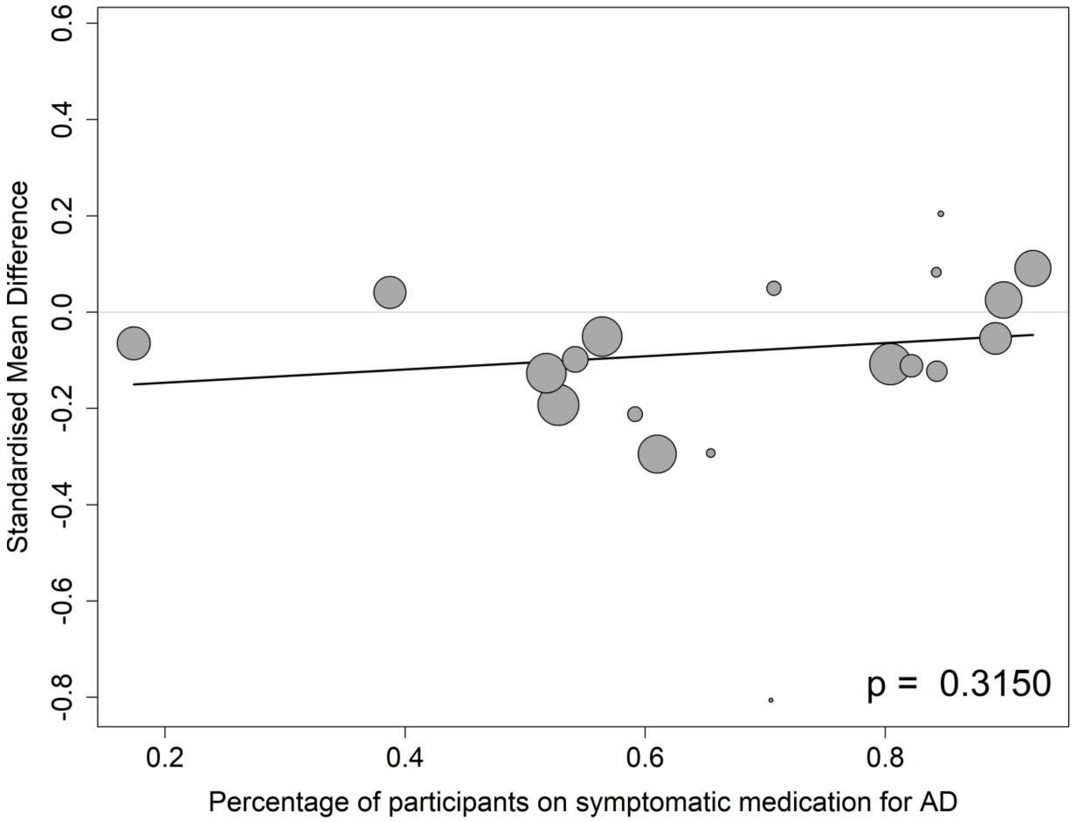

#### 8.3 MMSE

##### 8.3.1 Meta-regressions of MMSE by baseline CDR-SB score

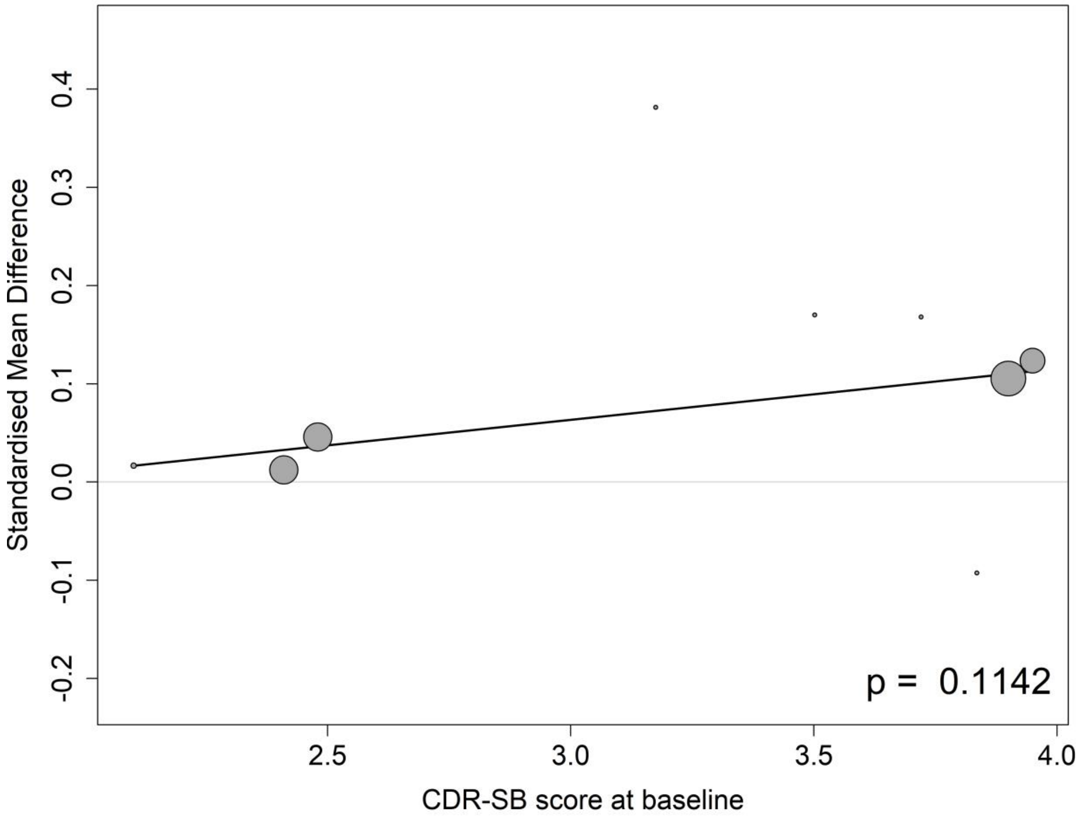

##### 8.3.2 Meta-regressions of MMSE by baseline MMSE score

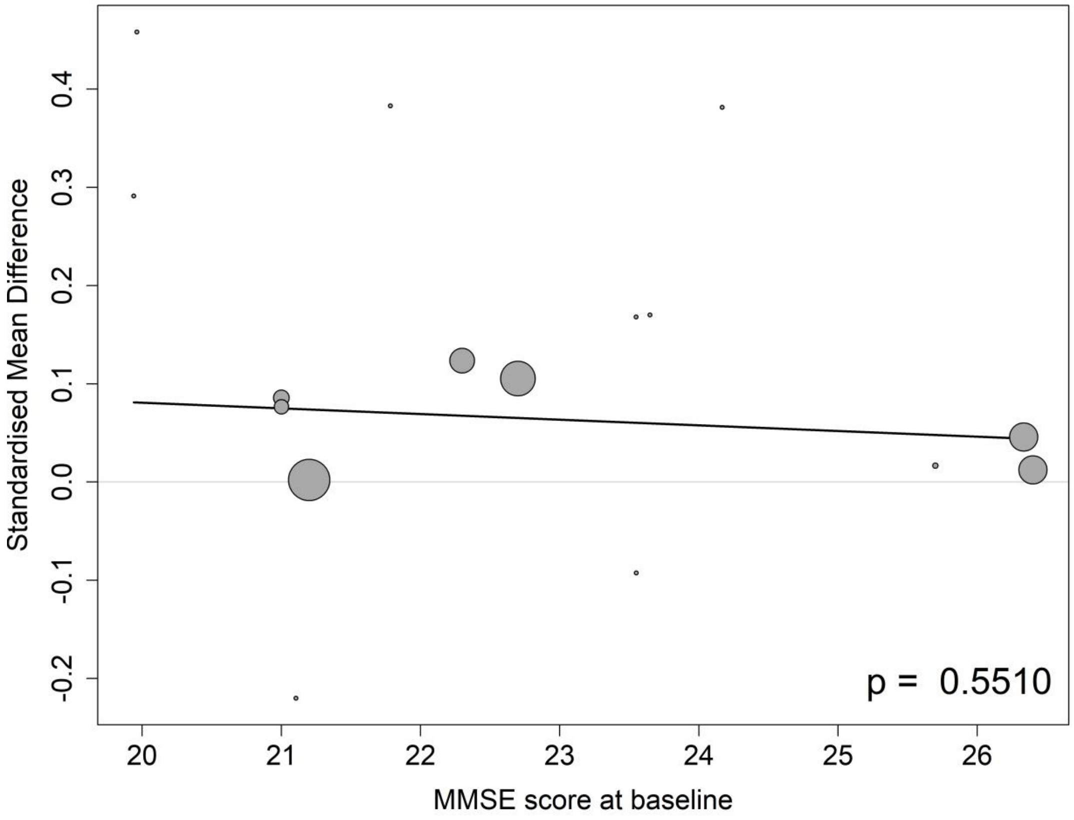

##### 8.3.3 Meta-regressions of MMSE by the mean age at the baseline

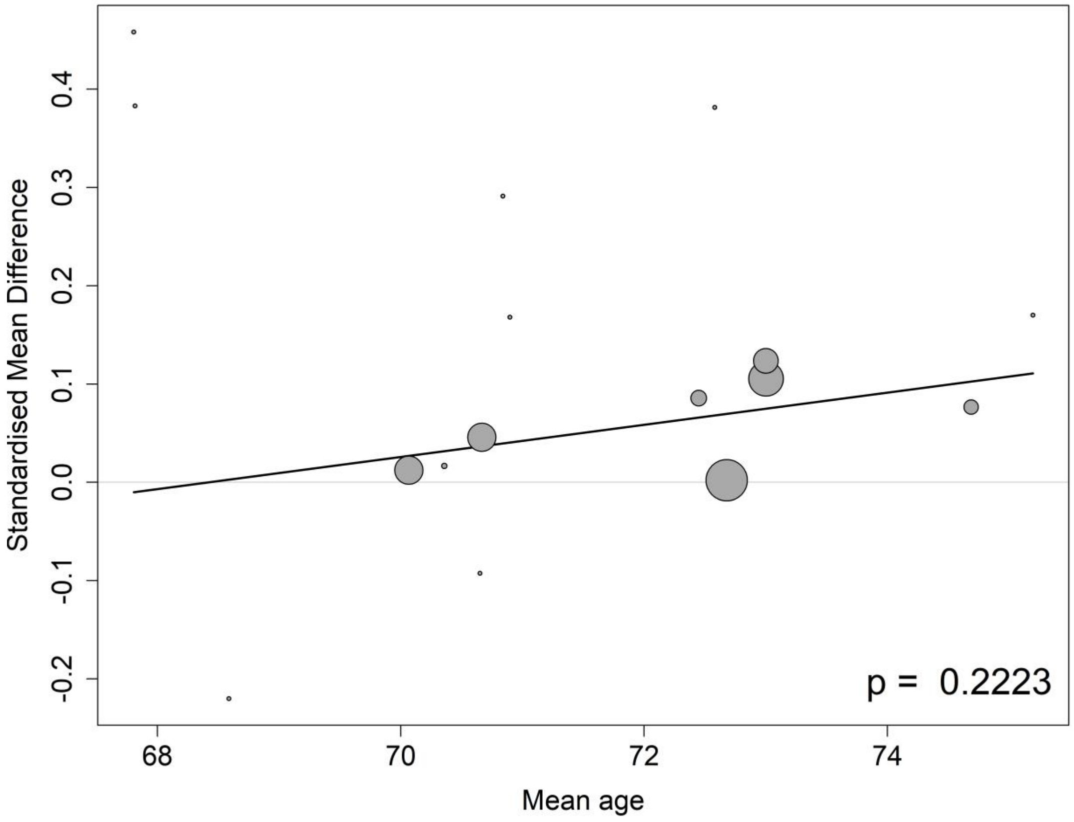

##### 8.3.4 Meta-regressions of MMSE by the percentage of APOE4 carrier at the baseline

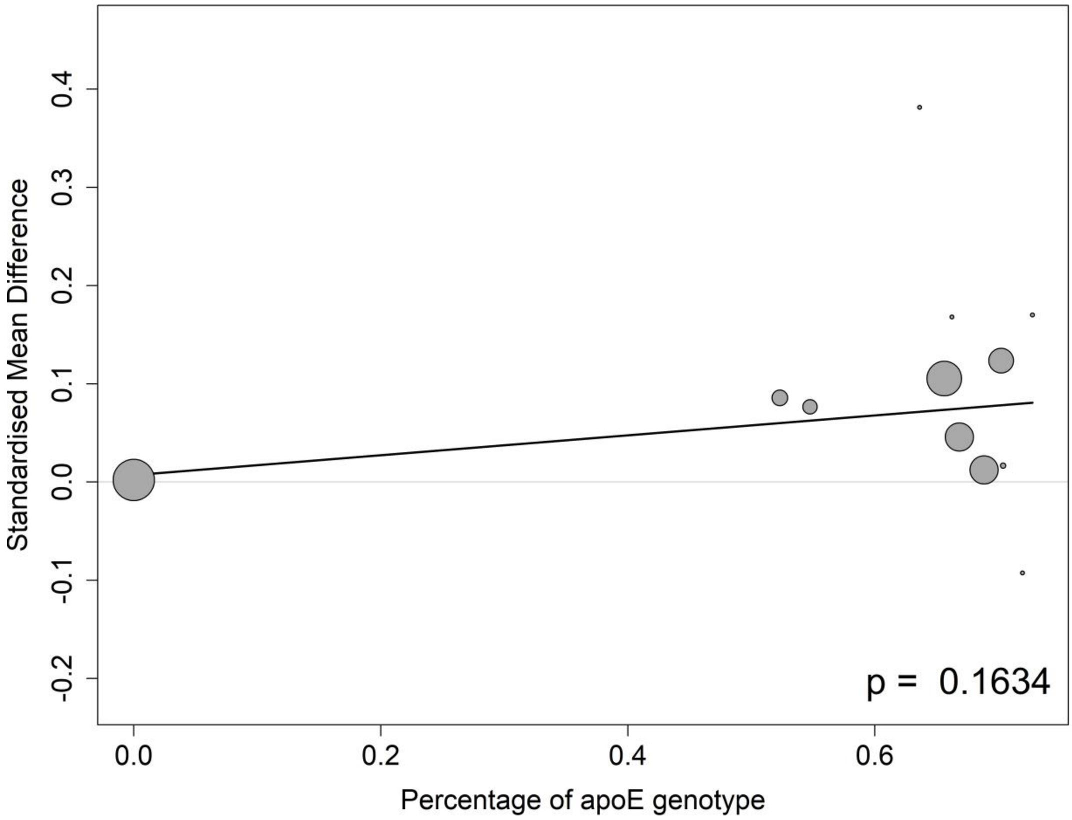

**8.3.5 Meta-regressions of MMSE by the percentage of female participants at the baseline**

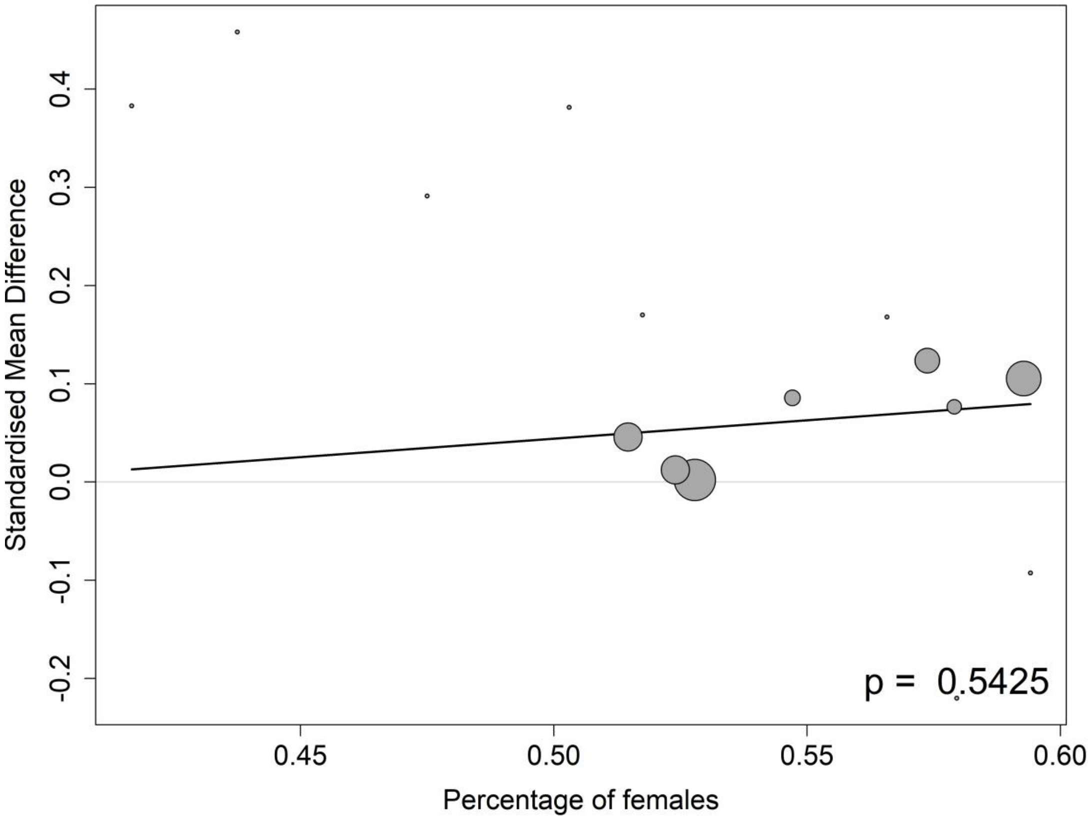

##### 8.3.6 Meta-regressions of MMSE by the percentage of participants on anti-AD medication at the baseline

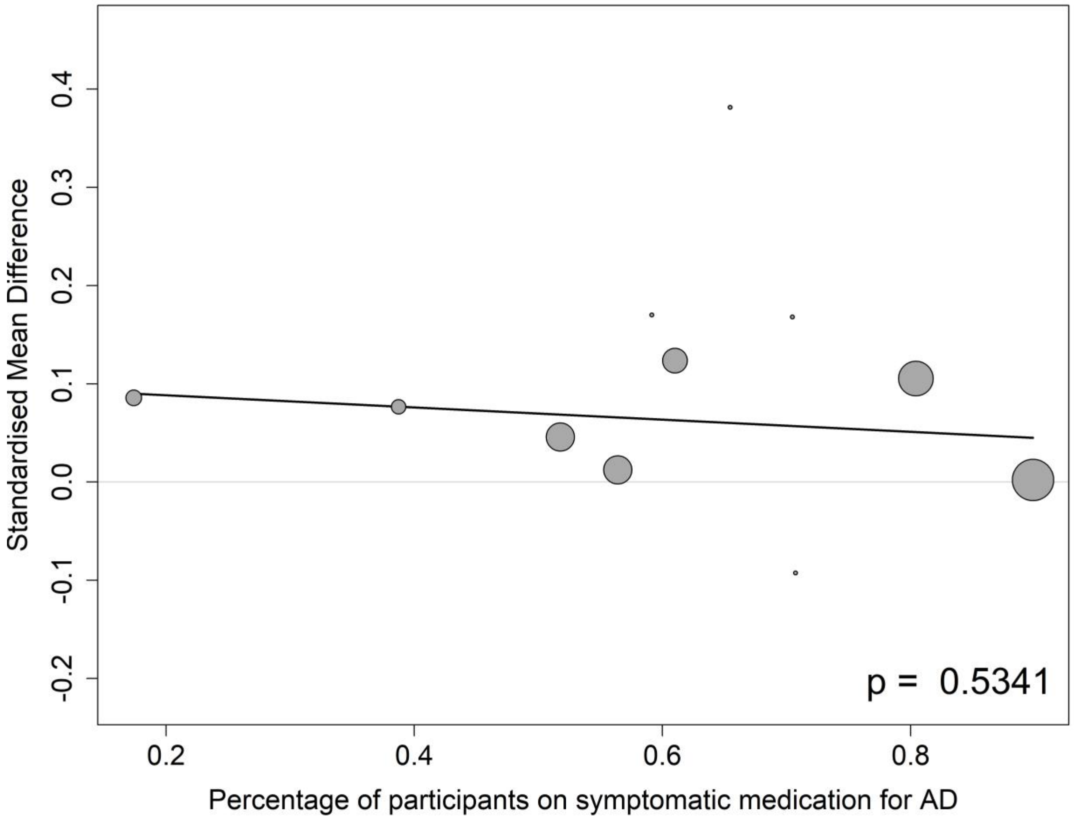

#### 8.4 Amyloid PET

##### 8.4.1 Meta-regressions of amyloid PET by baseline CDR-SB score

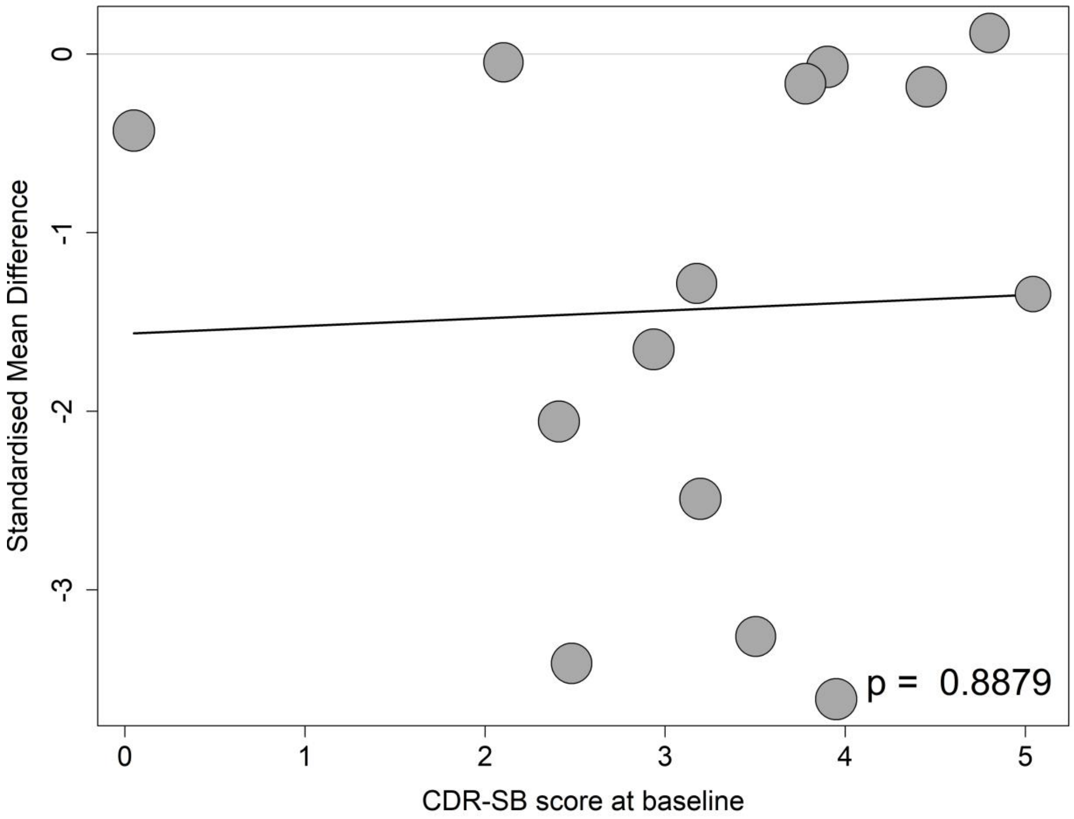

##### 8.4.2 Meta-regressions of amyloid PET by baseline MMSE score

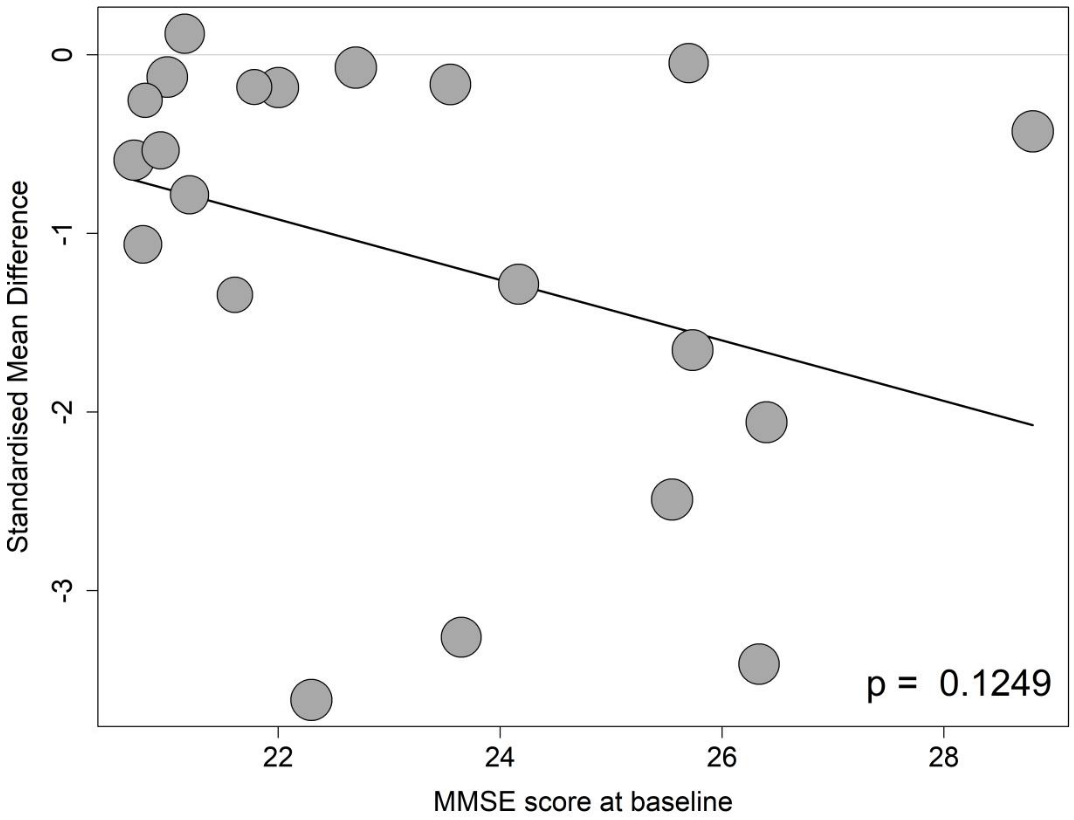

##### 8.4.3 Meta-regressions of amyloid PET by the mean age at the baseline

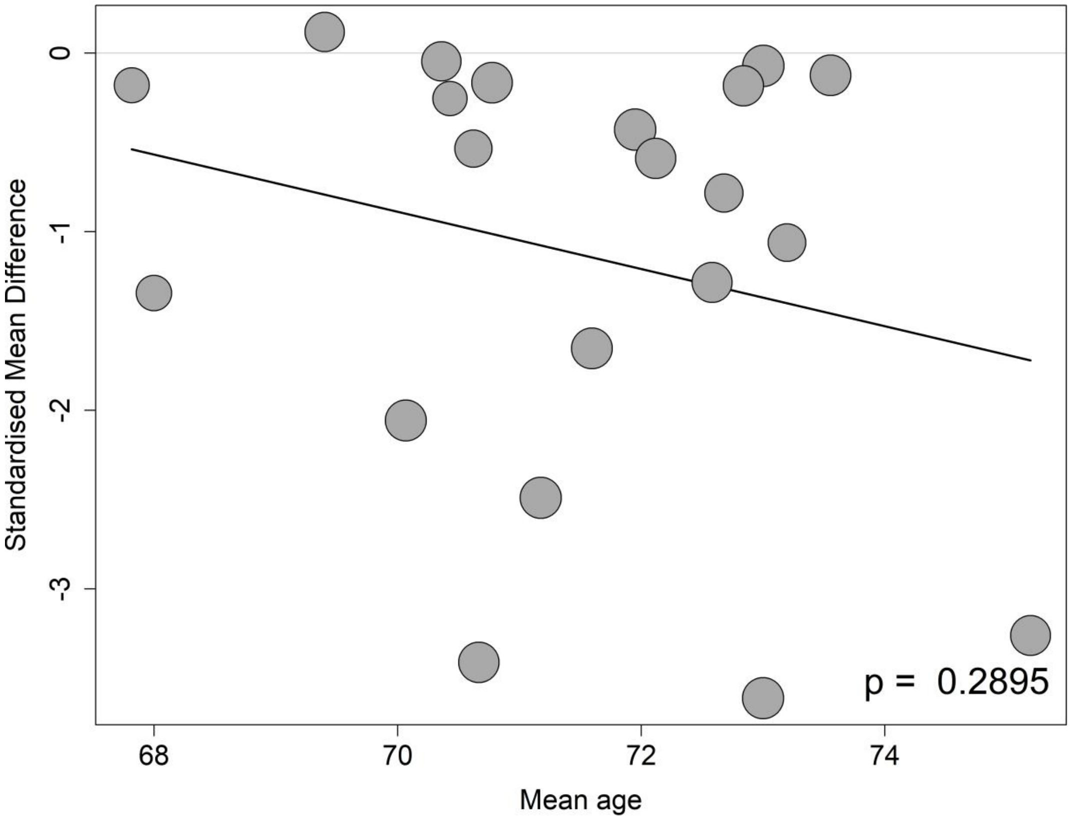

##### 8.4.4 Meta-regressions of amyloid PET by the percentage of APOE4 carrier at the baseline

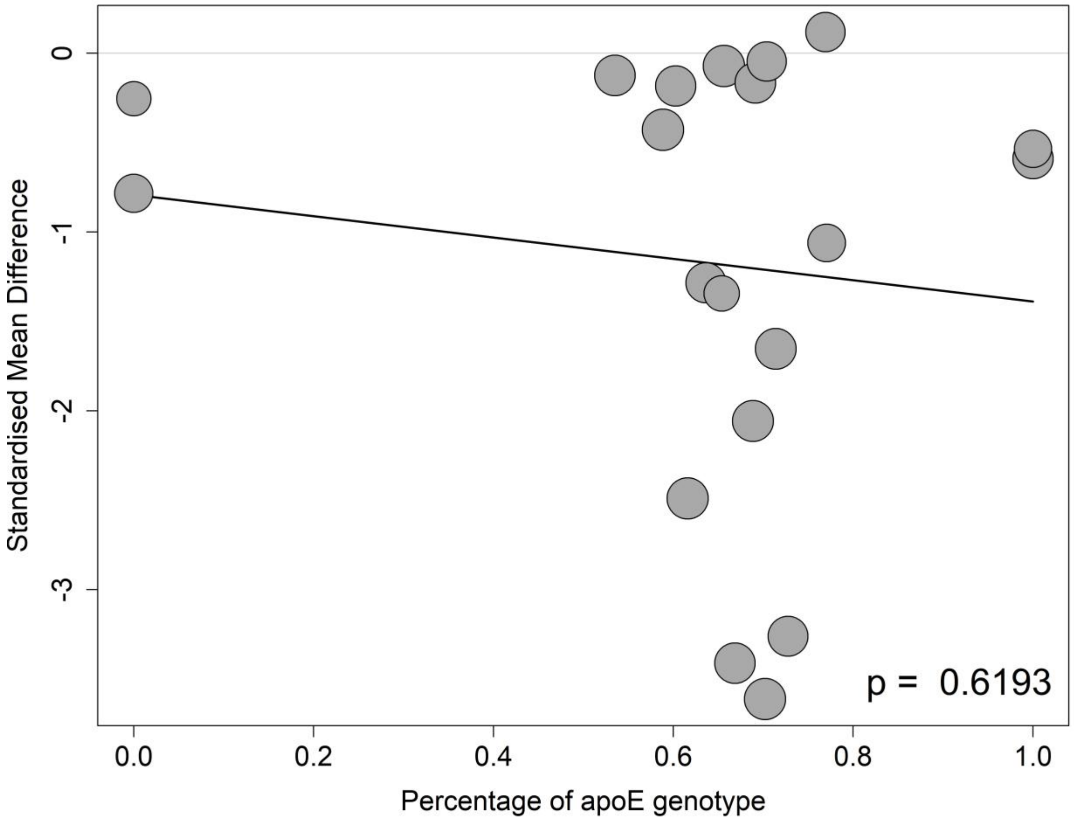

##### 8.4.5 Meta-regressions of amyloid PET by the percentage of female participants at the baseline

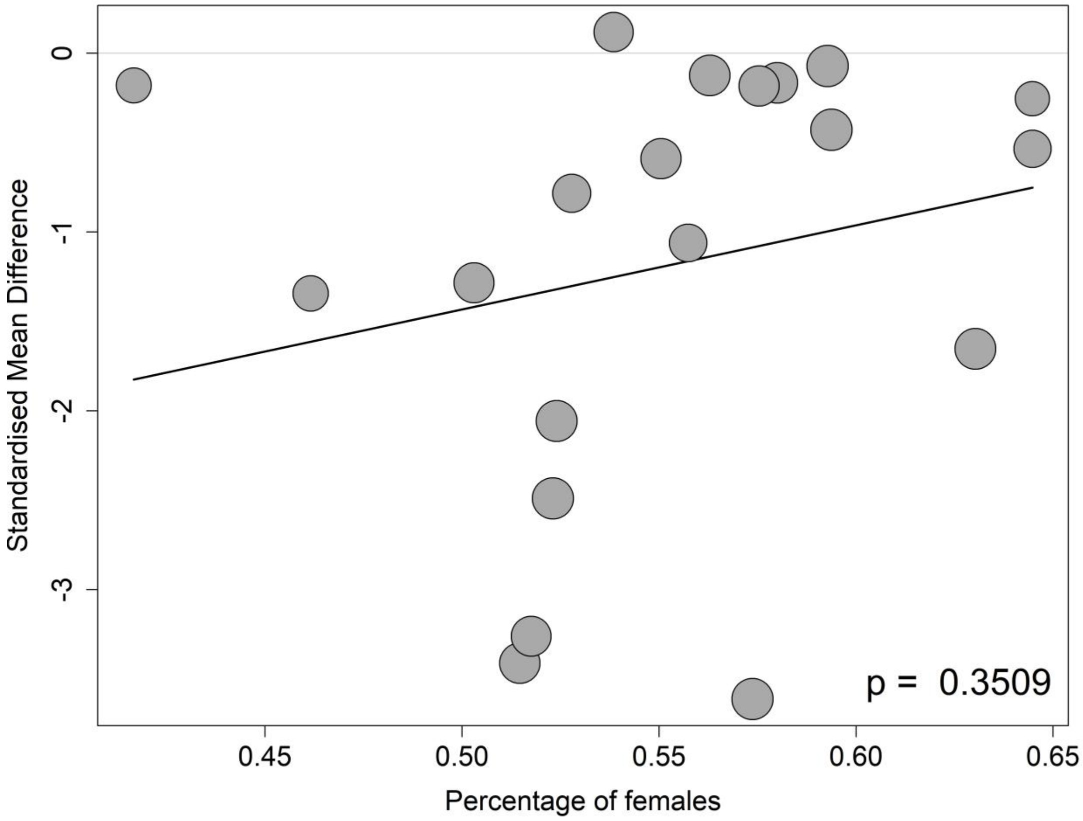

##### 8.4.6 Meta-regressions of amyloid PET by the percentage of participants on anti-AD medication at the baseline

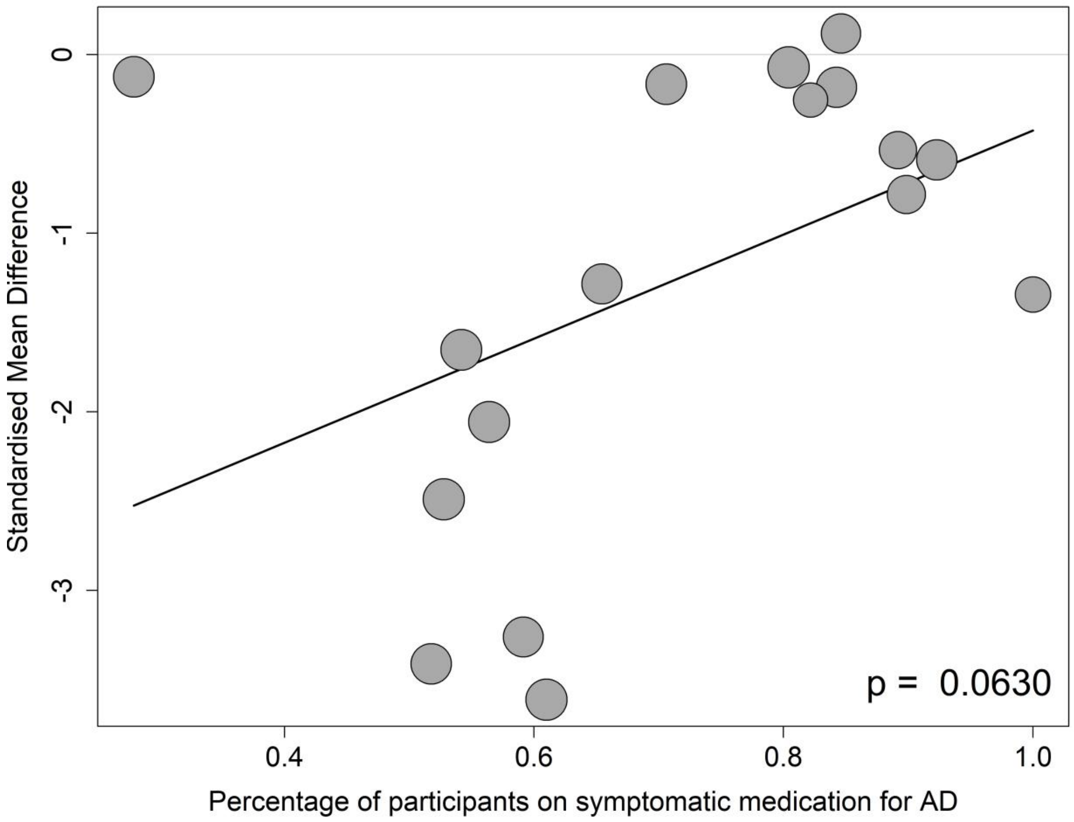

#### 8.5 Tau PET

##### 8.5.1 Meta-regressions of tau PET by baseline CDR-SB score

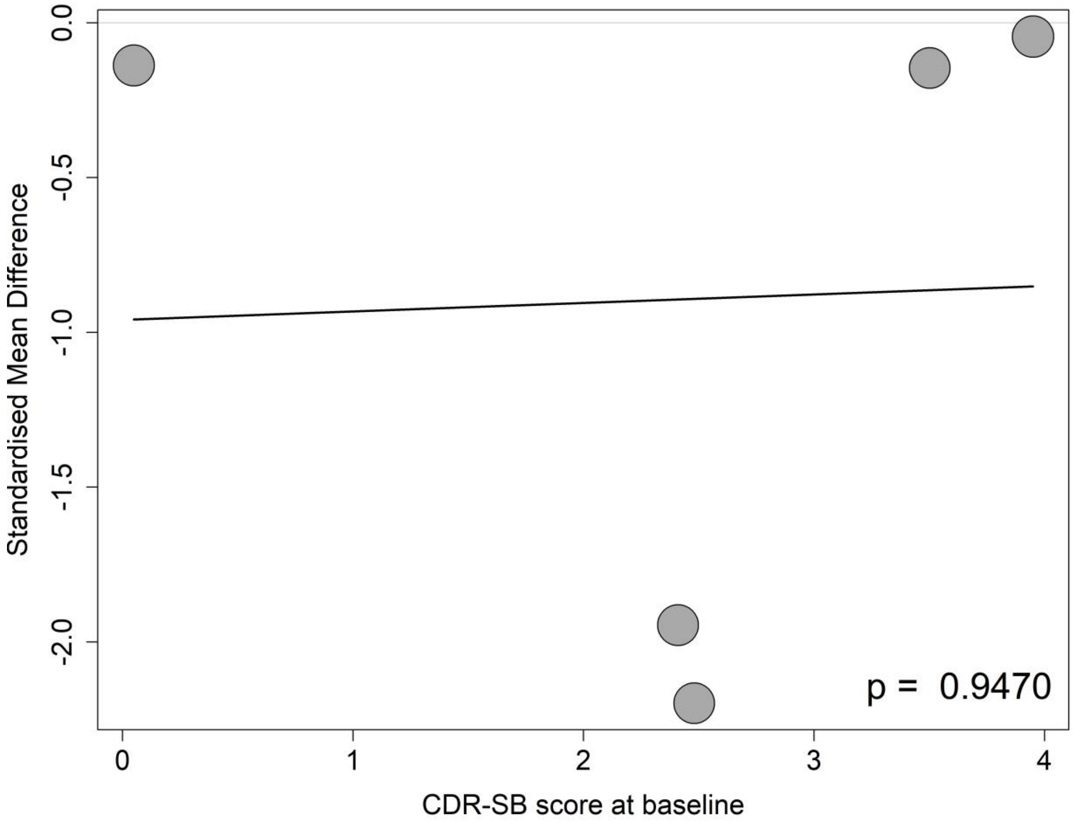

##### 8.5.2 Meta-regressions of tau PET by baseline MMSE score

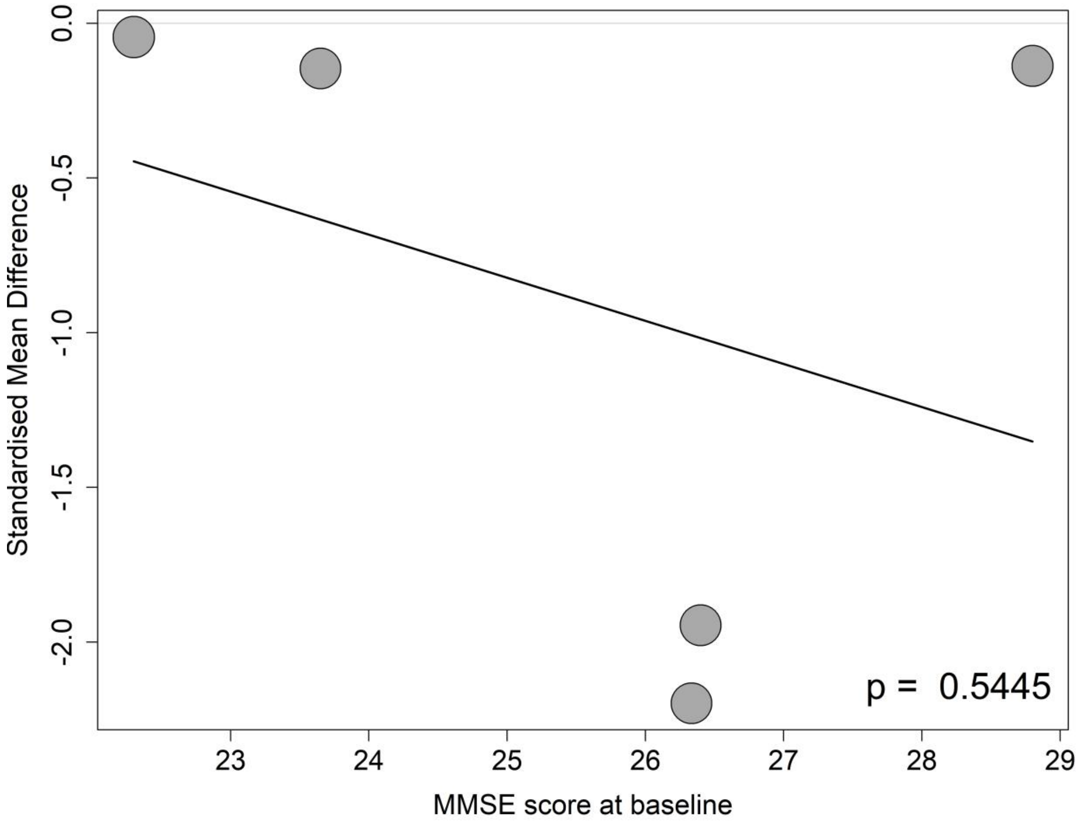

##### 8.5.3 Meta-regressions of tau PET by the mean age at the baseline

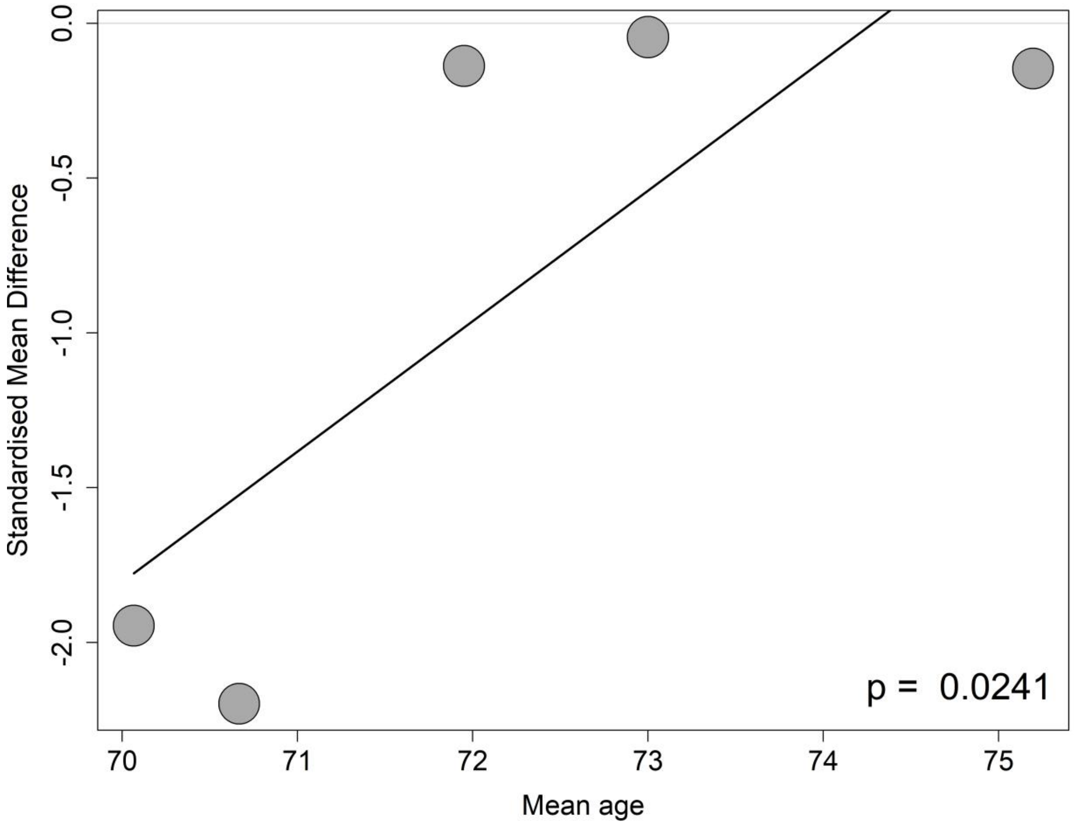

##### 8.5.4 Meta-regressions of tau PET by the percentage of APOE4 carrier at the baseline

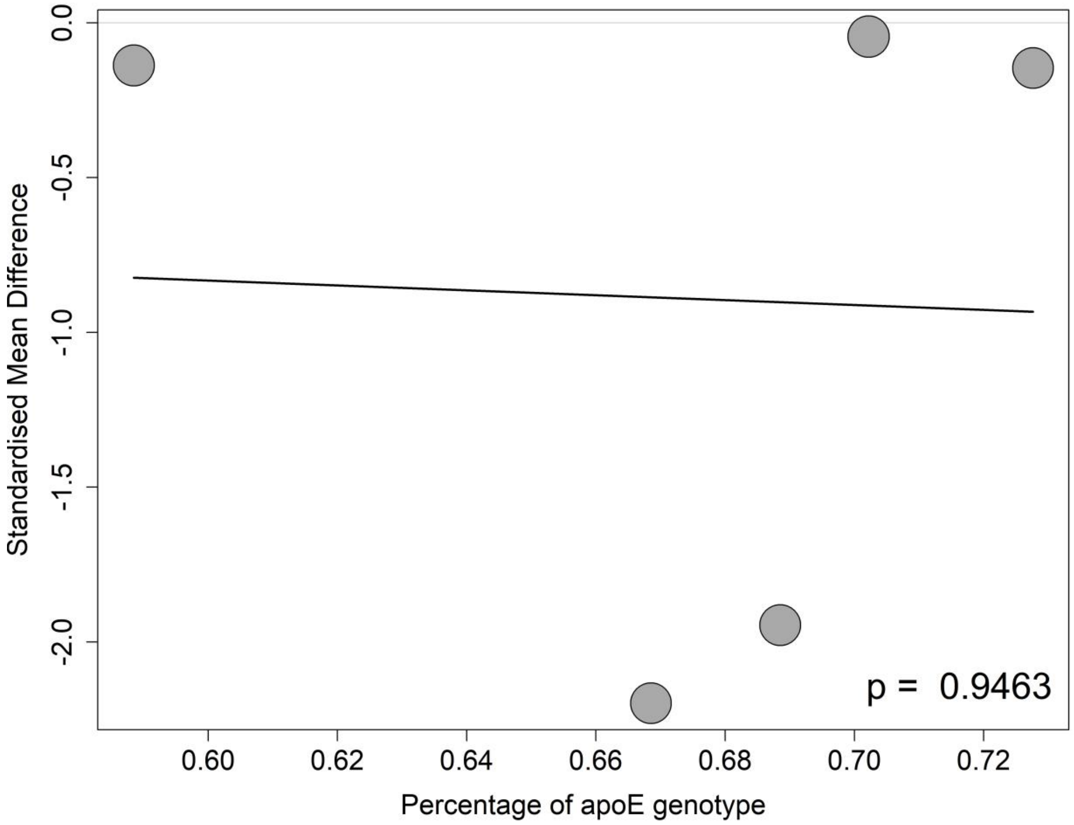

##### 8.5.5 Meta-regressions of tau PET by the percentage of female participants at the baseline

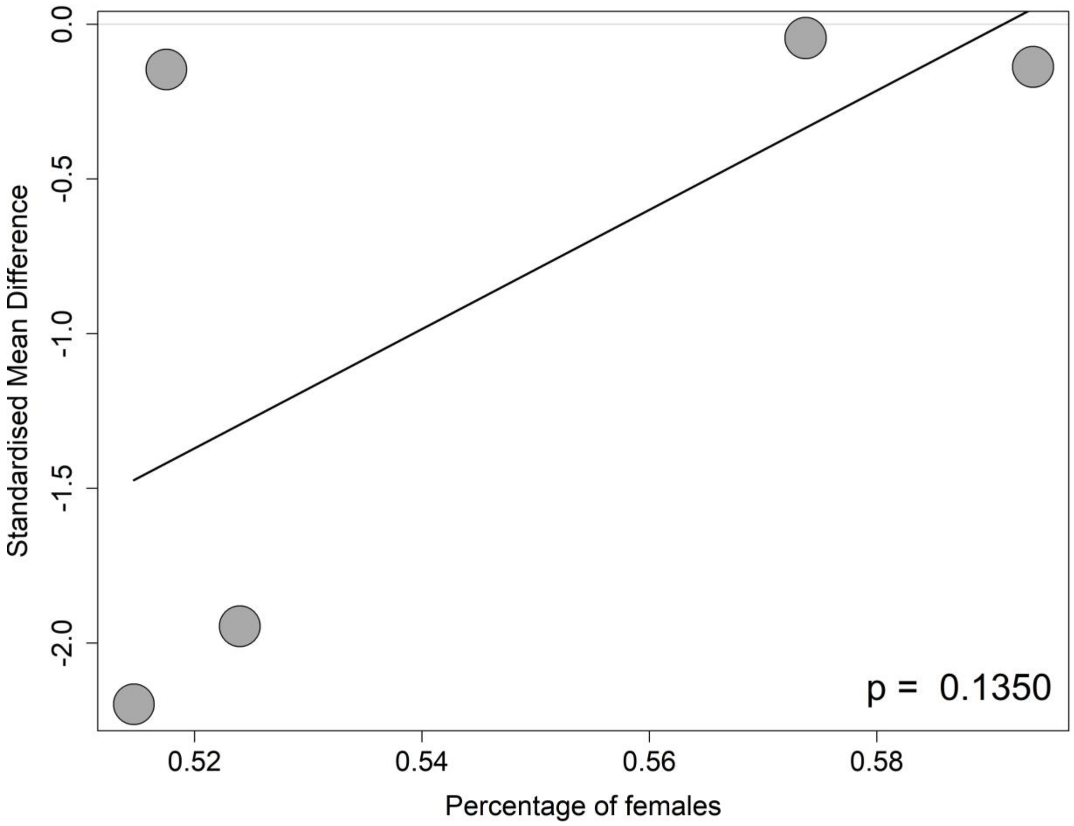

##### 8.5.6 Meta-regressions of tau PET by the percentage of participants on anti-AD medication at the baseline

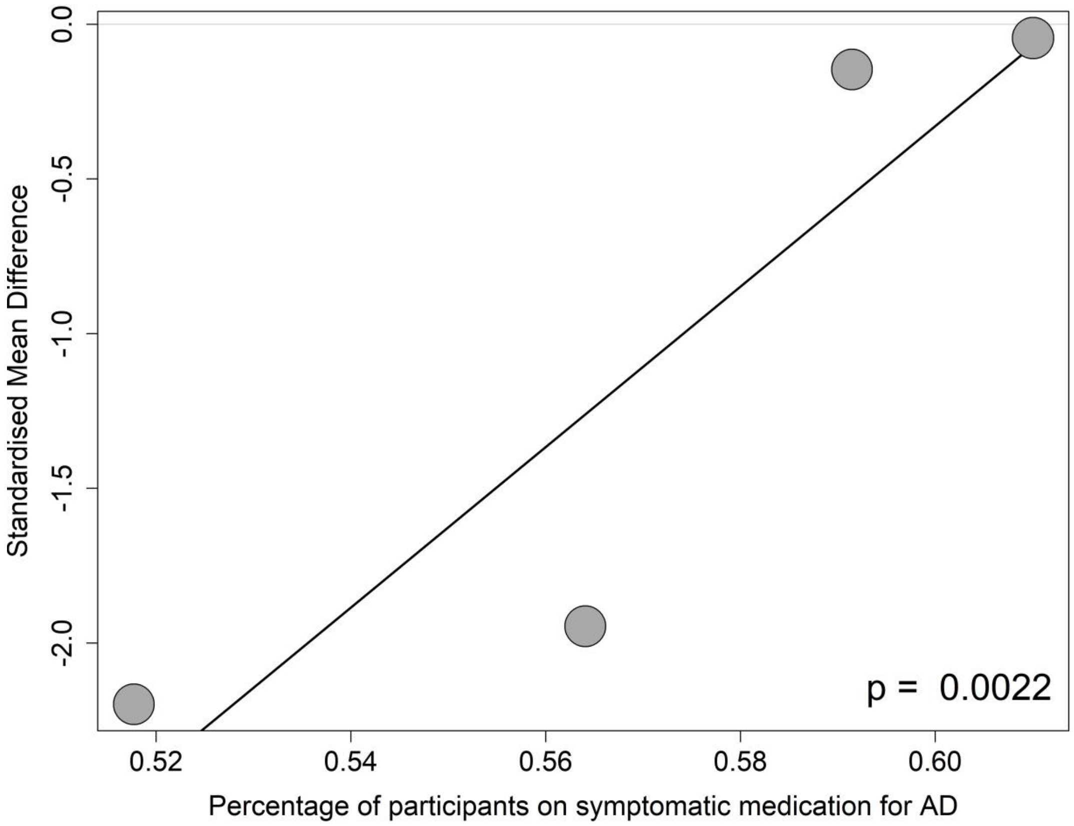

#### 8.6 Volumes of the whole brain

##### 8.6.1 Meta-regressions of volumes of the whole brain by baseline CDR-SB score

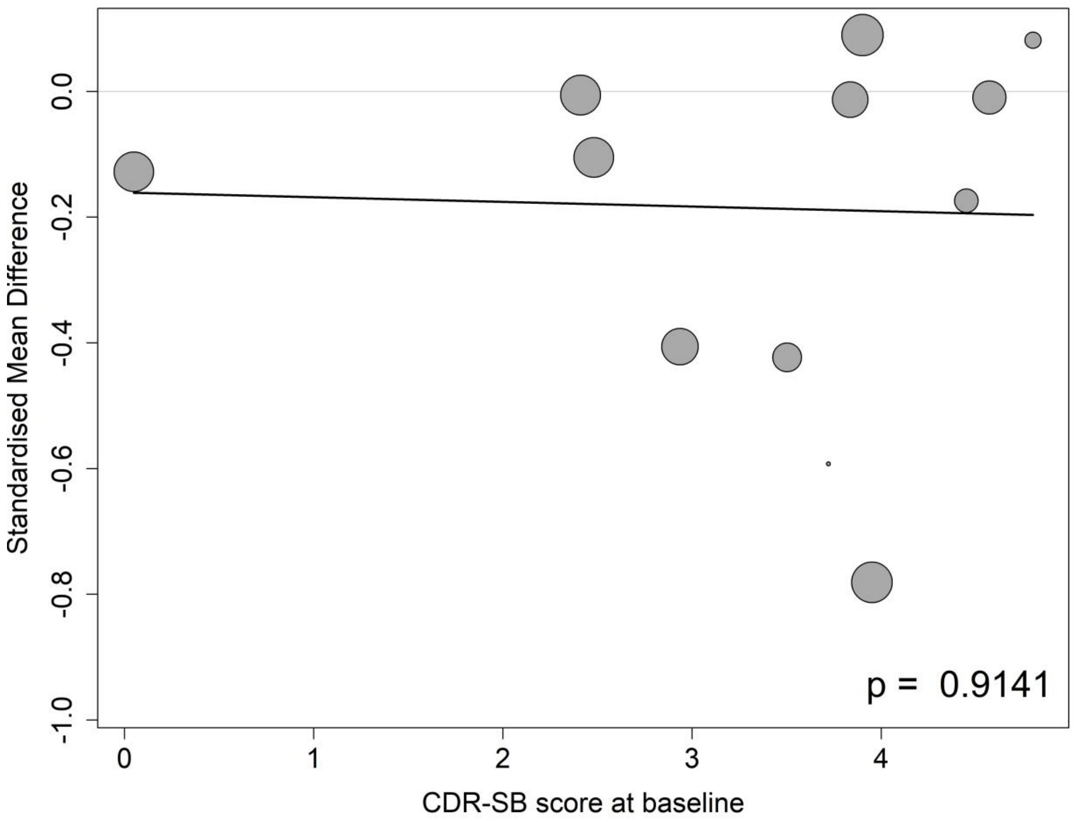

##### 8.6.2 Meta-regressions of volumes of the whole brain by baseline MMSE score

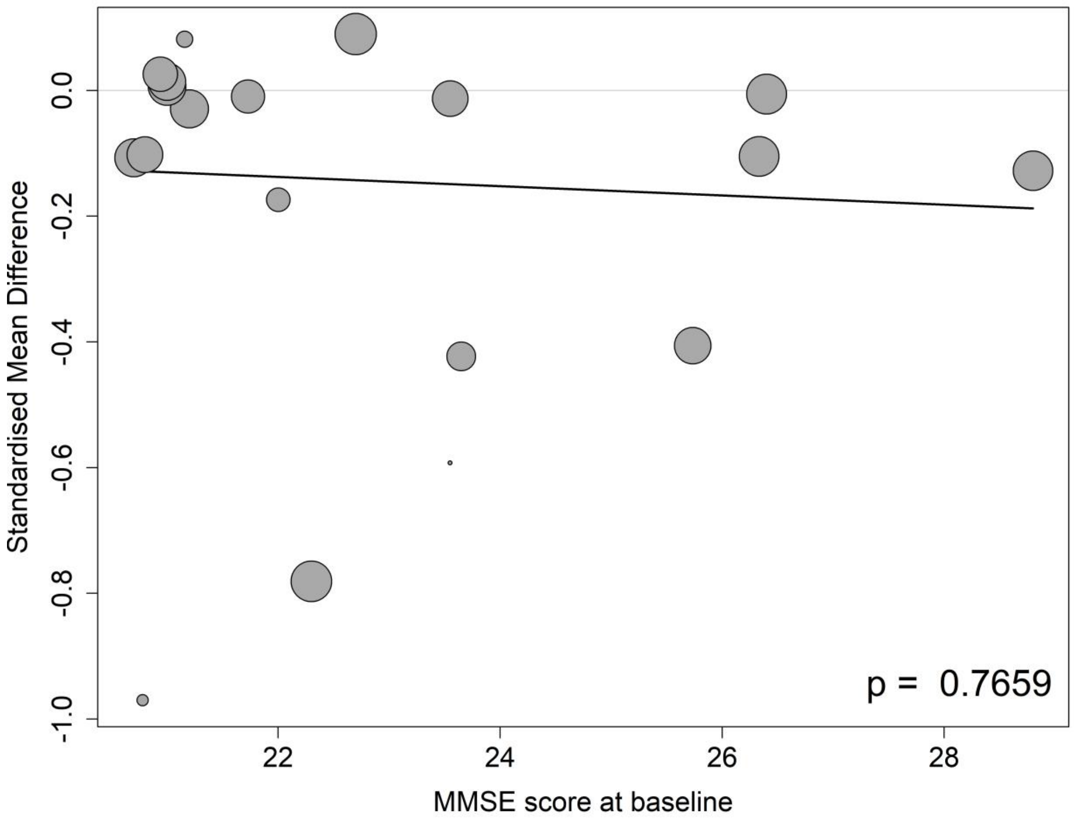

##### 8.6.3 Meta-regressions of volumes of the whole brain by the mean age at the baseline

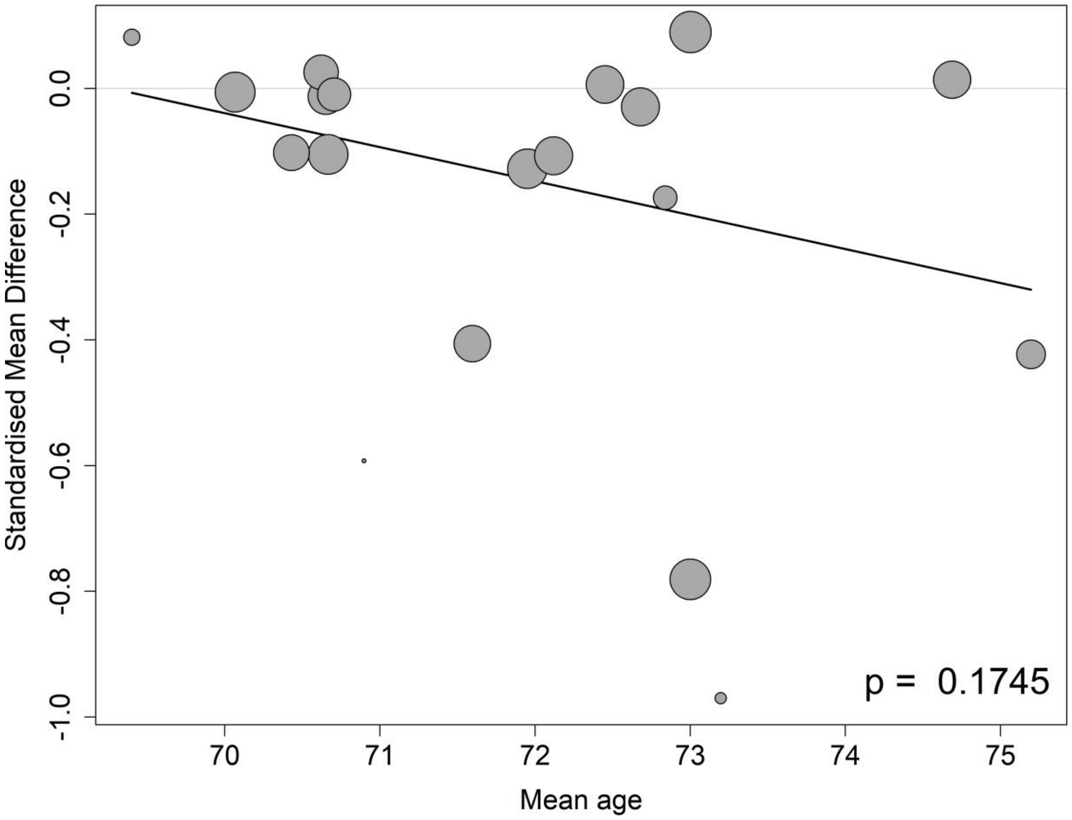

##### 8.6.4 Meta-regressions of volumes of the whole brain by the percentage of APOE4 carrier at the baseline

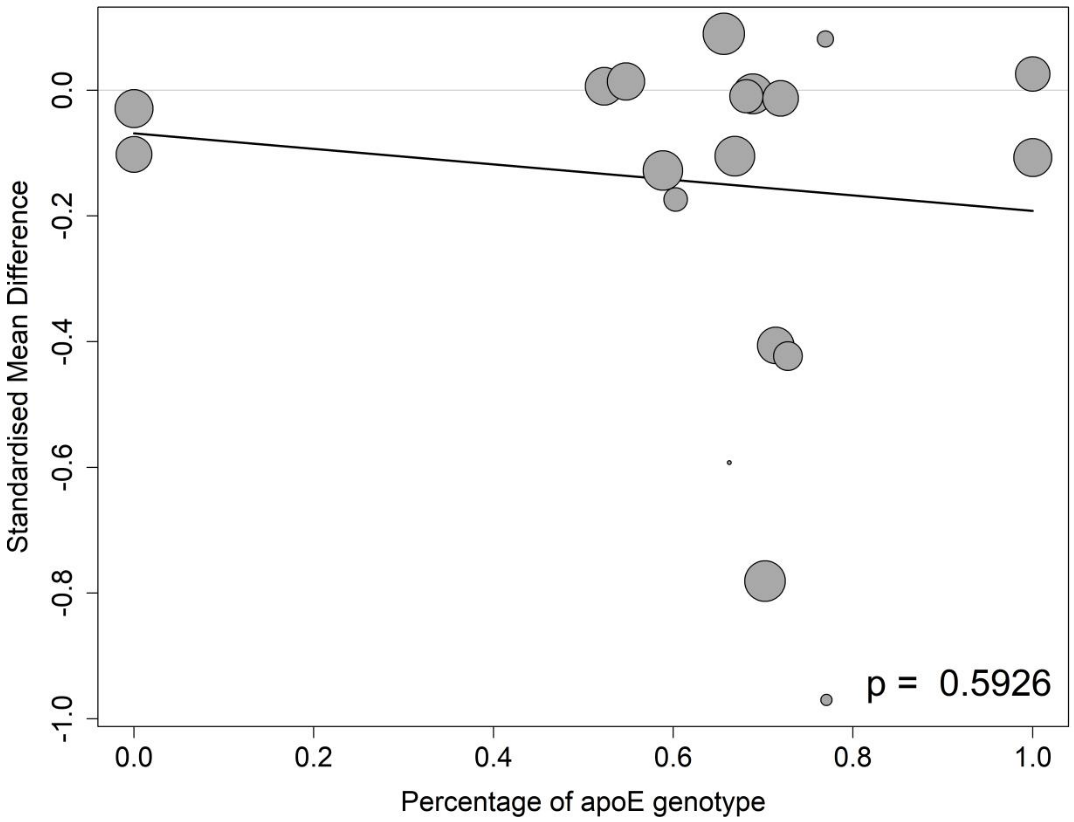

**8.6.5 Meta-regressions of volumes of the whole brain by the percentage of female participants at the baseline**

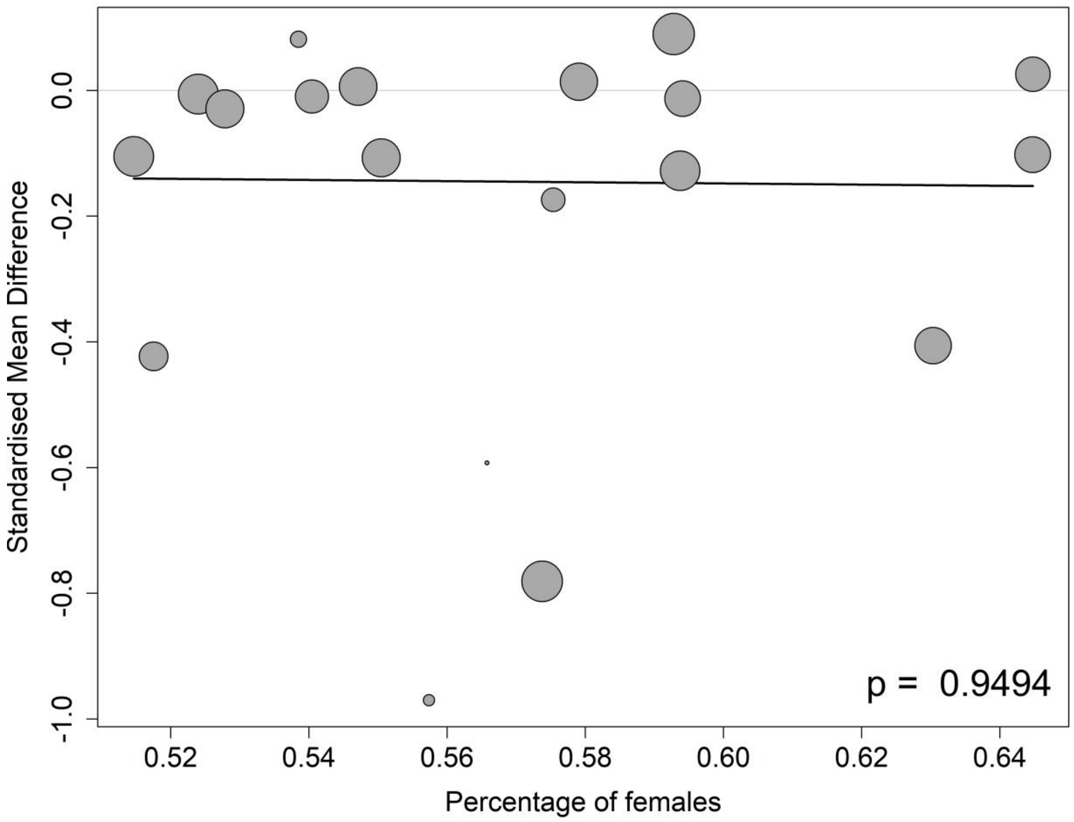

##### 8.6.6 Meta-regressions of volumes of the whole brain by the percentage of participants on anti-AD medication at the baseline

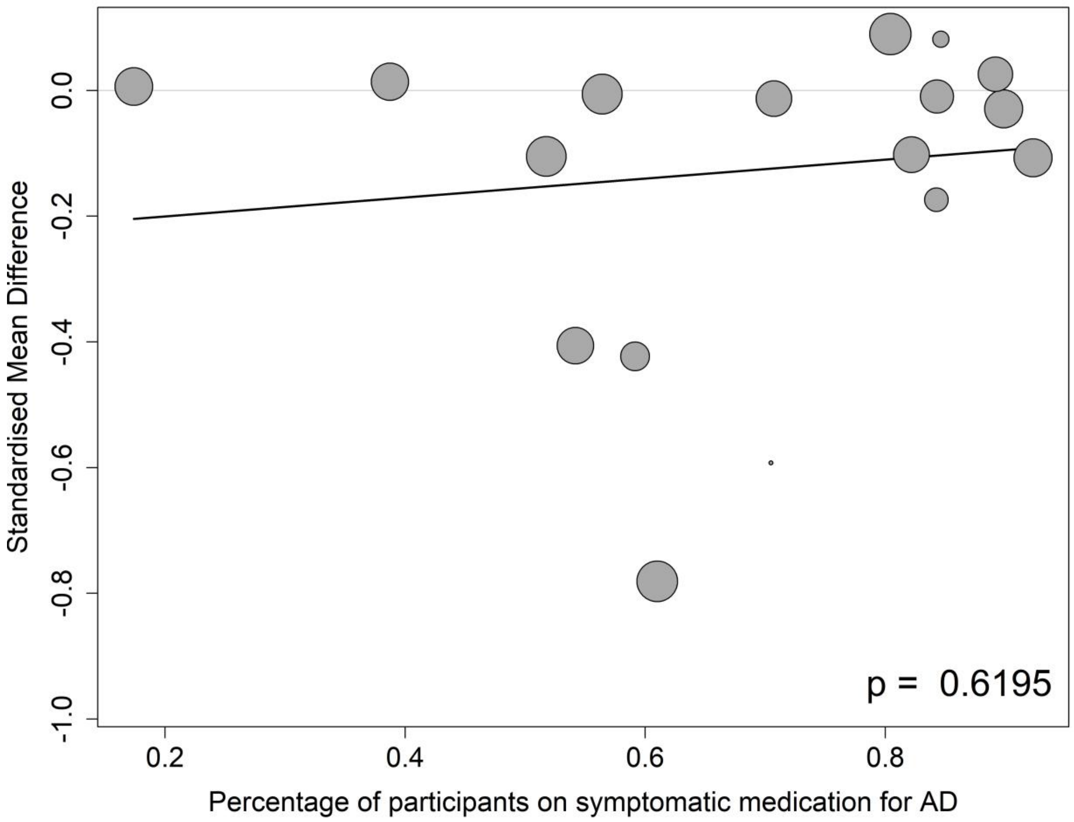

#### 8.7 Volumes of hippocampus

##### 8.7.1 Meta-regressions of volumes of hippocampus by baseline CDR-SB score

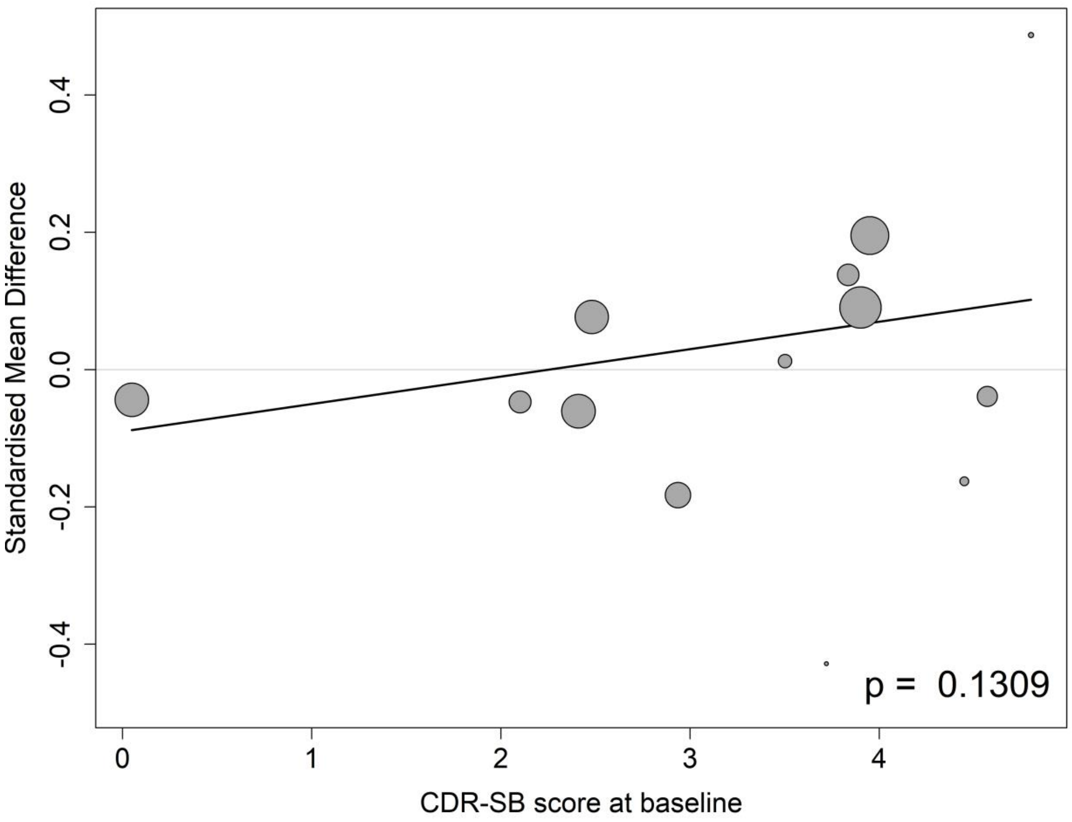

##### 8.7.2 Meta-regressions of volumes of hippocampus by baseline MMSE score

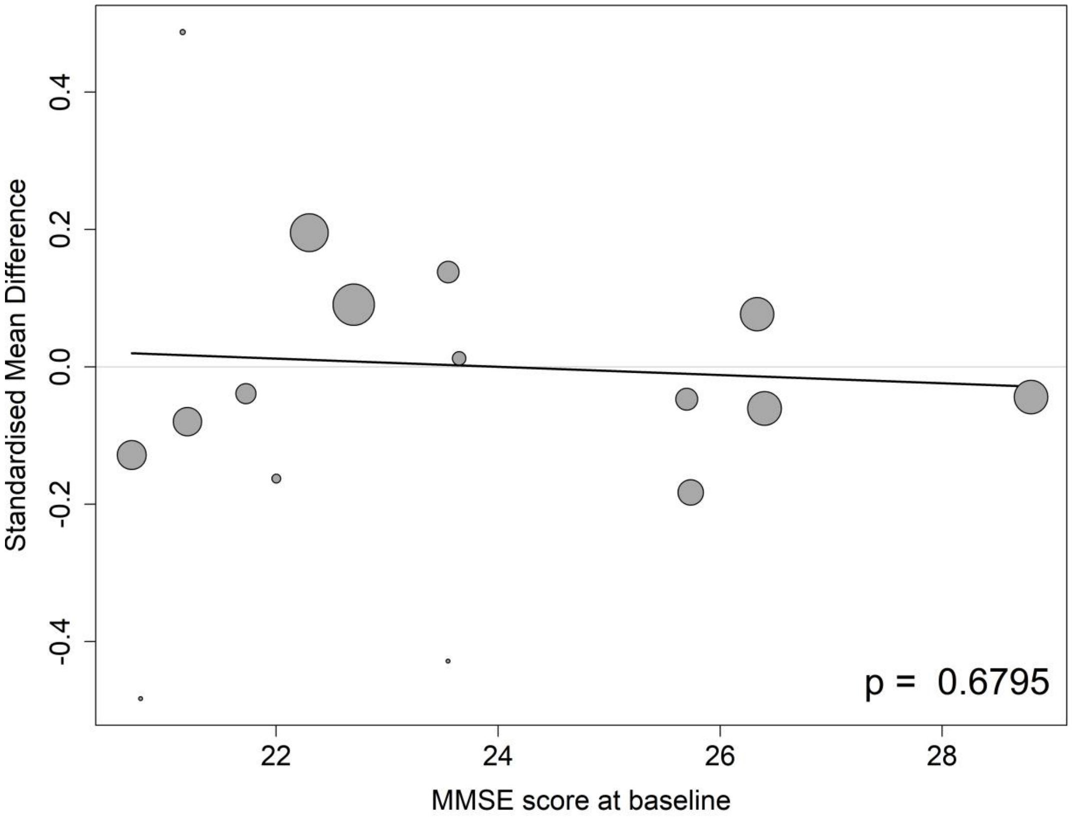

##### 8.7.3 Meta-regressions of volumes of hippocampus by the mean age at the baseline

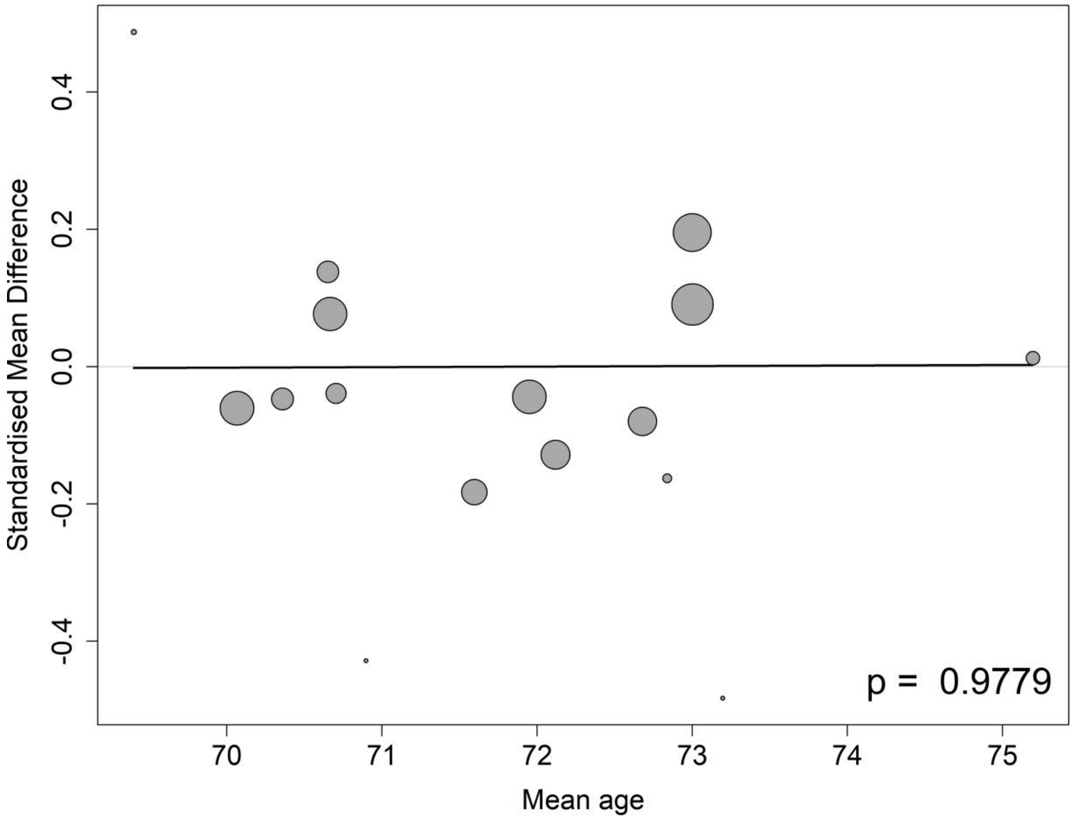

##### 8.7.4 Meta-regressions of volumes of hippocampus by the percentage of APOE4 carrier at the baseline

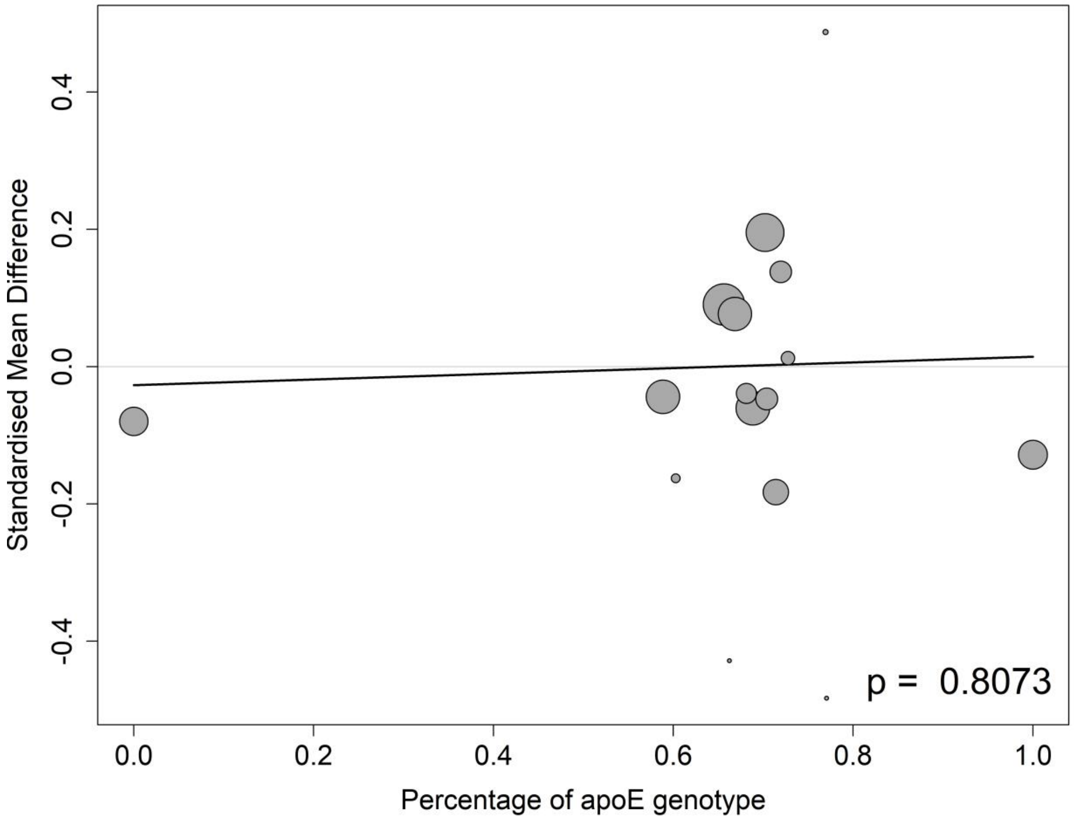

##### 8.7.5 Meta-regressions of volumes of hippocampus by the percentage of female participants at the baseline

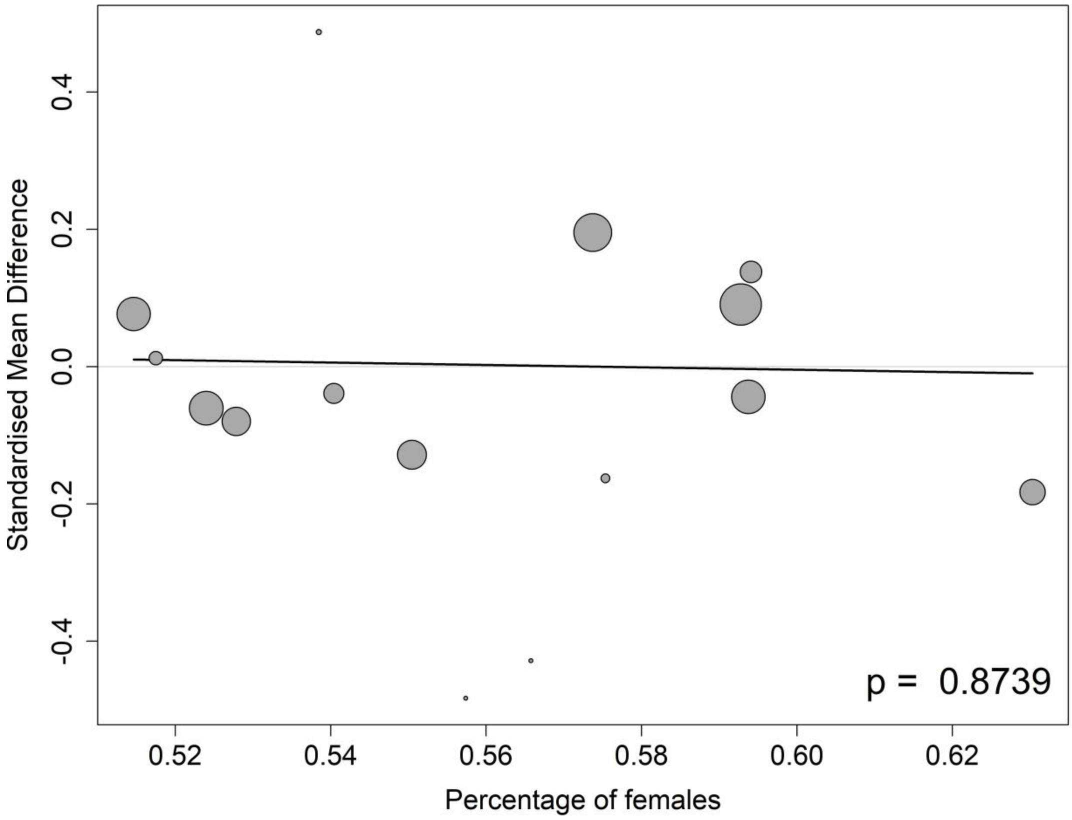

##### 8.7.6 Meta-regressions of volumes of hippocampus volumes of hippocampus by the percentage of participants on anti-AD medication at the baseline

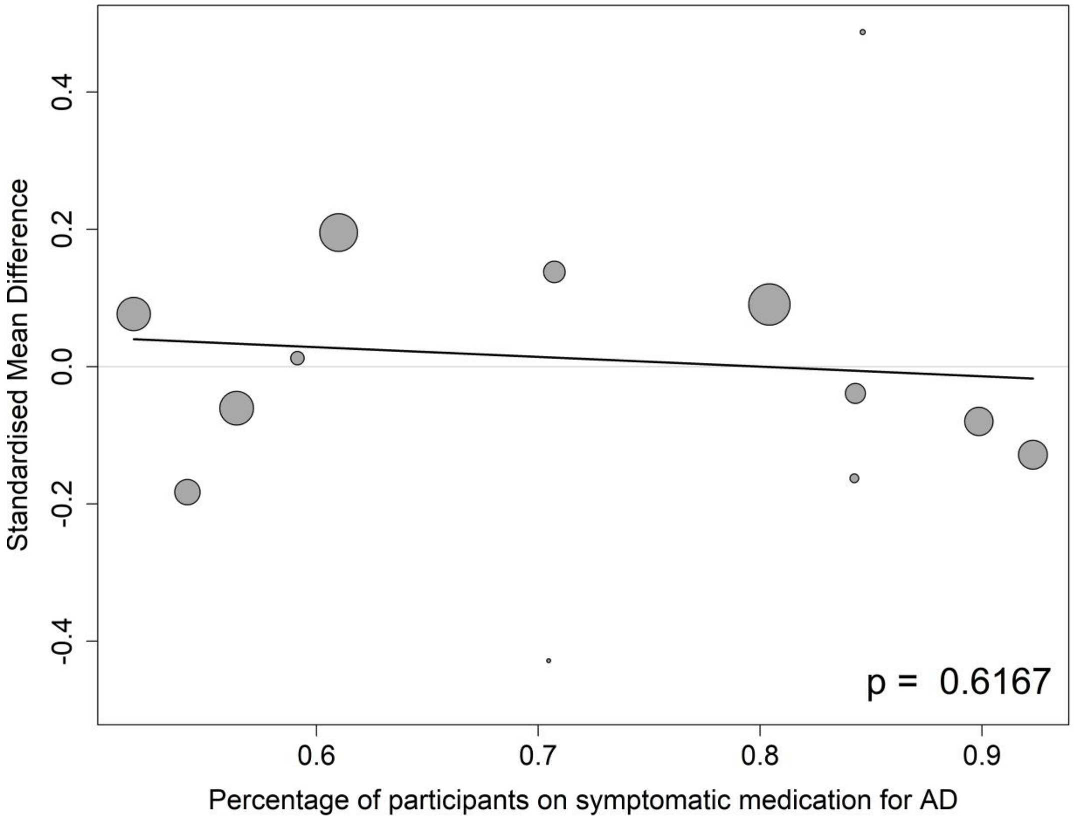

#### 8.8 Volumes of ventricle

##### 8.8.1 Meta-regressions of volumes of ventricle by baseline CDR-SB score

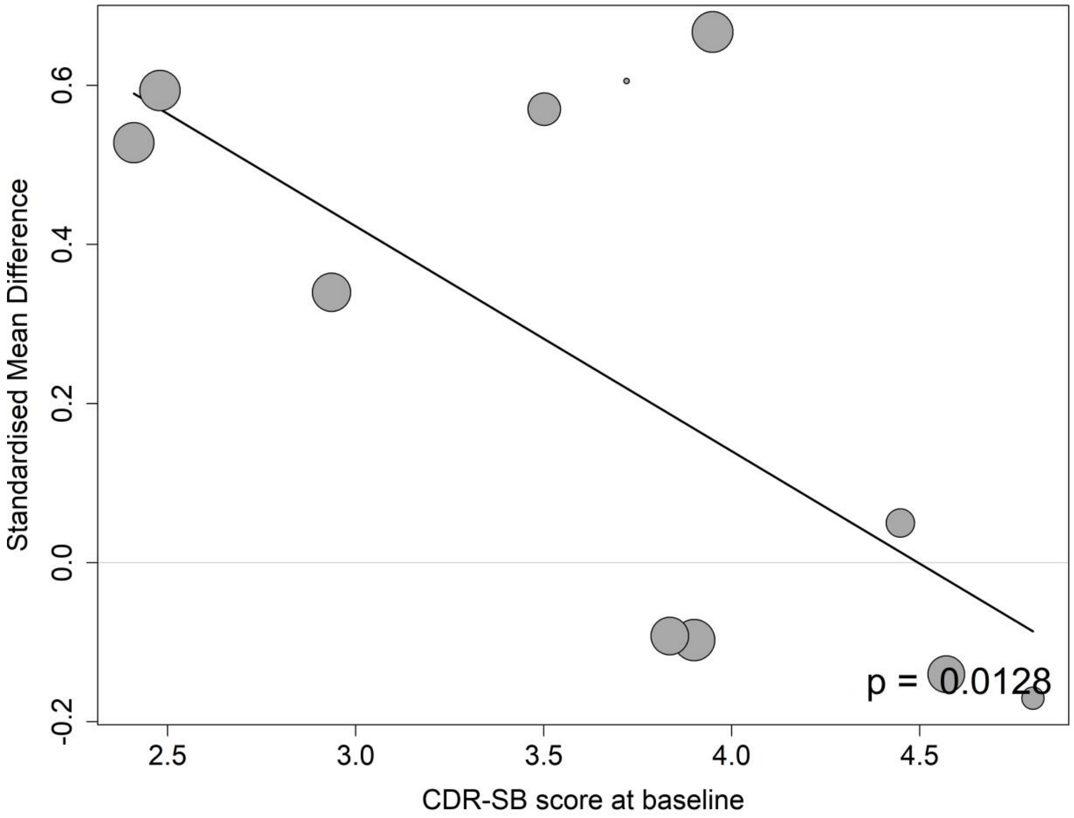

##### 8.8.2 Meta-regressions of volumes of ventricle by baseline MMSE score

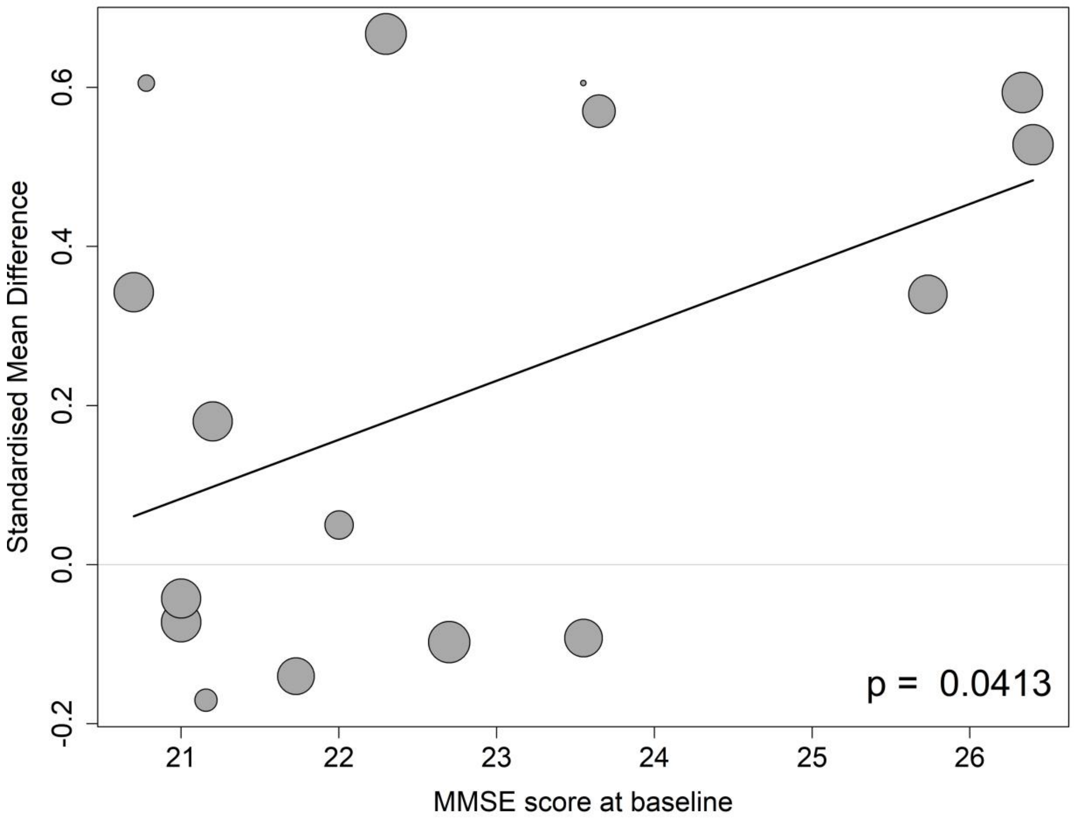

##### 8.8.3 Meta-regressions of volumes of ventricle by the mean age at the baseline

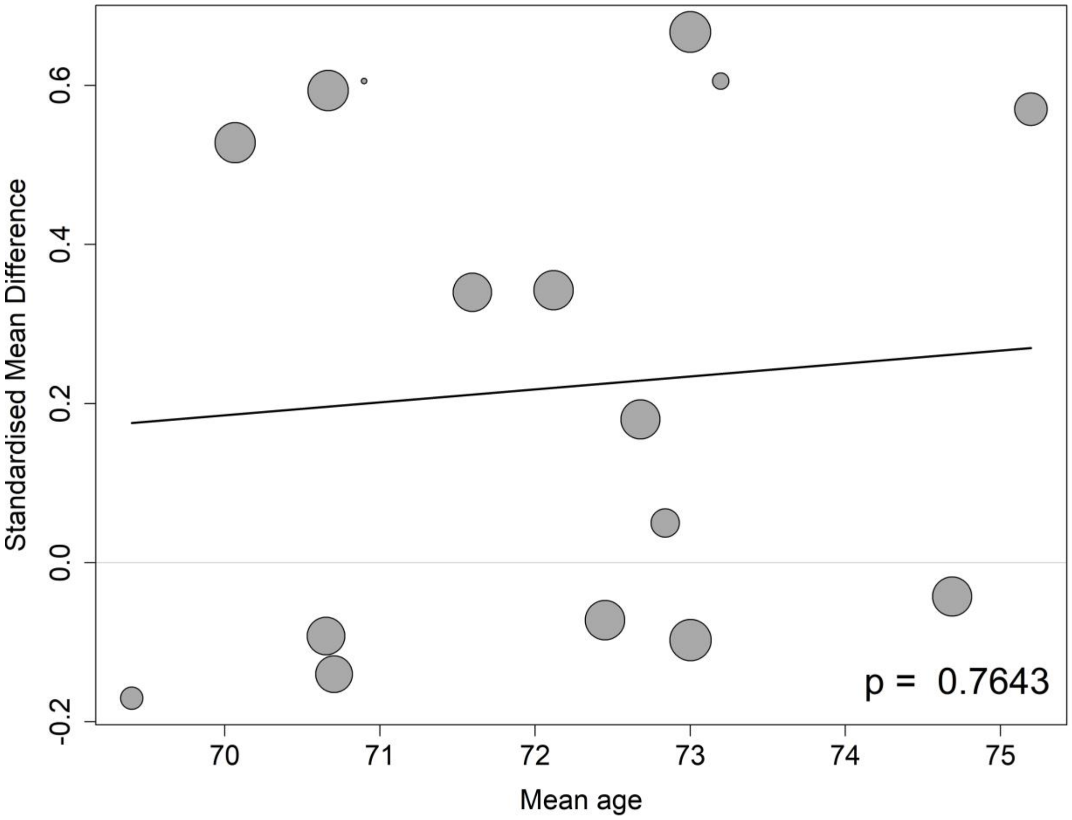

##### 8.8.4 Meta-regressions of volumes of ventricle by the percentage of APOE4 carrier at the baseline

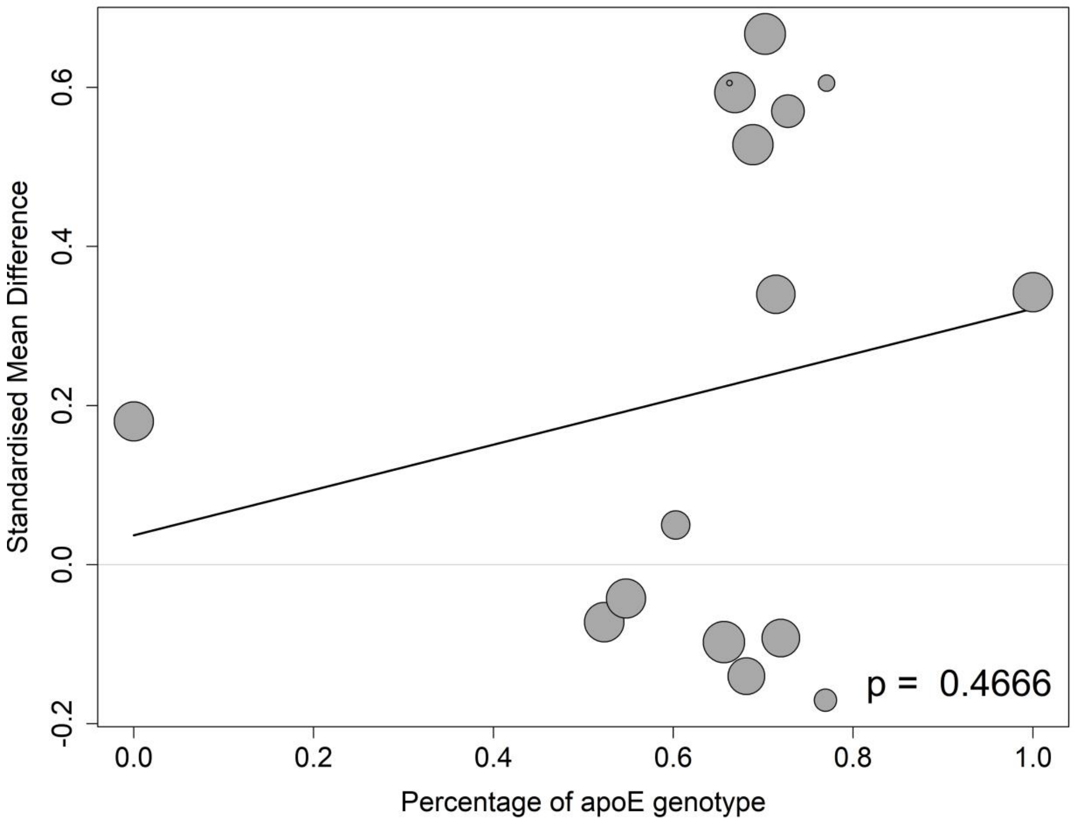

##### 8.8.5 Meta-regressions of volumes of ventricle by the percentage of female participants at the baseline

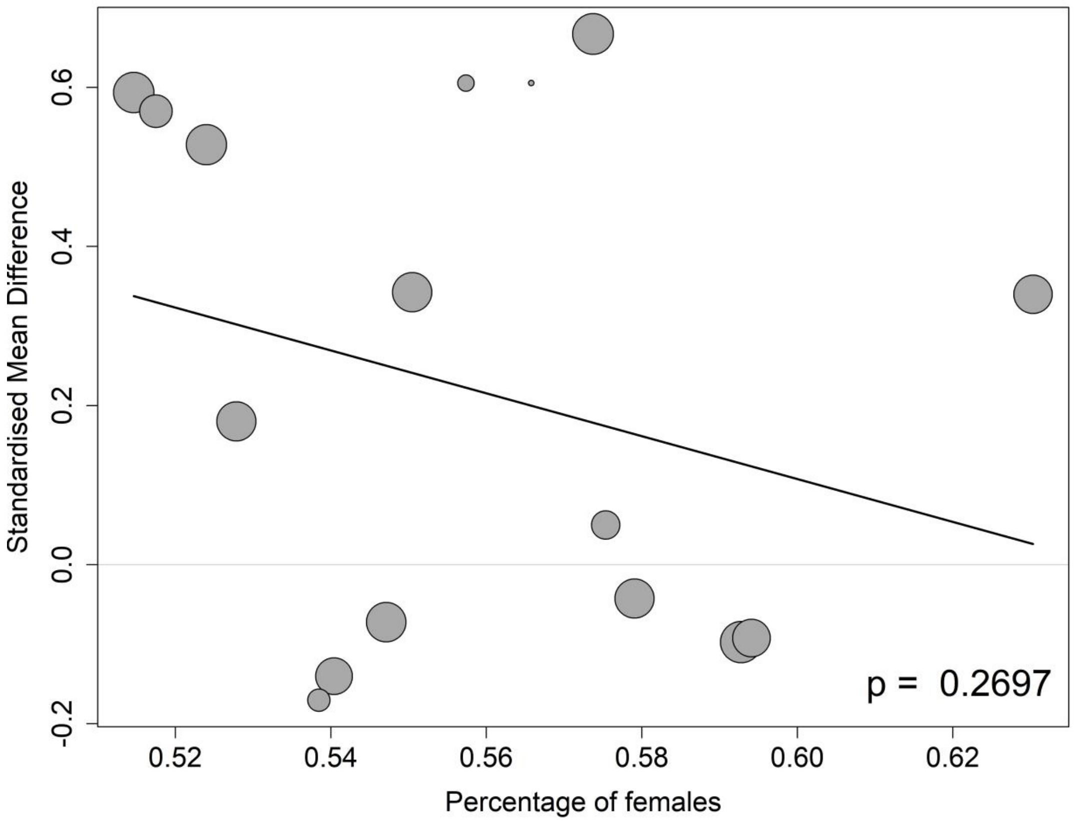

##### 8.8.6 Meta-regressions of volumes of ventricle by the percentage of participants on anti-AD medication at the baseline

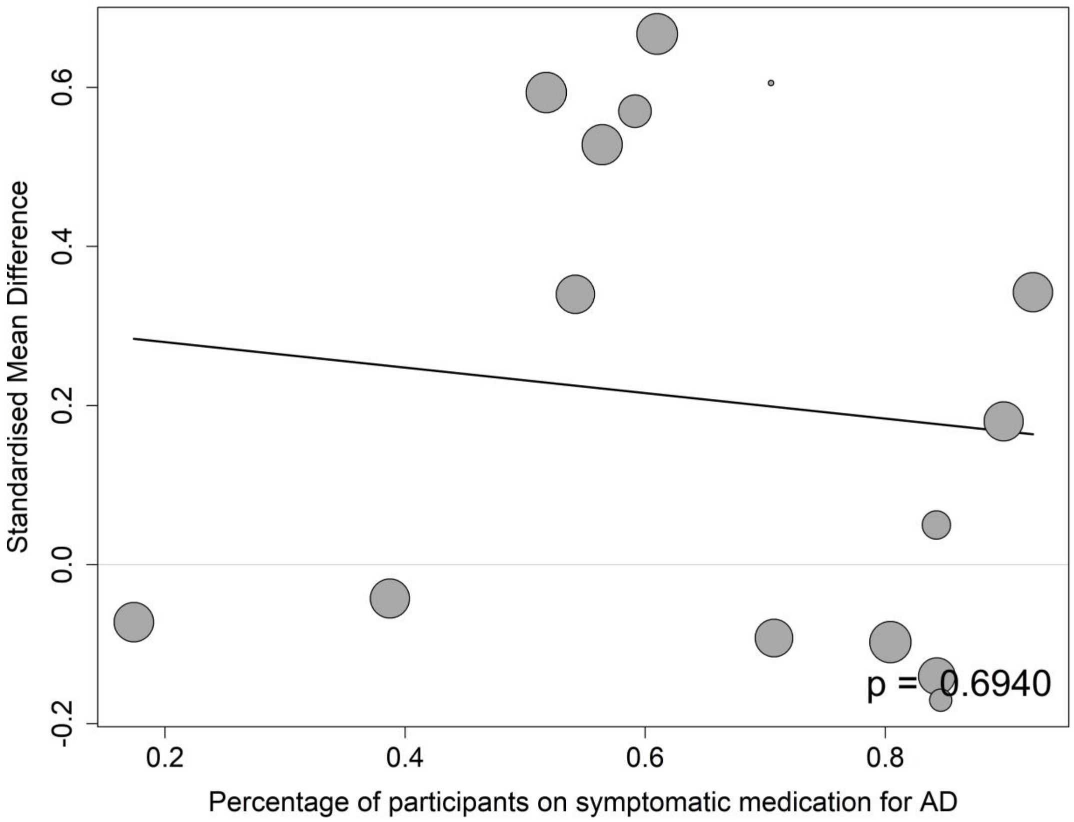

#### 8.9 CSF Aβ40

##### 8.9.1 Meta-regressions of CSF Aβ40 by baseline CDR-SB score

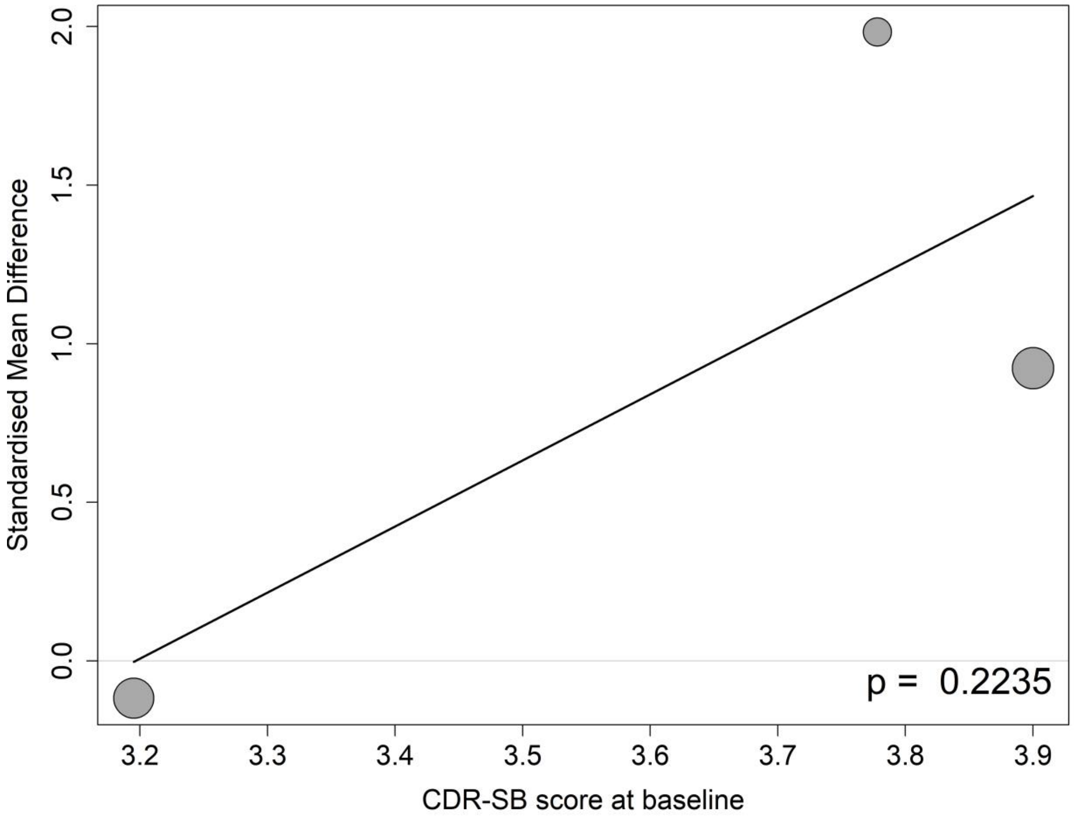

##### 8.9.2 Meta-regressions of CSF Aβ40 by baseline MMSE score

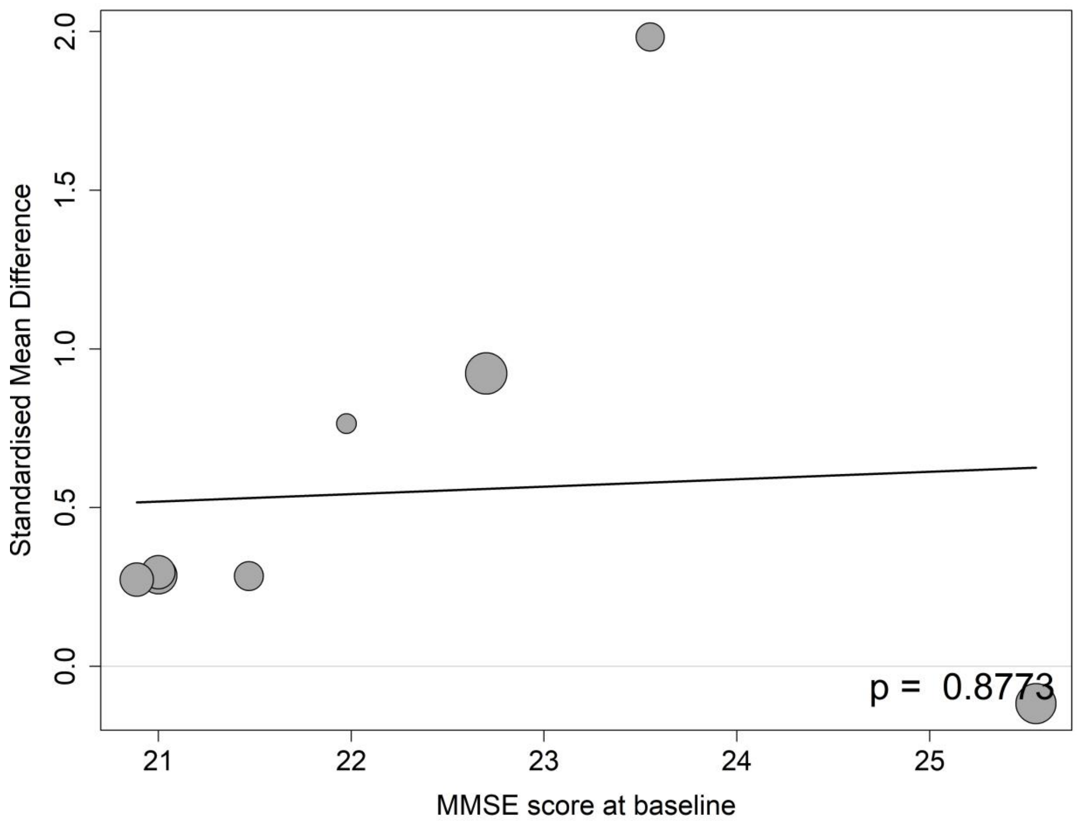

##### 8.9.3 Meta-regressions of CSF Aβ40 by the mean age at the baseline

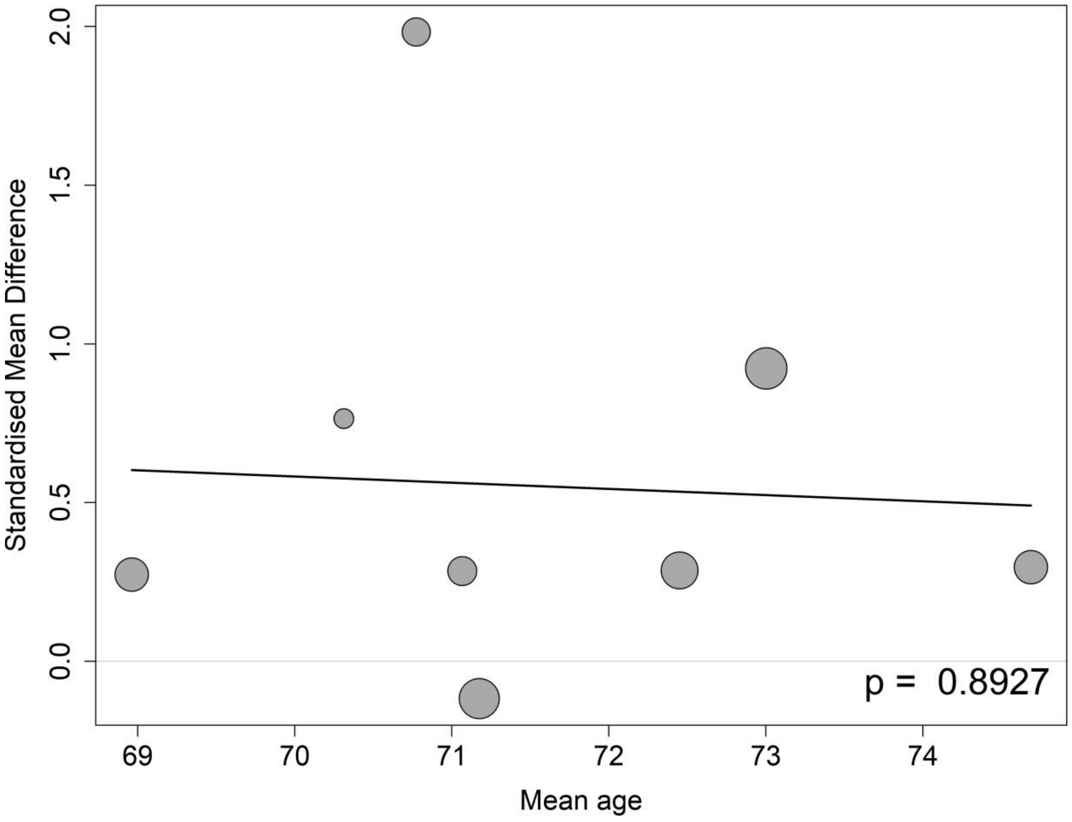

##### 8.9.4 Meta-regressions of CSF Aβ40 by the percentage of APOE4 carrier at the baseline

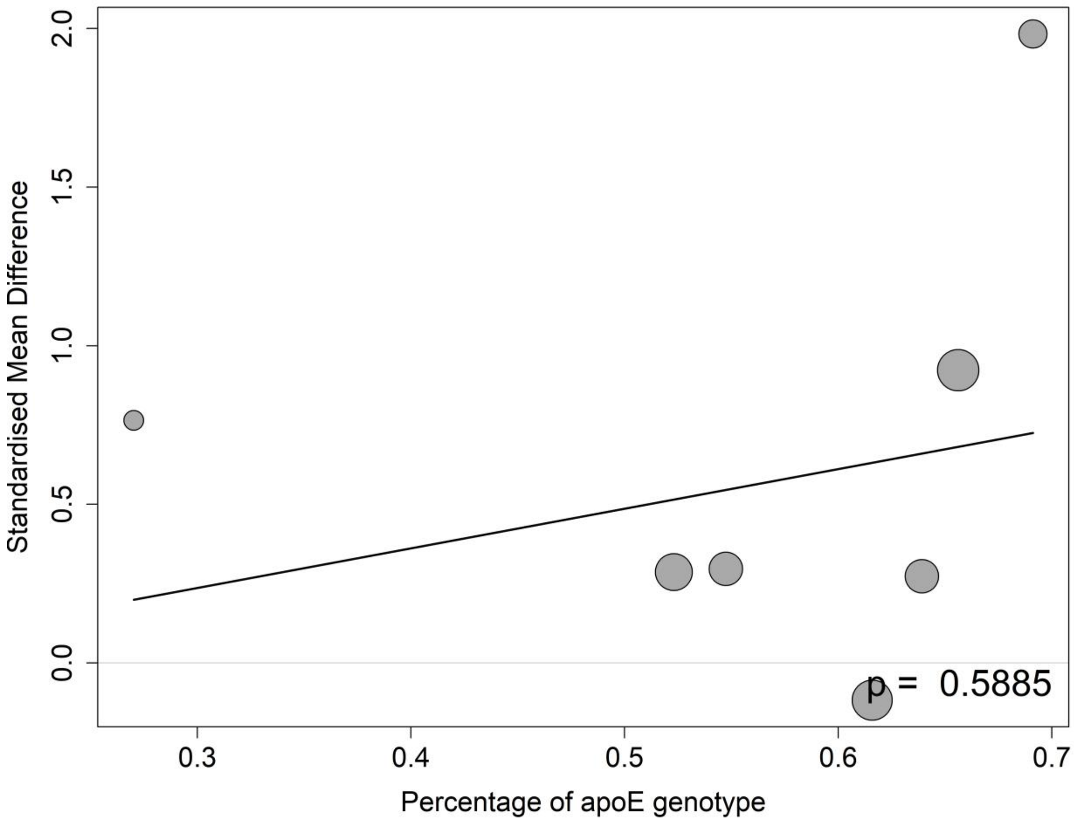

**8.9.5 Meta-regressions of CSF Aβ40 by the percentage of female participants at the baseline**

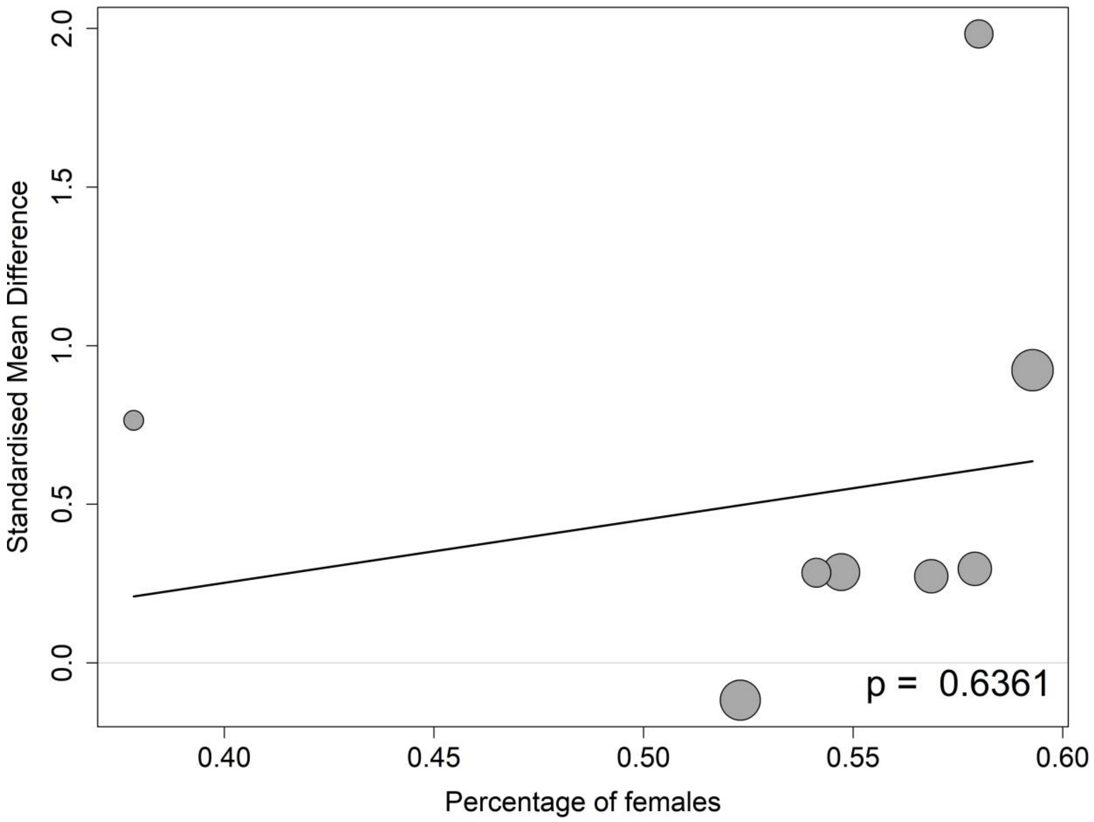

##### 8.9.6 Meta-regressions of CSF Aβ40 by the percentage of participants on anti-AD medication at the baseline

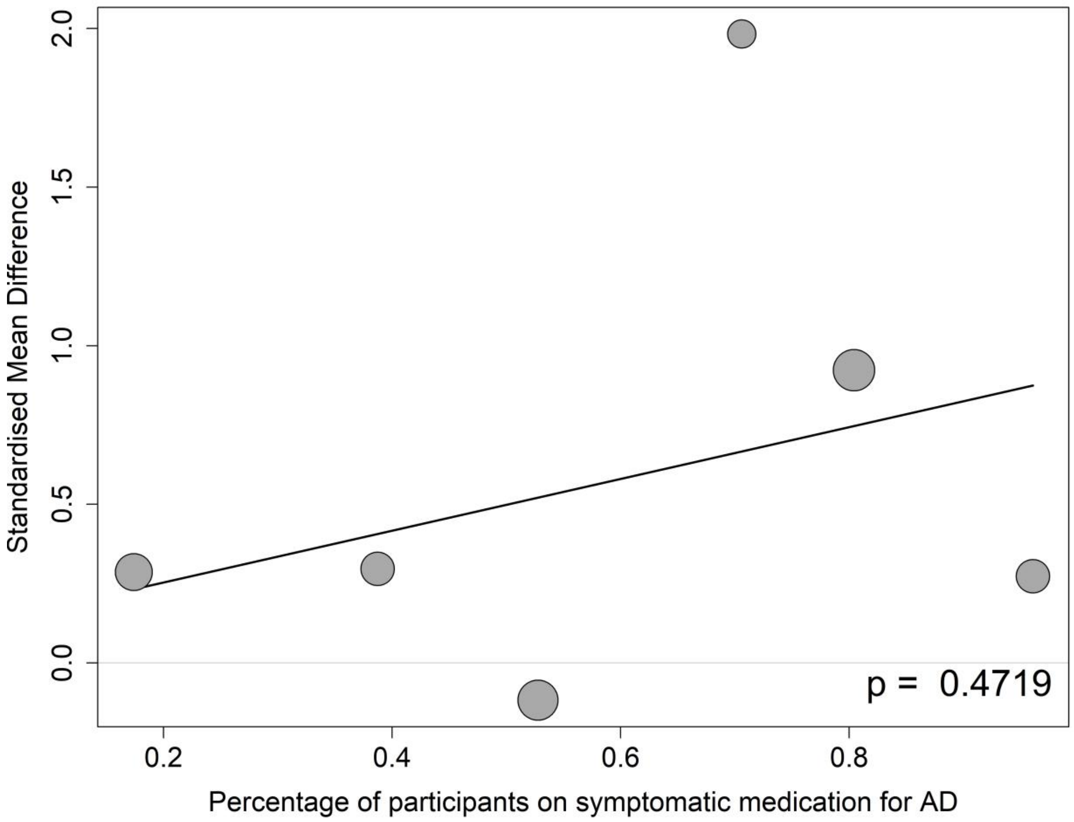

#### 8.10 CSF Aβ42

##### 8.10.1 Meta-regressions of CSF Aβ42 by baseline CDR-SB score

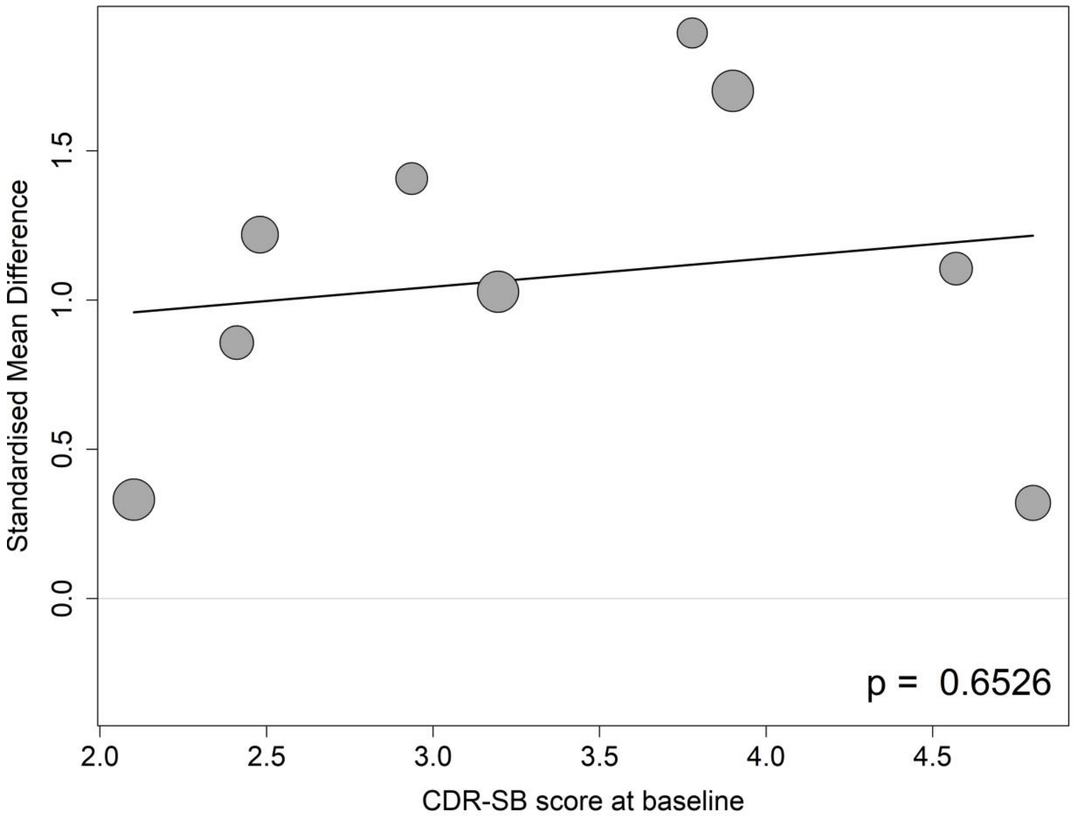

##### 8.10.2 Meta-regressions of CSF Aβ42 by baseline MMSE score

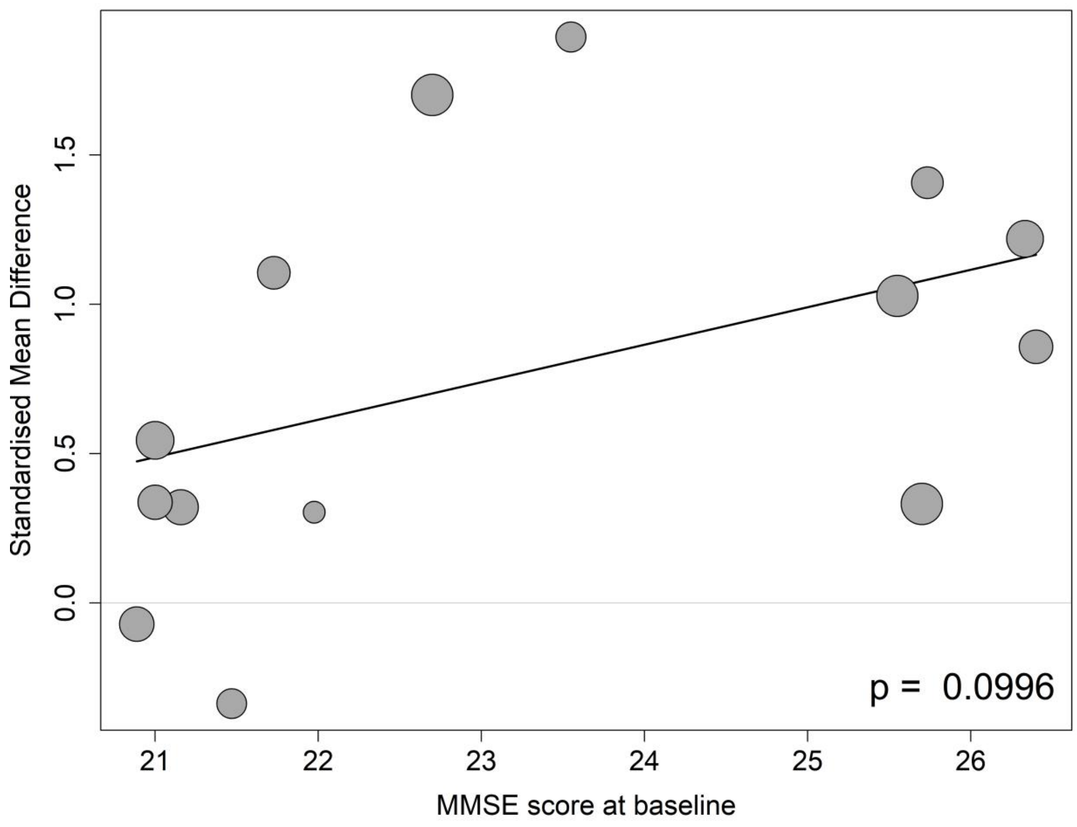

**8.10.3 Meta-regressions of CSF Aβ42 by the mean age at the baseline**

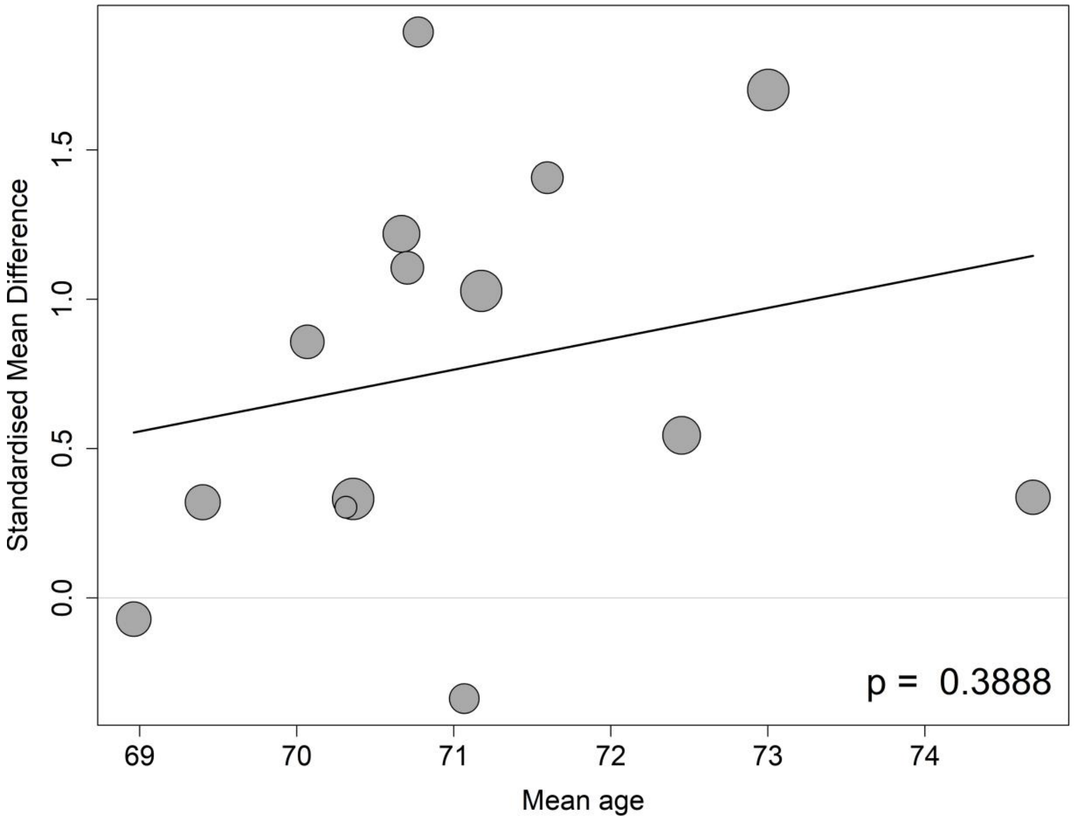

##### 8.10.4 Meta-regressions of CSF Aβ42 by the percentage of APOE4 carrier at the baseline

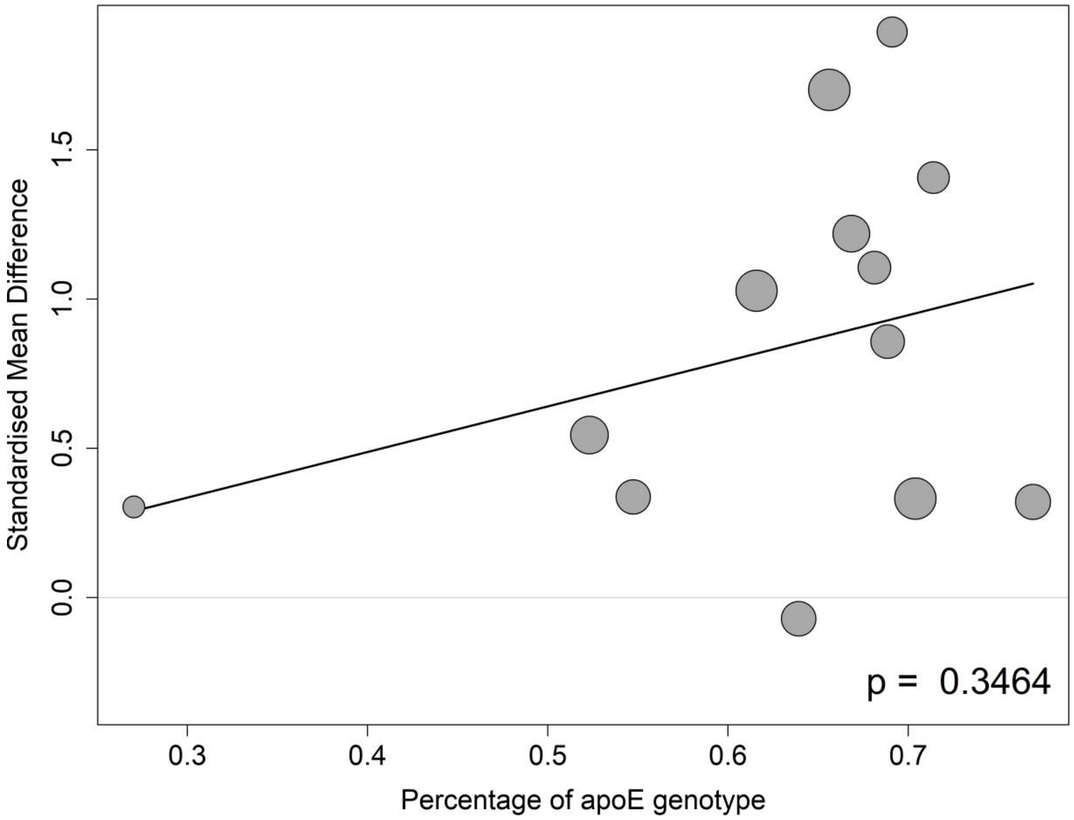

##### 8.10.5 Meta-regressions of CSF Aβ42 by the percentage of female participants at the baseline

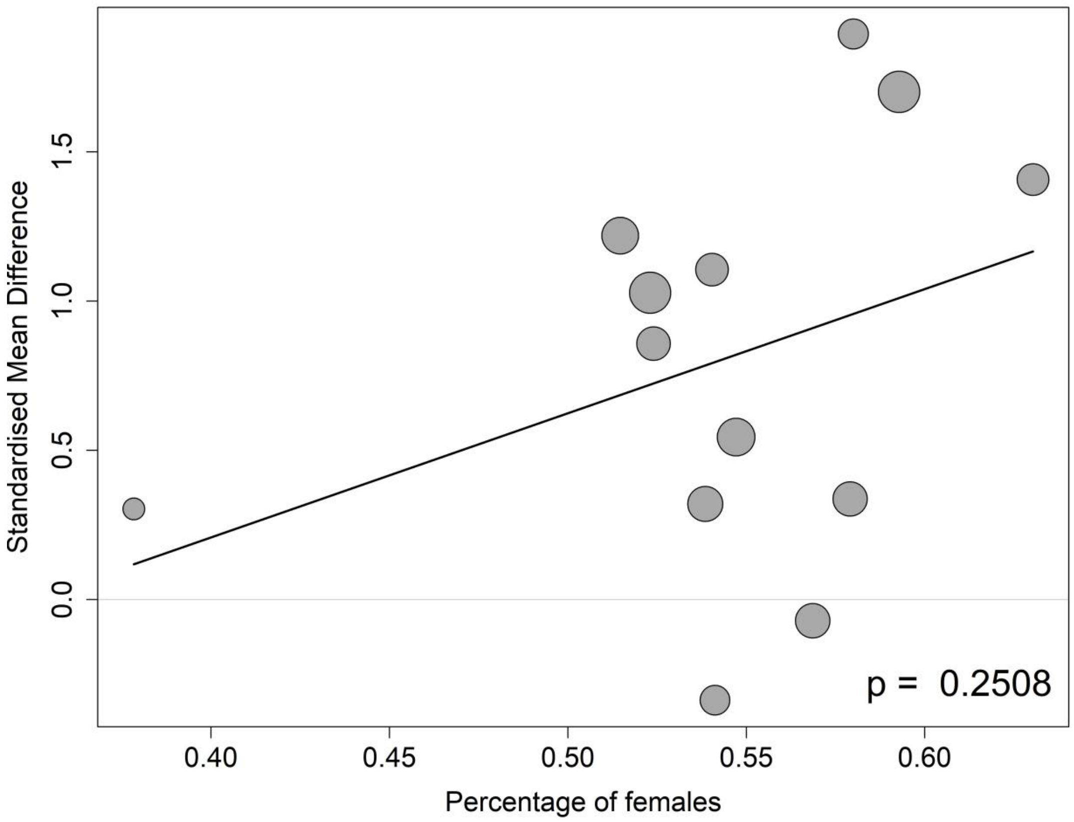

##### 8.10.6 Meta-regressions of CSF Aβ42 by the percentage of participants on anti-AD medication at the baseline

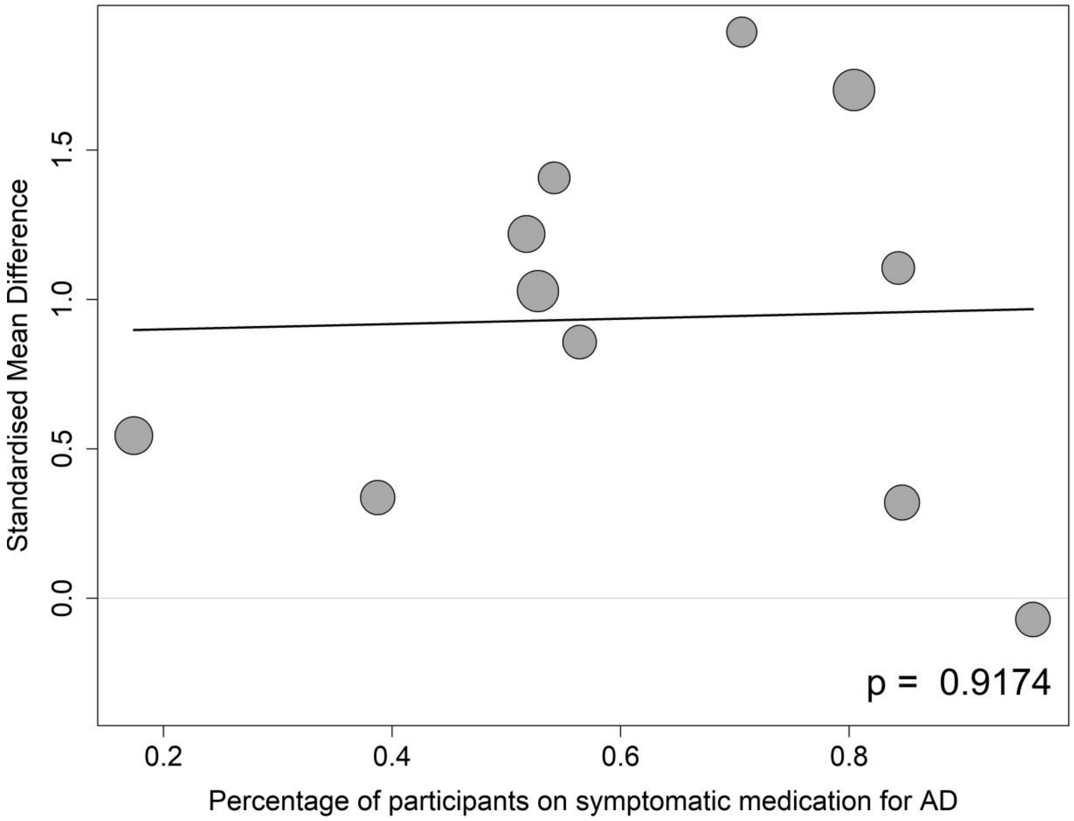

#### 8.11 CSF p-tau

##### 8.11.1 Meta-regressions of CSF p-tau by baseline CDR-SB score

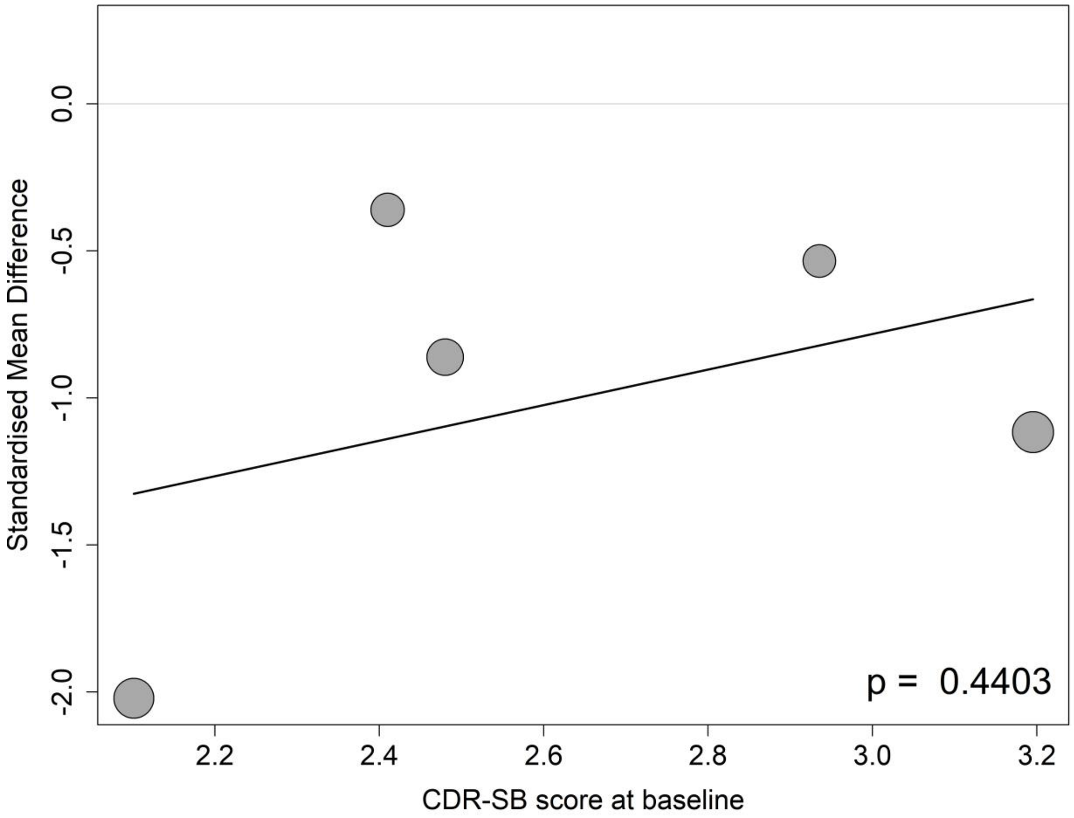

##### 8.11.2 Meta-regressions of CSF p-tau by baseline MMSE score

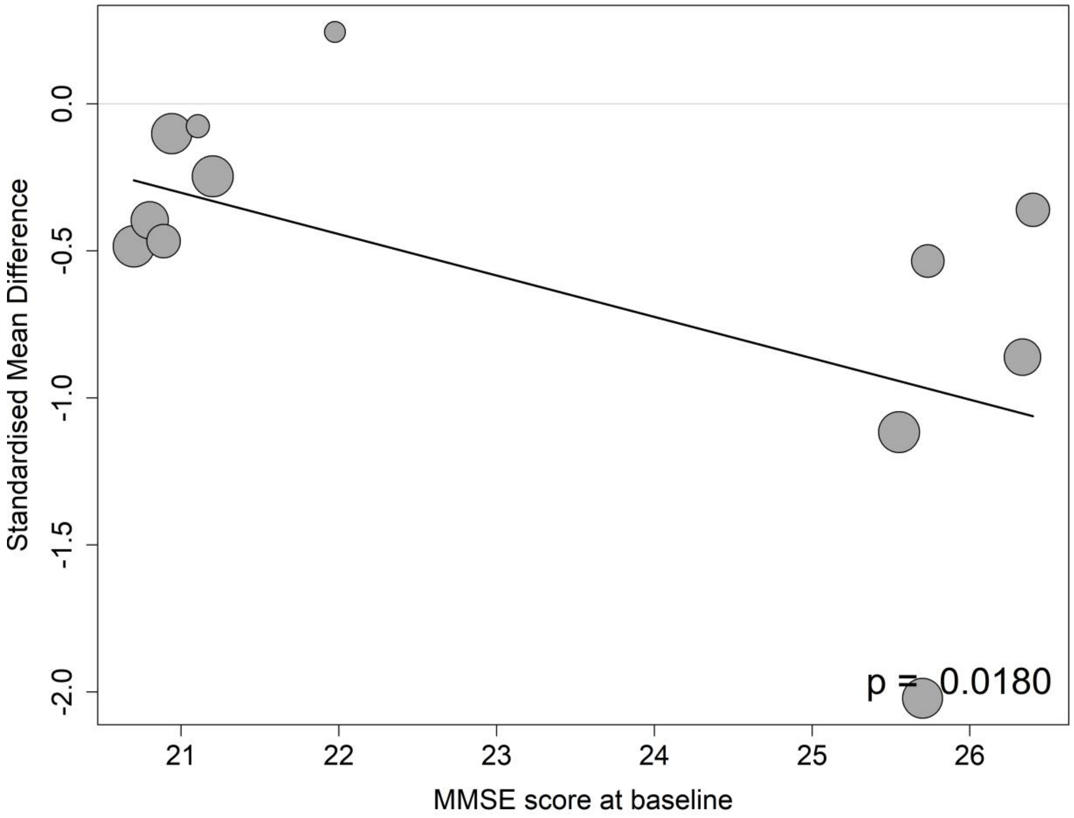

##### 8.11.3 Meta-regressions of CSF p-tau by the mean age at the baseline

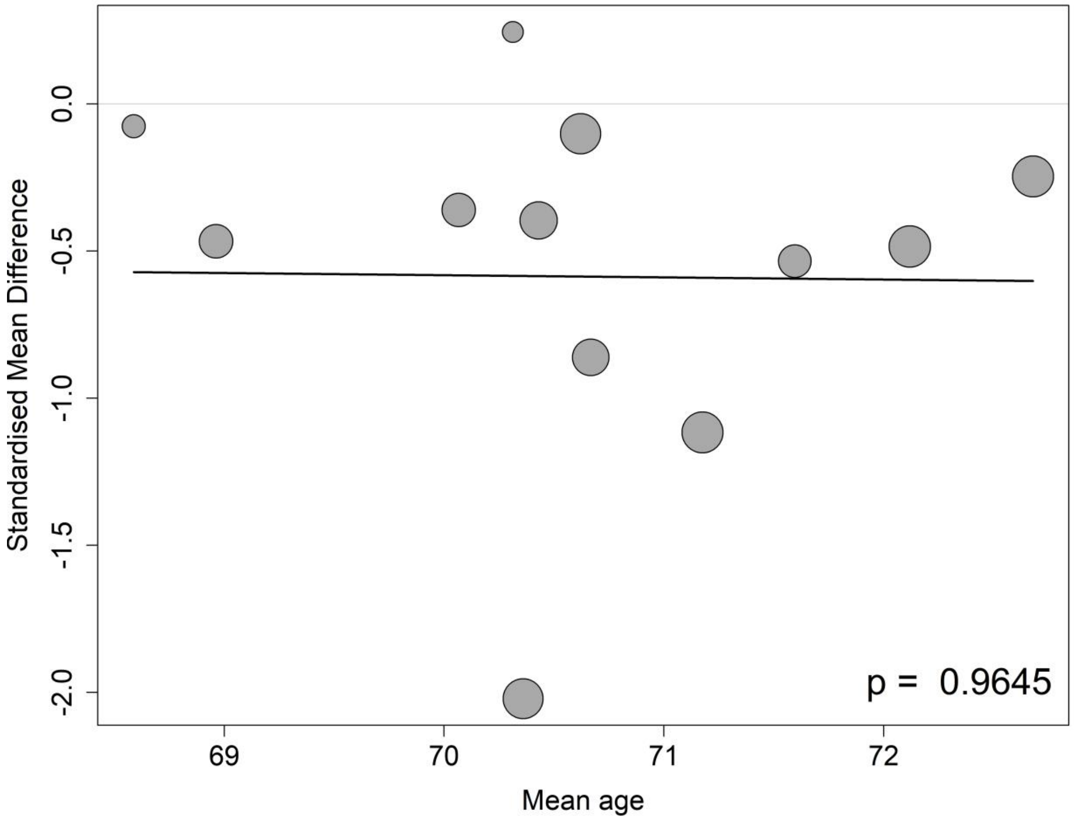

**8.11.4 Meta-regressions of CSF p-tau by the percentage of APOE4 carrier at the baseline**

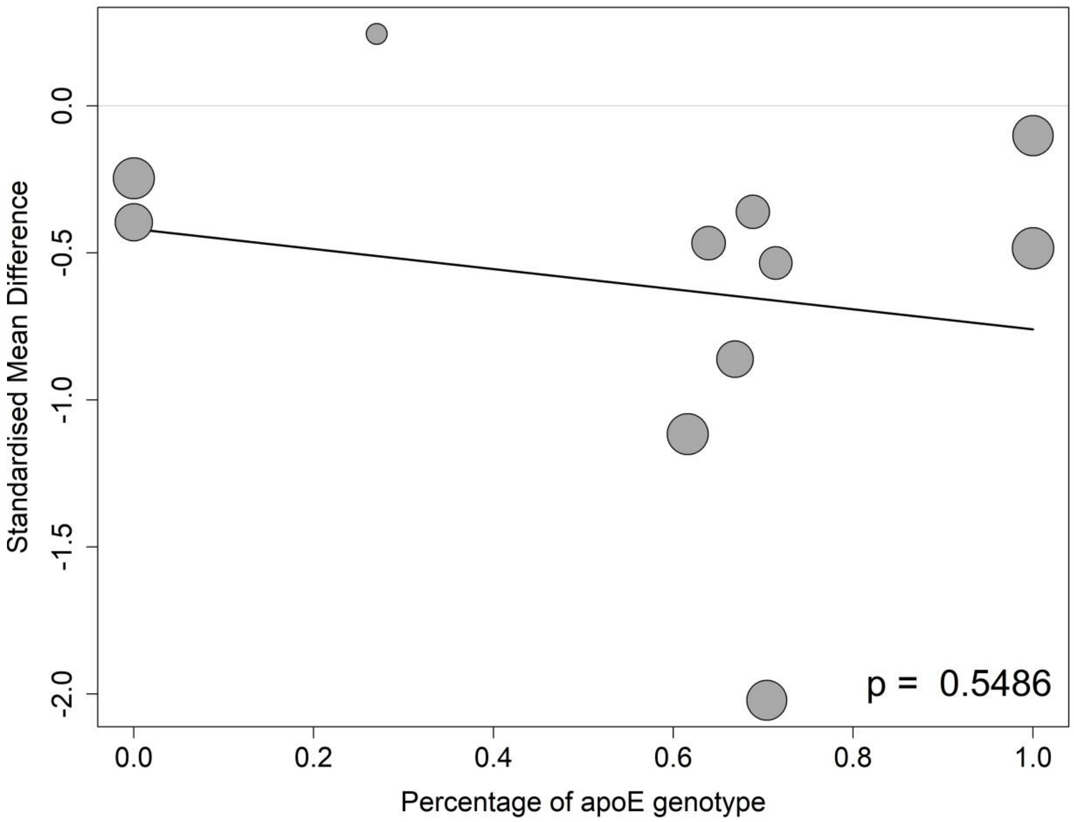

##### 8.11.5 Meta-regressions of CSF p-tau by the percentage of female participants at the baseline

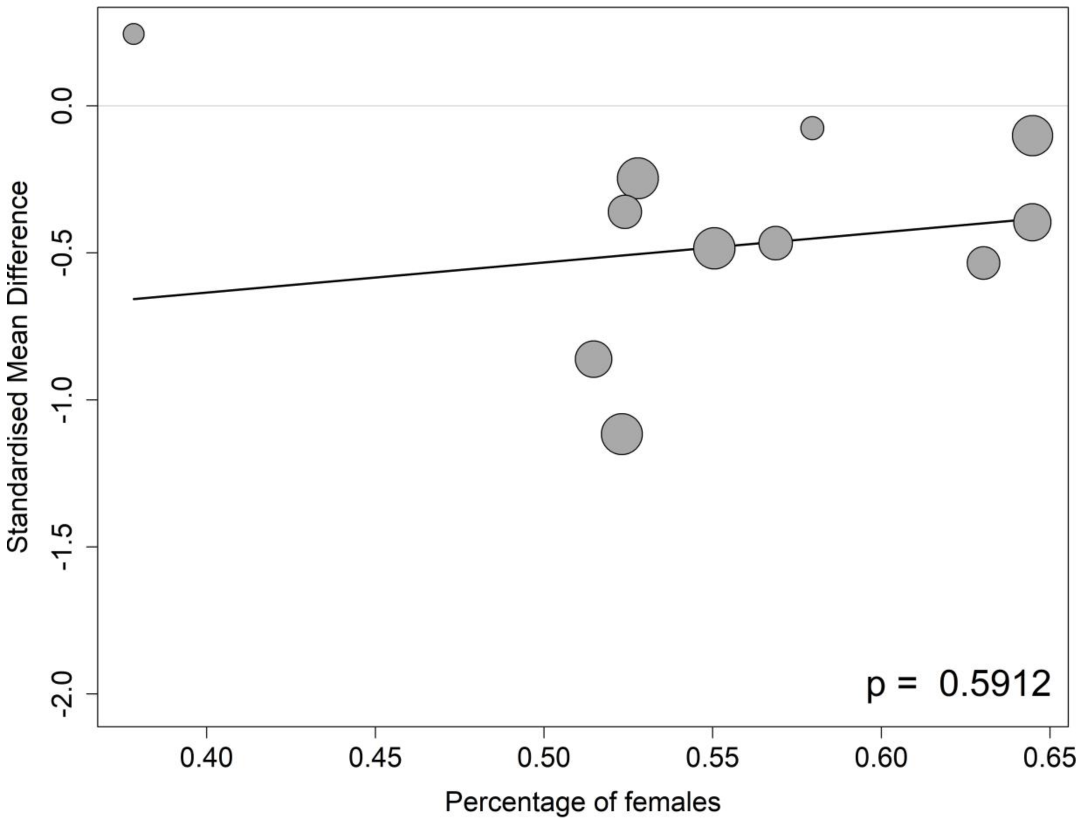

**8.11.6 Meta-regressions of CSF p-tau by the percentage of participants on anti-AD medication at the baseline**

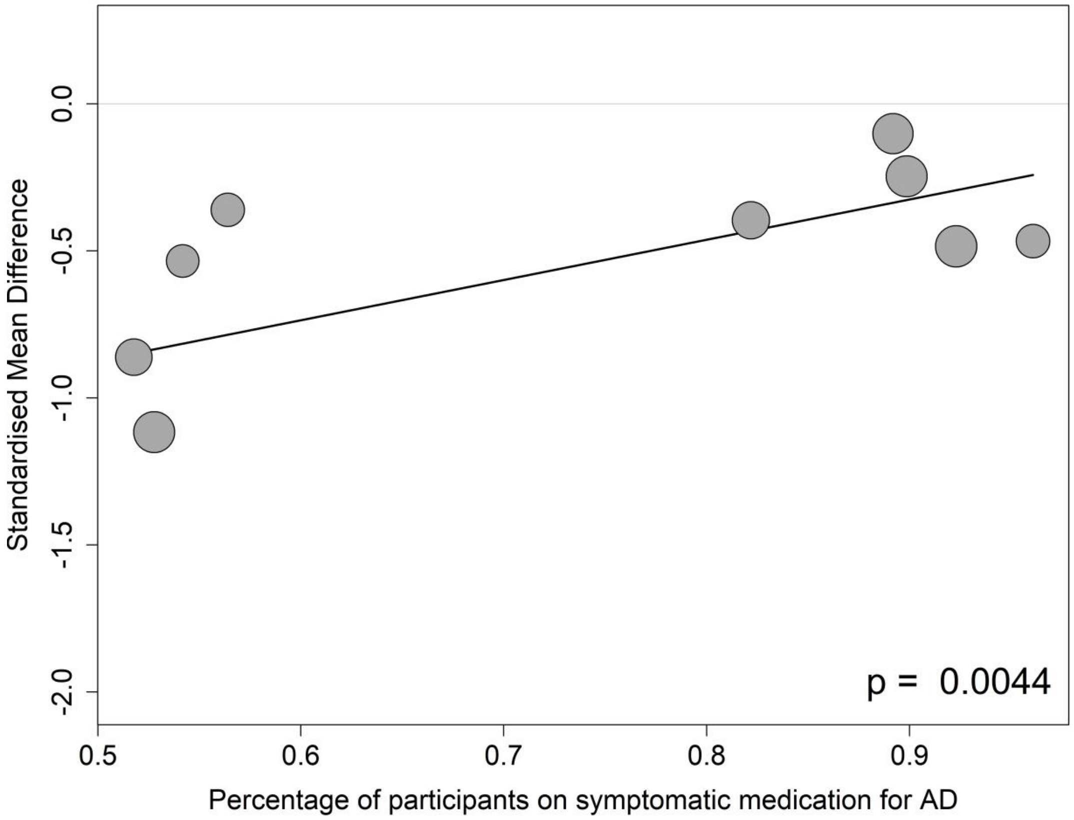

#### 8.12 CSF total tau

##### 8.12.1 Meta-regressions of CSF total tau by baseline CDR-SB score

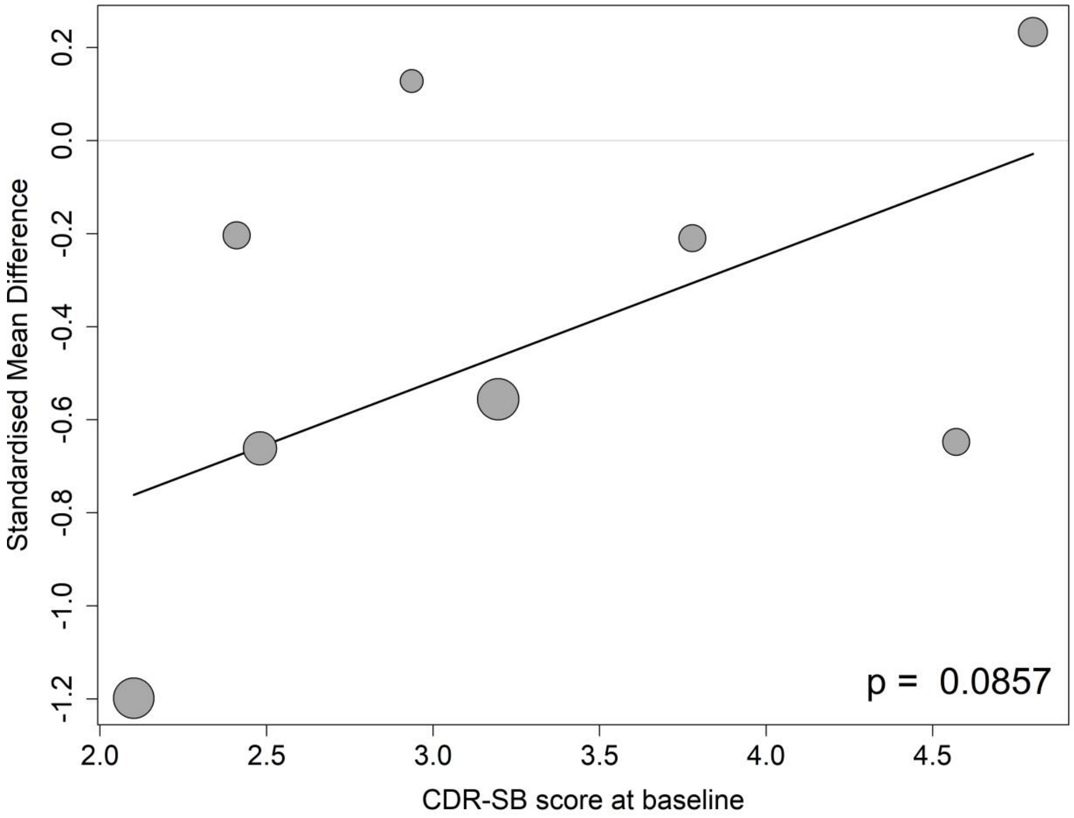

##### 8.12.2 Meta-regressions of CSF total tau by baseline MMSE score

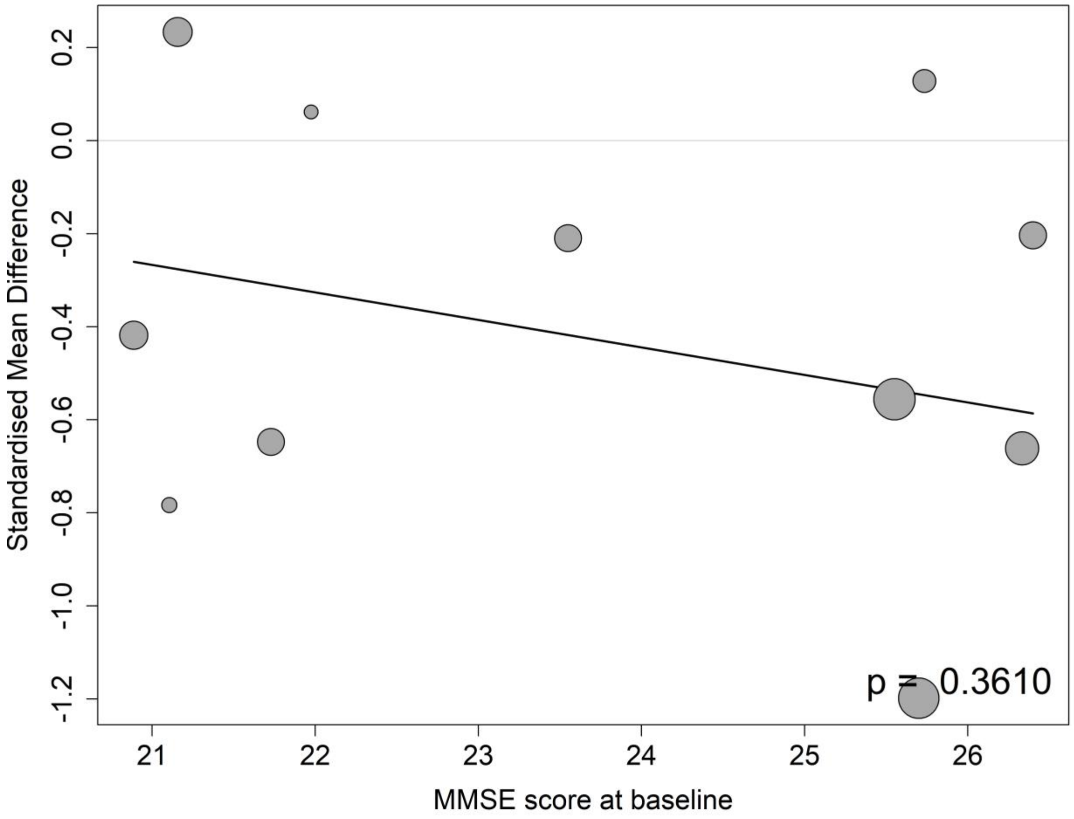

##### 8.12.3 Meta-regressions of CSF total tau by the mean age at the baseline

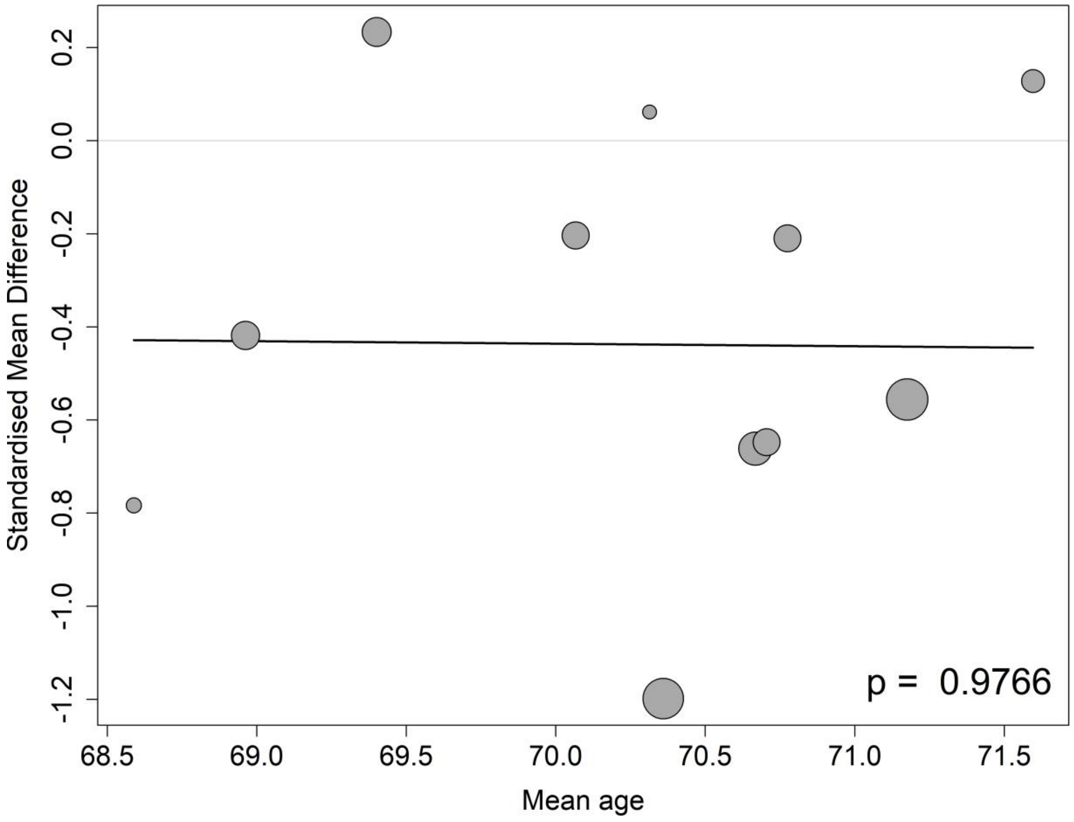

##### 8.12.4 Meta-regressions of CSF total tau by the percentage of APOE4 carrier at the baseline

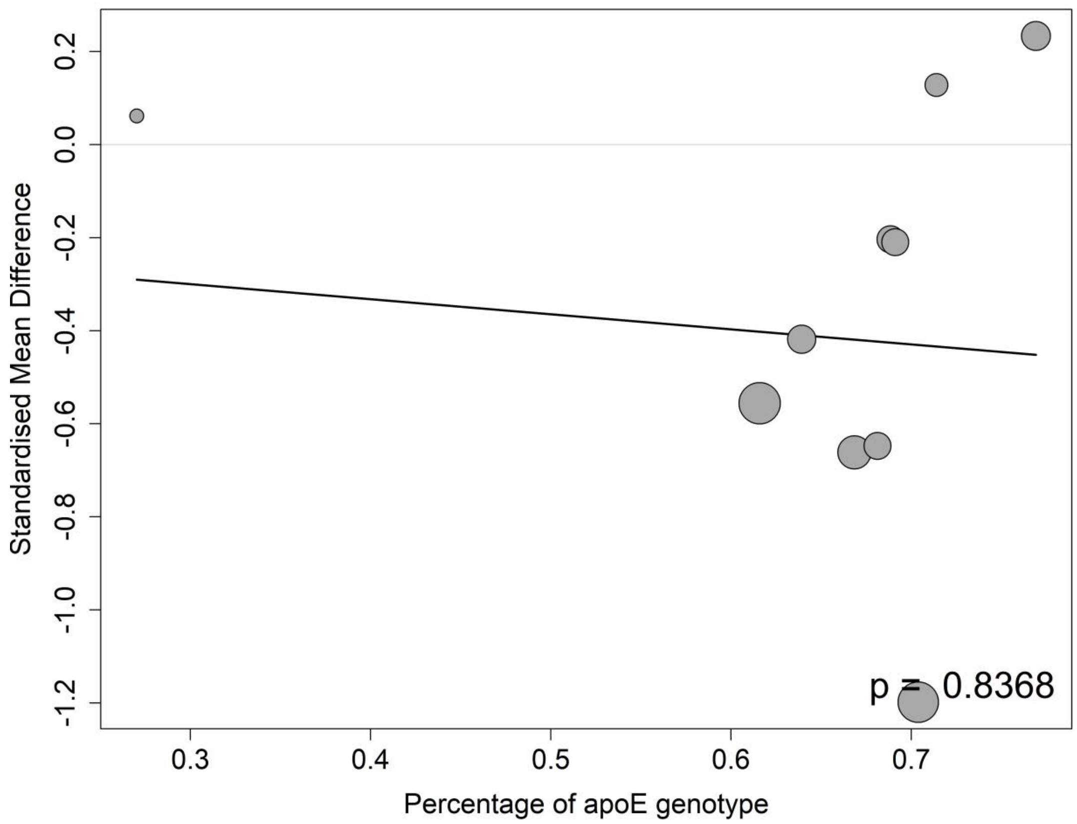

##### 8.12.5 Meta-regressions of CSF total tau by the percentage of female participants at the baseline

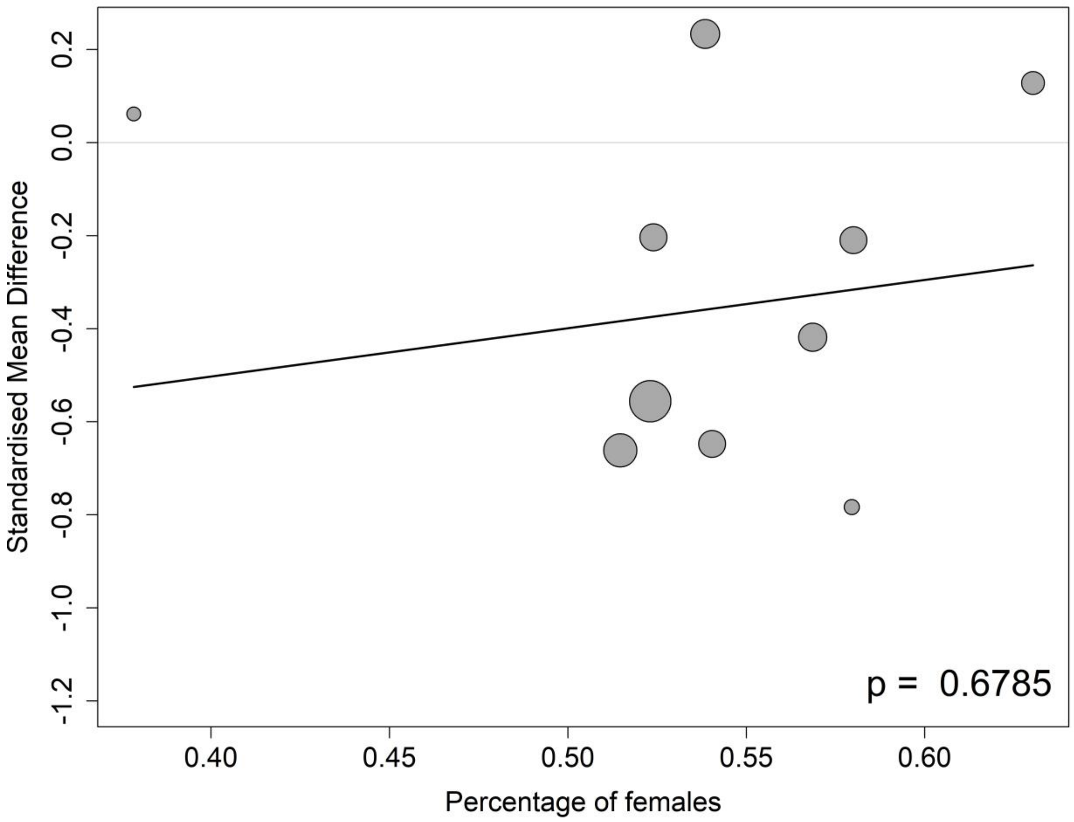

##### 8.12.6 Meta-regressions of CSF total tau by the percentage of participants on anti-AD medication at the baseline

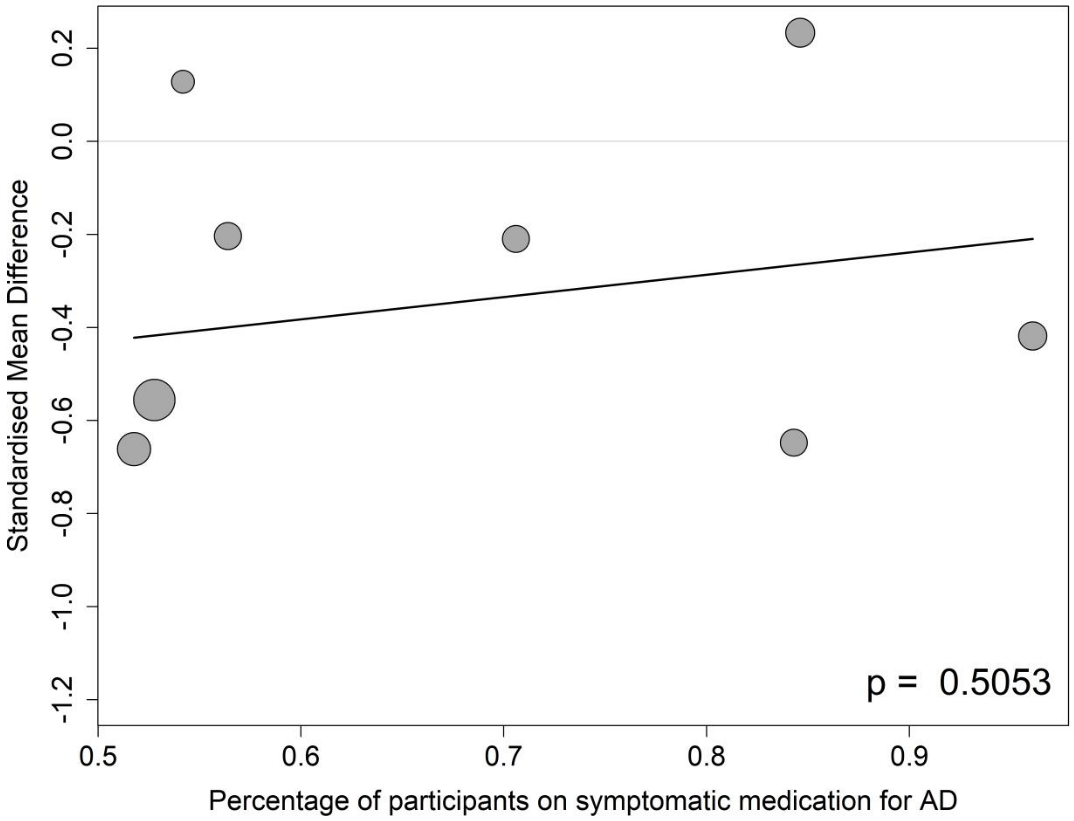

#### 8.13 CSF p-tau181

##### 8.13.1 Meta-regressions of CSF p-tau181 by baseline CDR-SB score

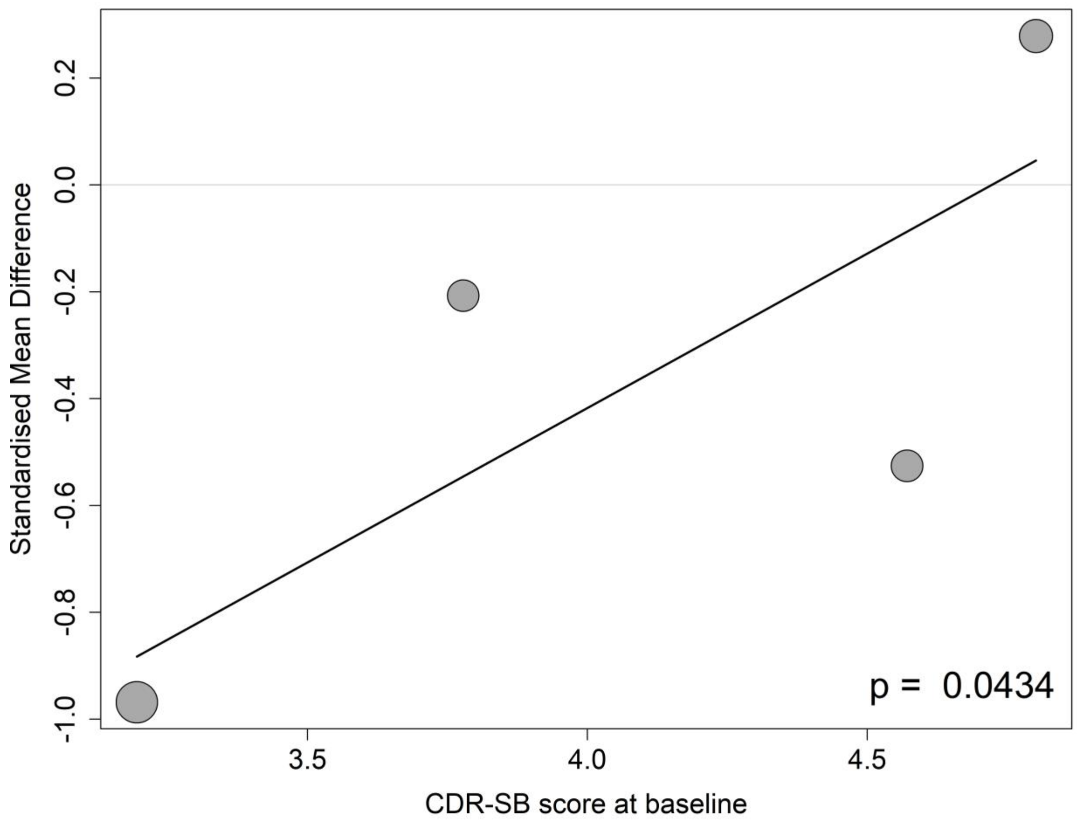

##### 8.13.2 Meta-regressions of CSF p-tau181 by baseline MMSE score

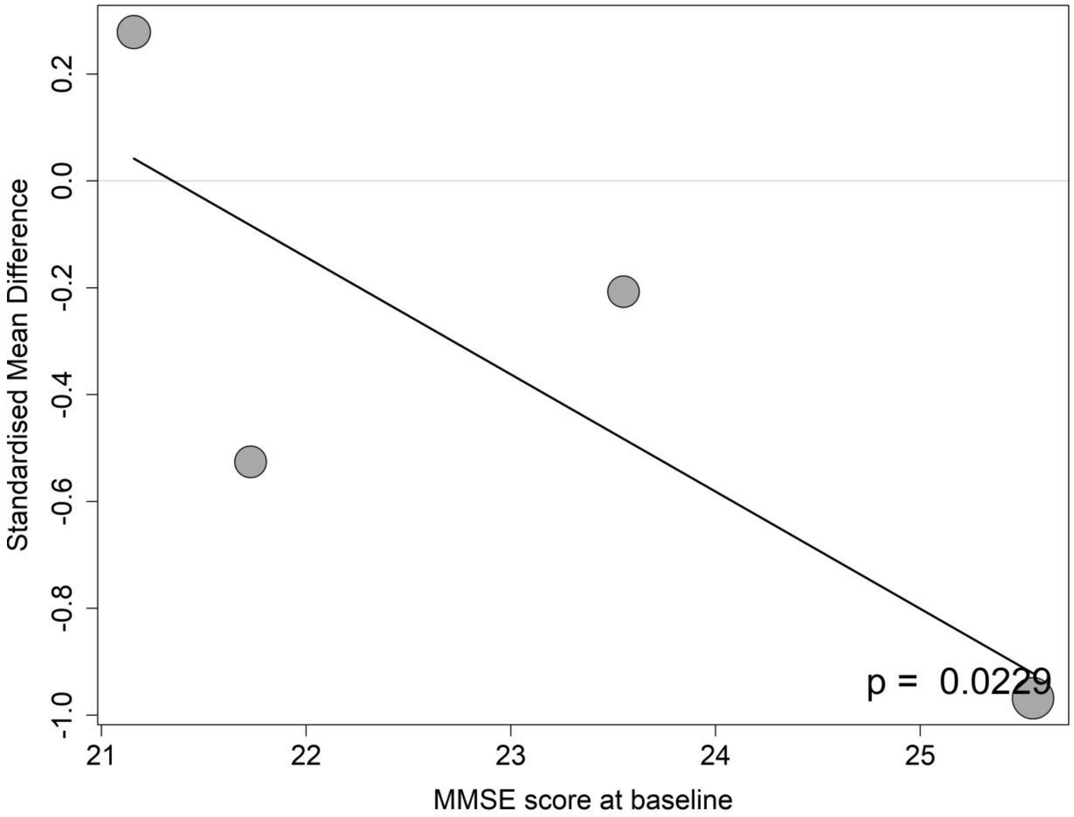

**8.13.3 Meta-regressions of CSF p-tau181 by the mean age at the baseline**

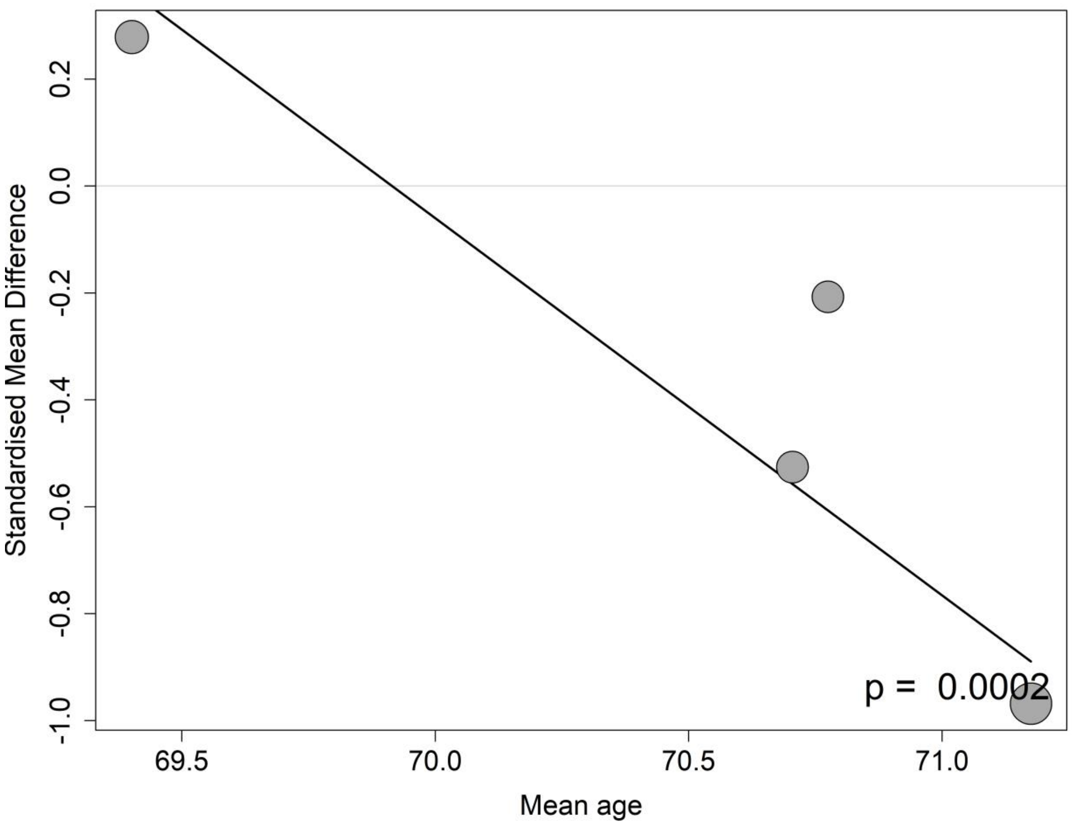

**8.13.4 Meta-regressions of CSF p-tau181 by the percentage of APOE4 carrier at the baseline**

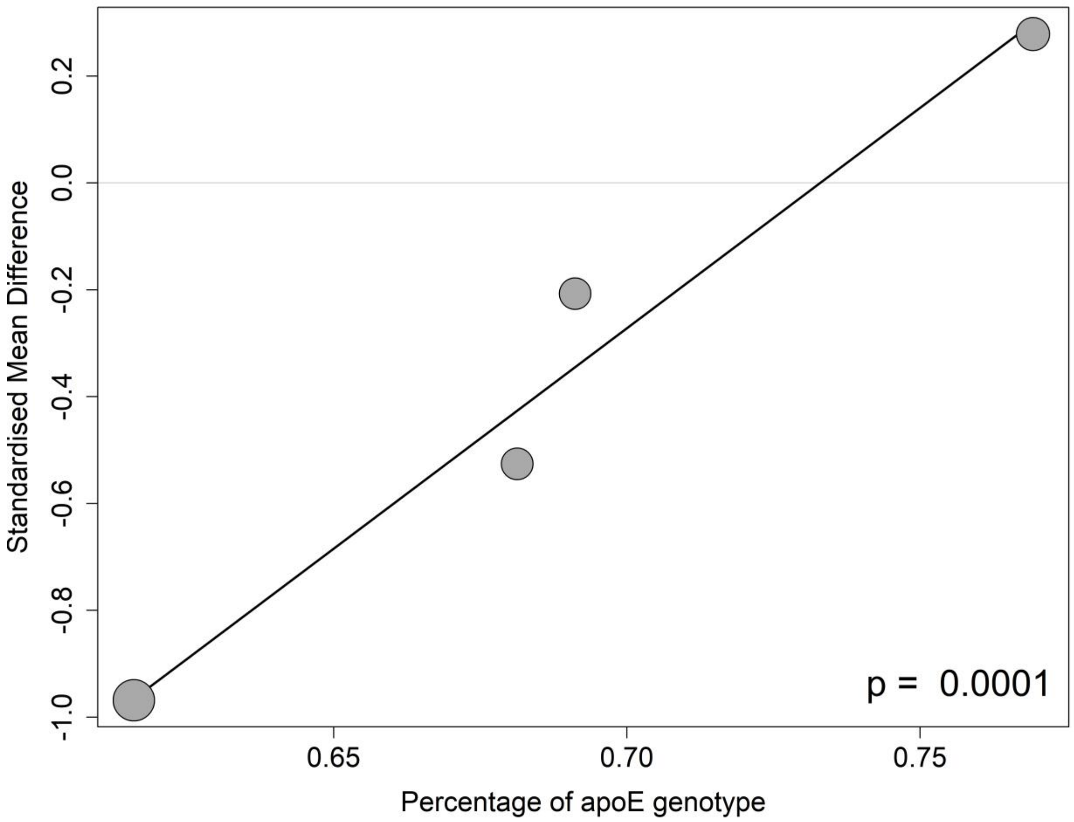

**8.13.5 Meta-regressions of CSF p-tau181 by the percentage of female participants at the baseline**

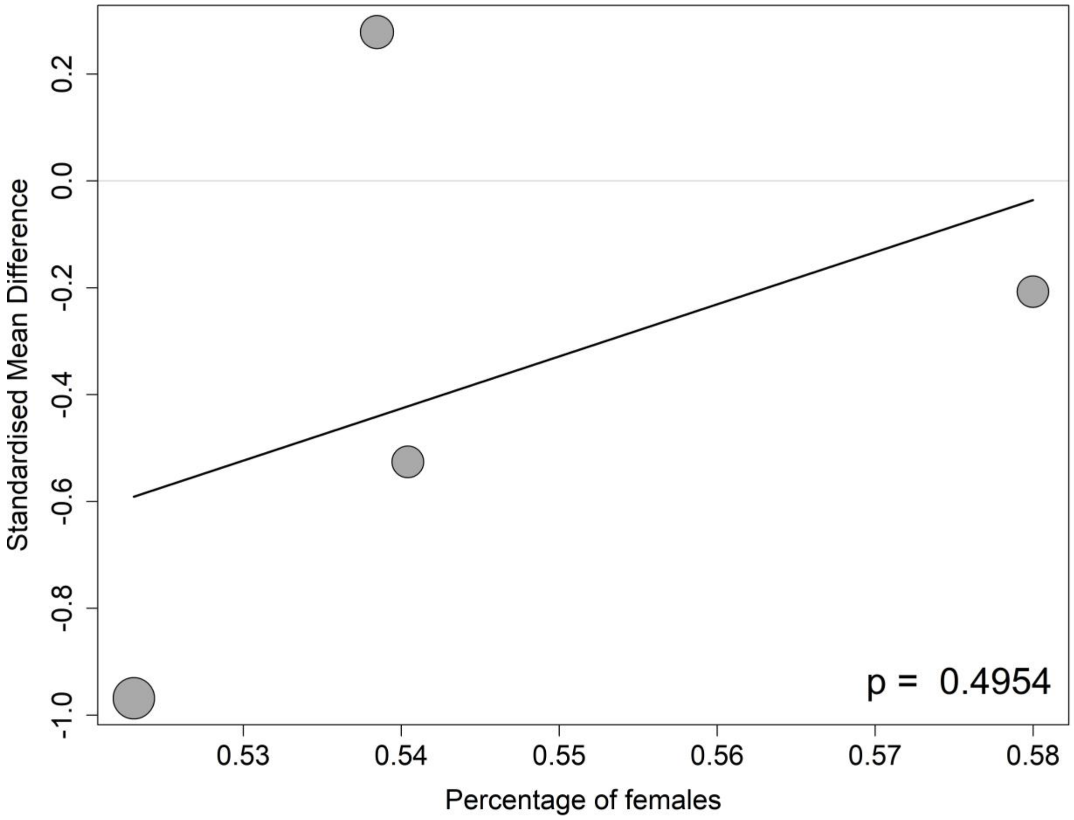

##### 8.13.6 Meta-regressions of CSF p-tau181 by the percentage of participants on anti-AD medication at the baseline

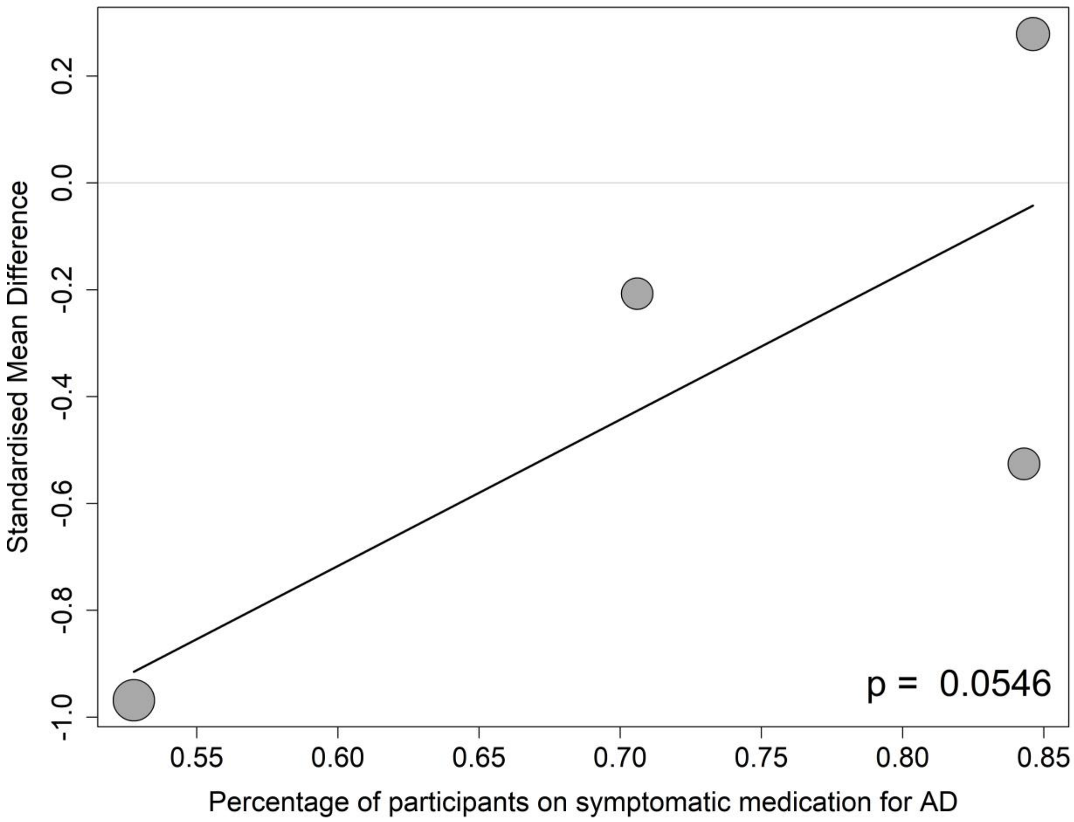

#### 8.14 CSF Nfl

##### 8.14.1 Meta-regressions of CSF Nfl by baseline CDR-SB score

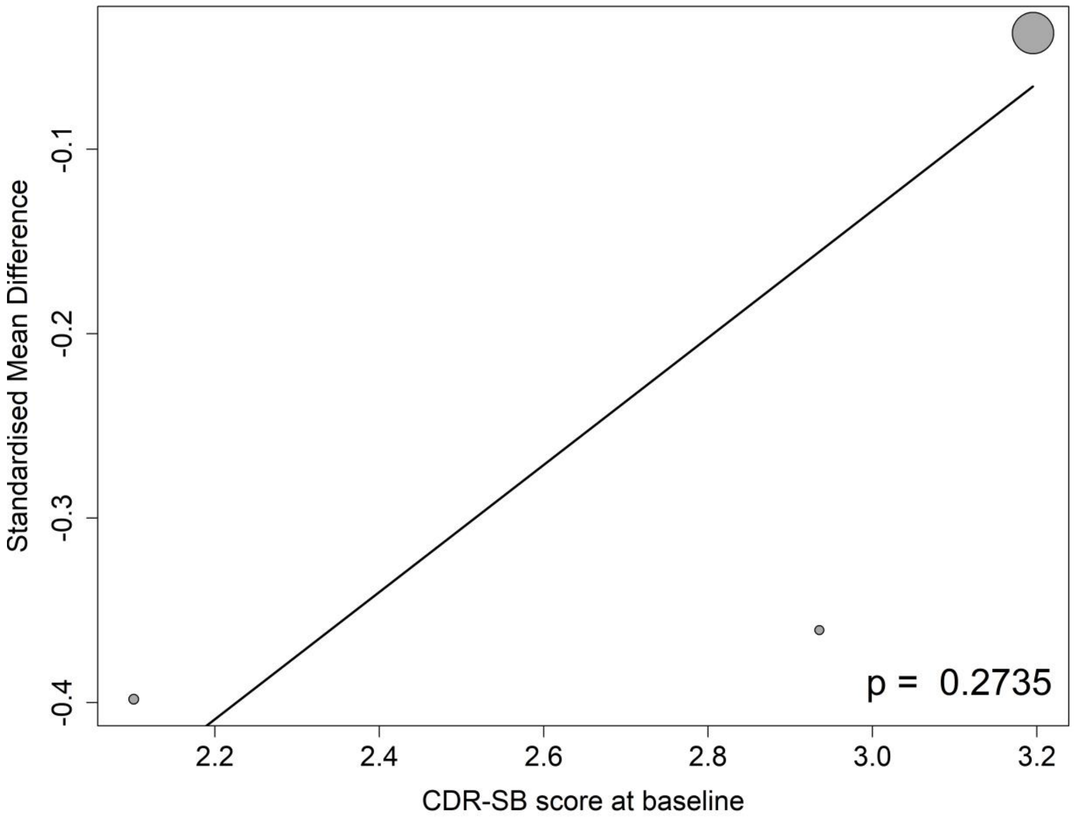

##### 8.14.2 Meta-regressions of CSF Nfl by baseline MMSE score

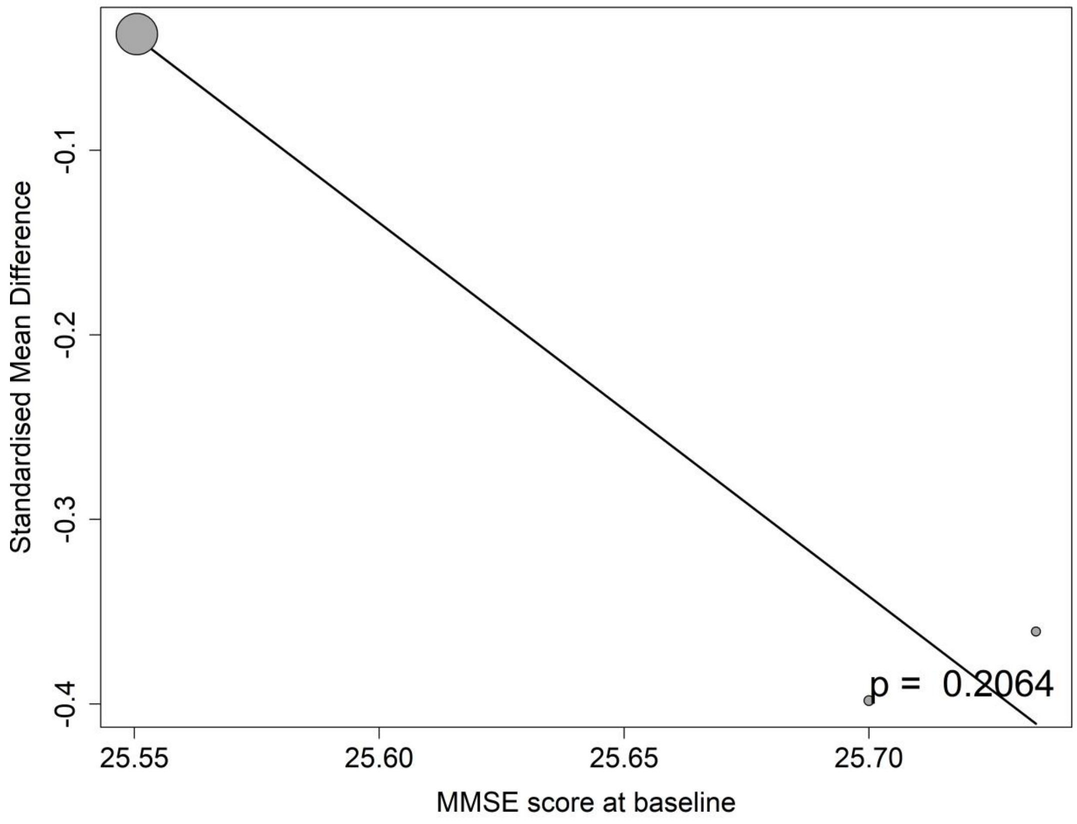

##### 8.14.3 Meta-regressions of CSF Nfl by the mean age at the baseline

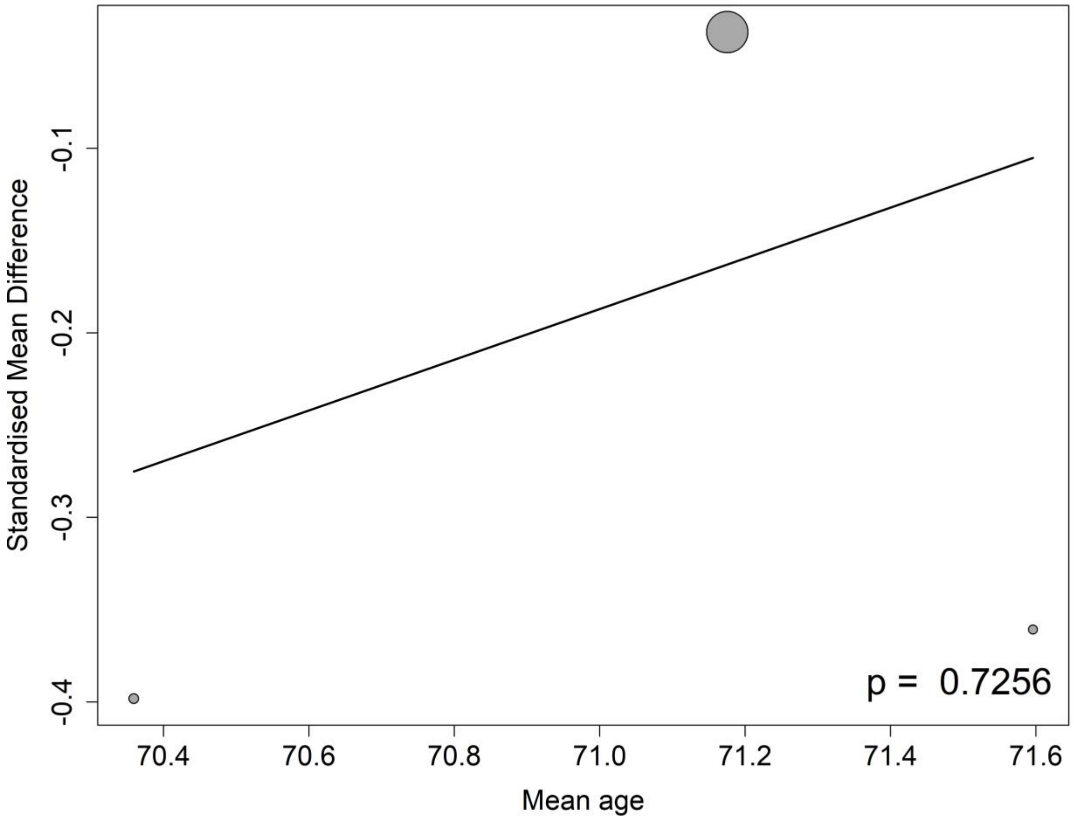

##### 8.14.4 Meta-regressions of CSF Nfl by the percentage of APOE4 carrier at the baseline

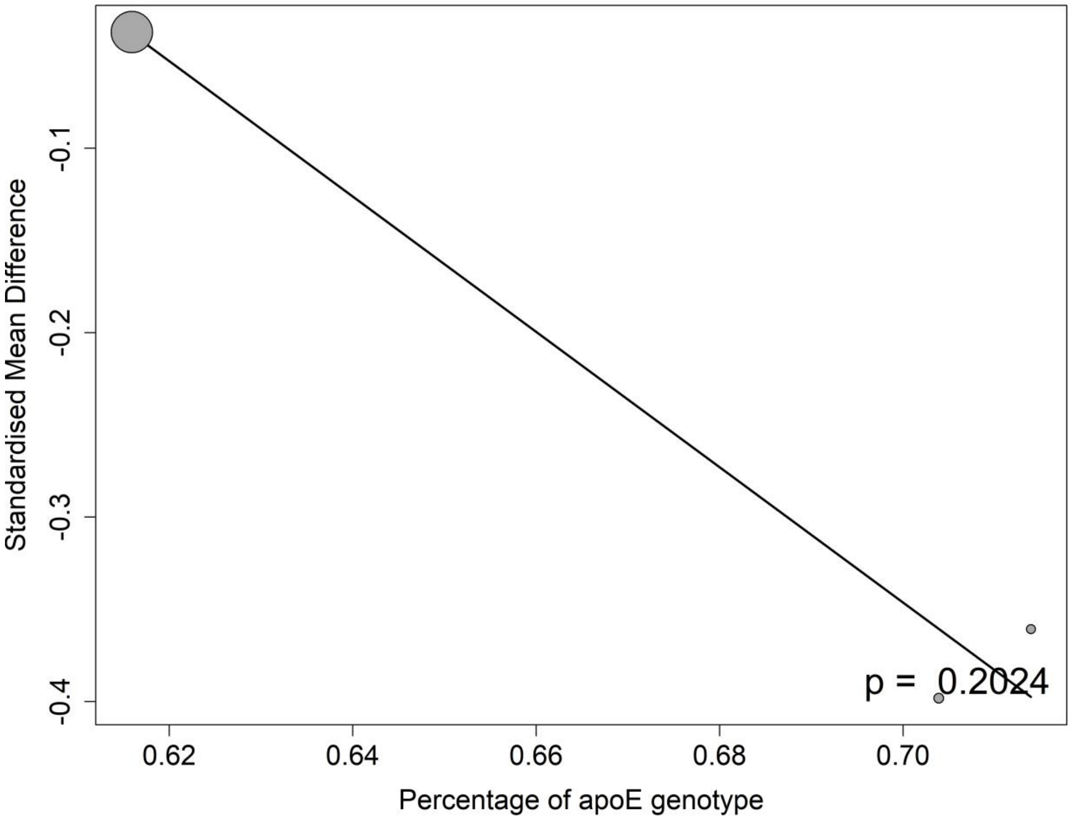

#### 8.15 CSF Neurogranin

##### 8.15.1 Meta-regressions of CSF Neurogranin by baseline CDR-SB score

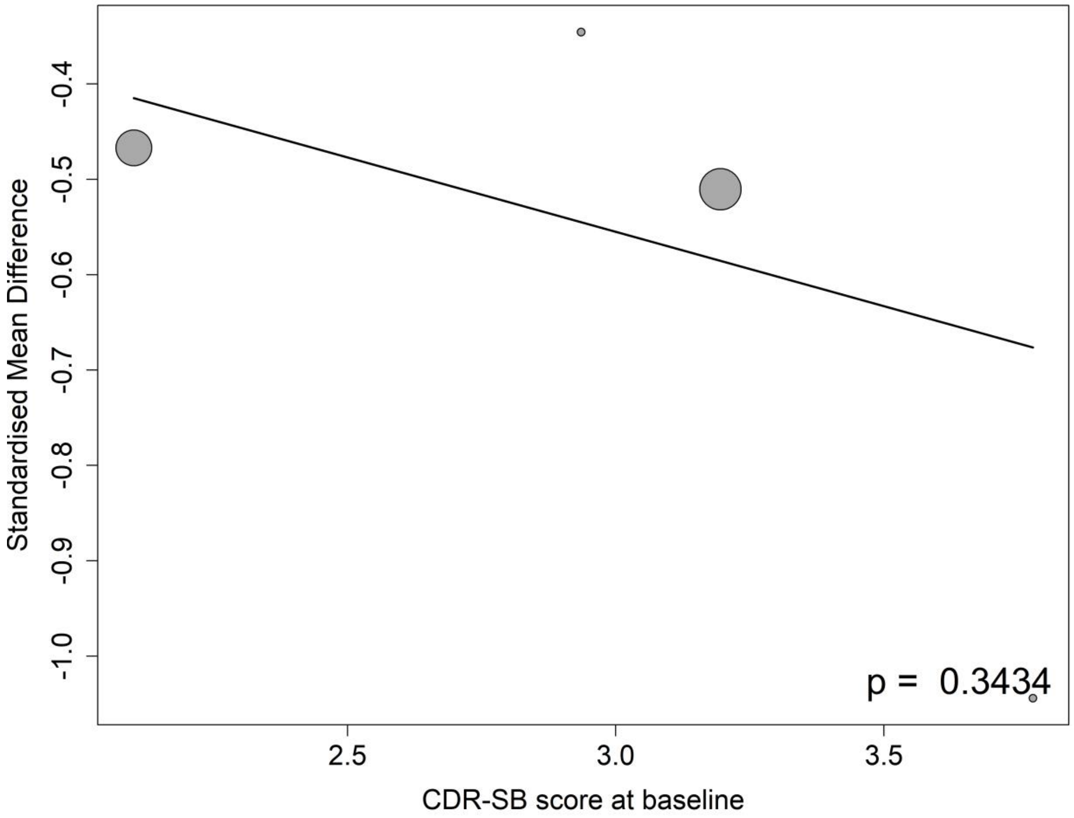

##### 8.15.2 Meta-regressions of CSF Neurogranin by baseline MMSE score

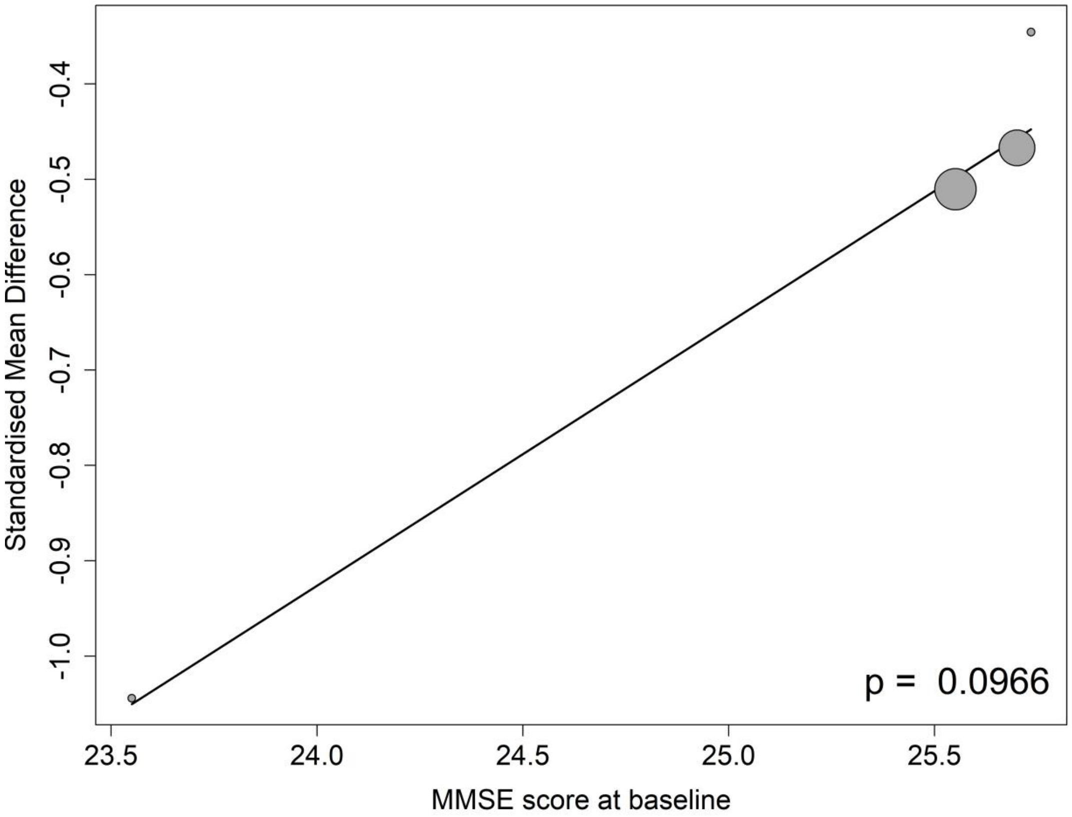

##### 8.15.3 Meta-regressions of CSF Neurogranin by the mean age at the baseline

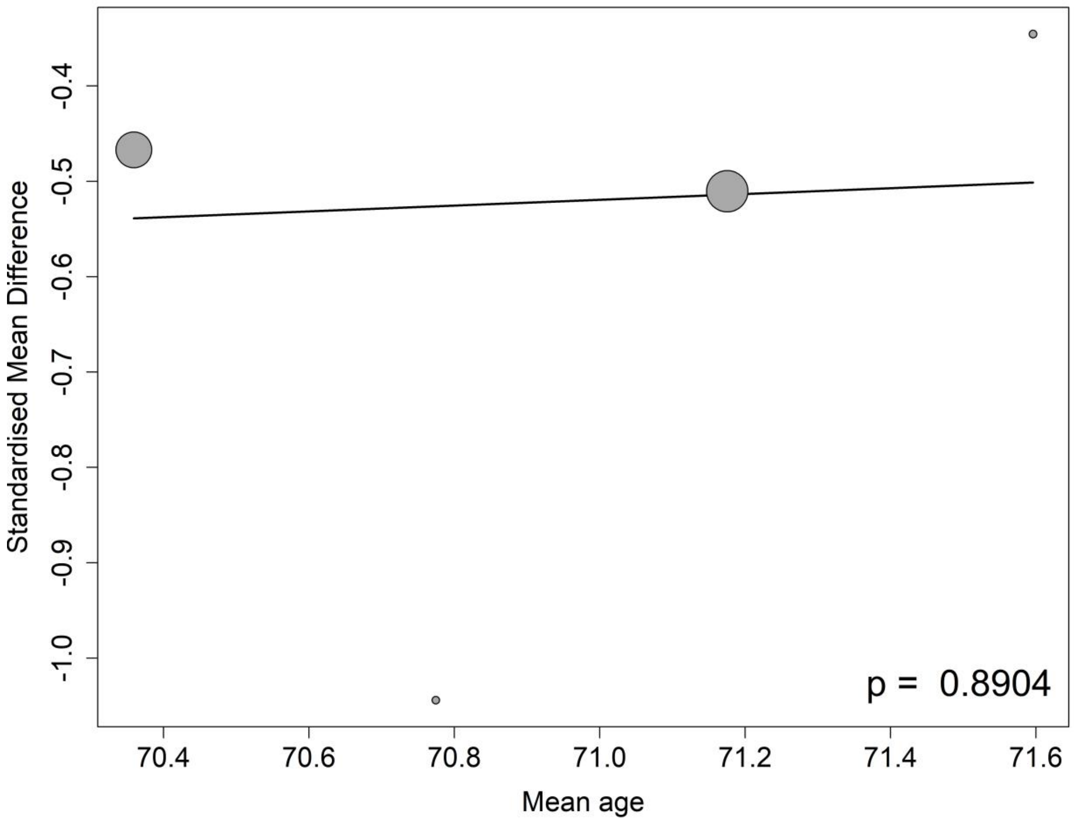

##### 8.15.4 Meta-regressions of CSF Neurogranin by the percentage of APOE4 carrier at the baseline

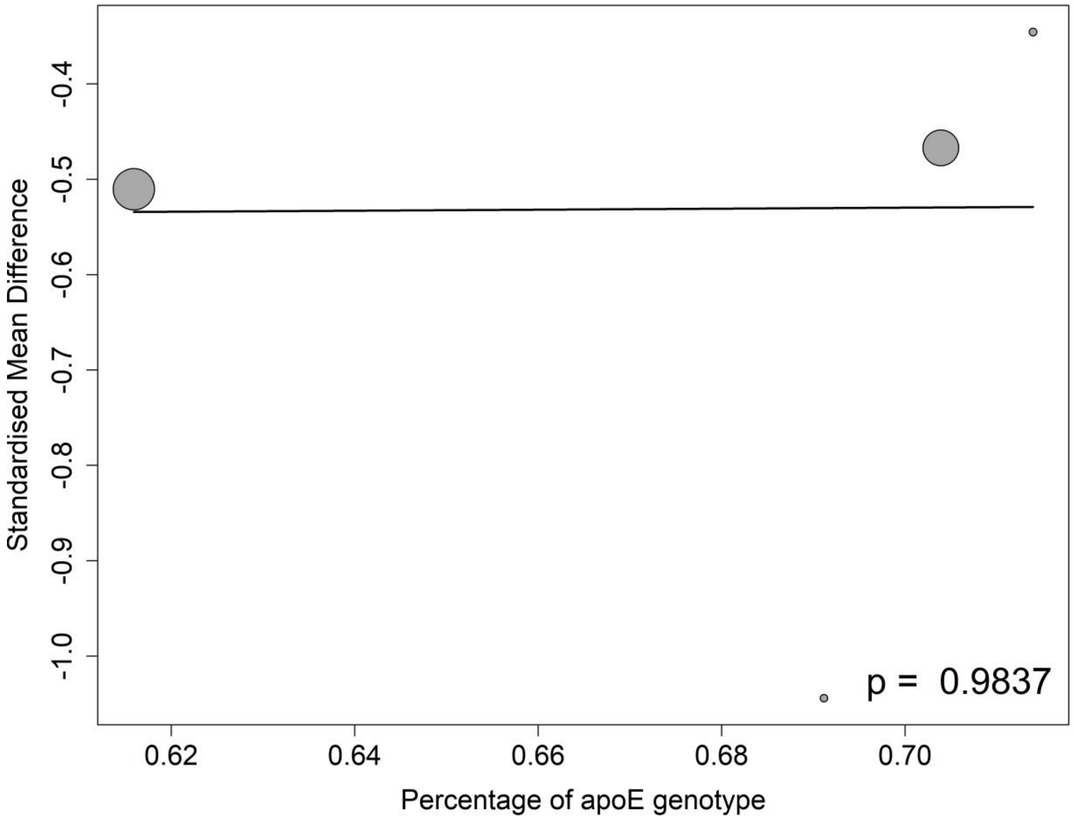

##### 8.15.5 Meta-regressions of CSF Neurogranin by the percentage of female participants at the baseline

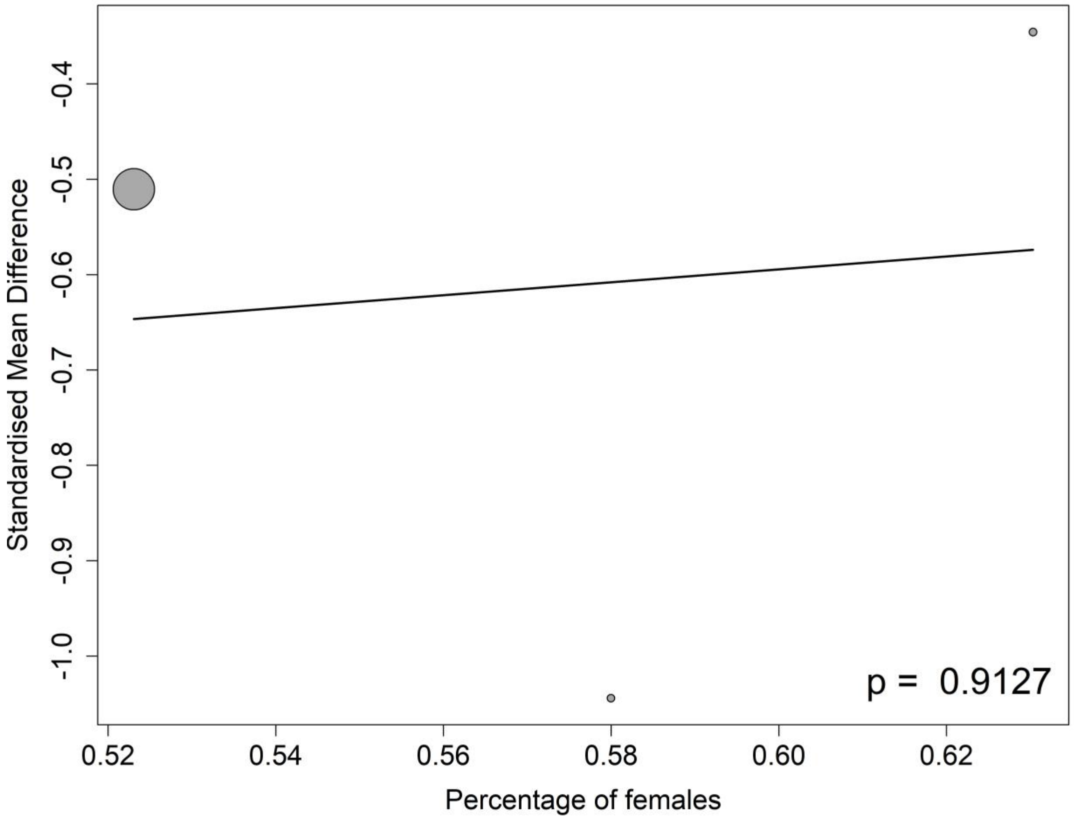

##### 8.15.6 Meta-regressions of CSF Neurogranin by the percentage of participants on anti-AD medication at the baseline

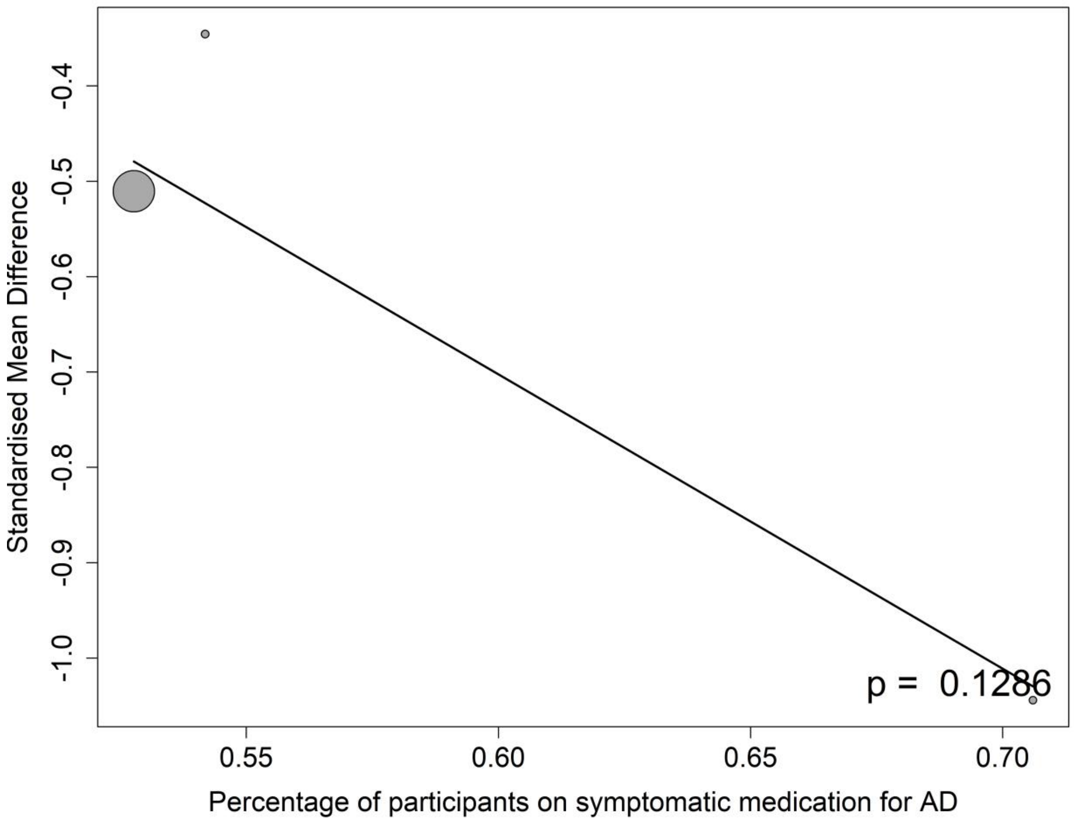

#### 8.16 Plasma Aβ40

##### 8.16.1 Meta-regressions of Plasma Aβ40 by baseline MMSE score

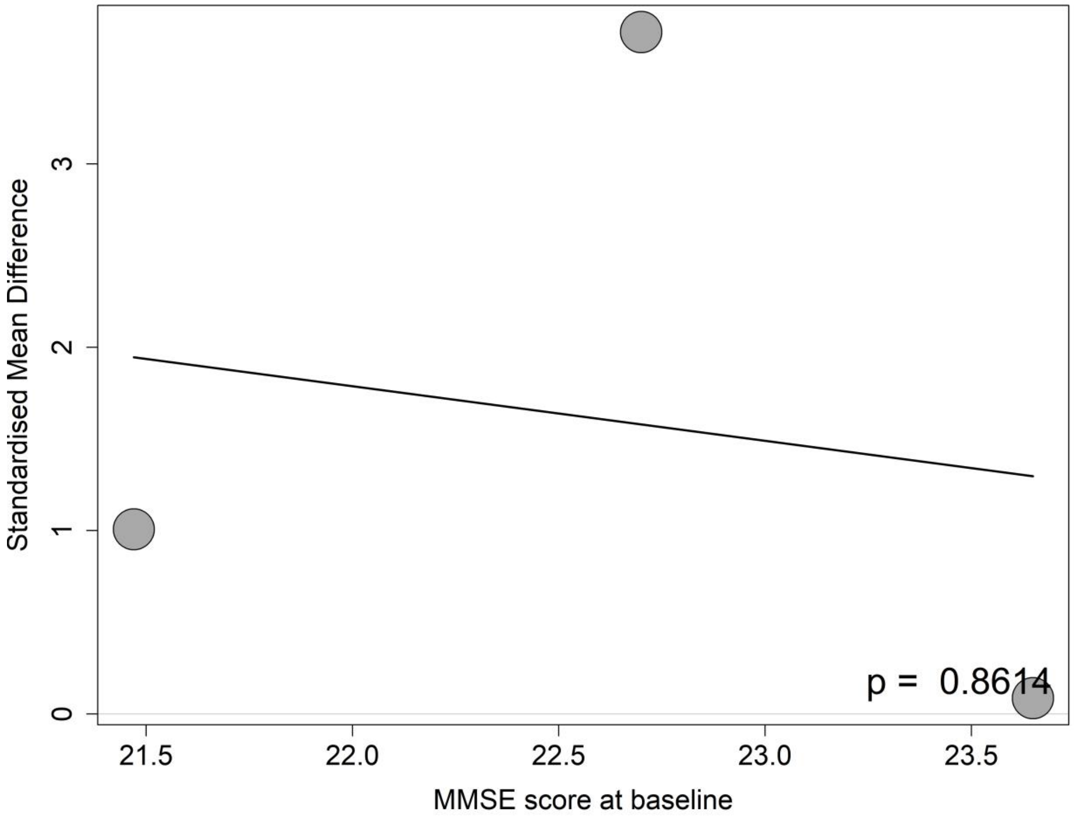

##### 8.16.2 Meta-regressions of Plasma Aβ40 by the mean age at the baseline

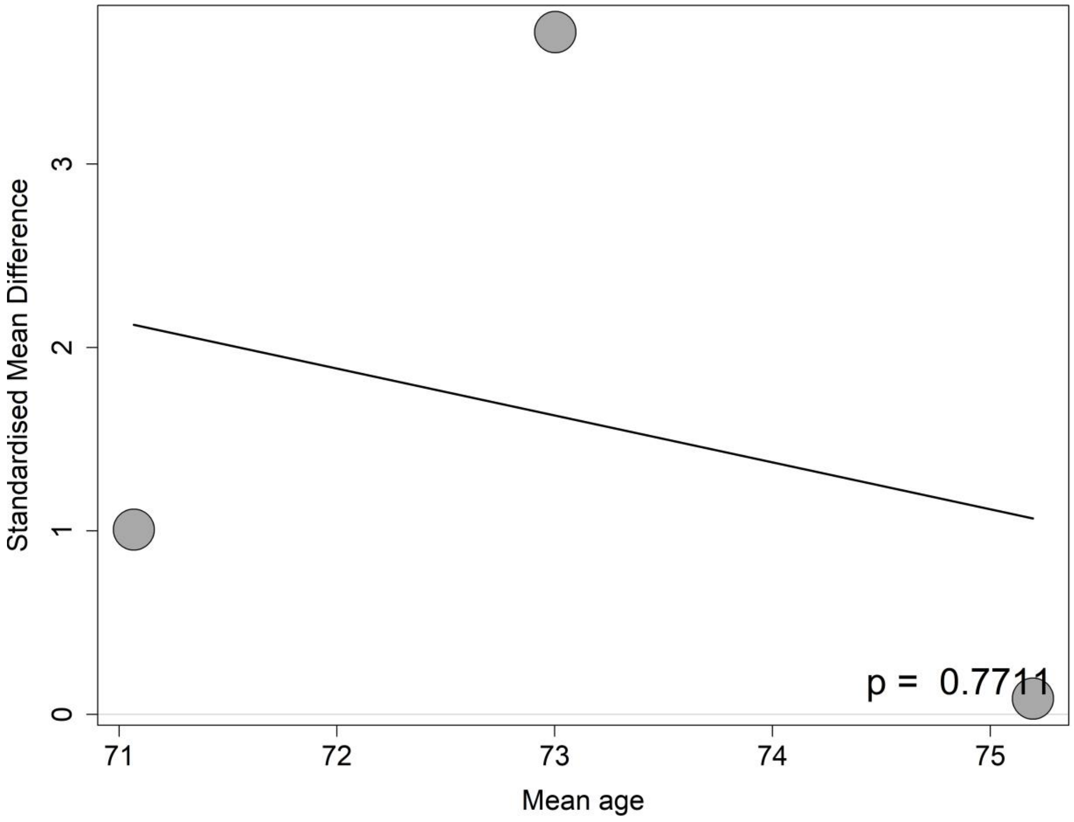

##### 8.16.3 Meta-regressions of Plasma Aβ40 by the percentage of female participants at the baseline

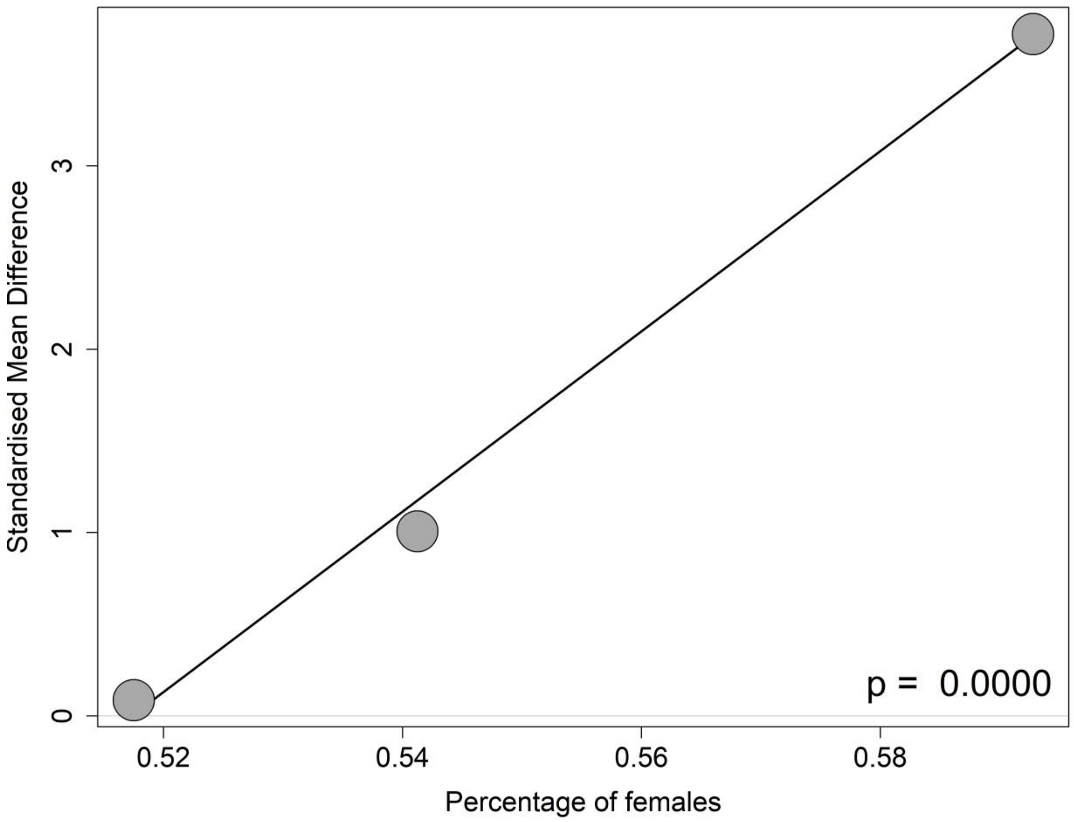

#### 8.17 Plasma Nfl

##### 8.17.1 Meta-regressions of Plasma Nfl by baseline CDR-SB score

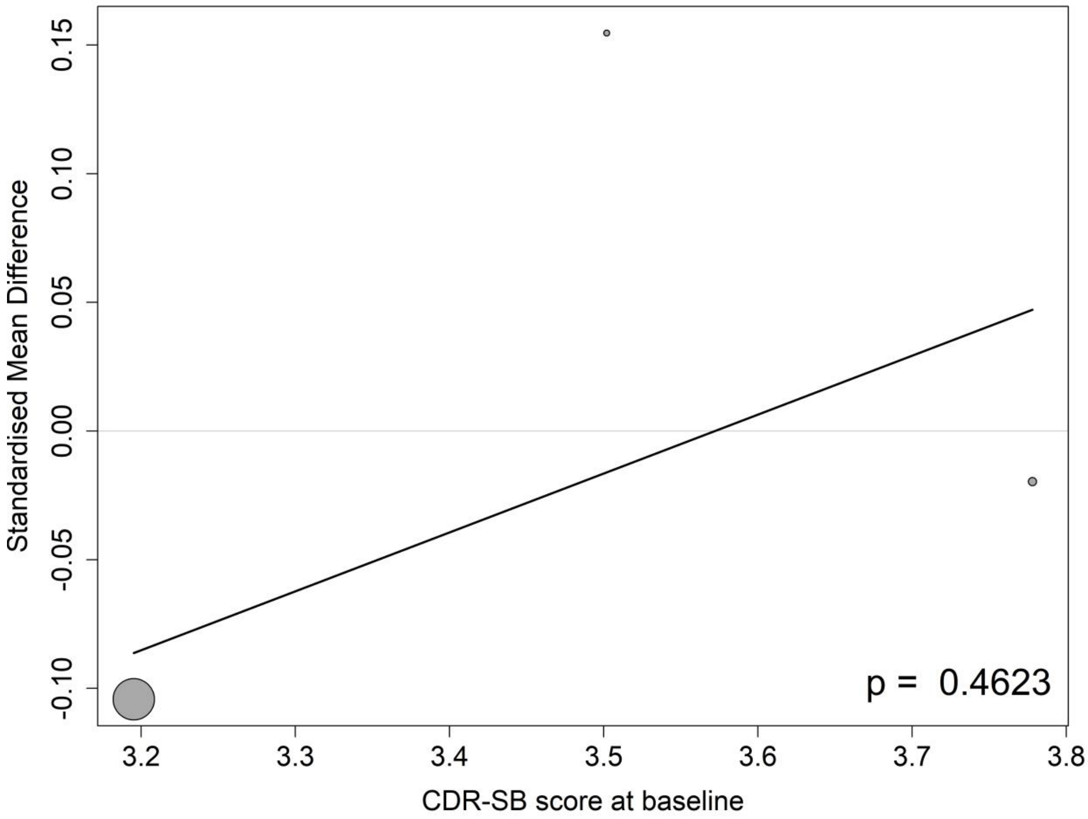

##### 8.17.2 Meta-regressions of Plasma Nfl by baseline MMSE score

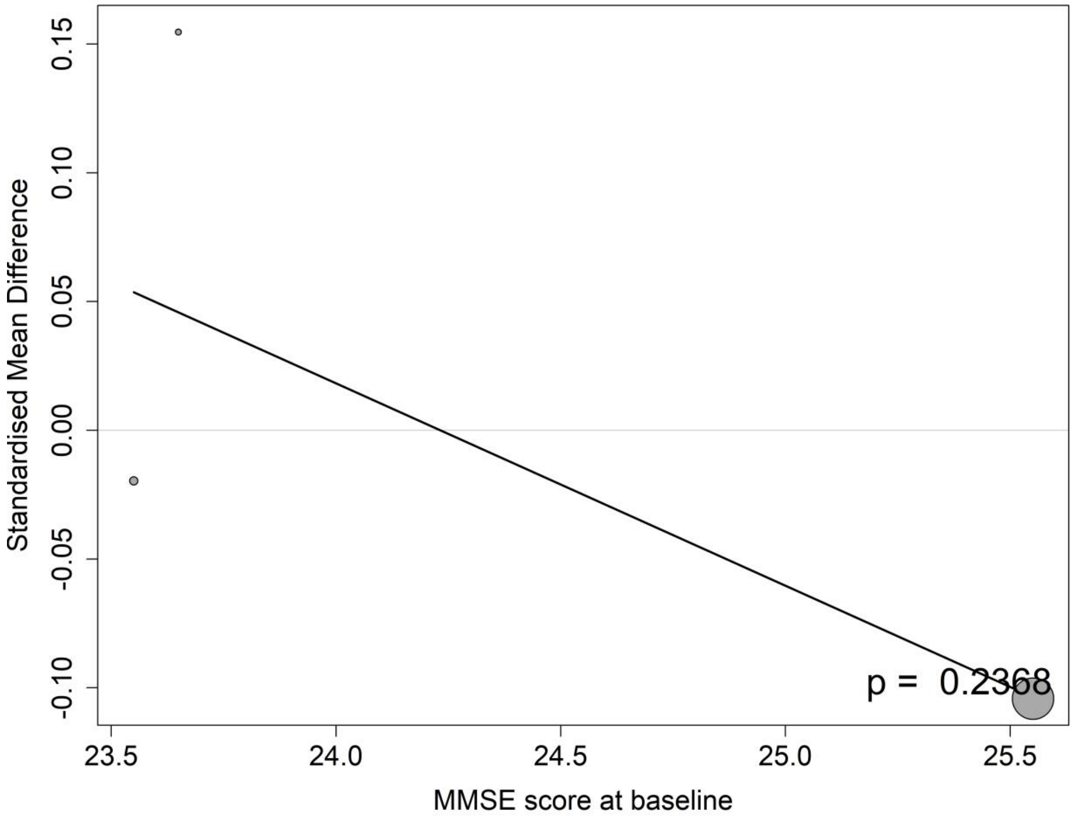

##### 8.17.3 Meta-regressions of Plasma Nfl by the mean age at the baseline

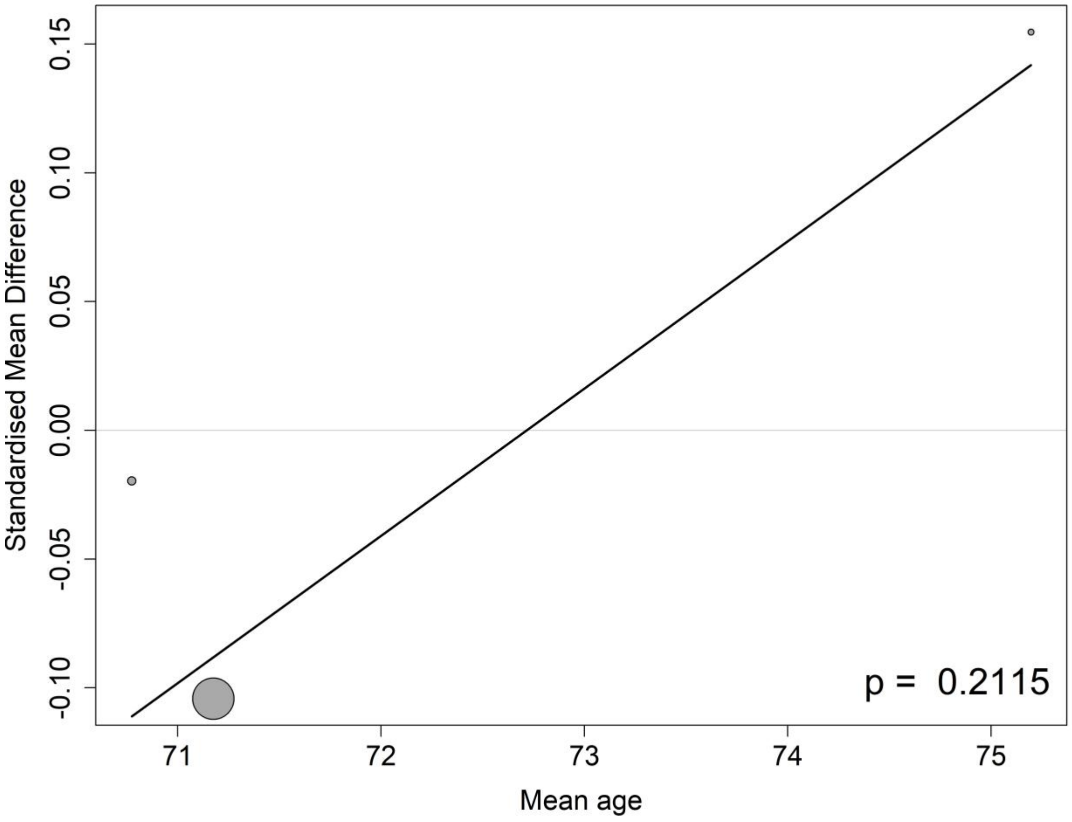

##### 8.17.4 Meta-regressions of Plasma Nfl by the percentage of APOE4 carrier at the baseline

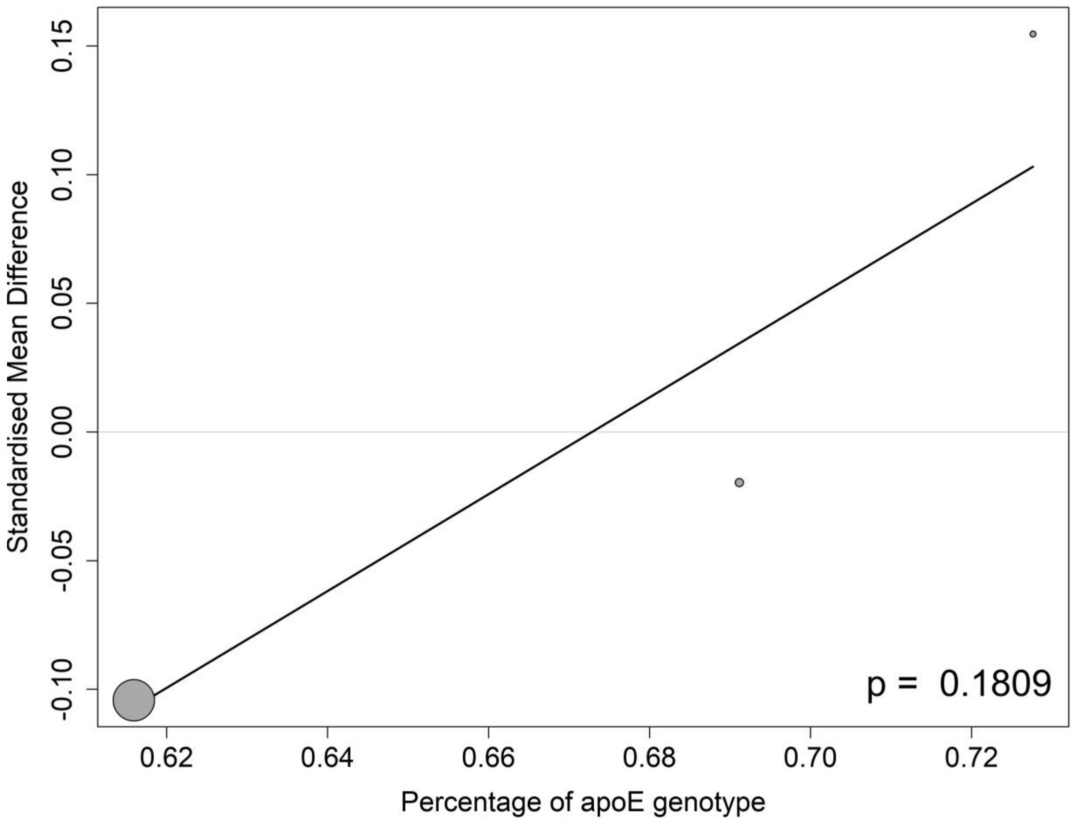

##### 8.17.5 Meta-regressions of Plasma Nfl by the percentage of female participants at the baseline

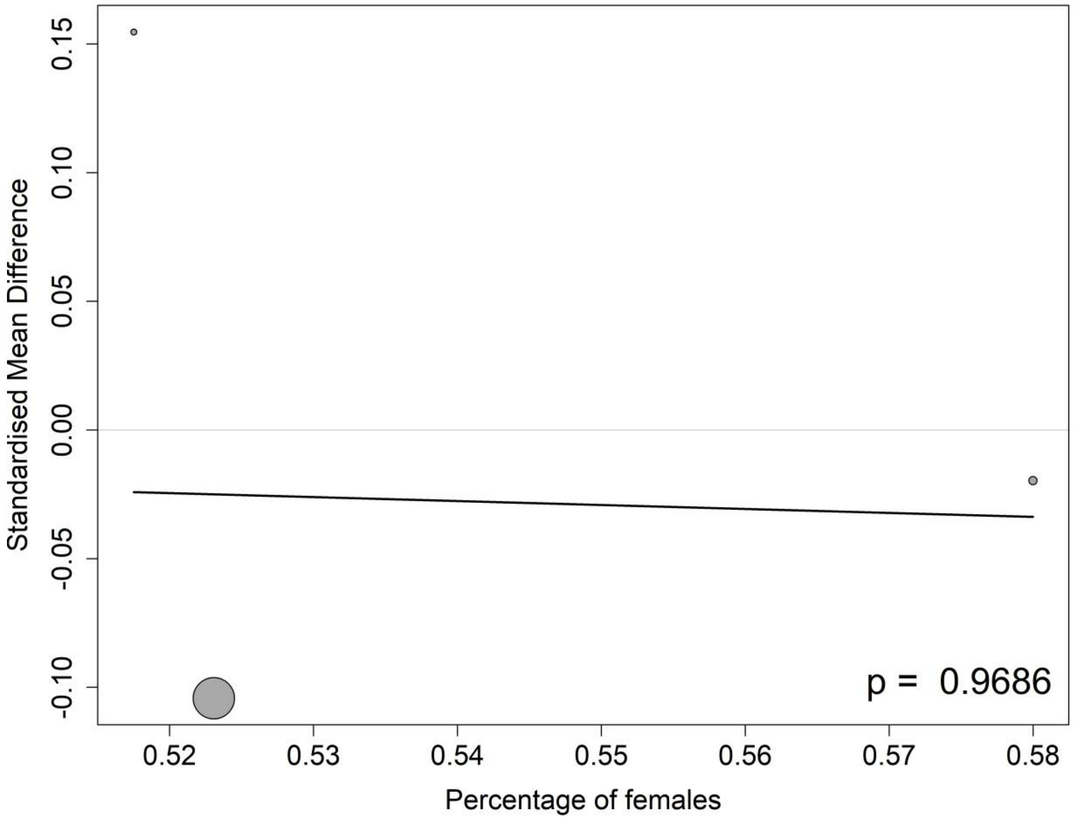

##### 8.17.6 Meta-regressions of Plasma Nfl by the percentage of participants on anti-AD medication at the baseline

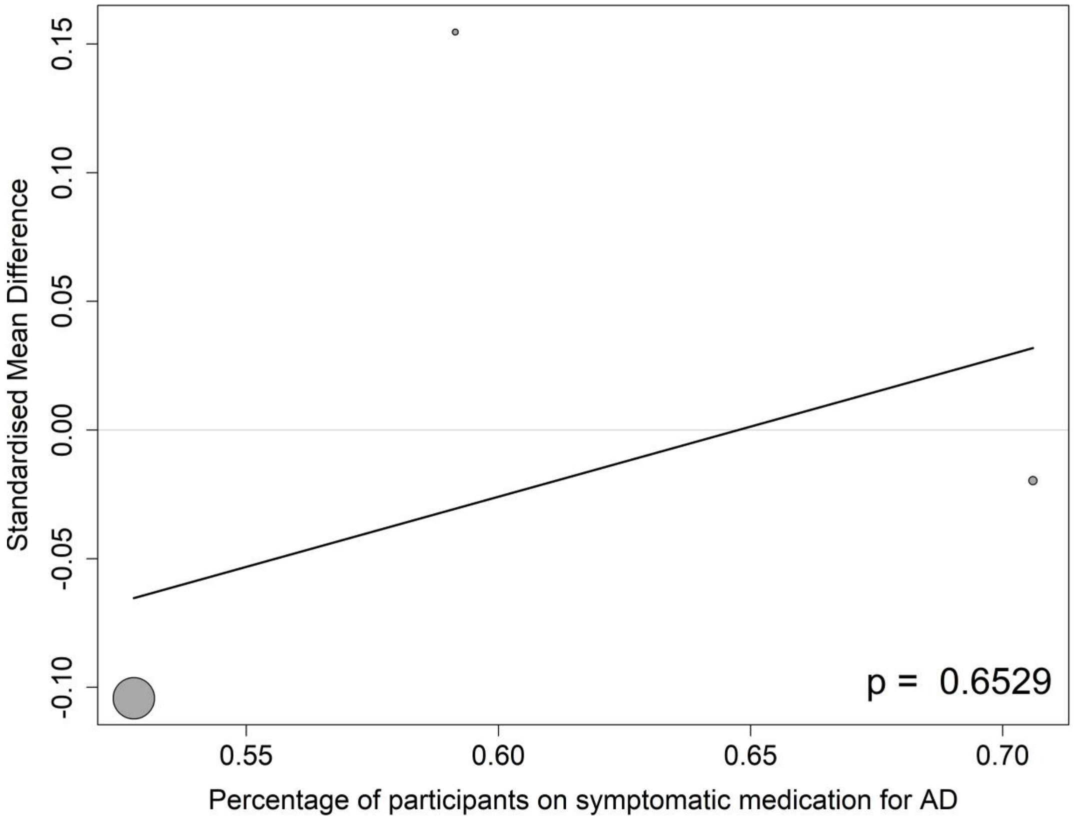

#### 8.18 Plasma p-tau181

##### 8.18.1 Meta-regressions of plasma p-tau181 by baseline CDR-SB score

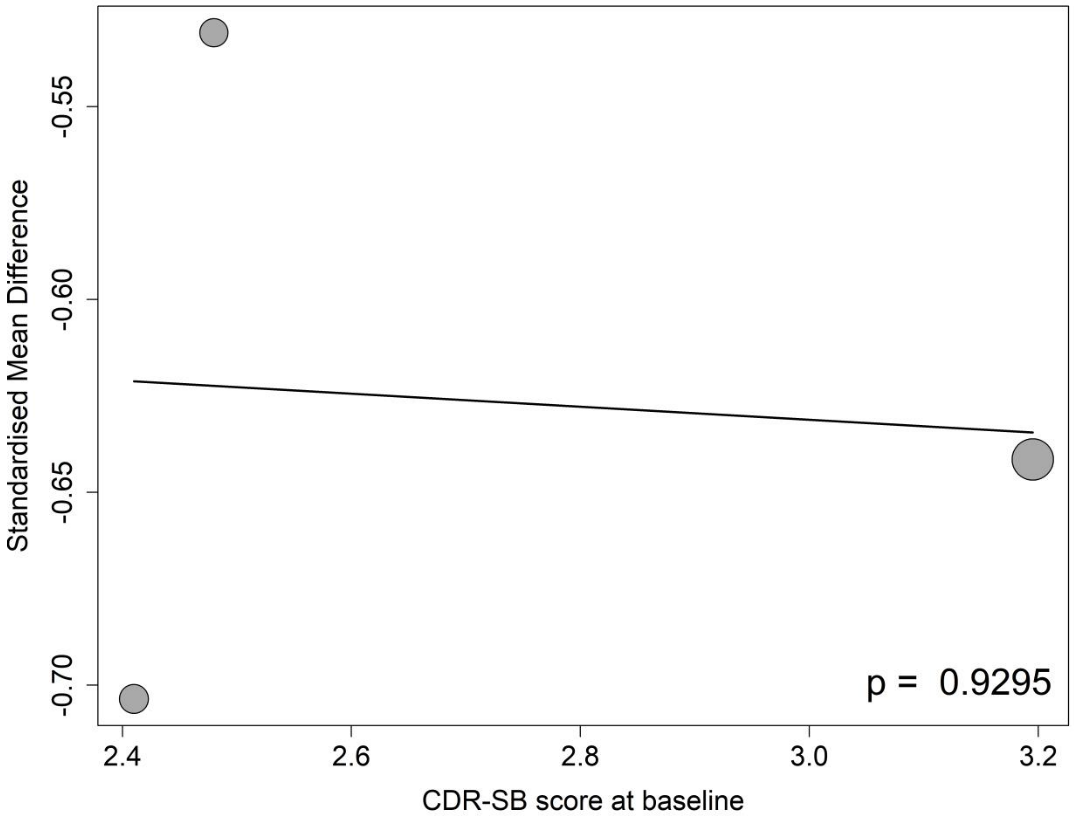

##### 8.18.2 Meta-regressions of plasma p-tau181 by baseline MMSE score

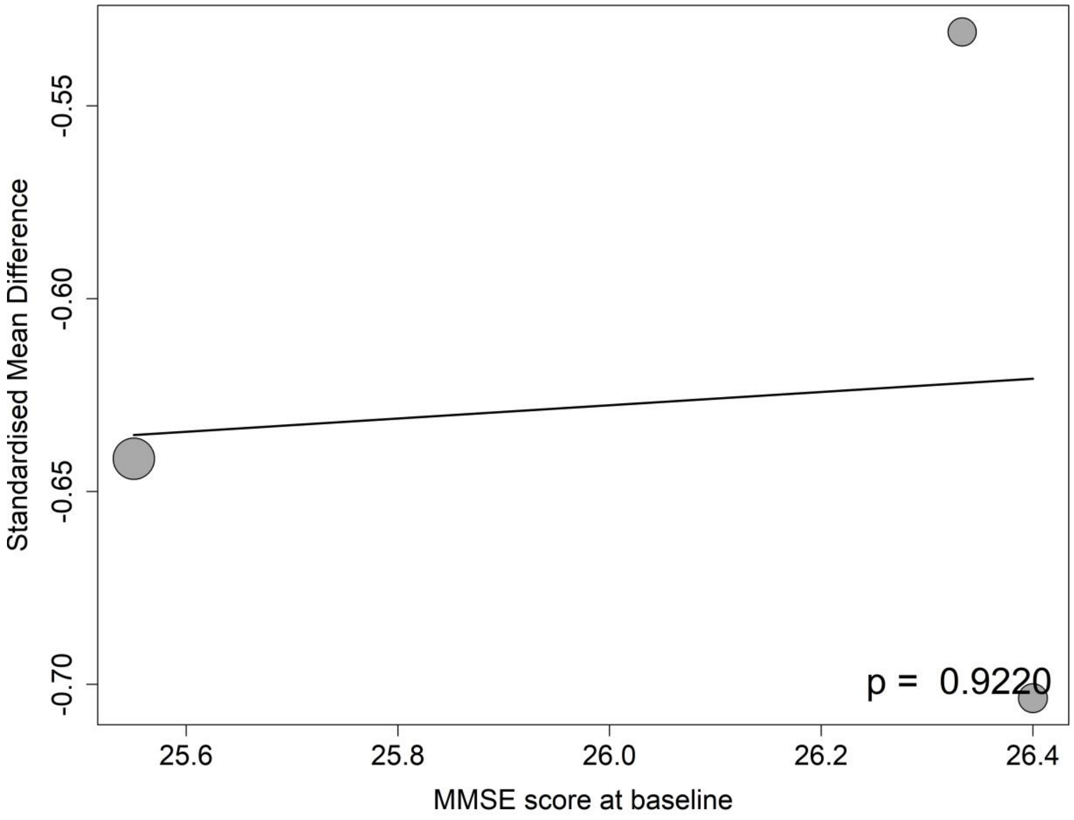

##### 8.18.3 Meta-regressions of plasma p-tau181 by the mean age at the baseline

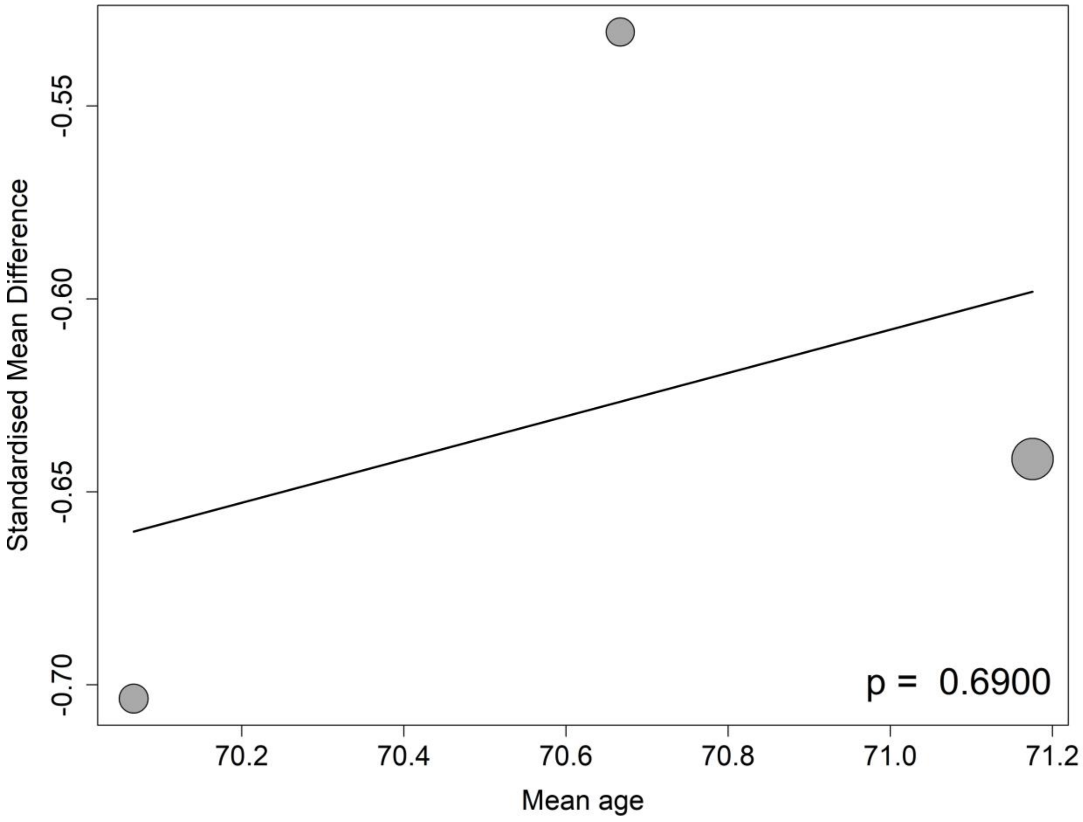

##### 8.18.4 Meta-regressions of plasma p-tau181 by the percentage of APOE4 carrier at the baseline

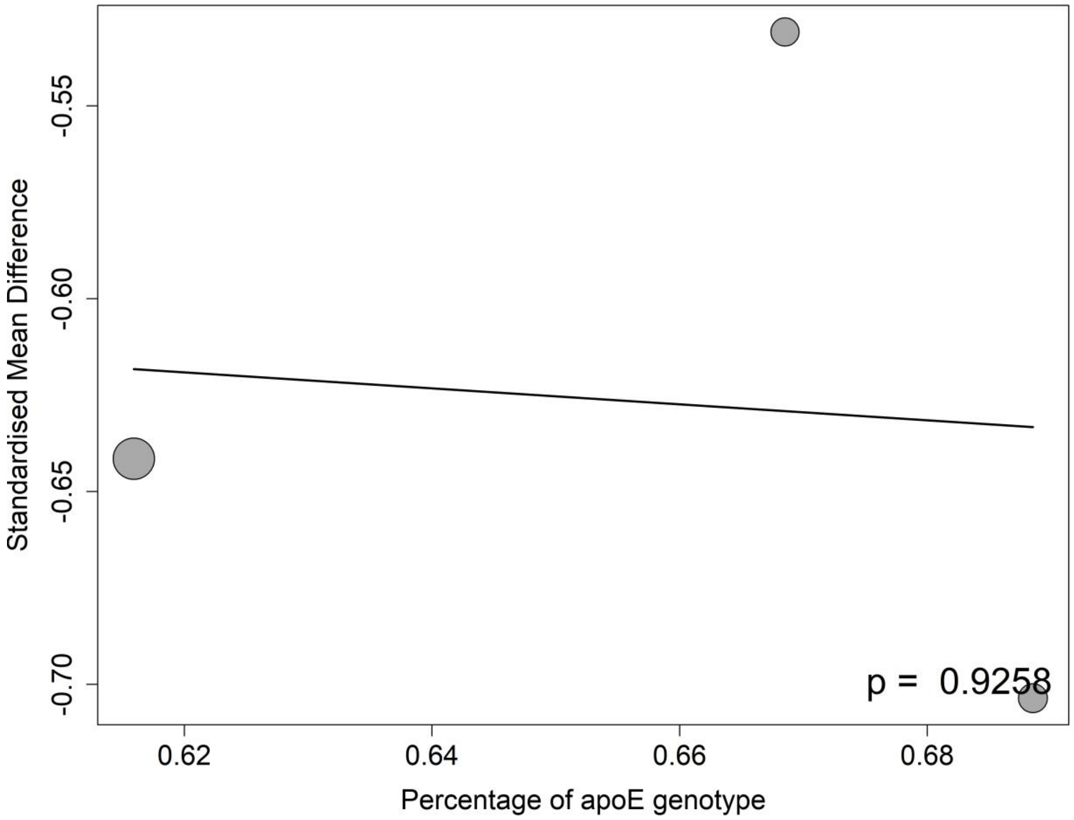

**8.18.5 Meta-regressions of plasma p-tau181 by the percentage of female participants at the baseline**

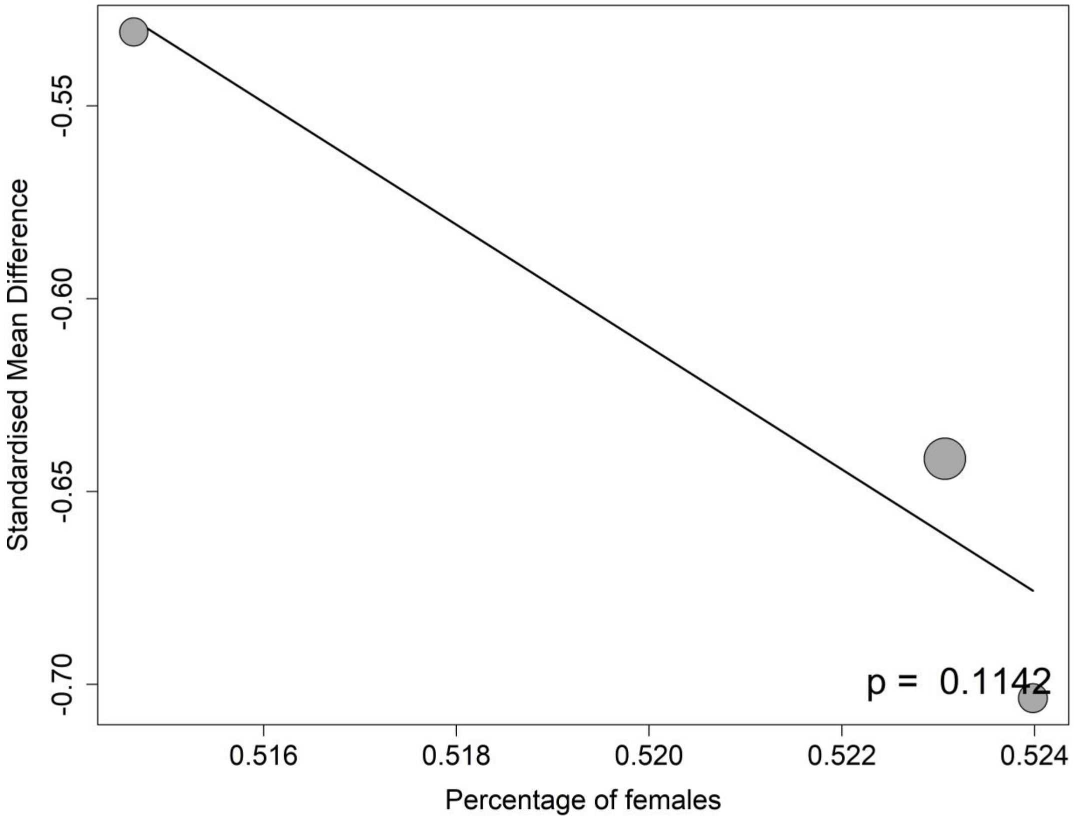

**8.18.6 Meta-regressions of plasma p-tau181 by the percentage of participants on anti-AD medication at the baseline**

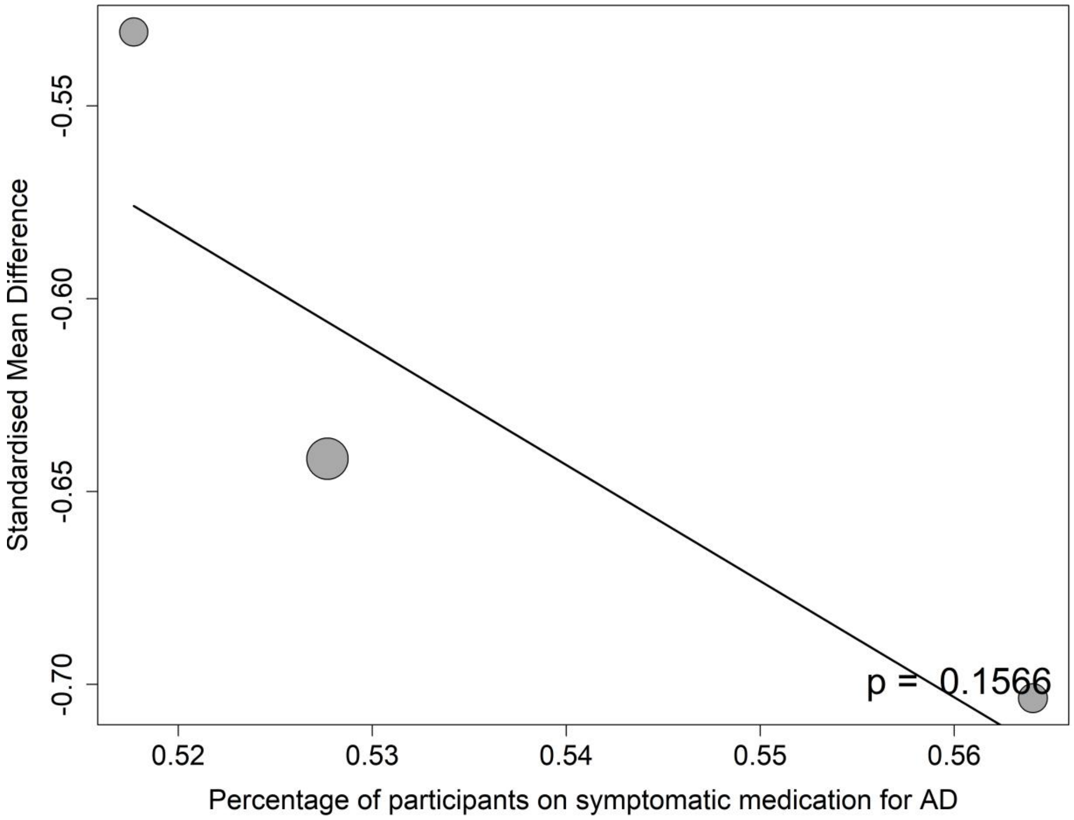

#### 8.19 ARIA-E

##### 8.19.1 Meta-regressions of ARIA-E by baseline CDR-SB score

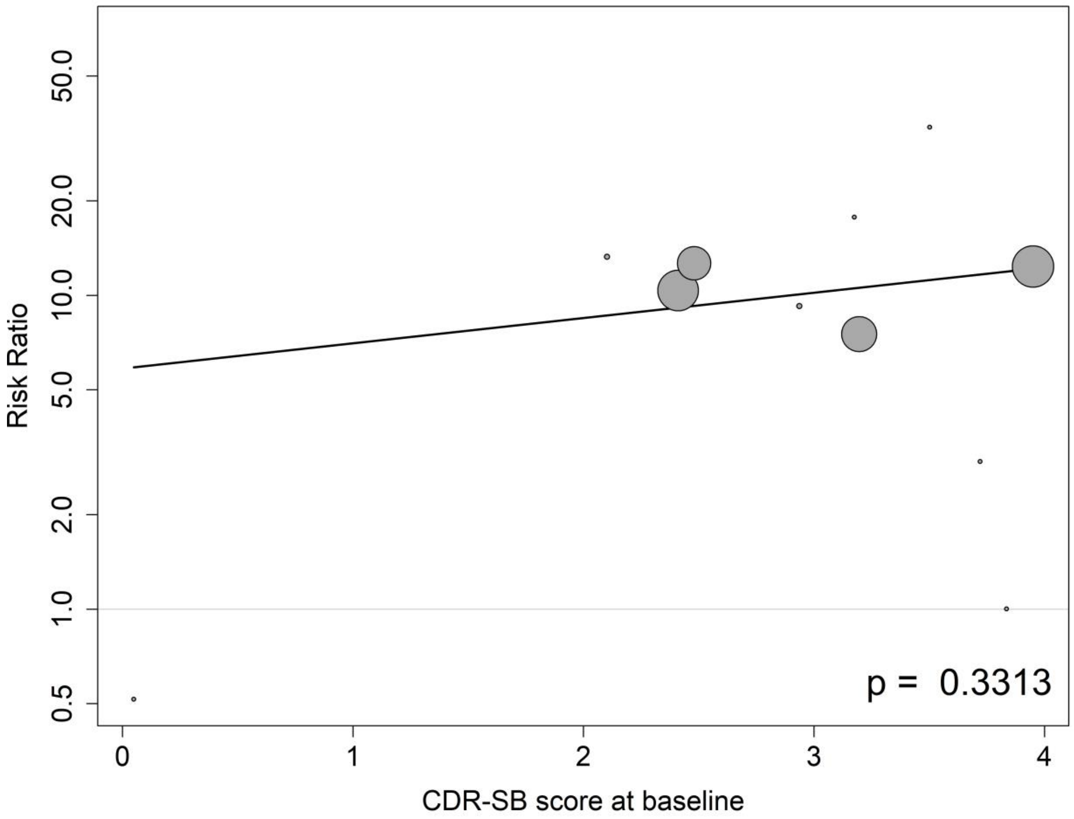

##### 8.19.2 Meta-regressions of ARIA-E by baseline MMSE score

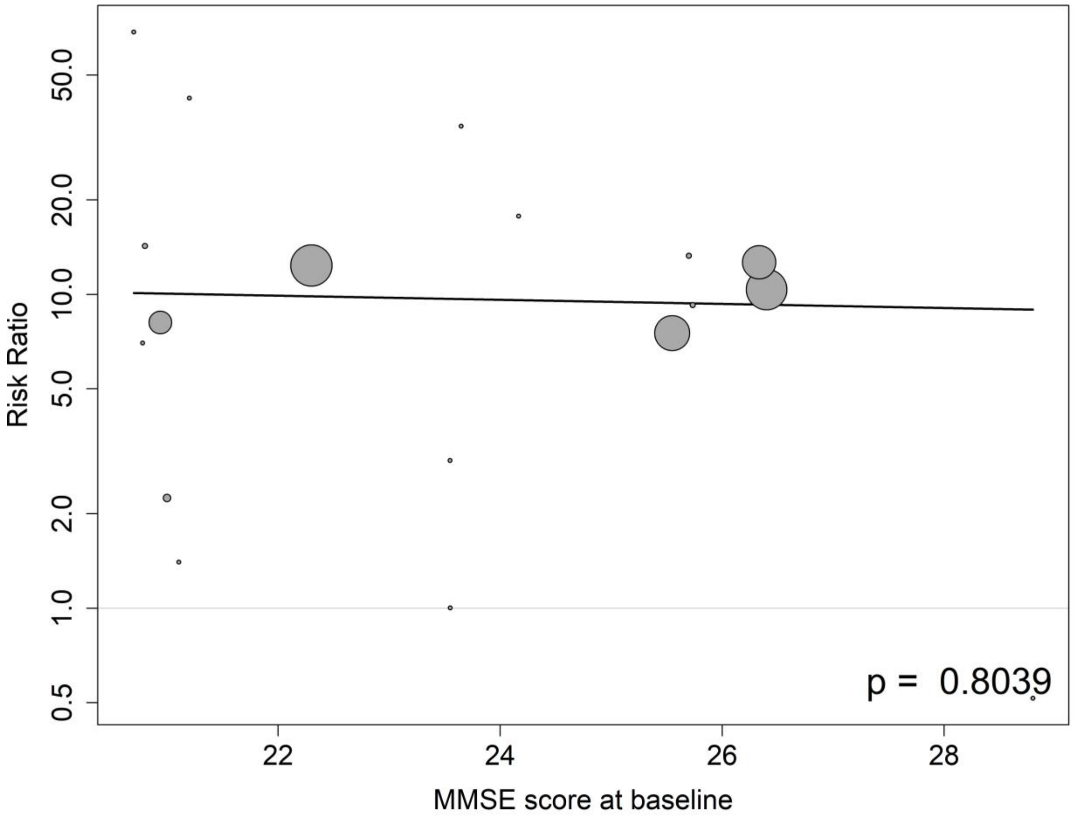

##### 8.19.3 Meta-regressions of ARIA-E by the mean age at the baseline

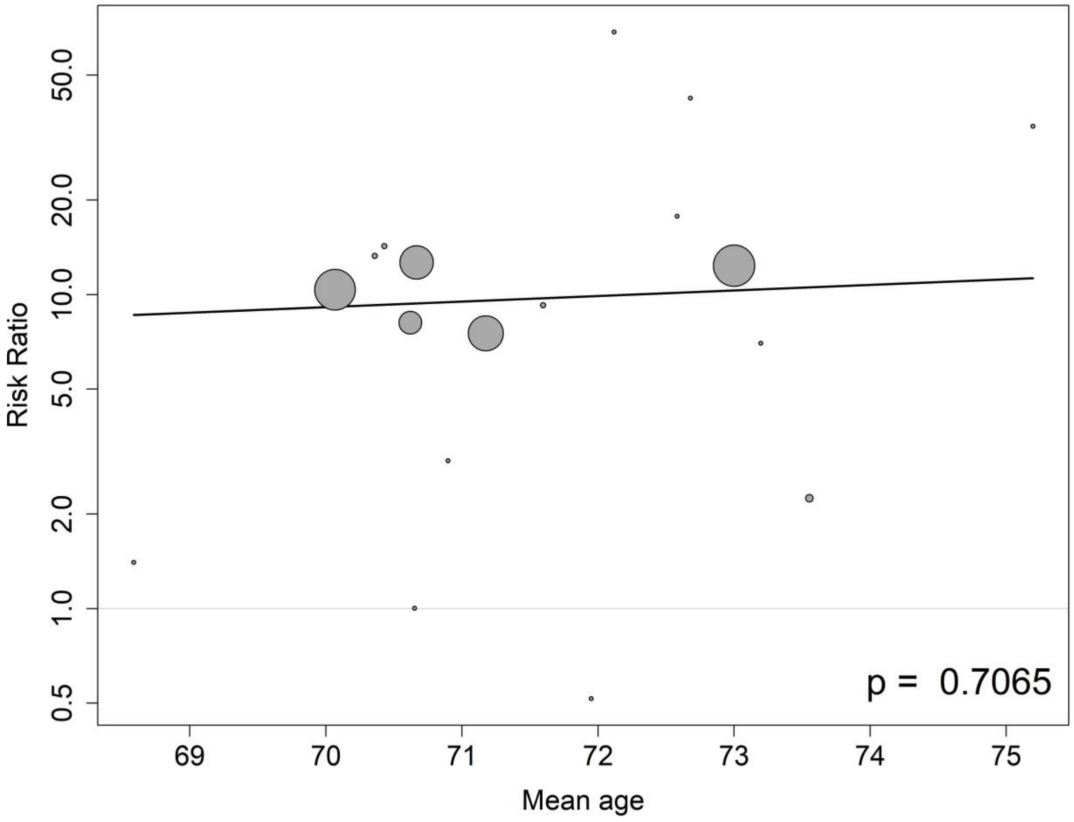

**8.19.4 Meta-regressions of ARIA-E by the percentage of APOE4 carrier at the baseline**

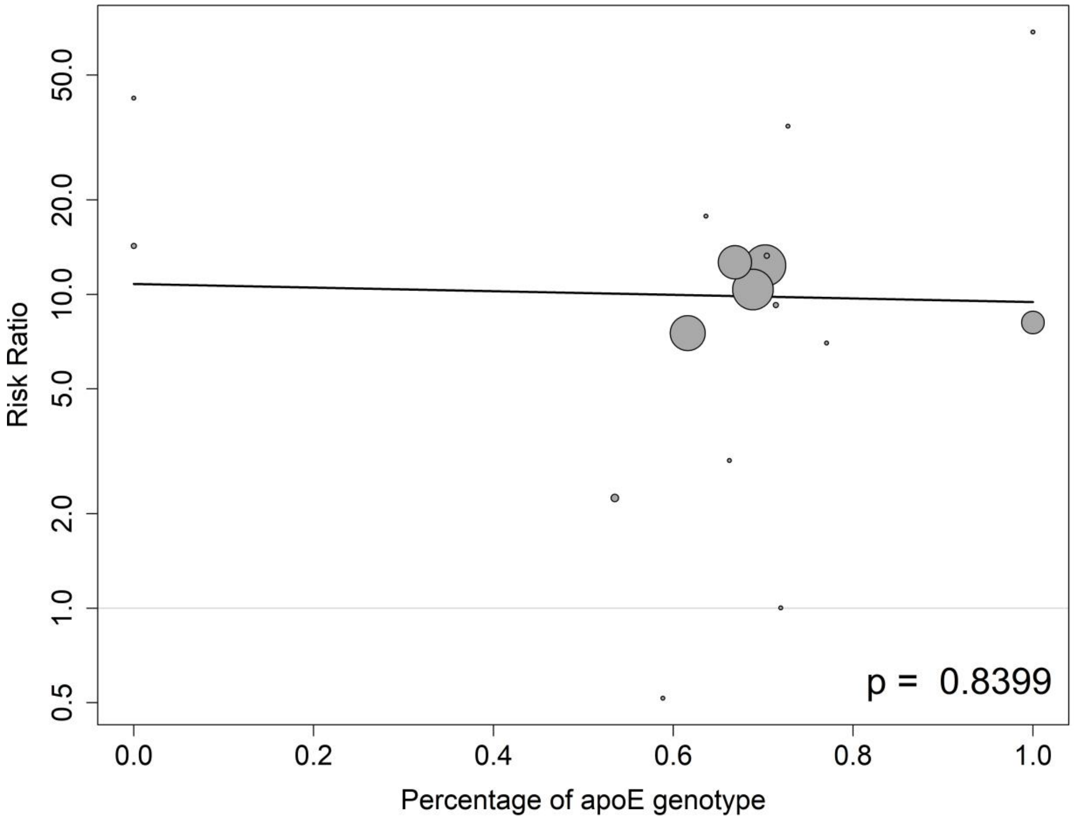

##### 8.19.5 Meta-regressions of ARIA-E by the percentage of female participants at the baseline

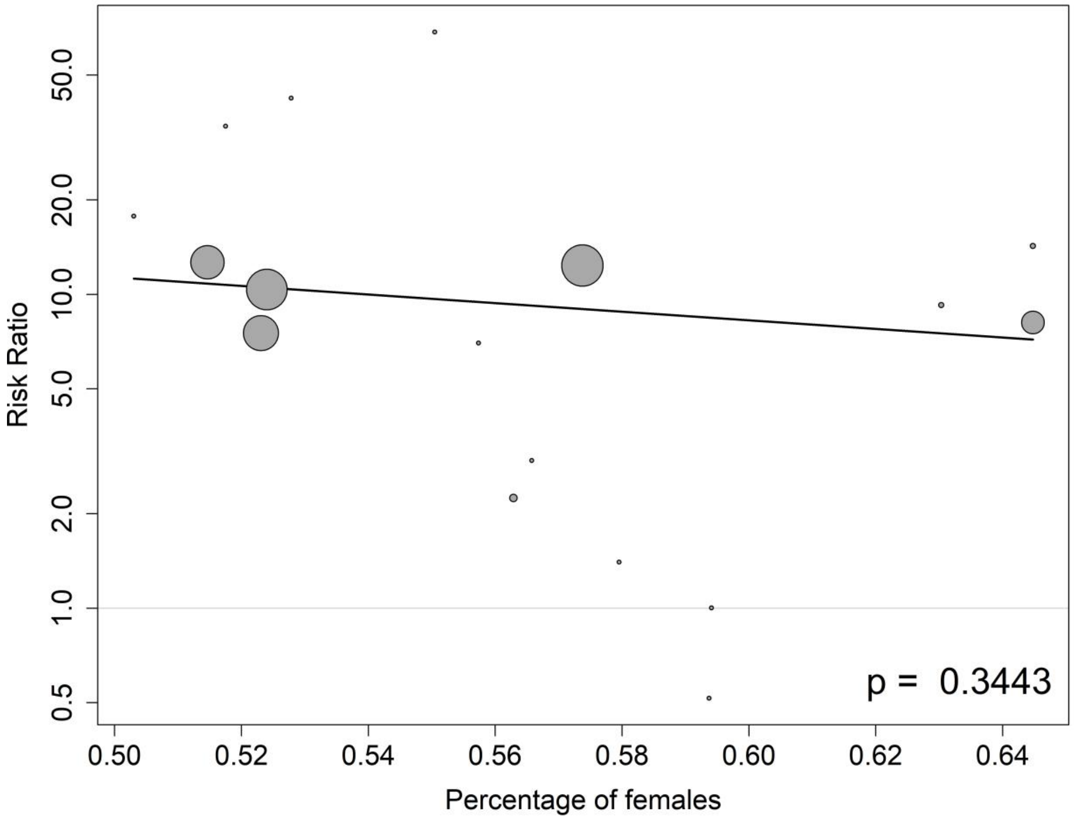

##### 8.19.6 Meta-regressions of ARIA-E by the percentage of participants on anti-AD medication at the baseline

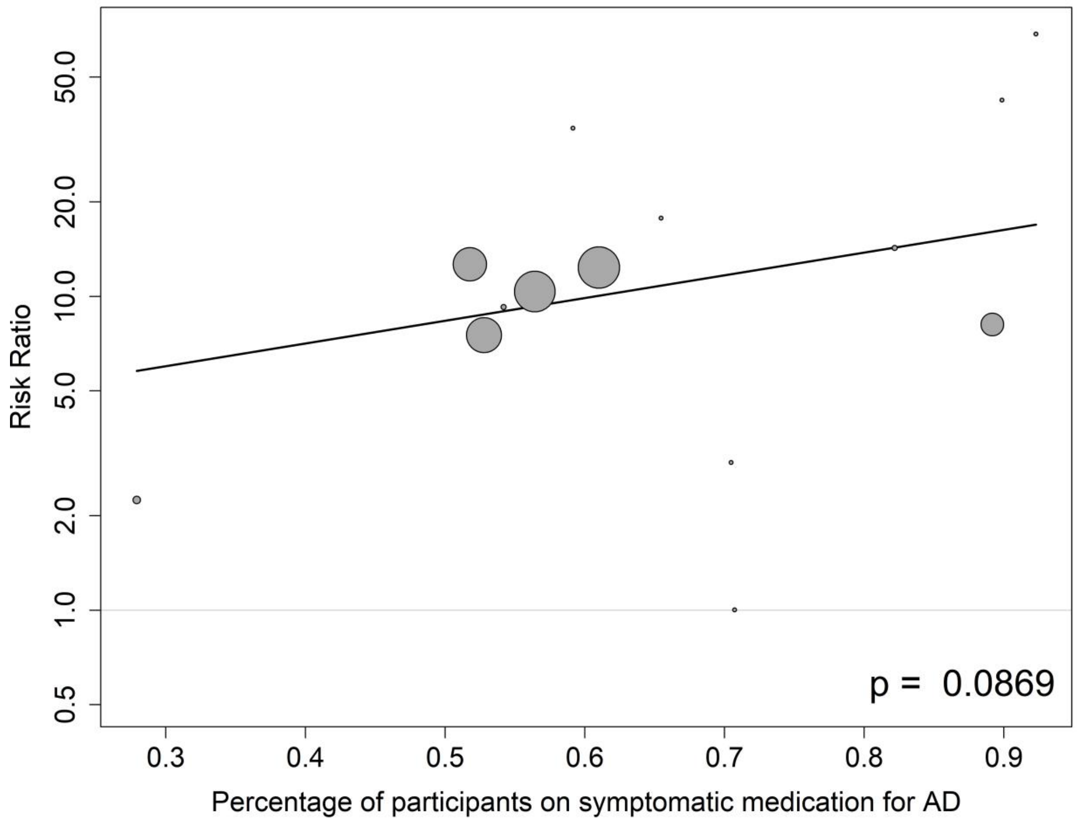

#### 8.20 ARIA-H

##### 8.20.1 Meta-regressions of ARIA-H by baseline CDR-SB score

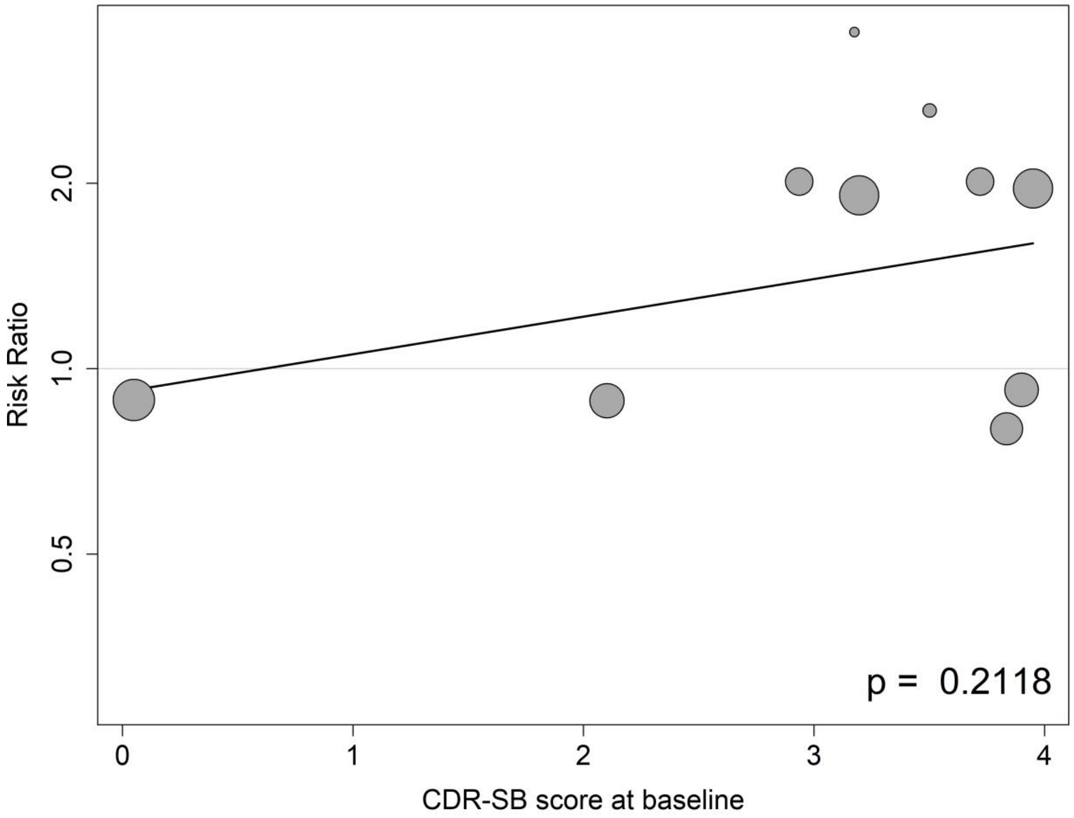

##### 8.20.2 Meta-regressions of ARIA-H by baseline MMSE score

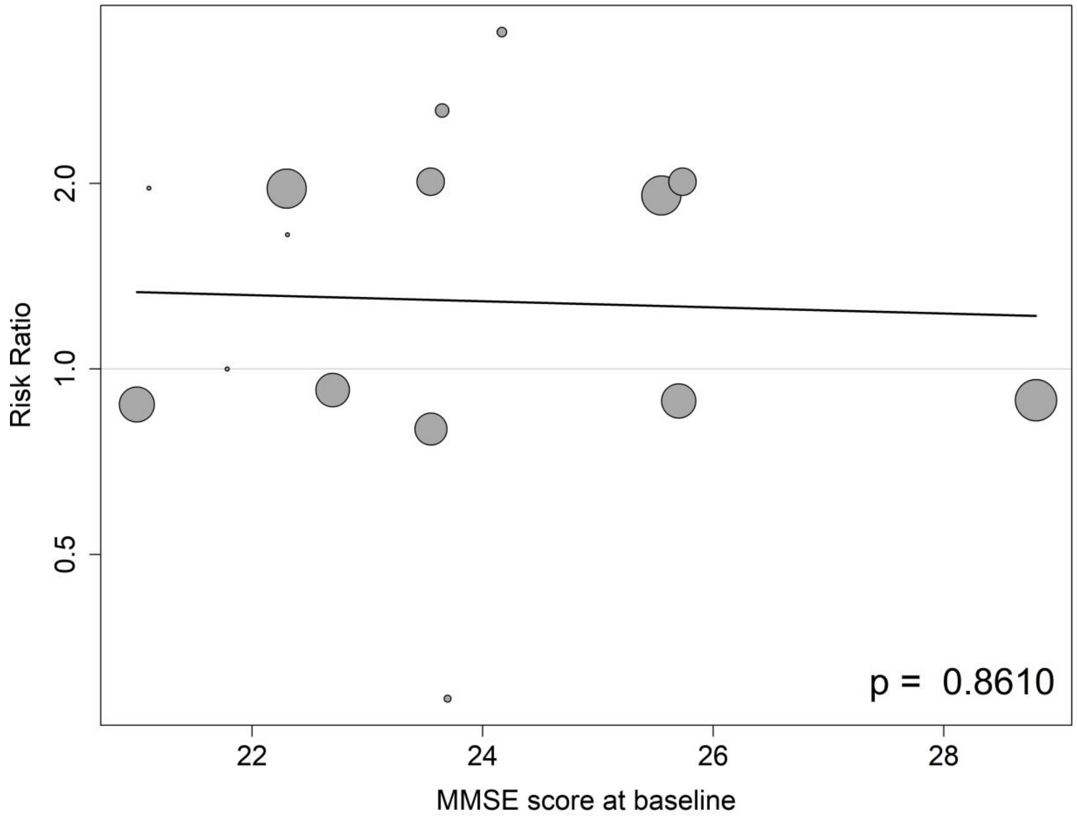

##### 8.20.3 Meta-regressions of ARIA-H by the mean age at the baseline

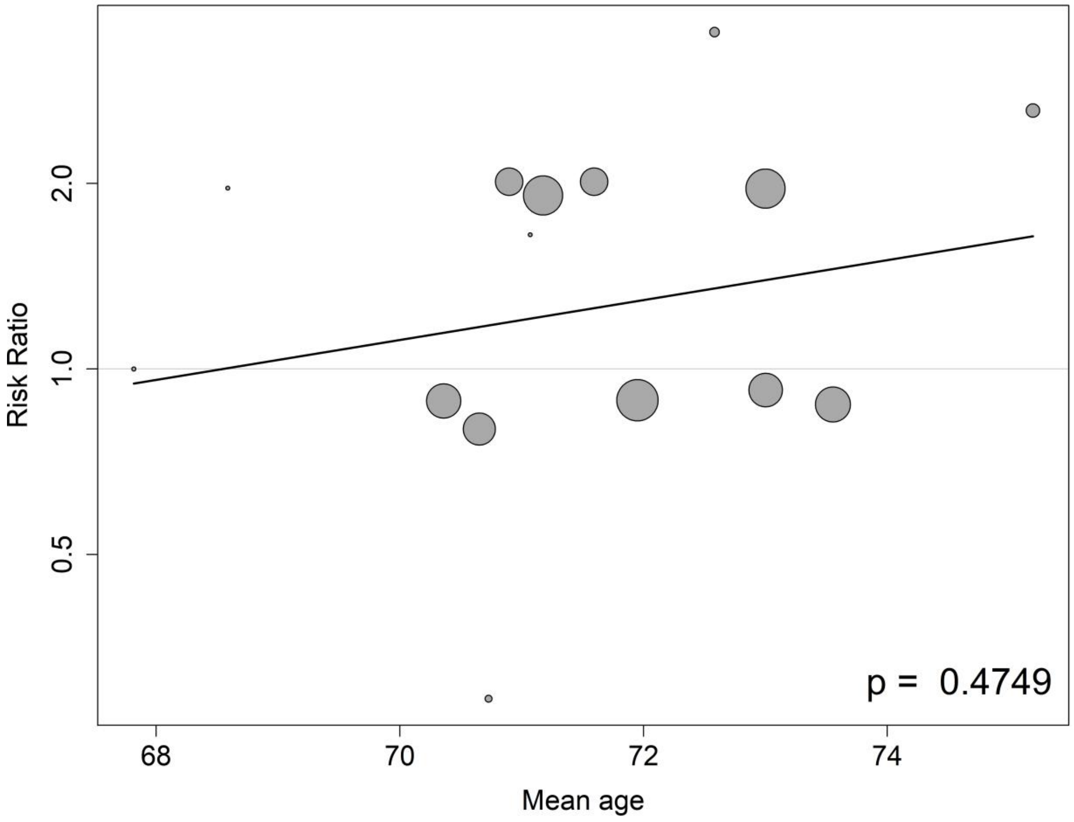

##### 8.20.4 Meta-regressions of ARIA-H by the percentage of APOE4 carrier at the baseline

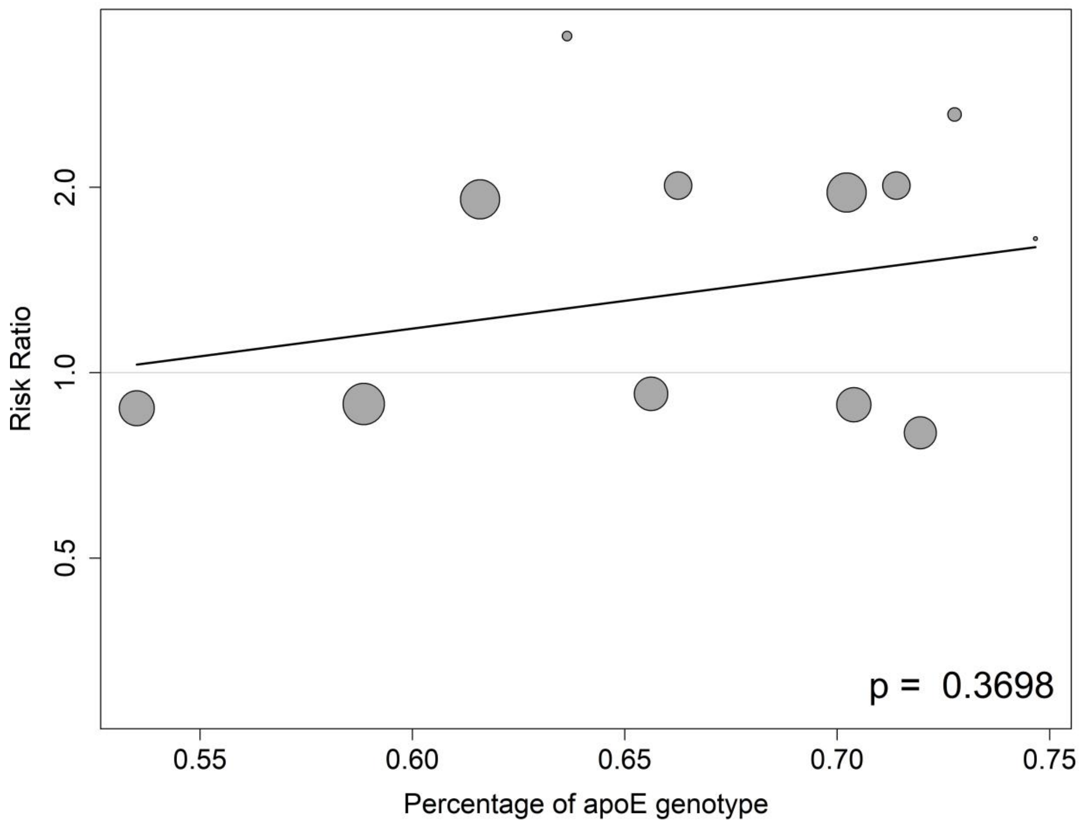

**8.20.5 Meta-regressions of ARIA-H by the percentage of female participants at the baseline**

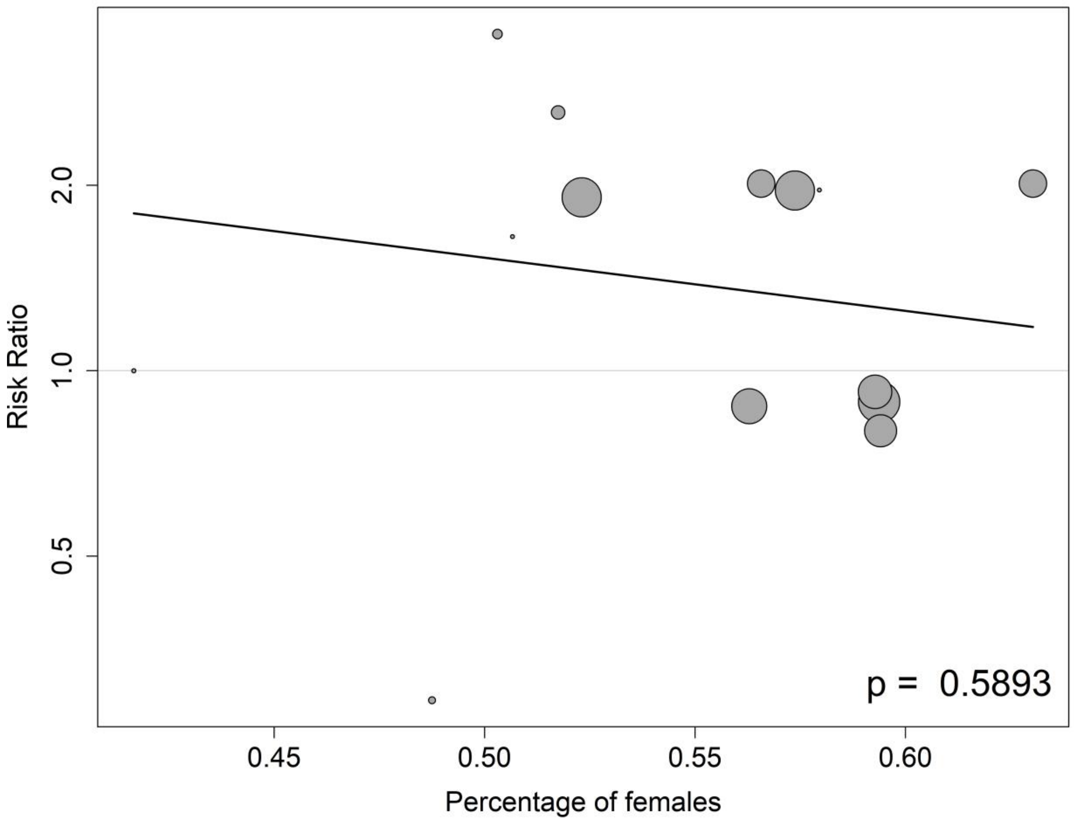

##### 8.20.6 Meta-regressions of ARIA-H by the percentage of participants on anti-AD medication at the baseline

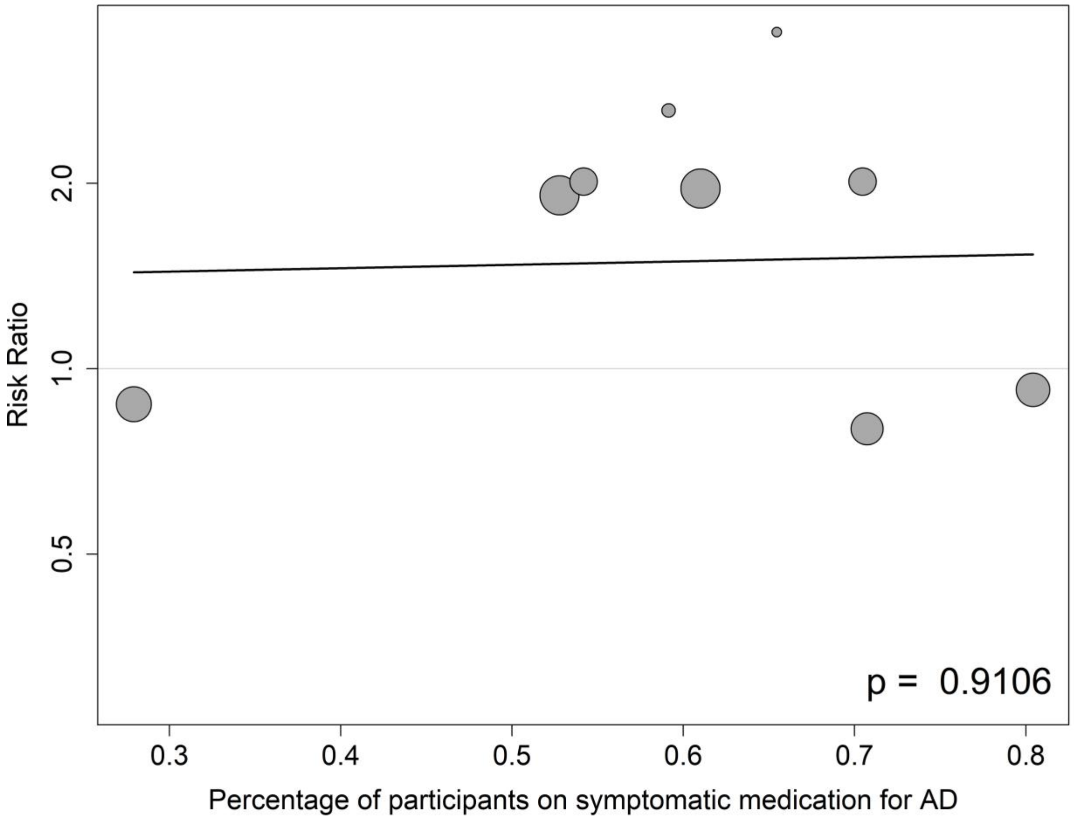

### Appendix 9: Sensitivity Analysis

#### 9.1 ADAS-cog

##### 9.1.1 Leave-one-out method

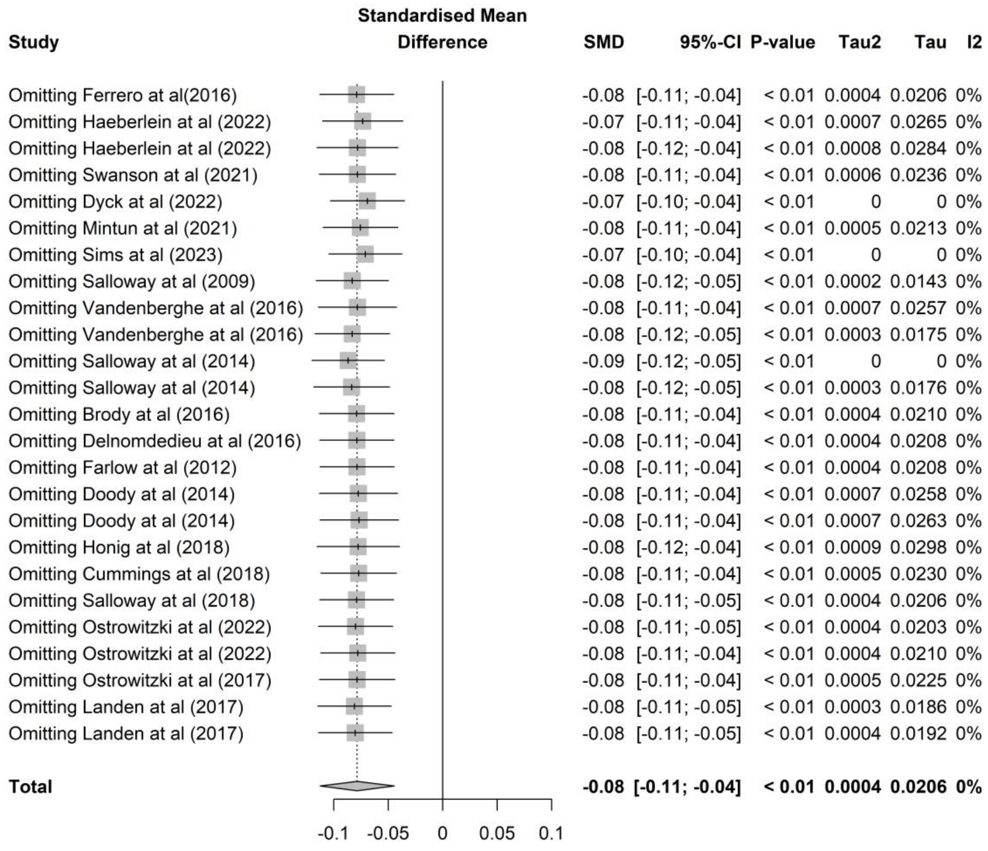

#### 9.1.2 Comparing the results between the fixed-effect and random-effects models

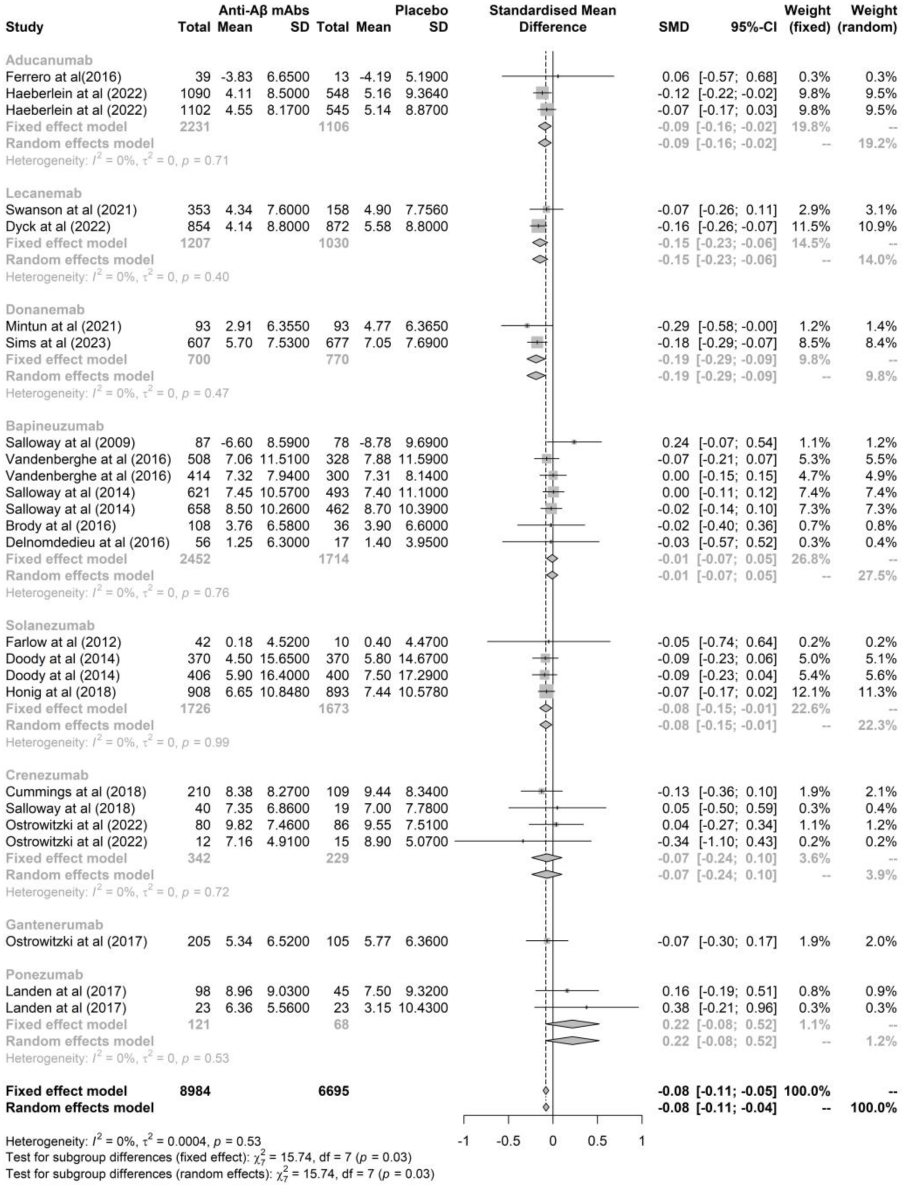

#### 9.2 CDR-SB

##### 9.2.1 Leave-one-out method

| Study | SMD | 95%-CI | P-value | Tau2 | Tau | I2 |
|---|---|---|---|---|---|---|
| Omitting Haeberlein at al (2022) | -0.06 | [-0.12; 0.00] | 0.07 | 0.0096 | 0.0978 | 64% |
| Omitting Haeberlein at al (2022) | -0.06 | [-0.12; -0.00] | 0.05 | 0.0099 | 0.0996 | 64% |
| Omitting Jeff sevigny at al (2016) | -0.06 | [-0.11; -0.00] | 0.05 | 0.0089 | 0.0943 | 64% |
| Omitting Swanson at al (2021) | -0.06 | [-0.12; -0.00] | 0.05 | 0.0095 | 0.0977 | 64% |
| Omitting Dyck at al (2022) | -0.05 | [-0.11; 0.01] | 0.08 | 0.0082 | 0.0904 | 60% |
| Omitting Mintun at al (2021) | -0.06 | [-0.11; 0.00] | 0.05 | 0.0090 | 0.0950 | 64% |
| Omitting Sims at al (2023) | -0.05 | [-0.10; 0.00] | 0.07 | 0.0049 | 0.0697 | 50% |
| Omitting Vandenberghe at al (2016) | -0.06 | [-0.12; -0.00] | 0.05 | 0.0094 | 0.0971 | 64% |
| Omitting Vandenberghe at al (2016) | -0.06 | [-0.12; -0.00] | 0.04 | 0.0097 | 0.0986 | 64% |
| Omitting Salloway at al (2014) | -0.07 | [-0.13; -0.01] | 0.02 | 0.0091 | 0.0953 | 62% |
| Omitting Salloway at al (2014) | -0.07 | [-0.13; -0.02] | 0.01 | 0.0076 | 0.0871 | 59% |
| Omitting Brody at al (2016) | -0.06 | [-0.12; -0.01] | 0.03 | 0.0089 | 0.0946 | 64% |
| Omitting Delnomdedieu at al (2016) | -0.07 | [-0.12; -0.01] | 0.02 | 0.0081 | 0.0903 | 60% |
| Omitting Doody at al (2014) | -0.07 | [-0.13; -0.01] | 0.02 | 0.0089 | 0.0944 | 63% |
| Omitting Doody at al (2014) | -0.06 | [-0.12; -0.00] | 0.05 | 0.0098 | 0.0989 | 64% |
| Omitting Honig at al (2018) | -0.06 | [-0.12; 0.00] | 0.06 | 0.0098 | 0.0990 | 64% |
| Omitting Sperling at al (2023) | -0.07 | [-0.13; -0.02] | 0.01 | 0.0079 | 0.0891 | 60% |
| Omitting Cummings at al (2018) | -0.06 | [-0.12; -0.00] | 0.05 | 0.0094 | 0.0967 | 64% |
| Omitting Salloway at al (2018) | -0.06 | [-0.12; -0.01] | 0.03 | 0.0088 | 0.0939 | 64% |
| Omitting Ostrowitzki at al (2022) | -0.06 | [-0.12; -0.01] | 0.03 | 0.0090 | 0.0950 | 64% |
| Omitting Ostrowitzki at al (2022) | -0.06 | [-0.11; -0.00] | 0.04 | 0.0088 | 0.0936 | 62% |
| Omitting Ostrowitzki at al (2017) | -0.07 | [-0.12; -0.01] | 0.02 | 0.0089 | 0.0944 | 64% |
| **Total** | **-0.06** | **[-0.12; -0.00]** | **0.03** | **0.0088** | **0.0940** | **63%** |

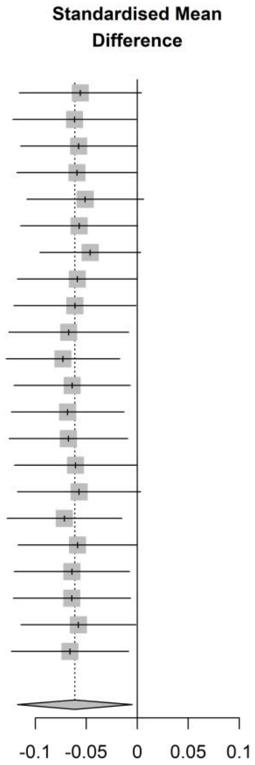

#### 9.2.2 Comparing the results between the fixed-effect and random-effects model

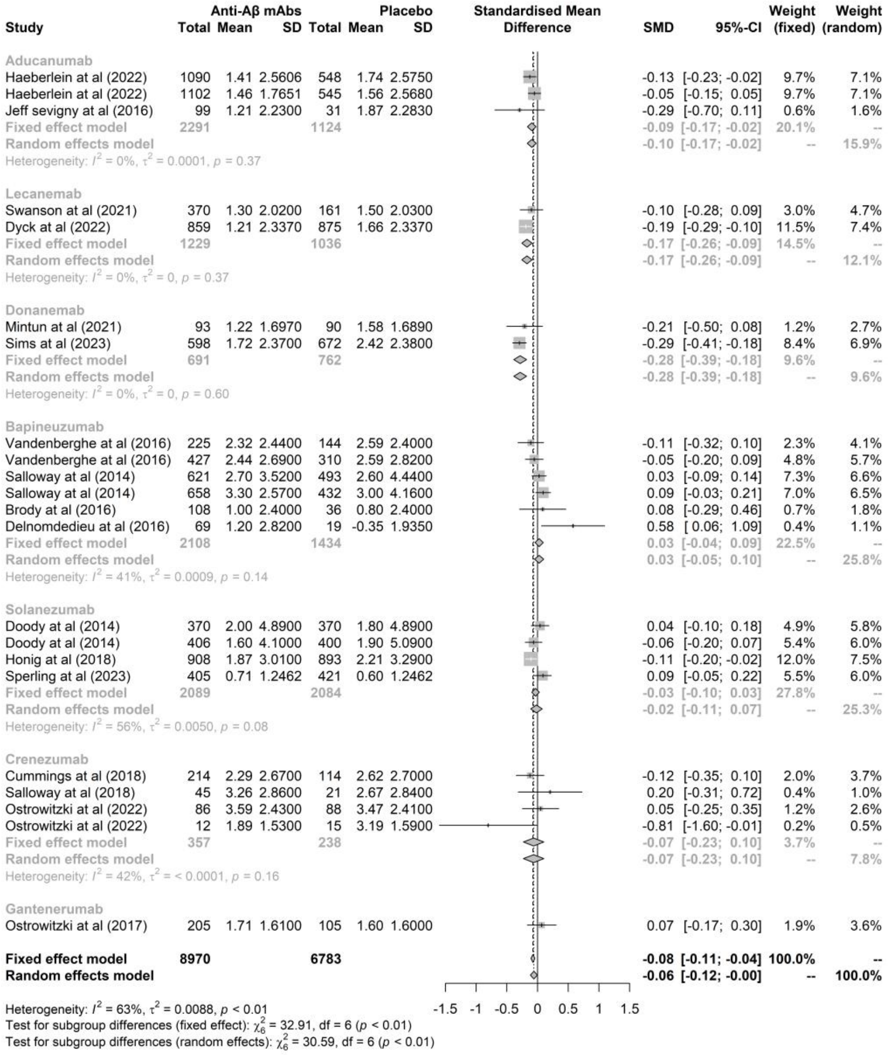

#### 9.3 MMSE

##### 9.3.1 Leave-one-out method

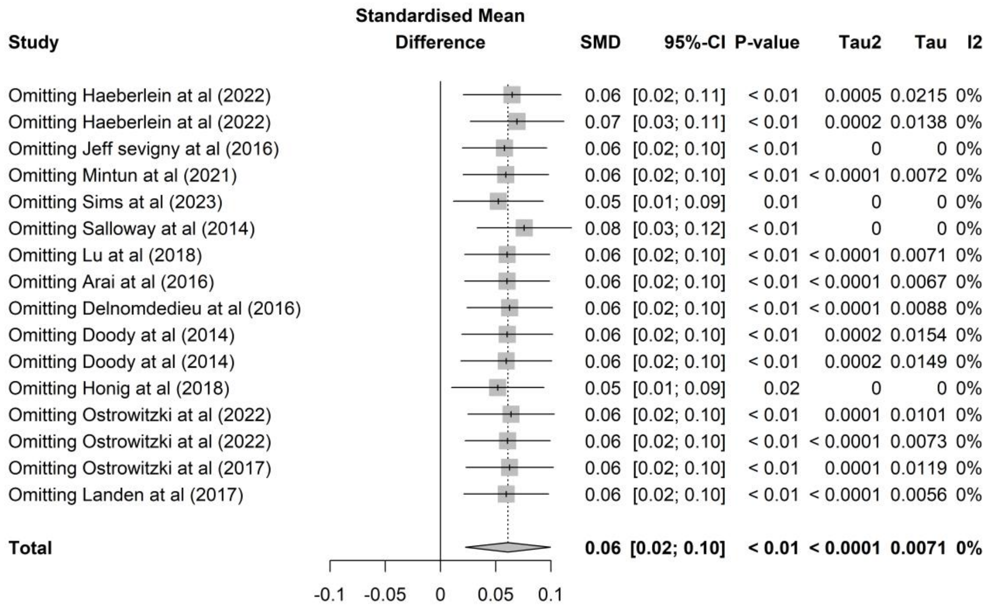

#### 9.3.2 Comparing the results between the fixed-effect and random-effects model

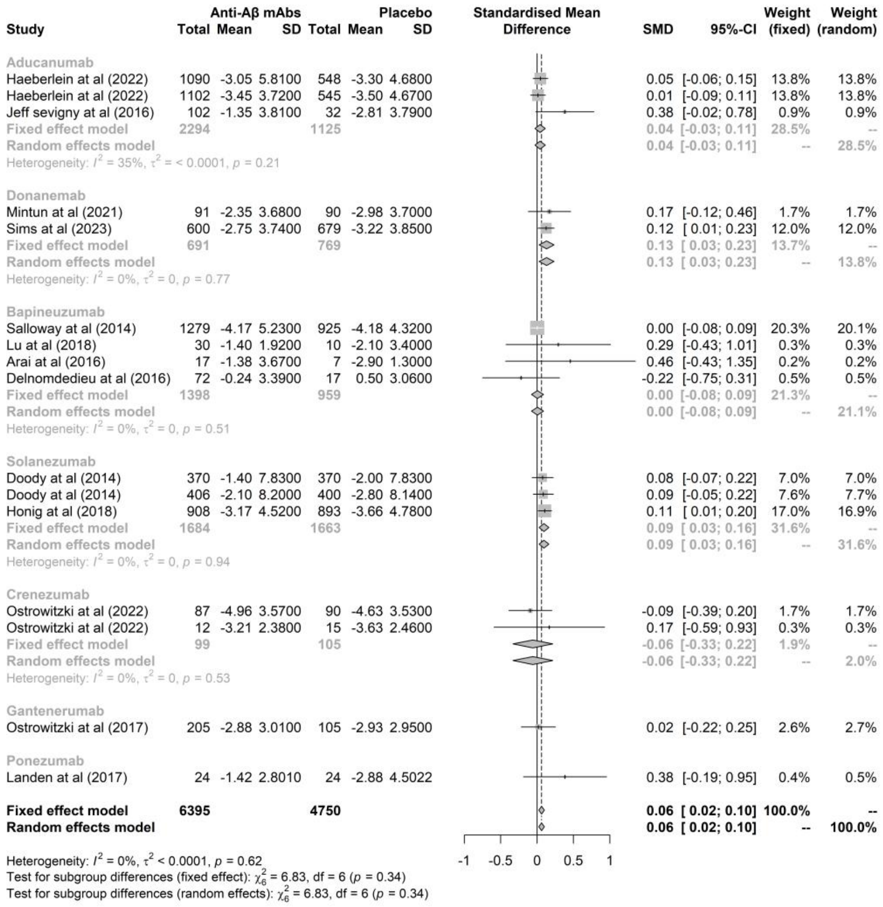

#### 9.4 Amyloid PET

##### 9.4.1 Leave-one-out method

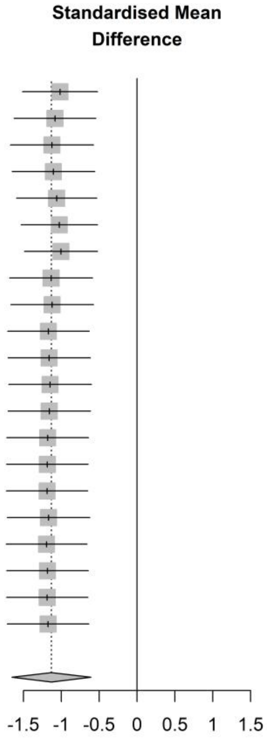

| Study | SMD | 95%-CI | P-value | Tau2 | Tau | I2 |
|---|---|---|---|---|---|---|
| Omitting Haeberlein at al (2022) | -1.02 | [-1.51; -0.52] | < 0.01 | 1.2074 | 1.0988 | 99% |
| Omitting Haeberlein at al (2022) | -1.08 | [-1.63; -0.54] | < 0.01 | 1.4471 | 1.2030 | 99% |
| Omitting Jeff sevigny at al (2016) | -1.12 | [-1.67; -0.58] | < 0.01 | 1.4918 | 1.2214 | 99% |
| Omitting Swanson at al (2021) | -1.10 | [-1.65; -0.56] | < 0.01 | 1.4797 | 1.2164 | 99% |
| Omitting Dyck at al (2022) | -1.06 | [-1.59; -0.53] | < 0.01 | 1.3904 | 1.1791 | 99% |
| Omitting Mintun at al (2021) | -1.03 | [-1.53; -0.52] | < 0.01 | 1.2504 | 1.1182 | 99% |
| Omitting Sims at al (2023) | -1.01 | [-1.49; -0.52] | < 0.01 | 1.1389 | 1.0672 | 98% |
| Omitting Lowe at al (2021) | -1.14 | [-1.68; -0.59] | < 0.01 | 1.4883 | 1.2200 | 99% |
| Omitting Rinne at al (2010) | -1.12 | [-1.67; -0.58] | < 0.01 | 1.4830 | 1.2178 | 99% |
| Omitting Vandenberghe at al (2016) | -1.17 | [-1.71; -0.63] | < 0.01 | 1.4501 | 1.2042 | 99% |
| Omitting Vandenberghe at al (2016) | -1.16 | [-1.70; -0.62] | < 0.01 | 1.4699 | 1.2124 | 99% |
| Omitting Salloway at al (2014) | -1.15 | [-1.70; -0.60] | < 0.01 | 1.4830 | 1.2178 | 99% |
| Omitting Salloway at al (2014) | -1.16 | [-1.71; -0.61] | < 0.01 | 1.4760 | 1.2149 | 99% |
| Omitting Brody at al (2016) | -1.18 | [-1.72; -0.64] | < 0.01 | 1.4414 | 1.2006 | 99% |
| Omitting Doody at al (2014_ | -1.18 | [-1.72; -0.65] | < 0.01 | 1.4346 | 1.1977 | 99% |
| Omitting Honig at al (2018) | -1.19 | [-1.73; -0.65] | < 0.01 | 1.4270 | 1.1946 | 99% |
| Omitting Sperling at al (2023) | -1.17 | [-1.71; -0.62] | < 0.01 | 1.4648 | 1.2103 | 99% |
| Omitting Salloway at al (2018) | -1.20 | [-1.73; -0.66] | < 0.01 | 1.4063 | 1.1859 | 99% |
| Omitting Ostrowitzki at al (2022) | -1.18 | [-1.72; -0.64] | < 0.01 | 1.4391 | 1.1996 | 99% |
| Omitting Ostrowitzki at al (2017) | -1.19 | [-1.72; -0.65] | < 0.01 | 1.4272 | 1.1947 | 99% |
| Omitting Landen at al (2017) | -1.17 | [-1.71; -0.64] | < 0.01 | 1.4443 | 1.2018 | 99% |
| **Total** | **-1.13** | **[-1.66; -0.61]** | **< 0.01** | **1.4164** | **1.1901** | **99%** |

##### 9.4.2 Comparing the results between the fixed-effect and random-effects model

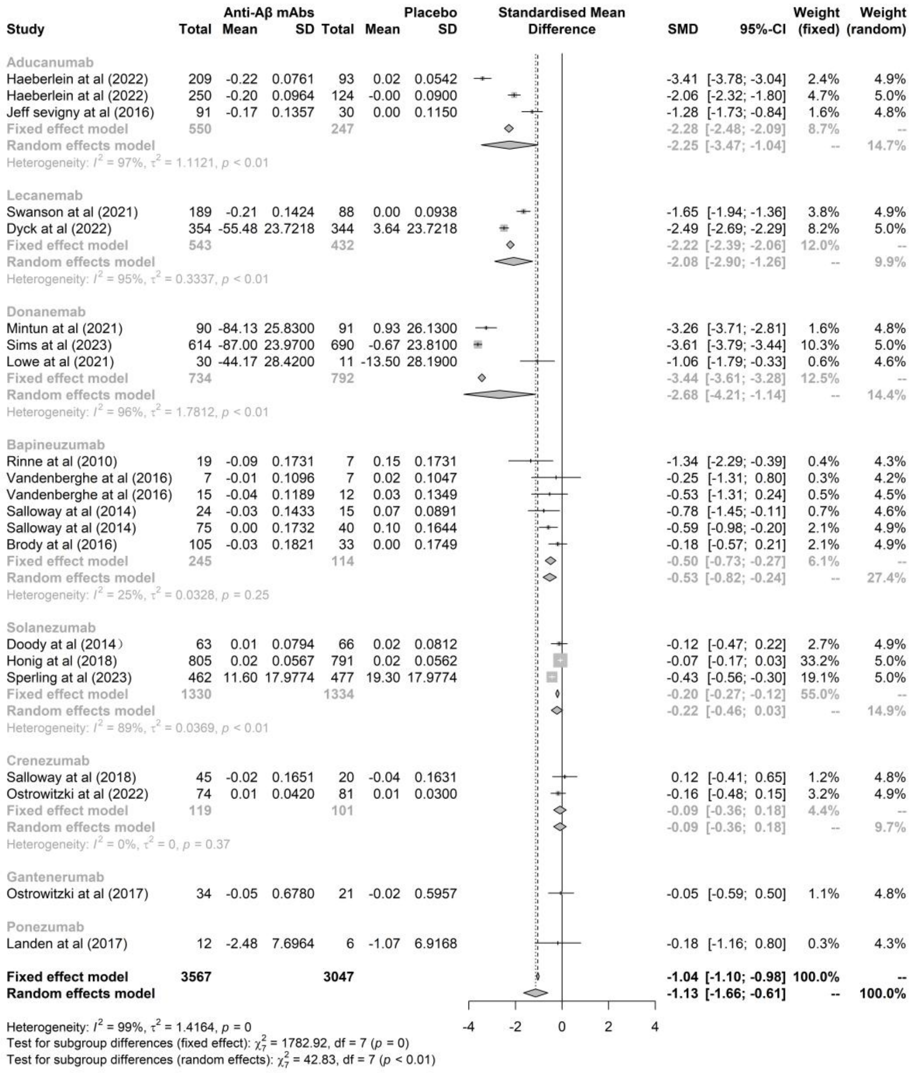

#### 9.5 Tau PET

##### 9.5.1 Leave-one-out method

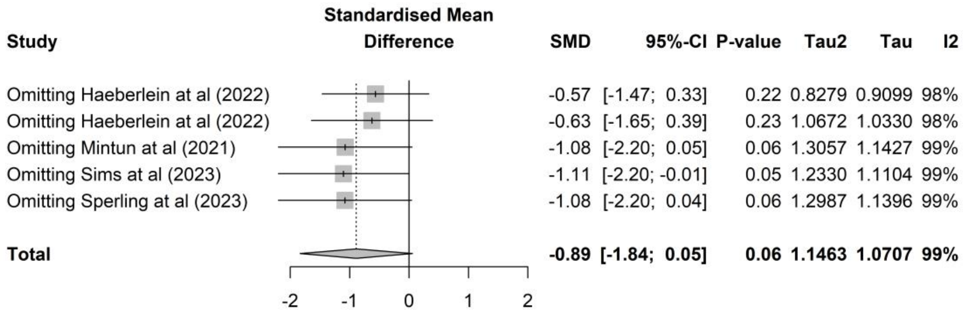

#### 9.5.2 Comparing the results between the fixed-effect and random-effects model

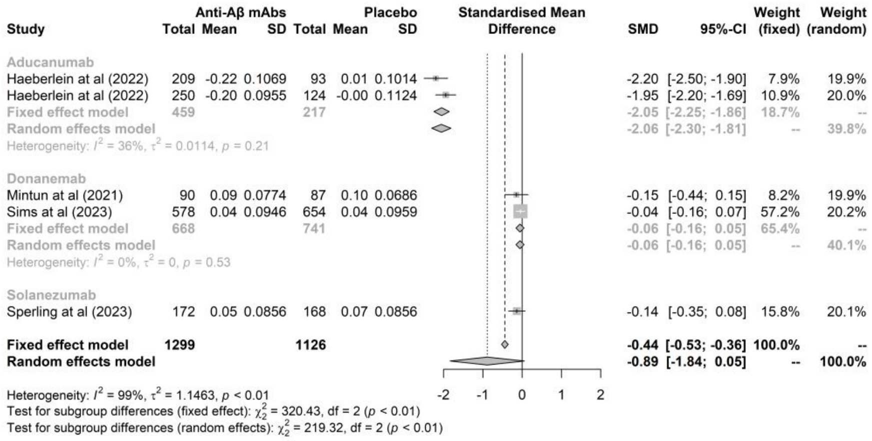

#### 9.6 Volumes of the whole brain

##### 9.6.1 Leave-one-out method

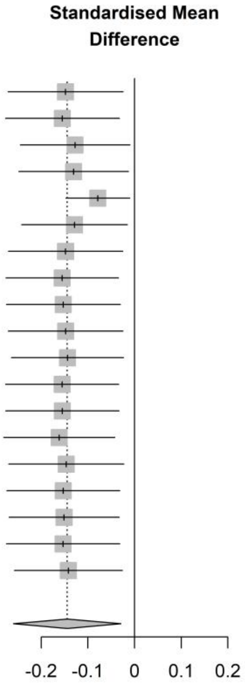

| Study | SMD | 95%-CI | P-value | Tau2 | Tau | I2 |
|---|---|---|---|---|---|---|
| Omitting Haeberlein at al (2022) | -0.15 | [-0.27; -0.02] | 0.02 | 0.0527 | 0.2295 | 90% |
| Omitting Haeberlein at al (2022) | -0.15 | [-0.28; -0.03] | 0.01 | 0.0513 | 0.2266 | 89% |
| Omitting Swanson at al (2021) | -0.13 | [-0.25; -0.01] | 0.03 | 0.0472 | 0.2172 | 89% |
| Omitting Mintun at al (2021) | -0.13 | [-0.25; -0.01] | 0.03 | 0.0480 | 0.2191 | 89% |
| Omitting Sims at al (2023) | -0.08 | [-0.15; -0.01] | 0.02 | 0.0098 | 0.0990 | 56% |
| Omitting Lowe at al (2021) | -0.13 | [-0.24; -0.02] | 0.03 | 0.0456 | 0.2135 | 89% |
| Omitting Vandenberghe at al (2016) | -0.15 | [-0.27; -0.03] | 0.02 | 0.0523 | 0.2286 | 90% |
| Omitting Vandenberghe at al (2016) | -0.16 | [-0.28; -0.03] | 0.01 | 0.0506 | 0.2250 | 89% |
| Omitting Salloway at al (2014) | -0.15 | [-0.27; -0.03] | 0.01 | 0.0517 | 0.2274 | 89% |
| Omitting Salloway at al (2014) | -0.15 | [-0.27; -0.02] | 0.02 | 0.0525 | 0.2291 | 90% |
| Omitting Brody at al (2016) | -0.14 | [-0.26; -0.02] | 0.02 | 0.0510 | 0.2259 | 90% |
| Omitting Doody at al (2014) | -0.16 | [-0.28; -0.03] | 0.01 | 0.0509 | 0.2256 | 89% |
| Omitting Doody at al (2014) | -0.15 | [-0.28; -0.03] | 0.01 | 0.0510 | 0.2259 | 89% |
| Omitting Honig at al (2018) | -0.16 | [-0.28; -0.04] | < 0.01 | 0.0482 | 0.2197 | 88% |
| Omitting Sperling at al (2023) | -0.15 | [-0.27; -0.02] | 0.02 | 0.0527 | 0.2296 | 90% |
| Omitting Cummings at al (2018) | -0.15 | [-0.27; -0.03] | 0.01 | 0.0512 | 0.2263 | 89% |
| Omitting Salloway at al (2018) | -0.15 | [-0.27; -0.03] | 0.01 | 0.0498 | 0.2231 | 90% |
| Omitting Ostrowitzki at al (2022) | -0.15 | [-0.27; -0.03] | 0.01 | 0.0514 | 0.2266 | 89% |
| Omitting Ostrowitzki at al (2022) | -0.14 | [-0.26; -0.03] | 0.02 | 0.0488 | 0.2209 | 90% |
| **Total** | **-0.14** | **[-0.26; -0.03]** | **0.01** | **0.0486** | **0.2204** | **89%** |

#### 9.6.2 Comparing the results between the fixed-effect and random-effects model

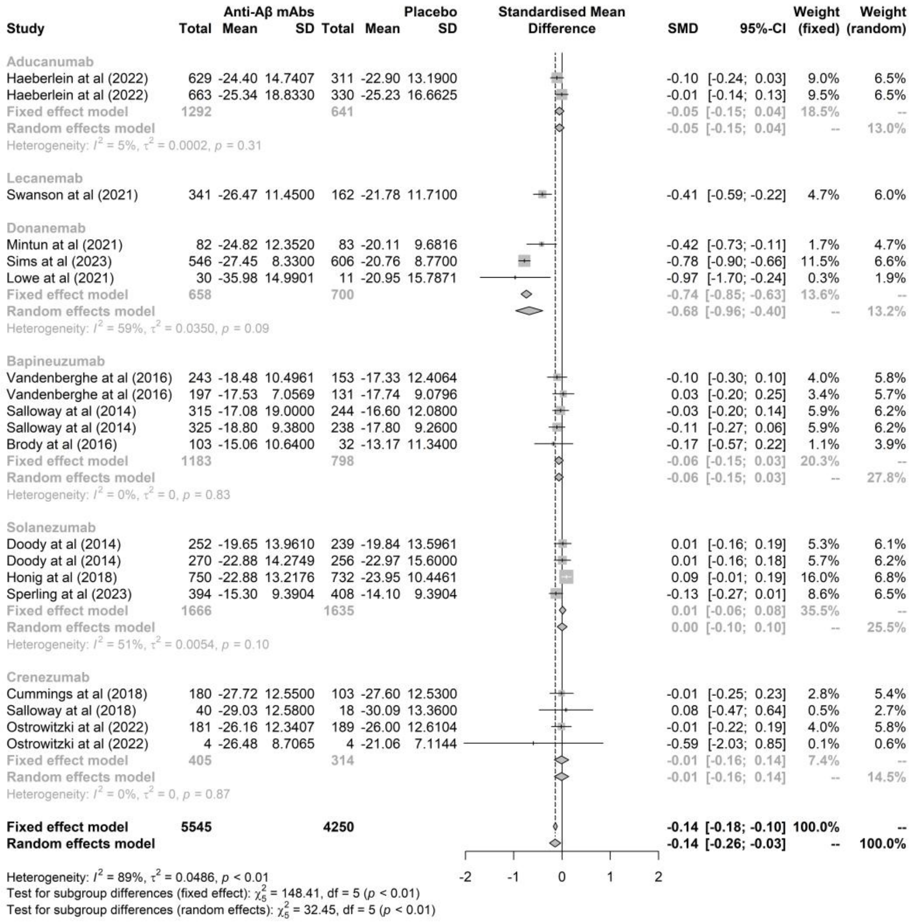

#### 9.7 Volumes of hippocampus

##### 9.7.1 Leave-one-out method

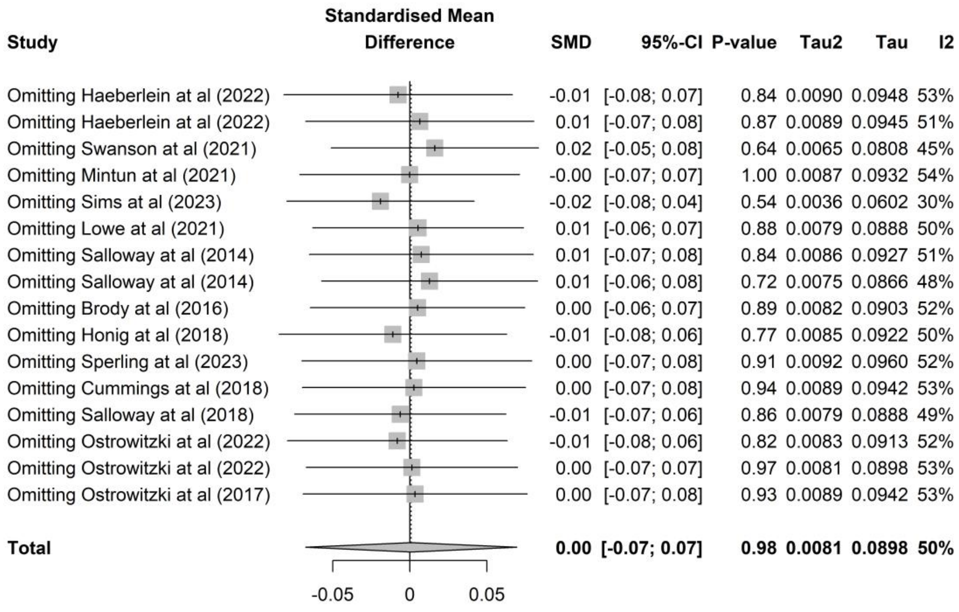

#### 9.7.2 Comparing the results between the fixed-effect and random-effects model

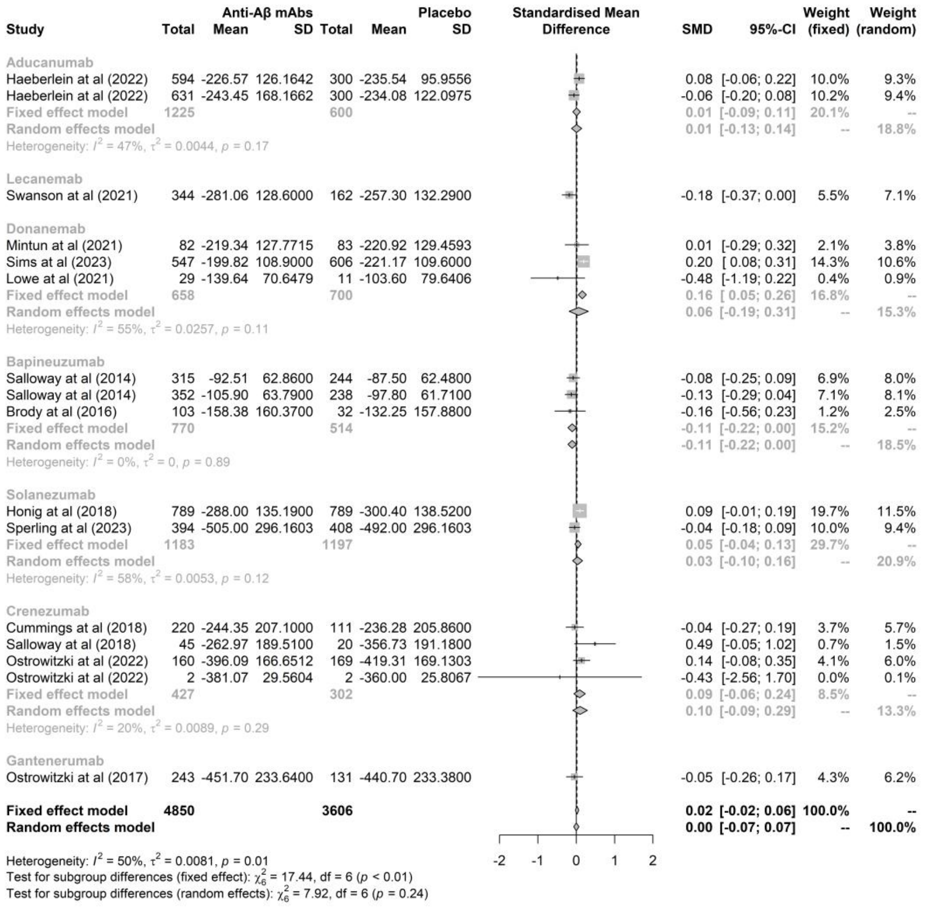

#### 9.8 Volumes of ventricle

##### 9.8.1 Leave-one-out method

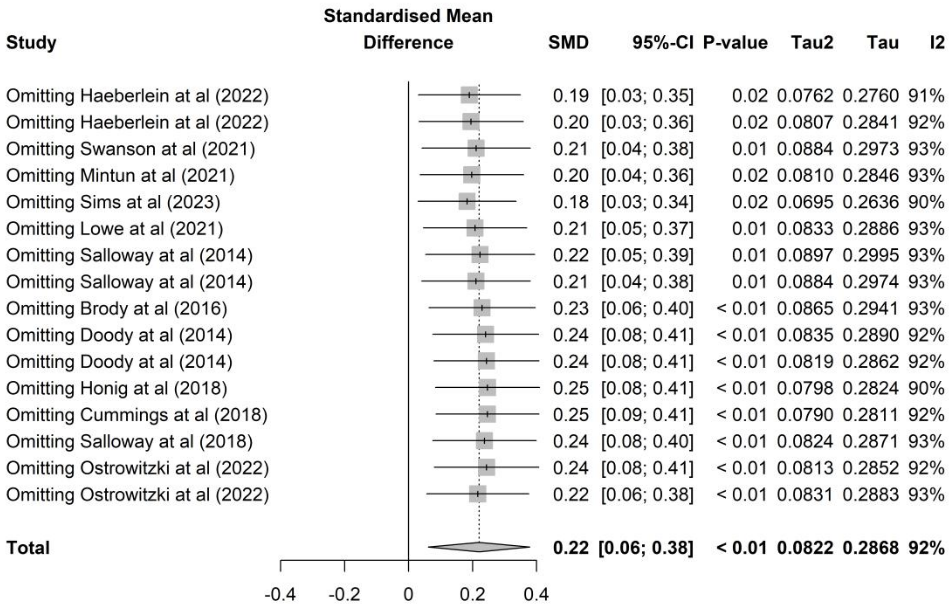

#### 9.8.2 Comparing the results between the fixed-effect and random-effects model

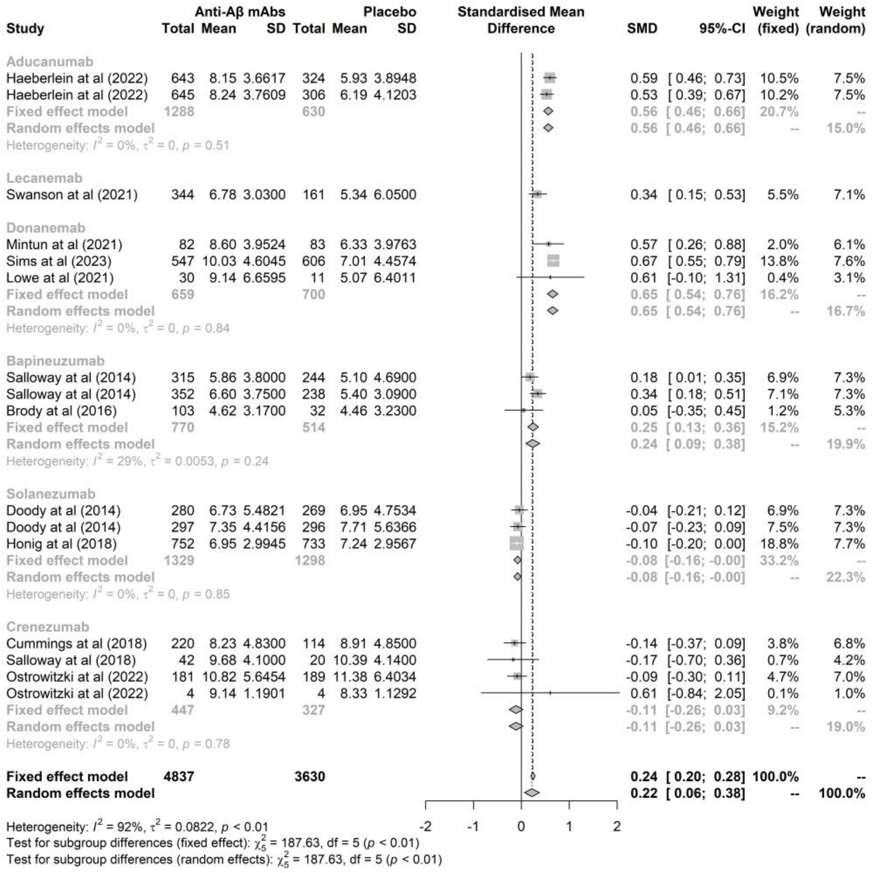

#### 9.9 CSF Aβ40

##### 9.9.1 Leave-one-out method

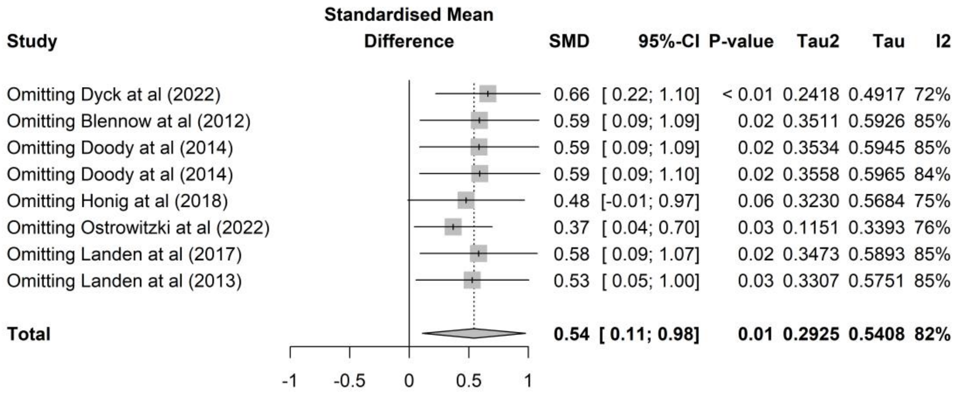

#### 9.9.2 Comparing the results between the fixed-effect and random-effects model

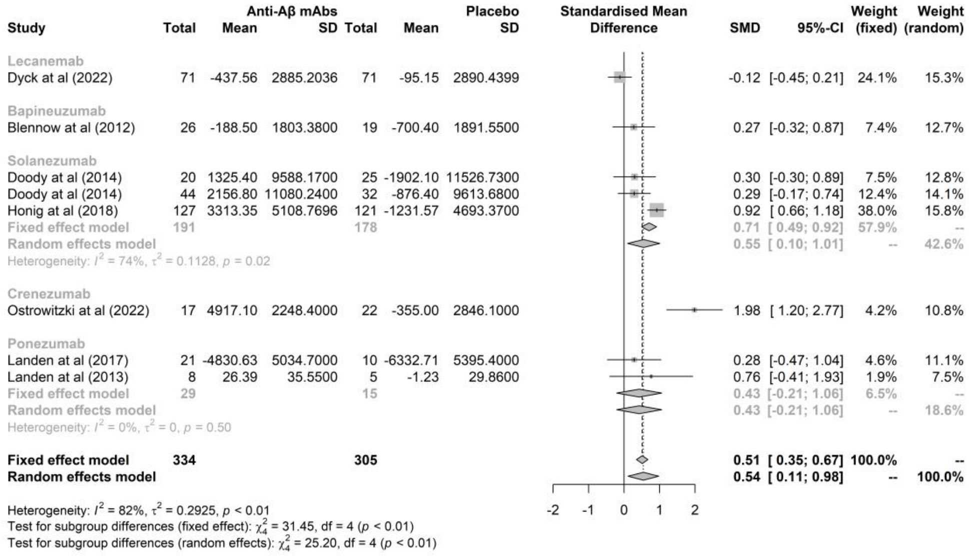

#### 9.10 CSF Aβ42

##### 9.10.1 Leave-one-out method

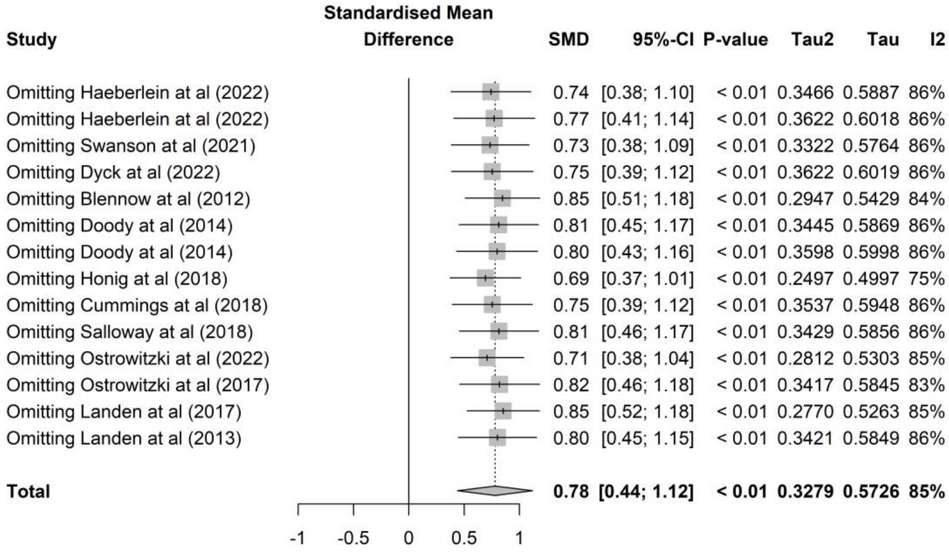

#### 9.10.2 Comparing the results between the fixed-effect and random-effects model

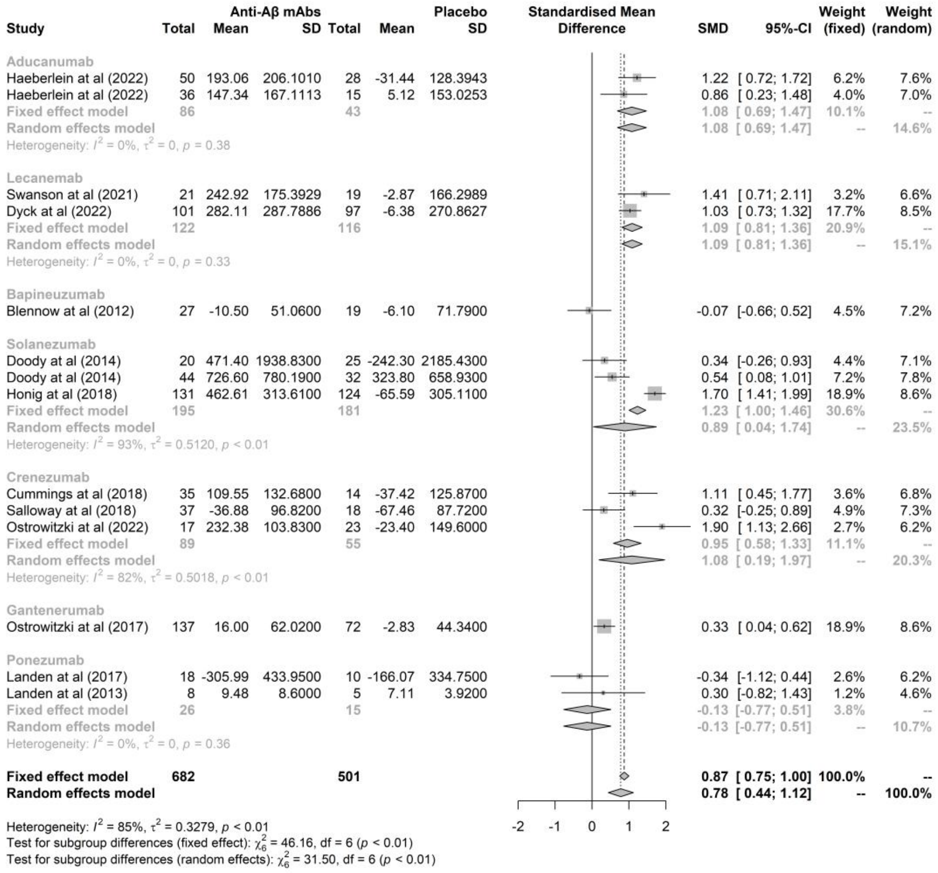

#### 9.11 CSF p-tau

##### 9.11.1 Leave-one-out method

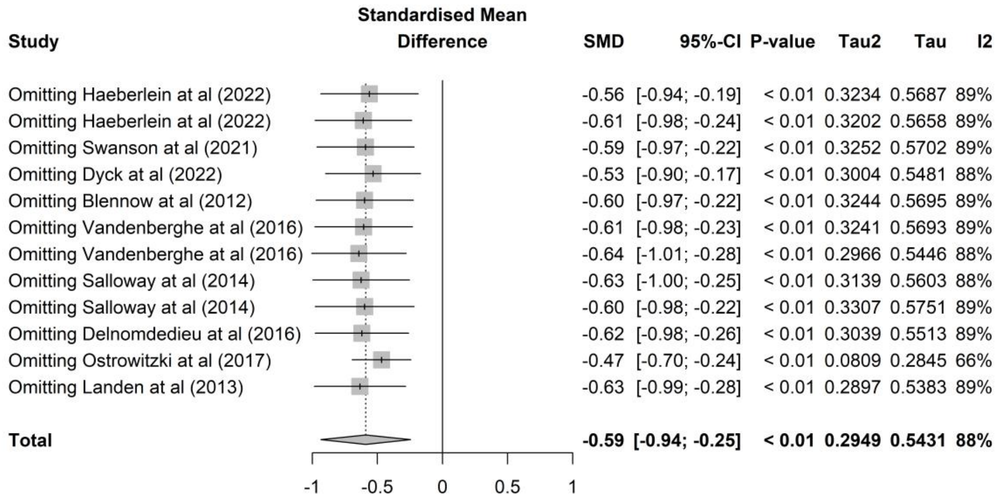

##### 9.11.2 Comparing the results between the fixed-effect and random-effects model

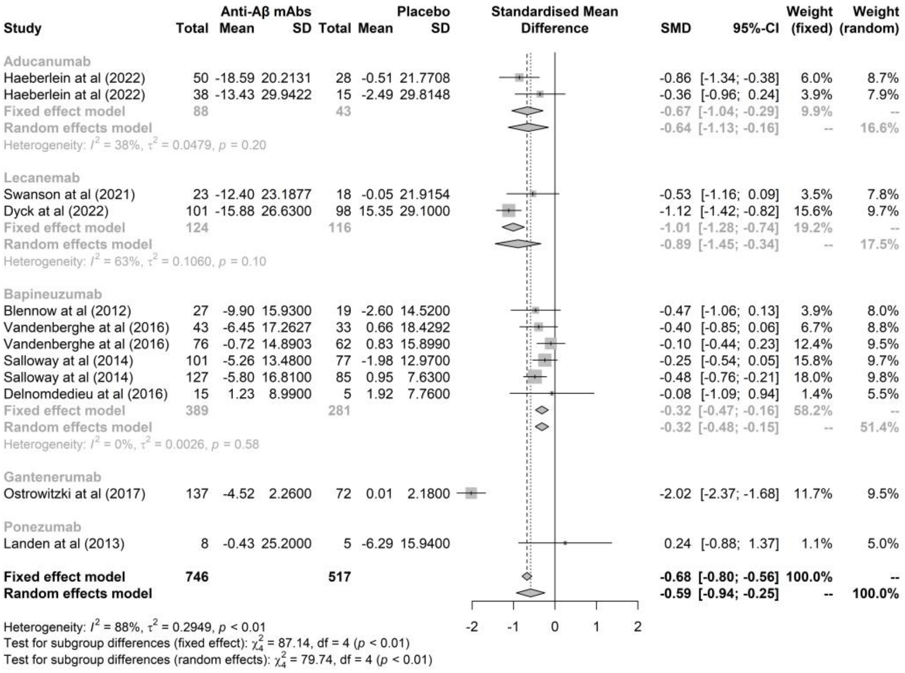

### 9.12 CSF total tau

#### 9.12.1 Leave-one-out method

| Study | SMD | 95%-CI | P-value | Tau2 | Tau | I2 |
|---|---|---|---|---|---|---|
| Omitting Haeberlein at al (2022) | -0.41 | [-0.72; -0.10] | 0.01 | 0.1522 | 0.3901 | 71% |
| Omitting Haeberlein at al (2022) | -0.46 | [-0.77; -0.16] | < 0.01 | 0.1445 | 0.3801 | 70% |
| Omitting Swanson at al (2021) | -0.49 | [-0.78; -0.20] | < 0.01 | 0.1241 | 0.3522 | 67% |
| Omitting Dyck at al (2022) | -0.42 | [-0.74; -0.10] | 0.01 | 0.1597 | 0.3996 | 71% |
| Omitting Blennow at al (2012) | -0.44 | [-0.75; -0.13] | < 0.01 | 0.1527 | 0.3908 | 71% |
| Omitting Delnomdedieu at al (2016) | -0.42 | [-0.72; -0.12] | < 0.01 | 0.1429 | 0.3780 | 71% |
| Omitting Cummings at al (2018) | -0.42 | [-0.73; -0.11] | < 0.01 | 0.1505 | 0.3879 | 71% |
| Omitting Salloway at al (2018) | -0.53 | [-0.80; -0.26] | < 0.01 | 0.0980 | 0.3130 | 61% |
| Omitting Ostrowitzki at al (2022) | -0.46 | [-0.77; -0.16] | < 0.01 | 0.1448 | 0.3805 | 70% |
| Omitting Ostrowitzki at al (2017) | -0.36 | [-0.57; -0.14] | < 0.01 | 0.0299 | 0.1728 | 21% |
| Omitting Landen at al (2013) | -0.47 | [-0.76; -0.18] | < 0.01 | 0.1349 | 0.3672 | 70% |
| **Total** | **-0.44** | **[-0.73; -0.16]** | **< 0.01** | **0.1330** | **0.3647** | **68%** |

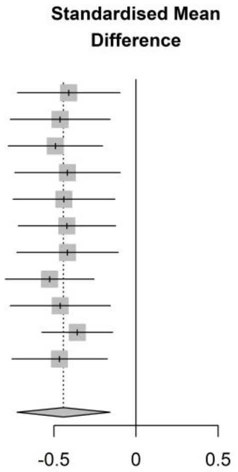

#### 9.12.2 Comparing the results between the fixed-effect and random-effects model

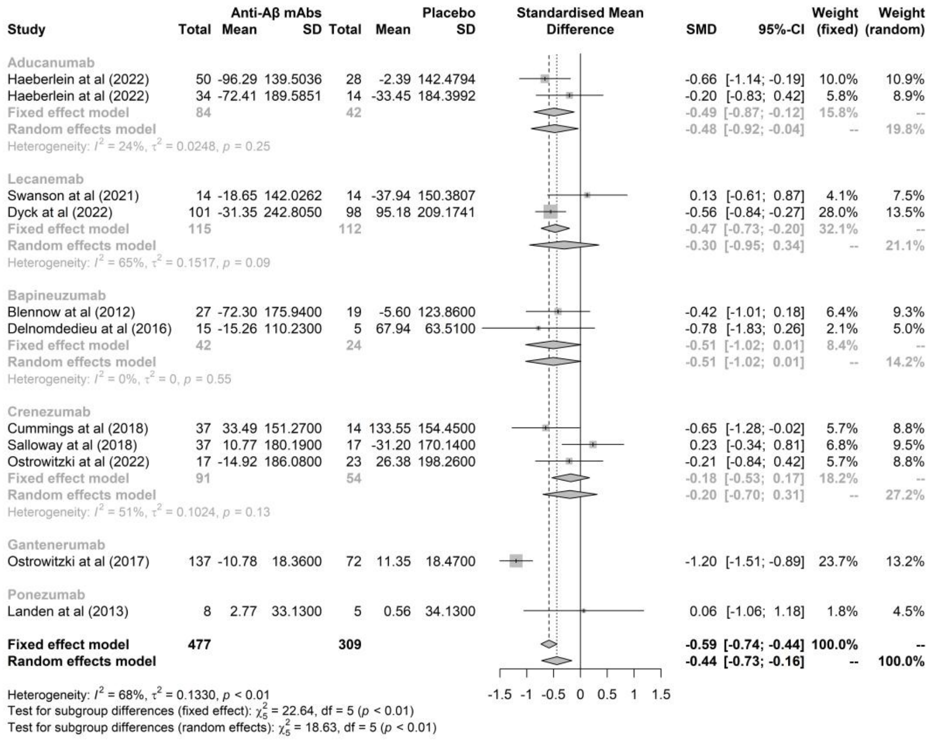

#### 9.13 CSF p-tau181

##### 9.13.1 Leave-one-out method

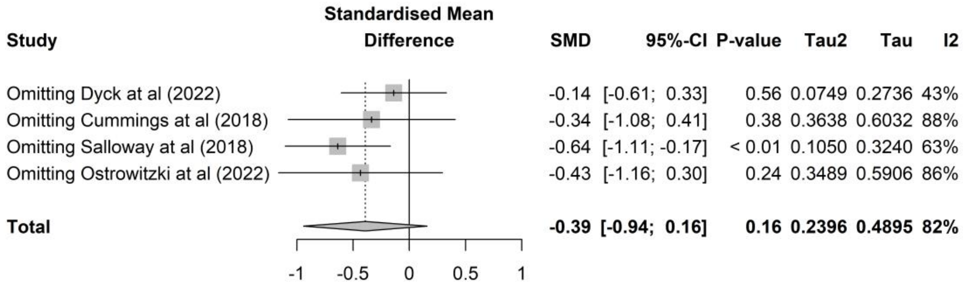

#### 9.13.2 Comparing the results between the fixed-effect and random-effects model

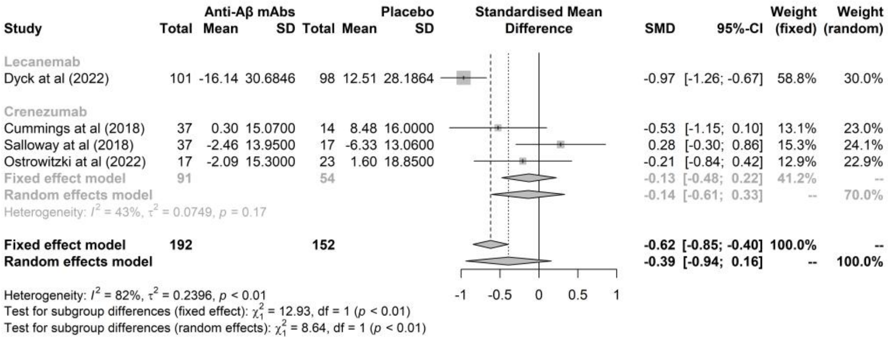

#### 9.14 CSF Nfl

##### 9.14.1 Leave-one-out method

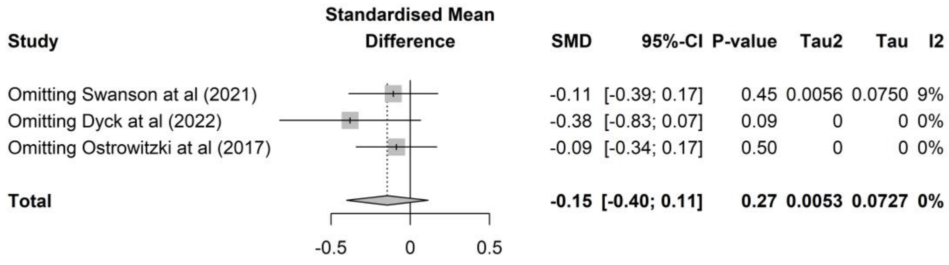

##### 9.14.2 Comparing the results between the fixed-effect and random-effects model

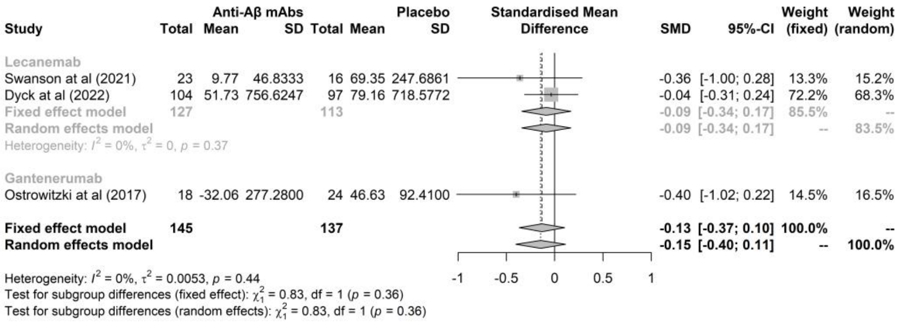

#### 9.15 CSF Neurogranin

##### 9.15.1 Leave-one-out method

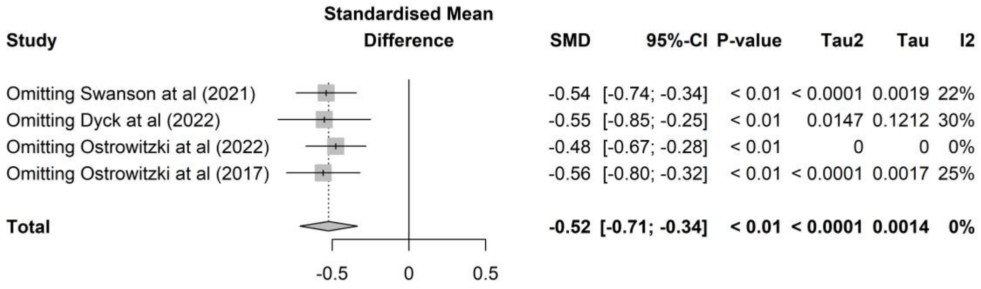

#### 9.15.2 Comparing the results between the fixed-effect and random-effects model

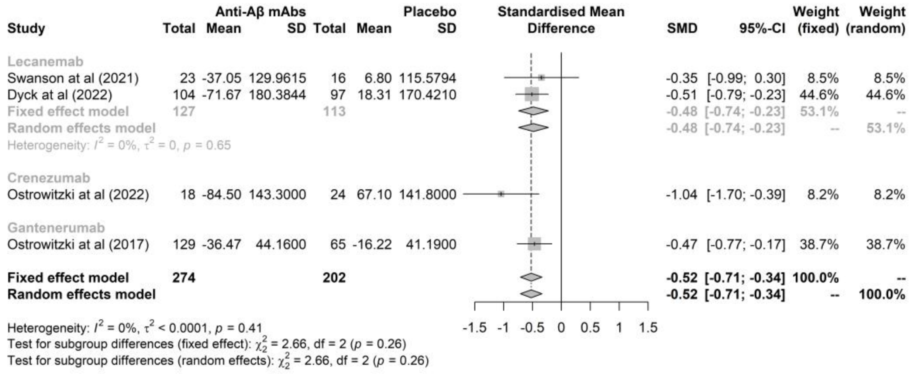

#### 9.16 Plasma Aβ40

##### 9.16.1 Leave-one-out method

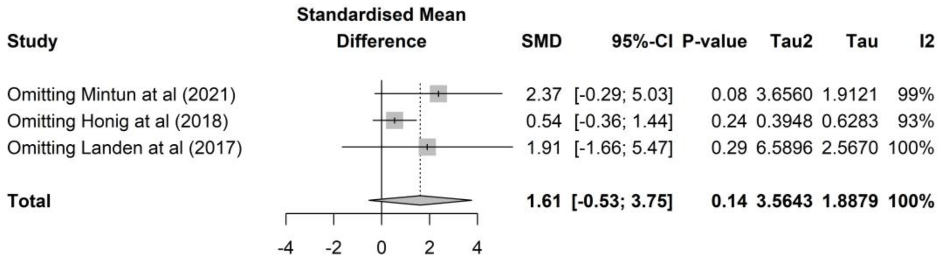

##### 9.16.2 Comparing the results between the fixed-effect and random-effects model

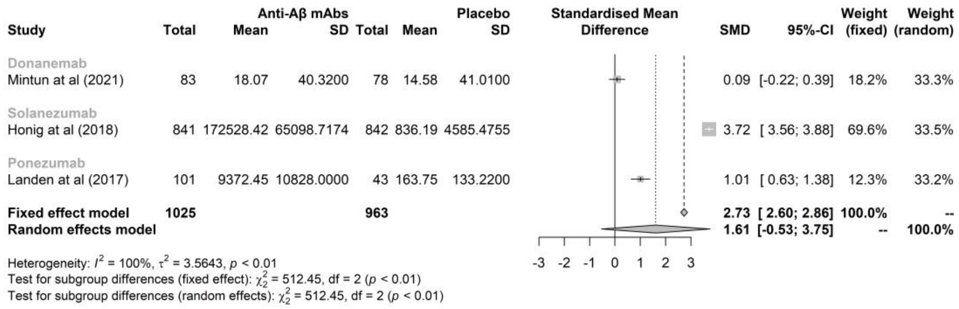

#### 9.17 Plasma Nfl

##### 9.17.1 Leave-one-out method

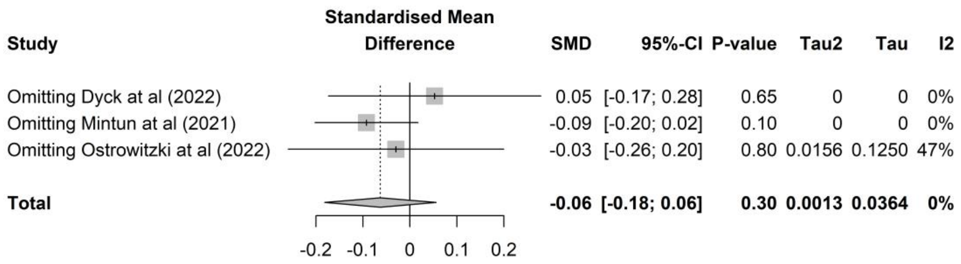

#### 9.17.2 Comparing the results between the fixed-effect and random-effects model

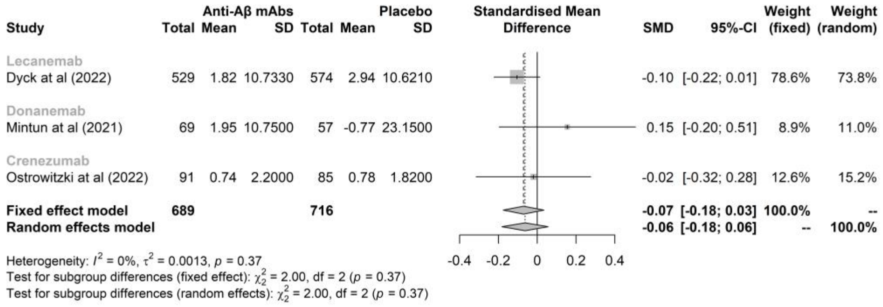

#### 9.18 Plasma p-tau181

##### 9.18.1 Leave-one-out method

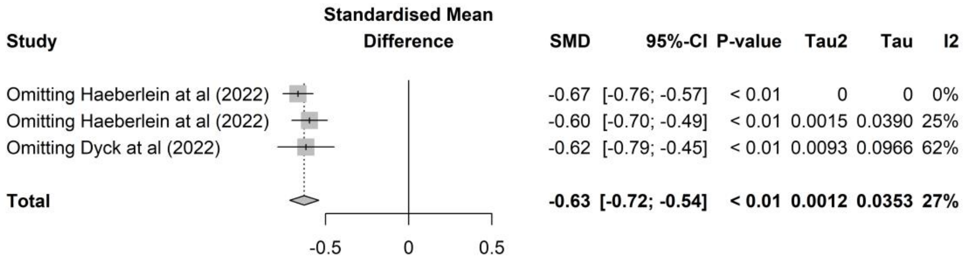

##### 9.18.2 Comparing the results between the fixed-effect and random-effects model

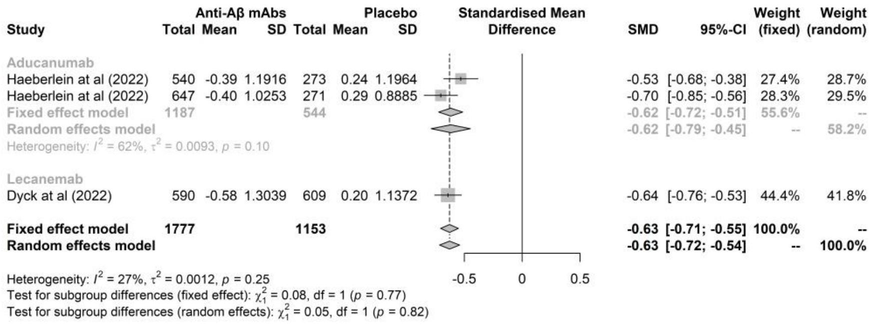

#### 9.19 ARIA-E

##### 9.19.1 Leave-one-out method

| Study | RR | 95%-CI | P-value | Tau2 | Tau | I2 |
| --- | --- | --- | --- | --- | --- | --- |
| Omitting Haeberlein at al (2022) | 9.26 | [7.10; 12.08] | < 0.01 | 0.0246 | 0.1568 | 40% |
| Omitting Haeberlein at al (2022) | 9.41 | [6.82; 12.96] | < 0.01 | 0.0836 | 0.2891 | 42% |
| Omitting Jeff sevigny at al (2016) | 9.74 | [7.75; 12.24] | < 0.01 | 0.0089 | 0.0945 | 42% |
| Omitting Swanson at al (2021) | 9.77 | [7.72; 12.38] | < 0.01 | 0.0138 | 0.1176 | 42% |
| Omitting Dyck at al (2022) | 10.36 | [8.18; 13.14] | < 0.01 | < 0.0001 | 0.0019 | 40% |
| Omitting Logovinsky at al (2016) | 9.79 | [7.83; 12.26] | < 0.01 | 0.0063 | 0.0797 | 39% |
| Omitting Mintun at al (2021) | 9.61 | [7.64; 12.10] | < 0.01 | 0.0096 | 0.0981 | 39% |
| Omitting Sims at al (2023) | 9.22 | [7.06; 12.04] | < 0.01 | 0.0213 | 0.1460 | 40% |
| Omitting Lowe at al (2021) | 9.81 | [7.82; 12.30] | < 0.01 | 0.0075 | 0.0868 | 42% |
| Omitting Vandenberghe at al (2016) | 9.65 | [7.61; 12.24] | < 0.01 | 0.0151 | 0.1230 | 42% |
| Omitting Vandenberghe at al (2016) | 9.91 | [7.61; 12.91] | < 0.01 | 0.0292 | 0.1710 | 42% |
| Omitting Salloway at al (2014) | 9.59 | [7.63; 12.06] | < 0.01 | 0.0087 | 0.0933 | 38% |
| Omitting Salloway at al (2014) | 9.55 | [7.62; 11.96] | < 0.01 | 0.0059 | 0.0767 | 33% |
| Omitting Delnomdedieu at al (2016) | 9.91 | [7.94; 12.37] | < 0.01 | 0.0035 | 0.0588 | 39% |
| Omitting Farlow at al (2012) | 9.79 | [7.83; 12.26] | < 0.01 | 0.0063 | 0.0797 | 39% |
| Omitting Doody at al (2014） | 10.35 | [8.30; 12.90] | < 0.01 | < 0.0001 | 0.0027 | 25% |
| Omitting Sperling at al (2023) | 10.07 | [8.10; 12.51] | < 0.01 | < 0.0001 | 0.0024 | 27% |
| Omitting Salloway at al (2018) | 9.79 | [7.83; 12.26] | < 0.01 | 0.0063 | 0.0797 | 39% |
| Omitting Ostrowitzki at al (2022) | 9.96 | [8.00; 12.40] | < 0.01 | 0.0014 | 0.0372 | 36% |
| Omitting Ostrowitzki at al (2022) | 9.85 | [7.88; 12.33] | < 0.01 | 0.0057 | 0.0755 | 41% |
| Omitting Guthrie at al (2020) | 9.79 | [7.83; 12.26] | < 0.01 | 0.0063 | 0.0797 | 39% |
| Omitting Ostrowitzki at al (2017) | 9.67 | [7.62; 12.27] | < 0.01 | 0.0154 | 0.1240 | 42% |
| **Total** | **9.79** | **[7.83; 12.26]** | **< 0.01** | **0.0063** | **0.0797** | **39%** |

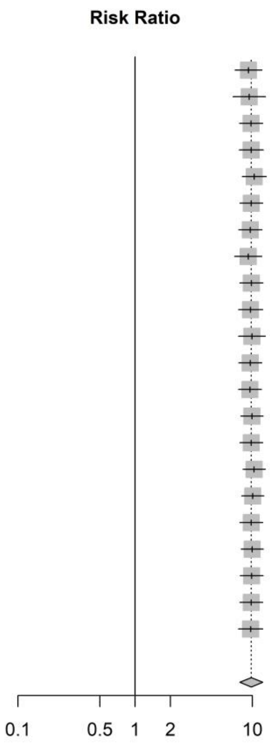

#### 9.19.2 Comparing the results between the fixed-effect and random-effects model

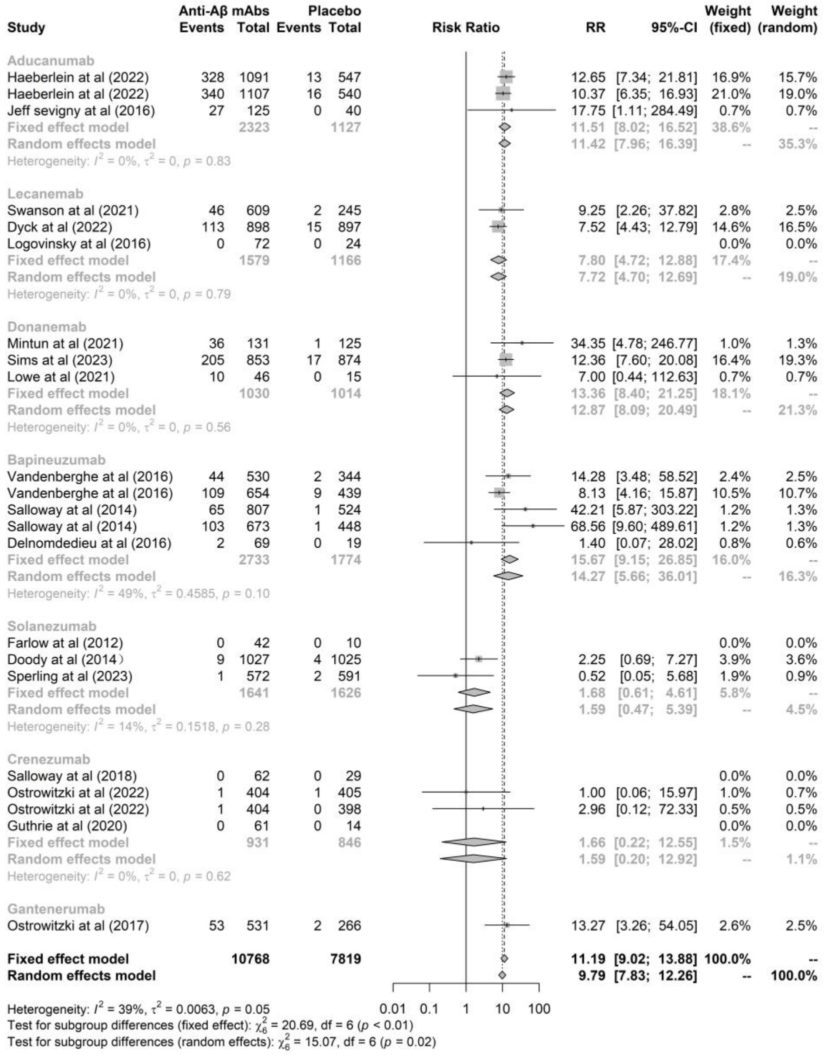

#### 9.20 ARIA-H

##### 9.20.1 Leave-one-out method

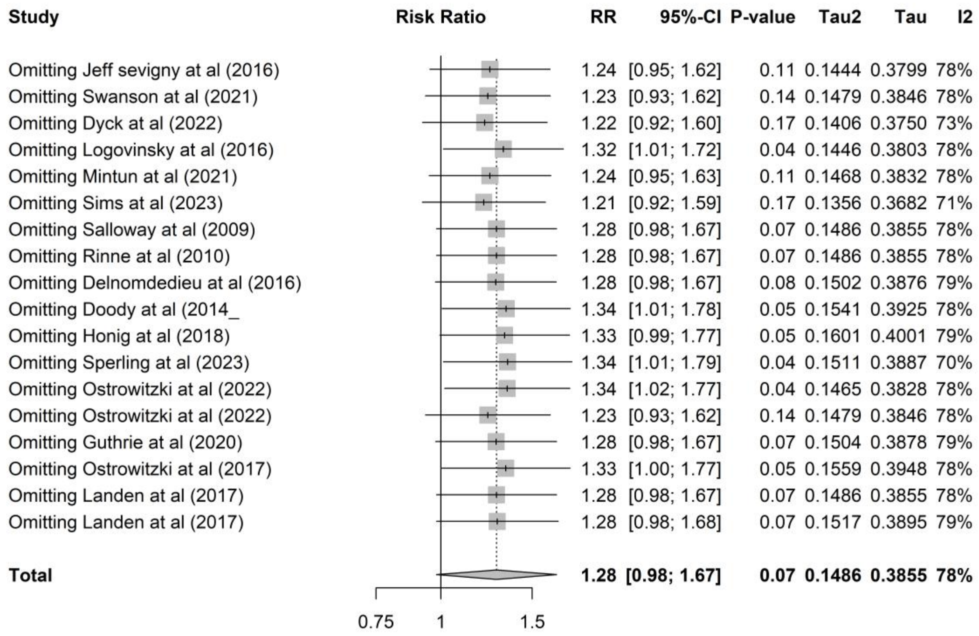

#### 9.20.2 Comparing the results between the fixed-effect and random-effects model

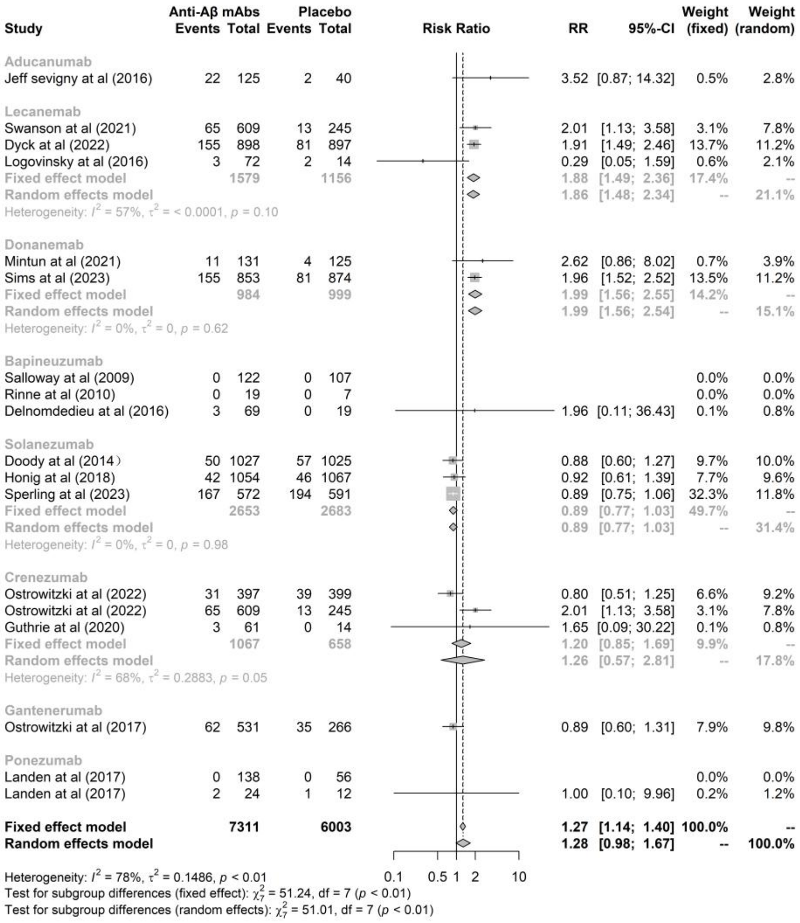

#### Appendix 10: Interrelationships among changes in Aβ deposition, variations in AD biomarkers, alterations in cognitive performance, and the risks of ARIA-E and ARIA-H across all mAbs

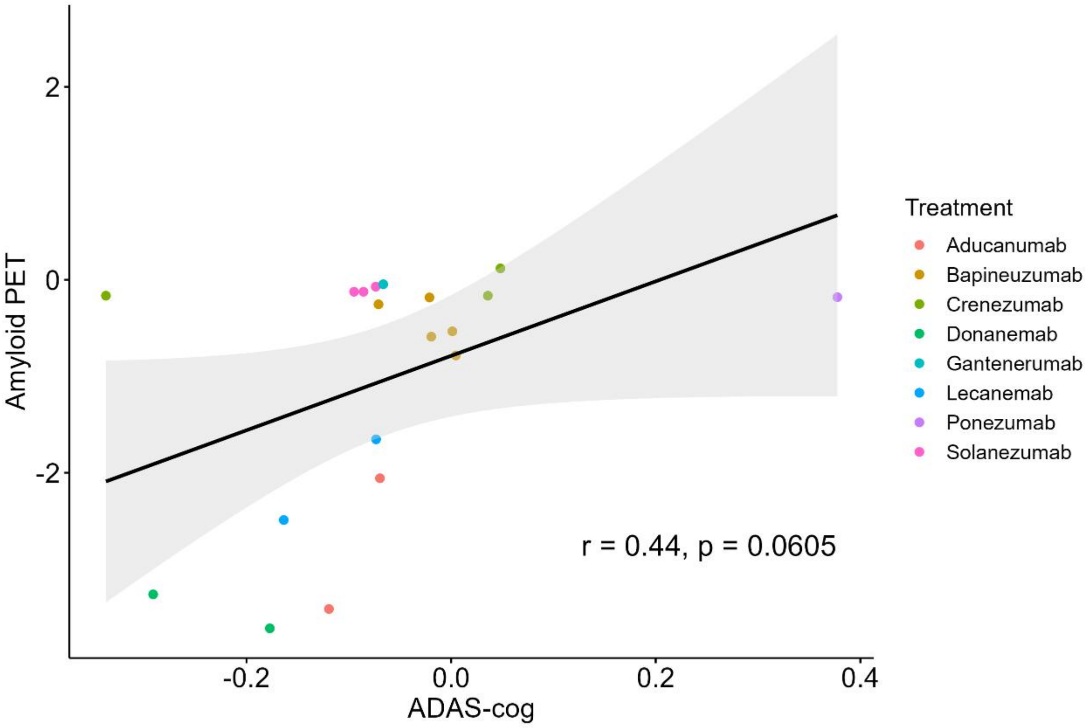

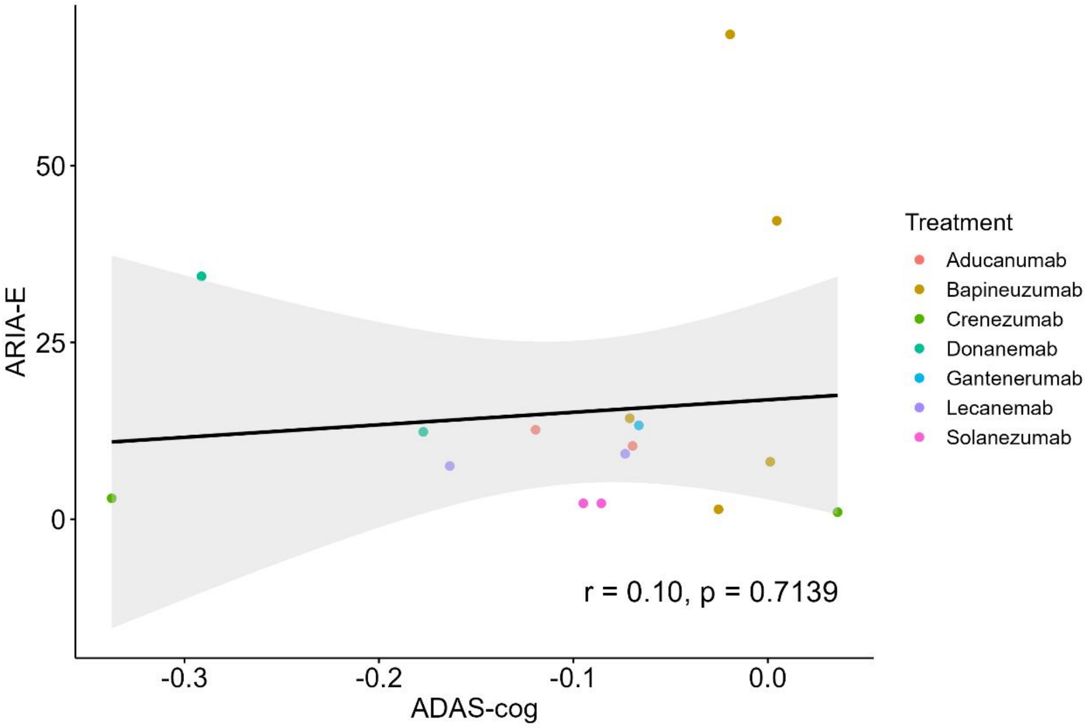

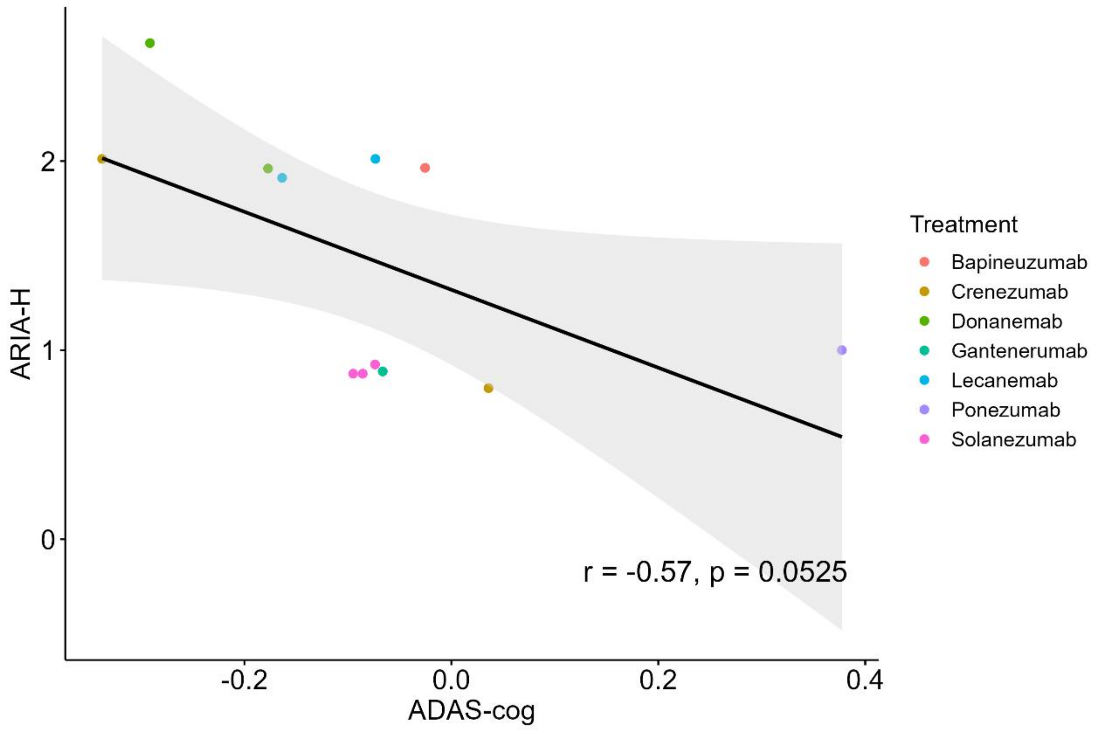
ARIA-H
ADAS-cog
-0.2
0.0
0.2
0.4
0
1
2
r = -0.57, p = 0.0525
Treatment
Bapineuzumab
Crenezumab
Donanemab
Gantenerumab
Lecanemab
Ponezumab
Solanezumab

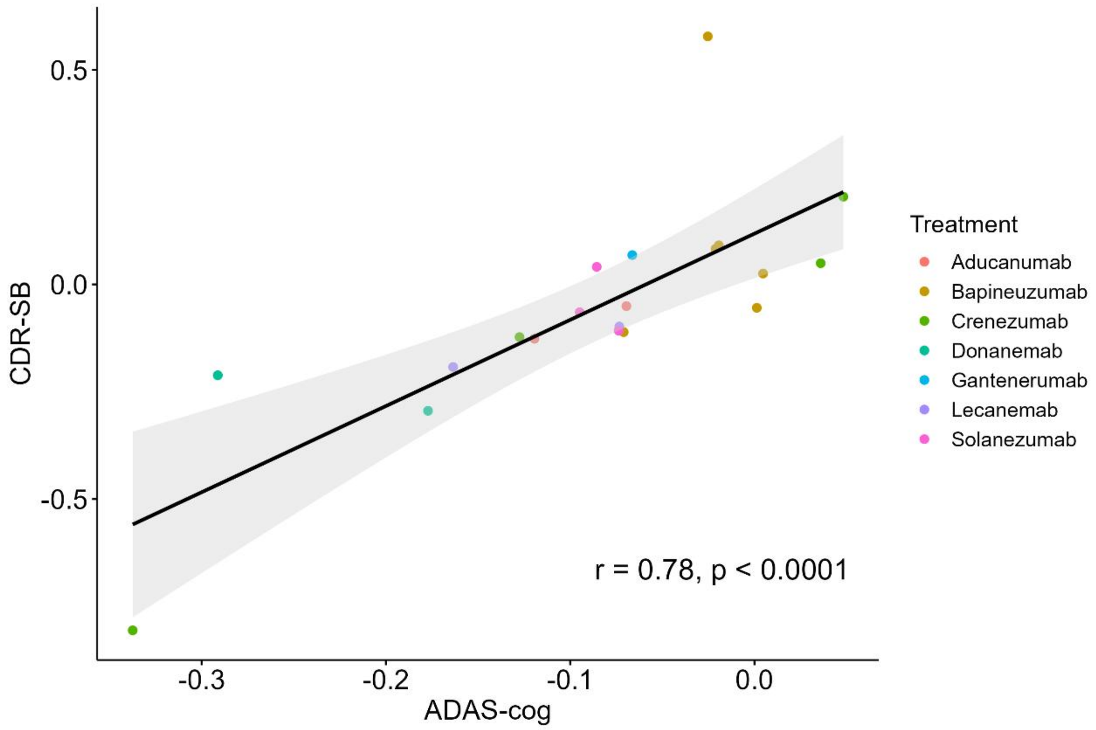
CDR-SB
ADAS-cog
-0.3
-0.2
-0.1
0.0
-0.5
0.0
0.5
r = 0.78, p < 0.0001
Treatment
Aducanumab
Bapineuzumab
Crenezumab
Donanemab
Gantenerumab
Lecanemab
Solanezumab

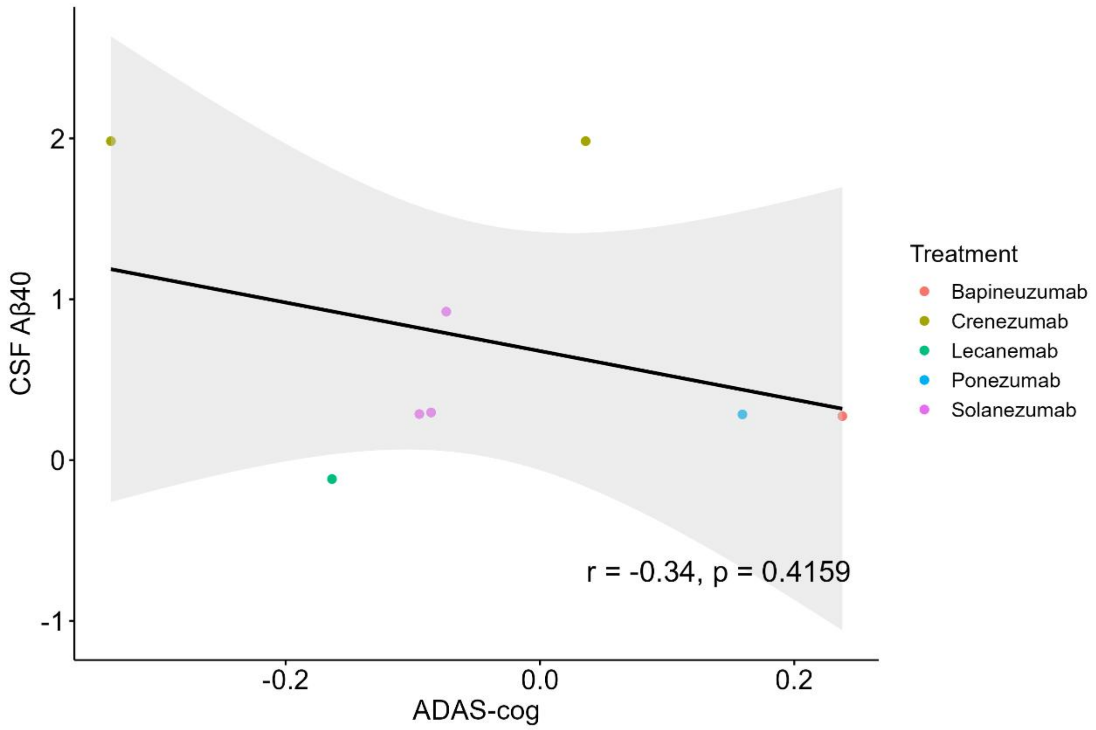

CSF Aβ40
ADAS-cog
-0.2
0.0
0.2
2
1
0
-1
r = -0.34, p = 0.4159
Treatment
Bapineuzumab
Crenezumab
Lecanemab
Ponezumab
Solanezumab

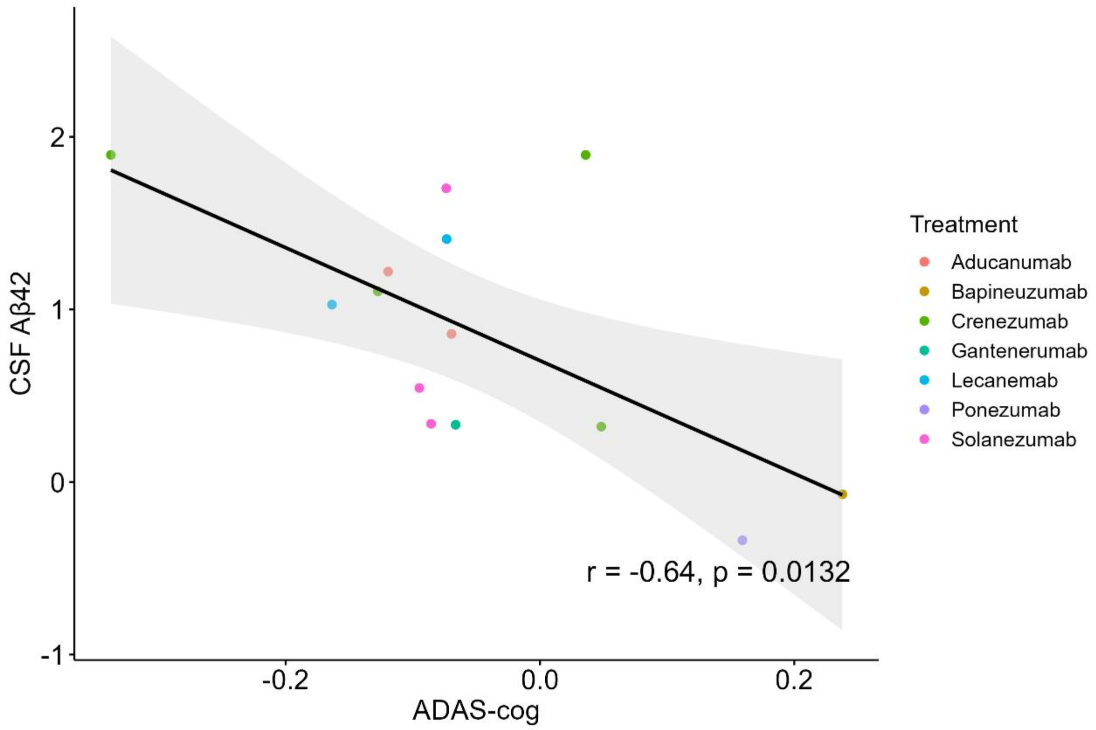

CSF Aβ42
ADAS-cog
-0.2
0.0
0.2
2
1
0
-1
r = -0.64, p = 0.0132
Treatment
Aducanumab
Bapineuzumab
Crenezumab
Gantenerumab
Lecanemab
Ponezumab
Solanezumab

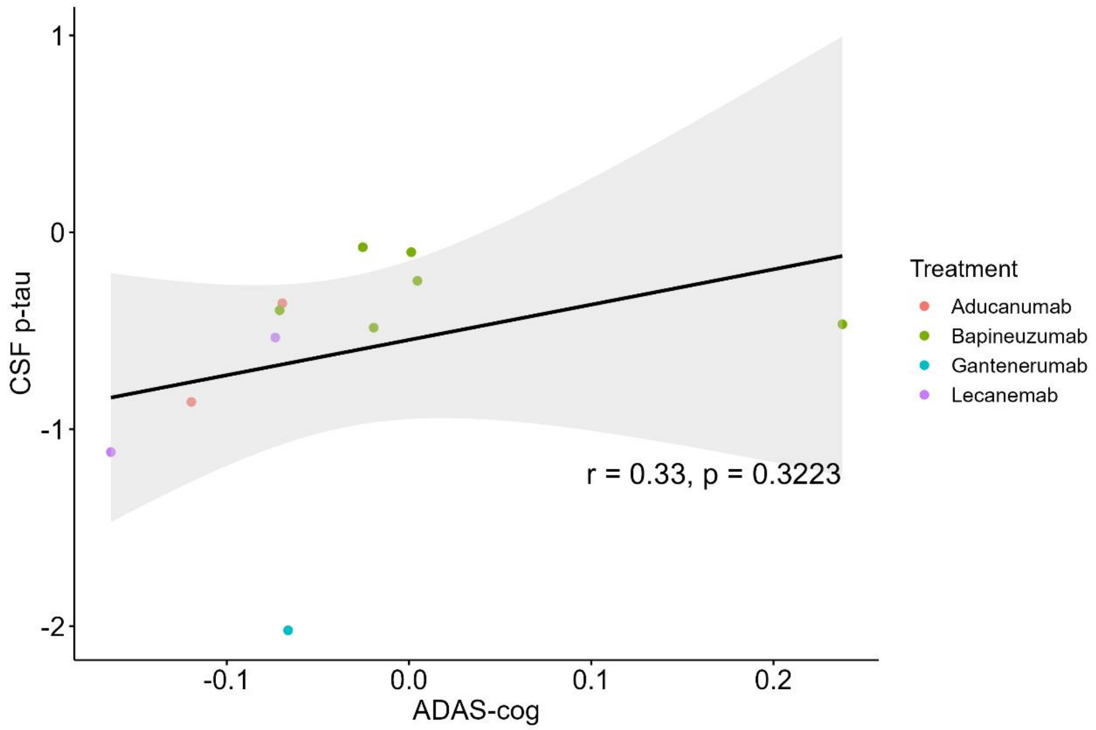
CSF p-tau
1
0
-1
-2
-0.1
0.0
0.1
0.2
ADAS-cog
r = 0.33, p = 0.3223
Treatment
Aducanumab
Bapineuzumab
Gantenerumab
Lecanemab

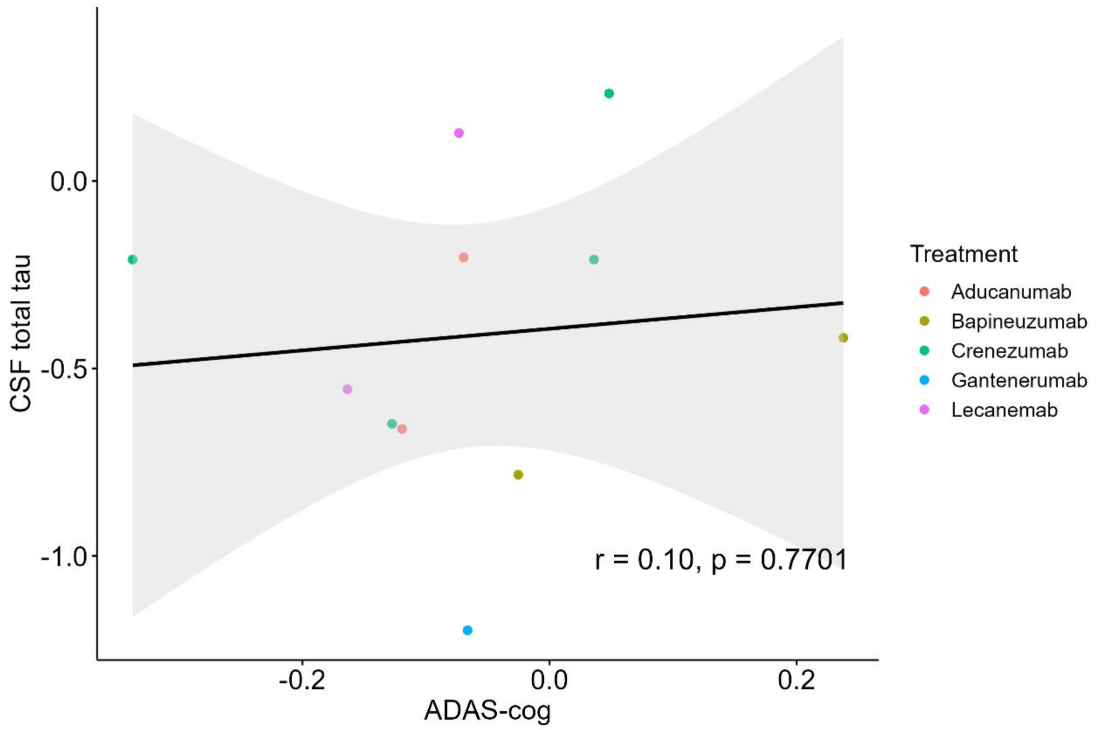
CSF total tau
0.0
-0.5
-1.0
-0.2
0.0
0.2
ADAS-cog
r = 0.10, p = 0.7701
Treatment
Aducanumab
Bapineuzumab
Crenezumab
Gantenerumab
Lecanemab

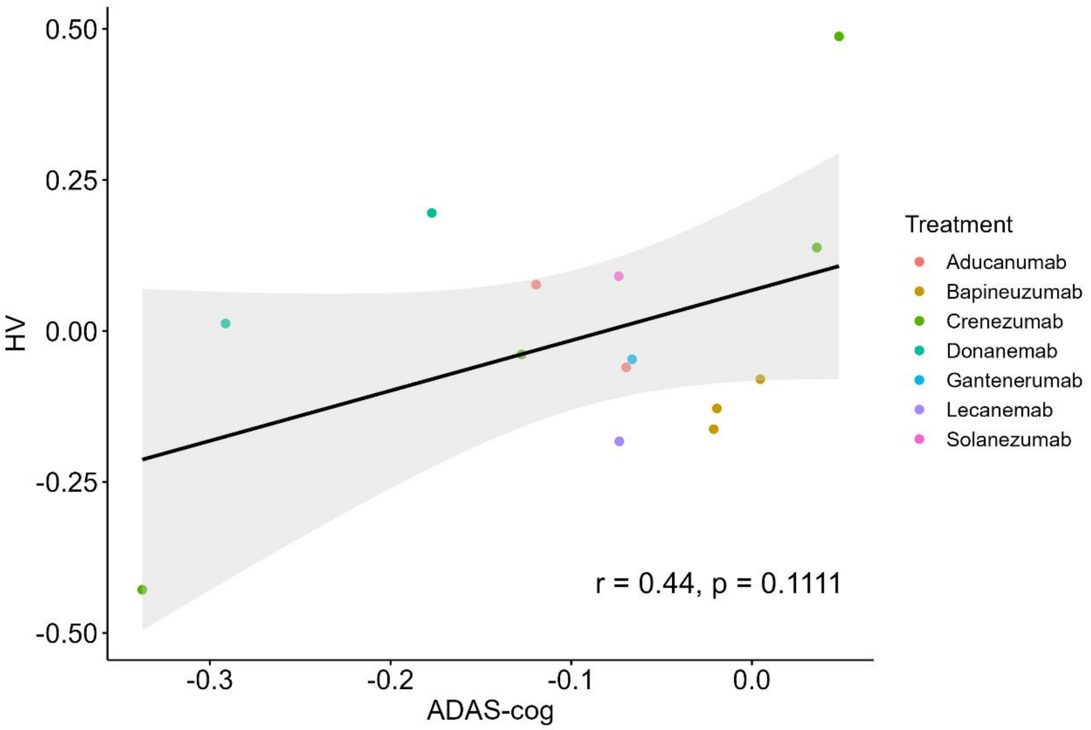

0.50
0.25
0.00
-0.25
-0.50
HV
-0.3
-0.2
-0.1
0.0
ADAS-cog
r = 0.44, p = 0.1111
Treatment
Aducanumab
Bapineuzumab
Crenezumab
Donanemab
Gantenerumab
Lecanemab
Solanezumab

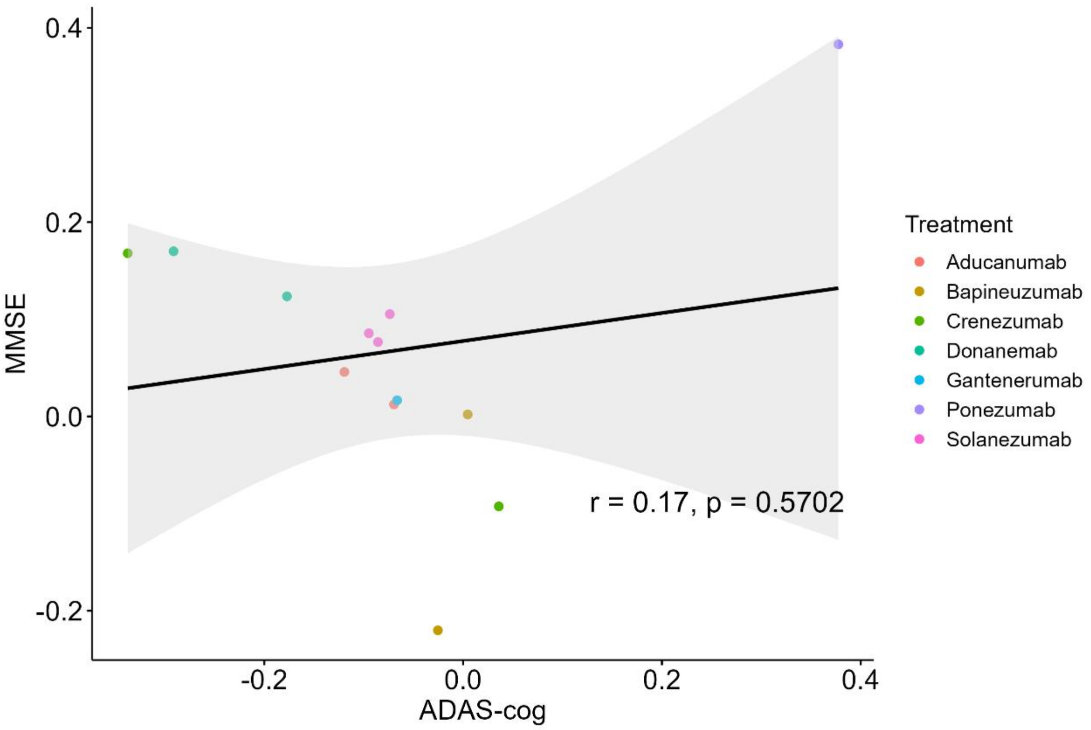

0.4
0.2
0.0
-0.2
MMSE
-0.2
0.0
0.2
0.4
ADAS-cog
r = 0.17, p = 0.5702
Treatment
Aducanumab
Bapineuzumab
Crenezumab
Donanemab
Gantenerumab
Ponezumab
Solanezumab

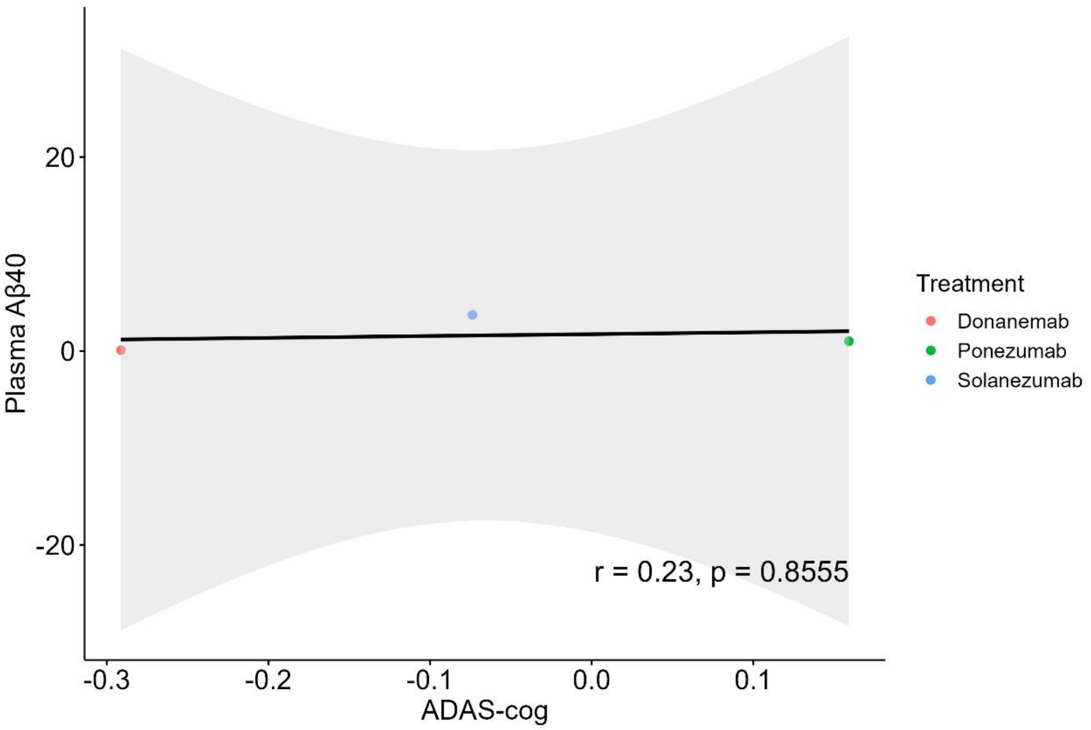

Plasma Aβ40
20
0
-20
-0.3
-0.2
-0.1
0.0
0.1
ADAS-cog
r = 0.23, p = 0.8555
Treatment
Donanemab
Ponezumab
Solanezumab

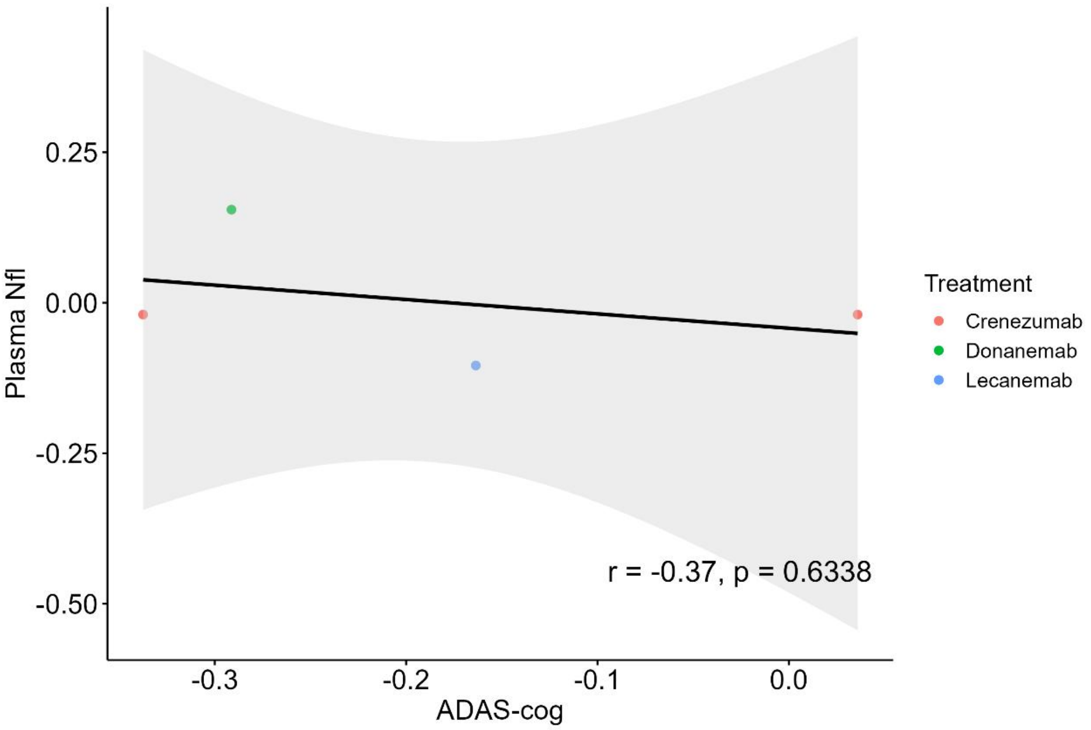

Plasma Nfl
0.25
0.00
-0.25
-0.50
-0.3
-0.2
-0.1
0.0
ADAS-cog
r = -0.37, p = 0.6338
Treatment
Crenezumab
Donanemab
Lecanemab

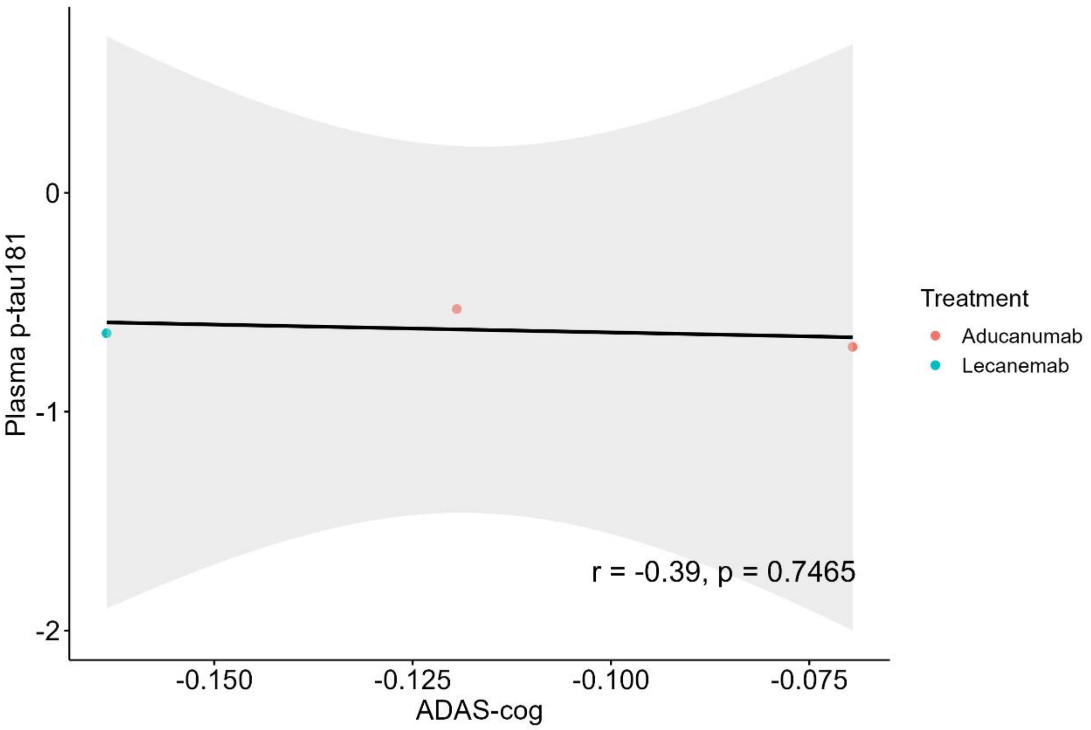
Plasma p-tau181
0
-1
-2
-0.150
-0.125
-0.100
-0.075
ADAS-cog
r = -0.39, p = 0.7465
Treatment
Aducanumab
Lecanemab

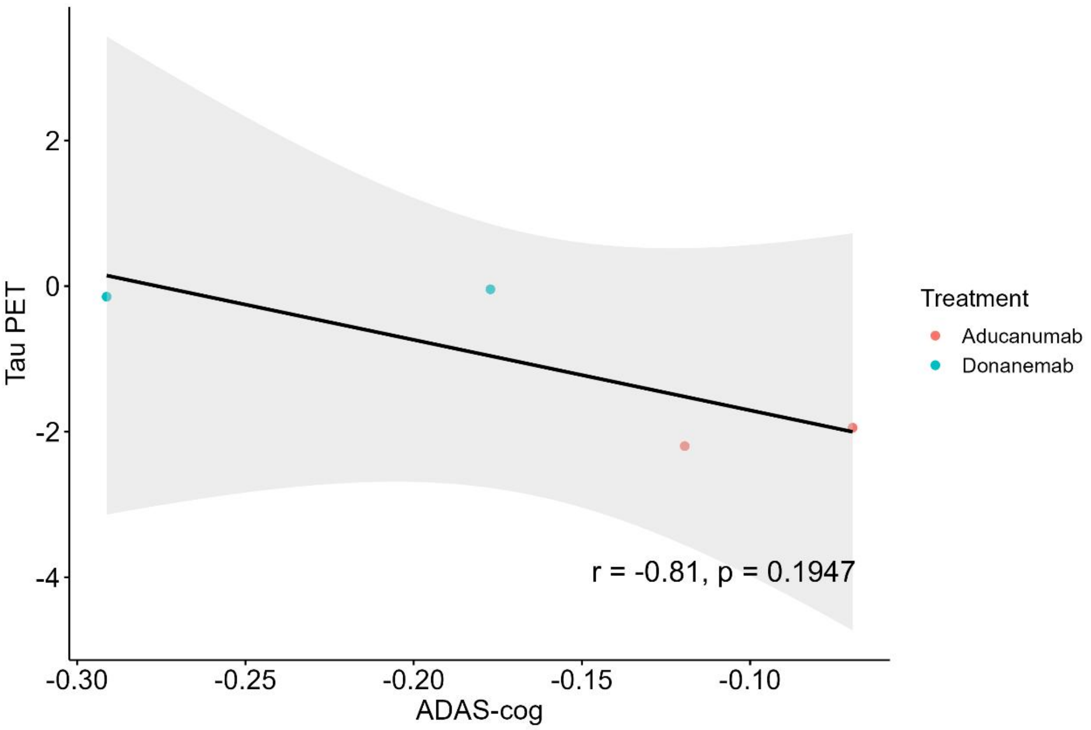
Tau PET
2
0
-2
-4
-0.30
-0.25
-0.20
-0.15
-0.10
ADAS-cog
r = -0.81, p = 0.1947
Treatment
Aducanumab
Donanemab

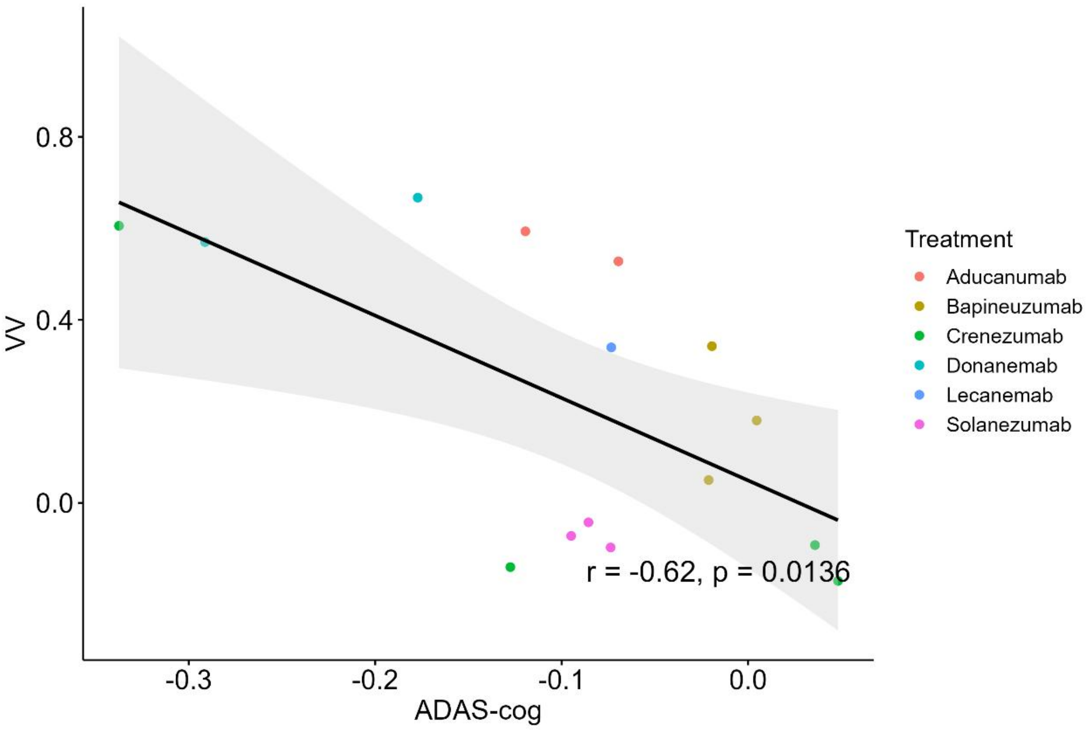
0.8
0.4
0.0
VV
-0.3
-0.2
-0.1
0.0
ADAS-cog
r = -0.62, p = 0.0136
Treatment
Aducanumab
Bapineuzumab
Crenezumab
Donanemab
Lecanemab
Solanezumab

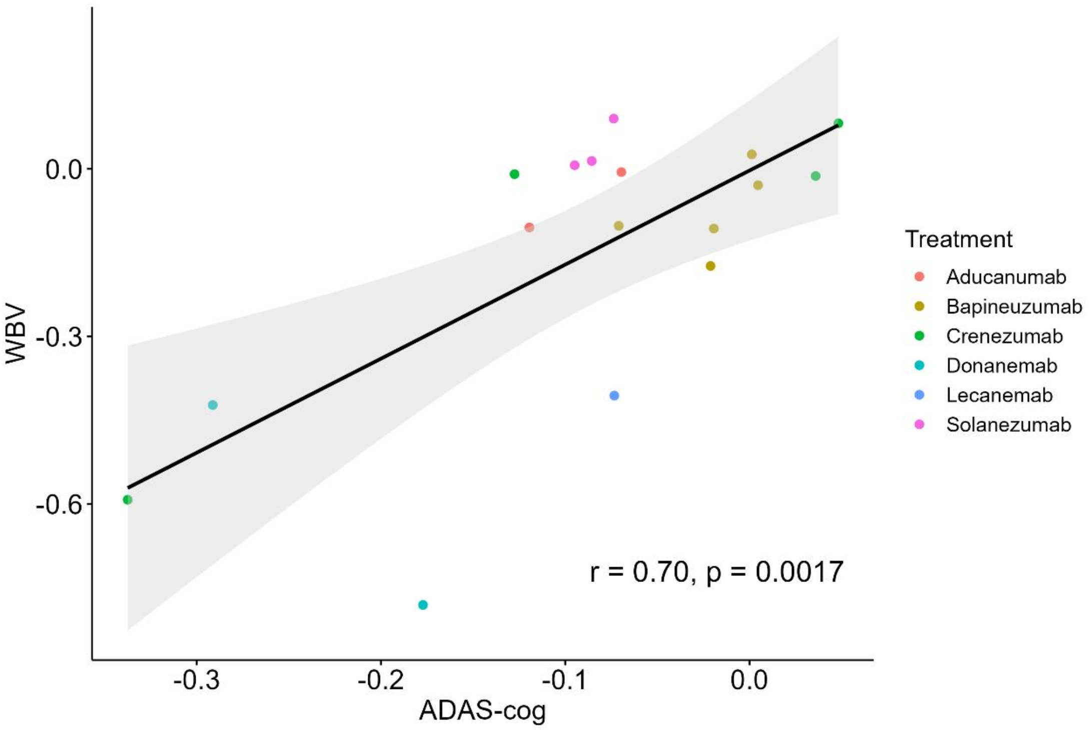
0.0
-0.3
-0.6
WBV
-0.3
-0.2
-0.1
0.0
ADAS-cog
r = 0.70, p = 0.0017
Treatment
Aducanumab
Bapineuzumab
Crenezumab
Donanemab
Lecanemab
Solanezumab

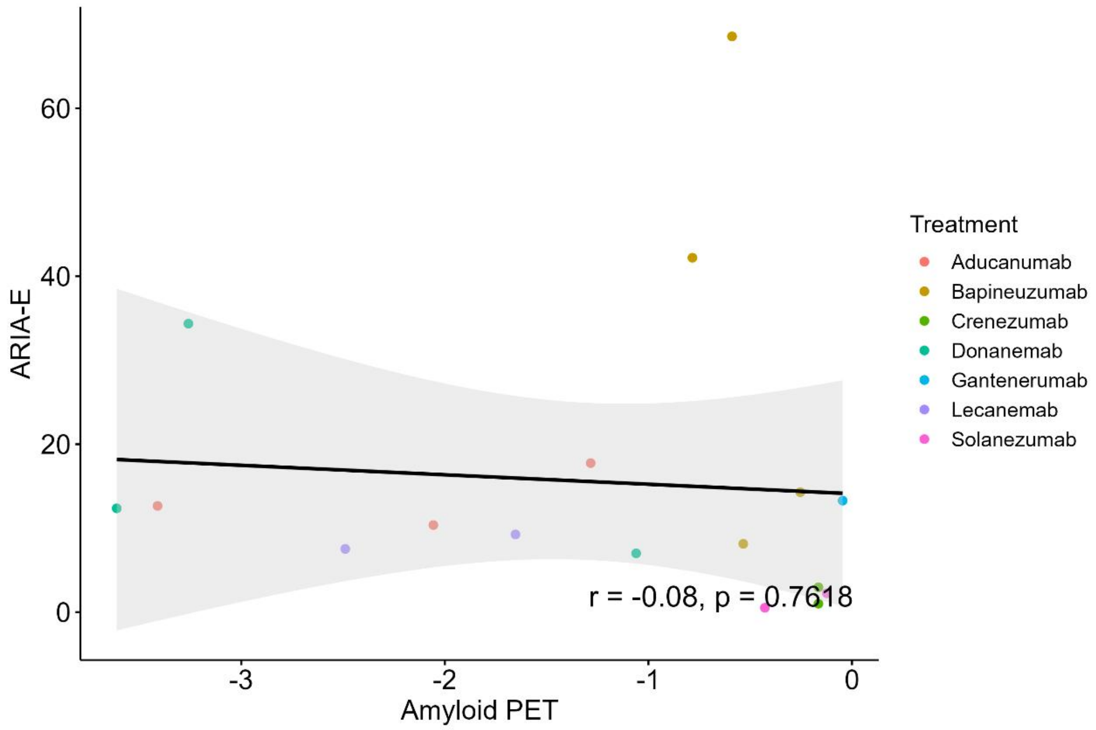
ARIA-E
Amyloid PET
r = -0.08, p = 0.7618
Treatment
Aducanumab
Bapineuzumab
Crenezumab
Donanemab
Gantenerumab
Lecanemab
Solanezumab

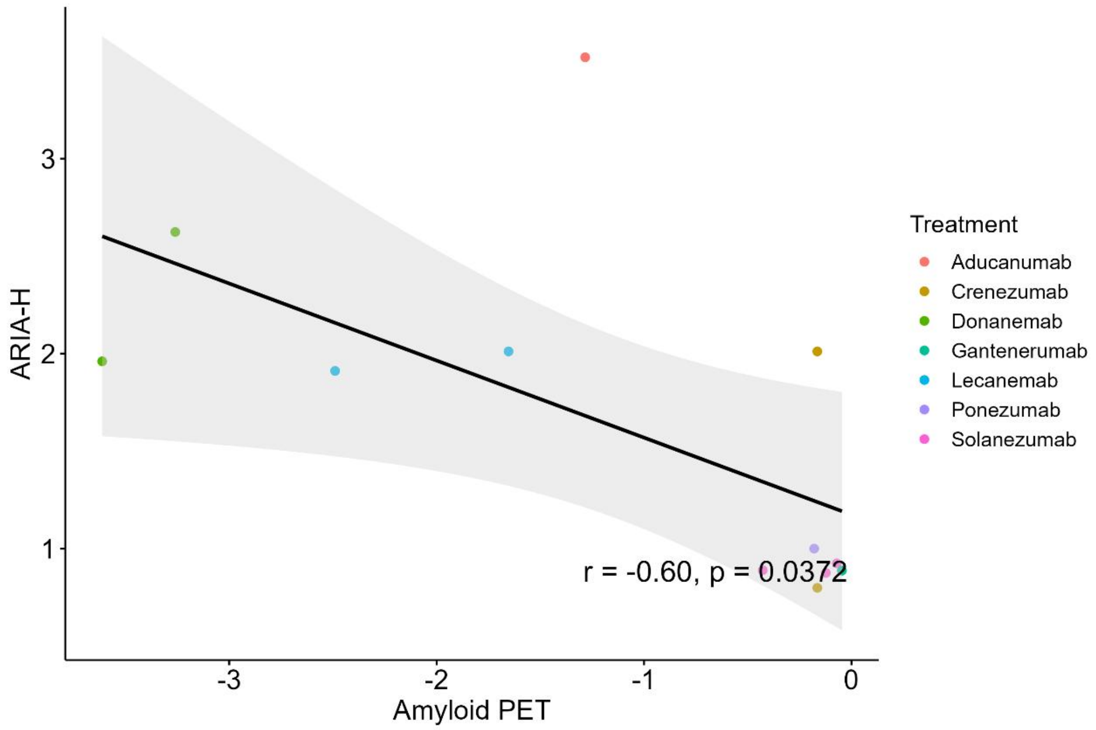
ARIA-H
Amyloid PET
r = -0.60, p = 0.0372
Treatment
Aducanumab
Crenezumab
Donanemab
Gantenerumab
Lecanemab
Ponezumab
Solanezumab

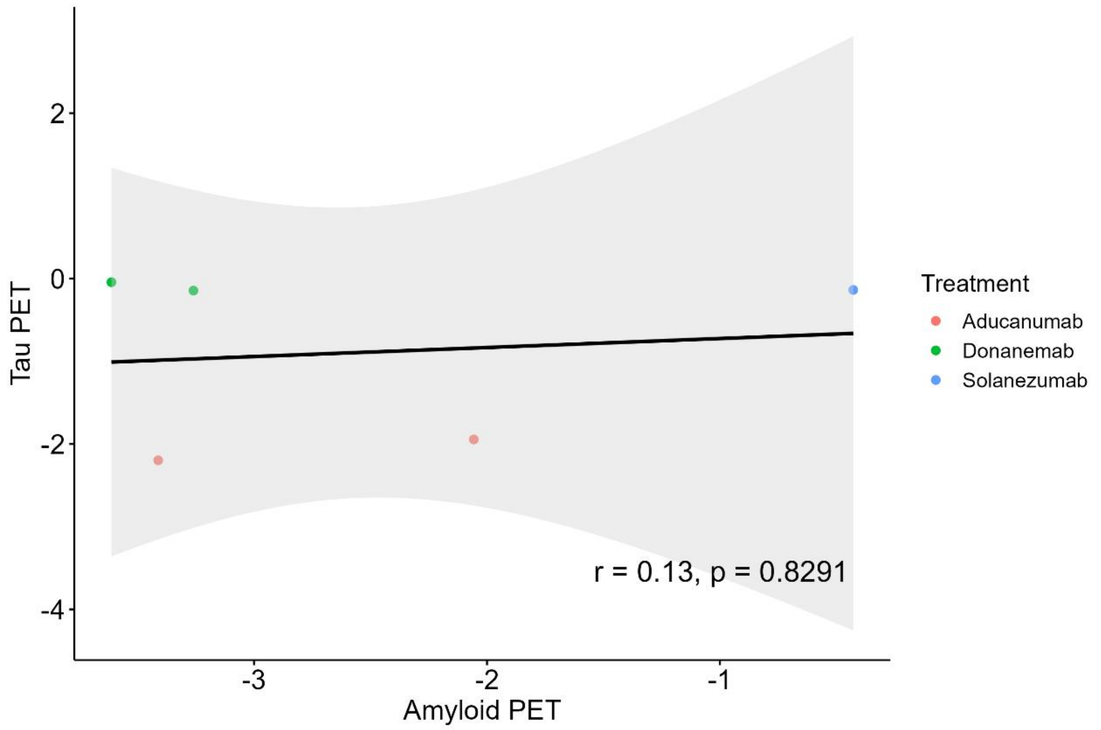
Tau PET
2
0
-2
-4
-3
-2
-1
Amyloid PET
r = 0.13, p = 0.8291
Treatment
Aducanumab
Donanemab
Solanezumab

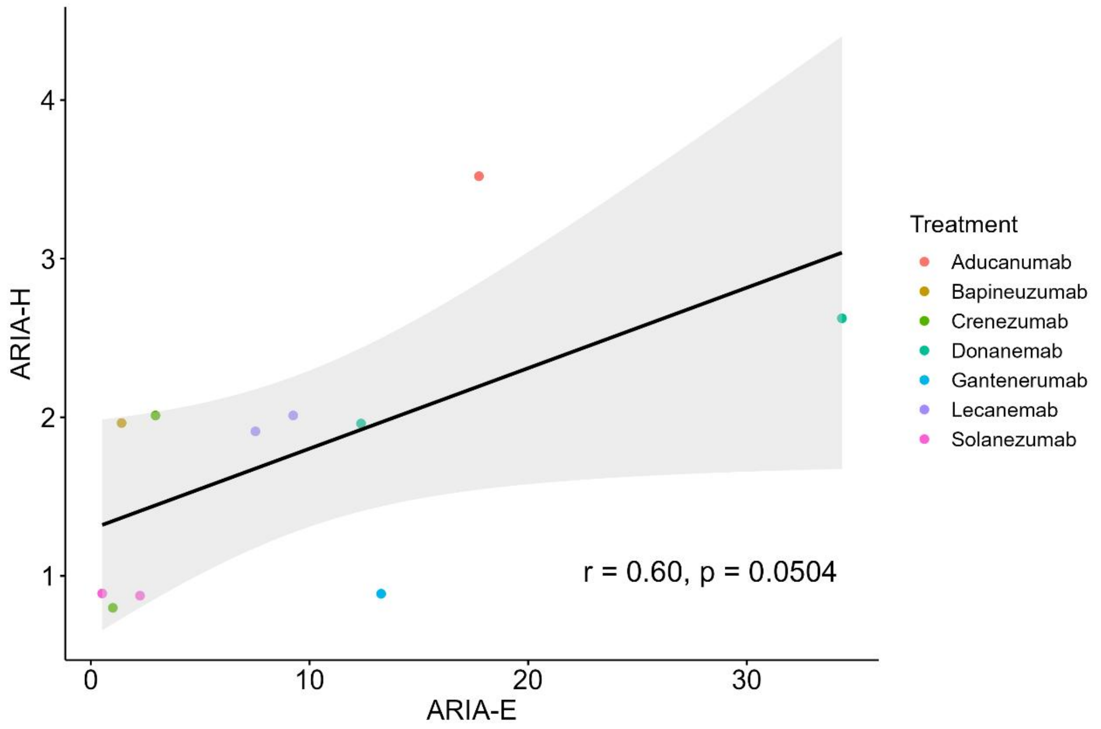
ARIA-H
4
3
2
1
0
10
20
30
ARIA-E
r = 0.60, p = 0.0504
Treatment
Aducanumab
Bapineuzumab
Crenezumab
Donanemab
Gantenerumab
Lecanemab
Solanezumab

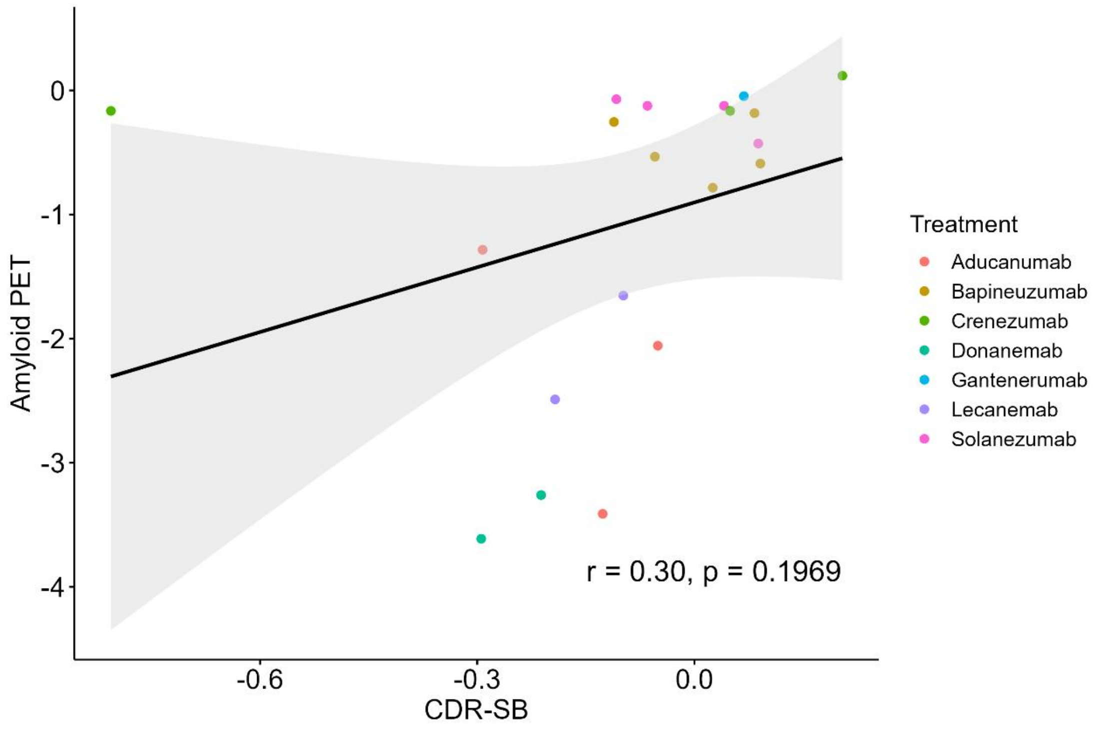
Amyloid PET
0
-1
-2
-3
-4
-0.6
-0.3
0.0
CDR-SB
r = 0.30, p = 0.1969
Treatment
Aducanumab
Bapineuzumab
Crenezumab
Donanemab
Gantenerumab
Lecanemab
Solanezumab

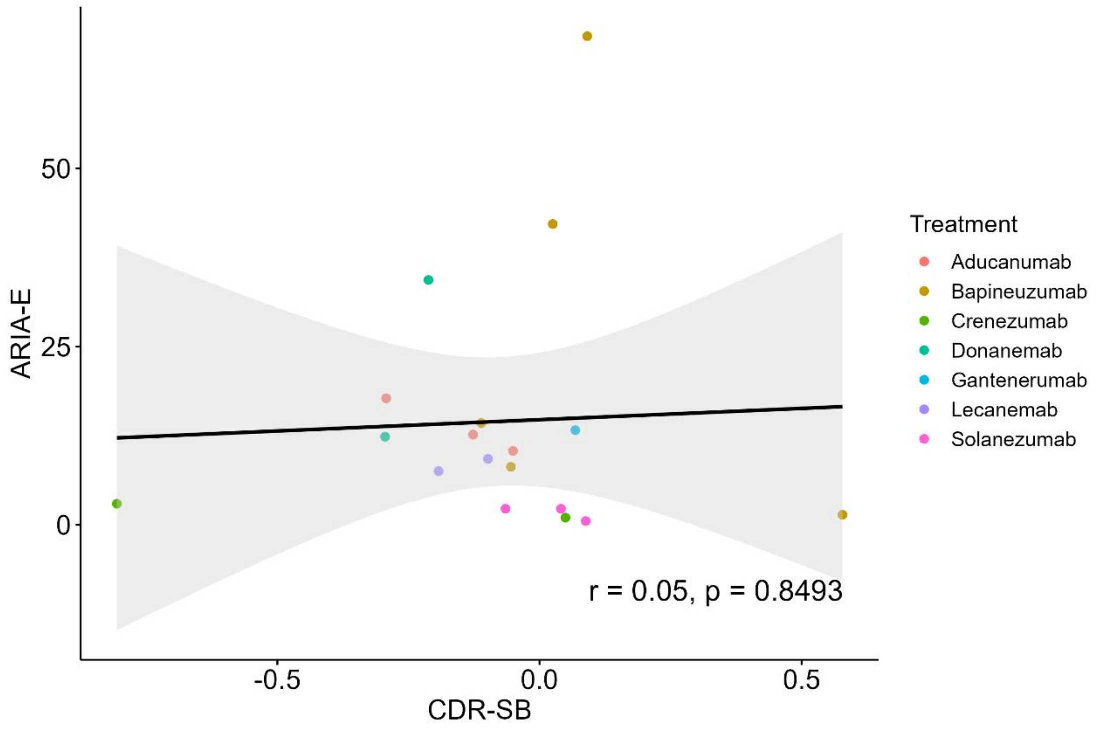
ARIA-E
50
25
0
-0.5
0.0
0.5
CDR-SB
r = 0.05, p = 0.8493
Treatment
Aducanumab
Bapineuzumab
Crenezumab
Donanemab
Gantenerumab
Lecanemab
Solanezumab

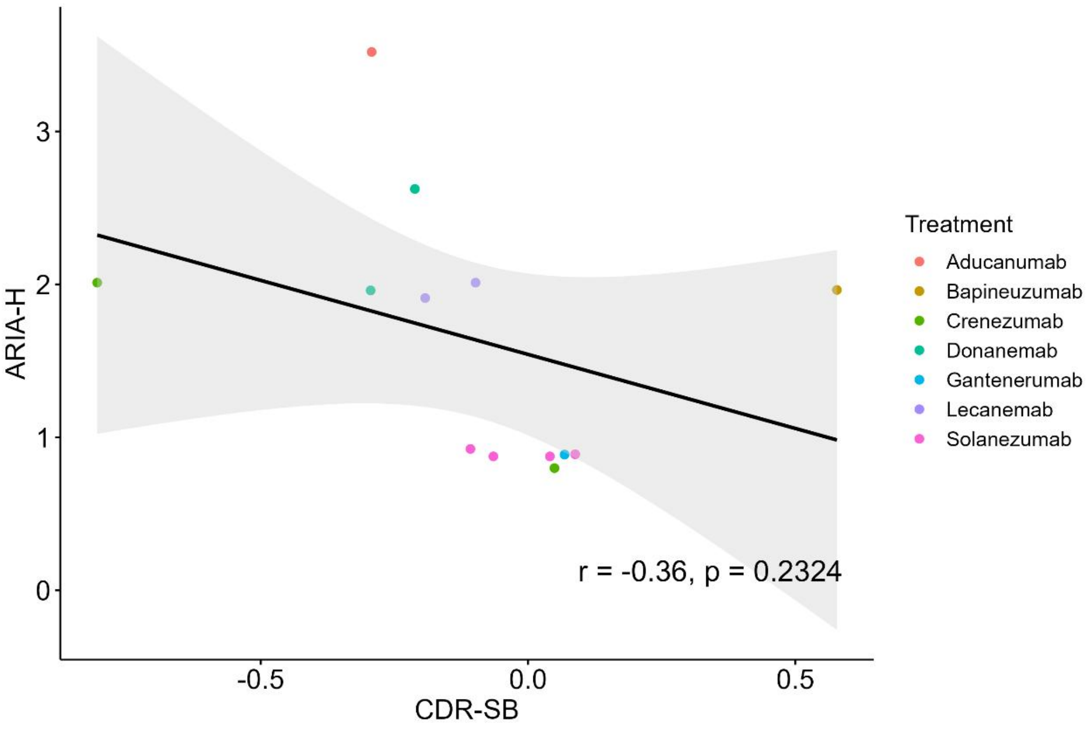

ARIA-H
CDR-SB
r = -0.36, p = 0.2324
Treatment
Aducanumab
Bapineuzumab
Crenezumab
Donanemab
Gantenerumab
Lecanemab
Solanezumab

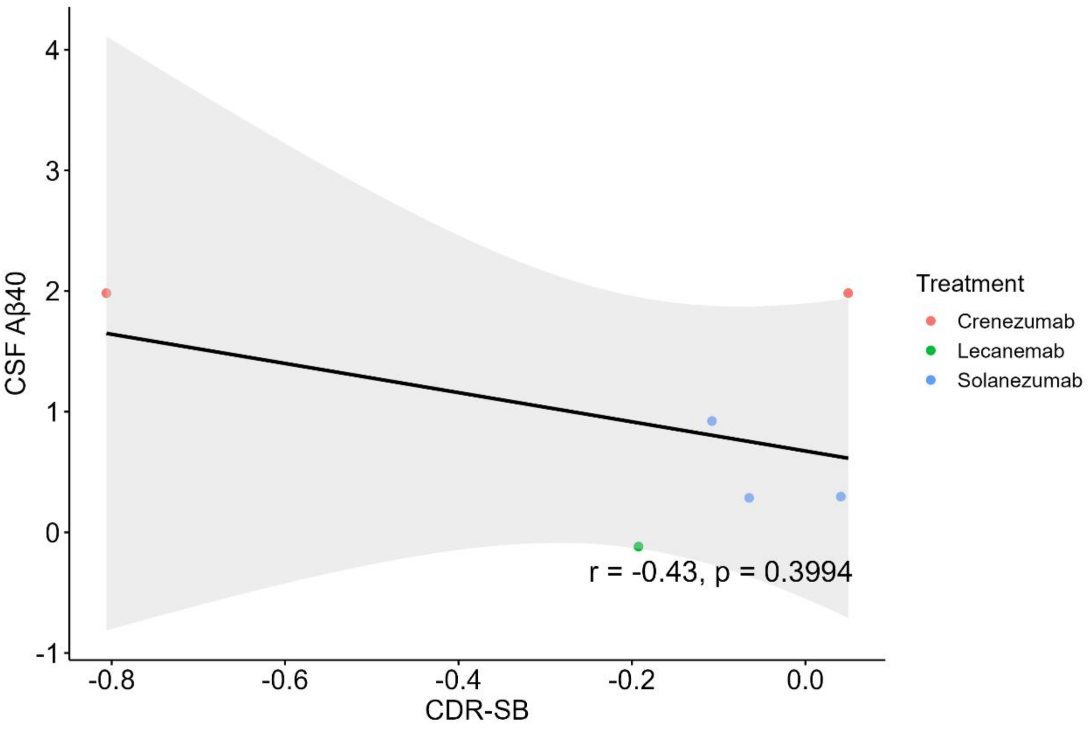

CSF Aβ40
CDR-SB
r = -0.43, p = 0.3994
Treatment
Crenezumab
Lecanemab
Solanezumab

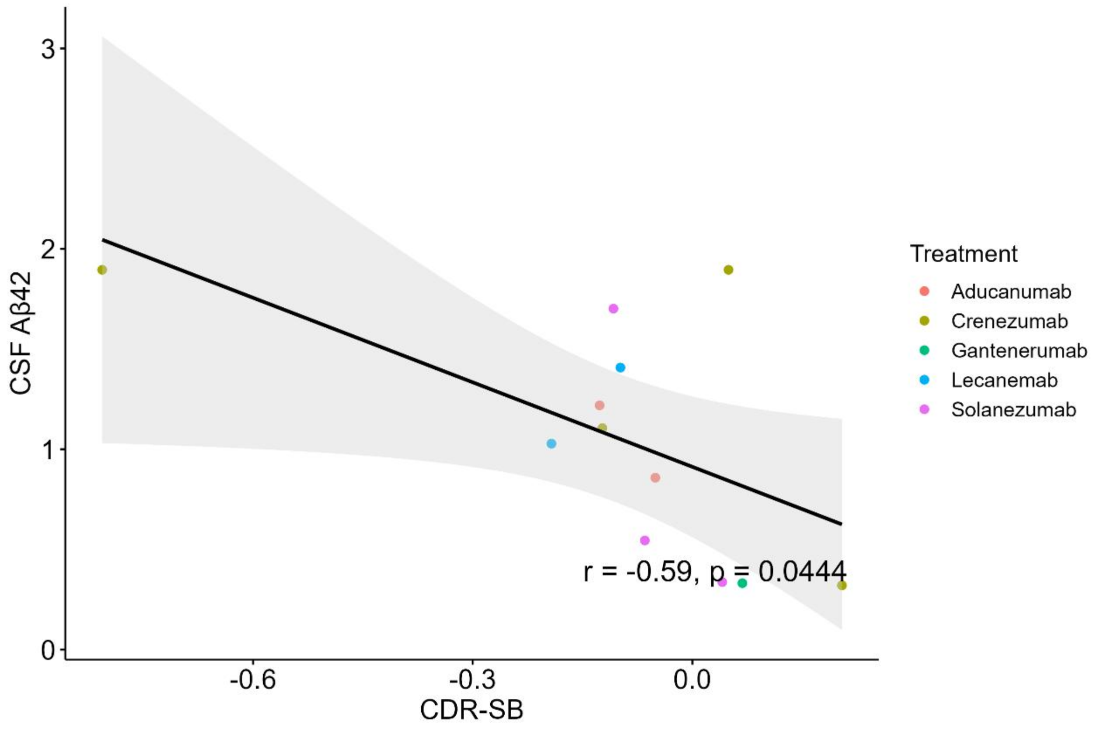
CSF Aβ42
CDR-SB
r = -0.59, p = 0.0444
Treatment
Aducanumab
Crenezumab
Gantenerumab
Lecanemab
Solanezumab

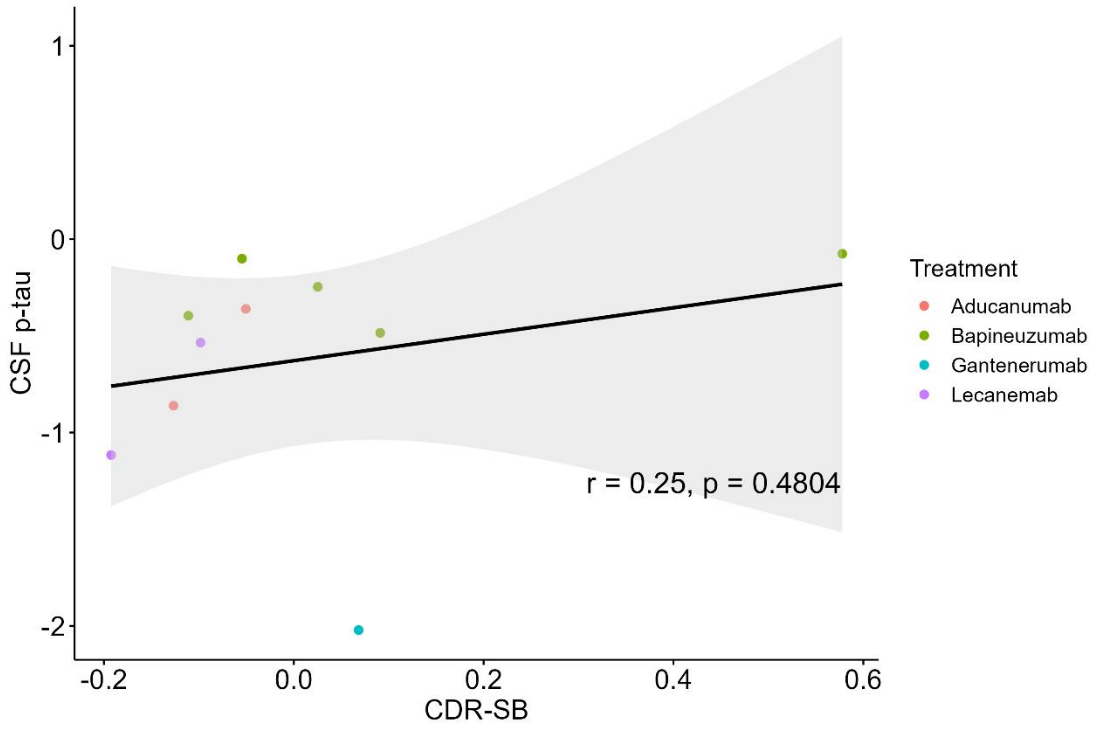
CSF p-tau
CDR-SB
r = 0.25, p = 0.4804
Treatment
Aducanumab
Bapineuzumab
Gantenerumab
Lecanemab

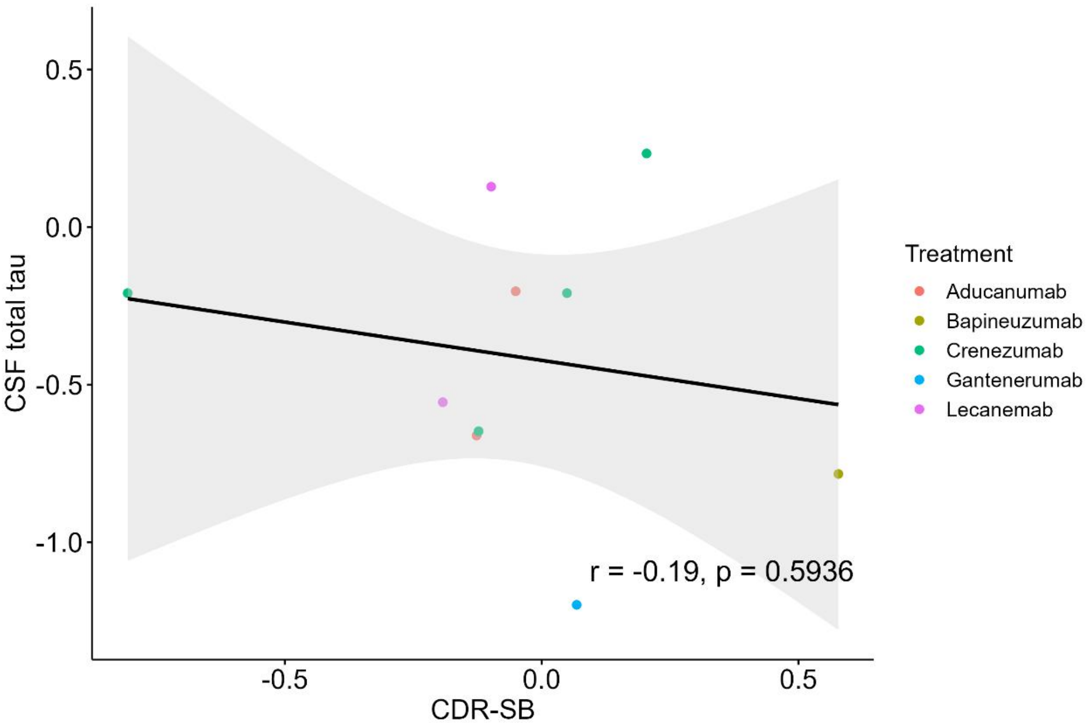
CSF total tau
CDR-SB
r = -0.19, p = 0.5936
Treatment
Aducanumab
Bapineuzumab
Crenezumab
Gantenerumab
Lecanemab
0.5
0.0
-0.5
-1.0
-0.5
0.0
0.5

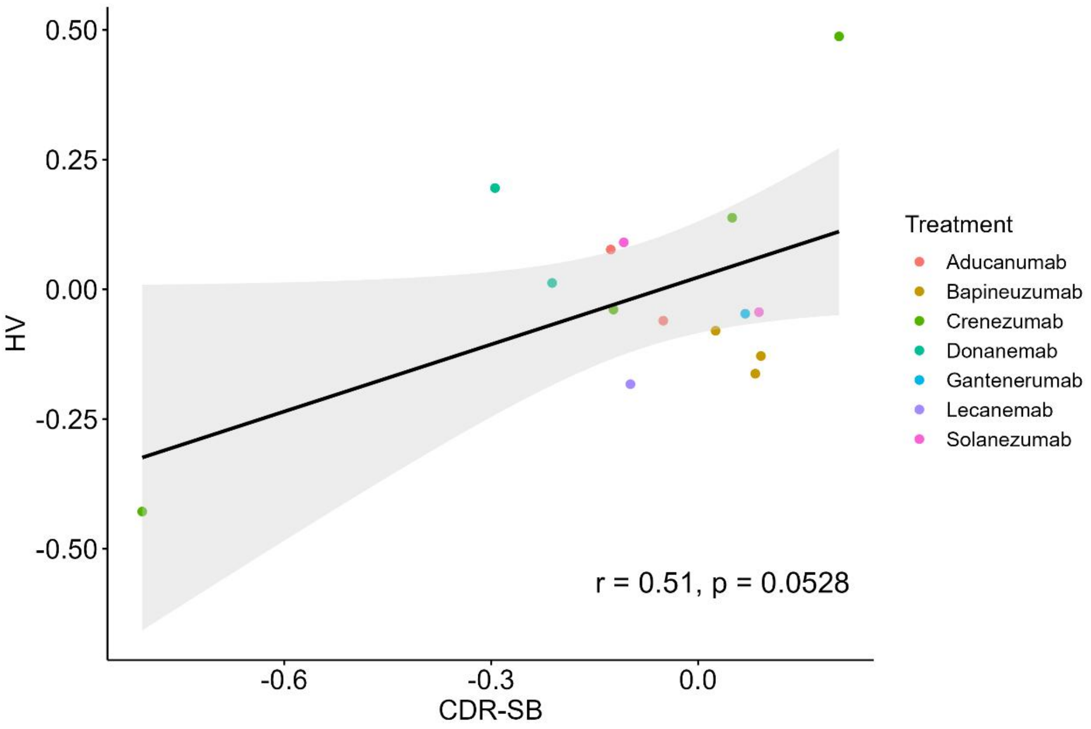
HV
CDR-SB
r = 0.51, p = 0.0528
Treatment
Aducanumab
Bapineuzumab
Crenezumab
Donanemab
Gantenerumab
Lecanemab
Solanezumab
0.50
0.25
0.00
-0.25
-0.50
-0.6
-0.3
0.0

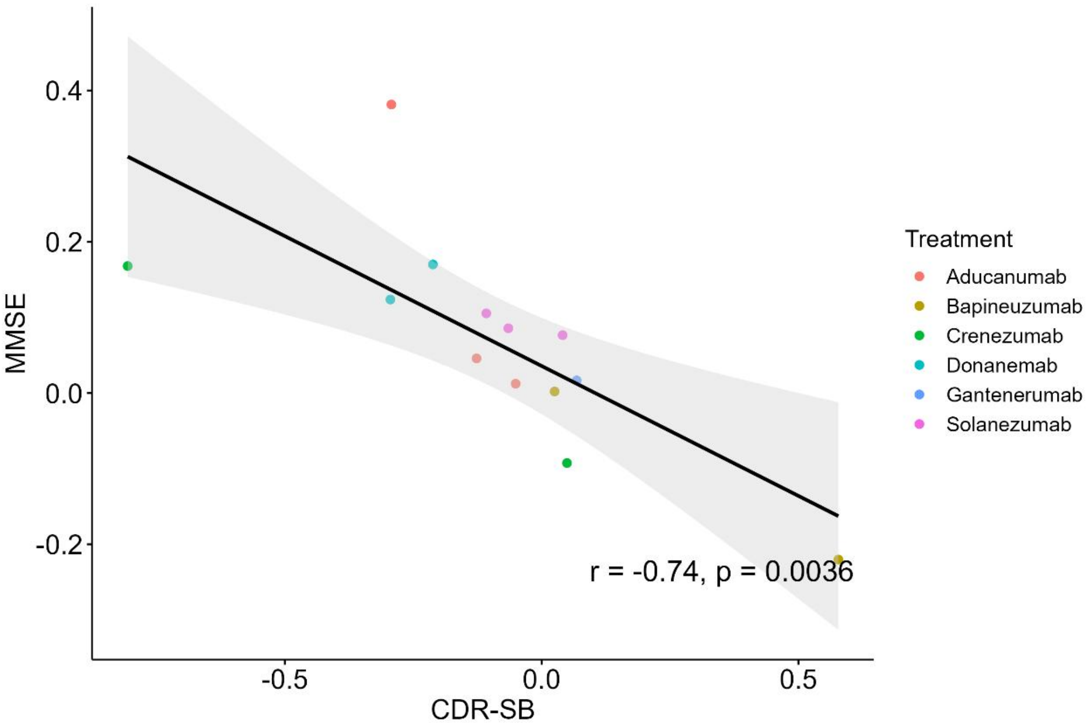

0.4
0.2
0.0
-0.2
MMSE
-0.5
0.0
0.5
CDR-SB
r = -0.74, p = 0.0036
Treatment
Aducanumab
Bapineuzumab
Crenezumab
Donanemab
Gantenerumab
Solanezumab

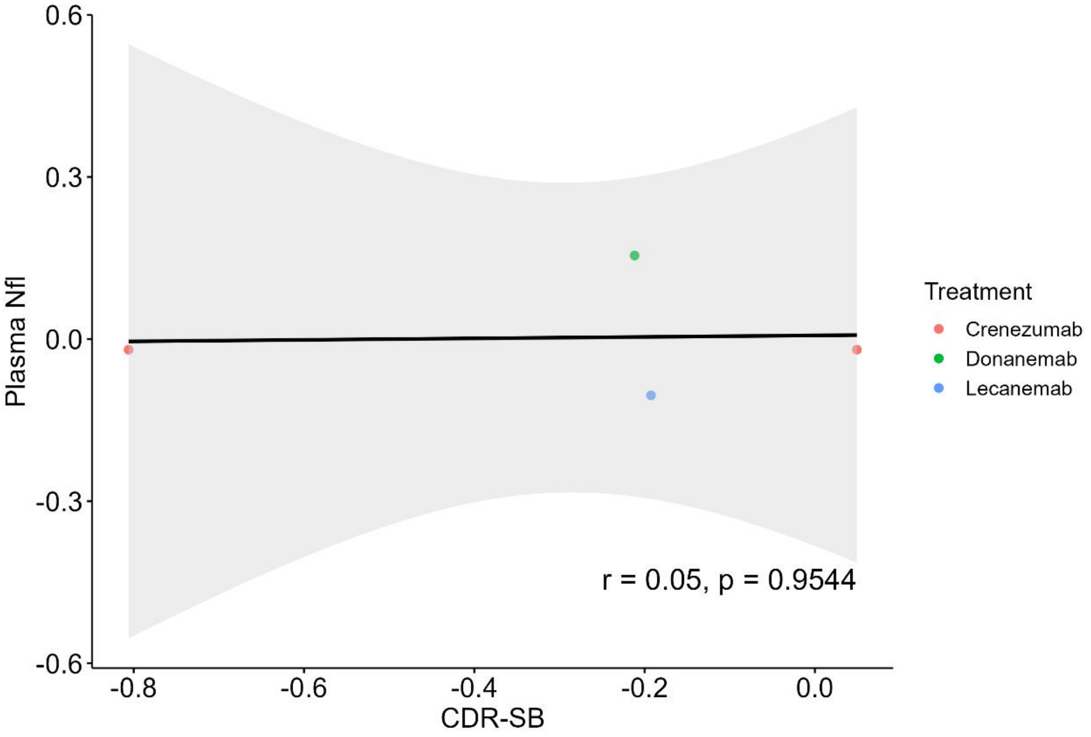

0.6
0.3
0.0
-0.3
-0.6
Plasma Nfl
-0.8
-0.6
-0.4
-0.2
0.0
CDR-SB
r = 0.05, p = 0.9544
Treatment
Crenezumab
Donanemab
Lecanemab

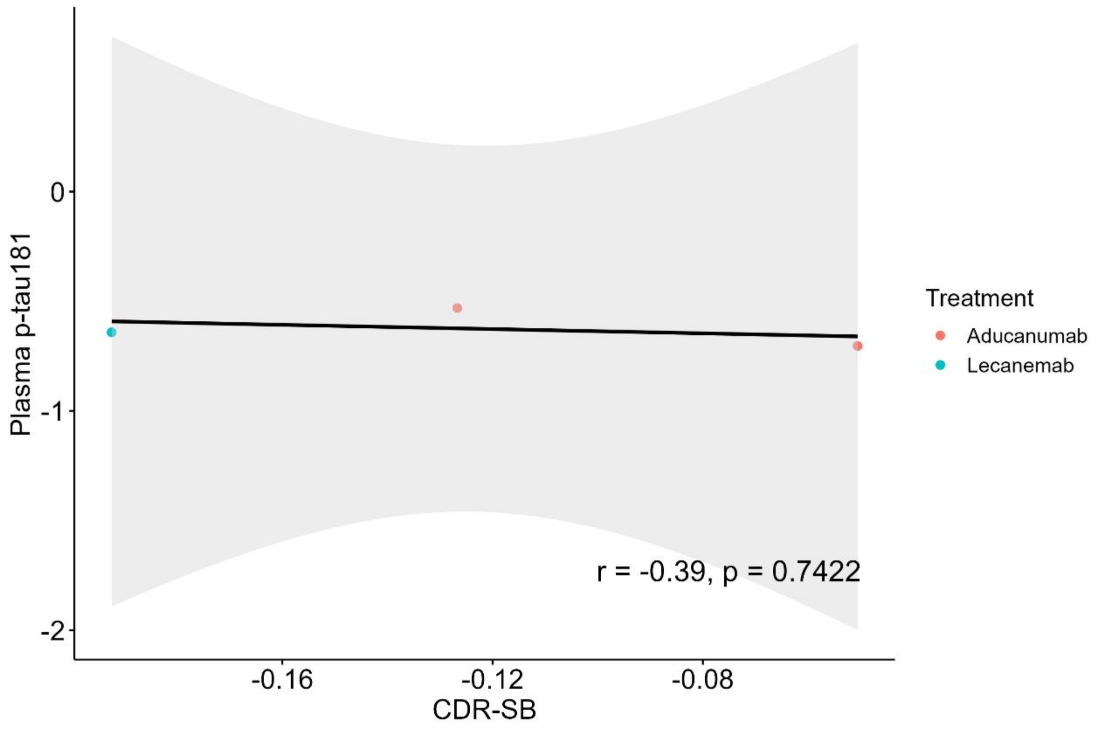
Plasma p-tau181
0
-1
-2
-0.16
-0.12
-0.08
CDR-SB
r = -0.39, p = 0.7422
Treatment
Aducanumab
Lecanemab

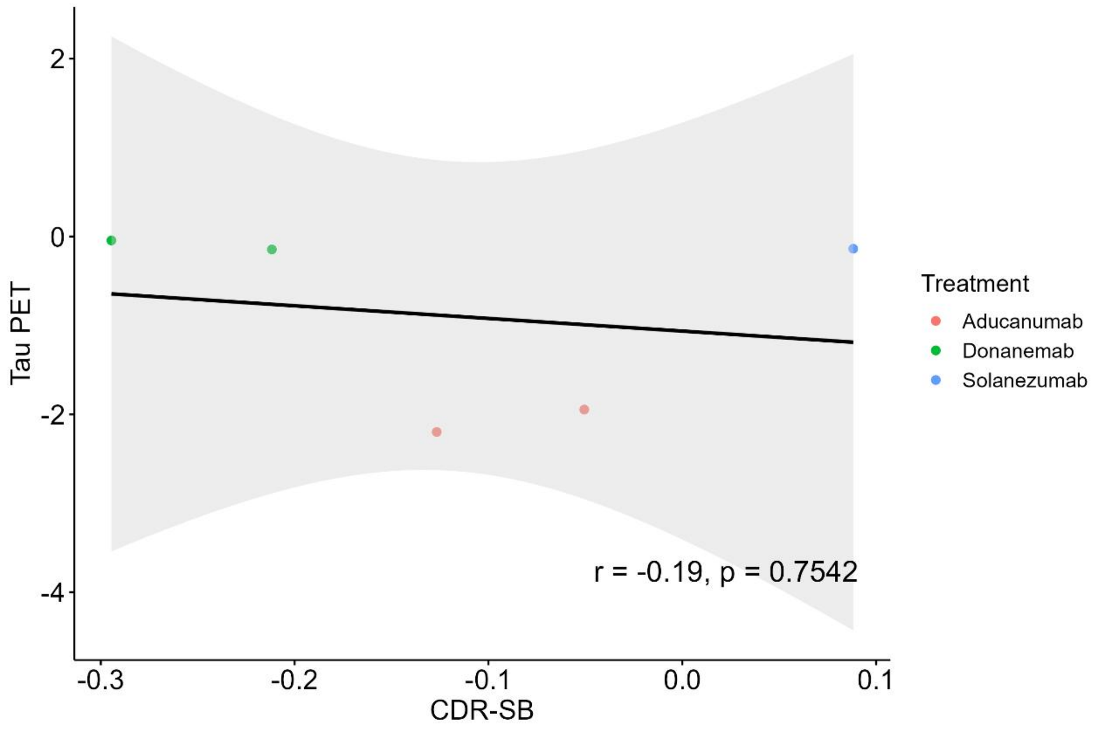
Tau PET
2
0
-2
-4
-0.3
-0.2
-0.1
0.0
0.1
CDR-SB
r = -0.19, p = 0.7542
Treatment
Aducanumab
Donanemab
Solanezumab

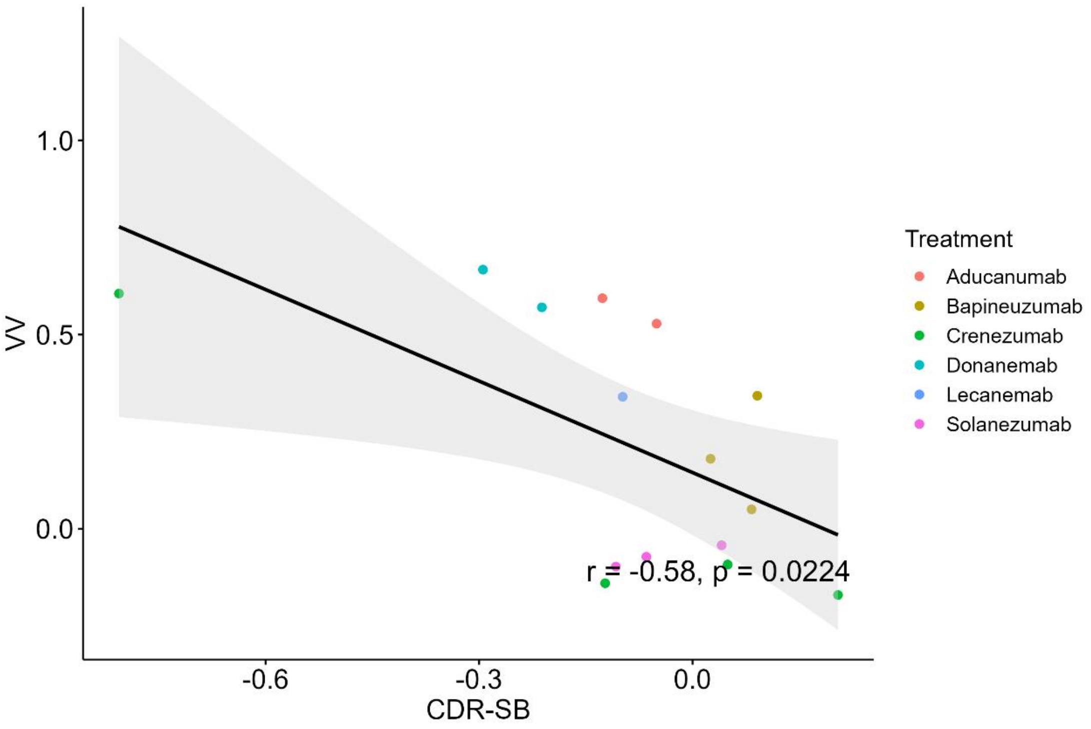
VV
1.0
0.5
0.0
-0.6
-0.3
0.0
CDR-SB
r = -0.58, p = 0.0224
Treatment
Aducanumab
Bapineuzumab
Crenezumab
Donanemab
Lecanemab
Solanezumab

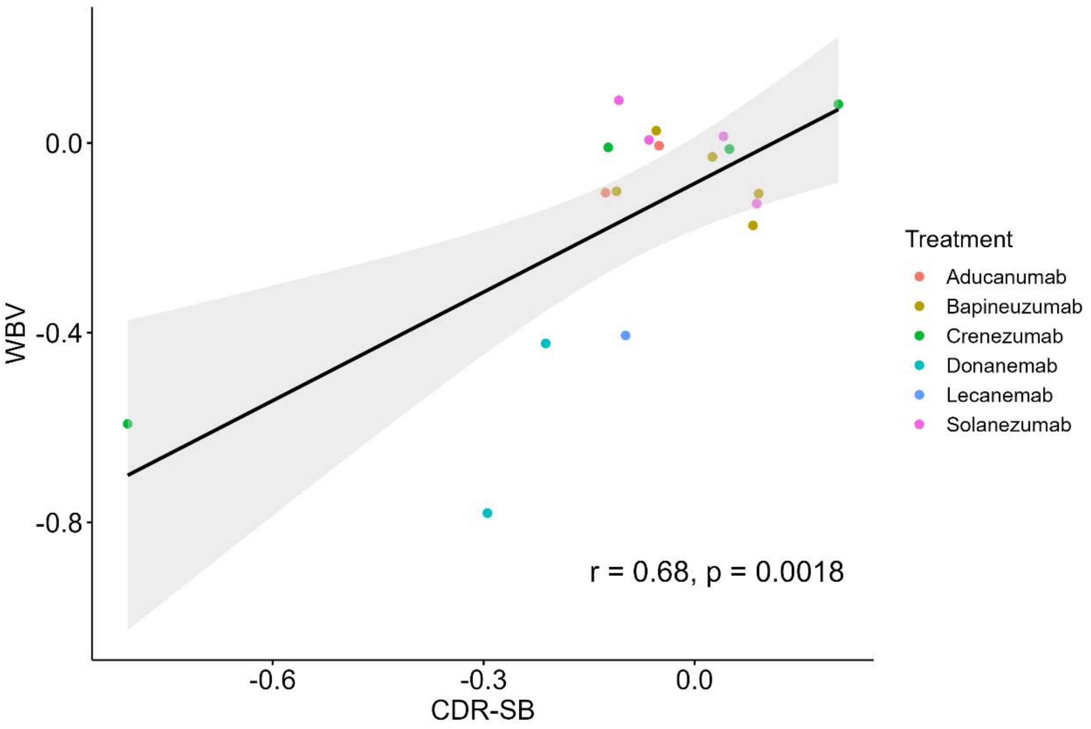
WBV
0.0
-0.4
-0.8
-0.6
-0.3
0.0
CDR-SB
r = 0.68, p = 0.0018
Treatment
Aducanumab
Bapineuzumab
Crenezumab
Donanemab
Lecanemab
Solanezumab

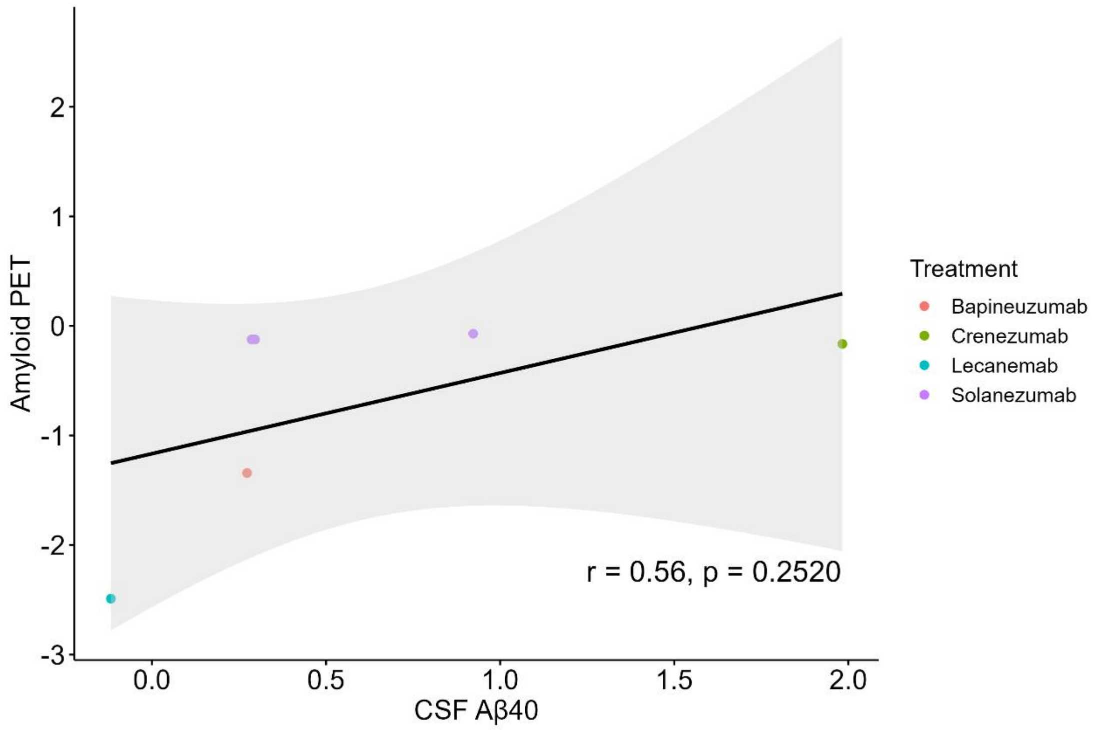

Amyloid PET
CSF Aβ40
Treatment
Bapineuzumab
Crenezumab
Lecanemab
Solanezumab
r = 0.56, p = 0.2520

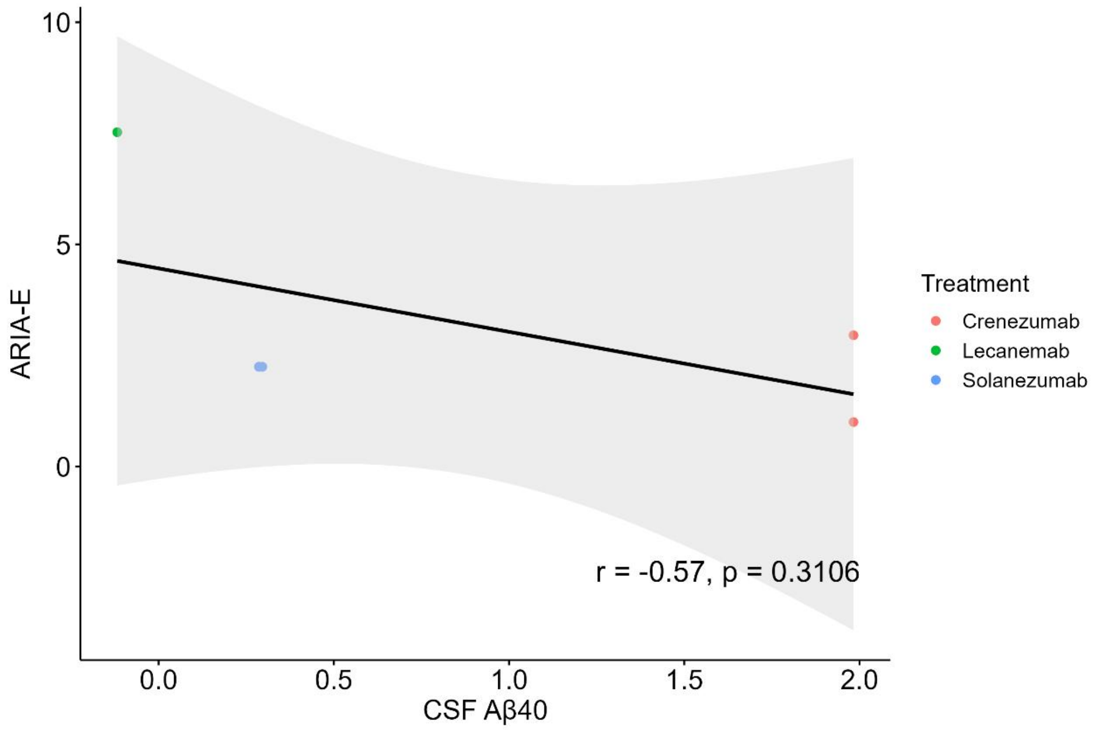

ARIA-E
CSF Aβ40
Treatment
Crenezumab
Lecanemab
Solanezumab
r = -0.57, p = 0.3106

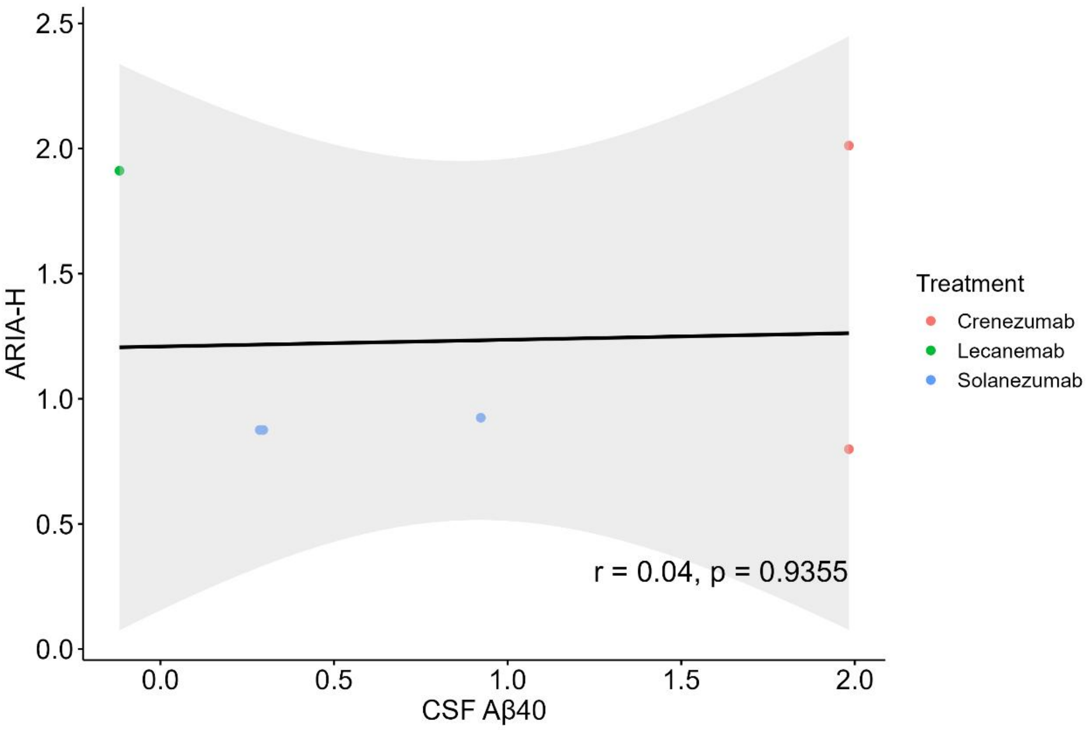
2.5
2.0
1.5
1.0
0.5
0.0
ARIA-H
0.0
0.5
1.0
1.5
2.0
CSF Aβ40
r = 0.04, p = 0.9355
Treatment
Crenezumab
Lecanemab
Solanezumab

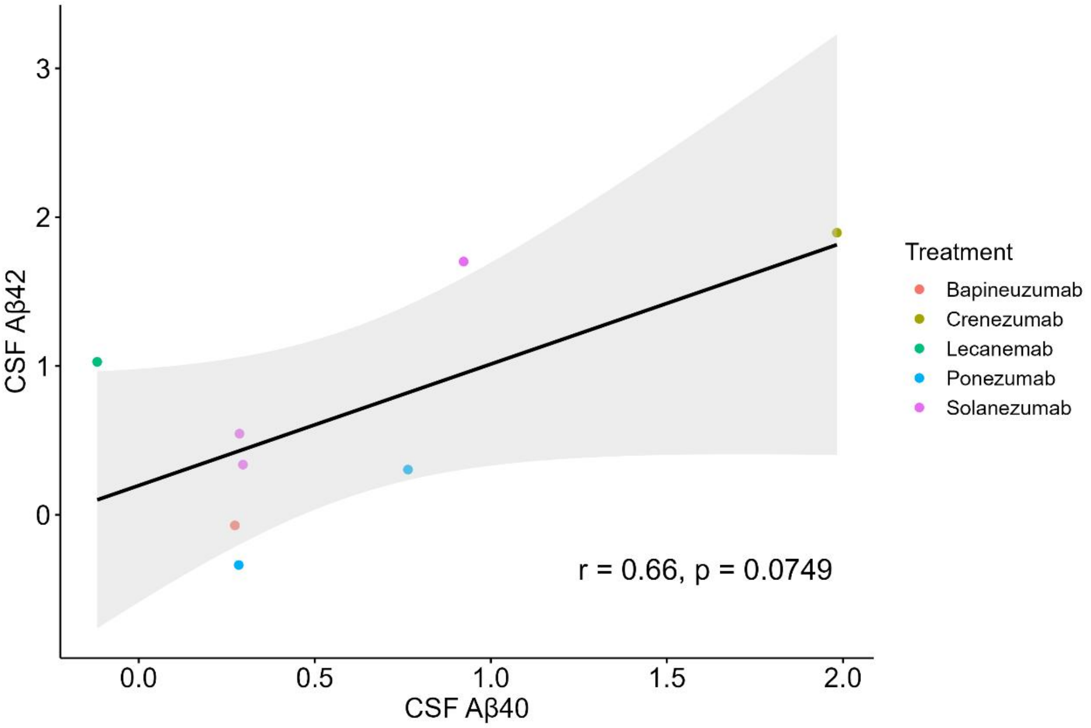
3
2
1
0
CSF Aβ42
0.0
0.5
1.0
1.5
2.0
CSF Aβ40
r = 0.66, p = 0.0749
Treatment
Bapineuzumab
Crenezumab
Lecanemab
Ponezumab
Solanezumab

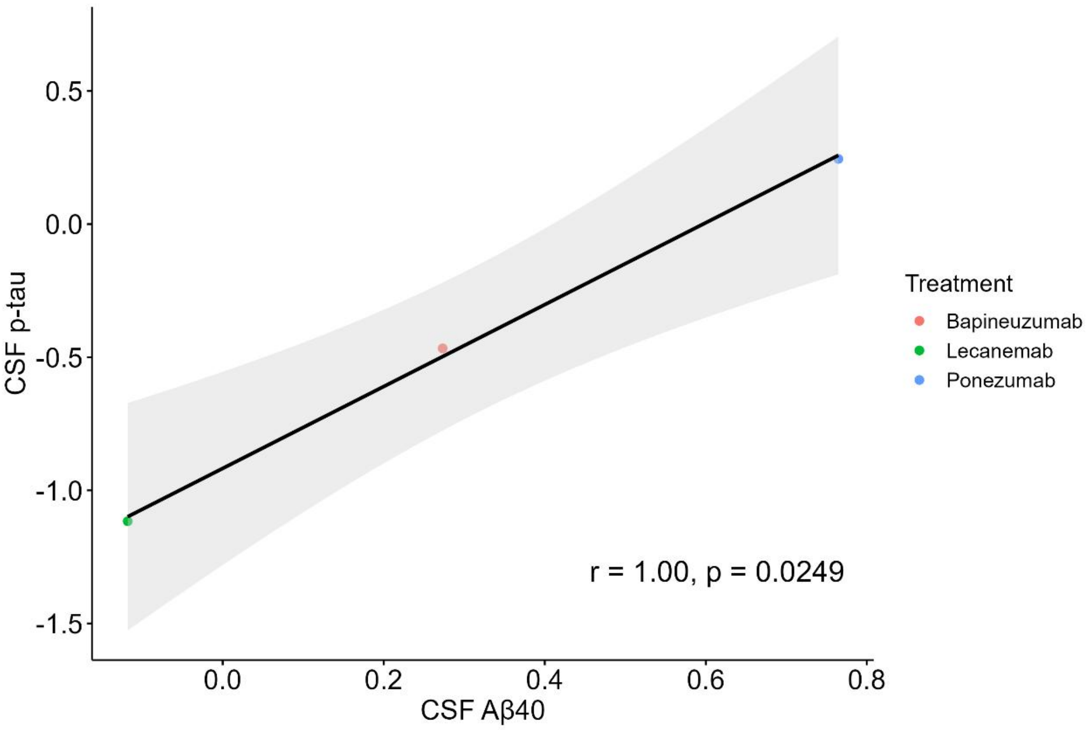

CSF p-tau
0.5
0.0
-0.5
-1.0
-1.5
0.0
0.2
0.4
0.6
0.8
CSF Aβ40
r = 1.00, p = 0.0249
Treatment
Bapineuzumab
Lecanemab
Ponezumab

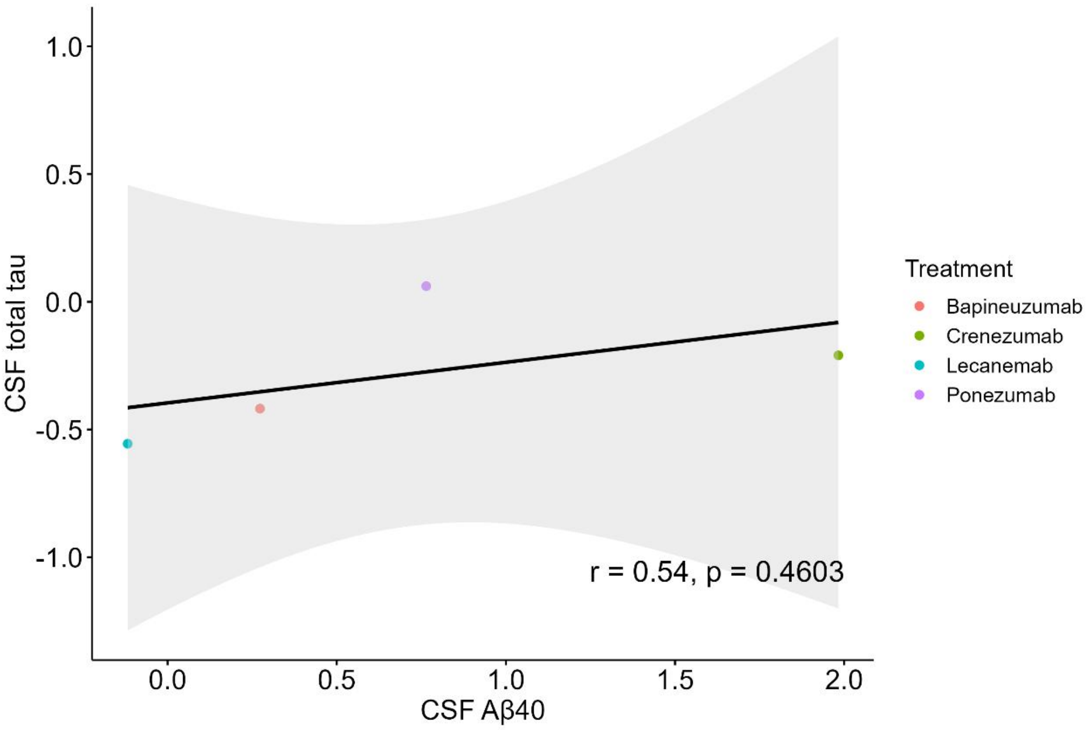

CSF total tau
1.0
0.5
0.0
-0.5
-1.0
0.0
0.5
1.0
1.5
2.0
CSF Aβ40
r = 0.54, p = 0.4603
Treatment
Bapineuzumab
Crenezumab
Lecanemab
Ponezumab

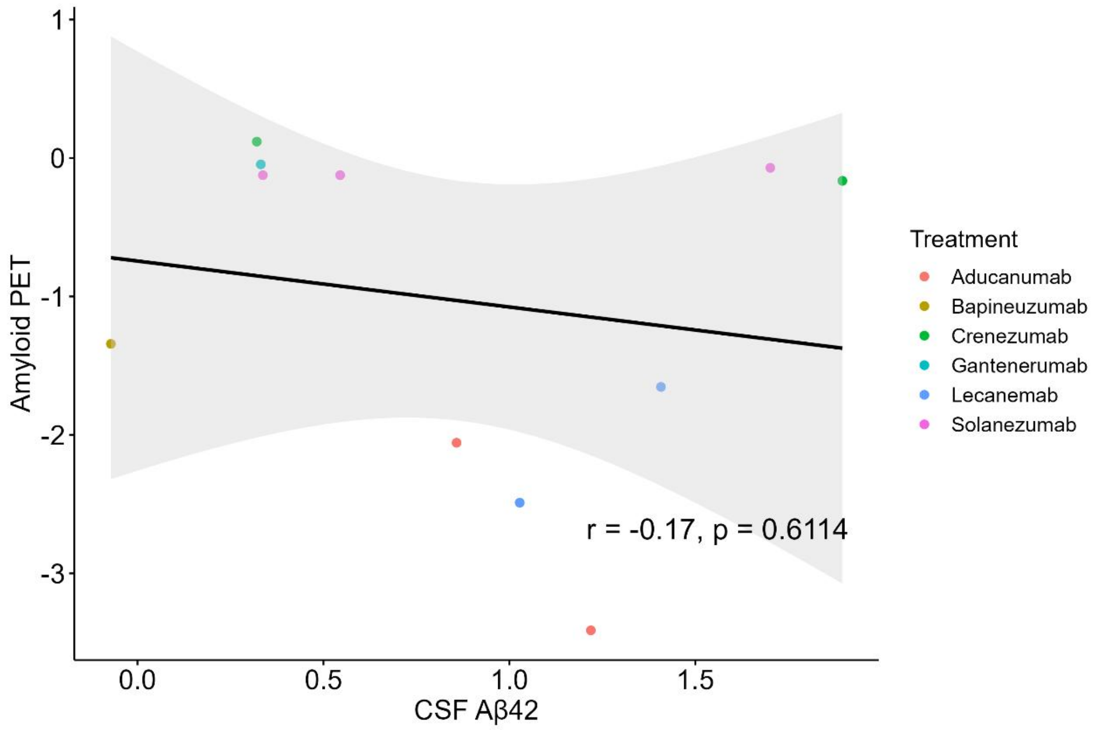
Amyloid PET
CSF Aβ42
r = -0.17, p = 0.6114
Treatment
Aducanumab
Bapineuzumab
Crenezumab
Gantenerumab
Lecanemab
Solanezumab

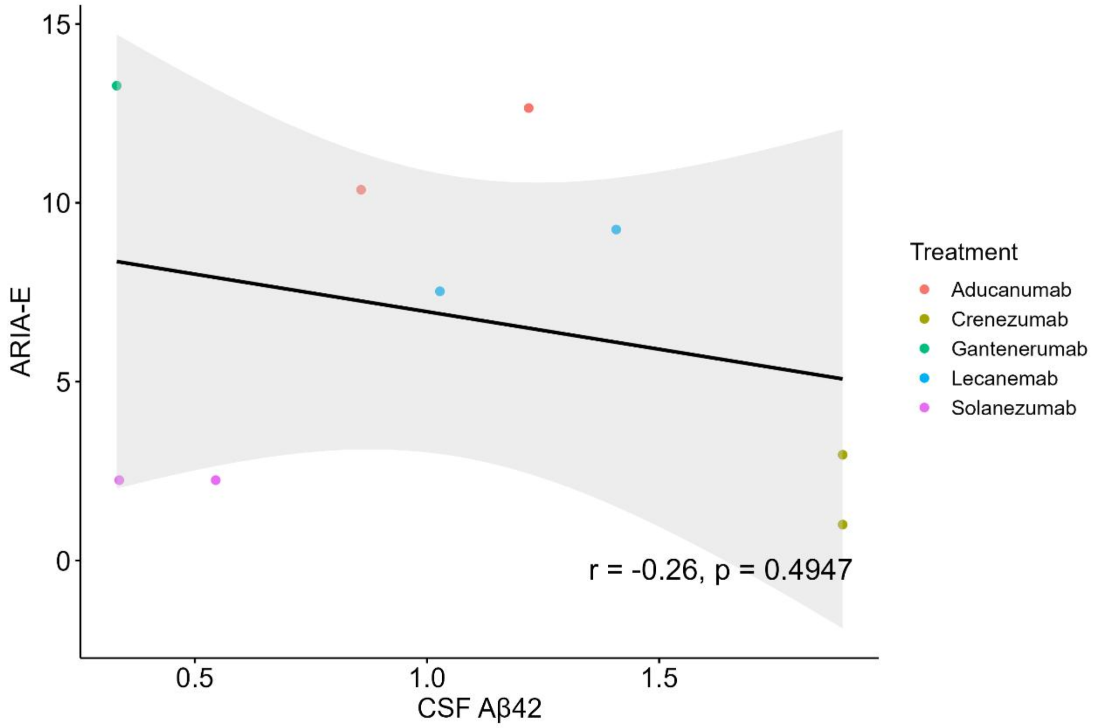
ARIA-E
CSF Aβ42
r = -0.26, p = 0.4947
Treatment
Aducanumab
Crenezumab
Gantenerumab
Lecanemab
Solanezumab

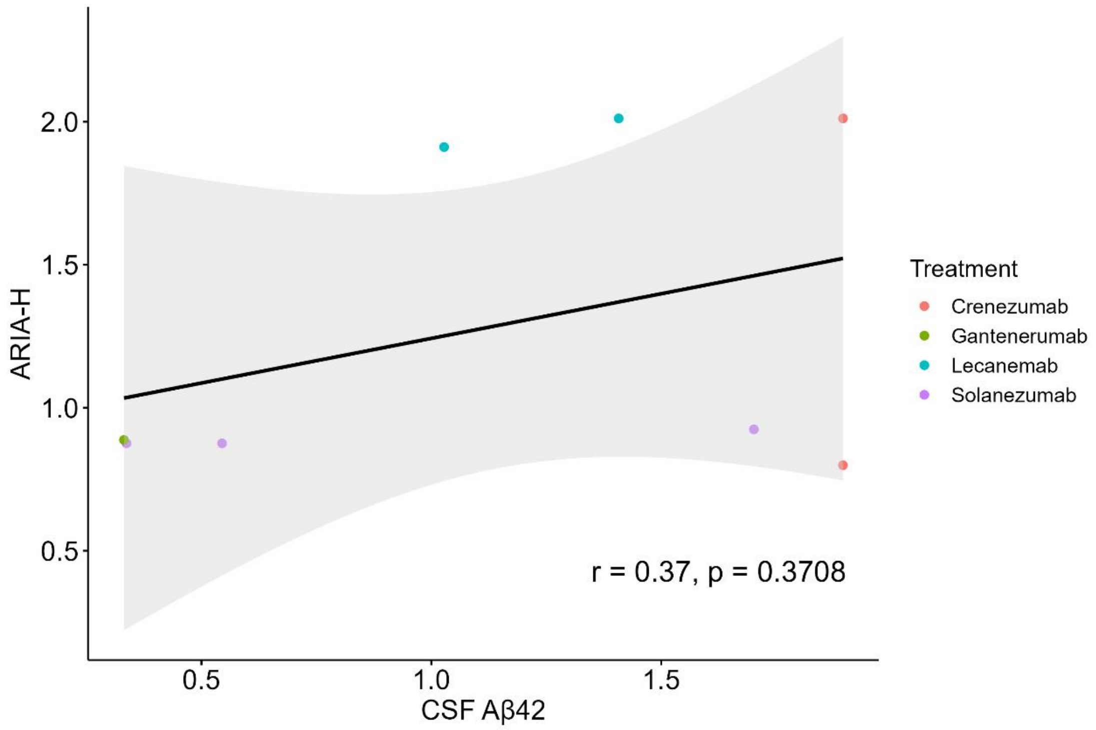

ARIA-H
2.0
1.5
1.0
0.5
r = 0.37, p = 0.3708
0.5
1.0
1.5
CSF Aβ42
Treatment
Crenezumab
Gantenerumab
Lecanemab
Solanezumab

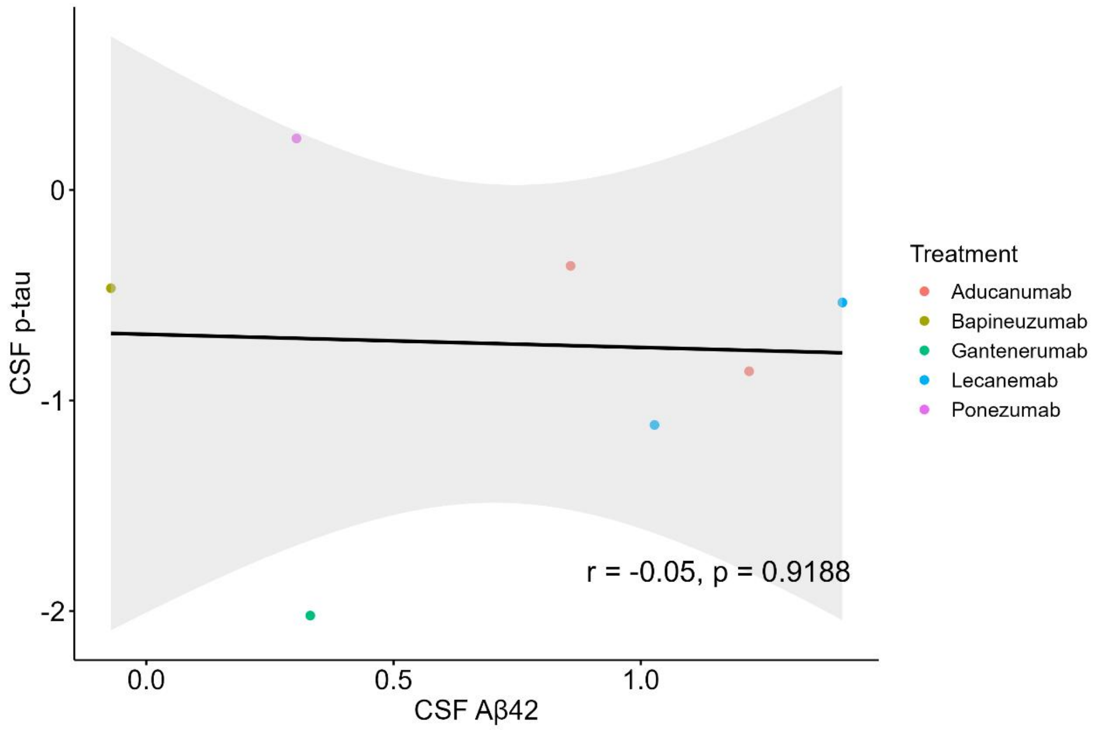

CSF p-tau
0
-1
-2
r = -0.05, p = 0.9188
0.0
0.5
1.0
CSF Aβ42
Treatment
Aducanumab
Bapineuzumab
Gantenerumab
Lecanemab
Ponezumab

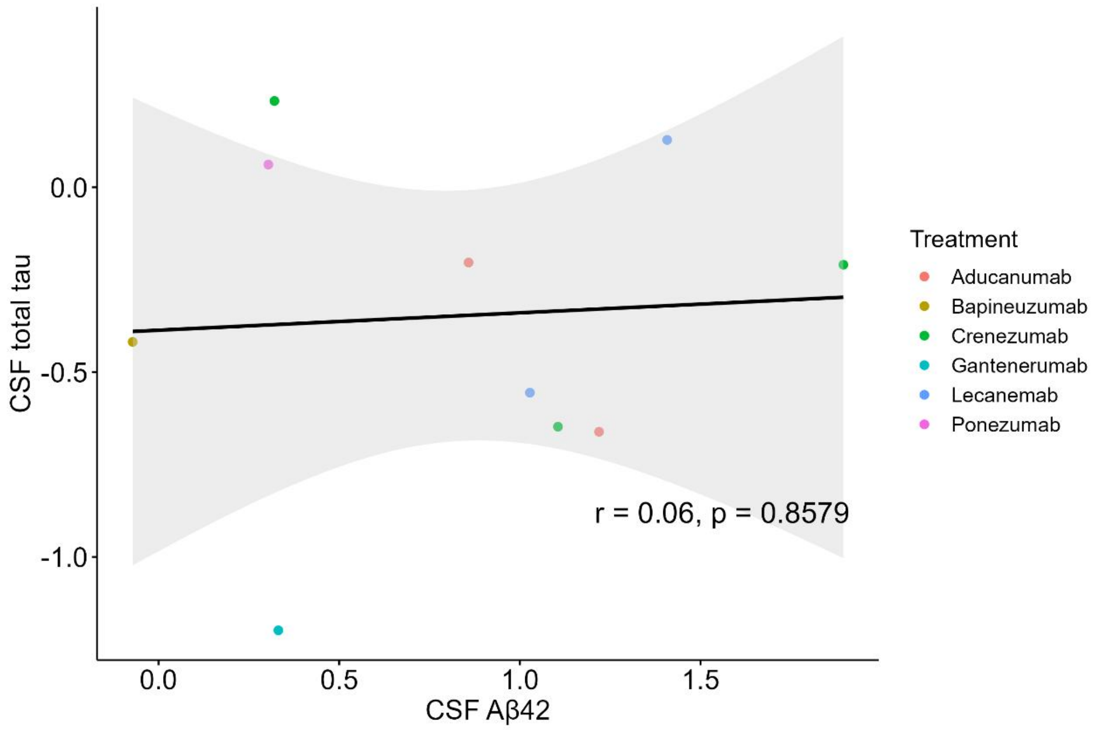
CSF total tau
0.0
-0.5
-1.0
0.0
0.5
1.0
1.5
CSF Aβ42
r = 0.06, p = 0.8579
Treatment
Aducanumab
Bapineuzumab
Crenezumab
Gantenerumab
Lecanemab
Ponezumab

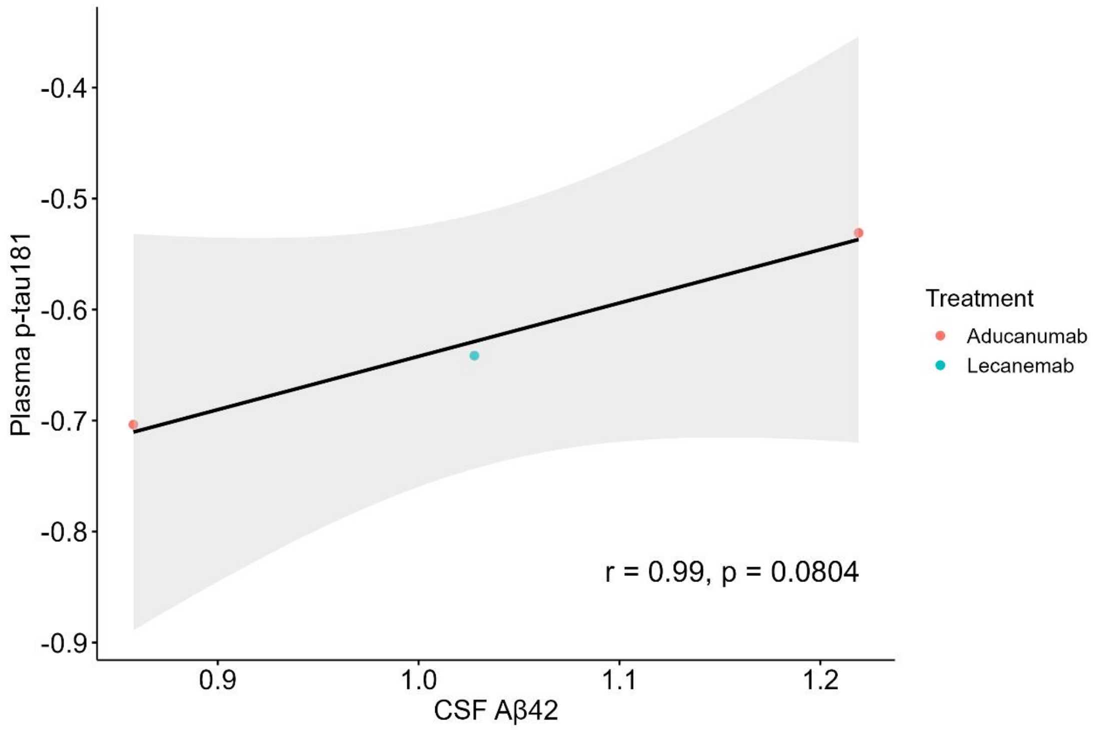
Plasma p-tau181
-0.4
-0.5
-0.6
-0.7
-0.8
-0.9
0.9
1.0
1.1
1.2
CSF Aβ42
r = 0.99, p = 0.0804
Treatment
Aducanumab
Lecanemab

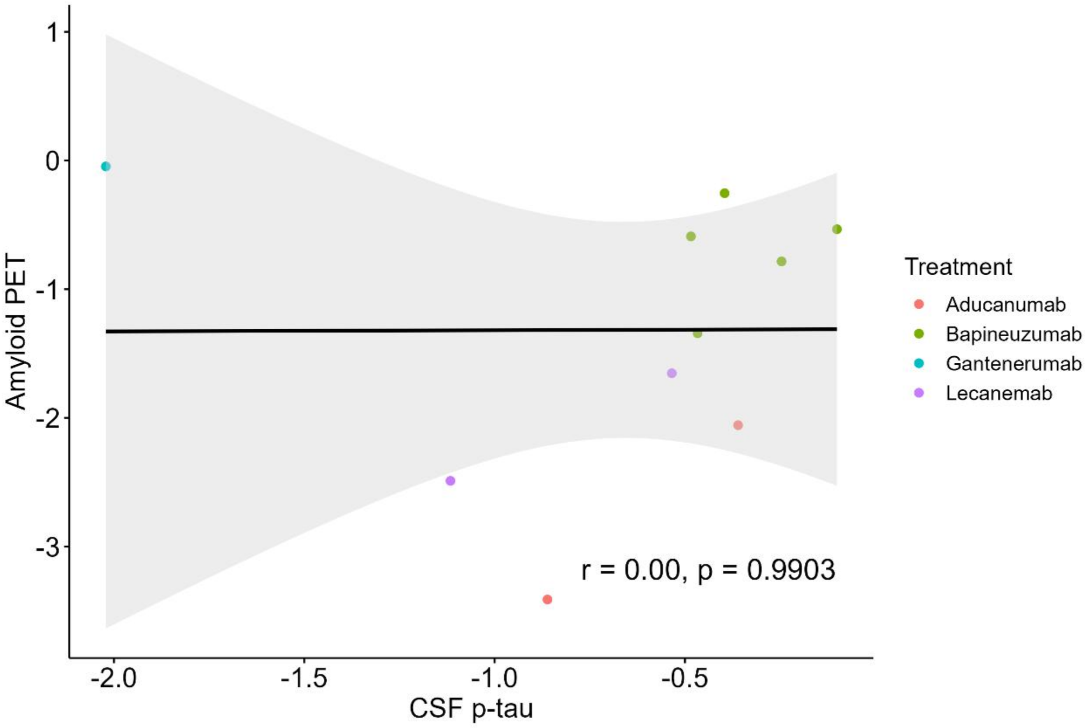
1
0
-1
-2
-3
Amyloid PET
r = 0.00, p = 0.9903
-2.0
-1.5
-1.0
-0.5
CSF p-tau
Treatment
Aducanumab
Bapineuzumab
Gantenerumab
Lecanemab

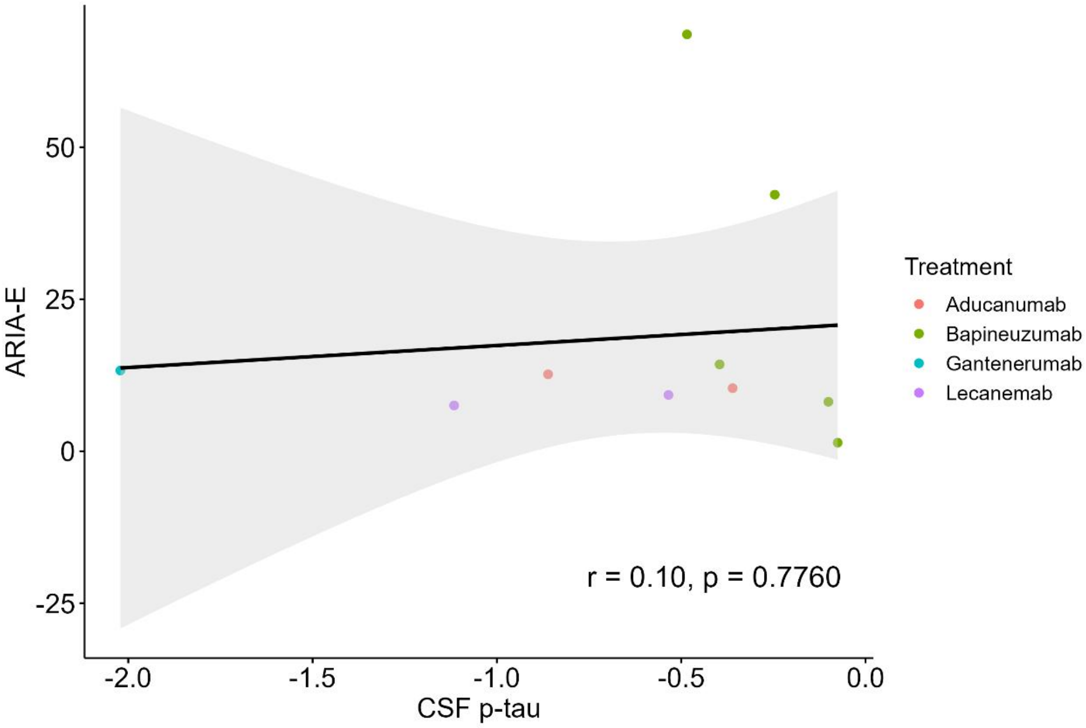
50
25
0
-25
ARIA-E
r = 0.10, p = 0.7760
-2.0
-1.5
-1.0
-0.5
0.0
CSF p-tau
Treatment
Aducanumab
Bapineuzumab
Gantenerumab
Lecanemab

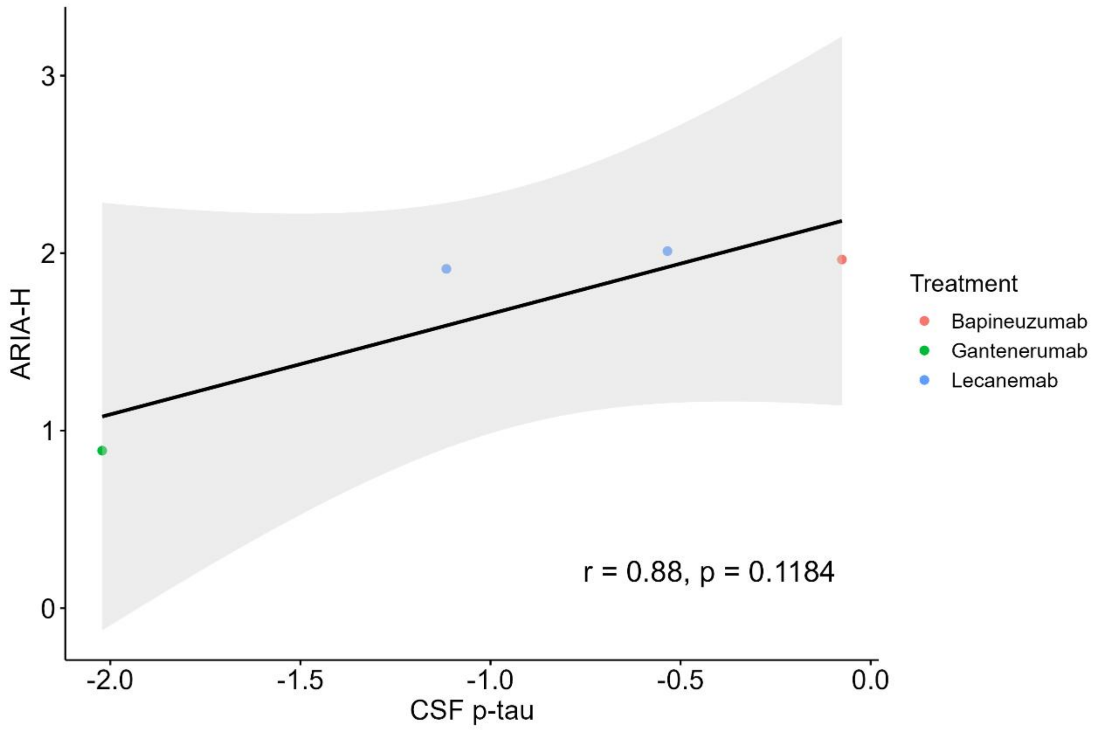
ARIA-H
CSF p-tau
Treatment
Bapineuzumab
Gantenerumab
Lecanemab
r = 0.88, p = 0.1184

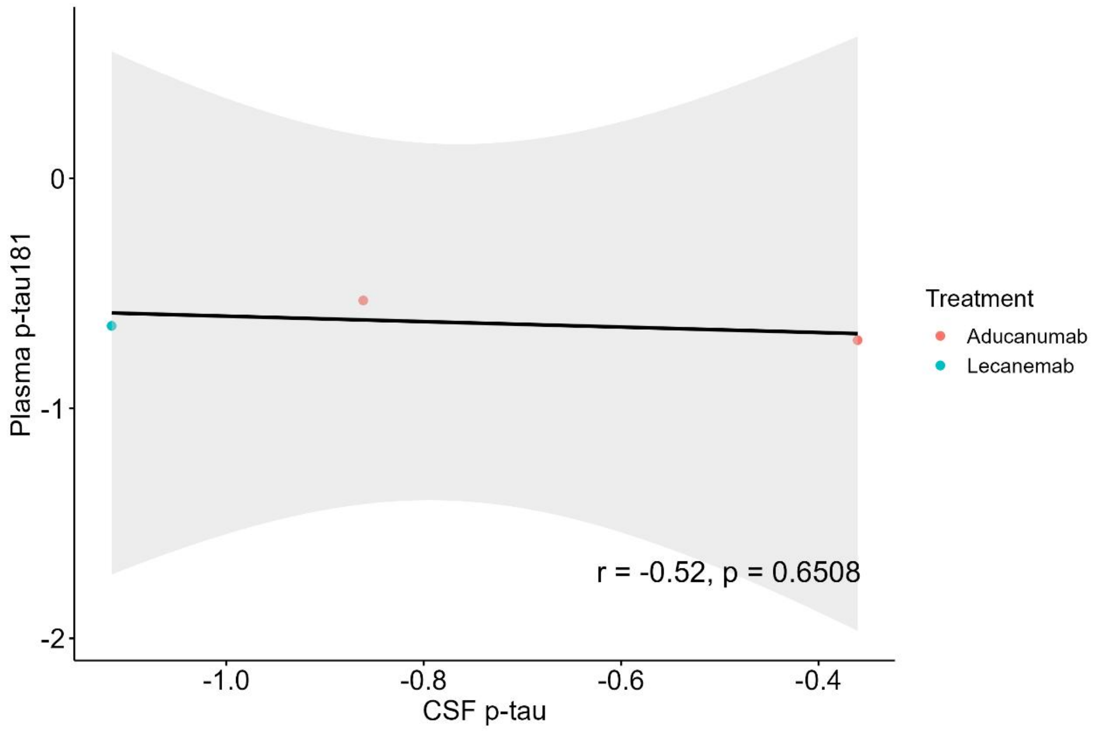
Plasma p-tau181
CSF p-tau
Treatment
Aducanumab
Lecanemab
r = -0.52, p = 0.6508

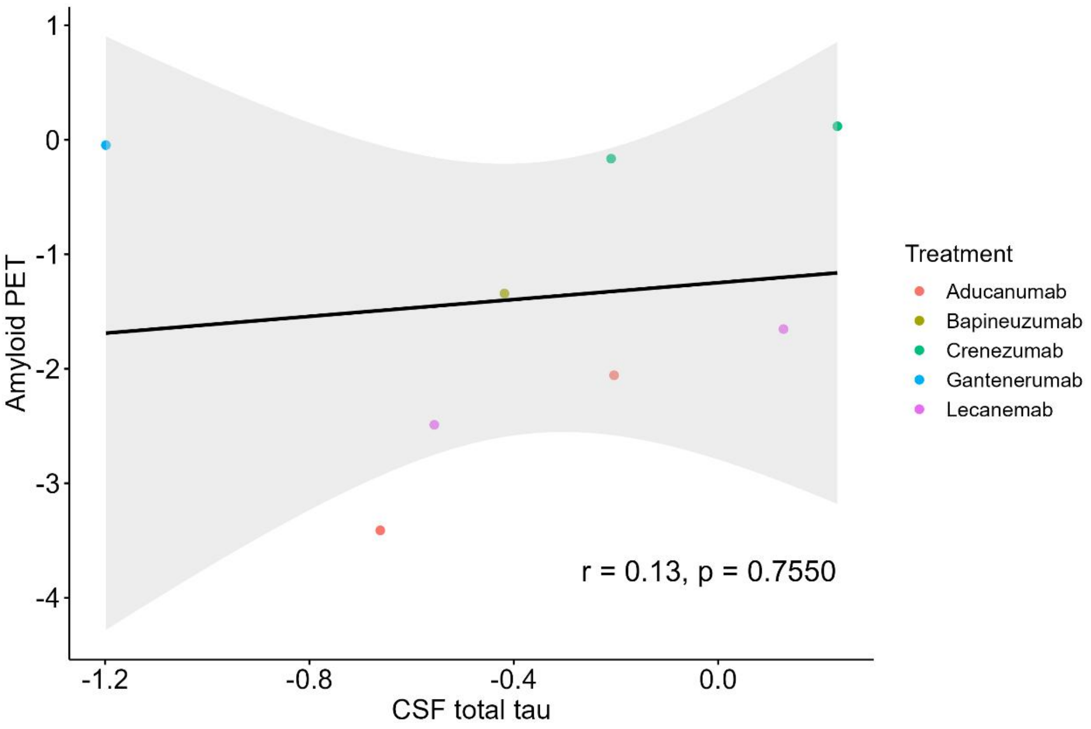
Amyloid PET
1
0
-1
-2
-3
-4
-1.2
-0.8
-0.4
0.0
CSF total tau
r = 0.13, p = 0.7550
Treatment
Aducanumab
Bapineuzumab
Crenezumab
Gantenerumab
Lecanemab

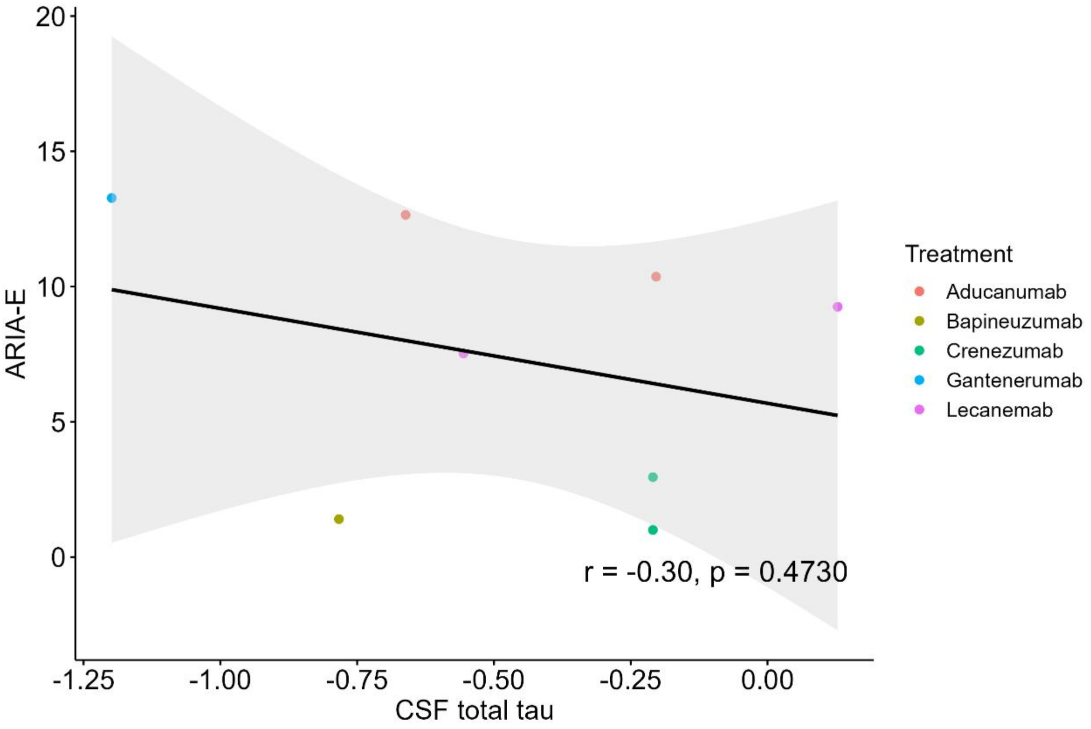
ARIA-E
20
15
10
5
0
-1.25
-1.00
-0.75
-0.50
-0.25
0.00
CSF total tau
r = -0.30, p = 0.4730
Treatment
Aducanumab
Bapineuzumab
Crenezumab
Gantenerumab
Lecanemab

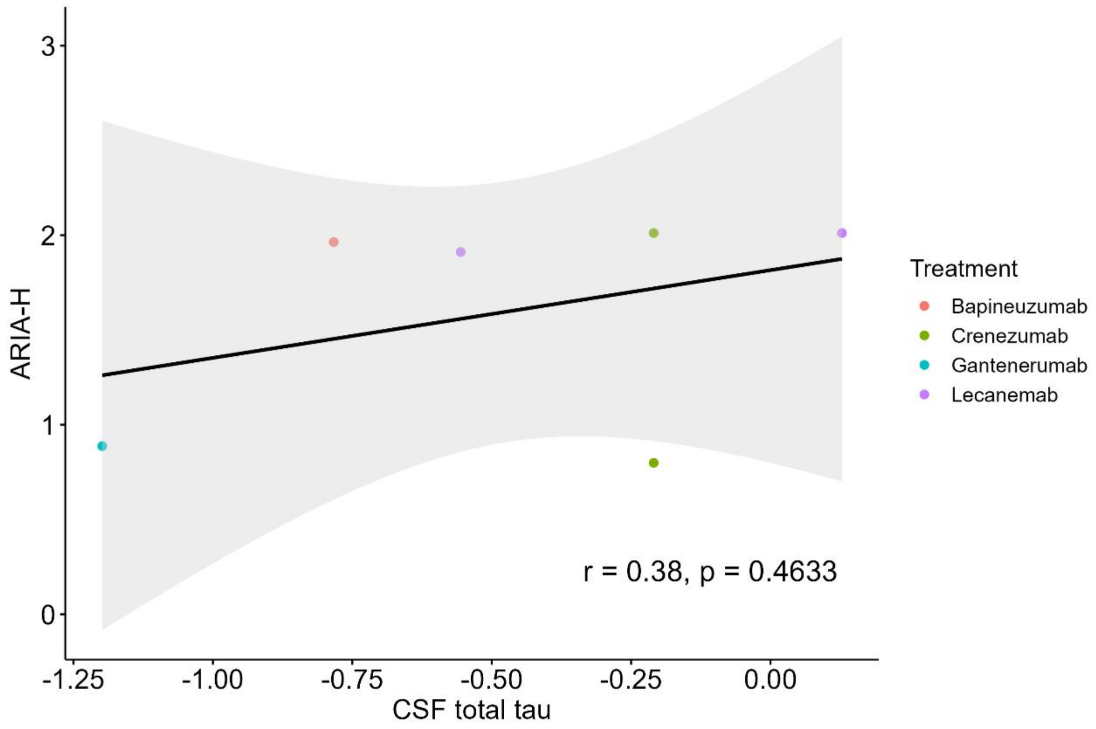

ARIA-H
3
2
1
0
-1.25
-1.00
-0.75
-0.50
-0.25
0.00
CSF total tau
r = 0.38, p = 0.4633
Treatment
Bapineuzumab
Crenezumab
Gantenerumab
Lecanemab

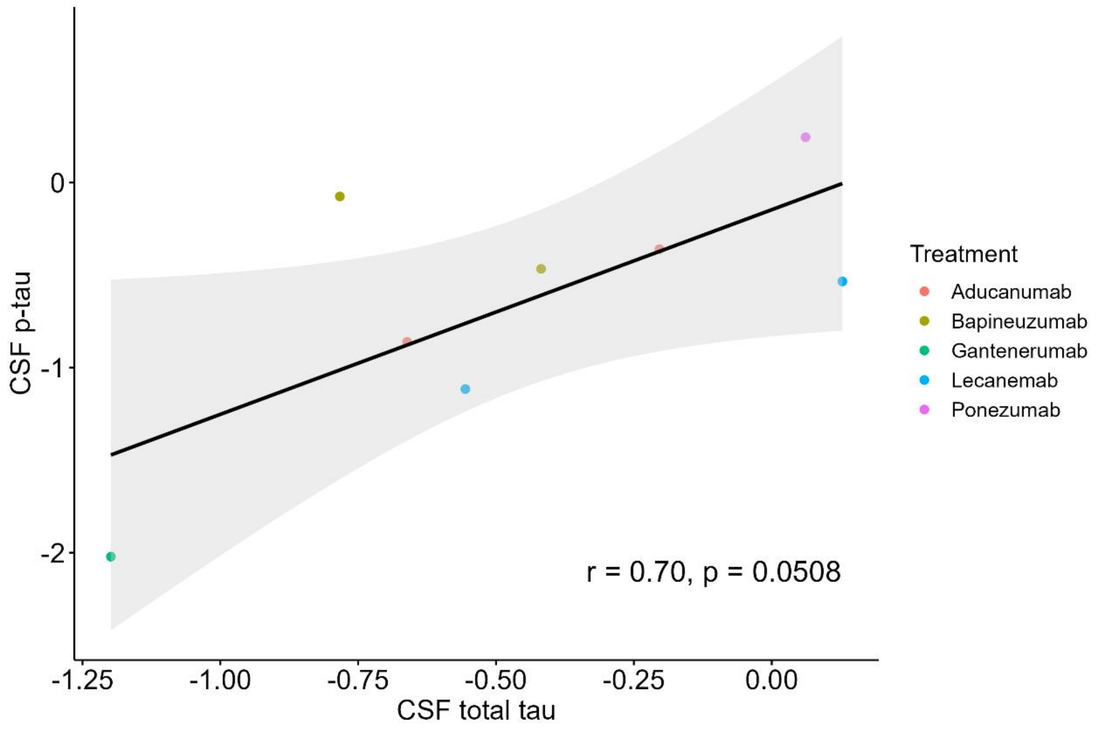

CSF p-tau
0
-1
-2
-1.25
-1.00
-0.75
-0.50
-0.25
0.00
CSF total tau
r = 0.70, p = 0.0508
Treatment
Aducanumab
Bapineuzumab
Gantenerumab
Lecanemab
Ponezumab

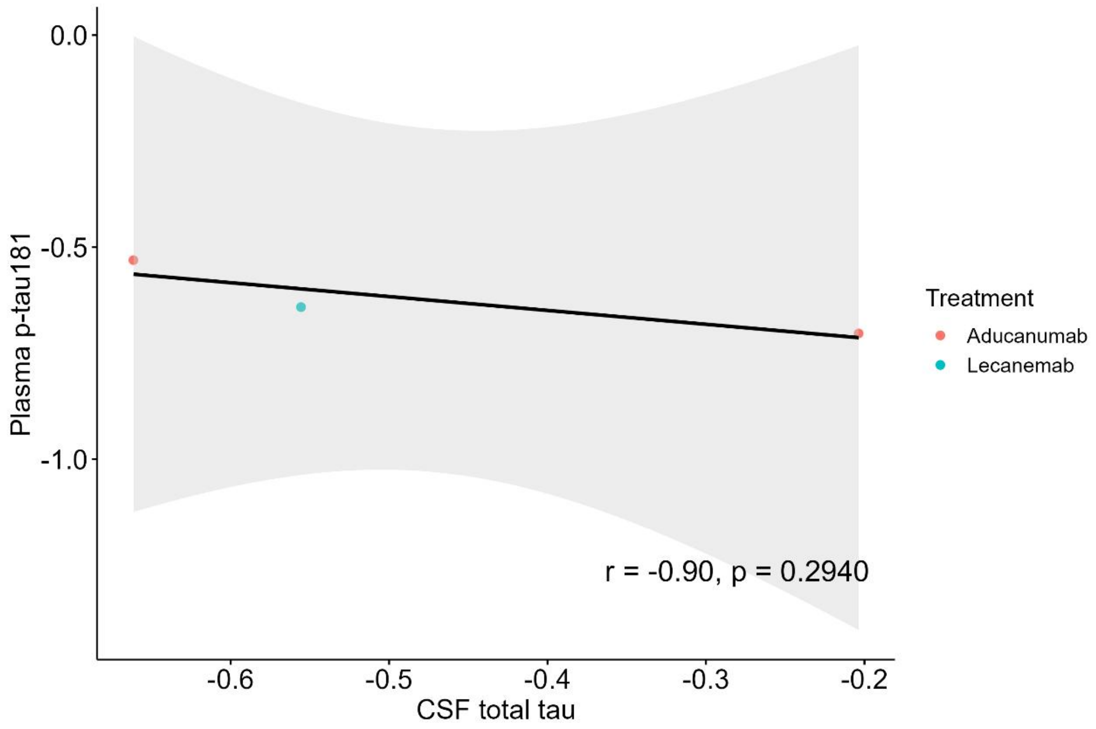
0.0
-0.5
-1.0
Plasma p-tau181
-0.6
-0.5
-0.4
-0.3
-0.2
CSF total tau
r = -0.90, p = 0.2940
Treatment
Aducanumab
Lecanemab

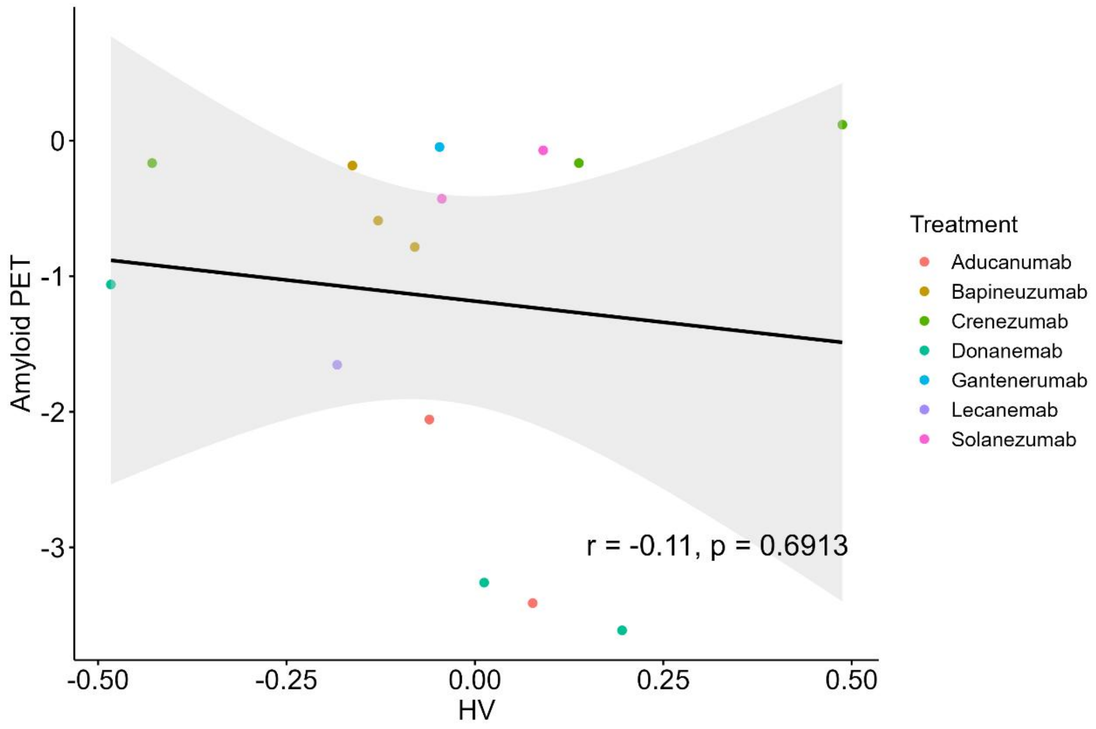
0
-1
-2
-3
Amyloid PET
-0.50
-0.25
0.00
0.25
0.50
HV
r = -0.11, p = 0.6913
Treatment
Aducanumab
Bapineuzumab
Crenezumab
Donanemab
Gantenerumab
Lecanemab
Solanezumab

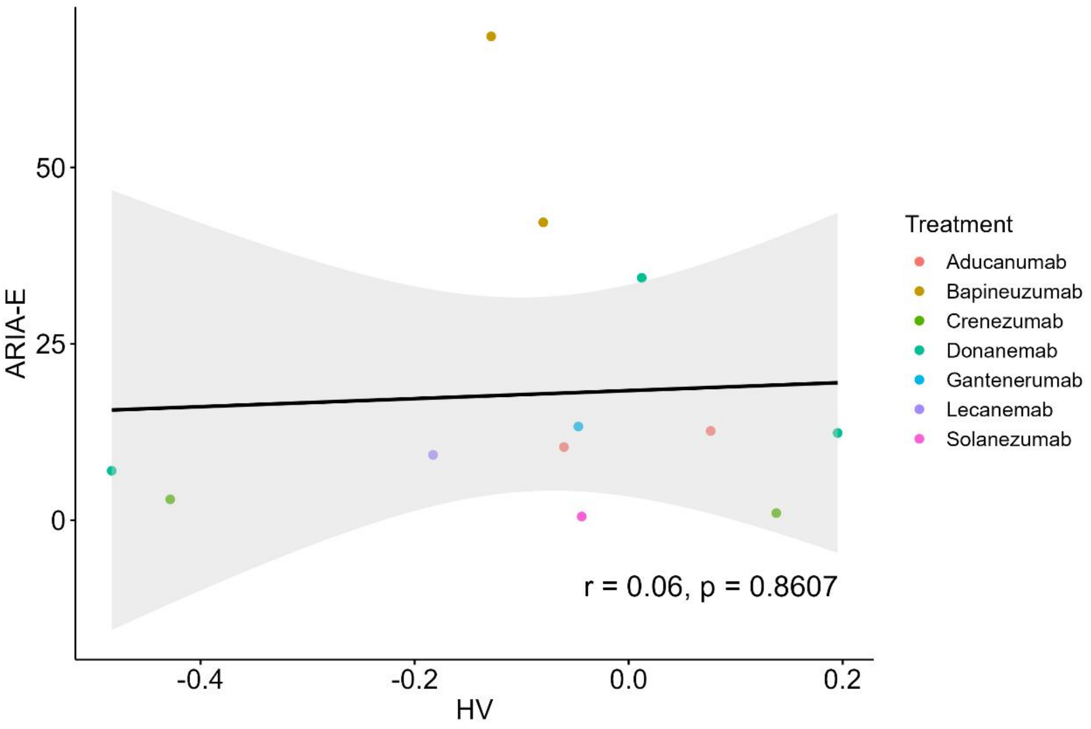
ARIA-E
50
25
0
-0.4
-0.2
0.0
0.2
HV
r = 0.06, p = 0.8607
Treatment
Aducanumab
Bapineuzumab
Crenezumab
Donanemab
Gantenerumab
Lecanemab
Solanezumab

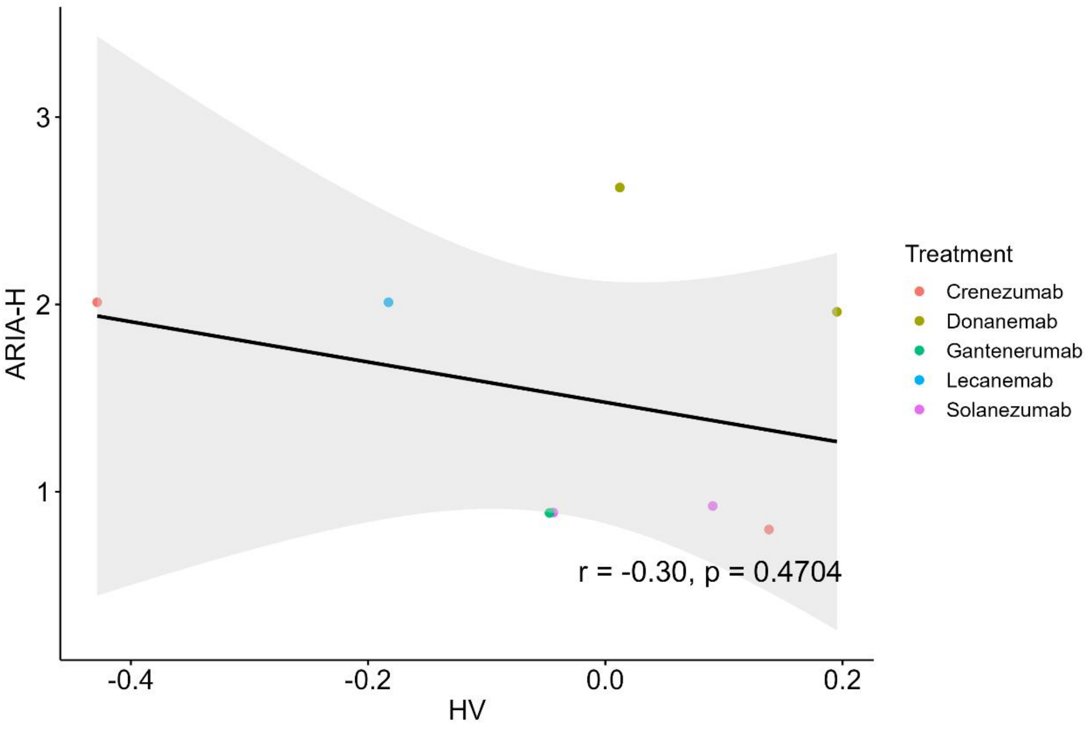
ARIA-H
3
2
1
-0.4
-0.2
0.0
0.2
HV
r = -0.30, p = 0.4704
Treatment
Crenezumab
Donanemab
Gantenerumab
Lecanemab
Solanezumab

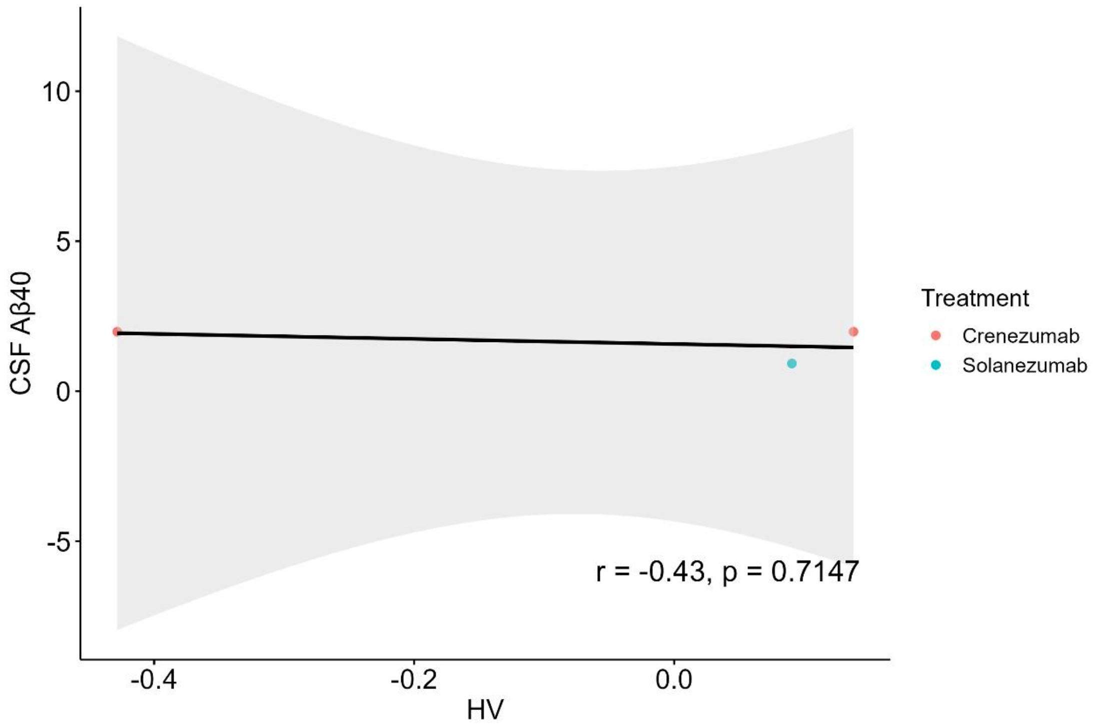
CSF Aβ40
10
5
0
-5
-0.4
-0.2
0.0
HV
r = -0.43, p = 0.7147
Treatment
Crenezumab
Solanezumab

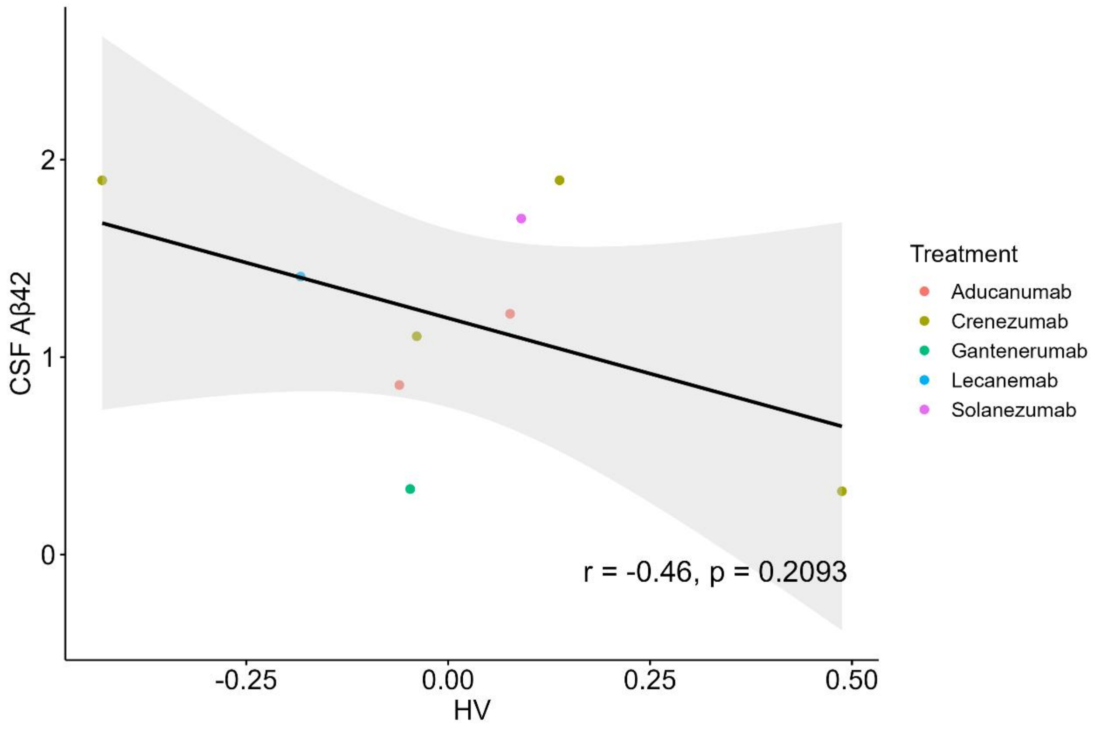
CSF Aβ42
2
1
0
-0.25
0.00
0.25
0.50
HV
r = -0.46, p = 0.2093
Treatment
Aducanumab
Crenezumab
Gantenerumab
Lecanemab
Solanezumab

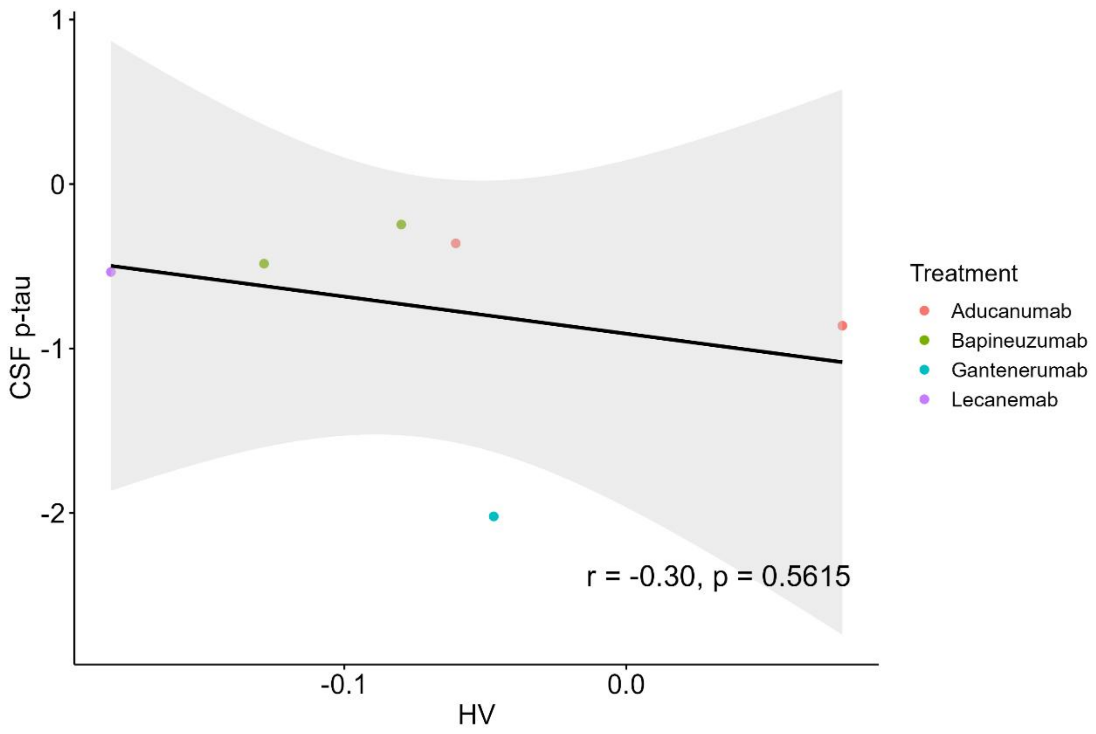
CSF p-tau
HV
-0.1
0.0
1
0
-1
-2
r = -0.30, p = 0.5615
Treatment
Aducanumab
Bapineuzumab
Gantenerumab
Lecanemab

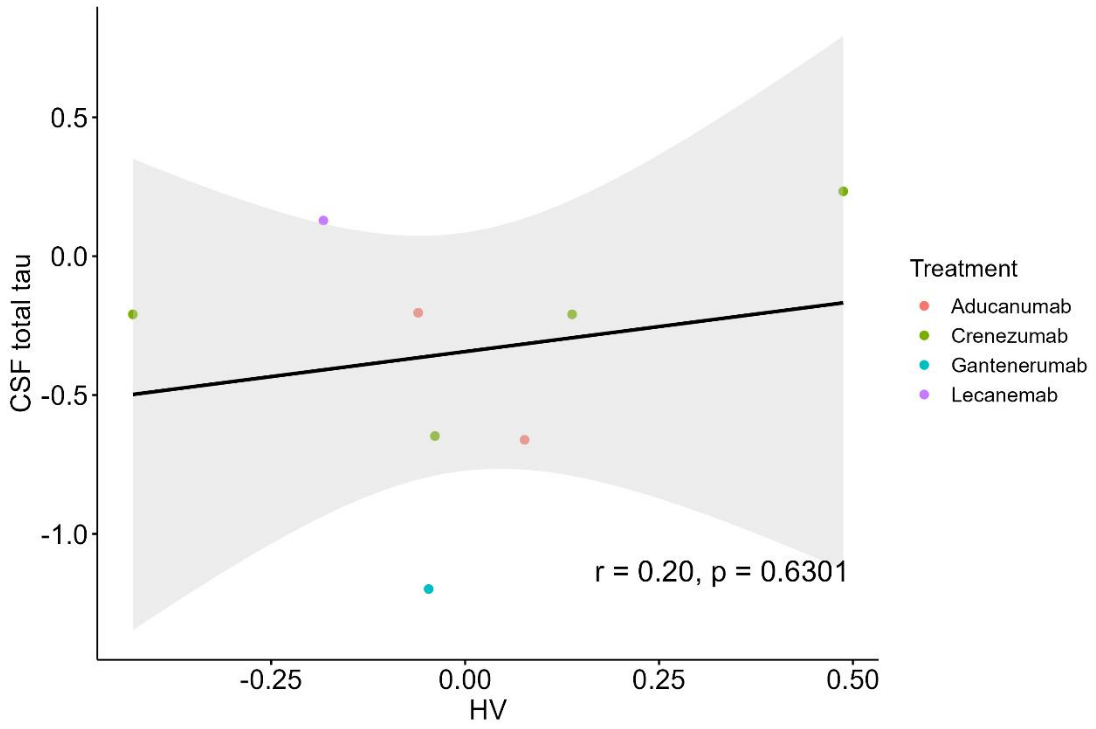
CSF total tau
HV
-0.25
0.00
0.25
0.50
0.5
0.0
-0.5
-1.0
r = 0.20, p = 0.6301
Treatment
Aducanumab
Crenezumab
Gantenerumab
Lecanemab

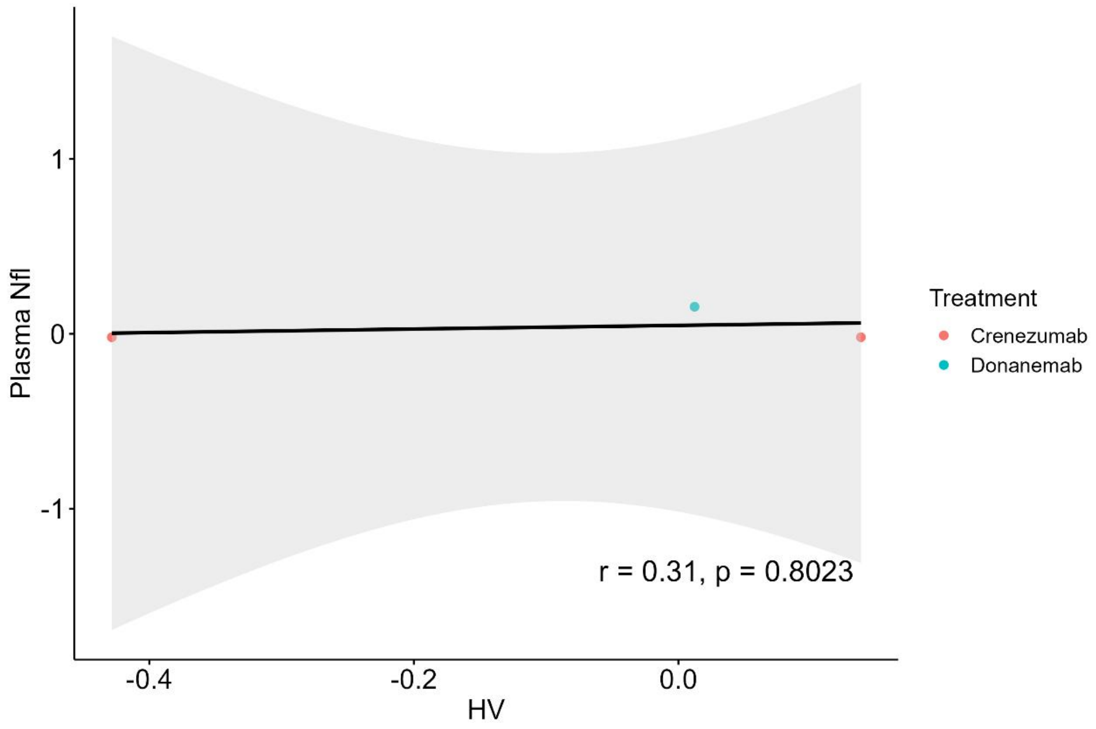
Plasma Nfl
1
0
-1
-0.4
-0.2
0.0
HV
r = 0.31, p = 0.8023
Treatment
Crenezumab
Donanemab

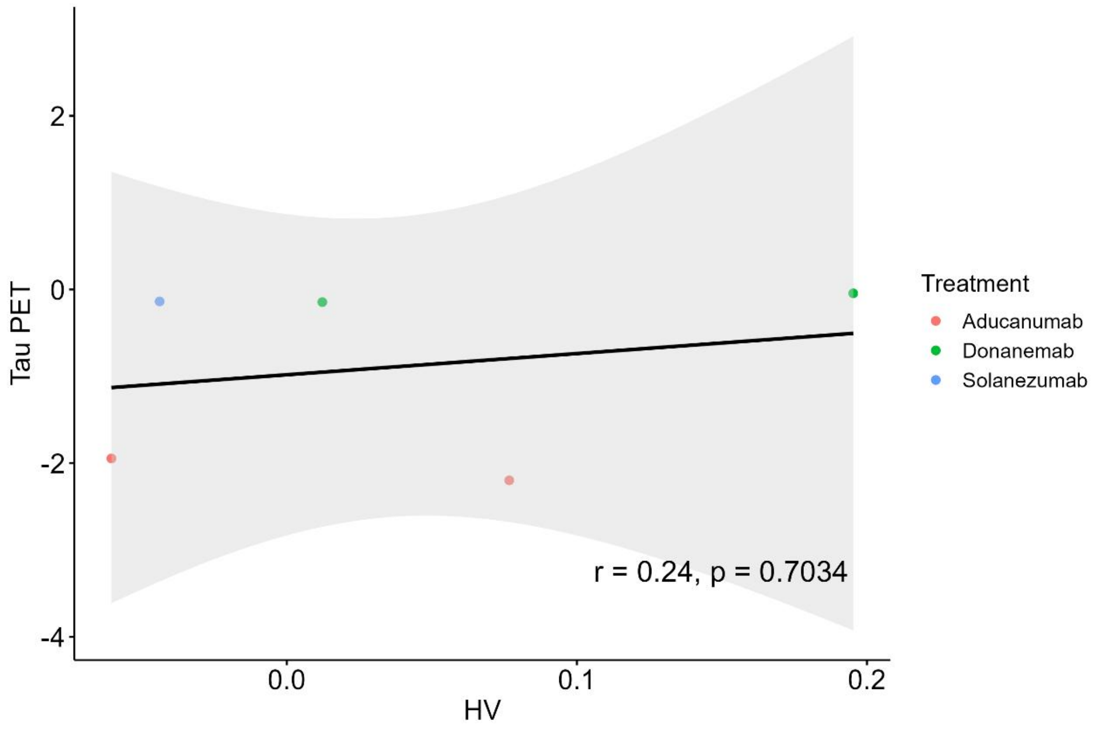
Tau PET
2
0
-2
-4
0.0
0.1
0.2
HV
r = 0.24, p = 0.7034
Treatment
Aducanumab
Donanemab
Solanezumab

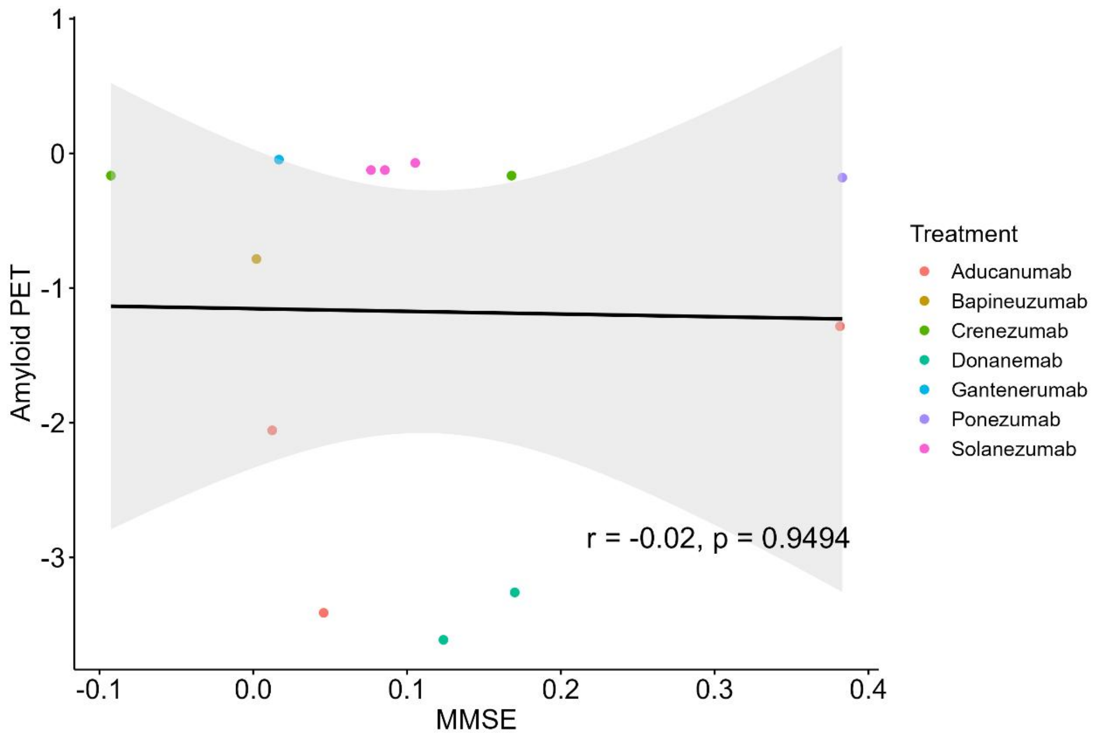
Amyloid PET
MMSE
Treatment
Aducanumab
Bapineuzumab
Crenezumab
Donanemab
Gantenerumab
Ponezumab
Solanezumab
r = -0.02, p = 0.9494

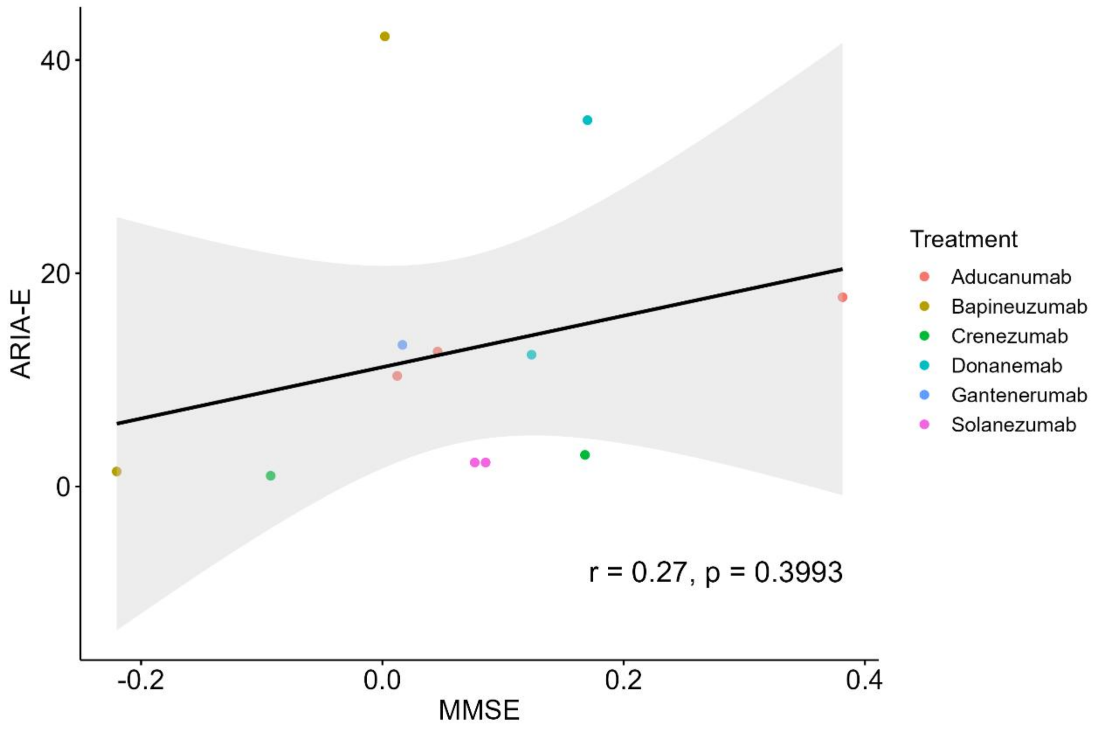
ARIA-E
MMSE
Treatment
Aducanumab
Bapineuzumab
Crenezumab
Donanemab
Gantenerumab
Solanezumab
r = 0.27, p = 0.3993

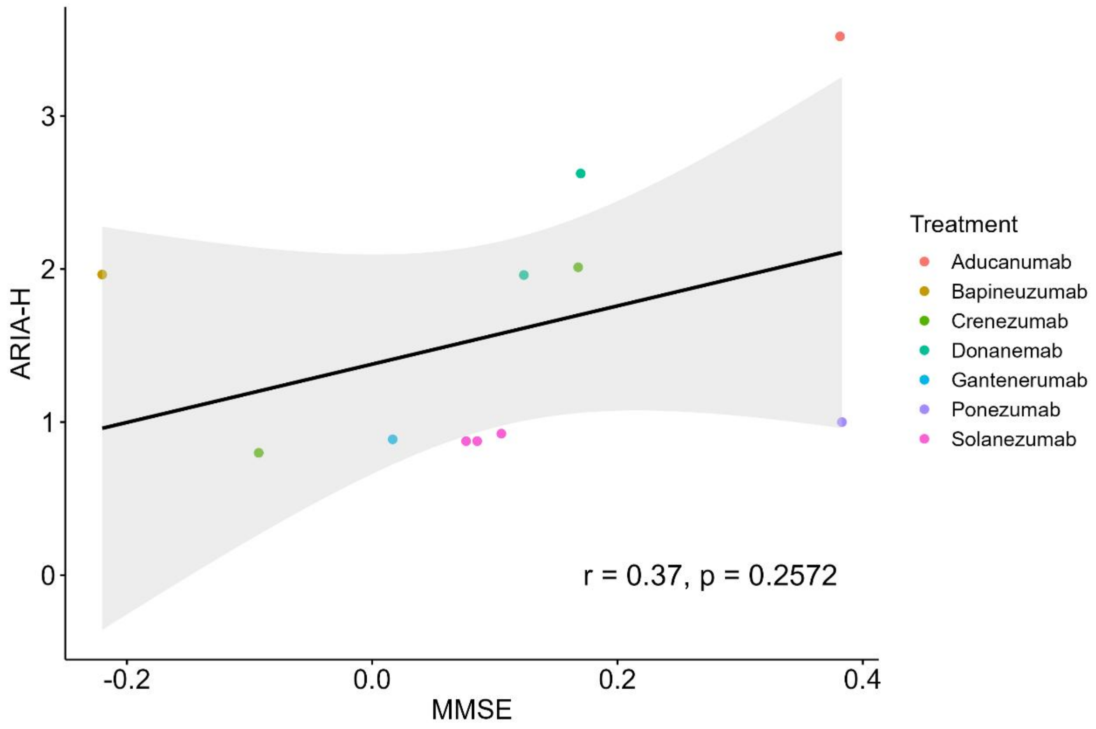
ARIA-H
MMSE
r = 0.37, p = 0.2572
Treatment
Aducanumab
Bapineuzumab
Crenezumab
Donanemab
Gantenerumab
Ponezumab
Solanezumab

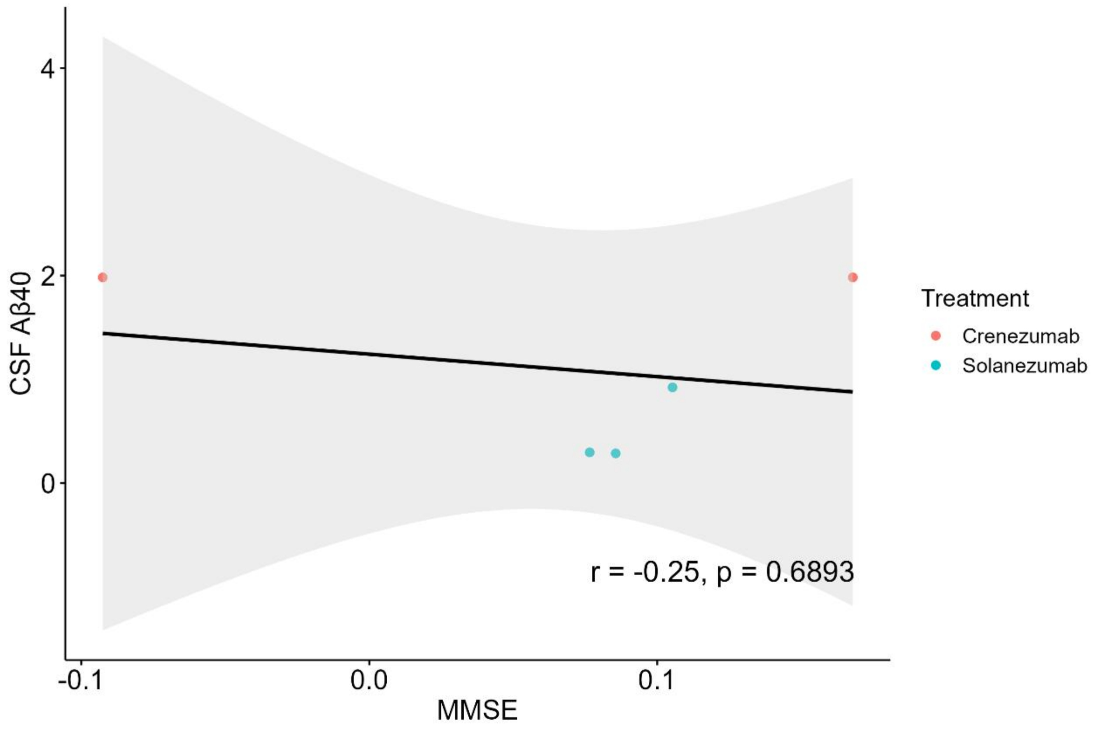
CSF Aβ40
MMSE
r = -0.25, p = 0.6893
Treatment
Crenezumab
Solanezumab

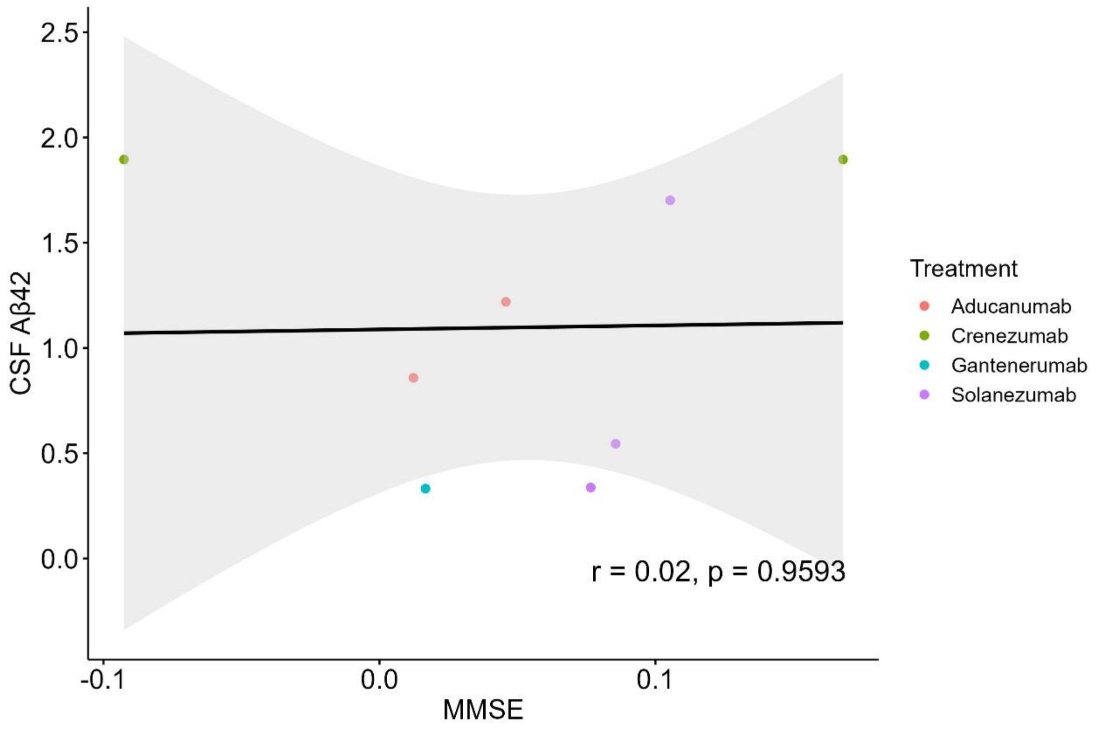

CSF Aβ42
MMSE
Treatment
Aducanumab
Crenezumab
Gantenerumab
Solanezumab
r = 0.02, p = 0.9593

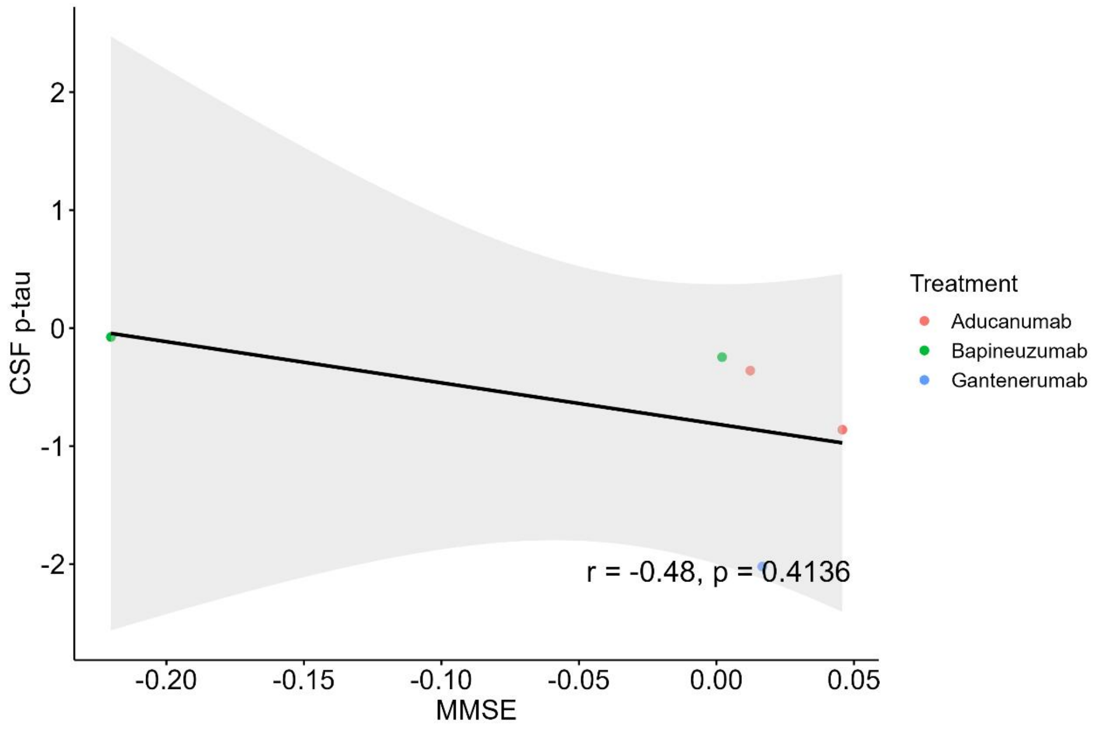

CSF p-tau
MMSE
Treatment
Aducanumab
Bapineuzumab
Gantenerumab
r = -0.48, p = 0.4136

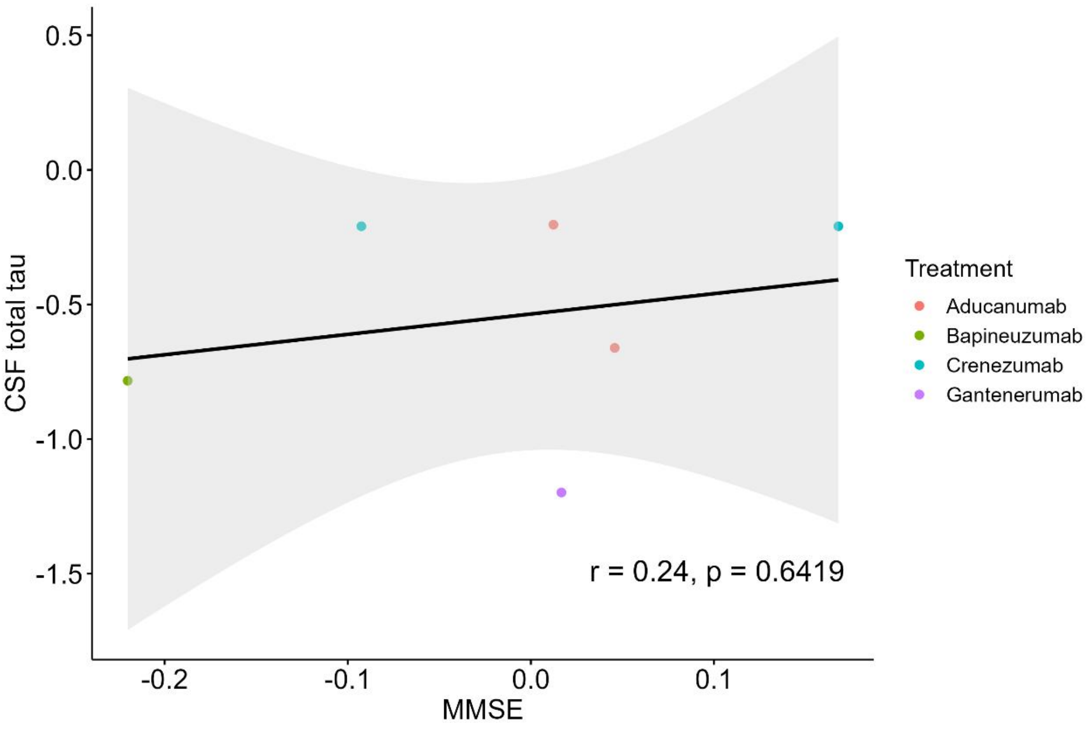
0.5
0.0
-0.5
-1.0
-1.5
CSF total tau
-0.2
-0.1
0.0
0.1
MMSE
r = 0.24, p = 0.6419
Treatment
Aducanumab
Bapineuzumab
Crenezumab
Gantenerumab

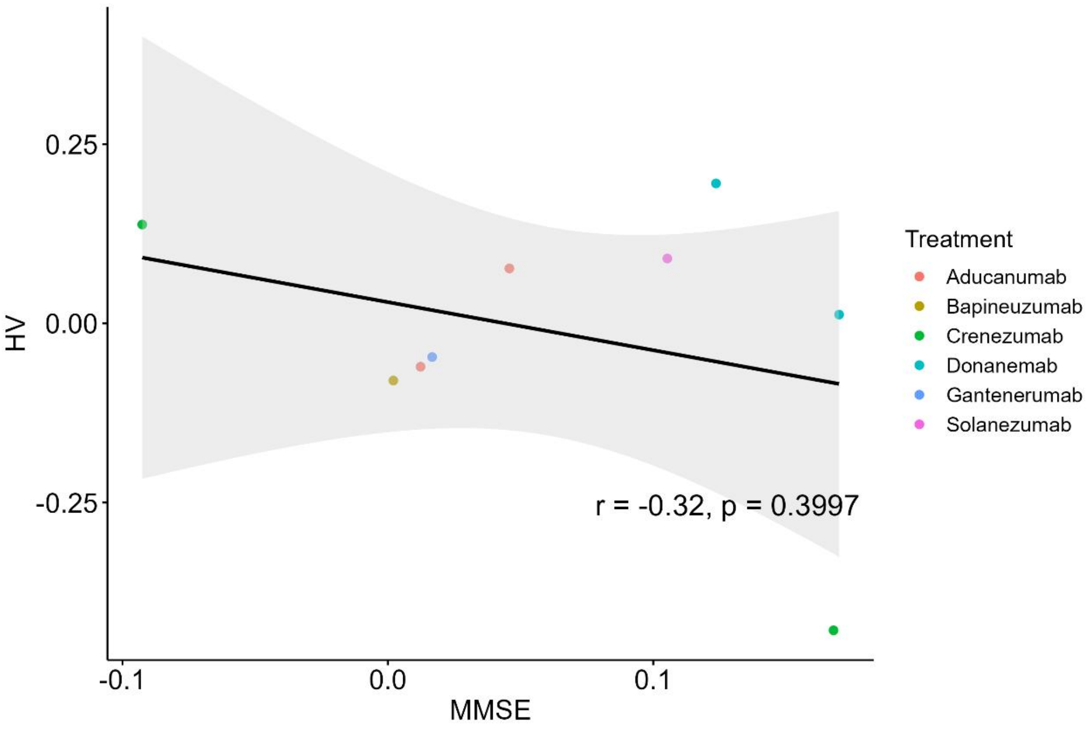
0.25
0.00
-0.25
HV
-0.1
0.0
0.1
MMSE
r = -0.32, p = 0.3997
Treatment
Aducanumab
Bapineuzumab
Crenezumab
Donanemab
Gantenerumab
Solanezumab

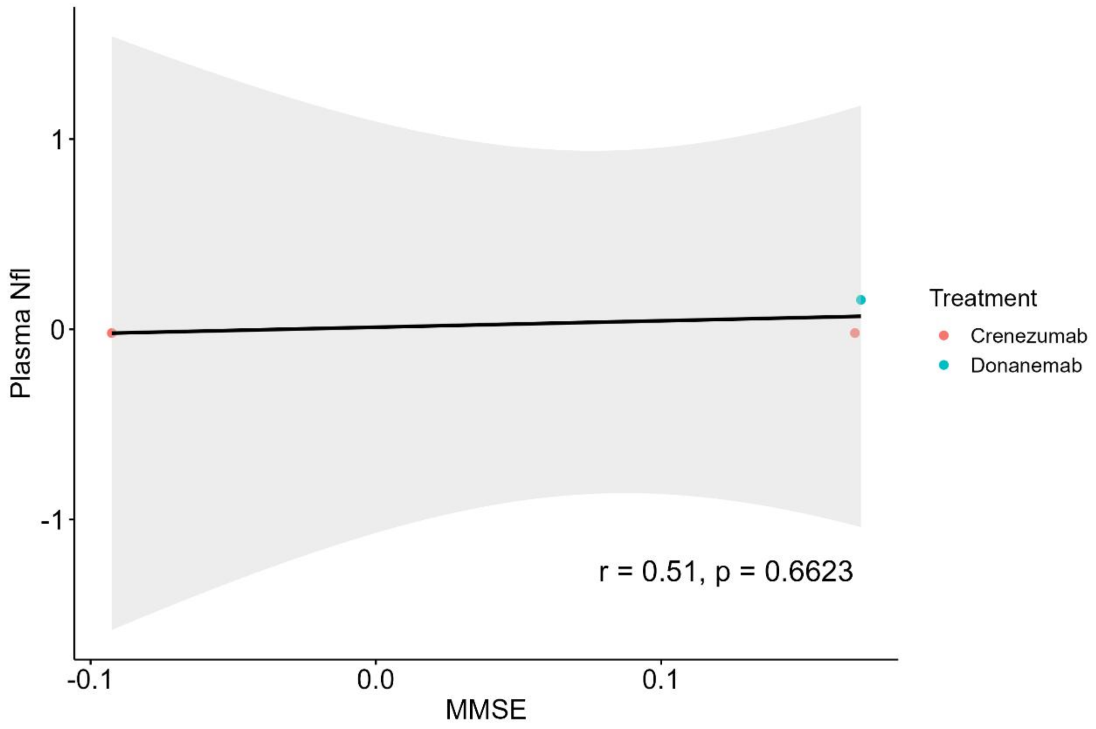
Plasma Nfl
1
0
-1
-0.1
0.0
0.1
MMSE
r = 0.51, p = 0.6623
Treatment
Crenezumab
Donanemab

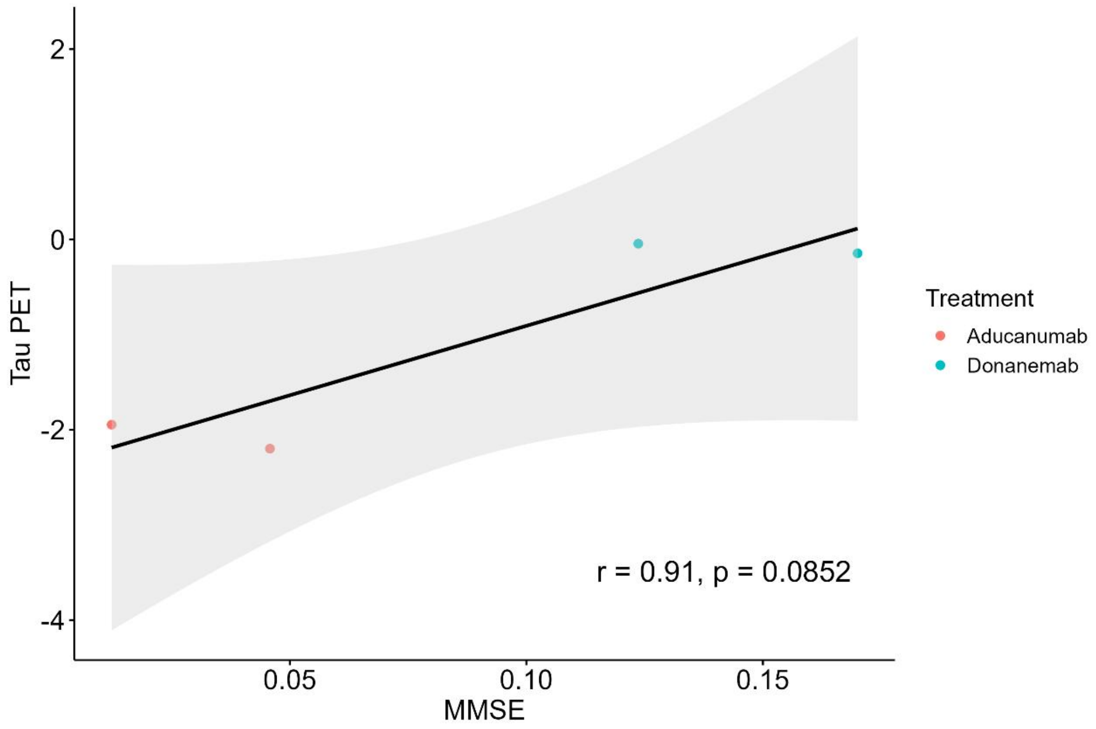
Tau PET
2
0
-2
-4
0.05
0.10
0.15
MMSE
r = 0.91, p = 0.0852
Treatment
Aducanumab
Donanemab

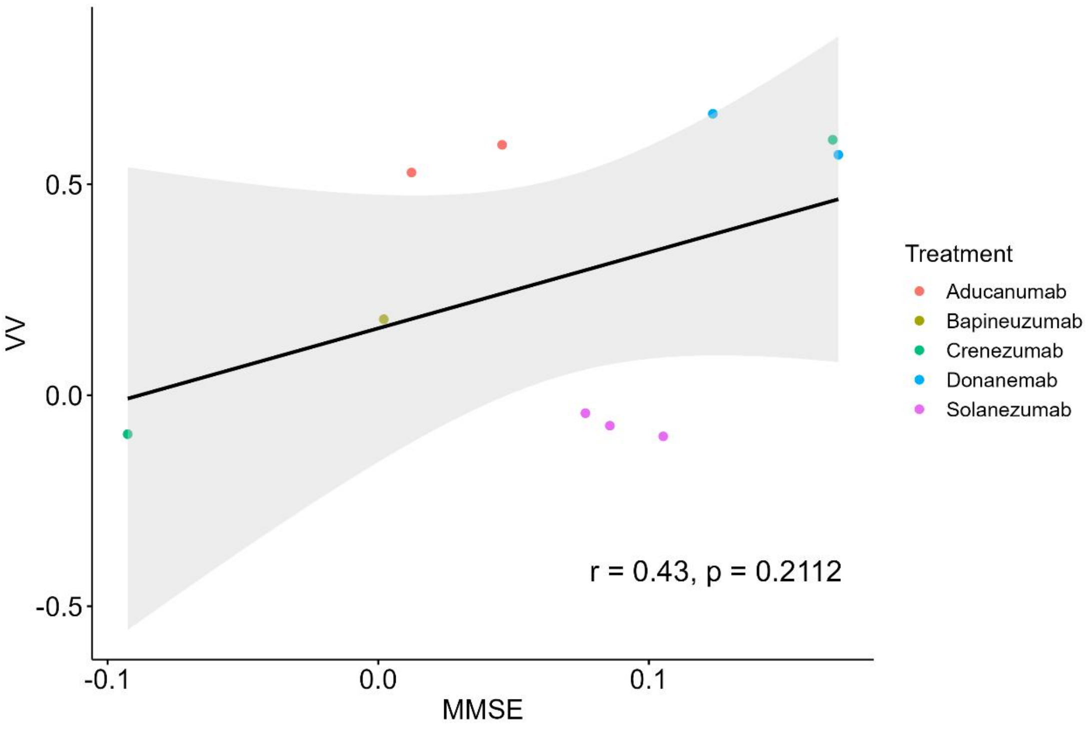

VV
0.5
0.0
-0.5
-0.1
0.0
0.1
MMSE
r = 0.43, p = 0.2112
Treatment
Aducanumab
Bapineuzumab
Crenezumab
Donanemab
Solanezumab

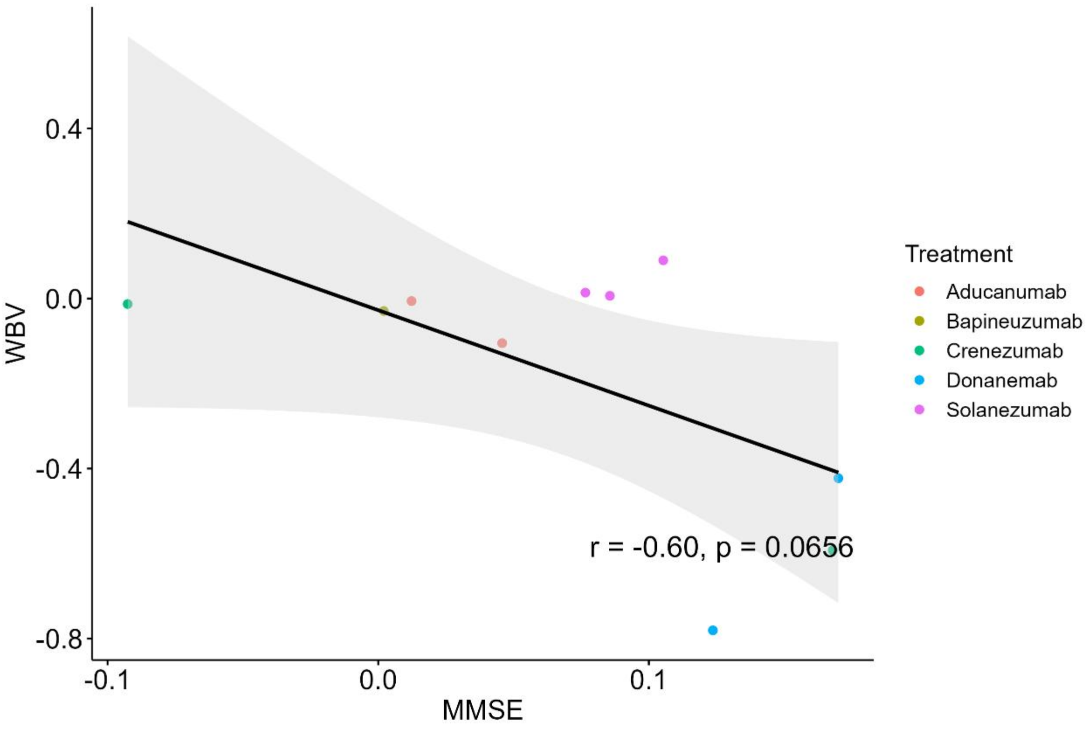

WBV
0.4
0.0
-0.4
-0.8
-0.1
0.0
0.1
MMSE
r = -0.60, p = 0.0656
Treatment
Aducanumab
Bapineuzumab
Crenezumab
Donanemab
Solanezumab

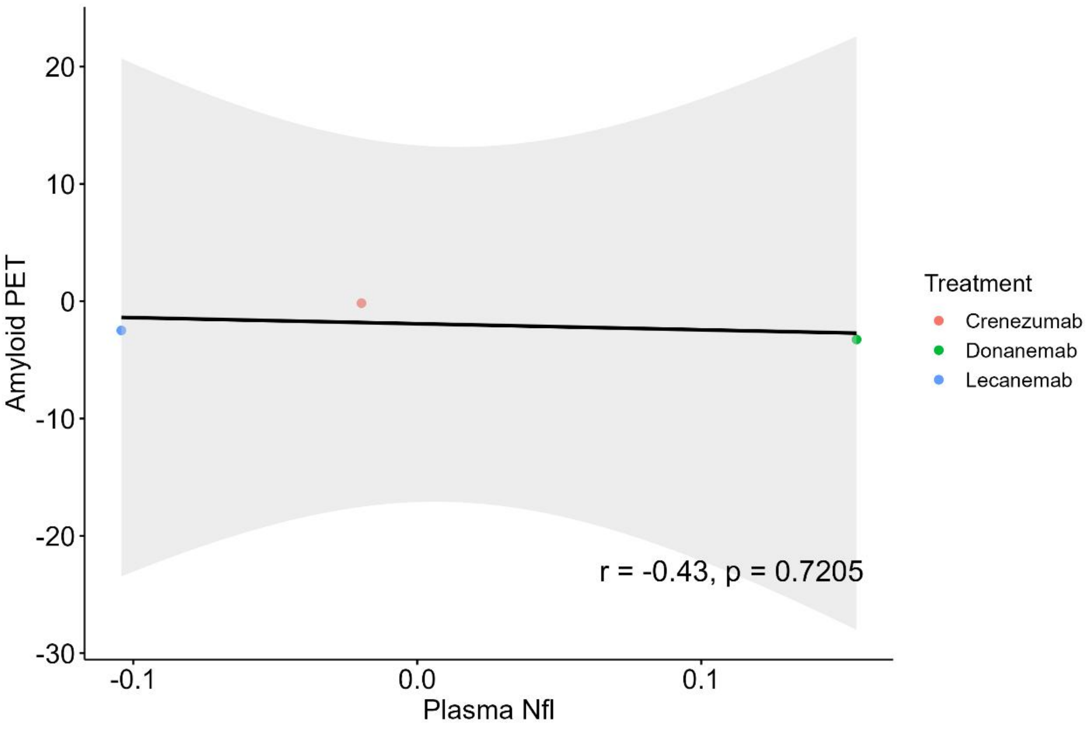
Amyloid PET
20
10
0
-10
-20
-30
-0.1
0.0
0.1
Plasma Nfl
Treatment
Crenezumab
Donanemab
Lecanemab
r = -0.43, p = 0.7205

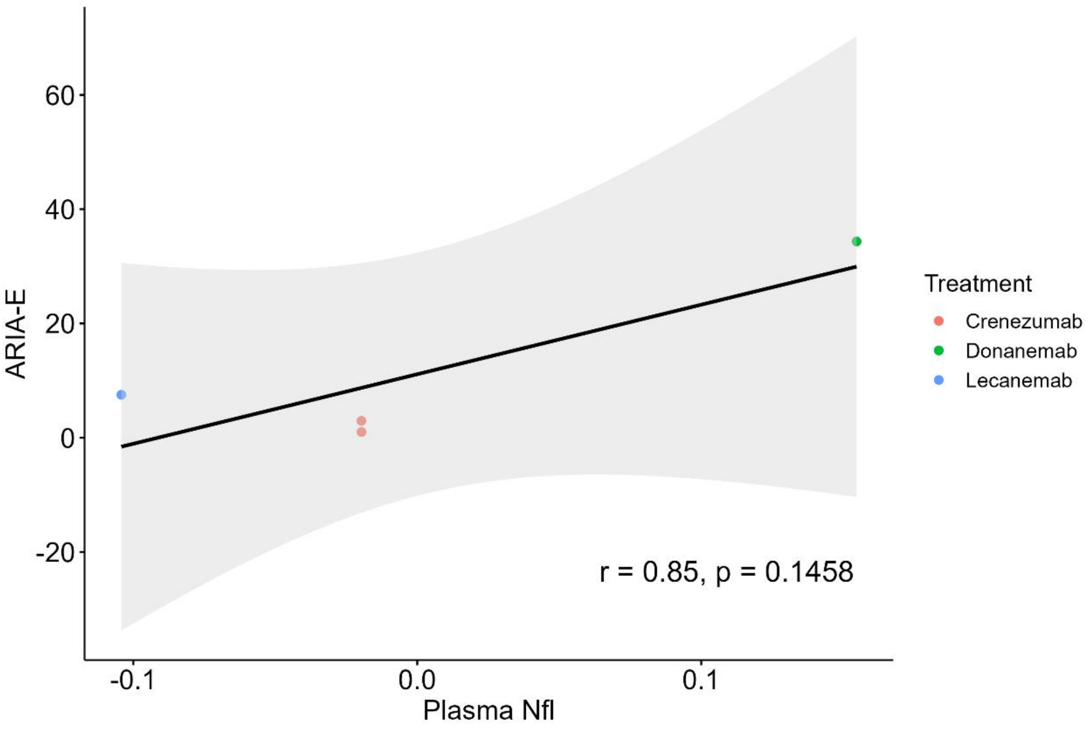
ARIA-E
60
40
20
0
-20
-0.1
0.0
0.1
Plasma Nfl
Treatment
Crenezumab
Donanemab
Lecanemab
r = 0.85, p = 0.1458

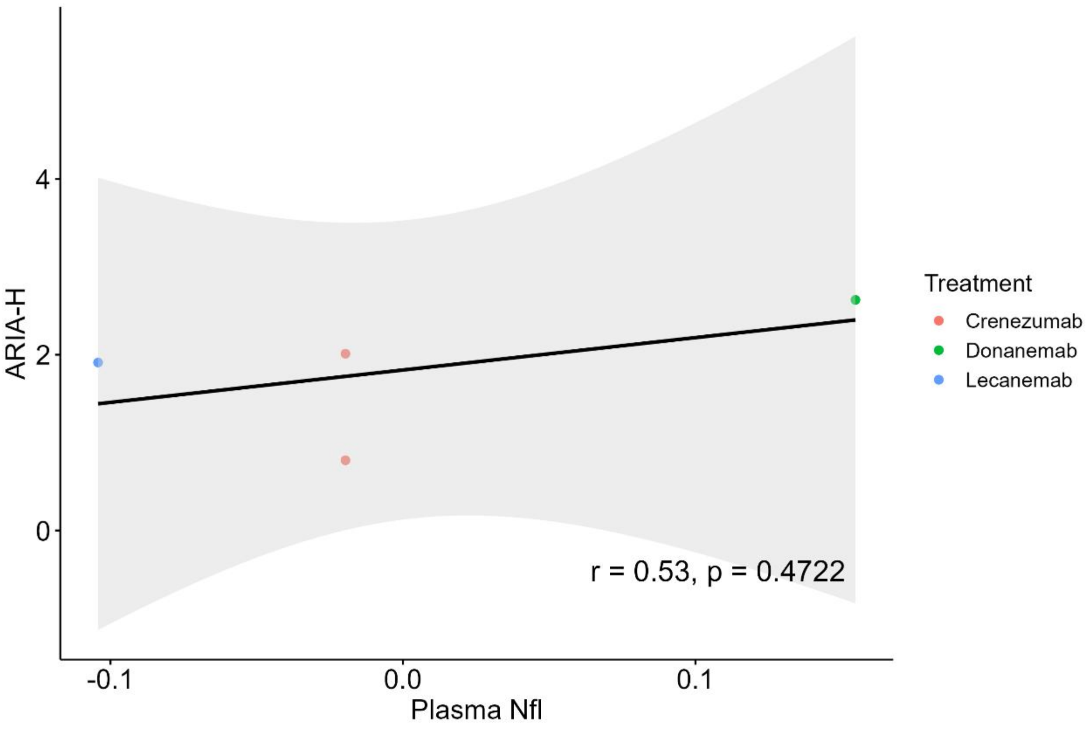
ARIA-H
4
2
0
-0.1
0.0
0.1
Plasma Nfl
r = 0.53, p = 0.4722
Treatment
Crenezumab
Donanemab
Lecanemab

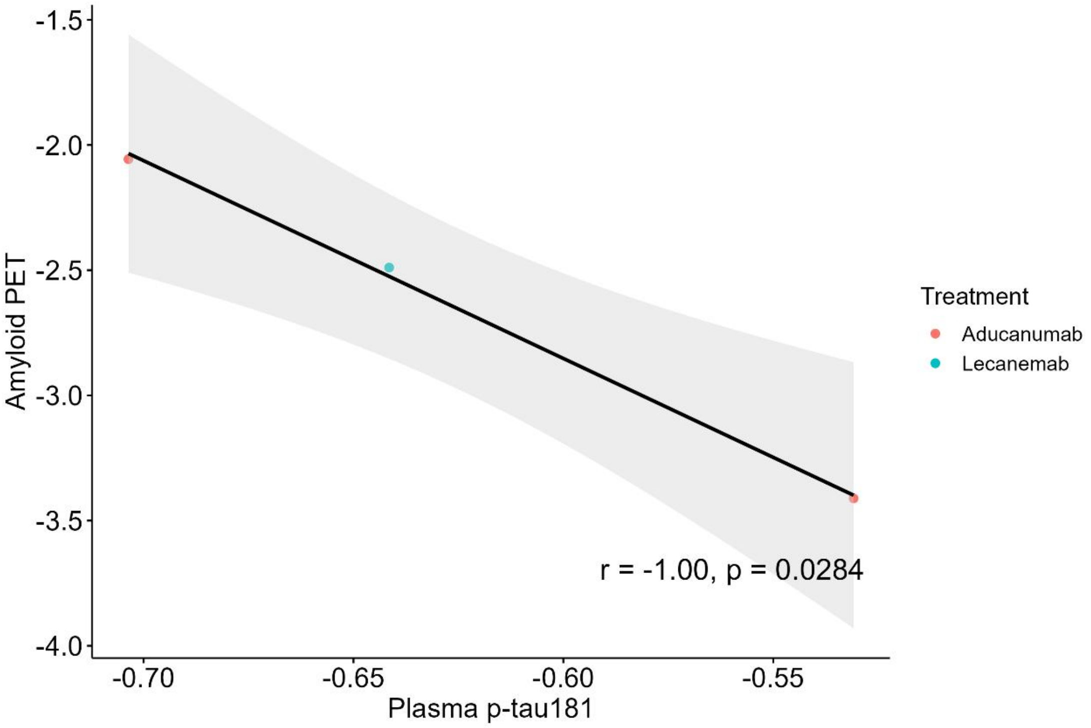
Amyloid PET
-1.5
-2.0
-2.5
-3.0
-3.5
-4.0
-0.70
-0.65
-0.60
-0.55
Plasma p-tau181
r = -1.00, p = 0.0284
Treatment
Aducanumab
Lecanemab

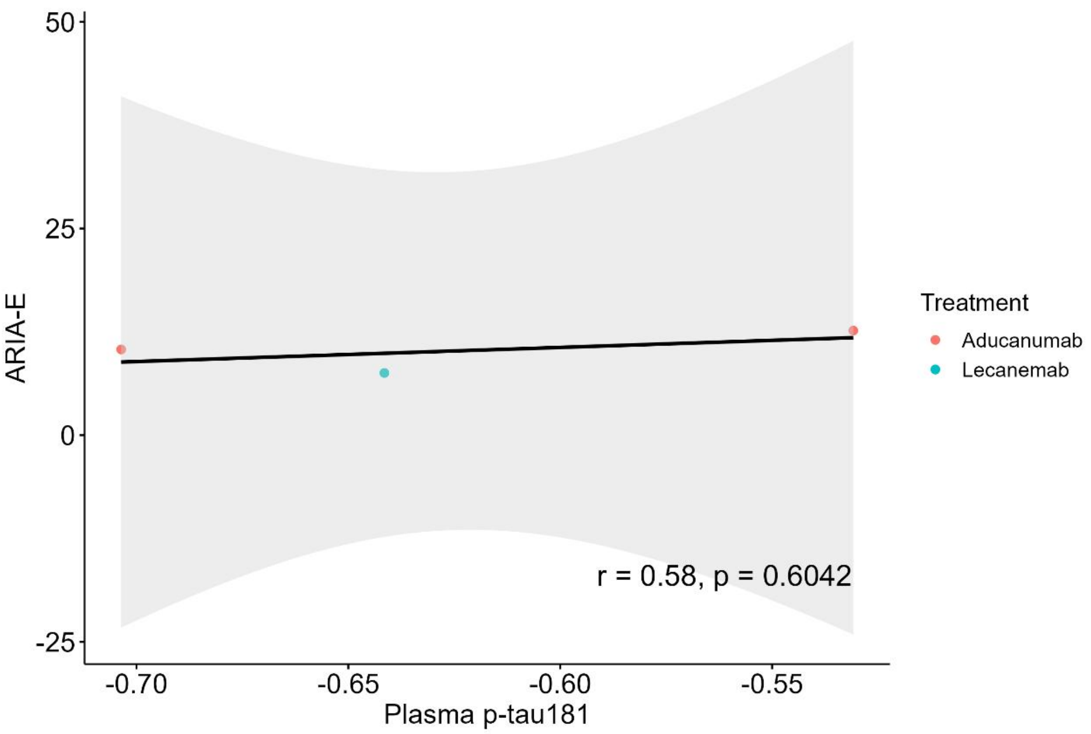
50
25
0
-25
ARIA-E
-0.70
-0.65
-0.60
-0.55
Plasma p-tau181
r = 0.58, p = 0.6042
Treatment
Aducanumab
Lecanemab

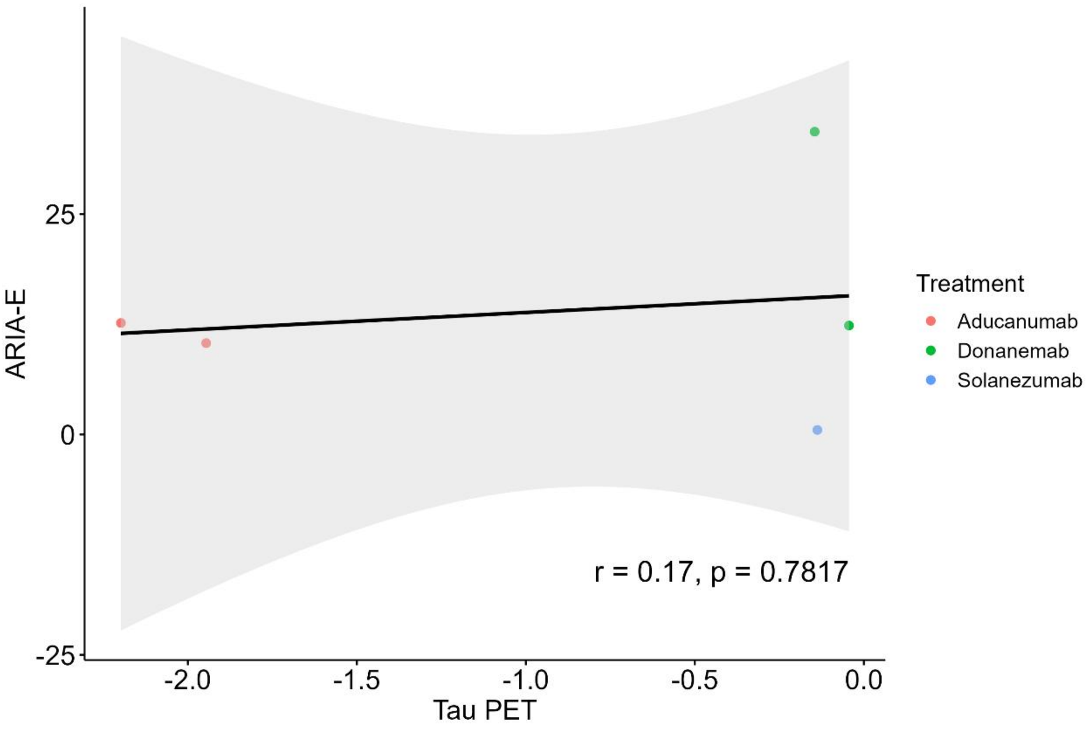
25
0
-25
ARIA-E
-2.0
-1.5
-1.0
-0.5
0.0
Tau PET
r = 0.17, p = 0.7817
Treatment
Aducanumab
Donanemab
Solanezumab

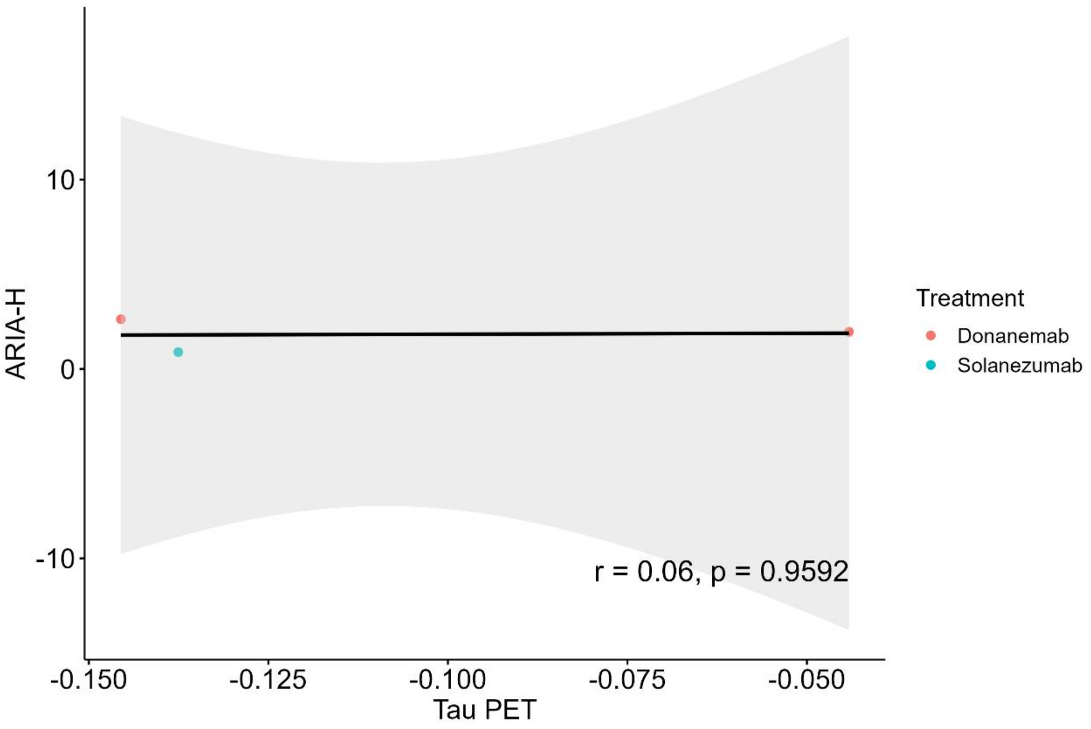
ARIA-H
10
0
-10
-0.150
-0.125
-0.100
-0.075
-0.050
Tau PET
r = 0.06, p = 0.9592
Treatment
Donanemab
Solanezumab

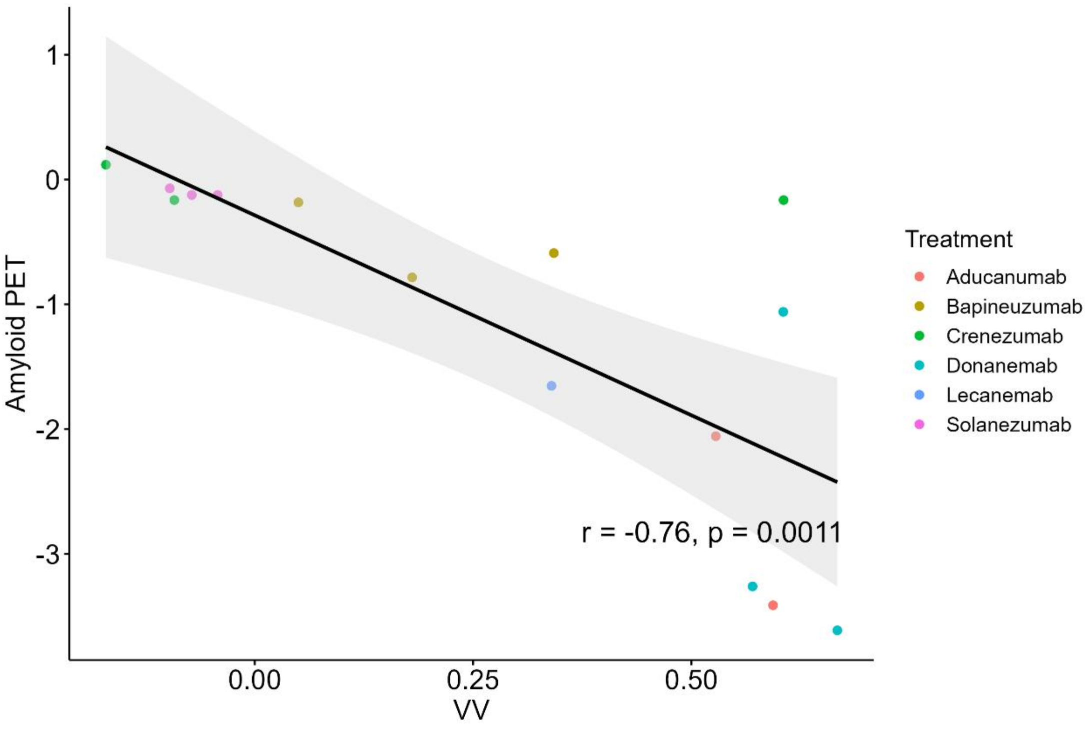
Amyloid PET
1
0
-1
-2
-3
0.00
0.25
0.50
VV
r = -0.76, p = 0.0011
Treatment
Aducanumab
Bapineuzumab
Crenezumab
Donanemab
Lecanemab
Solanezumab

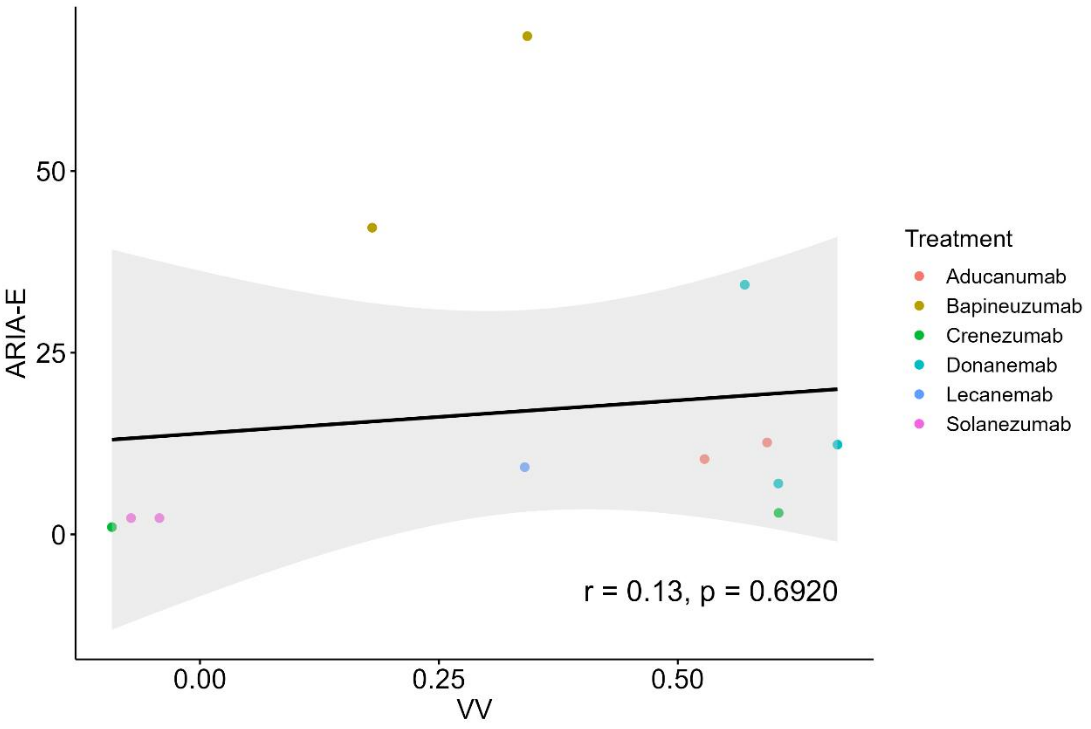
50
25
0
ARIA-E
0.00
0.25
0.50
VV
r = 0.13, p = 0.6920
Treatment
Aducanumab
Bapineuzumab
Crenezumab
Donanemab
Lecanemab
Solanezumab

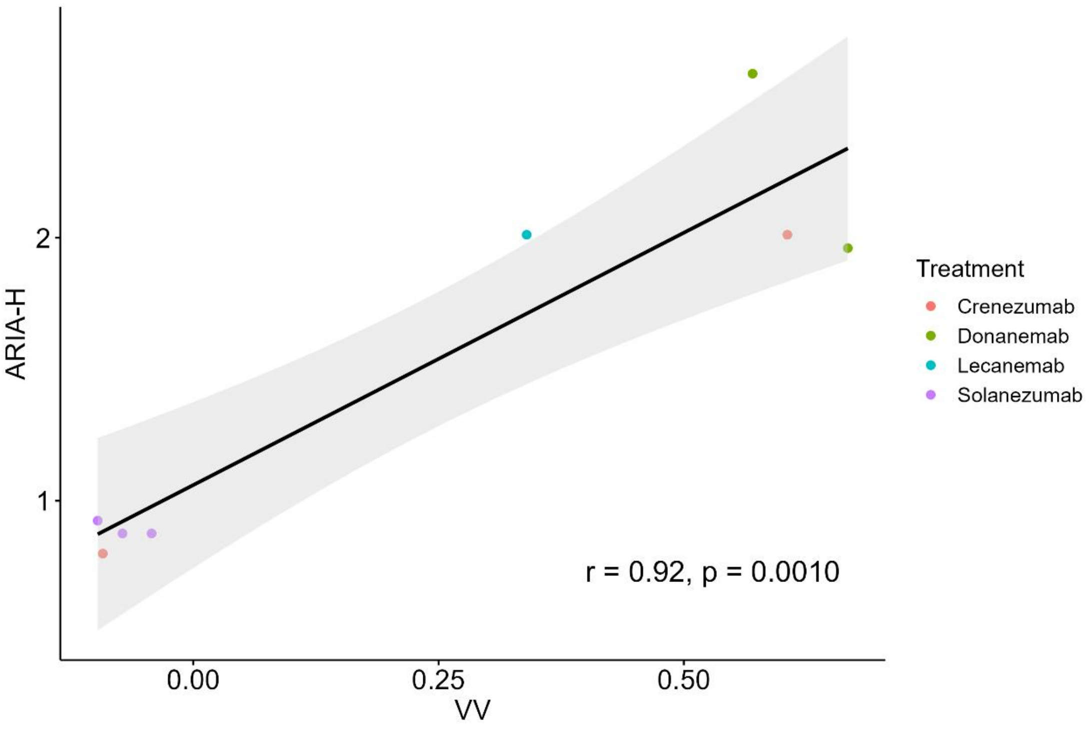
2
1
ARIA-H
0.00
0.25
0.50
VV
r = 0.92, p = 0.0010
Treatment
Crenezumab
Donanemab
Lecanemab
Solanezumab

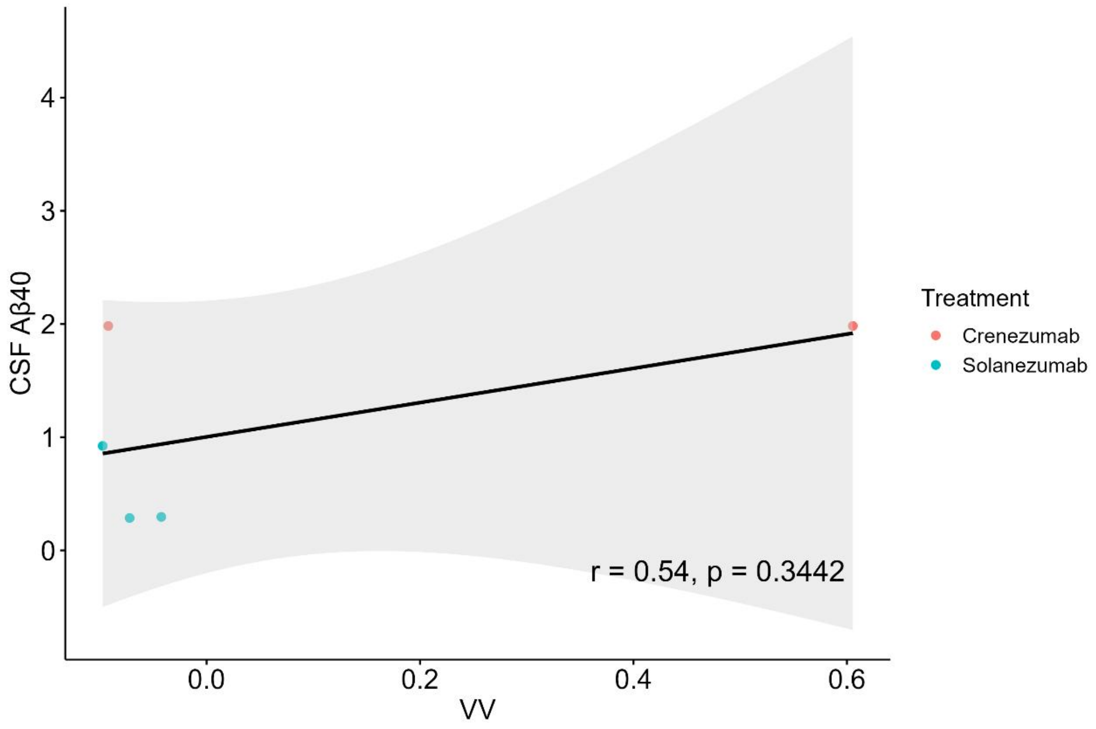
CSF Aβ40
VV
r = 0.54, p = 0.3442
Treatment
Crenezumab
Solanezumab
4
3
2
1
0
0.0
0.2
0.4
0.6

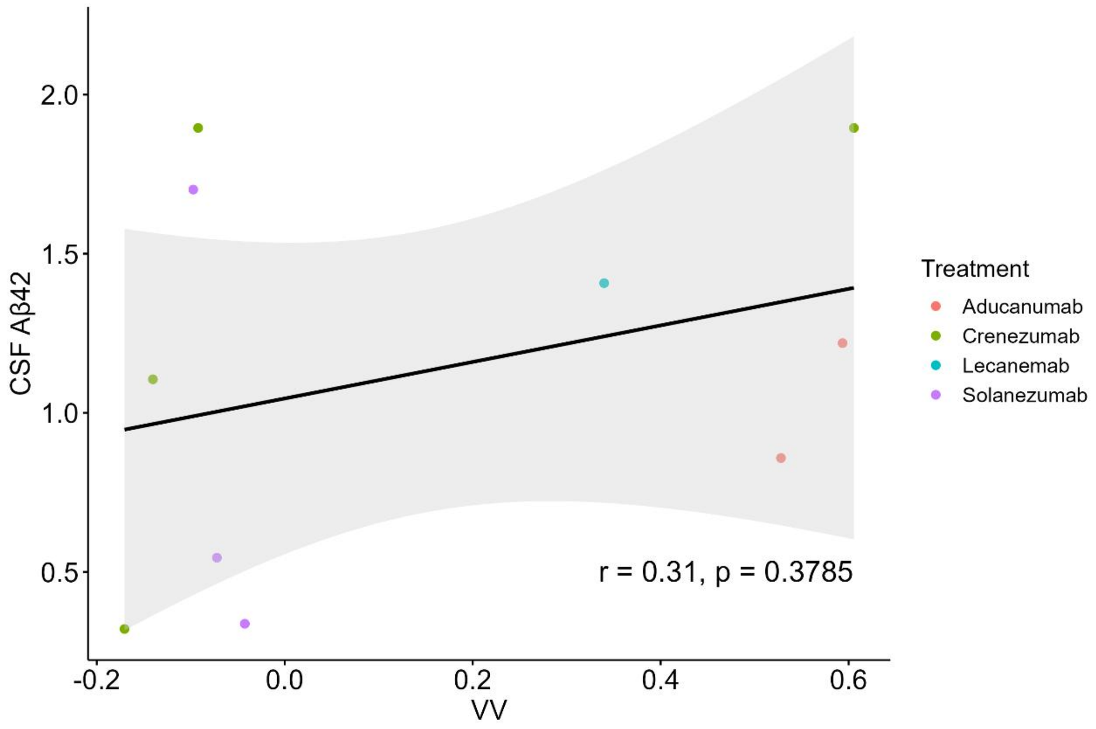
CSF Aβ42
VV
r = 0.31, p = 0.3785
Treatment
Aducanumab
Crenezumab
Lecanemab
Solanezumab
2.0
1.5
1.0
0.5
-0.2
0.0
0.2
0.4
0.6

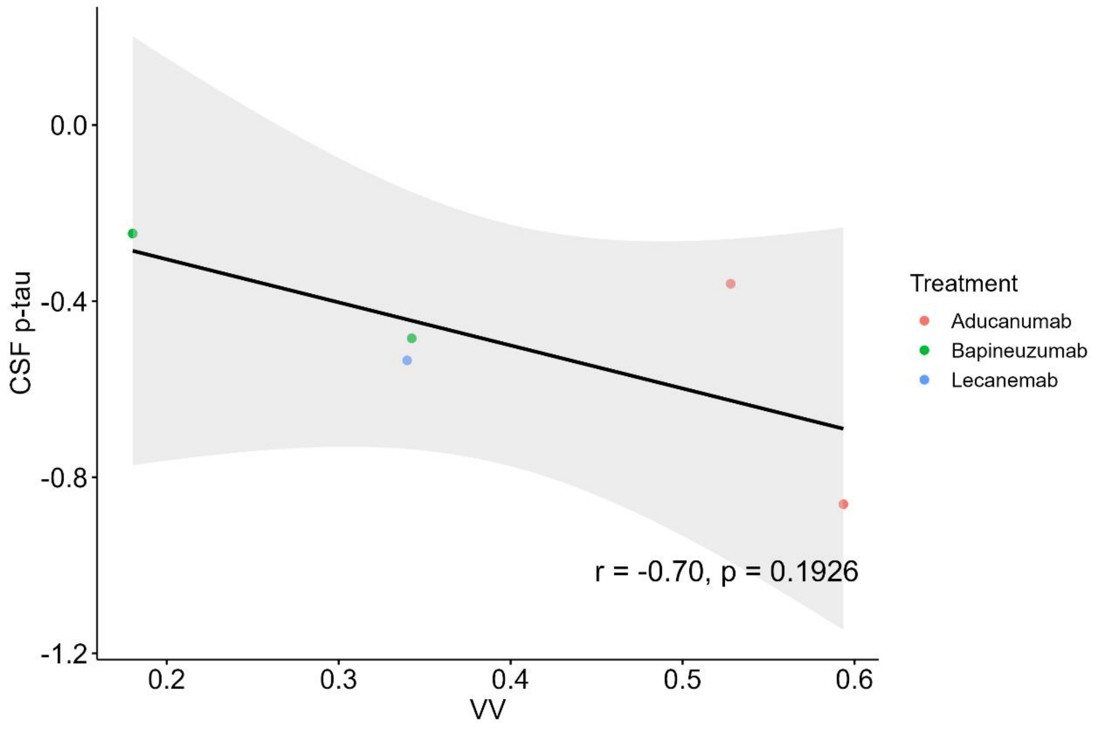

CSF p-tau
0.0
-0.4
-0.8
-1.2
0.2
0.3
0.4
0.5
0.6
VV
r = -0.70, p = 0.1926
Treatment
Aducanumab
Bapineuzumab
Lecanemab

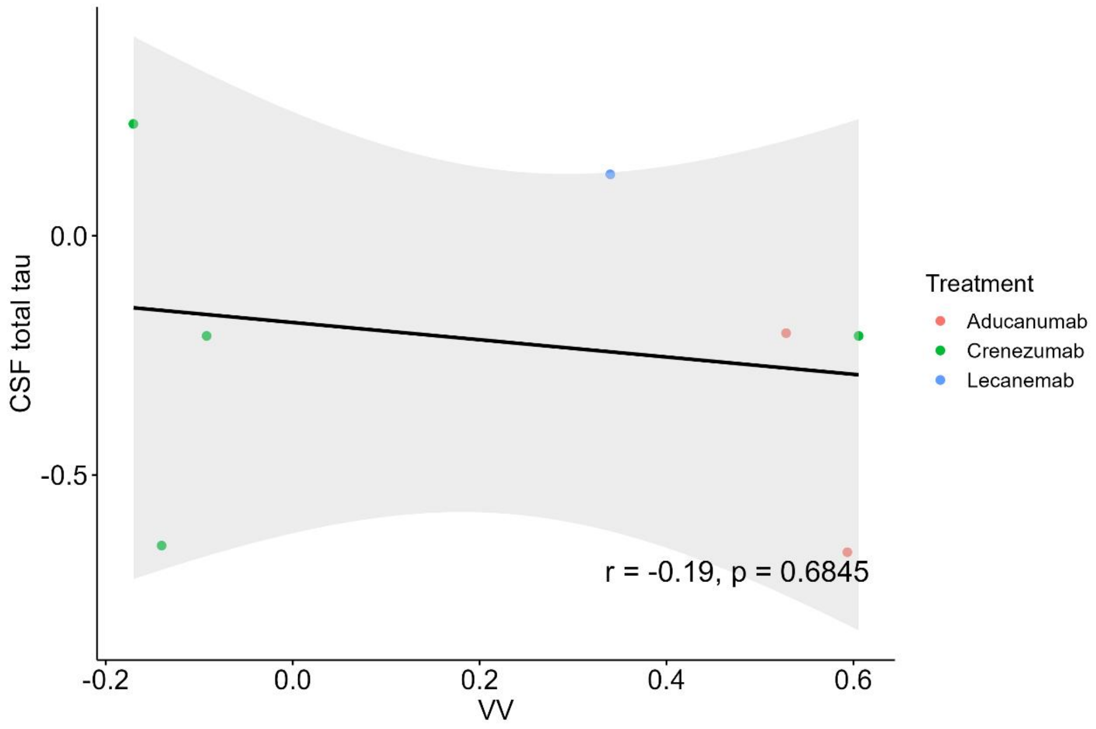

CSF total tau
0.0
-0.5
-0.2
0.0
0.2
0.4
0.6
VV
r = -0.19, p = 0.6845
Treatment
Aducanumab
Crenezumab
Lecanemab

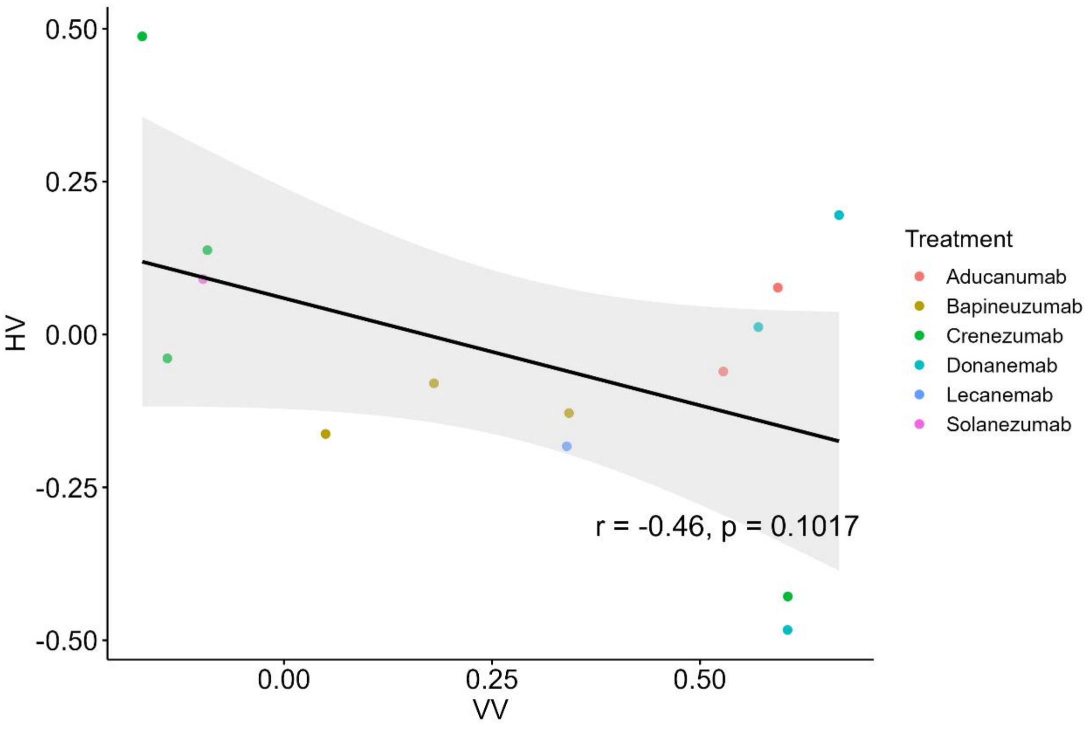

0.50
0.25
0.00
-0.25
-0.50
HV
0.00
0.25
0.50
VV
r = -0.46, p = 0.1017
Treatment
Aducanumab
Bapineuzumab
Crenezumab
Donanemab
Lecanemab
Solanezumab

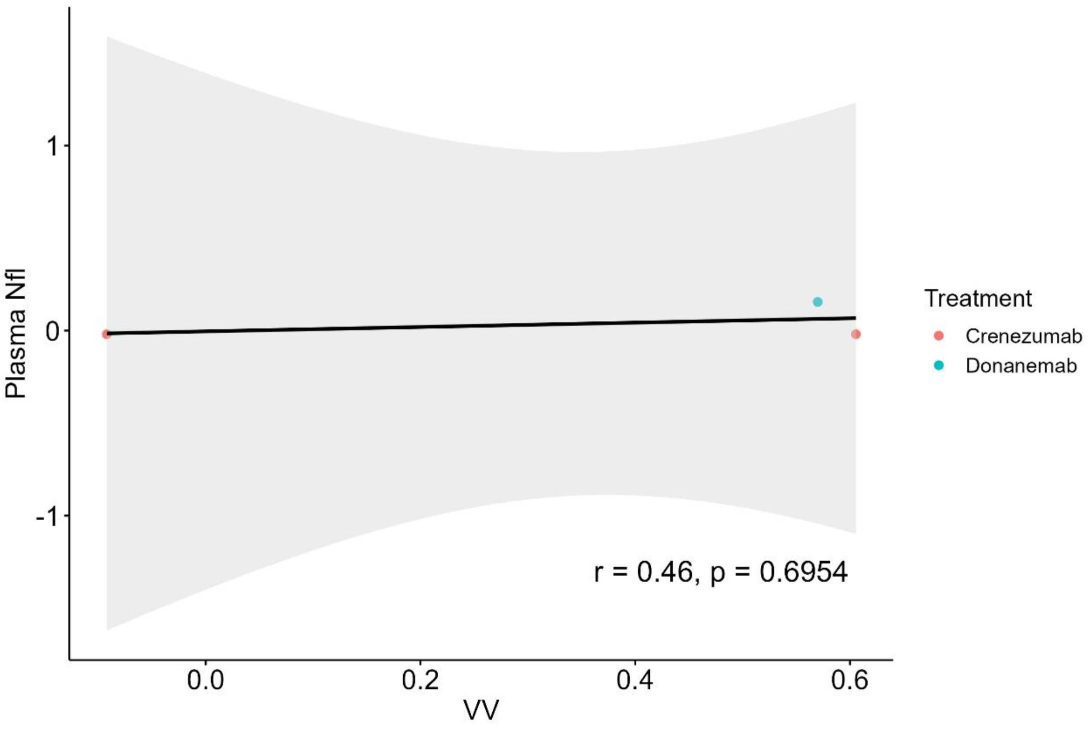

1
0
-1
Plasma Nfl
0.0
0.2
0.4
0.6
VV
r = 0.46, p = 0.6954
Treatment
Crenezumab
Donanemab

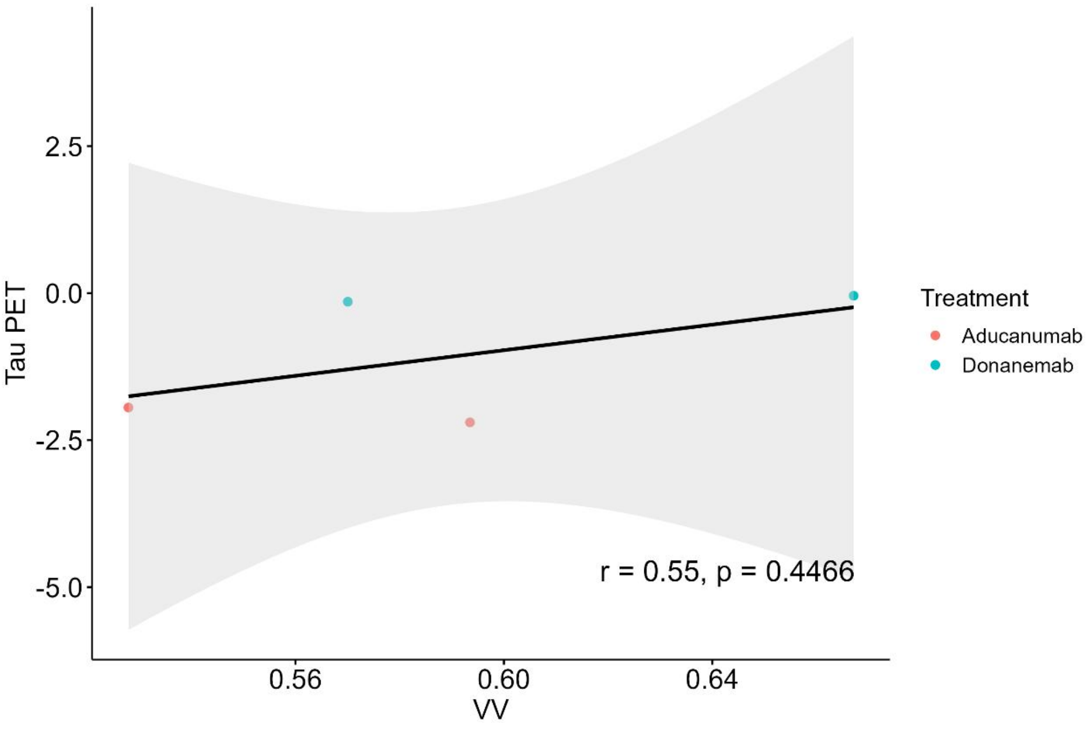
Tau PET
2.5
0.0
-2.5
-5.0
0.56
0.60
0.64
VV
r = 0.55, p = 0.4466
Treatment
Aducanumab
Donanemab

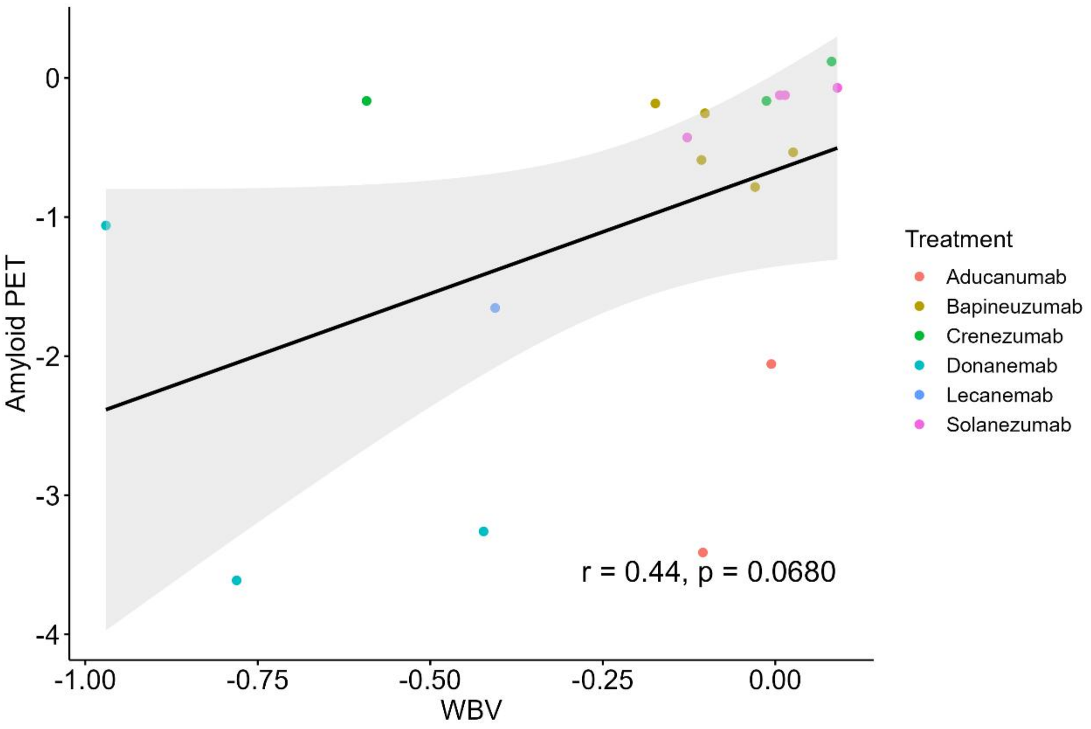
Amyloid PET
0
-1
-2
-3
-4
-1.00
-0.75
-0.50
-0.25
0.00
WBV
r = 0.44, p = 0.0680
Treatment
Aducanumab
Bapineuzumab
Crenezumab
Donanemab
Lecanemab
Solanezumab

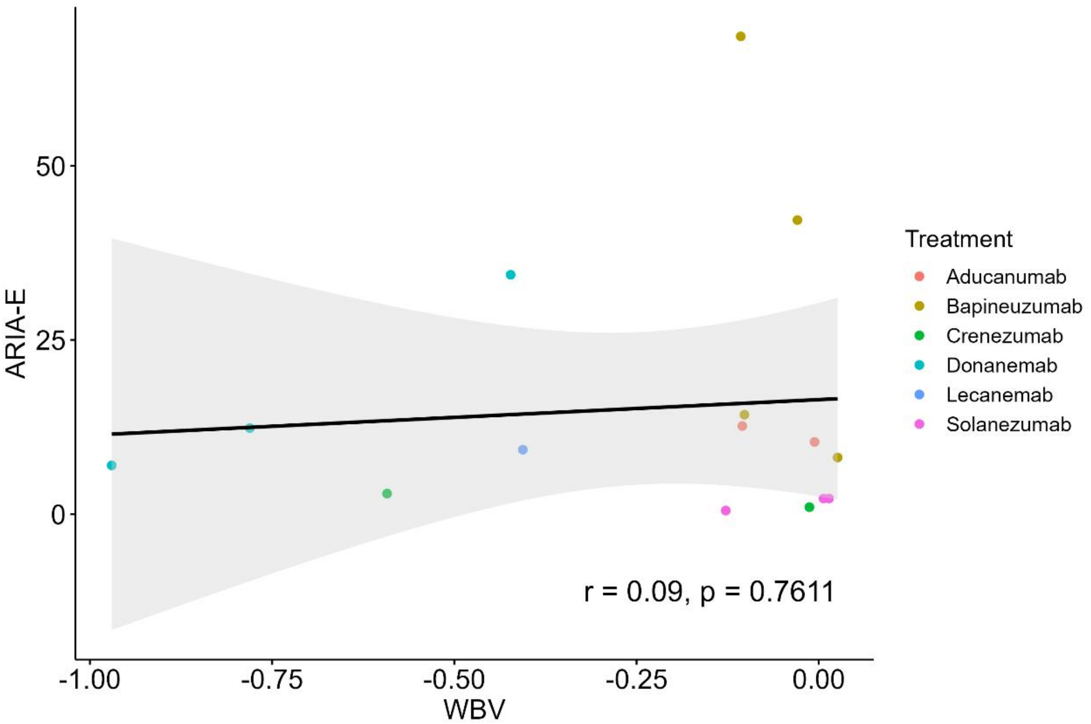

ARIA-E
50
25
0
-1.00
-0.75
-0.50
-0.25
0.00
WBV
r = 0.09, p = 0.7611
Treatment
Aducanumab
Bapineuzumab
Crenezumab
Donanemab
Lecanemab
Solanezumab

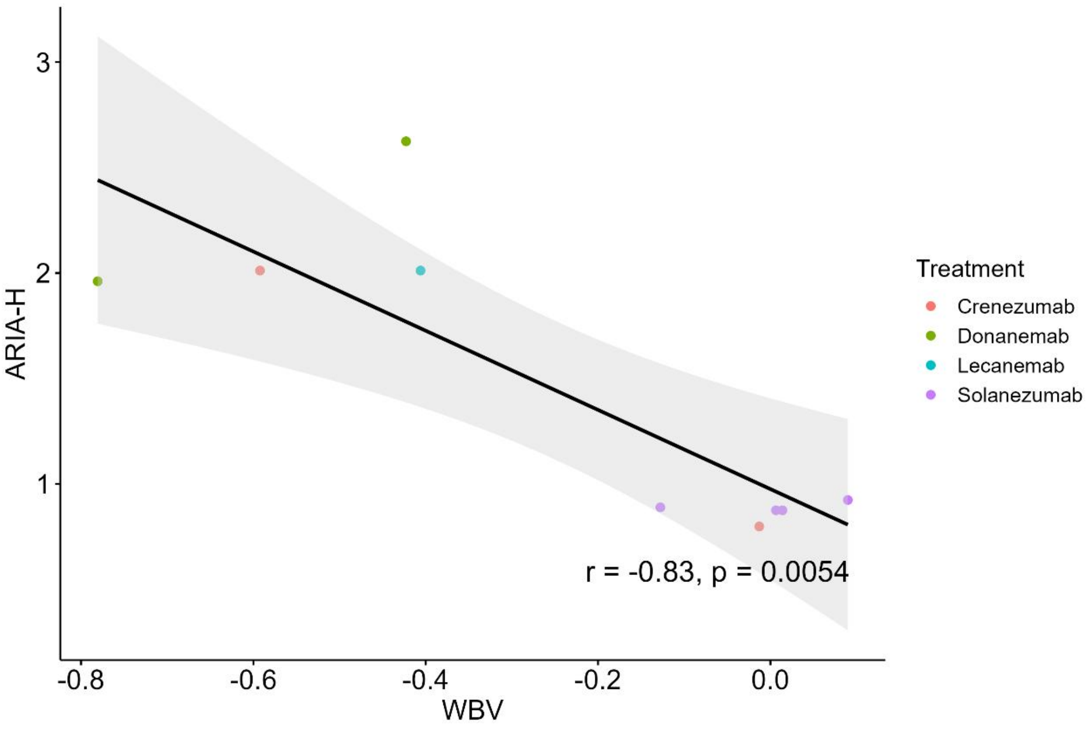

ARIA-H
3
2
1
-0.8
-0.6
-0.4
-0.2
0.0
WBV
r = -0.83, p = 0.0054
Treatment
Crenezumab
Donanemab
Lecanemab
Solanezumab

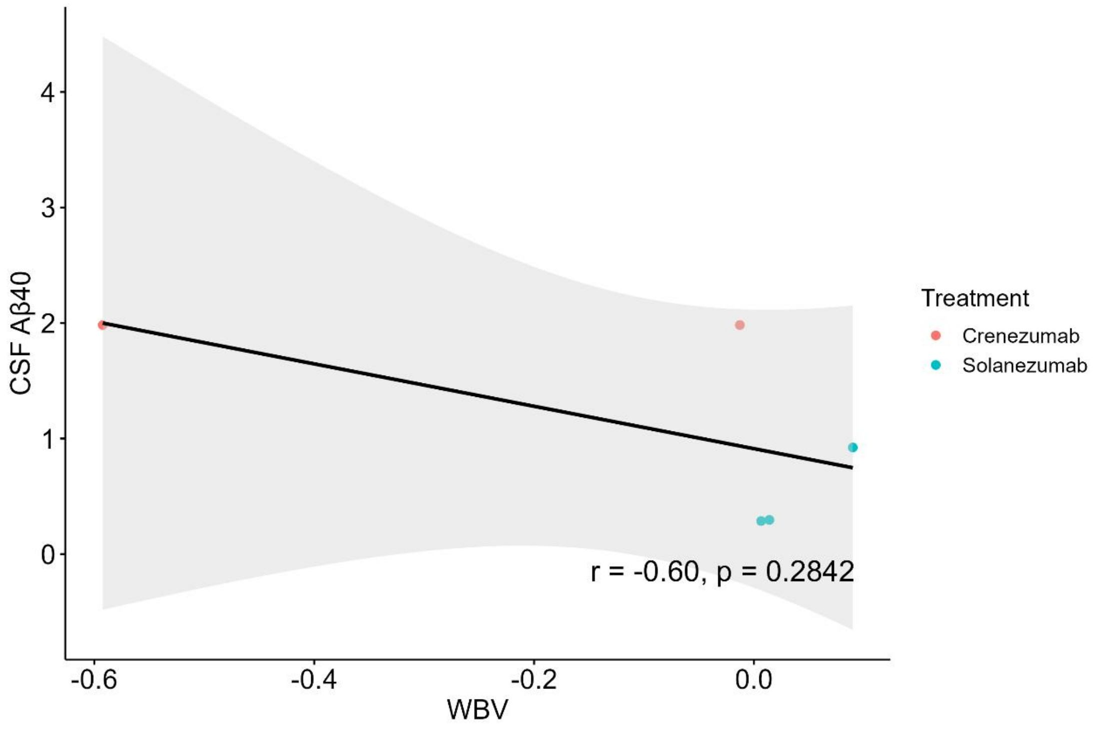

CSF Aβ40
4
3
2
1
0
-0.6
-0.4
-0.2
0.0
WBV
r = -0.60, p = 0.2842
Treatment
Crenezumab
Solanezumab

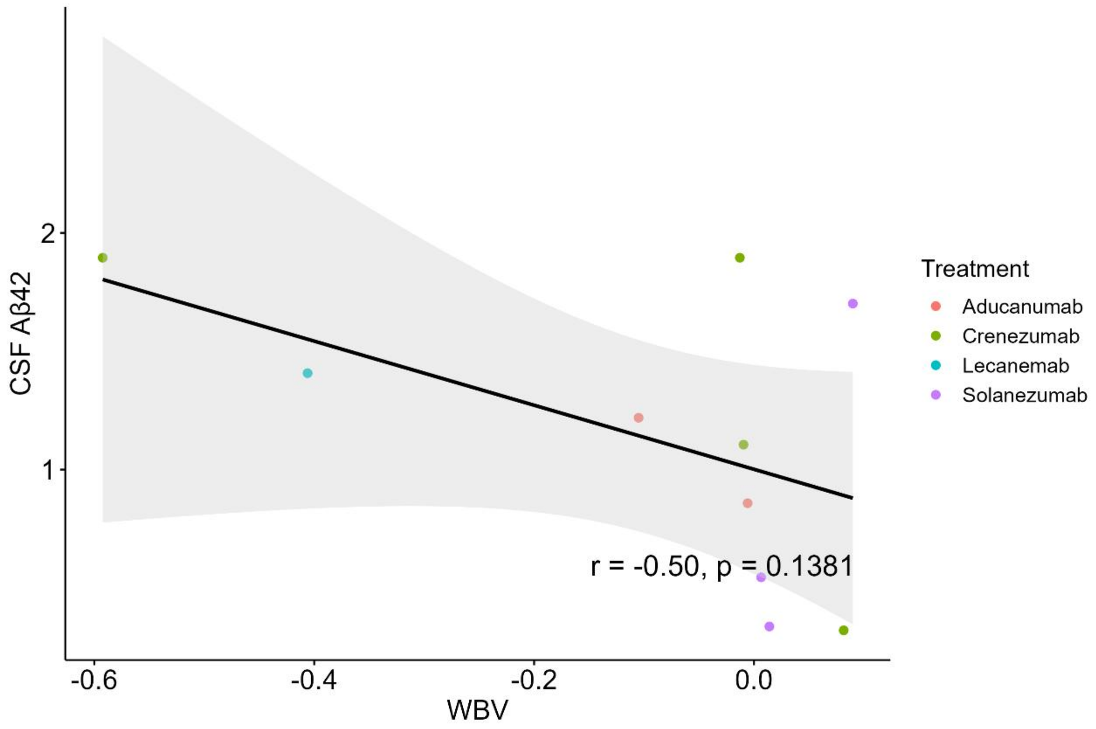

CSF Aβ42
2
1
-0.6
-0.4
-0.2
0.0
WBV
r = -0.50, p = 0.1381
Treatment
Aducanumab
Crenezumab
Lecanemab
Solanezumab

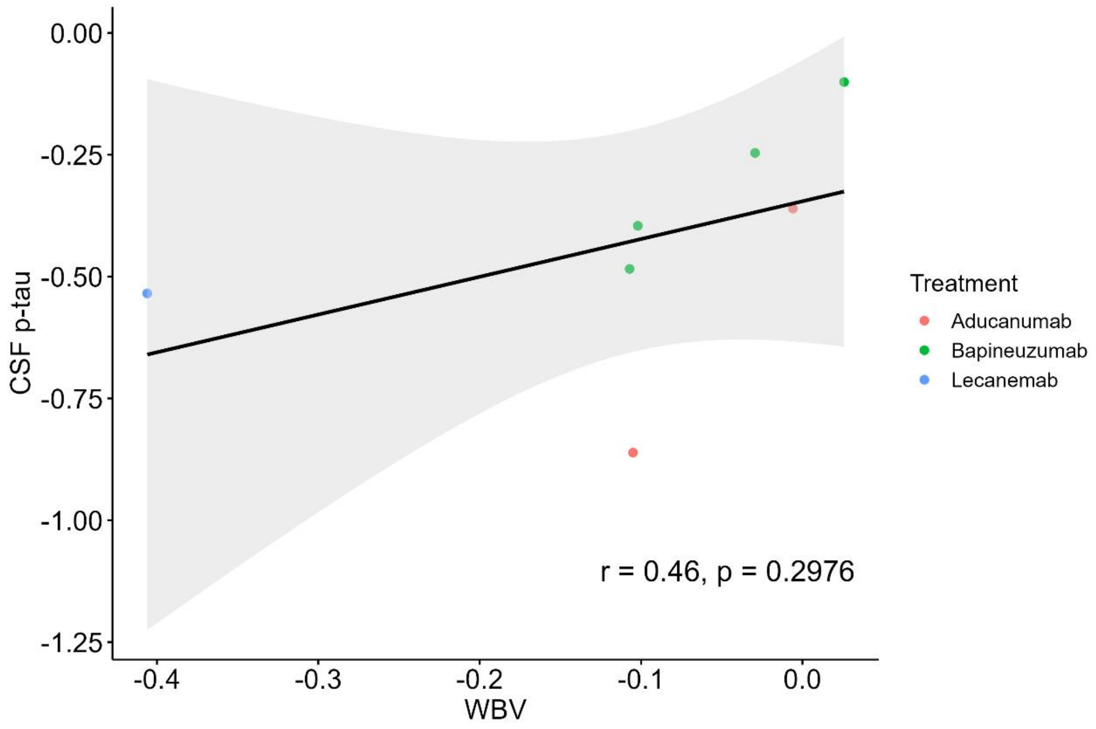
0.00
-0.25
-0.50
-0.75
-1.00
-1.25
CSF p-tau
-0.4
-0.3
-0.2
-0.1
0.0
WBV
r = 0.46, p = 0.2976
Treatment
Aducanumab
Bapineuzumab
Lecanemab

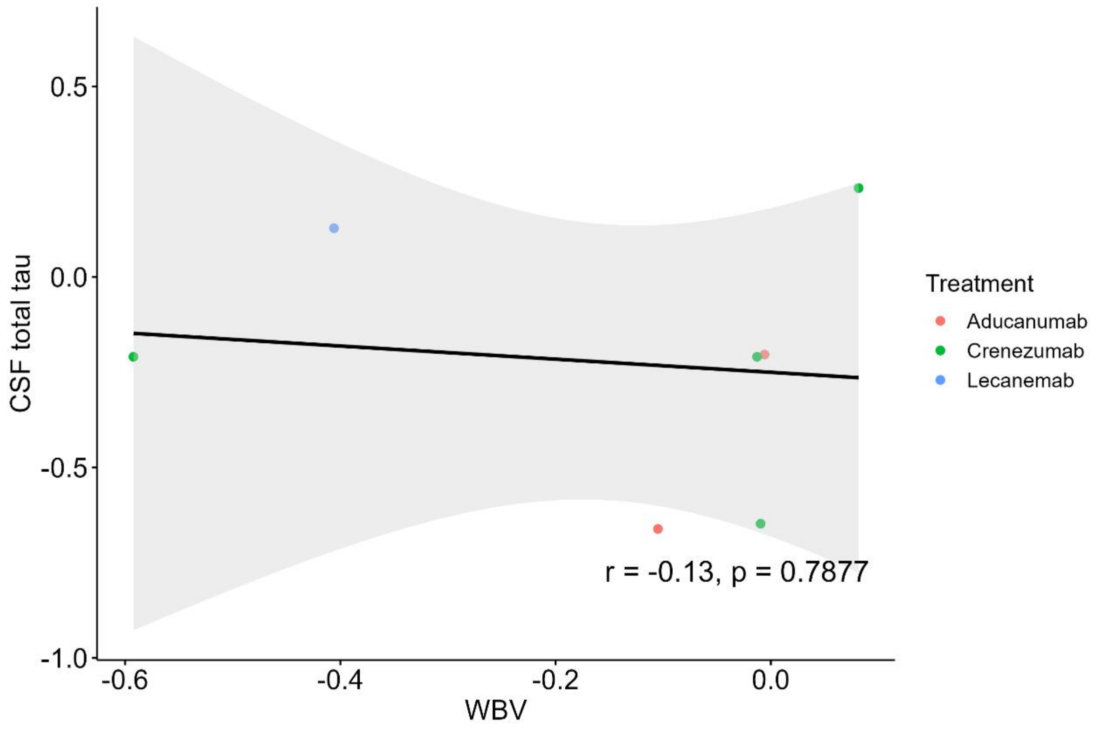
0.5
0.0
-0.5
-1.0
CSF total tau
-0.6
-0.4
-0.2
0.0
WBV
r = -0.13, p = 0.7877
Treatment
Aducanumab
Crenezumab
Lecanemab

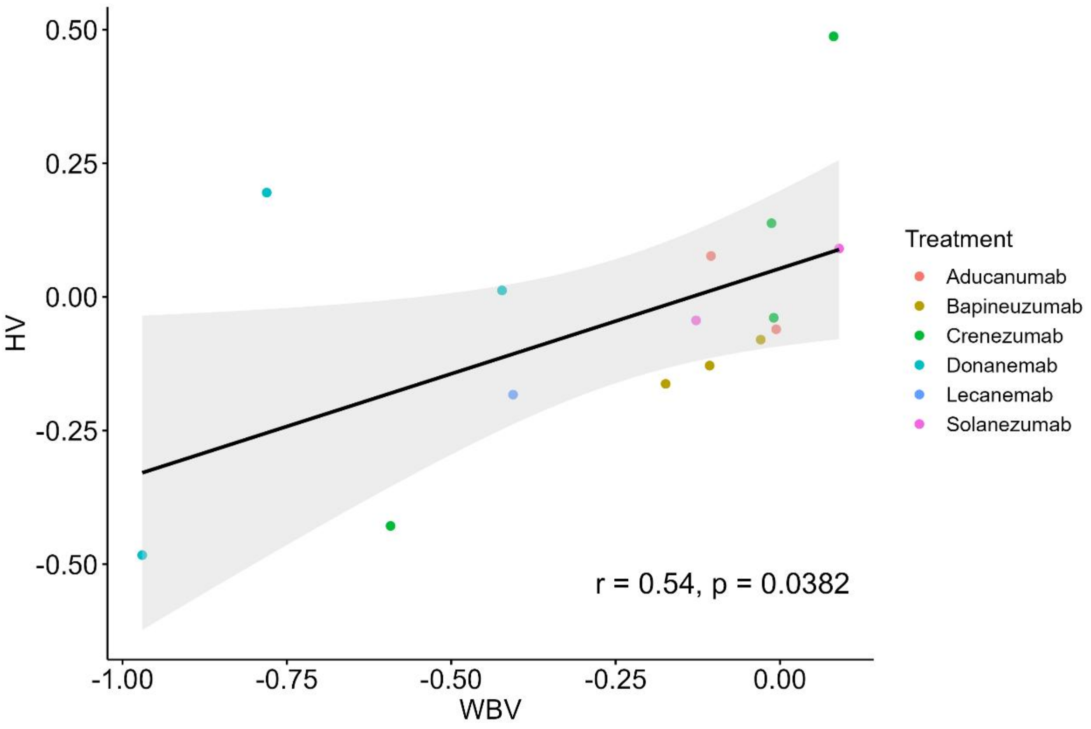
0.50
0.25
0.00
-0.25
-0.50
HV
-1.00
-0.75
-0.50
-0.25
0.00
WBV
r = 0.54, p = 0.0382
Treatment
Aducanumab
Bapineuzumab
Crenezumab
Donanemab
Lecanemab
Solanezumab

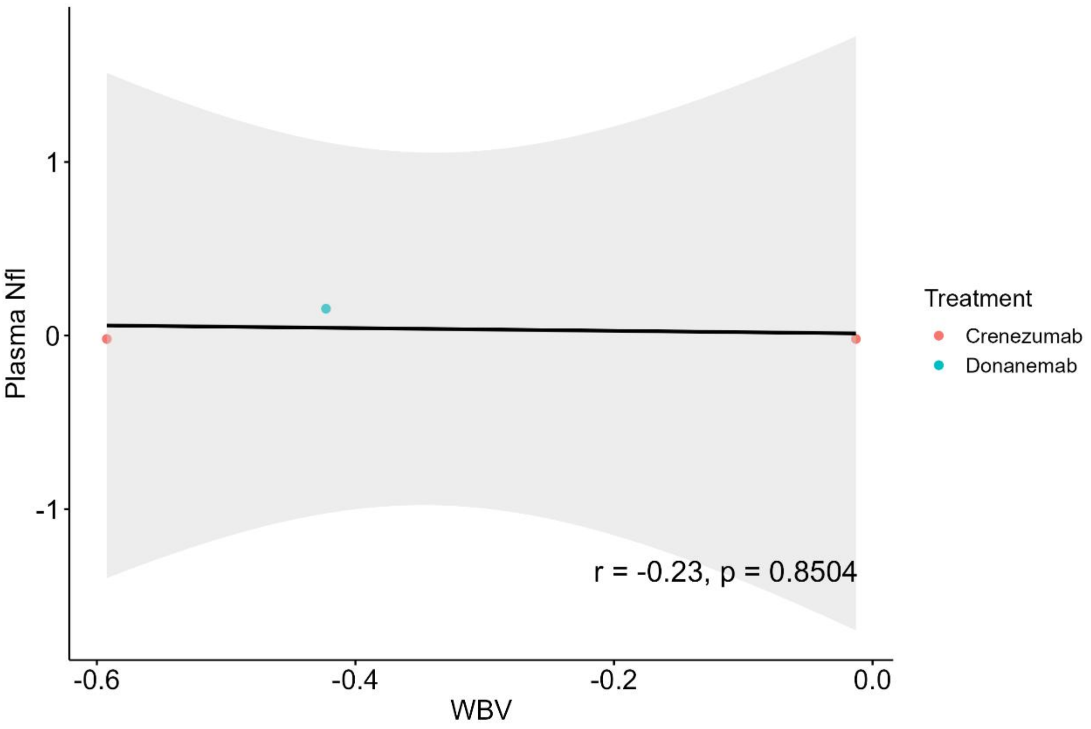
1
0
-1
Plasma Nfl
-0.6
-0.4
-0.2
0.0
WBV
r = -0.23, p = 0.8504
Treatment
Crenezumab
Donanemab

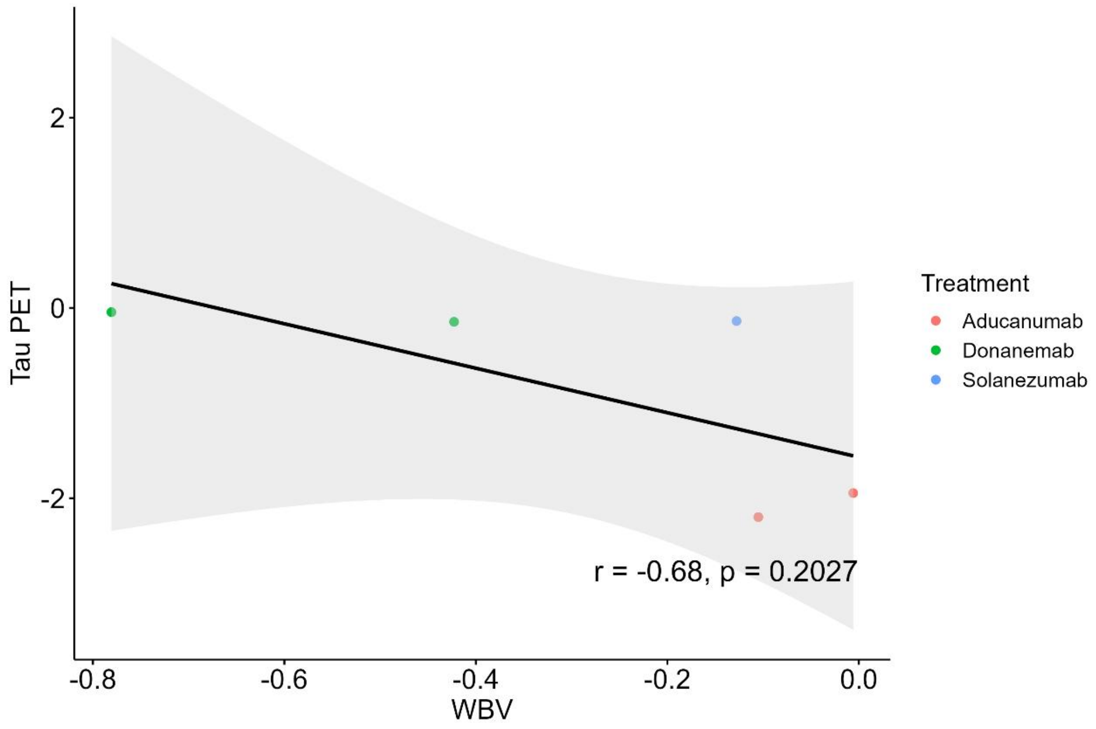

Tau PET
2
0
-2
-0.8
-0.6
-0.4
-0.2
0.0
WBV
r = -0.68, p = 0.2027
Treatment
Aducanumab
Donanemab
Solanezumab

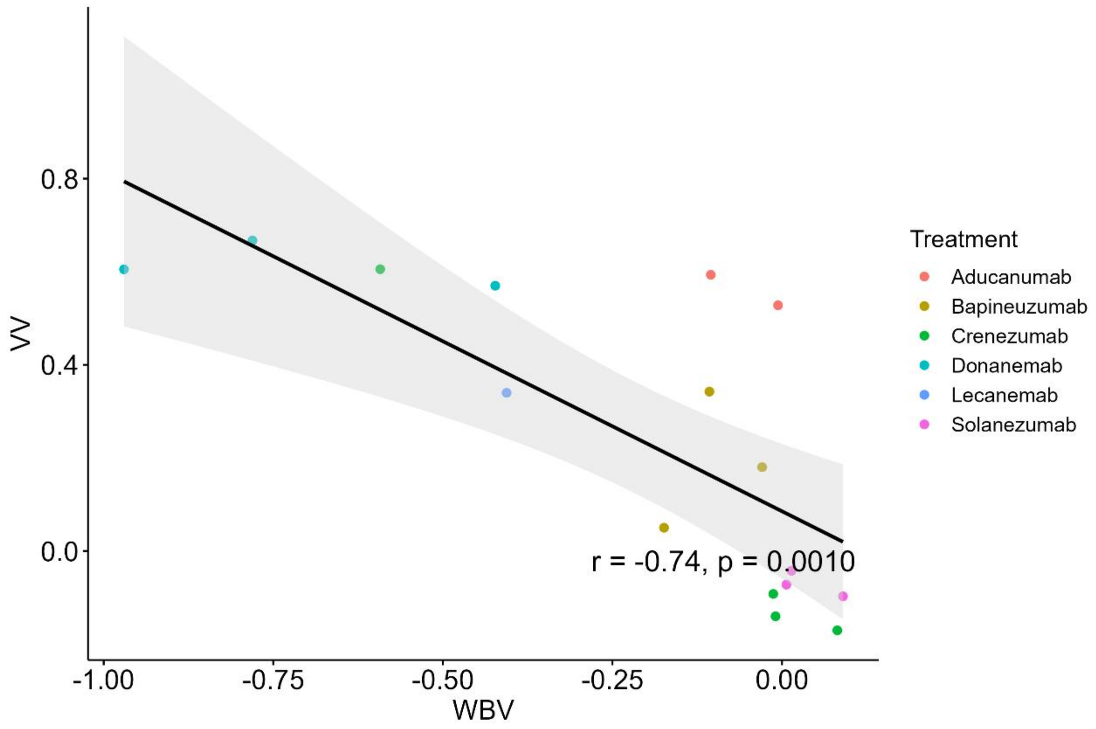

VV
0.8
0.4
0.0
-1.00
-0.75
-0.50
-0.25
0.00
WBV
r = -0.74, p = 0.0010
Treatment
Aducanumab
Bapineuzumab
Crenezumab
Donanemab
Lecanemab
Solanezumab

**Appendix 11: Interrelationships among changes in Aβ deposition, variations in AD biomarkers, alterations in cognitive performance, and the risks of ARIA-E and ARIA-H across N-terminal targeted mAbs**

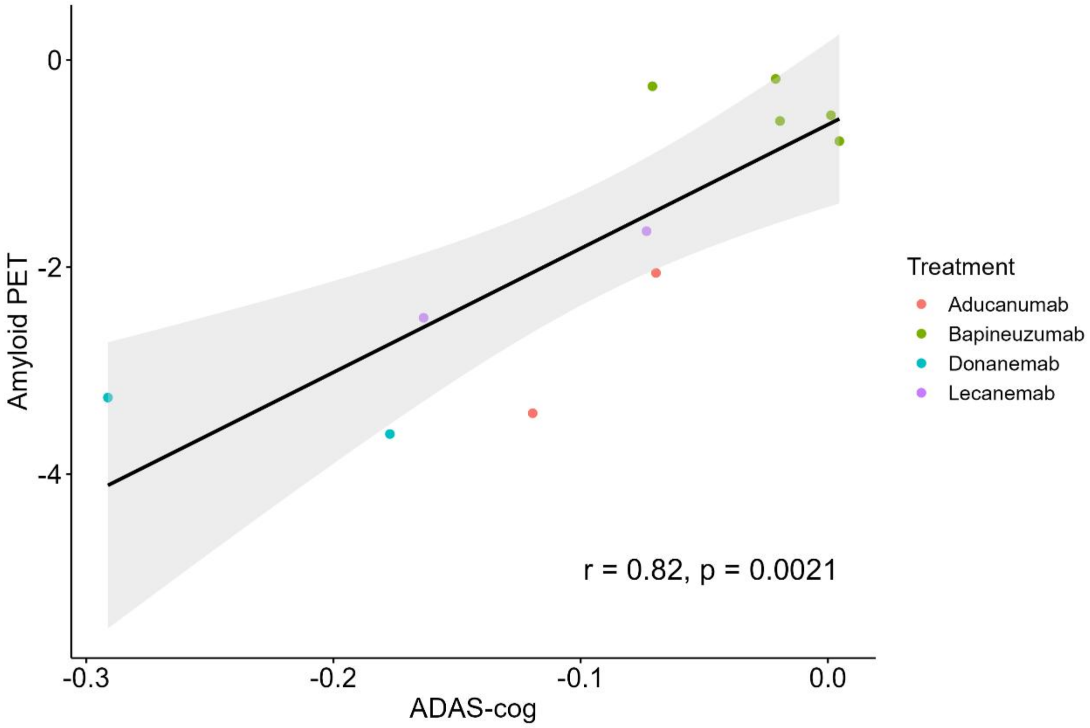

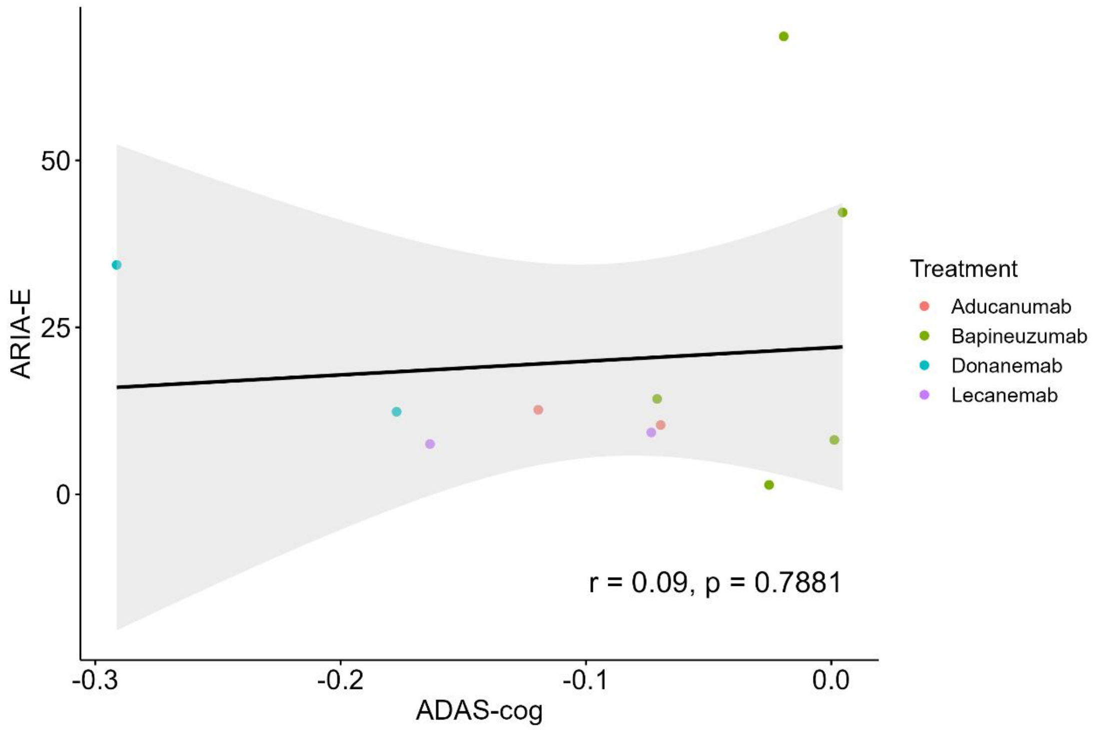
ARIA-E
50
25
0
-0.3
-0.2
-0.1
0.0
ADAS-cog
r = 0.09, p = 0.7881
Treatment
Aducanumab
Bapineuzumab
Donanemab
Lecanemab

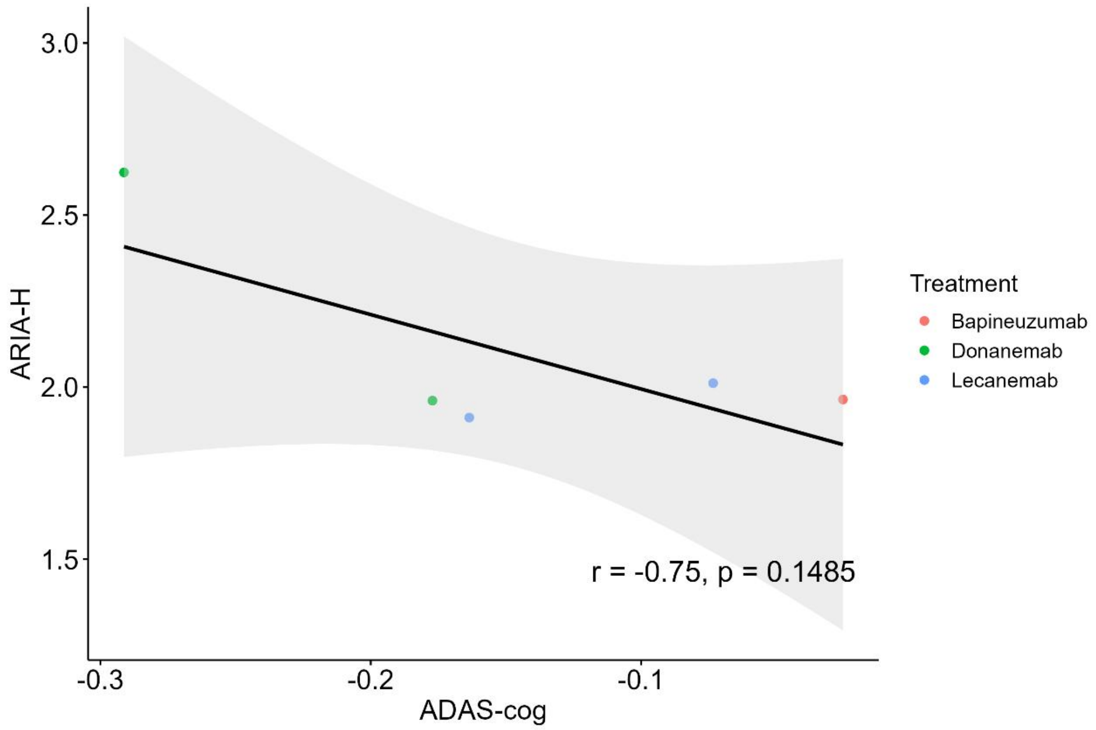
ARIA-H
3.0
2.5
2.0
1.5
-0.3
-0.2
-0.1
ADAS-cog
r = -0.75, p = 0.1485
Treatment
Bapineuzumab
Donanemab
Lecanemab

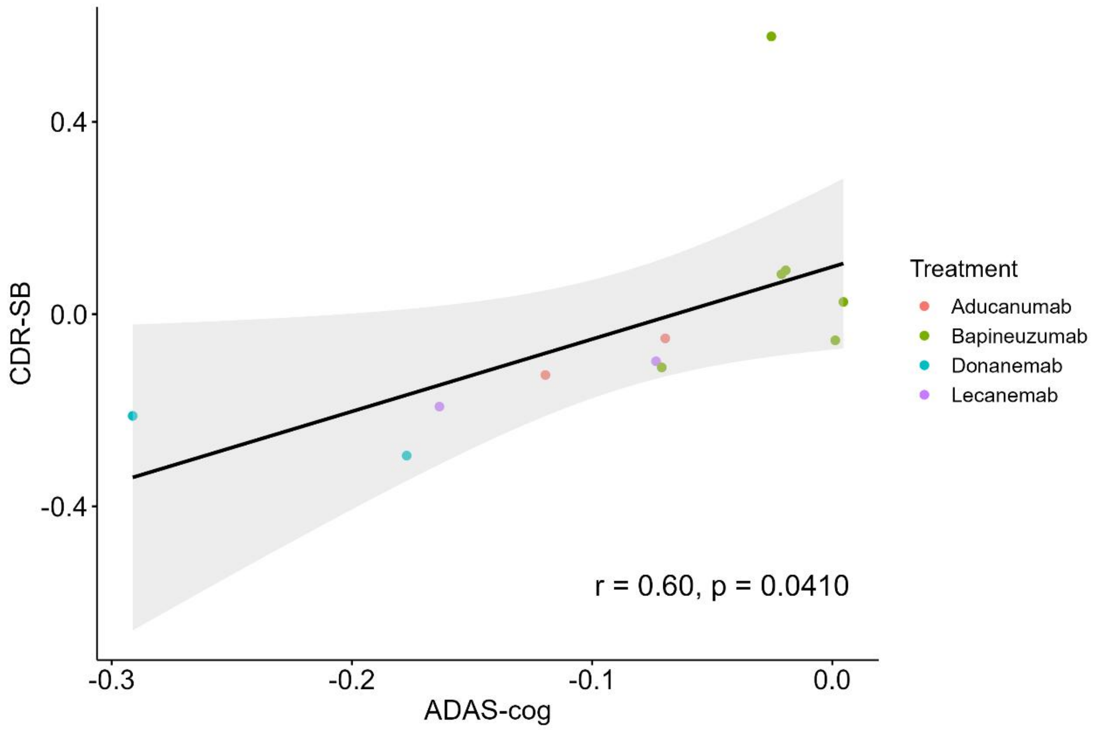
CDR-SB
ADAS-cog
0.4
0.0
-0.4
-0.3
-0.2
-0.1
0.0
r = 0.60, p = 0.0410
Treatment
Aducanumab
Bapineuzumab
Donanemab
Lecanemab

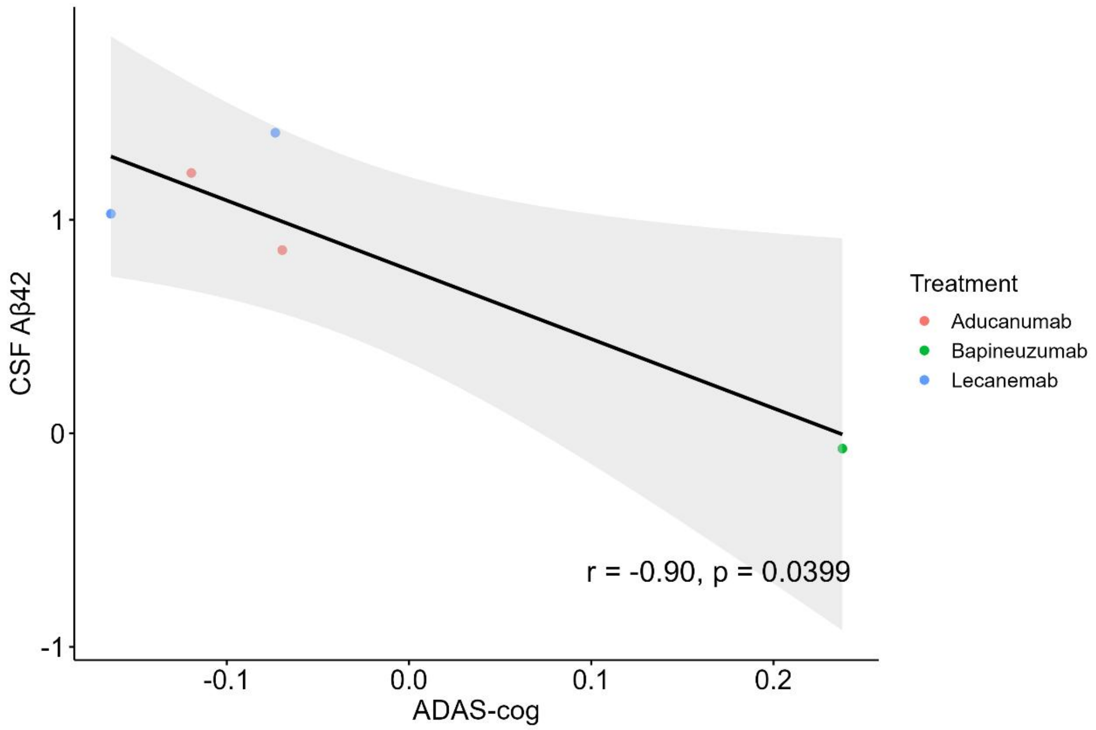
CSF Aβ42
ADAS-cog
1
0
-1
-0.1
0.0
0.1
0.2
r = -0.90, p = 0.0399
Treatment
Aducanumab
Bapineuzumab
Lecanemab

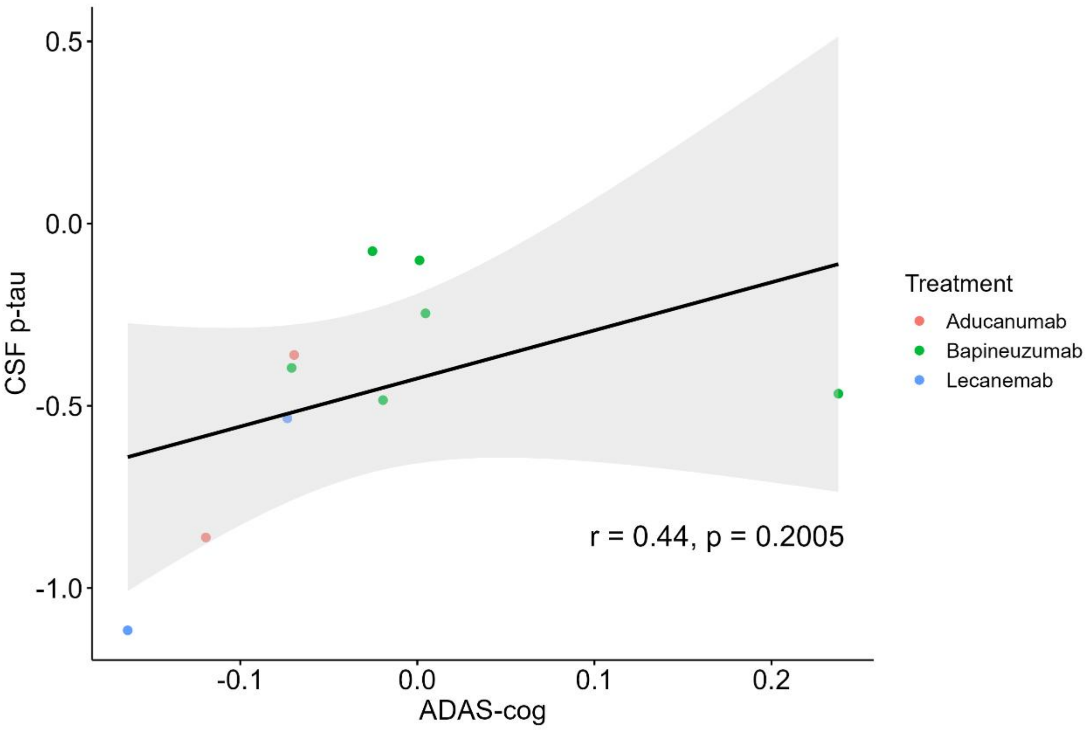
CSF p-tau
ADAS-cog
Treatment
Aducanumab
Bapineuzumab
Lecanemab
r = 0.44, p = 0.2005
0.5
0.0
-0.5
-1.0
-0.1
0.0
0.1
0.2

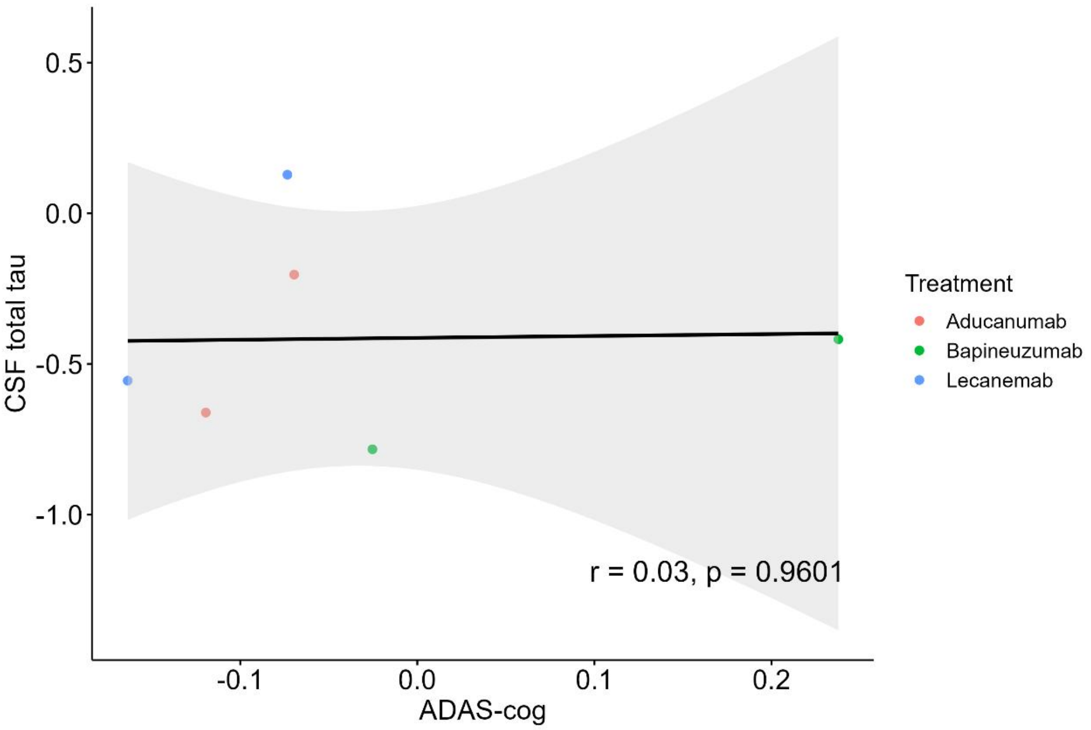
CSF total tau
ADAS-cog
Treatment
Aducanumab
Bapineuzumab
Lecanemab
r = 0.03, p = 0.9601
0.5
0.0
-0.5
-1.0
-0.1
0.0
0.1
0.2

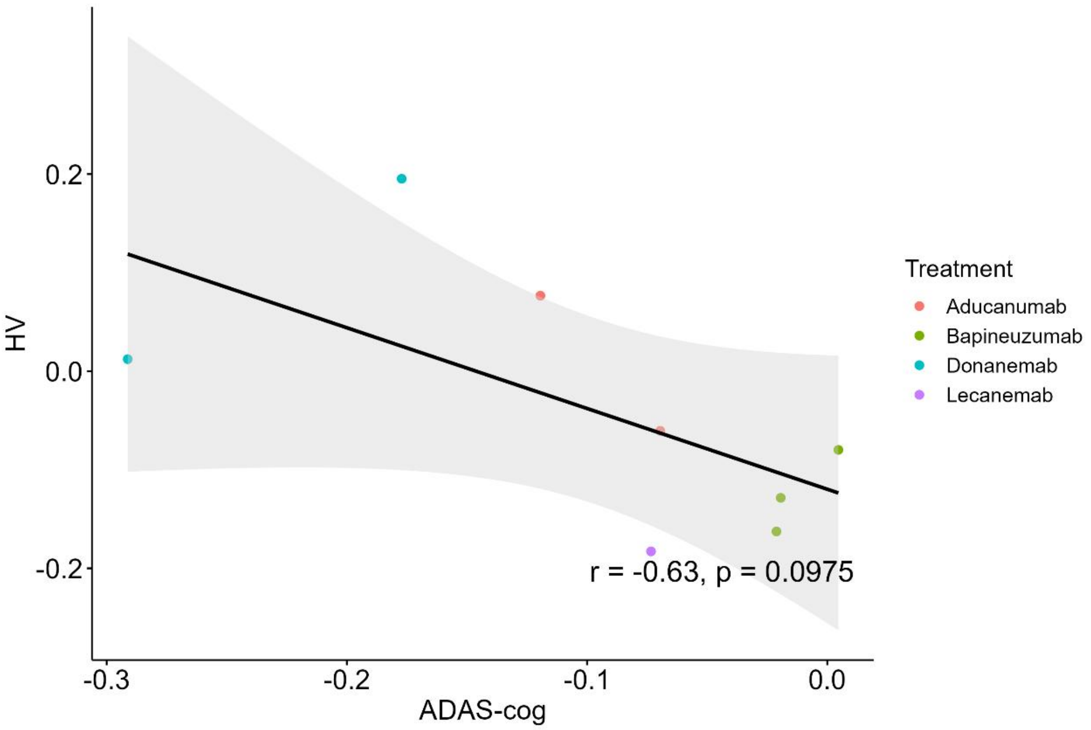

HV
0.2
0.0
-0.2
-0.3
-0.2
-0.1
0.0
ADAS-cog
r = -0.63, p = 0.0975
Treatment
Aducanumab
Bapineuzumab
Donanemab
Lecanemab

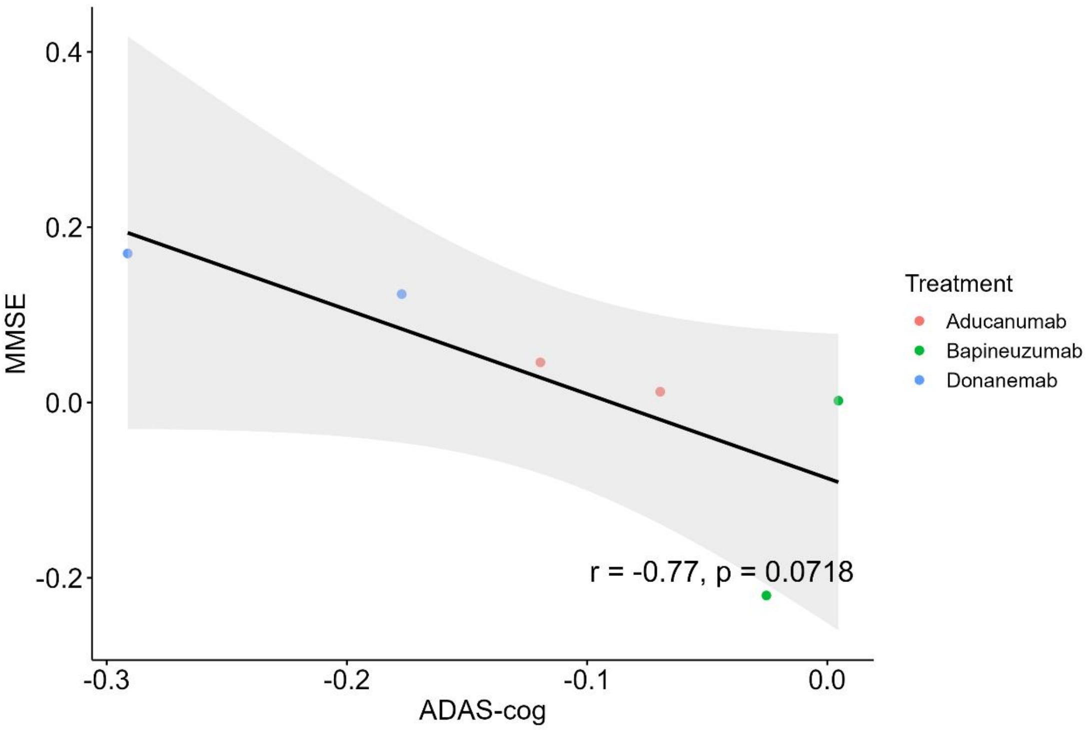

MMSE
0.4
0.2
0.0
-0.2
-0.3
-0.2
-0.1
0.0
ADAS-cog
r = -0.77, p = 0.0718
Treatment
Aducanumab
Bapineuzumab
Donanemab

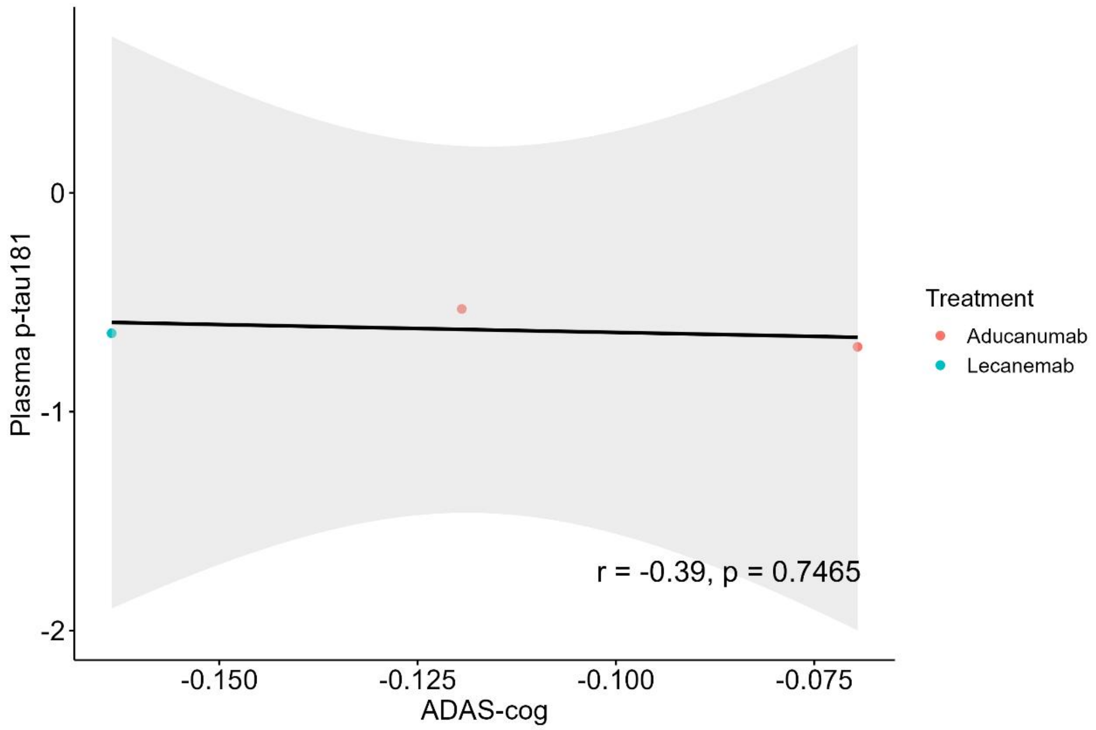
Plasma p-tau181
0
-1
-2
-0.150
-0.125
-0.100
-0.075
ADAS-cog
r = -0.39, p = 0.7465
Treatment
Aducanumab
Lecanemab

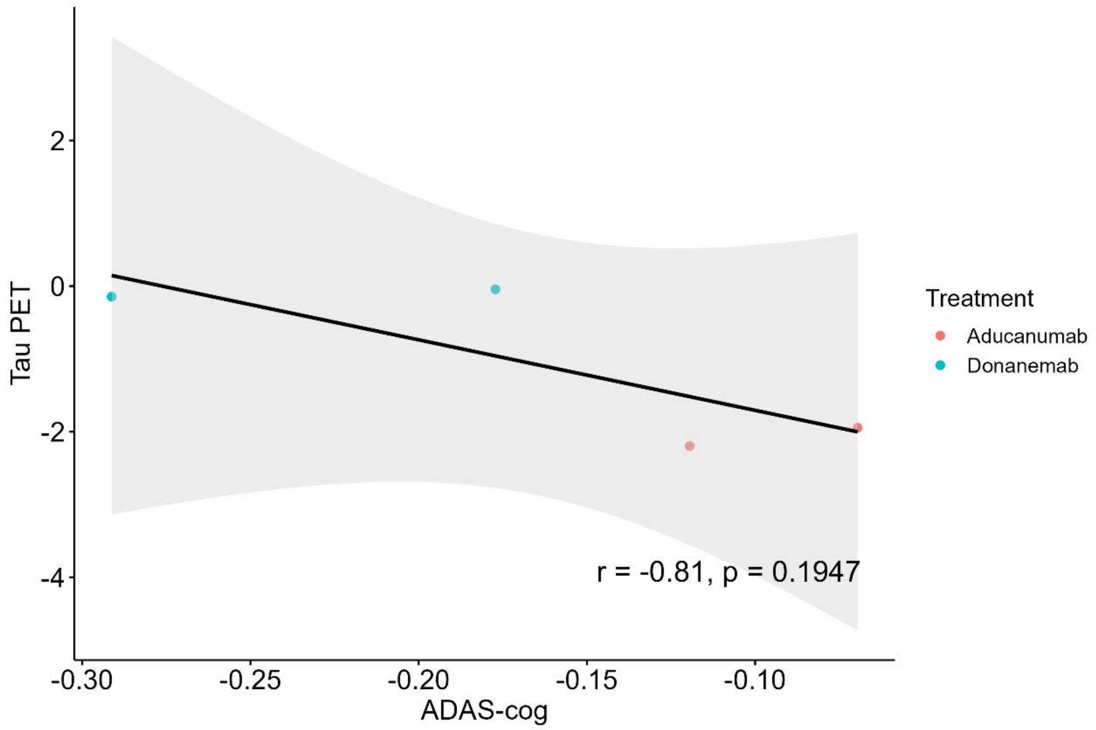
Tau PET
2
0
-2
-4
-0.30
-0.25
-0.20
-0.15
-0.10
ADAS-cog
r = -0.81, p = 0.1947
Treatment
Aducanumab
Donanemab

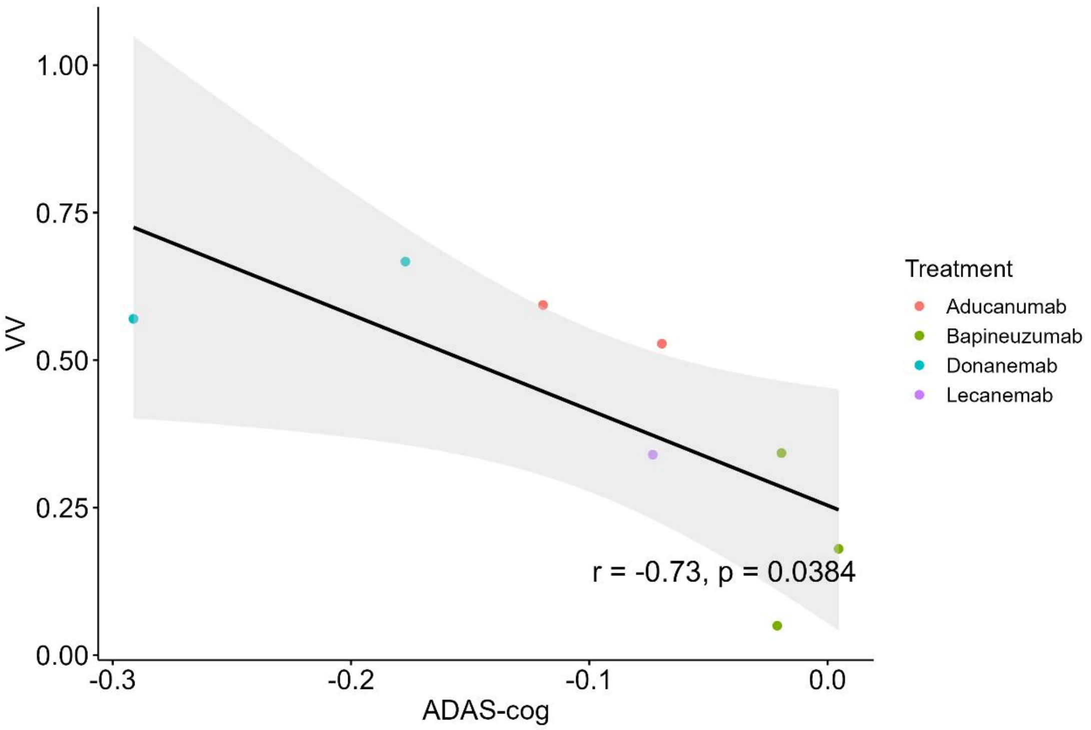
1.00
0.75
0.50
0.25
0.00
VV
-0.3
-0.2
-0.1
0.0
ADAS-cog
r = -0.73, p = 0.0384
Treatment
Aducanumab
Bapineuzumab
Donanemab
Lecanemab

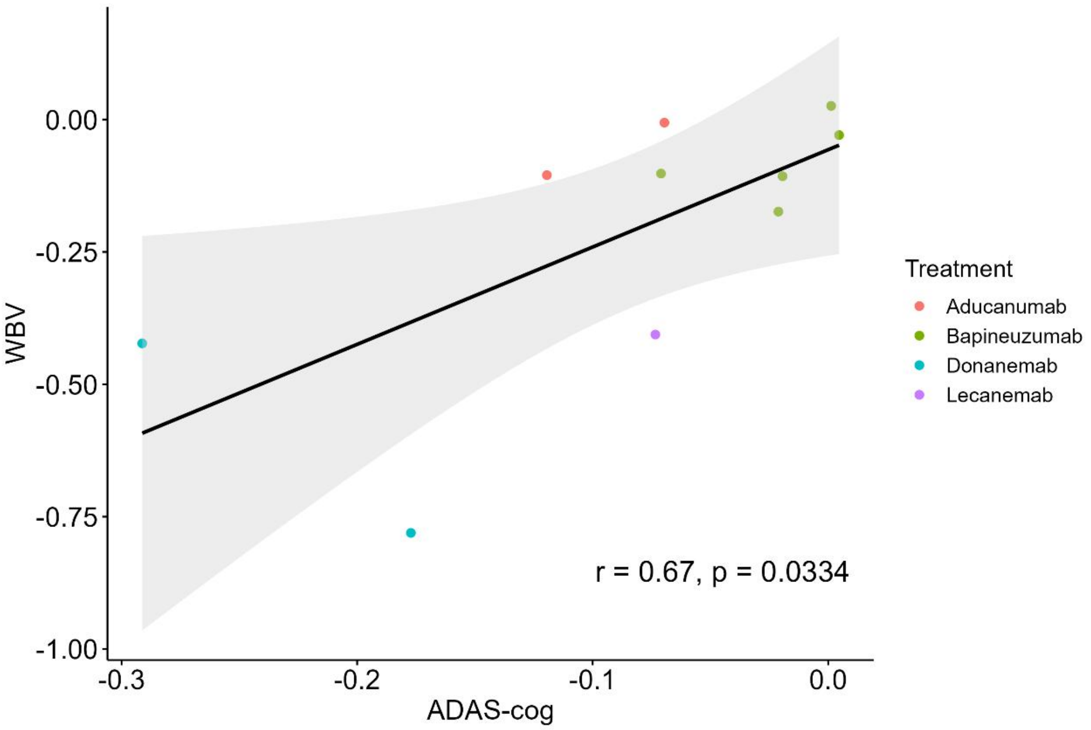
0.00
-0.25
-0.50
-0.75
-1.00
WBV
-0.3
-0.2
-0.1
0.0
ADAS-cog
r = 0.67, p = 0.0334
Treatment
Aducanumab
Bapineuzumab
Donanemab
Lecanemab

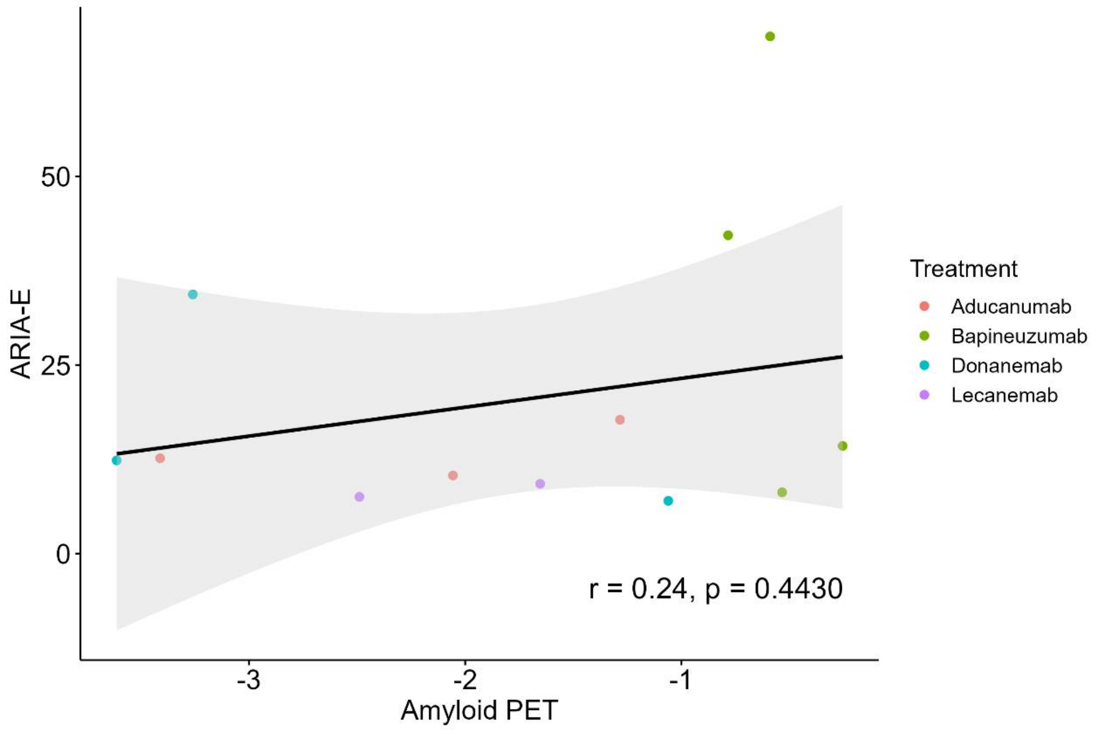
ARIA-E
50
25
0
-3
-2
-1
Amyloid PET
r = 0.24, p = 0.4430
Treatment
Aducanumab
Bapineuzumab
Donanemab
Lecanemab

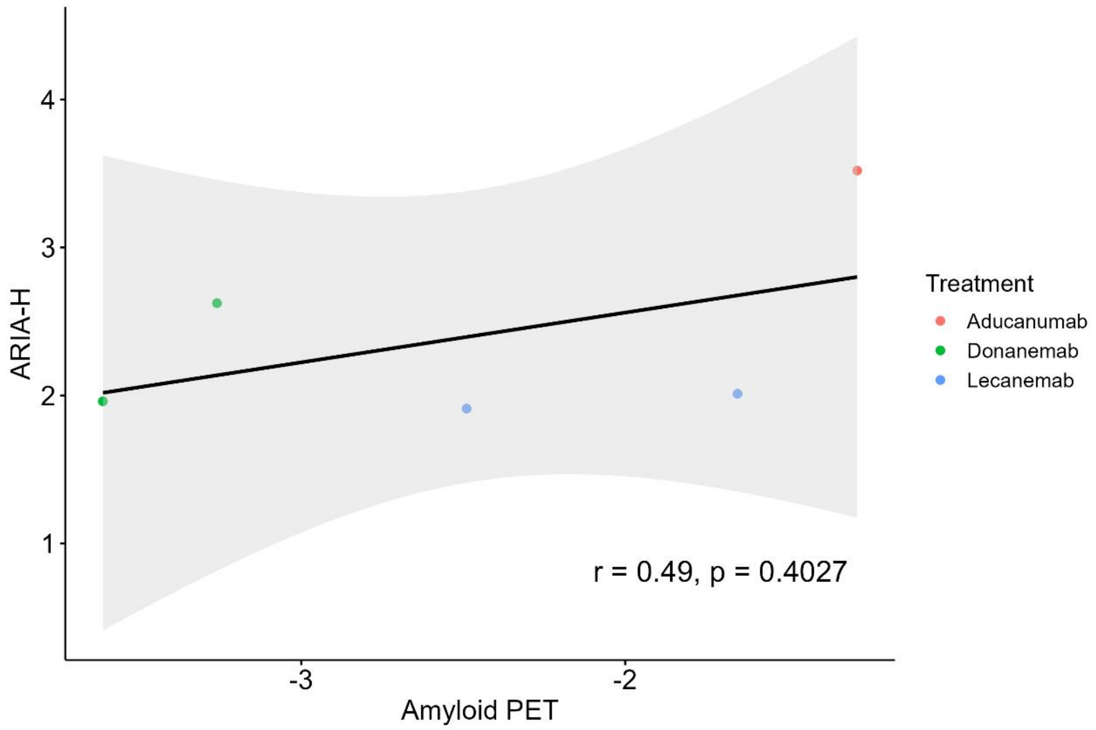
ARIA-H
4
3
2
1
-3
-2
Amyloid PET
r = 0.49, p = 0.4027
Treatment
Aducanumab
Donanemab
Lecanemab

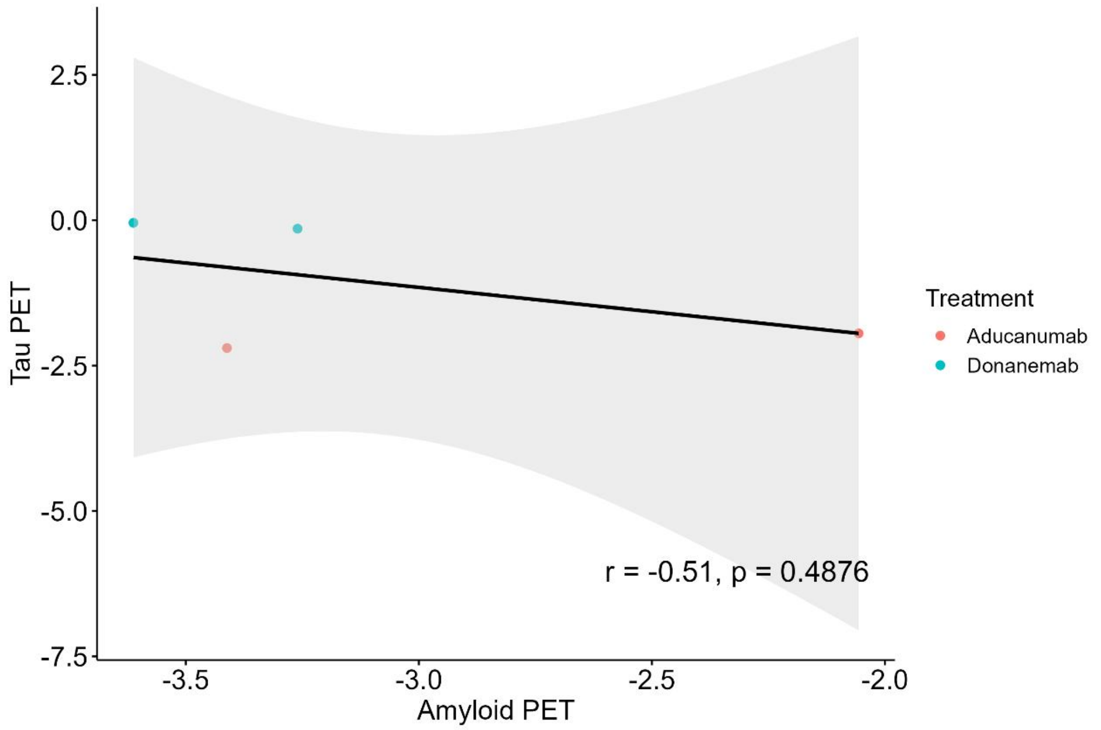

2.5
0.0
-2.5
-5.0
-7.5
Tau PET
-3.5
-3.0
-2.5
-2.0
Amyloid PET
r = -0.51, p = 0.4876
Treatment
Aducanumab
Donanemab

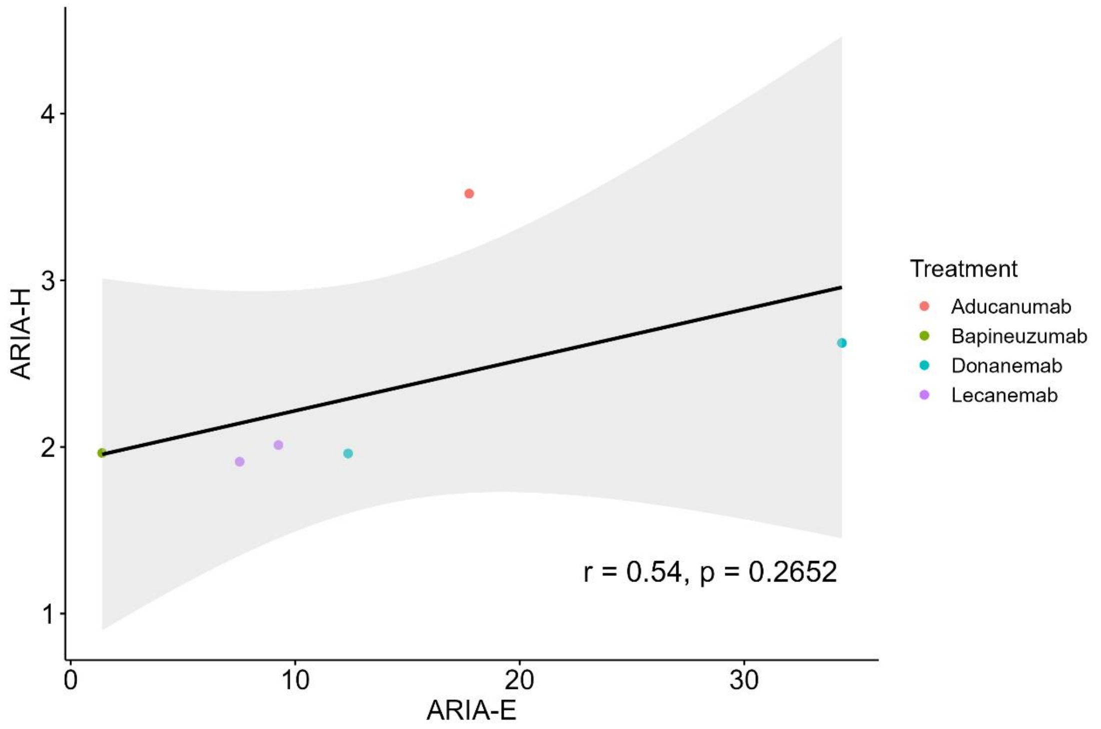

4
3
2
1
ARIA-H
0
10
20
30
ARIA-E
r = 0.54, p = 0.2652
Treatment
Aducanumab
Bapineuzumab
Donanemab
Lecanemab

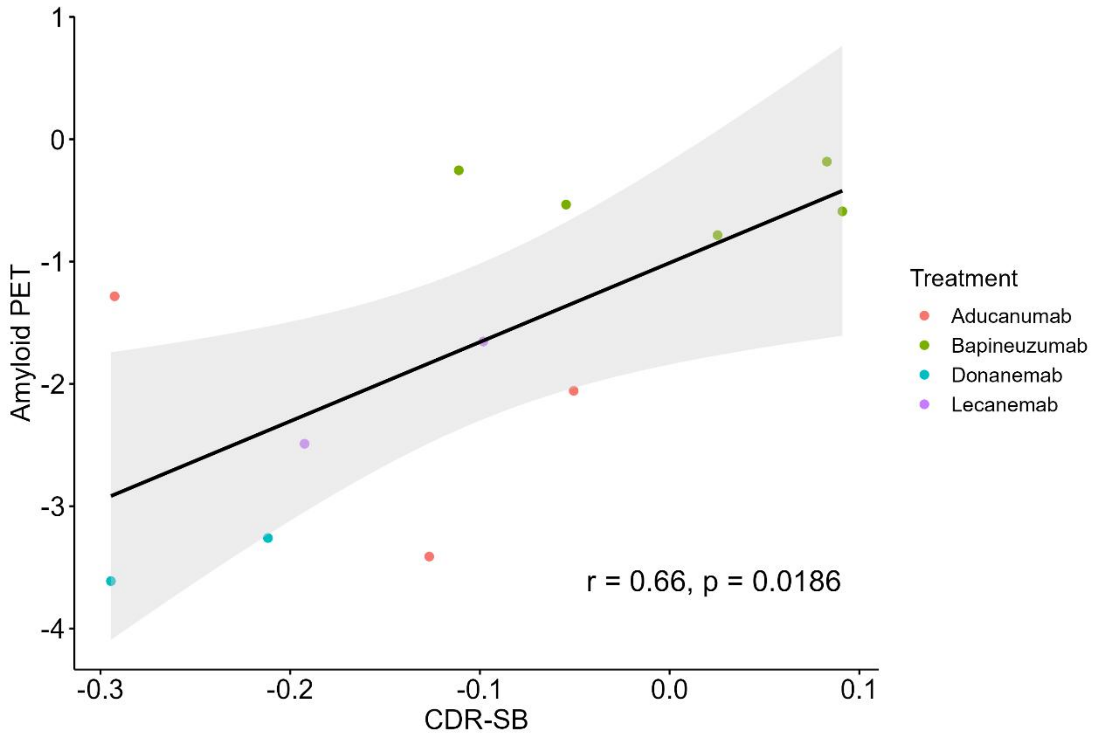
1
0
-1
-2
-3
-4
Amyloid PET
-0.3
-0.2
-0.1
0.0
0.1
CDR-SB
r = 0.66, p = 0.0186
Treatment
Aducanumab
Bapineuzumab
Donanemab
Lecanemab

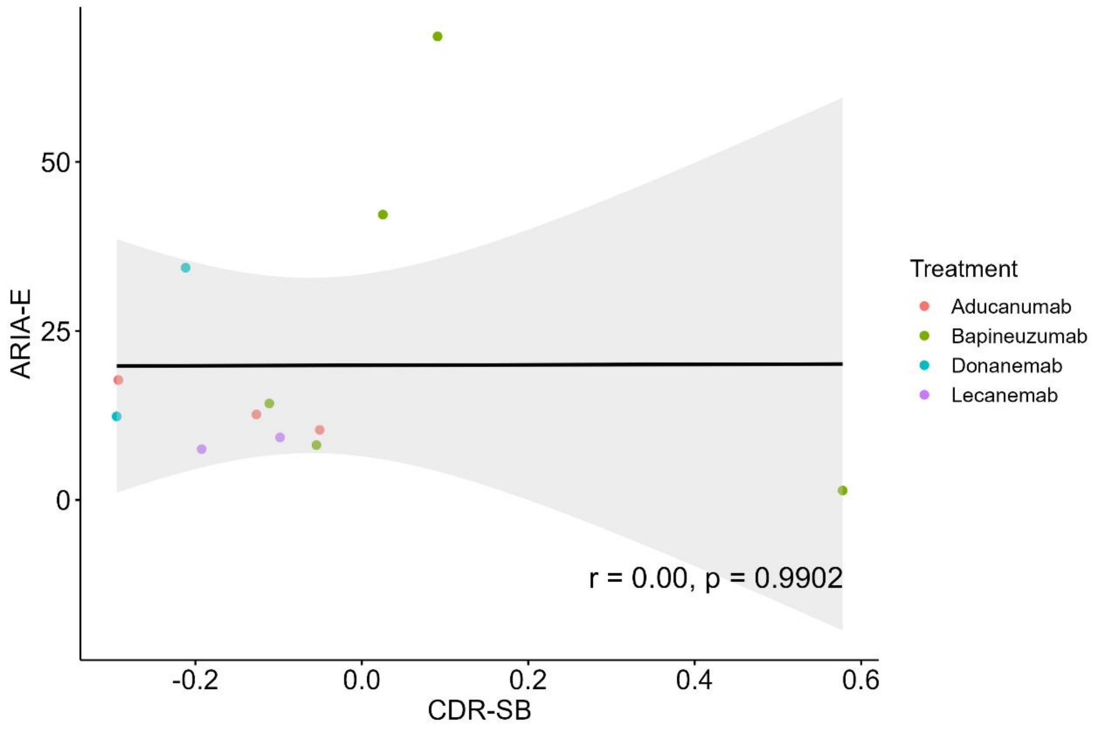
50
25
0
ARIA-E
-0.2
0.0
0.2
0.4
0.6
CDR-SB
r = 0.00, p = 0.9902
Treatment
Aducanumab
Bapineuzumab
Donanemab
Lecanemab

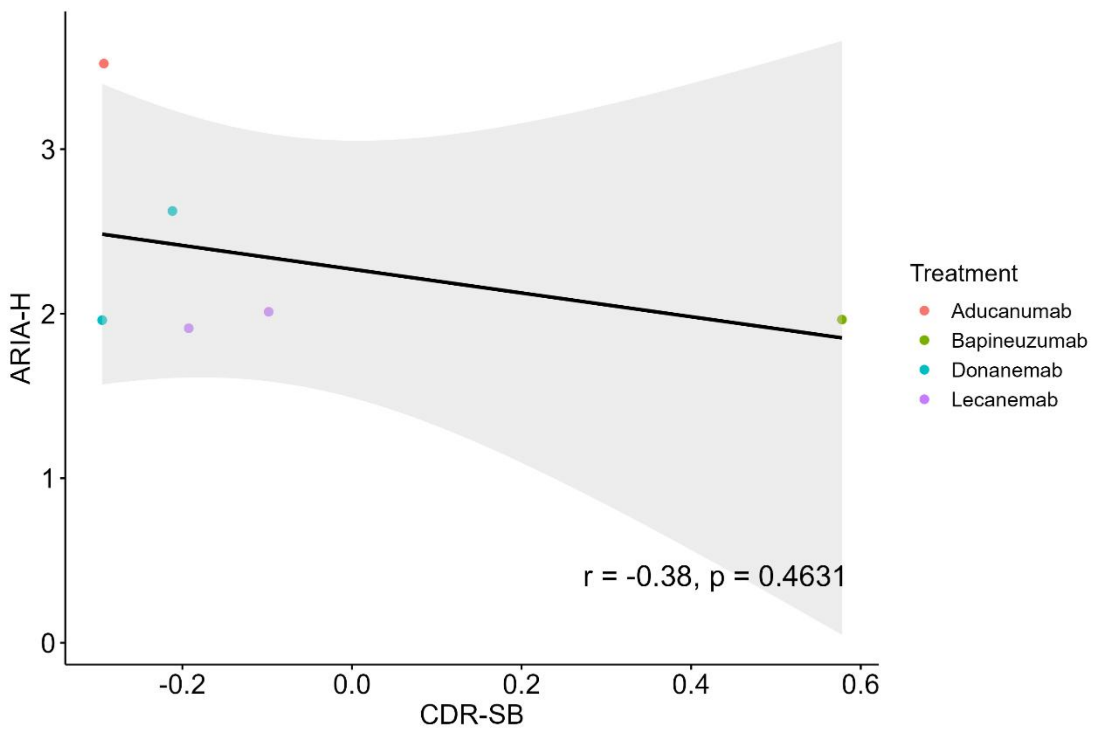
ARIA-H
CDR-SB
Treatment
Aducanumab
Bapineuzumab
Donanemab
Lecanemab
r = -0.38, p = 0.4631

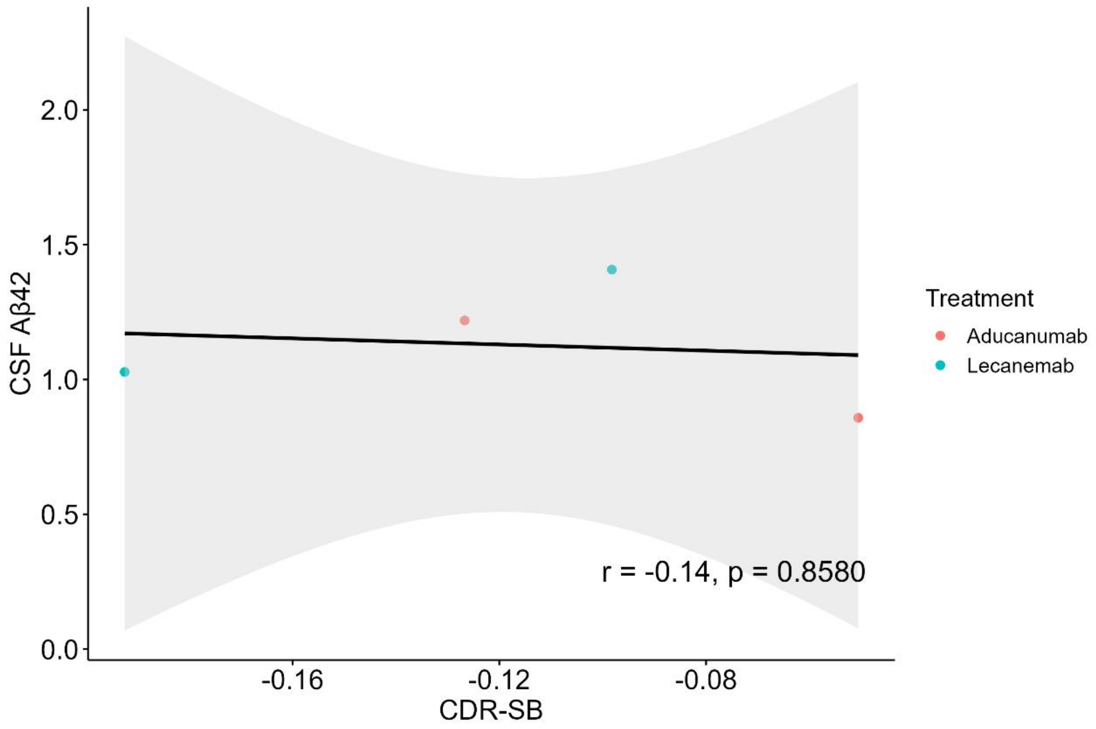
CSF Aβ42
CDR-SB
Treatment
Aducanumab
Lecanemab
r = -0.14, p = 0.8580

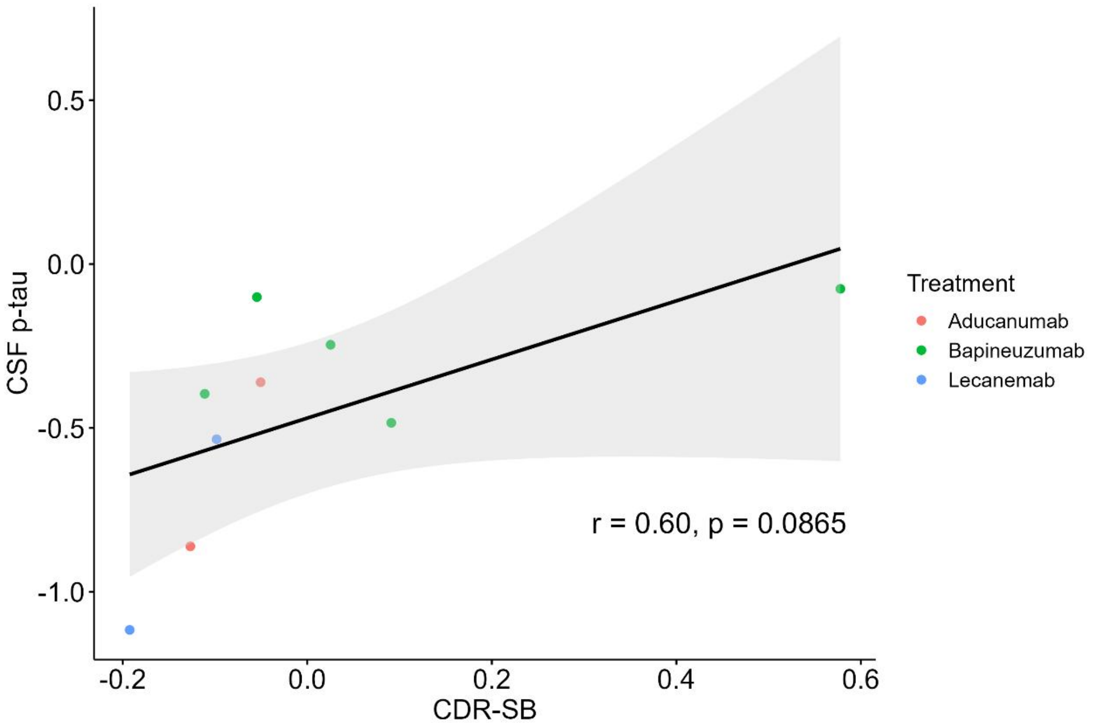
0.5
0.0
-0.5
-1.0
CSF p-tau
-0.2
0.0
0.2
0.4
0.6
CDR-SB
r = 0.60, p = 0.0865
Treatment
Aducanumab
Bapineuzumab
Lecanemab

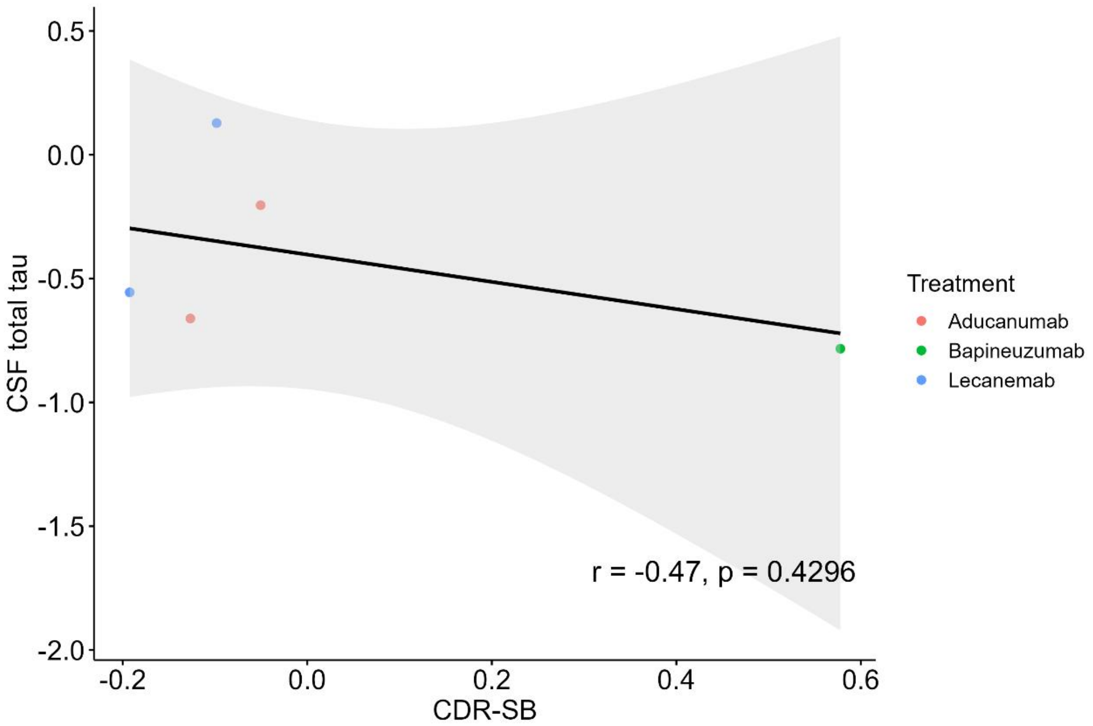
0.5
0.0
-0.5
-1.0
-1.5
-2.0
CSF total tau
-0.2
0.0
0.2
0.4
0.6
CDR-SB
r = -0.47, p = 0.4296
Treatment
Aducanumab
Bapineuzumab
Lecanemab

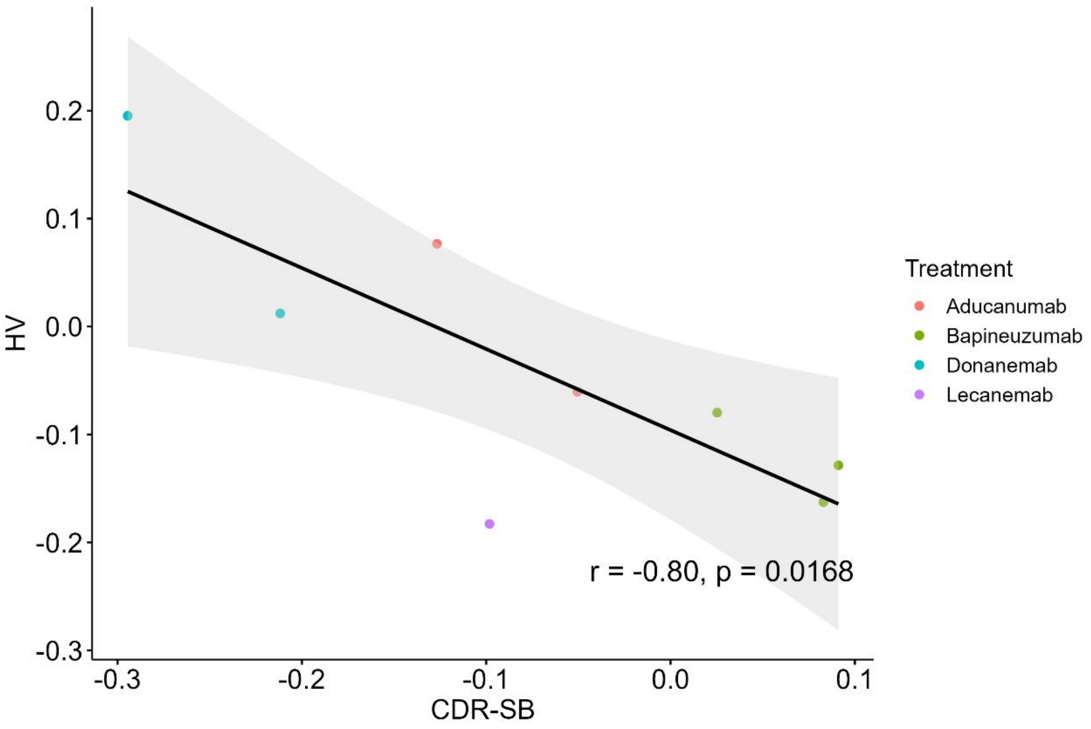
0.2
0.1
0.0
-0.1
-0.2
-0.3
HV
-0.3
-0.2
-0.1
0.0
0.1
CDR-SB
r = -0.80, p = 0.0168
Treatment
Aducanumab
Bapineuzumab
Donanemab
Lecanemab

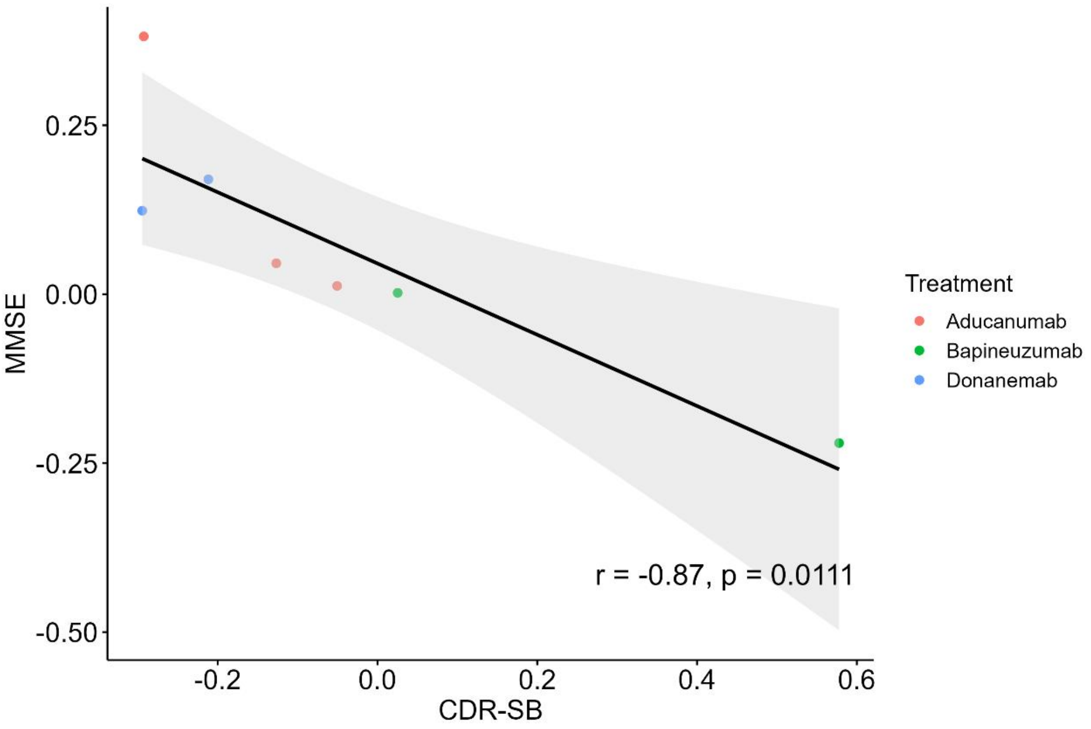
0.25
0.00
-0.25
-0.50
MMSE
-0.2
0.0
0.2
0.4
0.6
CDR-SB
r = -0.87, p = 0.0111
Treatment
Aducanumab
Bapineuzumab
Donanemab

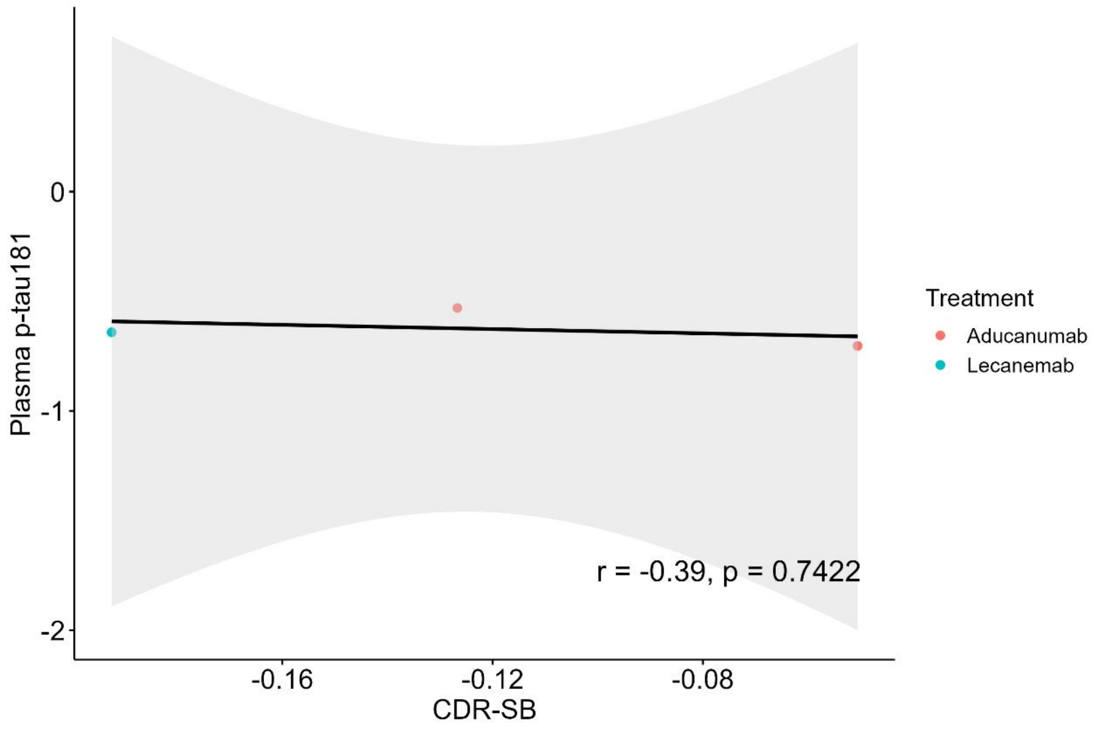
Plasma p-tau181
0
-1
-2
-0.16
-0.12
-0.08
CDR-SB
r = -0.39, p = 0.7422
Treatment
Aducanumab
Lecanemab

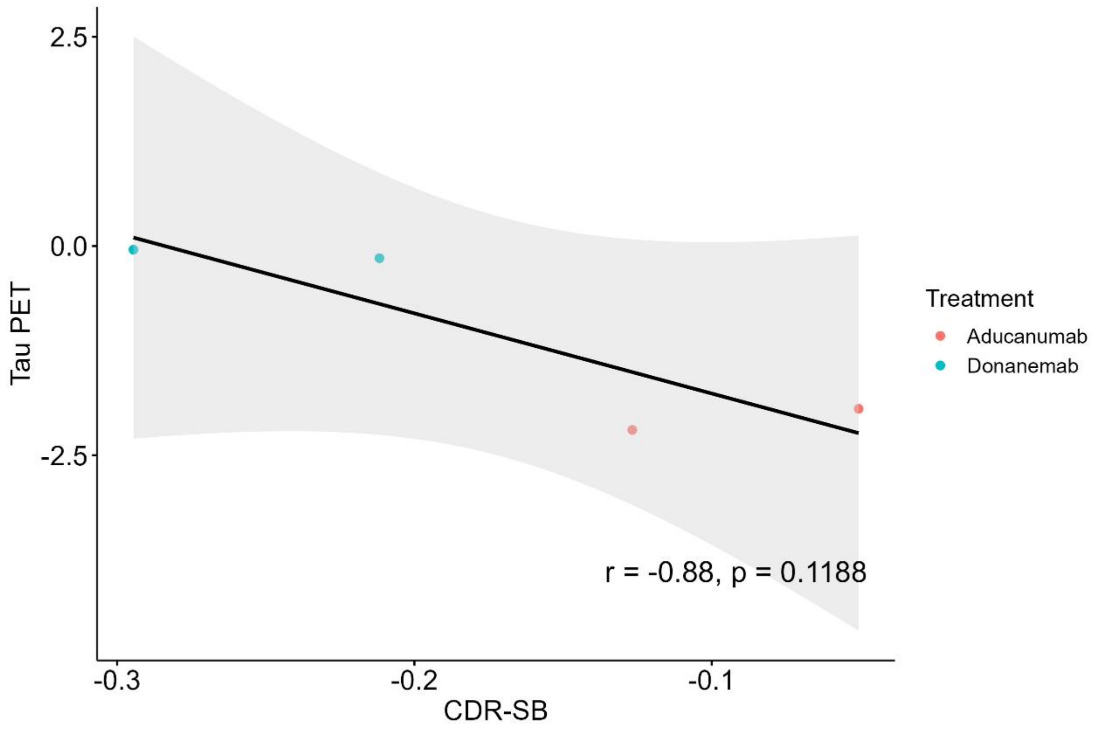
Tau PET
2.5
0.0
-2.5
-0.3
-0.2
-0.1
CDR-SB
r = -0.88, p = 0.1188
Treatment
Aducanumab
Donanemab

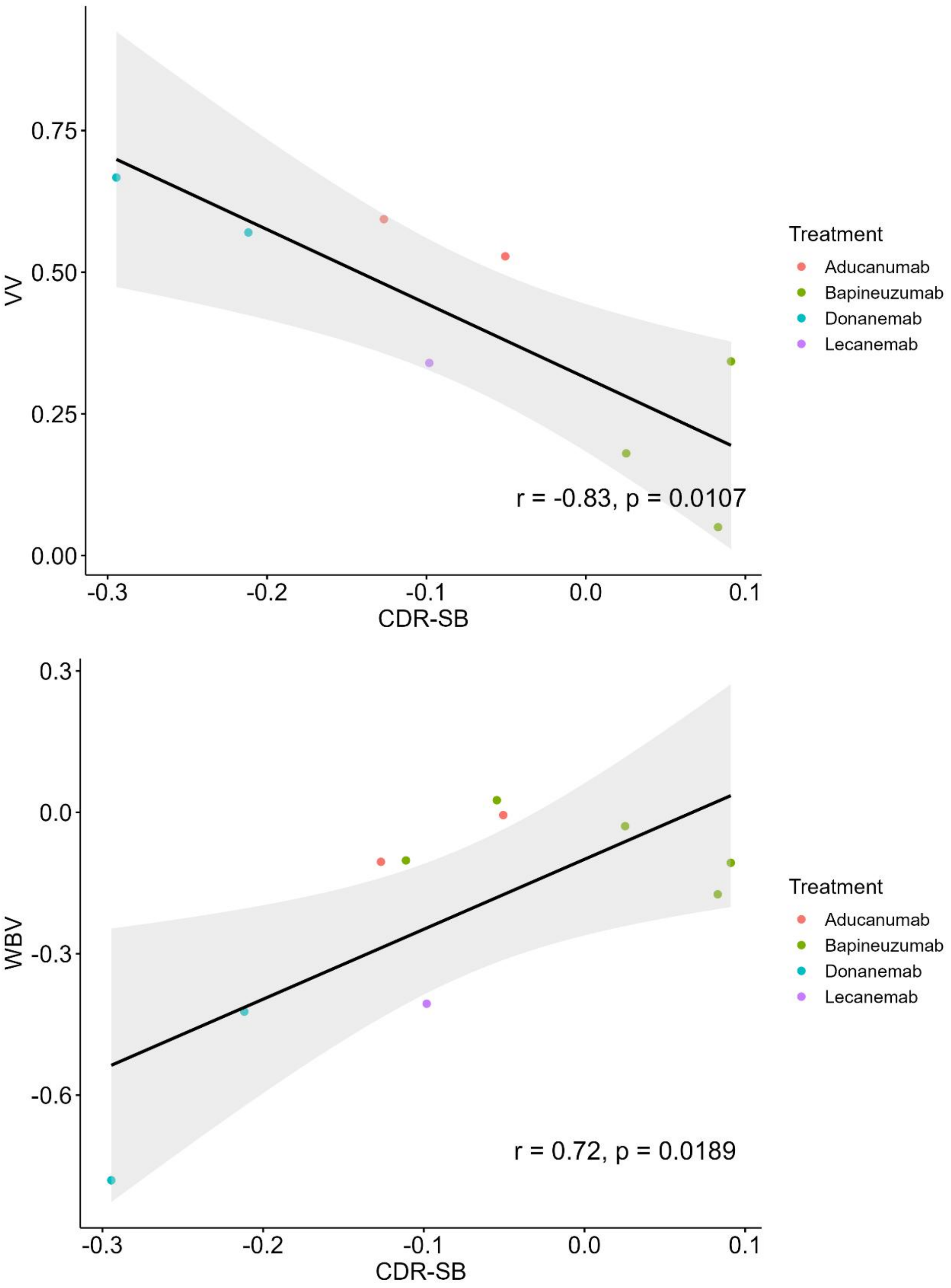
0.75
0.50
0.25
0.00
VV
-0.3
-0.2
-0.1
0.0
0.1
CDR-SB
r = -0.83, p = 0.0107
Treatment
Aducanumab
Bapineuzumab
Donanemab
Lecanemab
0.3
0.0
-0.3
-0.6
WBV
-0.3
-0.2
-0.1
0.0
0.1
CDR-SB
r = 0.72, p = 0.0189
Treatment
Aducanumab
Bapineuzumab
Donanemab
Lecanemab

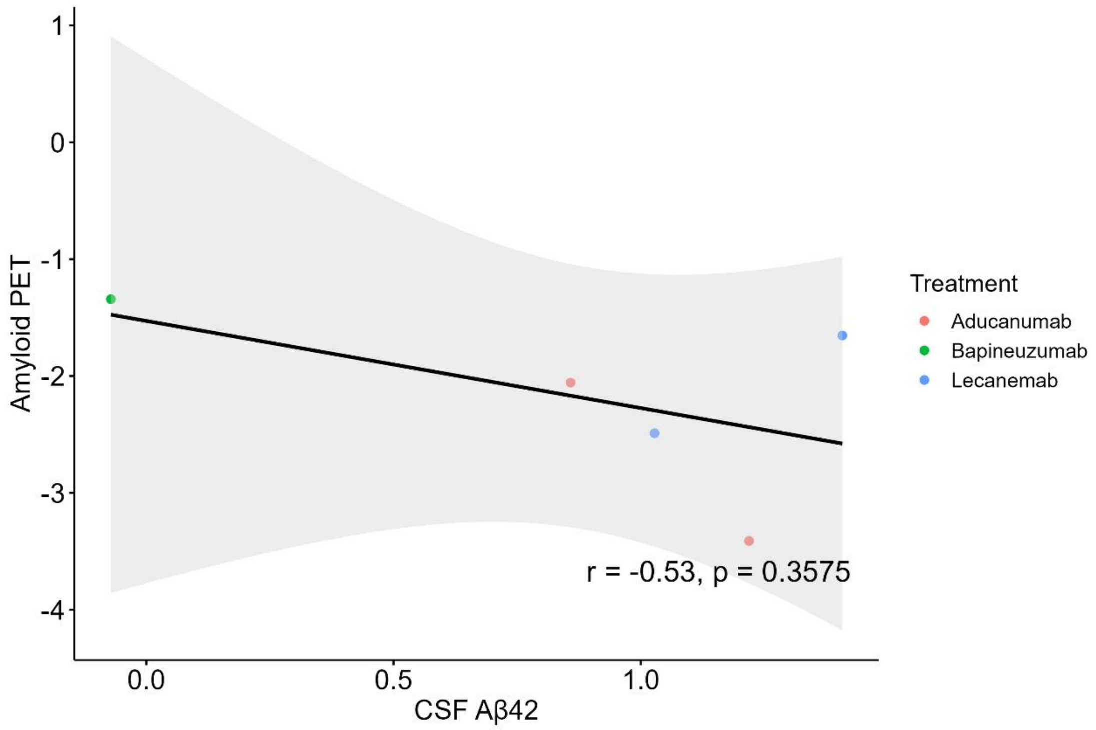
Amyloid PET
CSF Aβ42
Treatment
Aducanumab
Bapineuzumab
Lecanemab
r = -0.53, p = 0.3575
1
0
-1
-2
-3
-4
0.0
0.5
1.0

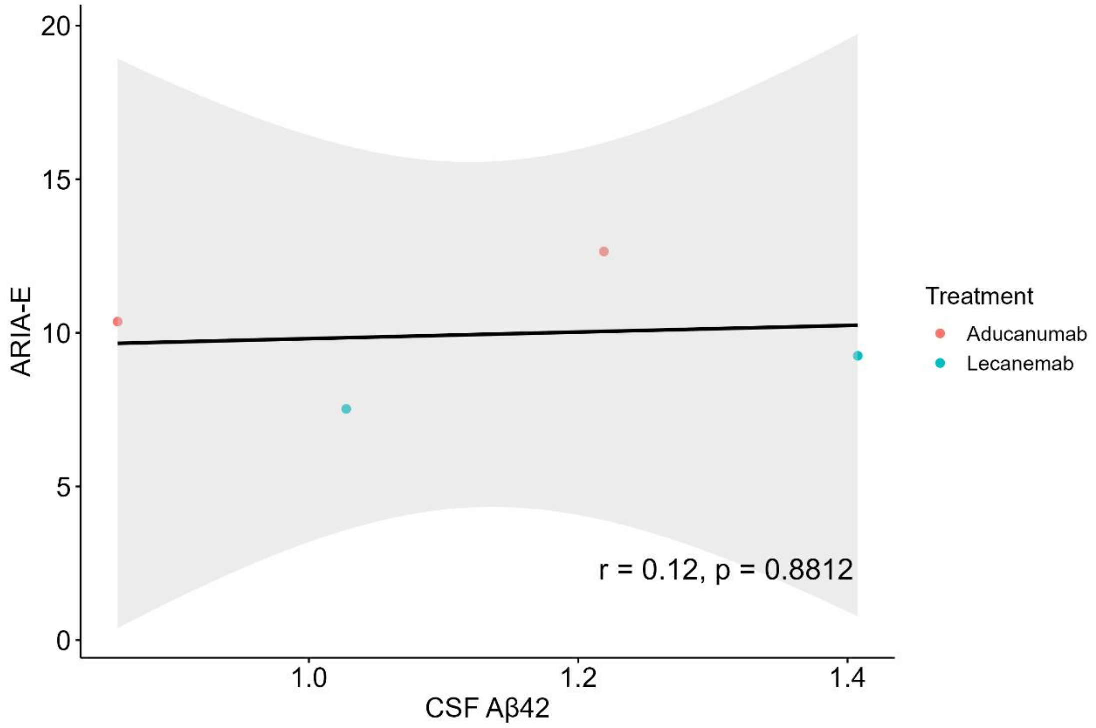
ARIA-E
CSF Aβ42
Treatment
Aducanumab
Lecanemab
r = 0.12, p = 0.8812
20
15
10
5
0
1.0
1.2
1.4

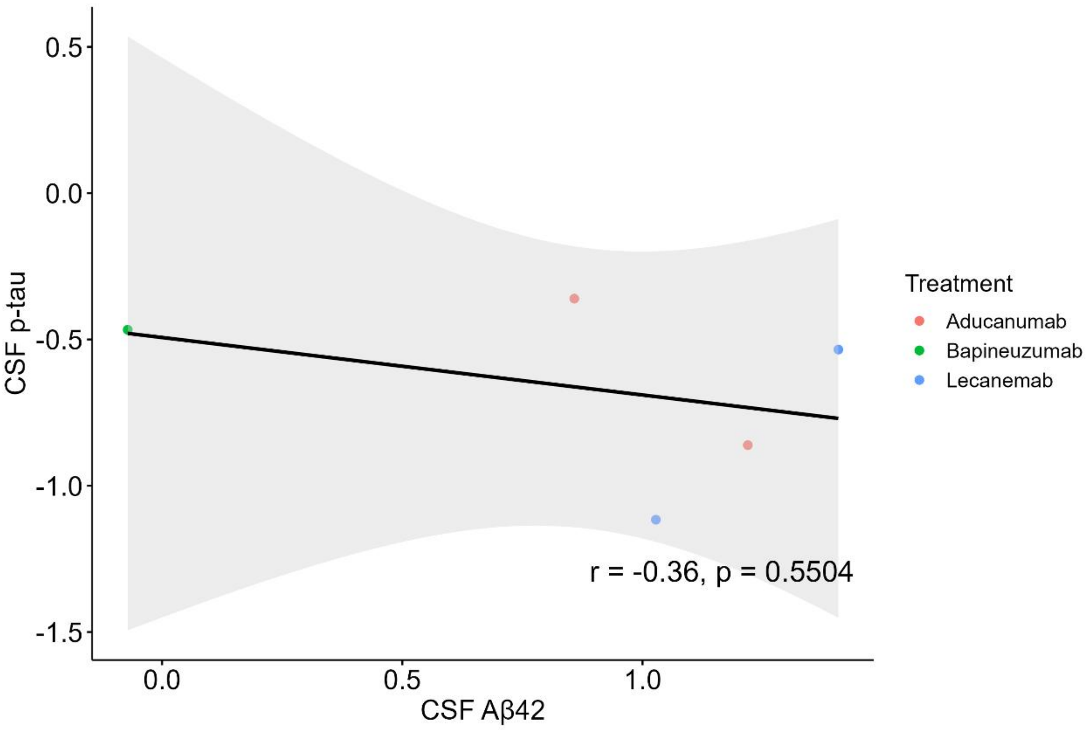

0.5
0.0
-0.5
-1.0
-1.5
CSF p-tau
r = -0.36, p = 0.5504
0.0
0.5
1.0
CSF Aβ42
Treatment
Aducanumab
Bapineuzumab
Lecanemab

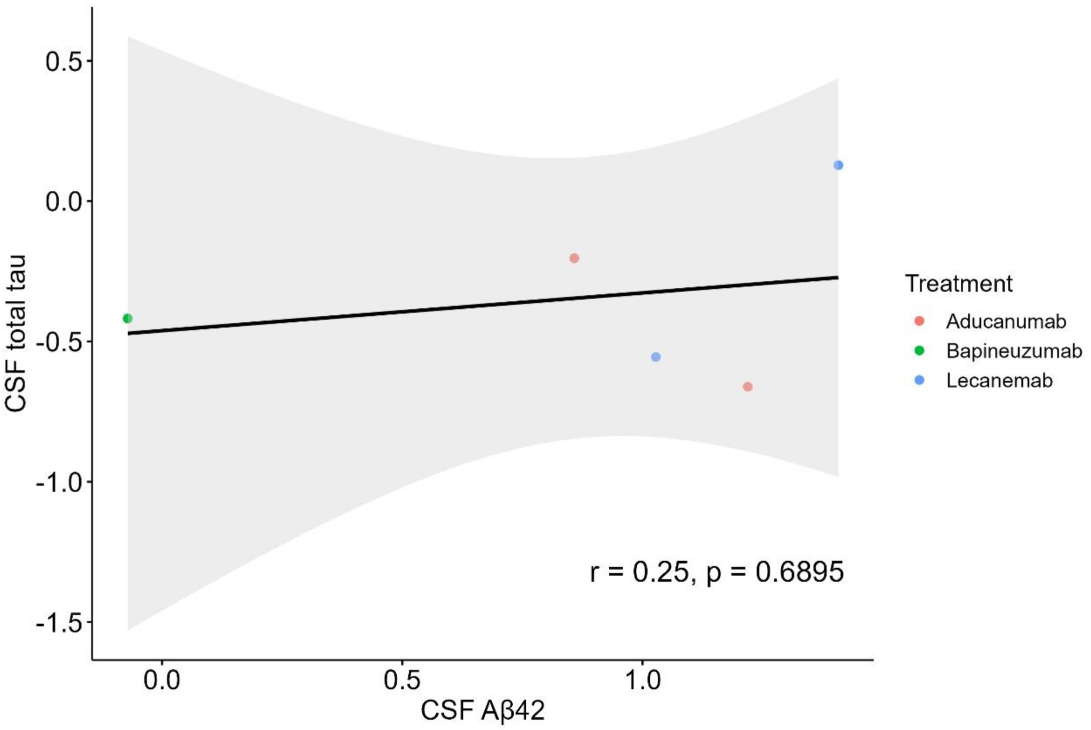

0.5
0.0
-0.5
-1.0
-1.5
CSF total tau
r = 0.25, p = 0.6895
0.0
0.5
1.0
CSF Aβ42
Treatment
Aducanumab
Bapineuzumab
Lecanemab

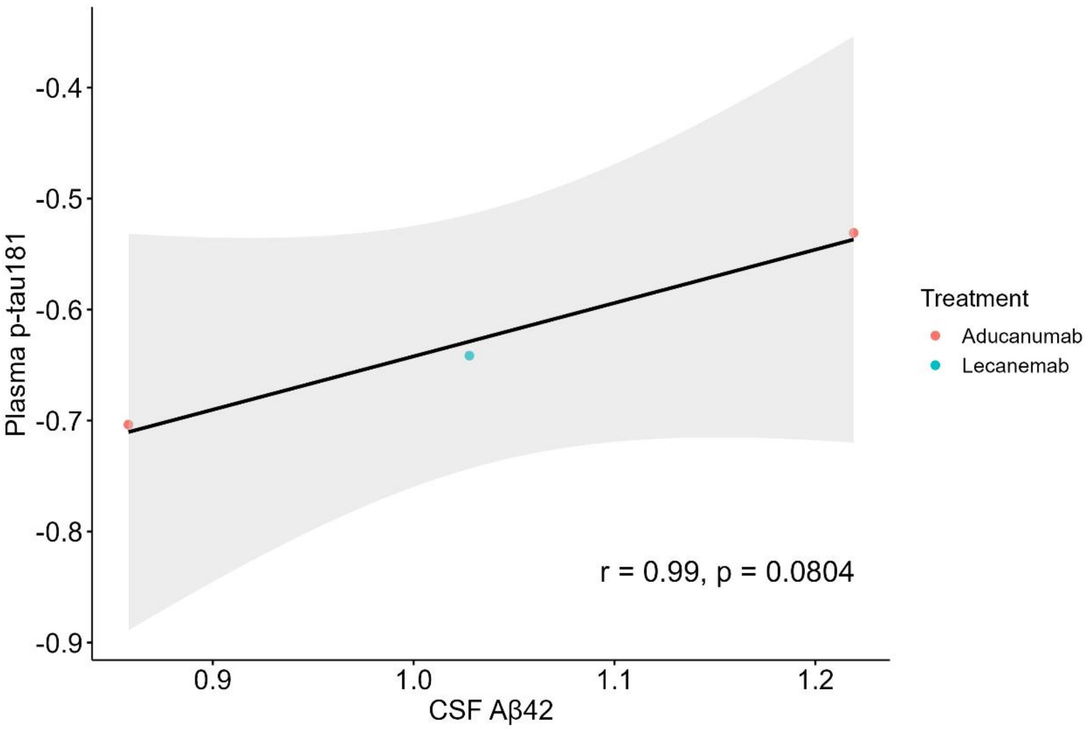
Plasma p-tau181
-0.4
-0.5
-0.6
-0.7
-0.8
-0.9
0.9
1.0
1.1
1.2
CSF Aβ42
r = 0.99, p = 0.0804
Treatment
Aducanumab
Lecanemab

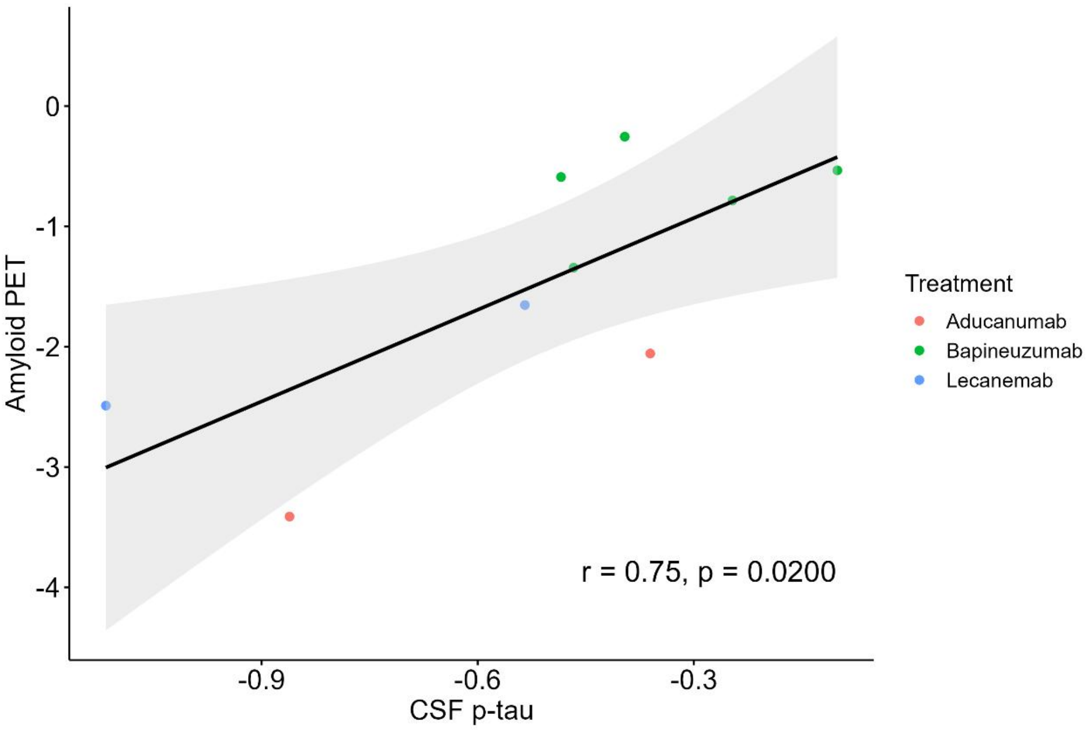
Amyloid PET
0
-1
-2
-3
-4
-0.9
-0.6
-0.3
CSF p-tau
r = 0.75, p = 0.0200
Treatment
Aducanumab
Bapineuzumab
Lecanemab

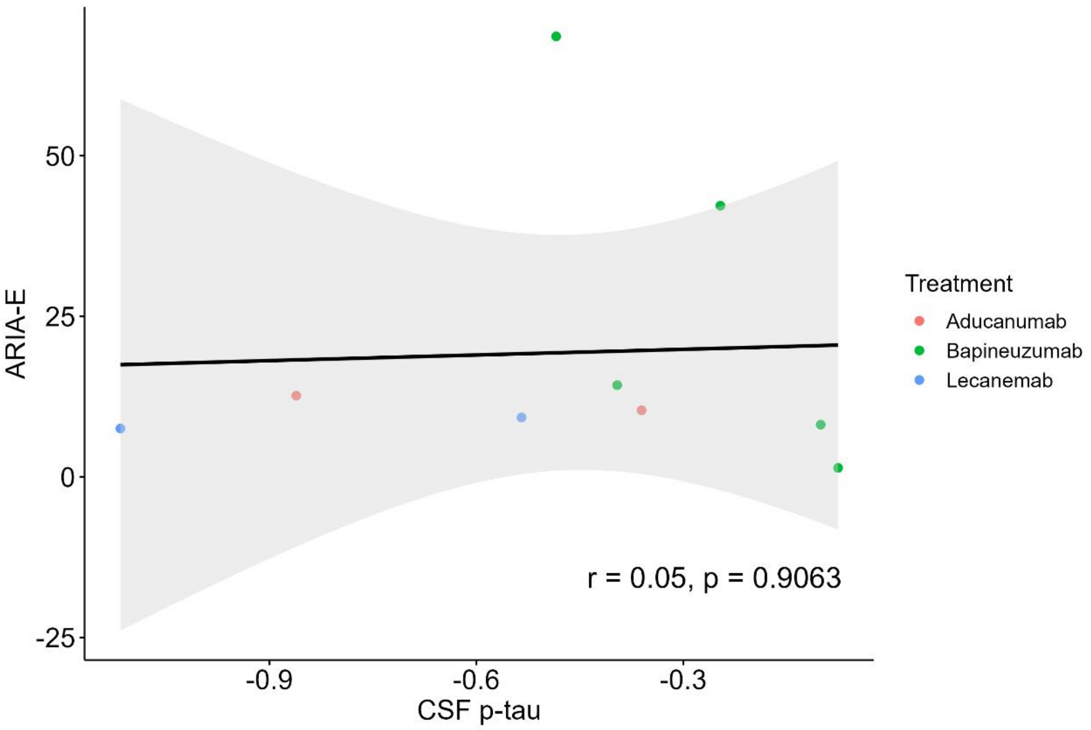
ARIA-E
50
25
0
-25
-0.9
-0.6
-0.3
CSF p-tau
r = 0.05, p = 0.9063
Treatment
Aducanumab
Bapineuzumab
Lecanemab

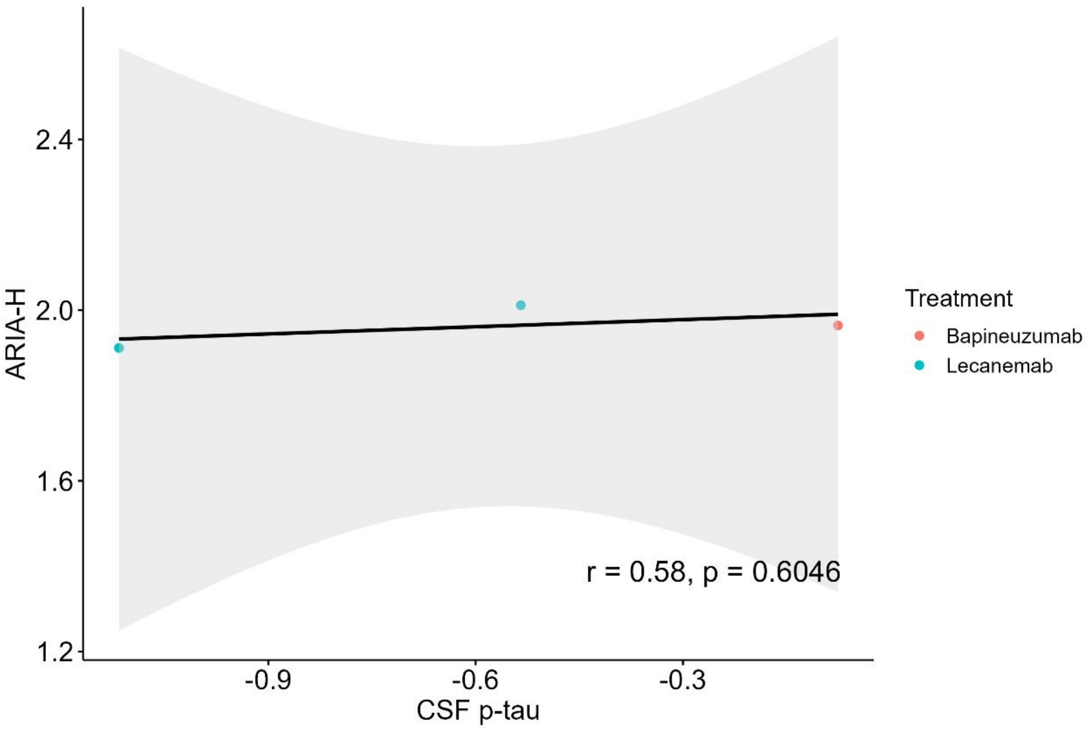
ARIA-H
2.4
2.0
1.6
1.2
-0.9
-0.6
-0.3
CSF p-tau
r = 0.58, p = 0.6046
Treatment
Bapineuzumab
Lecanemab

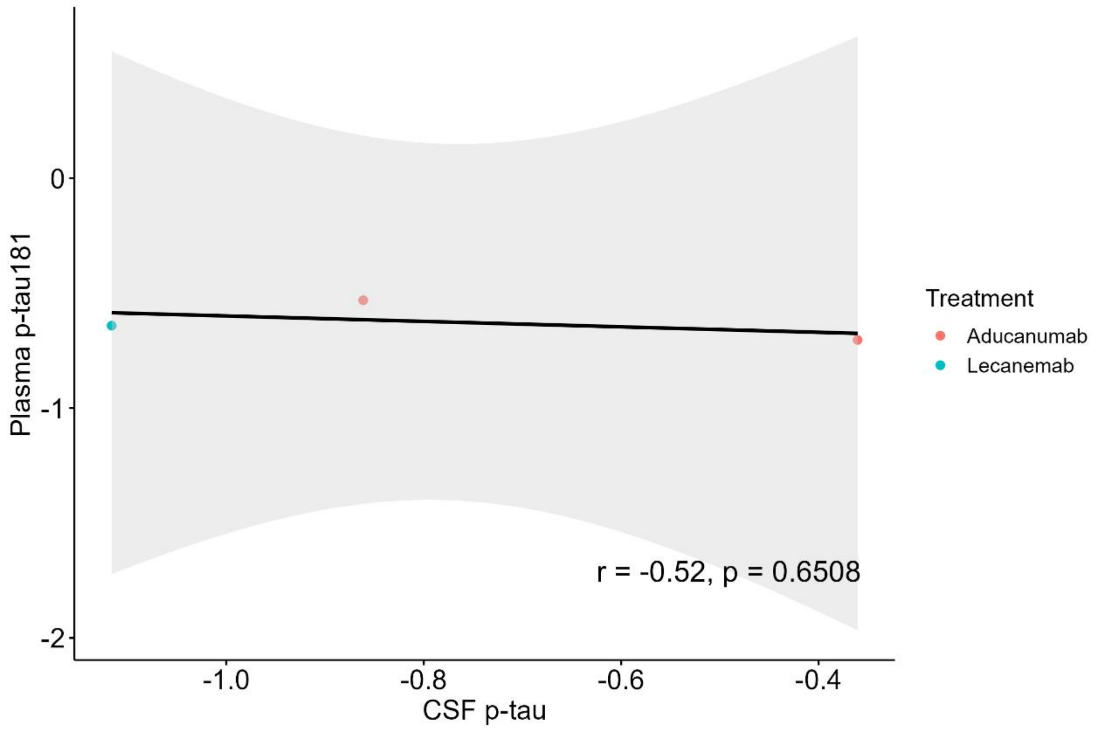
Plasma p-tau181
0
-1
-2
-1.0
-0.8
-0.6
-0.4
CSF p-tau
r = -0.52, p = 0.6508
Treatment
Aducanumab
Lecanemab

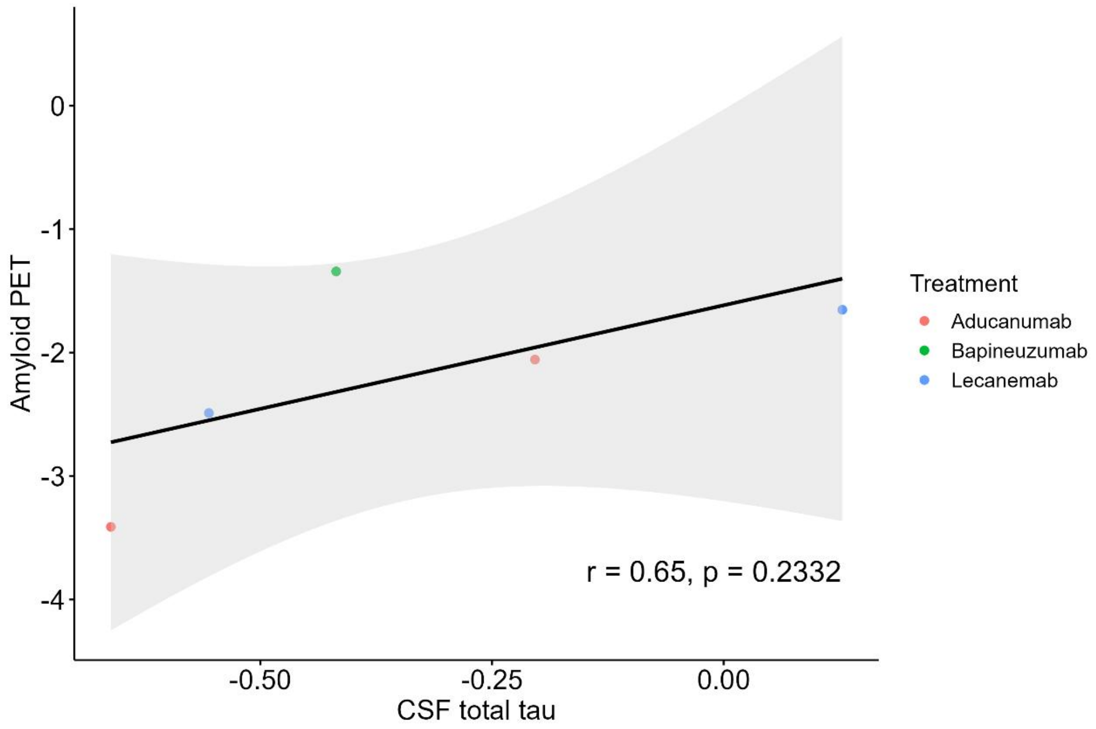
Amyloid PET
0
-1
-2
-3
-4
-0.50
-0.25
0.00
CSF total tau
r = 0.65, p = 0.2332
Treatment
Aducanumab
Bapineuzumab
Lecanemab

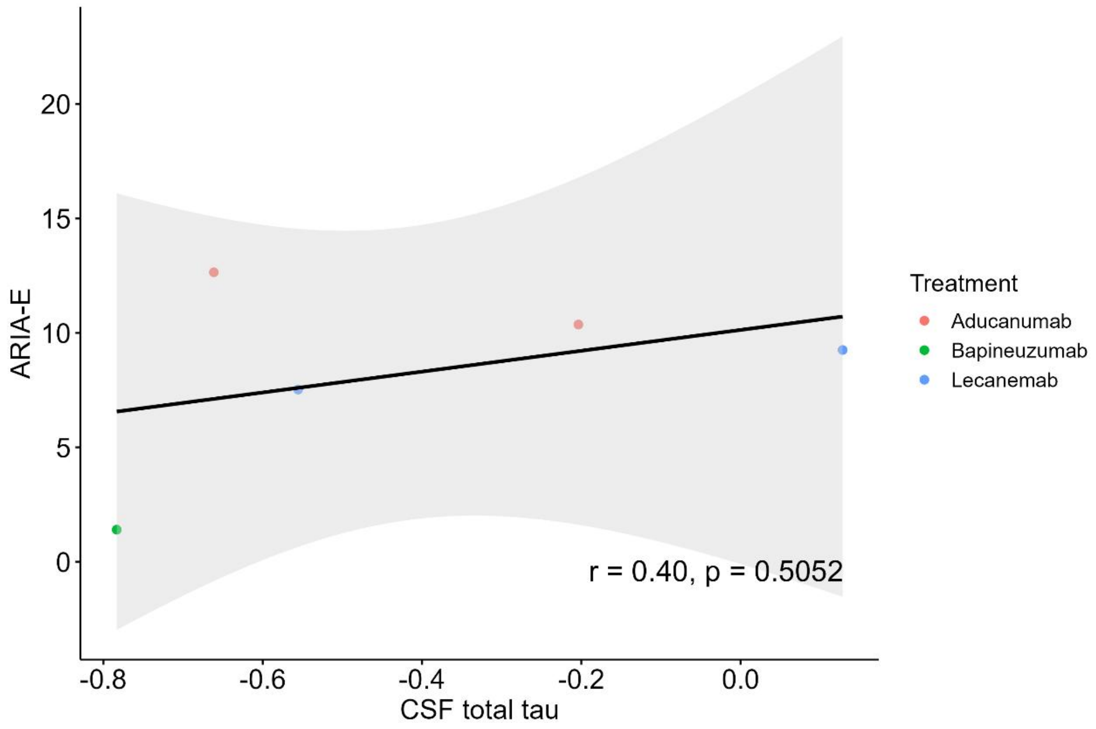
ARIA-E
CSF total tau
r = 0.40, p = 0.5052
Treatment
Aducanumab
Bapineuzumab
Lecanemab
20
15
10
5
0
-0.8
-0.6
-0.4
-0.2
0.0

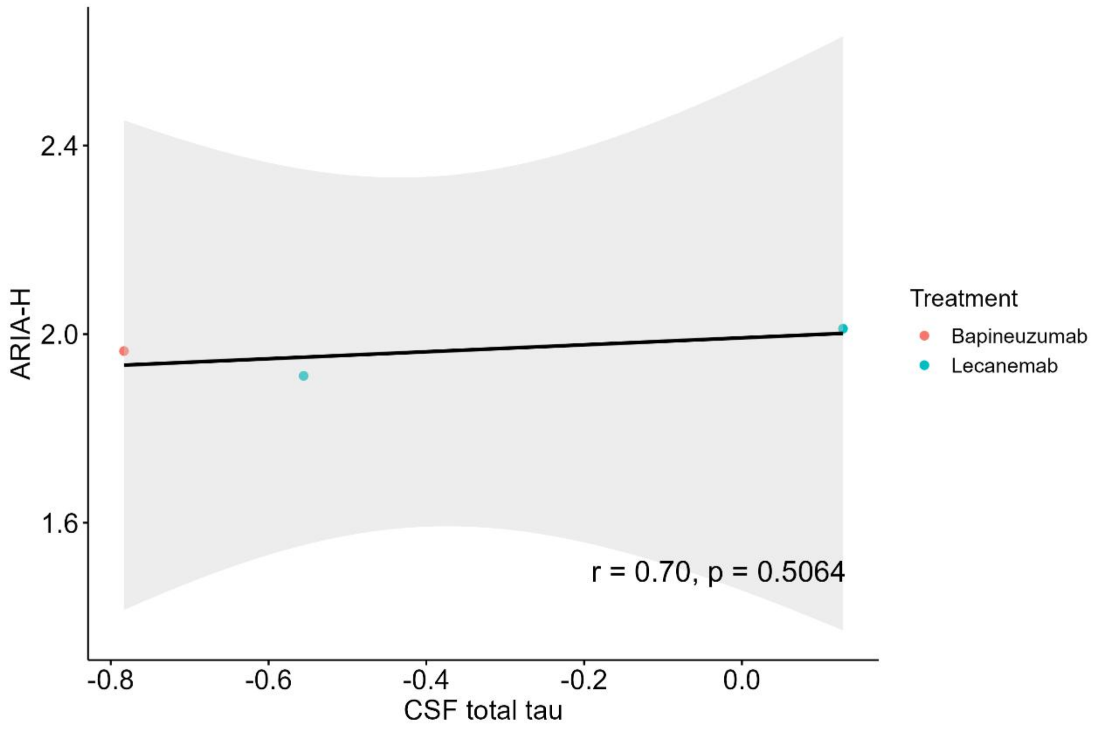
ARIA-H
CSF total tau
r = 0.70, p = 0.5064
Treatment
Bapineuzumab
Lecanemab
2.4
2.0
1.6
-0.8
-0.6
-0.4
-0.2
0.0

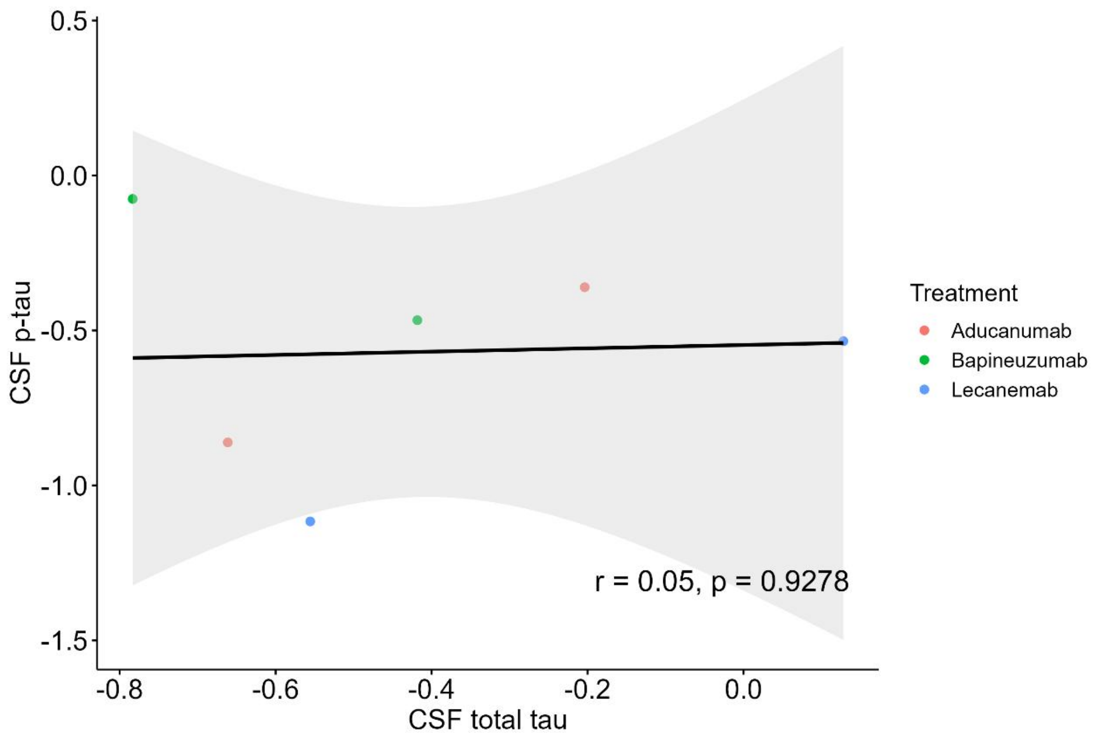

0.5
0.0
-0.5
-1.0
-1.5
CSF p-tau
-0.8
-0.6
-0.4
-0.2
0.0
CSF total tau
r = 0.05, p = 0.9278
Treatment
Aducanumab
Bapineuzumab
Lecanemab

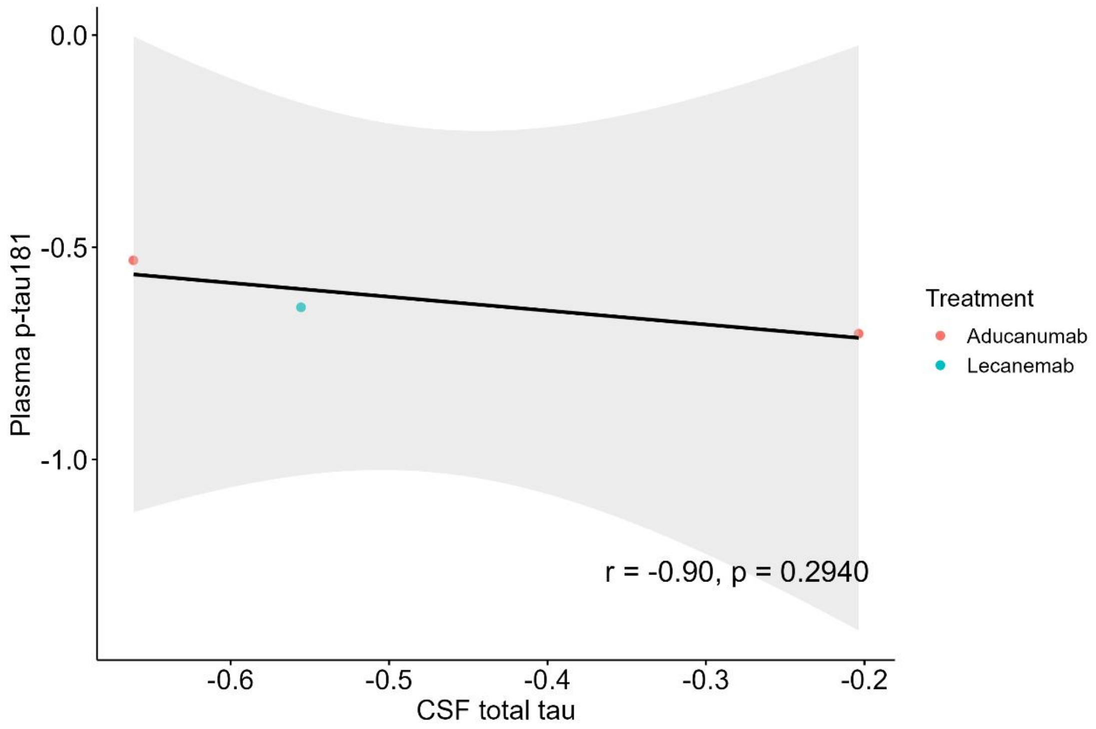

0.0
-0.5
-1.0
Plasma p-tau181
-0.6
-0.5
-0.4
-0.3
-0.2
CSF total tau
r = -0.90, p = 0.2940
Treatment
Aducanumab
Lecanemab

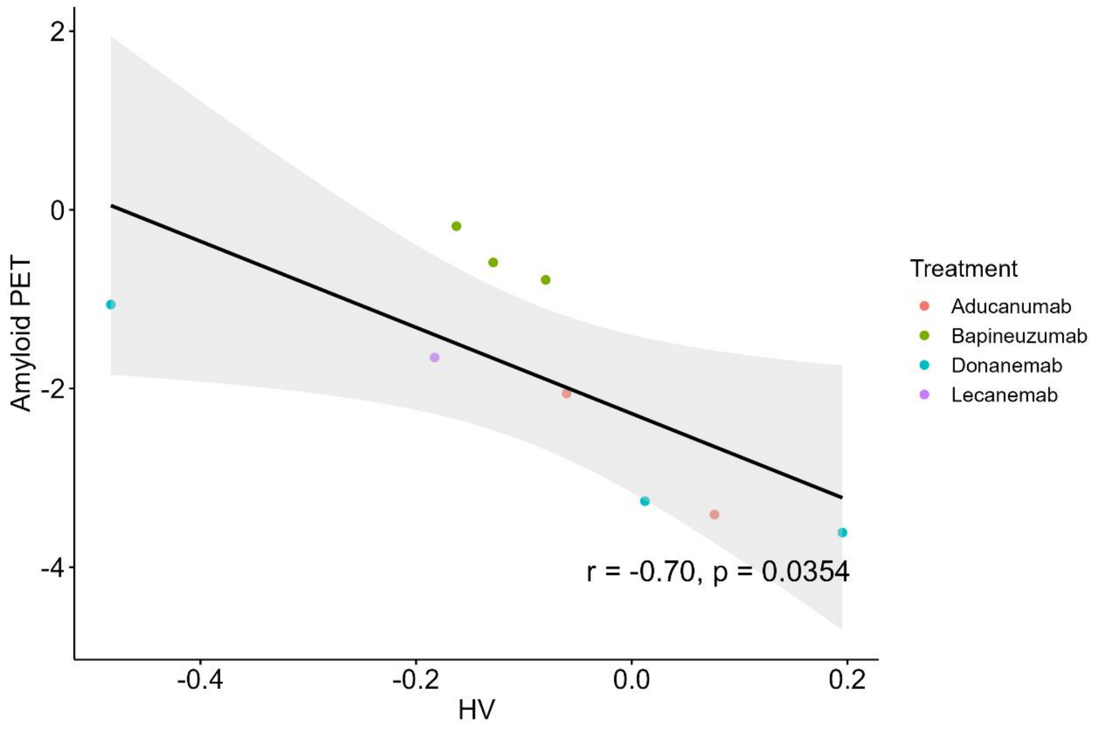
Amyloid PET
2
0
-2
-4
-0.4
-0.2
0.0
0.2
HV
r = -0.70, p = 0.0354
Treatment
Aducanumab
Bapineuzumab
Donanemab
Lecanemab

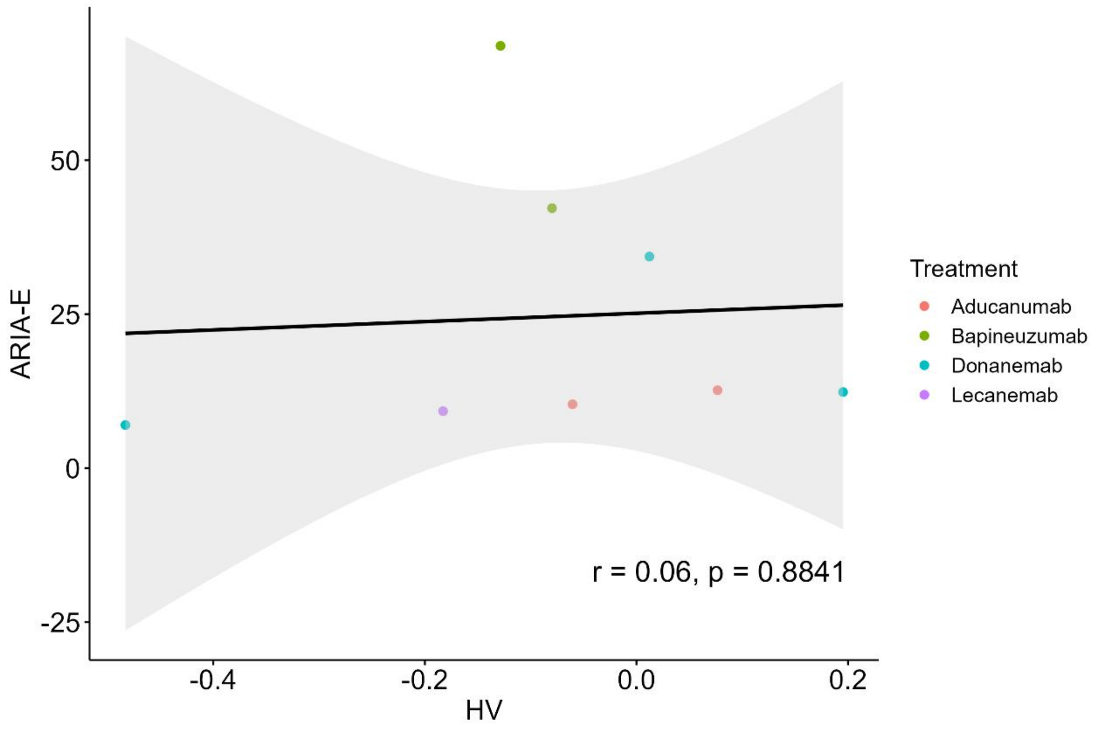
ARIA-E
50
25
0
-25
-0.4
-0.2
0.0
0.2
HV
r = 0.06, p = 0.8841
Treatment
Aducanumab
Bapineuzumab
Donanemab
Lecanemab

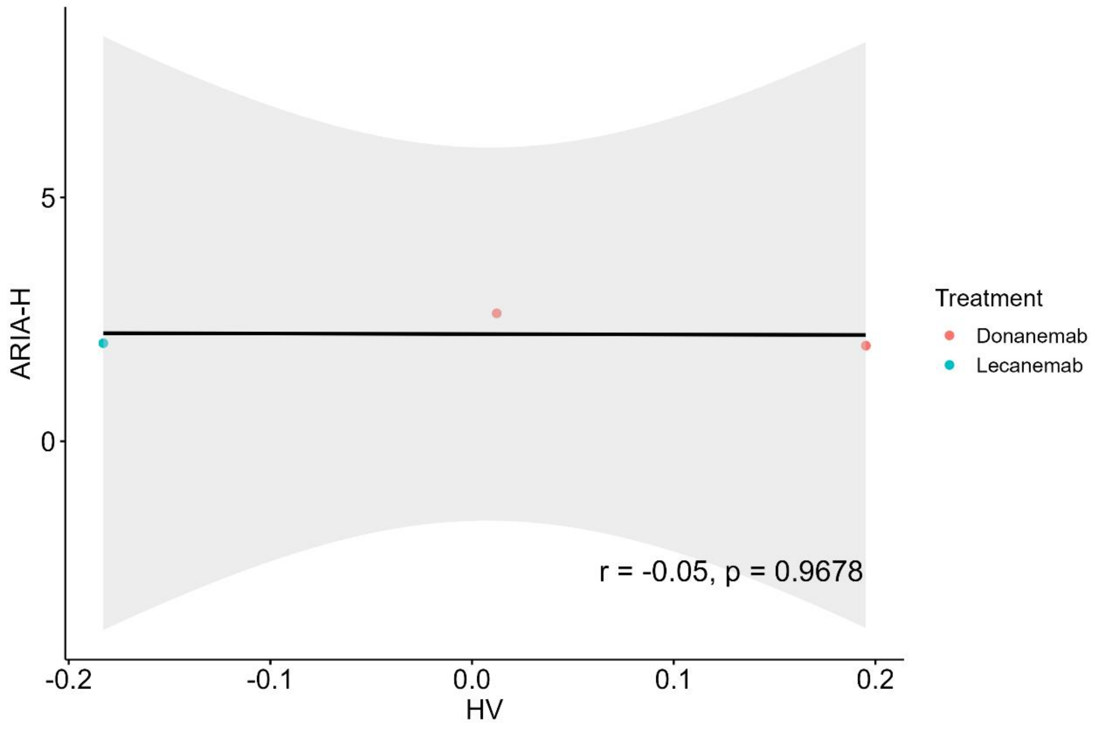

ARIA-H
5
0
-0.2
-0.1
0.0
0.1
0.2
HV
r = -0.05, p = 0.9678
Treatment
Donanemab
Lecanemab

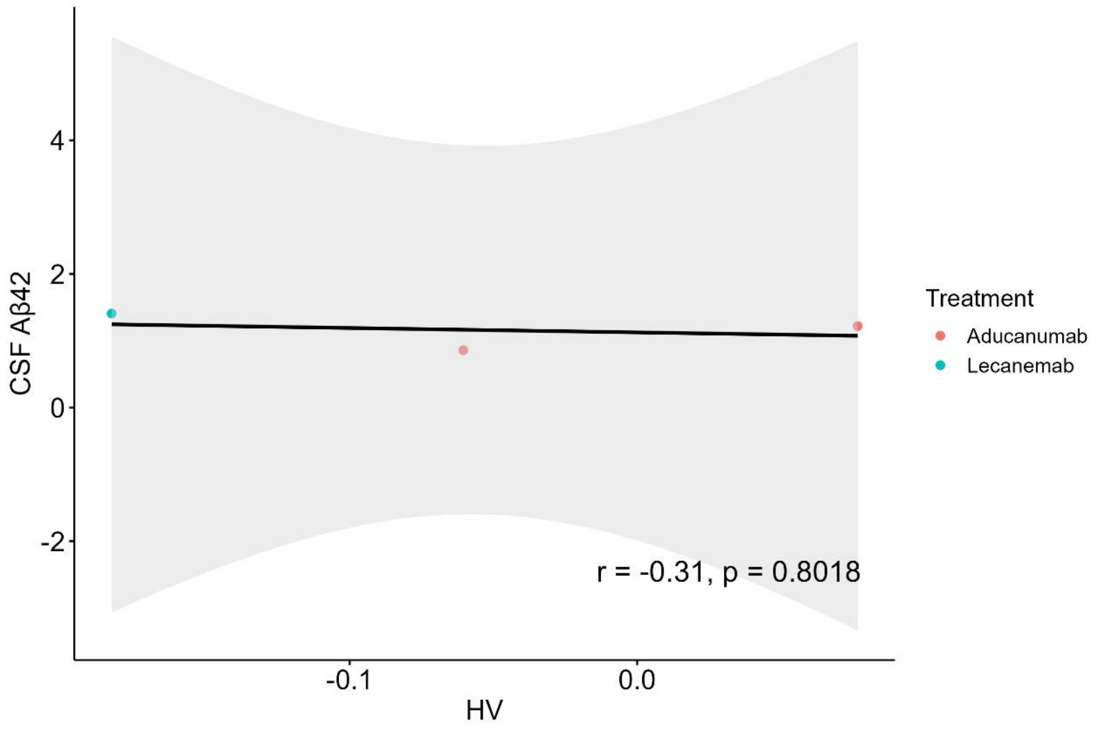

CSF Aβ42
4
2
0
-2
-0.1
0.0
HV
r = -0.31, p = 0.8018
Treatment
Aducanumab
Lecanemab

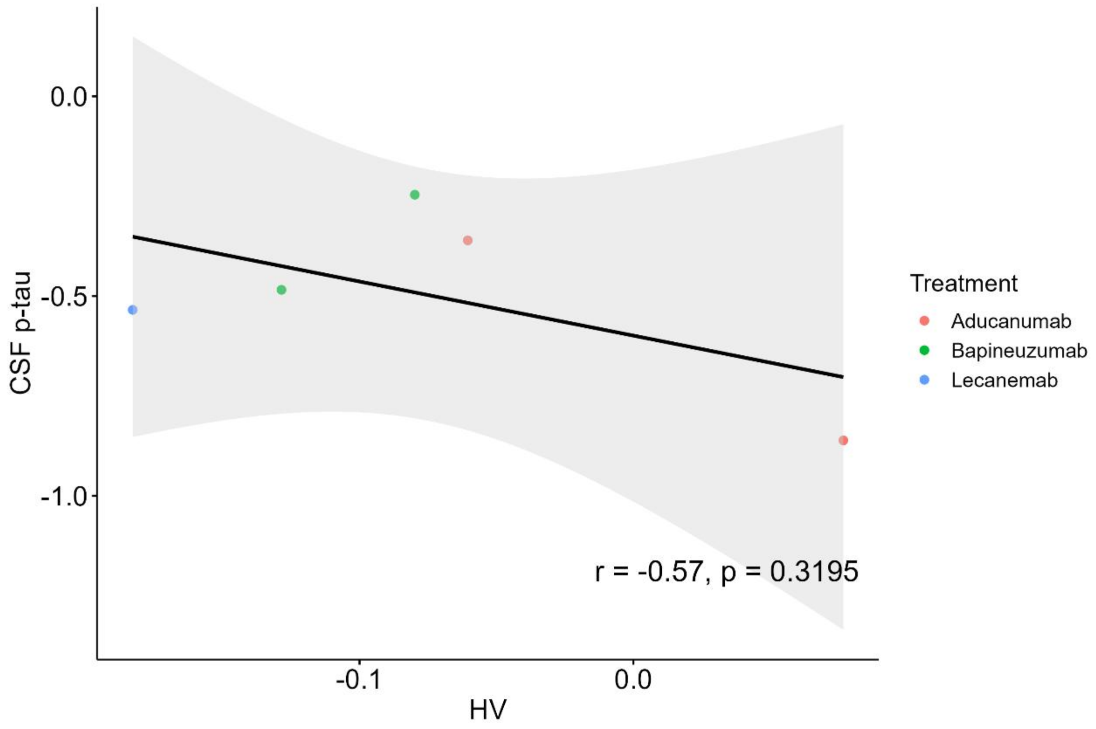
CSF p-tau
HV
Treatment
Aducanumab
Bapineuzumab
Lecanemab
r = -0.57, p = 0.3195
0.0
-0.5
-1.0
-0.1
0.0

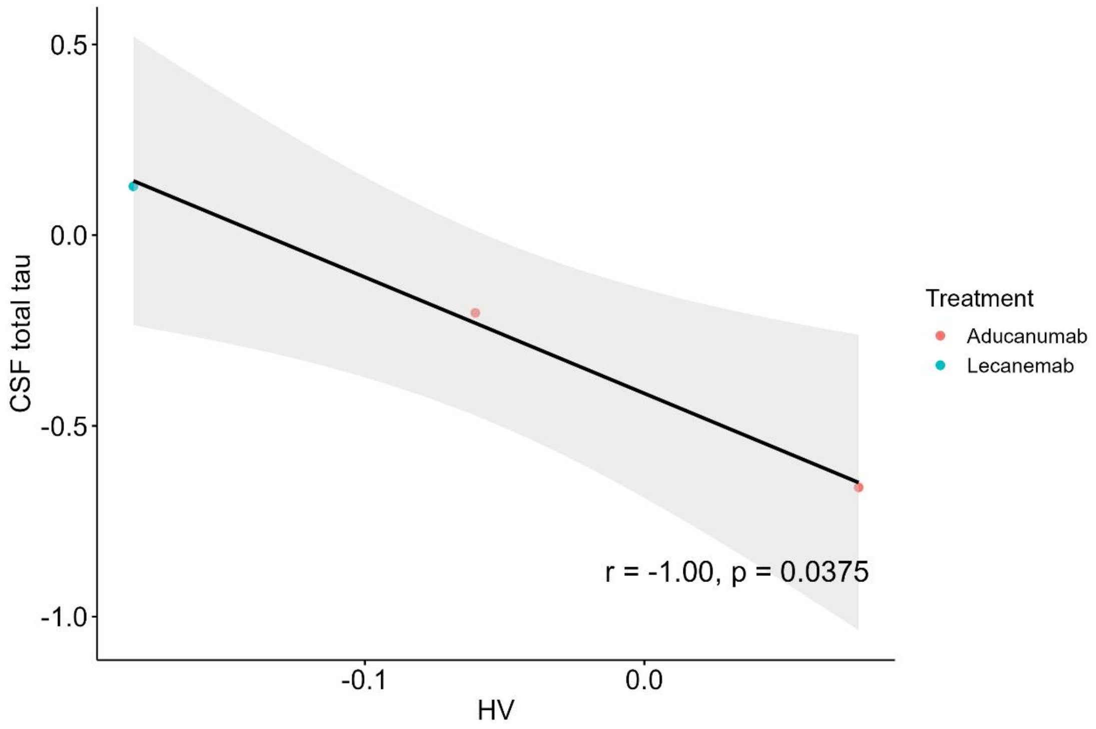
CSF total tau
HV
Treatment
Aducanumab
Lecanemab
r = -1.00, p = 0.0375
0.5
0.0
-0.5
-1.0
-0.1
0.0

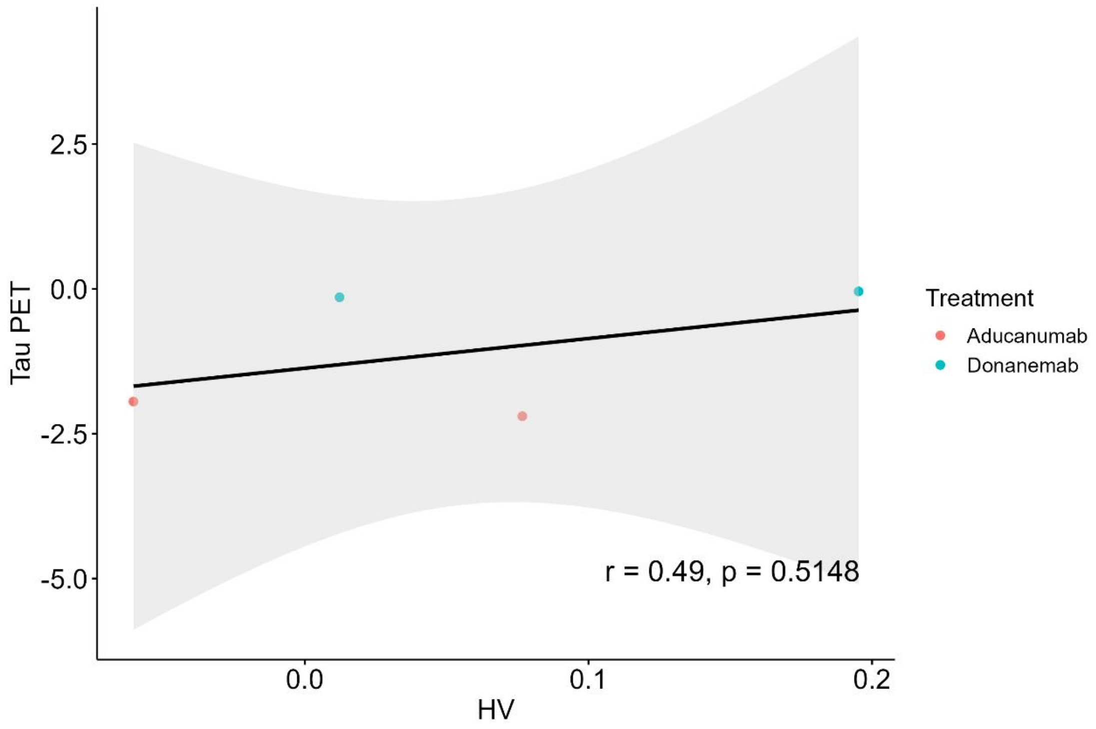

2.5
0.0
-2.5
-5.0
Tau PET
0.0
0.1
0.2
HV
r = 0.49, p = 0.5148
Treatment
Aducanumab
Donanemab

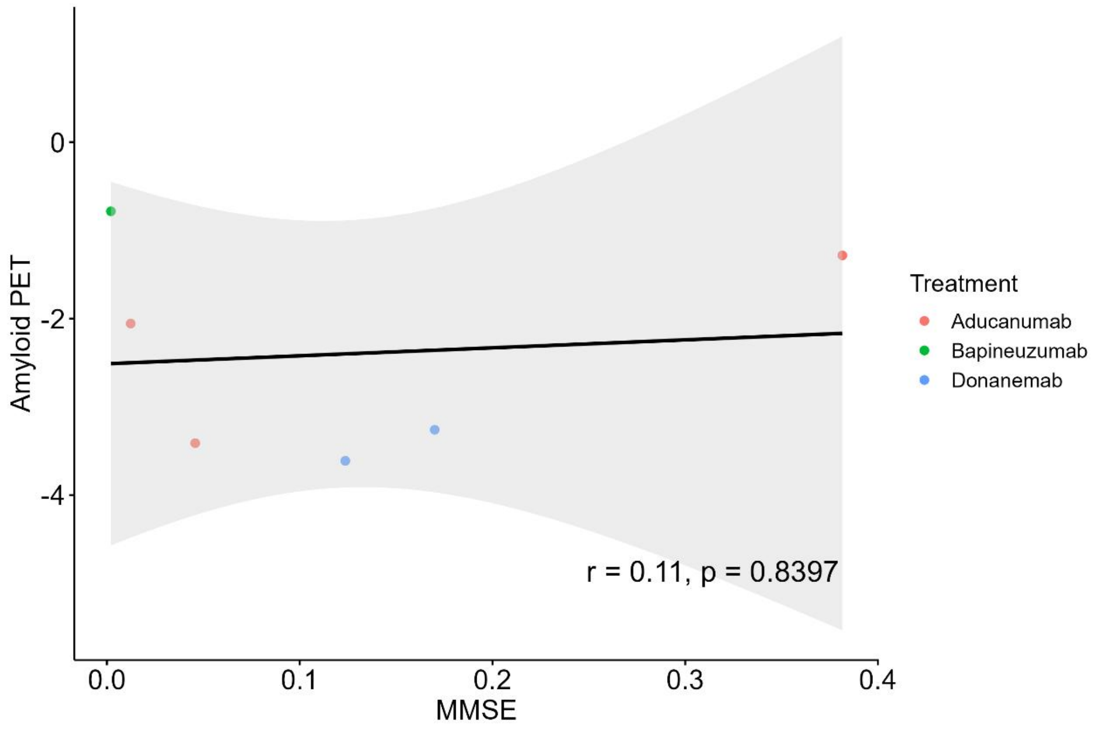

0
-2
-4
Amyloid PET
0.0
0.1
0.2
0.3
0.4
MMSE
r = 0.11, p = 0.8397
Treatment
Aducanumab
Bapineuzumab
Donanemab

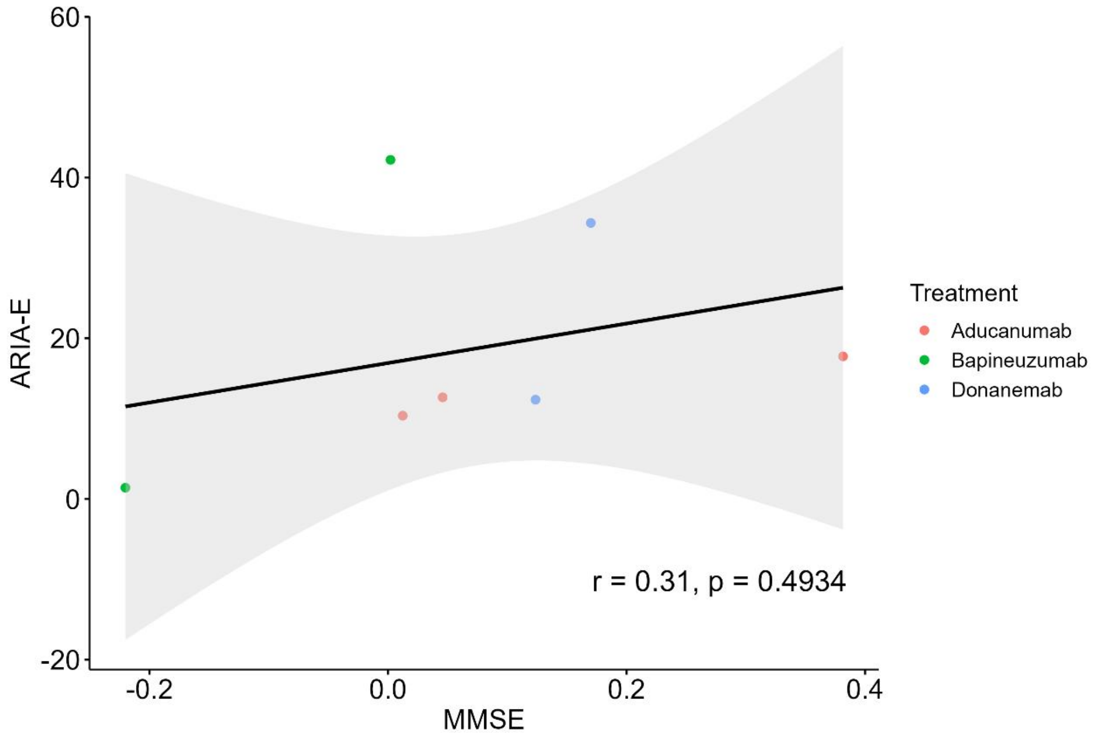
60
40
20
0
-20
ARIA-E
-0.2
0.0
0.2
0.4
MMSE
r = 0.31, p = 0.4934
Treatment
Aducanumab
Bapineuzumab
Donanemab

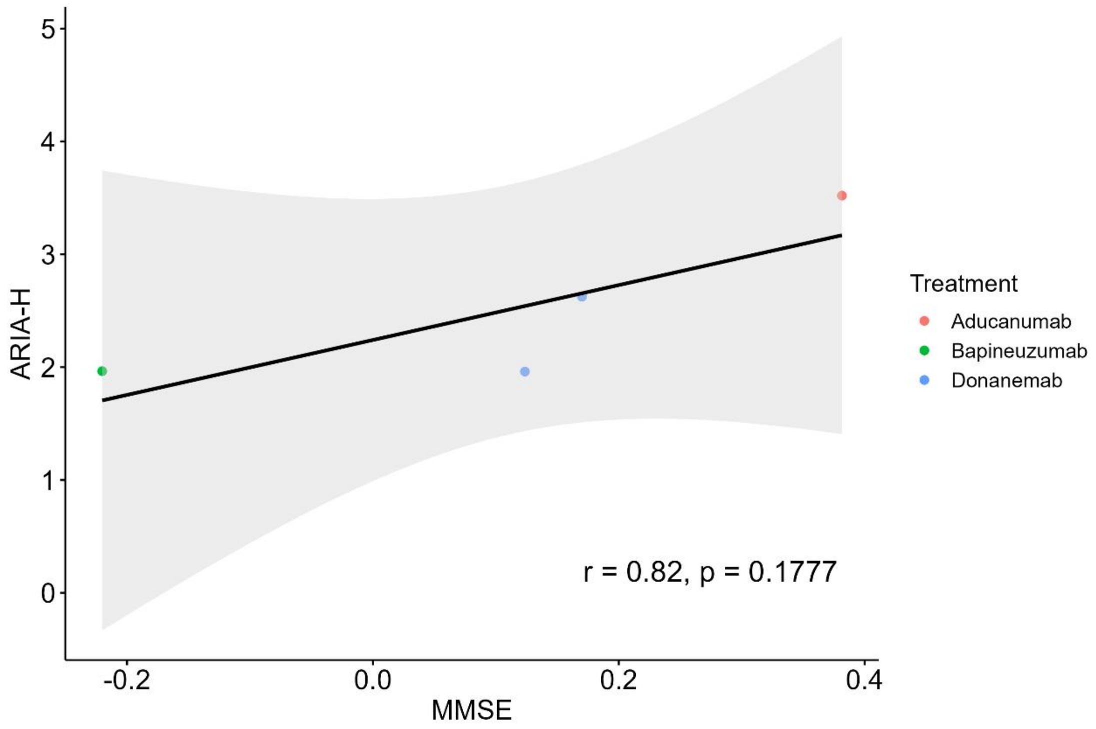
5
4
3
2
1
0
ARIA-H
-0.2
0.0
0.2
0.4
MMSE
r = 0.82, p = 0.1777
Treatment
Aducanumab
Bapineuzumab
Donanemab

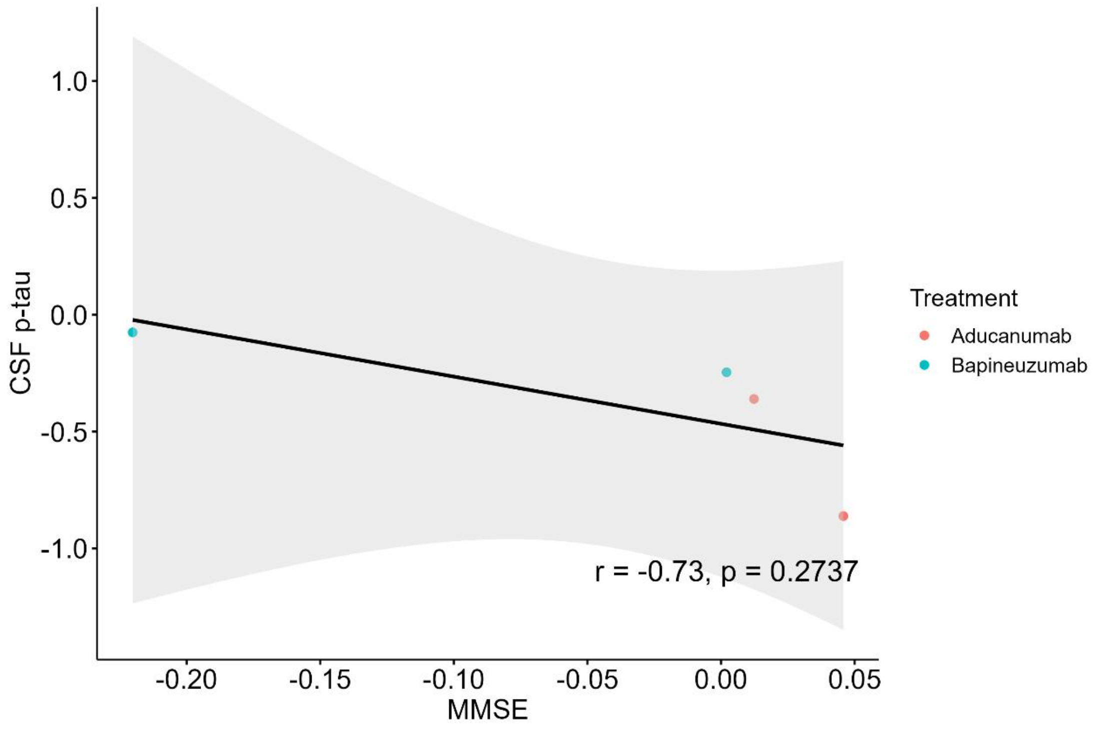
1.0
0.5
0.0
-0.5
-1.0
CSF p-tau
r = -0.73, p = 0.2737
-0.20
-0.15
-0.10
-0.05
0.00
0.05
MMSE
Treatment
Aducanumab
Bapineuzumab

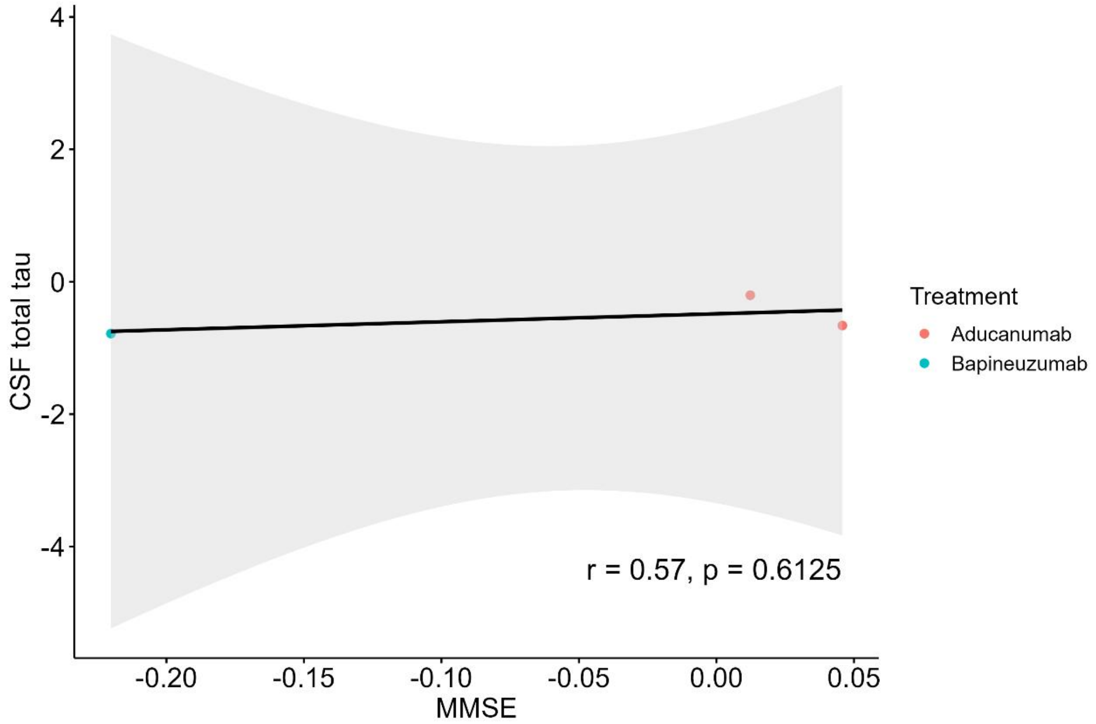
4
2
0
-2
-4
CSF total tau
r = 0.57, p = 0.6125
-0.20
-0.15
-0.10
-0.05
0.00
0.05
MMSE
Treatment
Aducanumab
Bapineuzumab

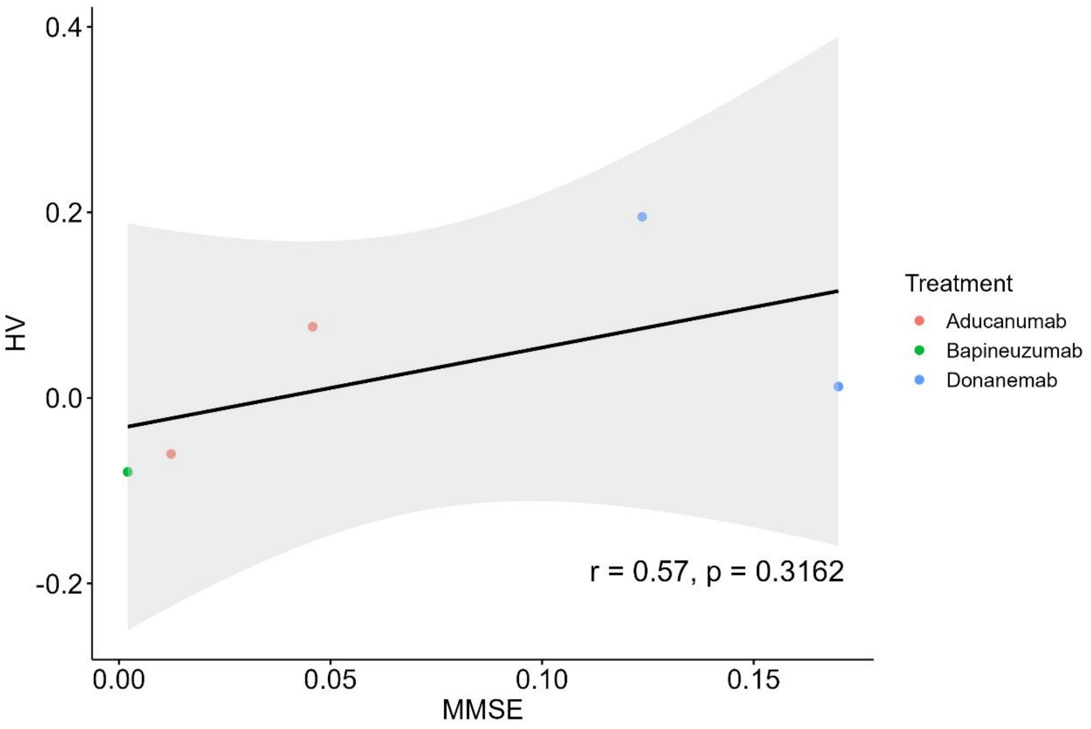
0.4
0.2
0.0
-0.2
HV
0.00
0.05
0.10
0.15
MMSE
Treatment
Aducanumab
Bapineuzumab
Donanemab
r = 0.57, p = 0.3162

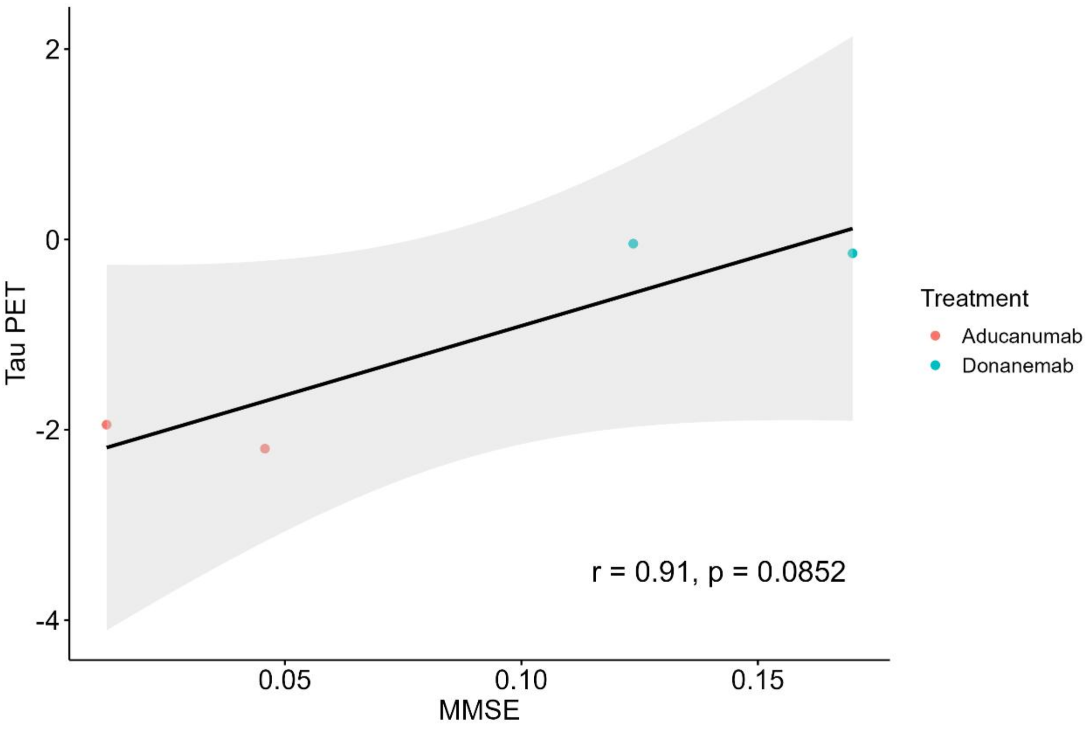
2
0
-2
-4
Tau PET
0.05
0.10
0.15
MMSE
Treatment
Aducanumab
Donanemab
r = 0.91, p = 0.0852

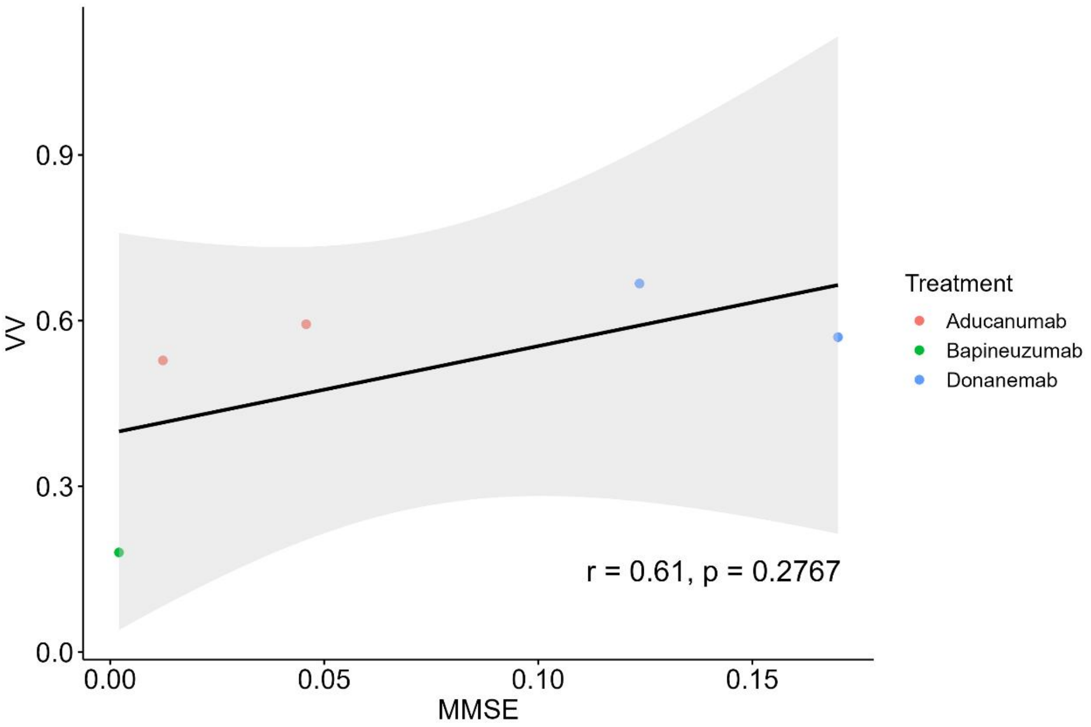

0.9
0.6
0.3
0.0
VV
0.00
0.05
0.10
0.15
MMSE
r = 0.61, p = 0.2767
Treatment
Aducanumab
Bapineuzumab
Donanemab

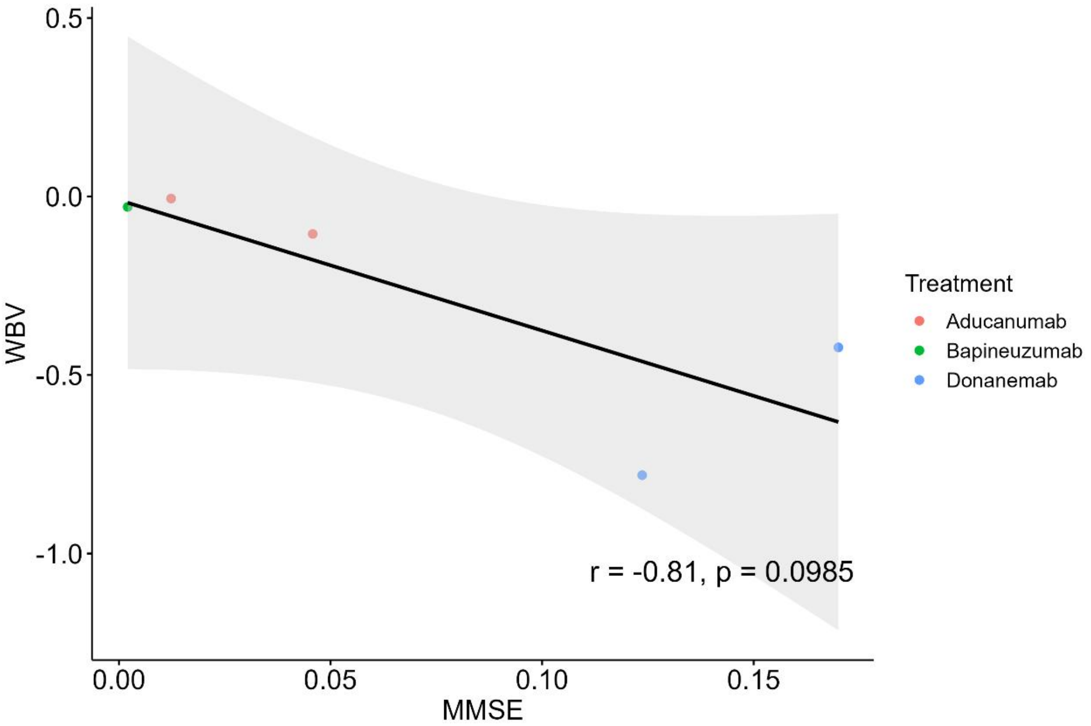

0.5
0.0
-0.5
-1.0
WBV
0.00
0.05
0.10
0.15
MMSE
r = -0.81, p = 0.0985
Treatment
Aducanumab
Bapineuzumab
Donanemab

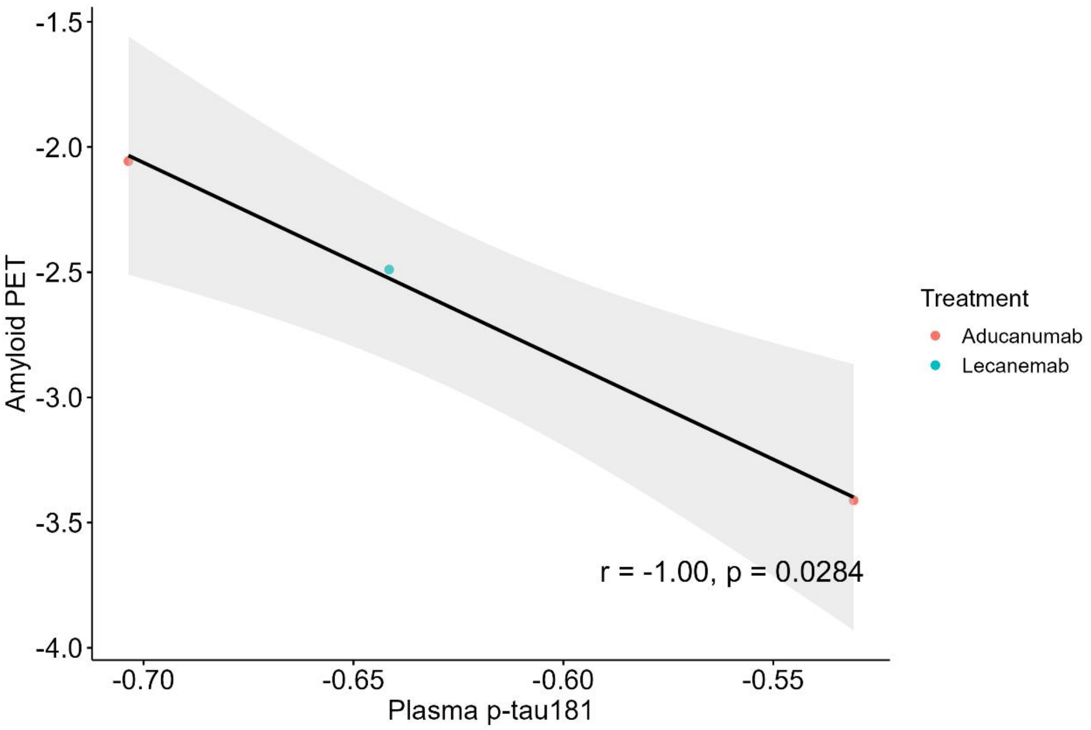

-1.5
-2.0
-2.5
-3.0
-3.5
-4.0
Amyloid PET
-0.70
-0.65
-0.60
-0.55
Plasma p-tau181
r = -1.00, p = 0.0284
Treatment
Aducanumab
Lecanemab

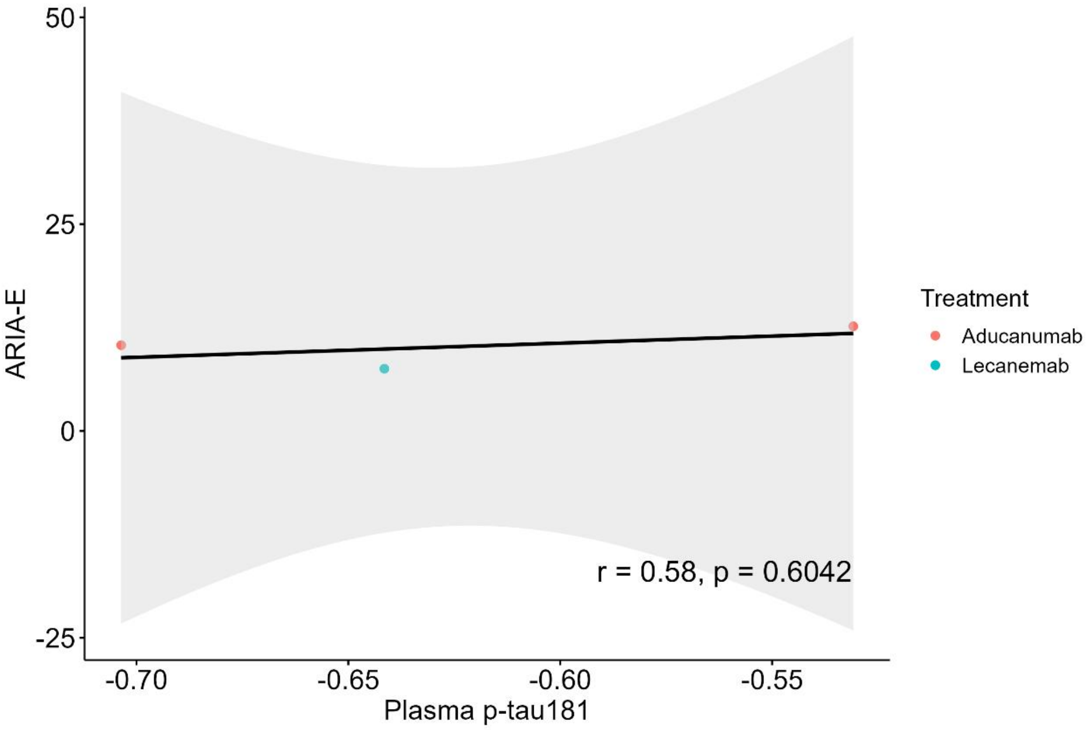

50
25
0
-25
ARIA-E
-0.70
-0.65
-0.60
-0.55
Plasma p-tau181
r = 0.58, p = 0.6042
Treatment
Aducanumab
Lecanemab

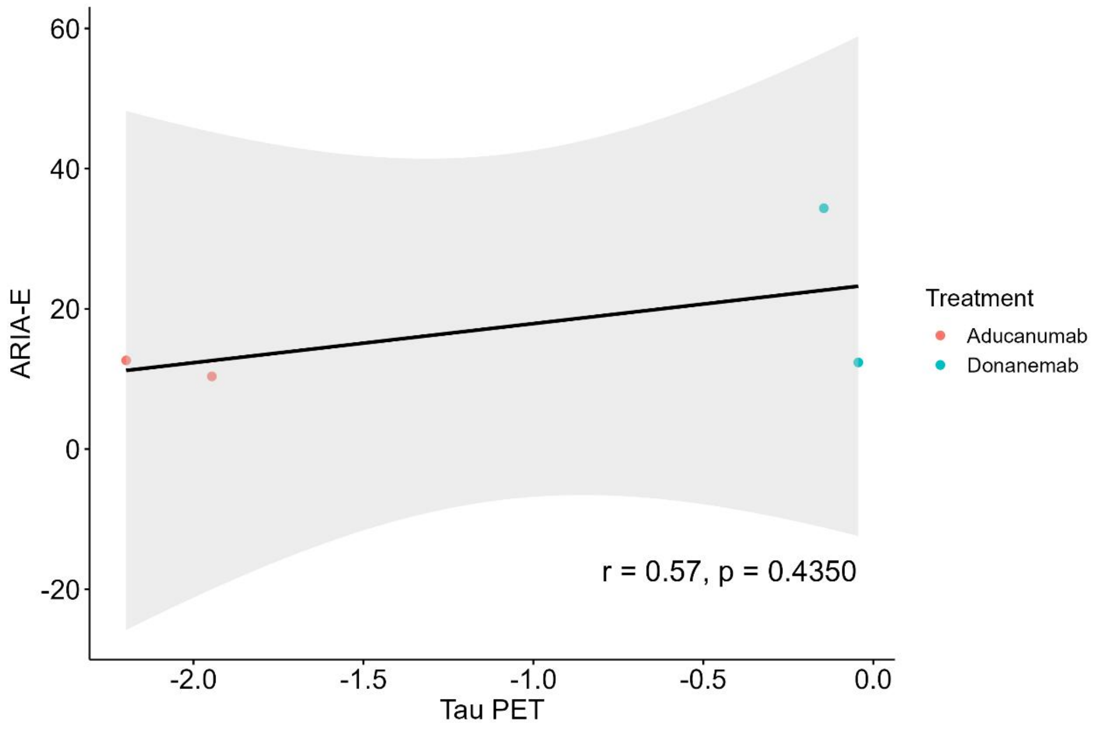
ARIA-E
60
40
20
0
-20
-2.0
-1.5
-1.0
-0.5
0.0
Tau PET
r = 0.57, p = 0.4350
Treatment
Aducanumab
Donanemab

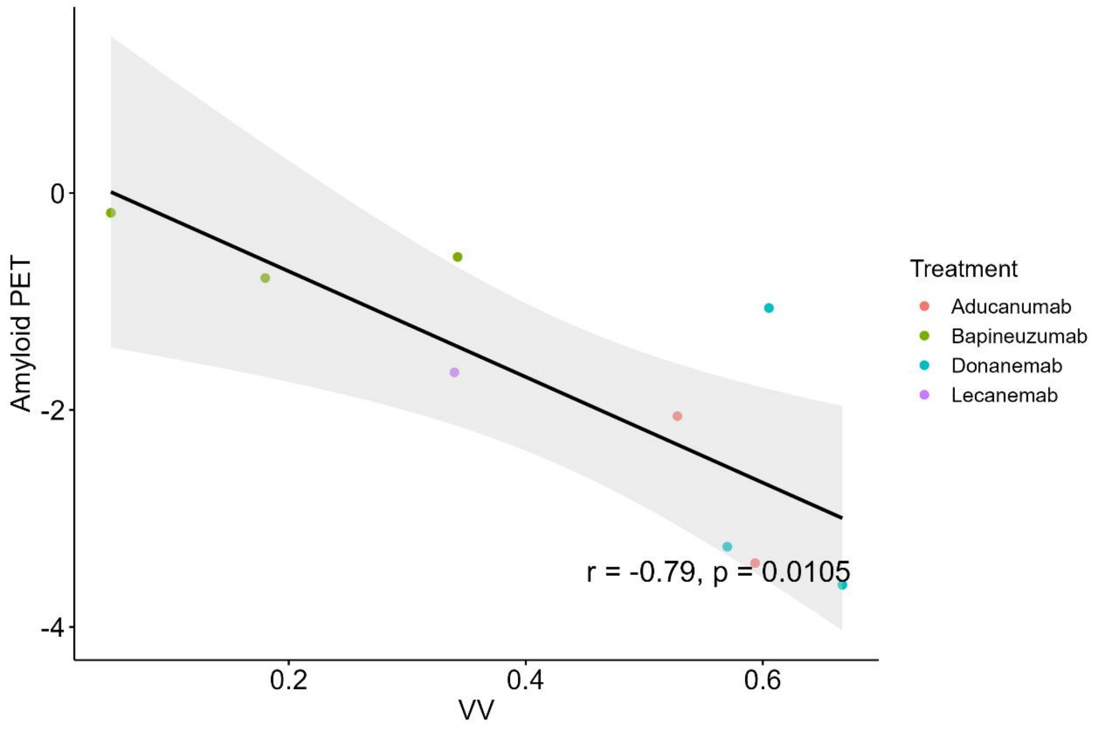
Amyloid PET
0
-2
-4
0.2
0.4
0.6
VV
r = -0.79, p = 0.0105
Treatment
Aducanumab
Bapineuzumab
Donanemab
Lecanemab

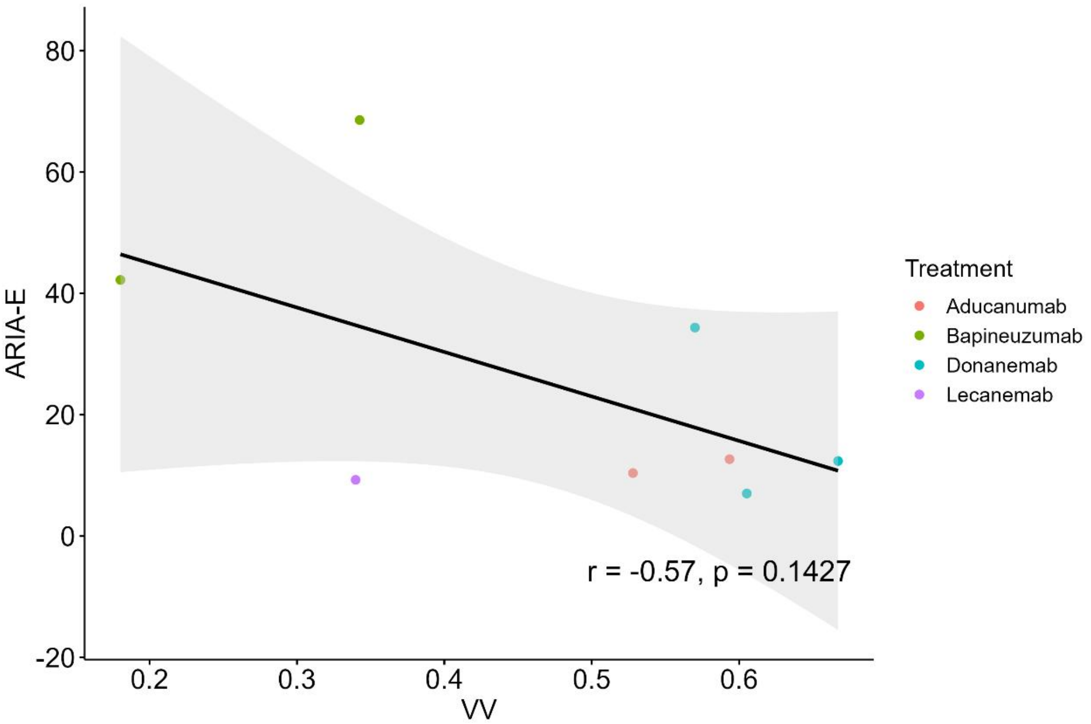
ARIA-E
VV
Treatment
Aducanumab
Bapineuzumab
Donanemab
Lecanemab
r = -0.57, p = 0.1427

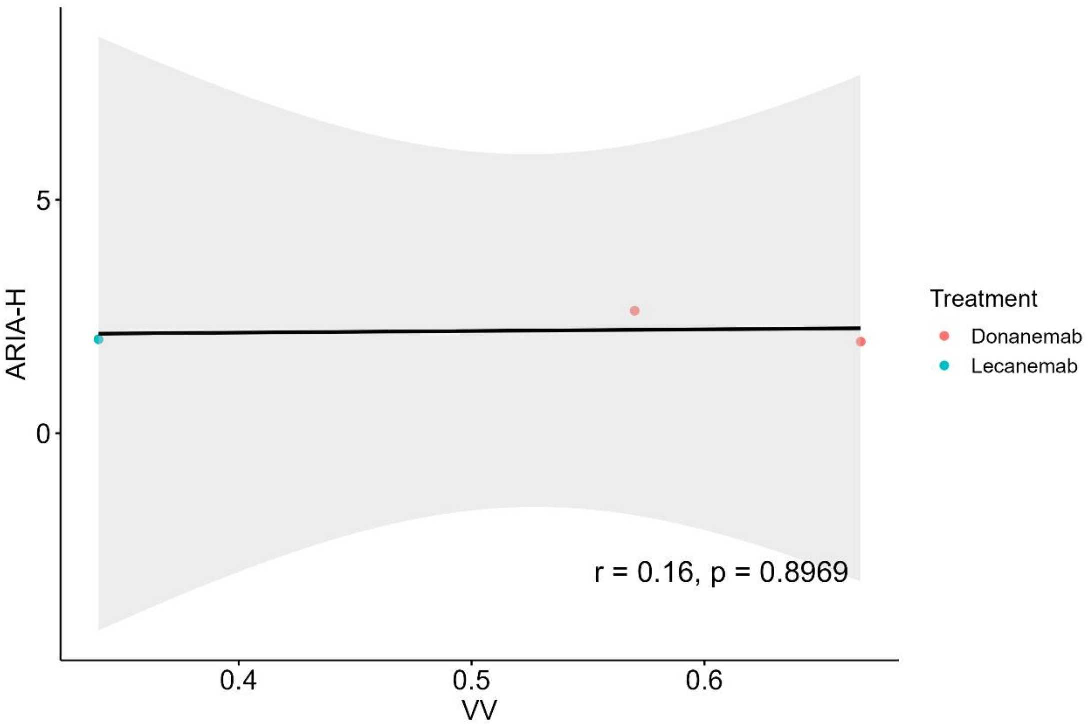
ARIA-H
VV
Treatment
Donanemab
Lecanemab
r = 0.16, p = 0.8969

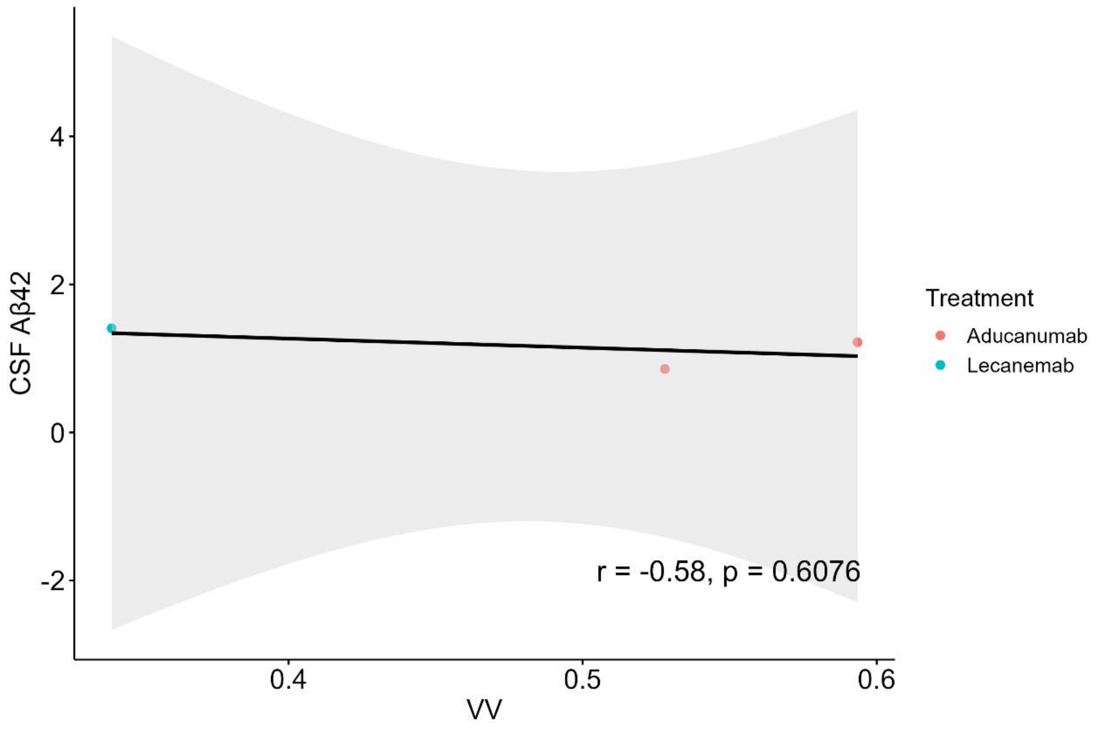
CSF Aβ42
4
2
0
-2
0.4
0.5
0.6
VV
r = -0.58, p = 0.6076
Treatment
Aducanumab
Lecanemab

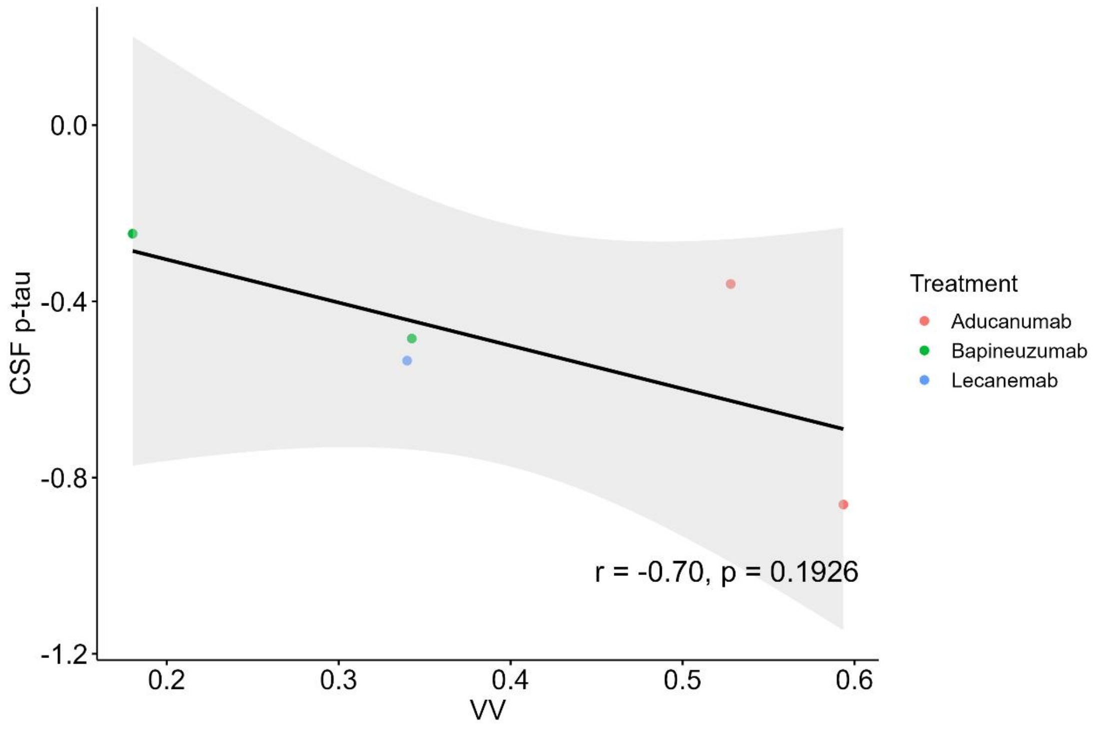
CSF p-tau
0.0
-0.4
-0.8
-1.2
0.2
0.3
0.4
0.5
0.6
VV
r = -0.70, p = 0.1926
Treatment
Aducanumab
Bapineuzumab
Lecanemab

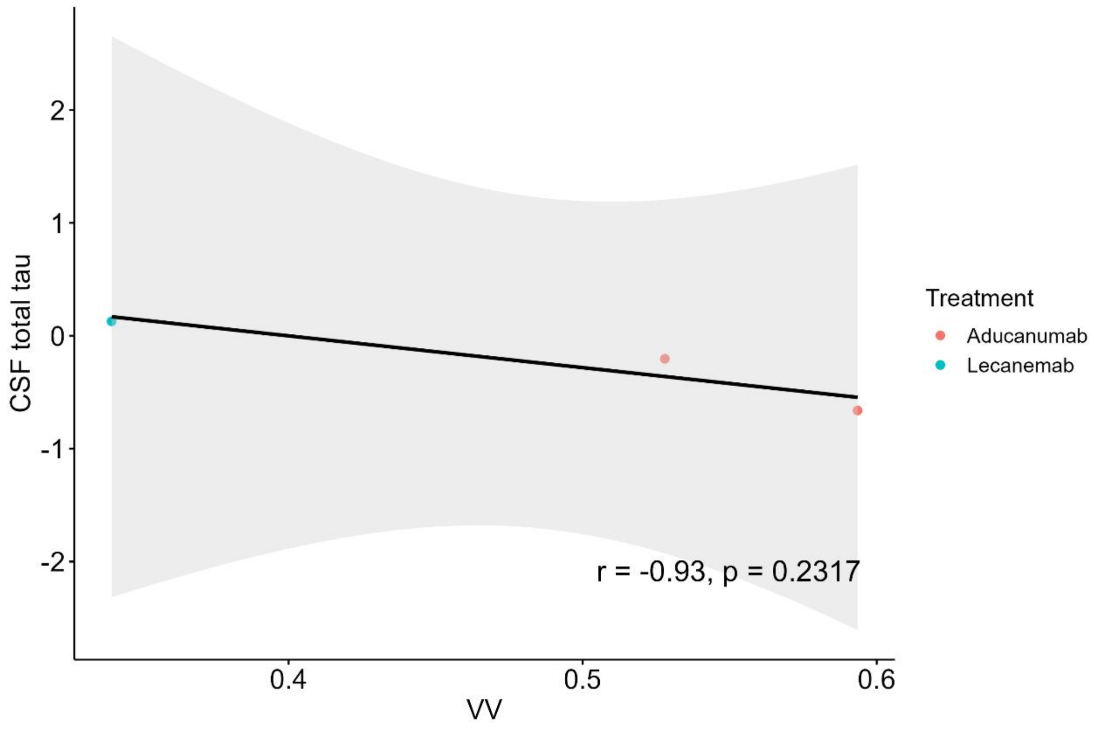
CSF total tau
VV
r = -0.93, p = 0.2317
Treatment
Aducanumab
Lecanemab

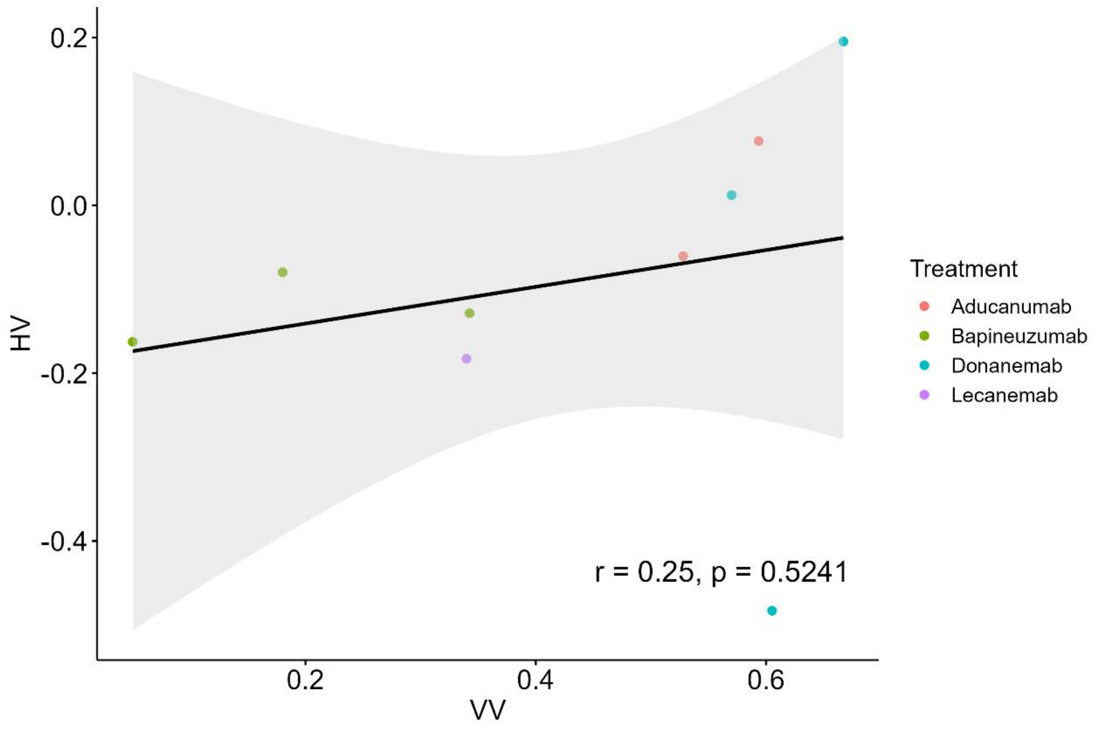
HV
VV
r = 0.25, p = 0.5241
Treatment
Aducanumab
Bapineuzumab
Donanemab
Lecanemab

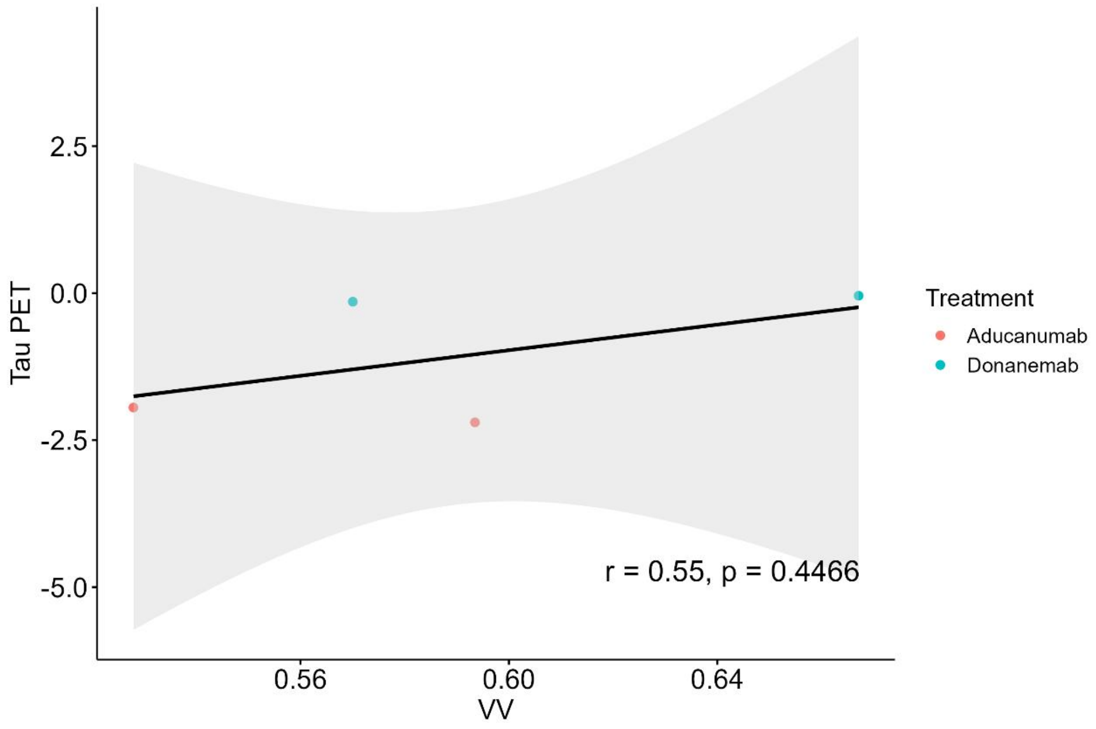
Tau PET
2.5
0.0
-2.5
-5.0
0.56
0.60
0.64
VV
r = 0.55, p = 0.4466
Treatment
Aducanumab
Donanemab

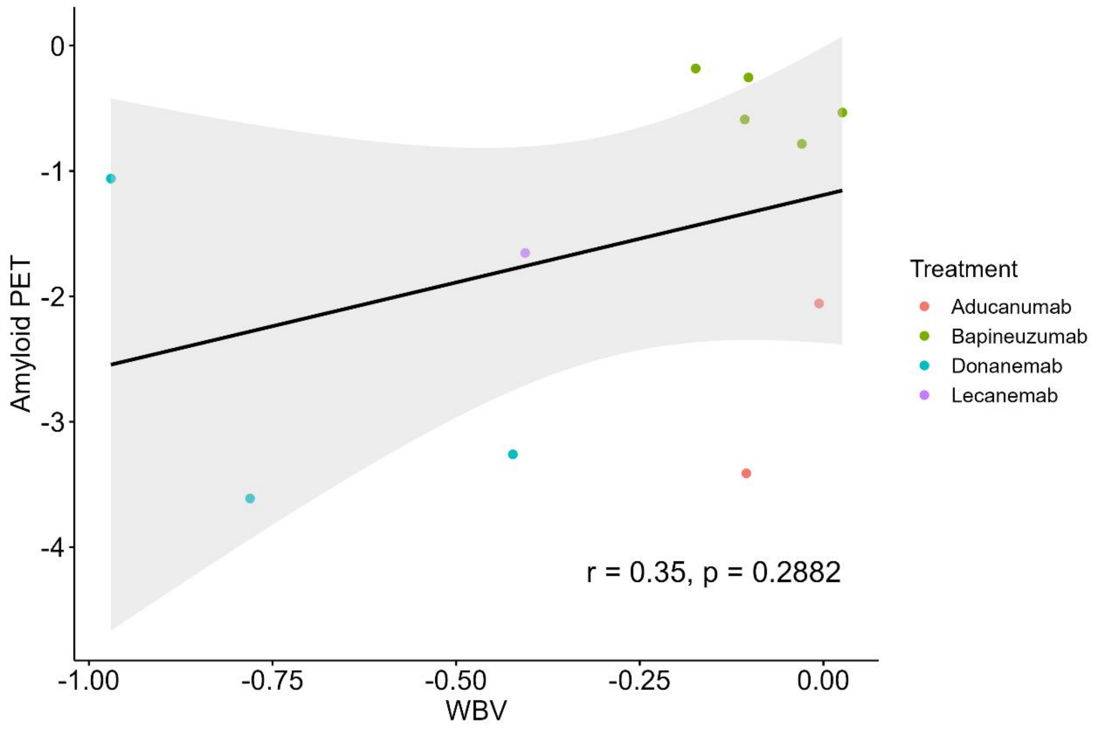
Amyloid PET
0
-1
-2
-3
-4
-1.00
-0.75
-0.50
-0.25
0.00
WBV
r = 0.35, p = 0.2882
Treatment
Aducanumab
Bapineuzumab
Donanemab
Lecanemab

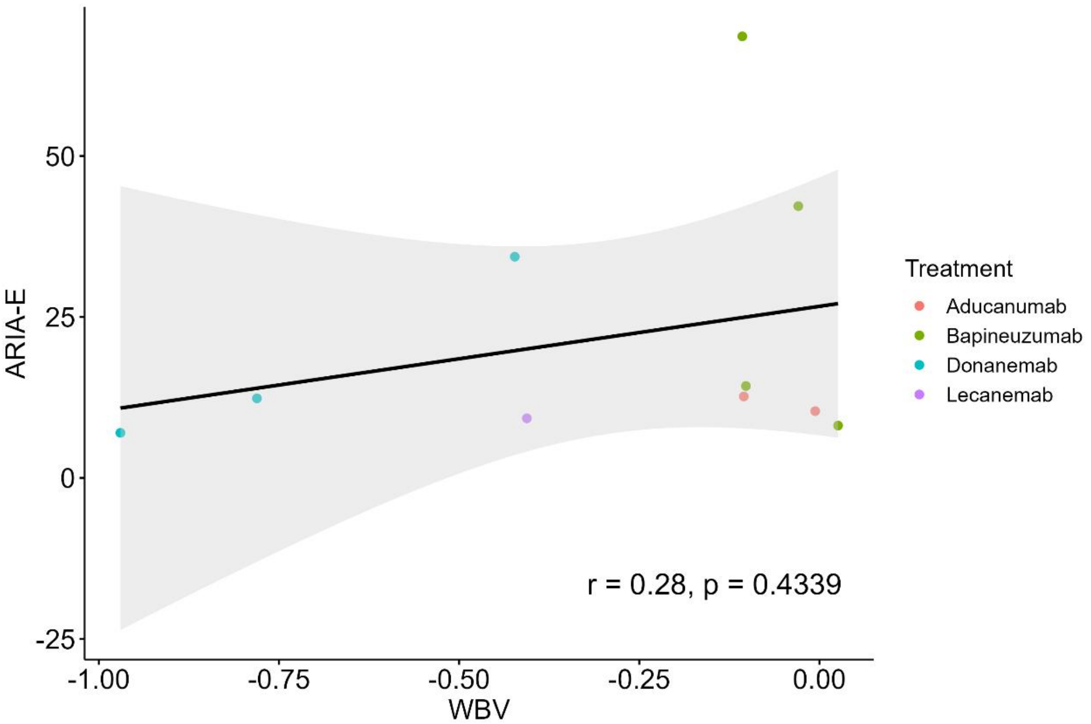
ARIA-E
50
25
0
-25
-1.00
-0.75
-0.50
-0.25
0.00
WBV
r = 0.28, p = 0.4339
Treatment
Aducanumab
Bapineuzumab
Donanemab
Lecanemab

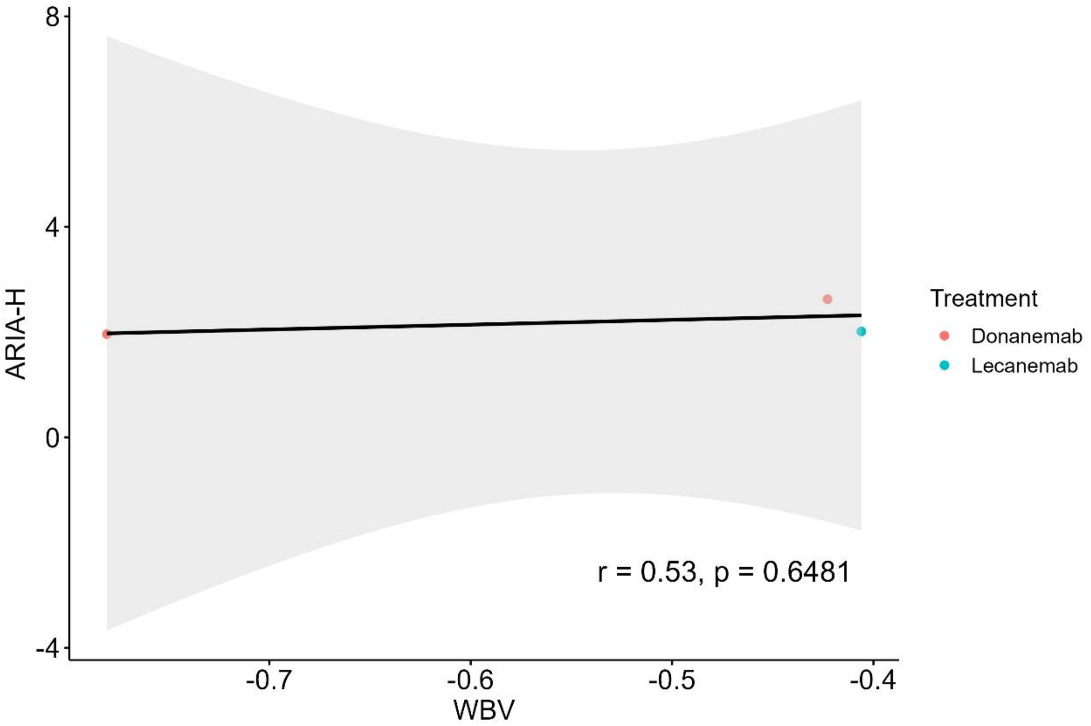
ARIA-H
8
4
0
-4
-0.7
-0.6
-0.5
-0.4
WBV
r = 0.53, p = 0.6481
Treatment
Donanemab
Lecanemab

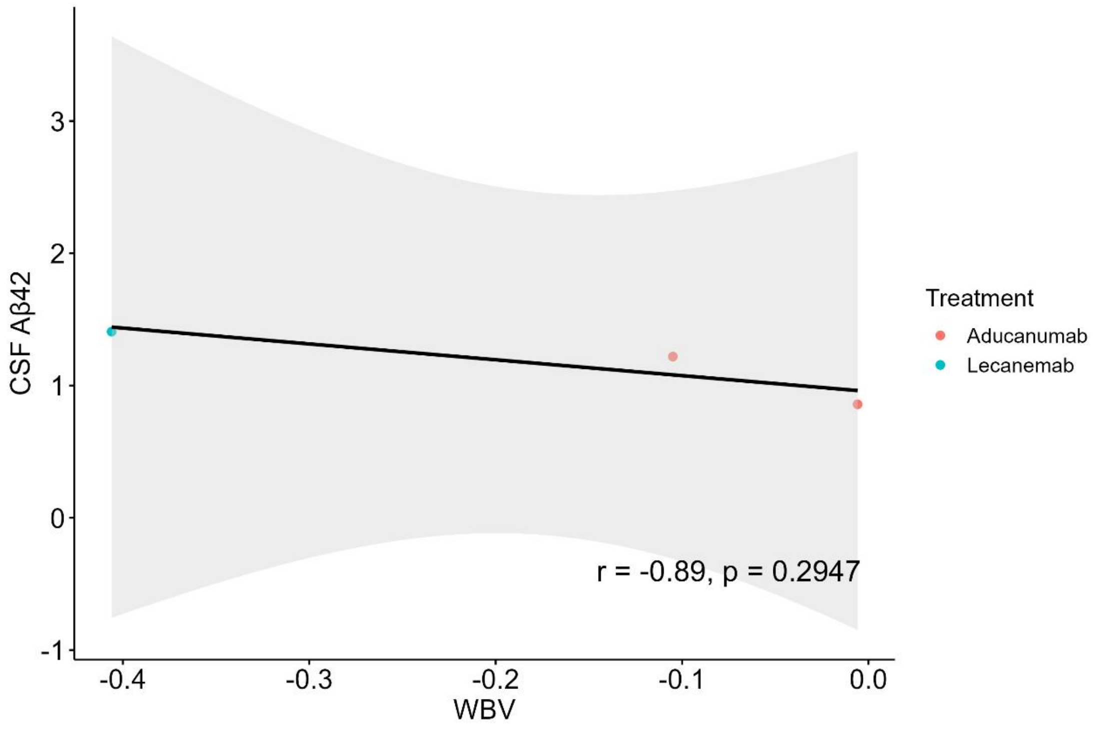

CSF Aβ42
WBV
Treatment
Aducanumab
Lecanemab
r = -0.89, p = 0.2947

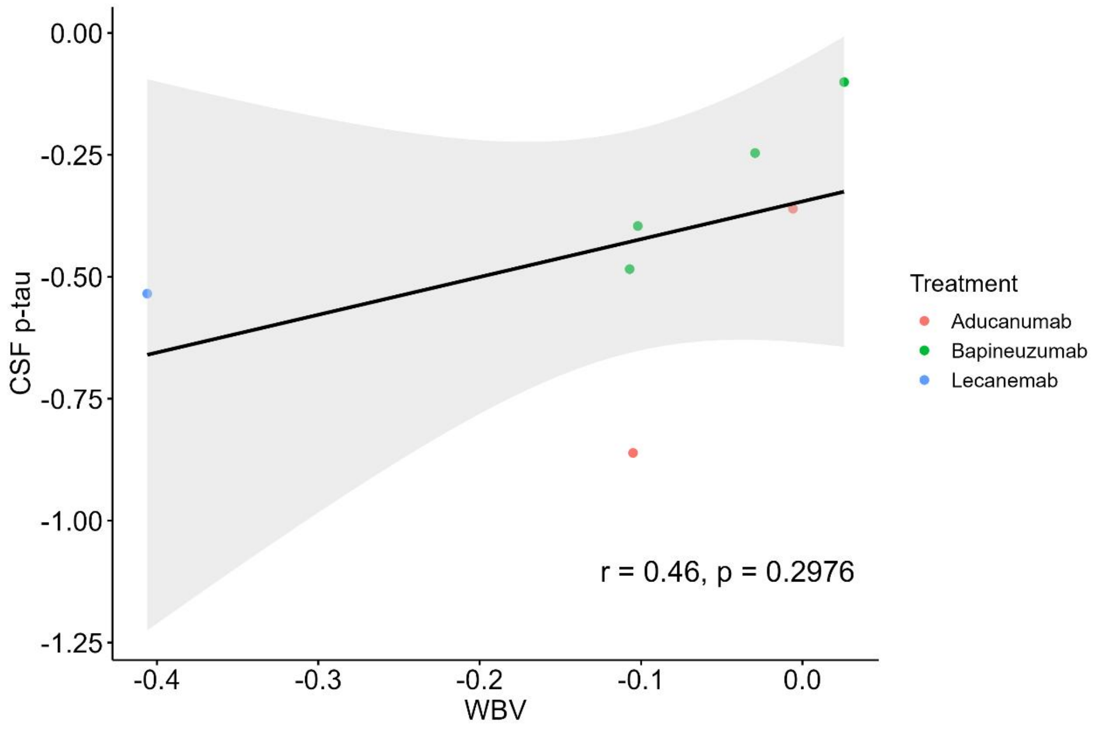

CSF p-tau
WBV
Treatment
Aducanumab
Bapineuzumab
Lecanemab
r = 0.46, p = 0.2976

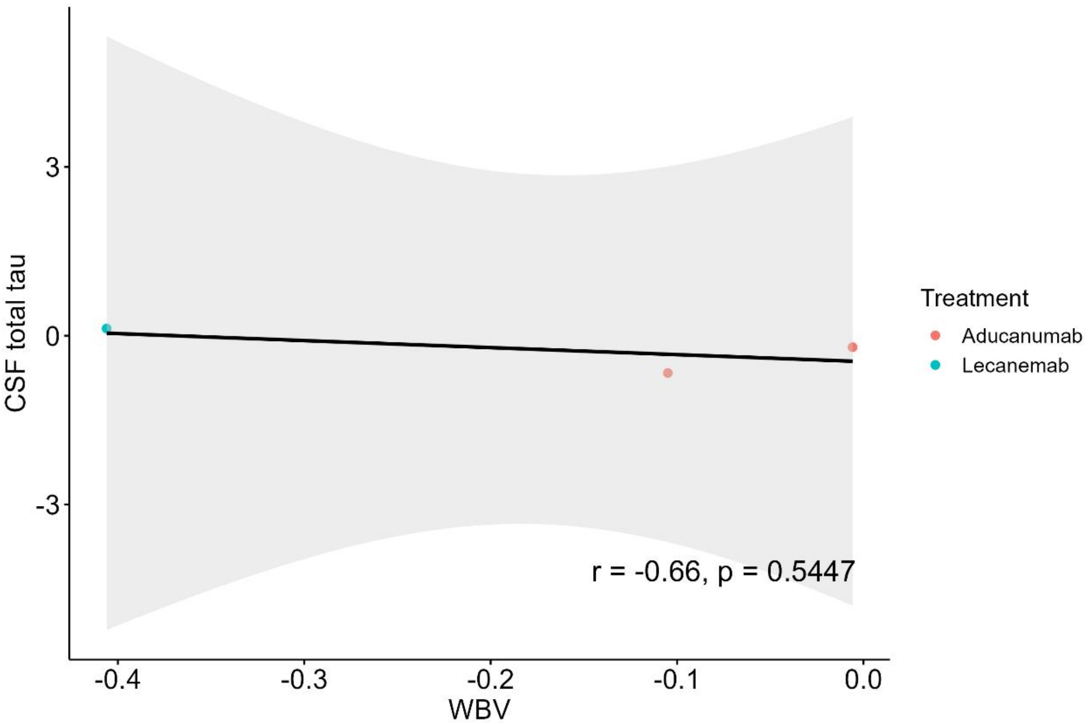
CSF total tau
3
0
-3
-0.4
-0.3
-0.2
-0.1
0.0
WBV
r = -0.66, p = 0.5447
Treatment
Aducanumab
Lecanemab

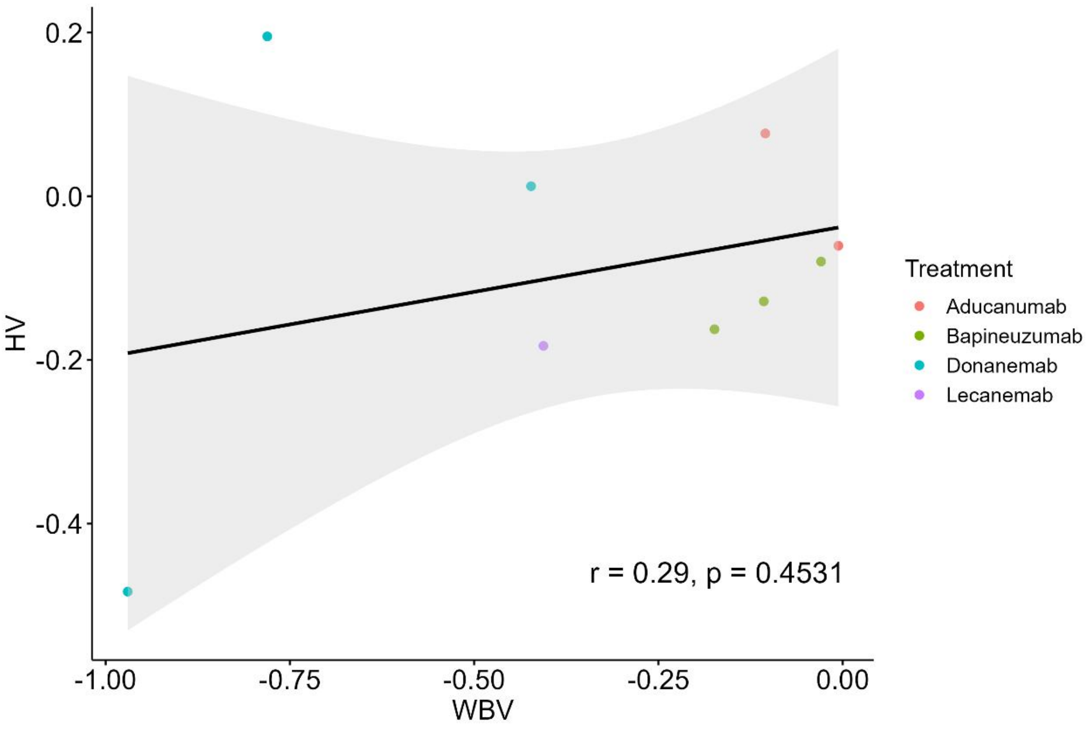
HV
0.2
0.0
-0.2
-0.4
-1.00
-0.75
-0.50
-0.25
0.00
WBV
r = 0.29, p = 0.4531
Treatment
Aducanumab
Bapineuzumab
Donanemab
Lecanemab

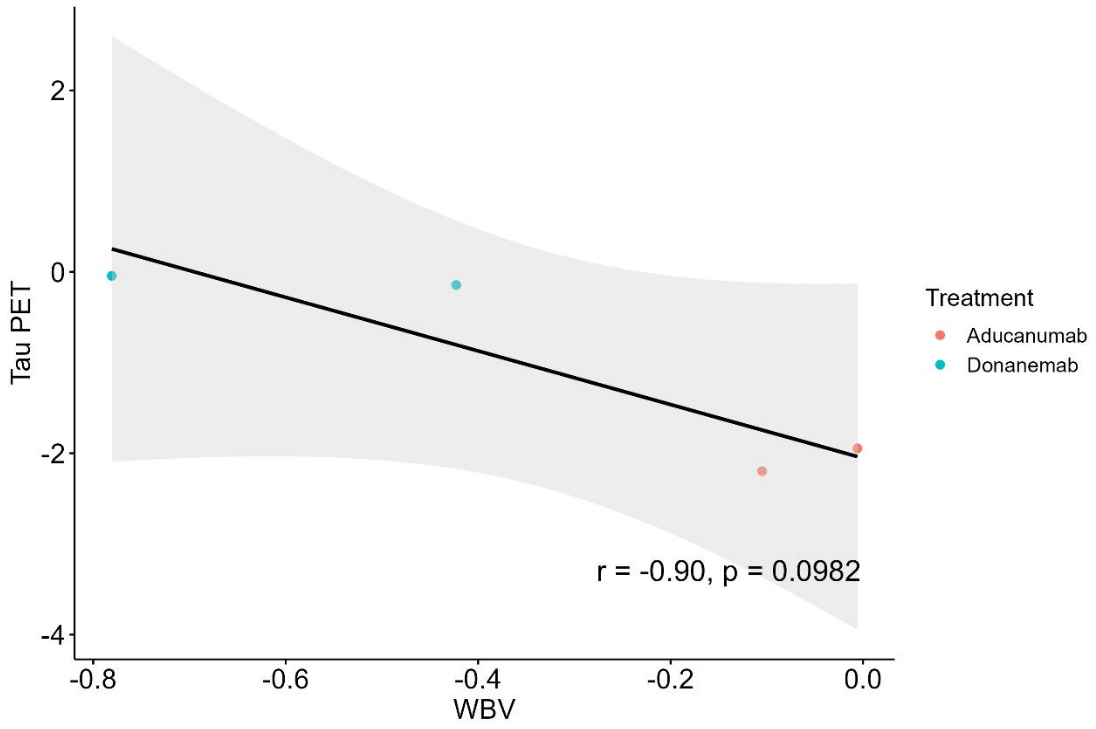
Tau PET
2
0
-2
-4
-0.8
-0.6
-0.4
-0.2
0.0
WBV
r = -0.90, p = 0.0982
Treatment
Aducanumab
Donanemab

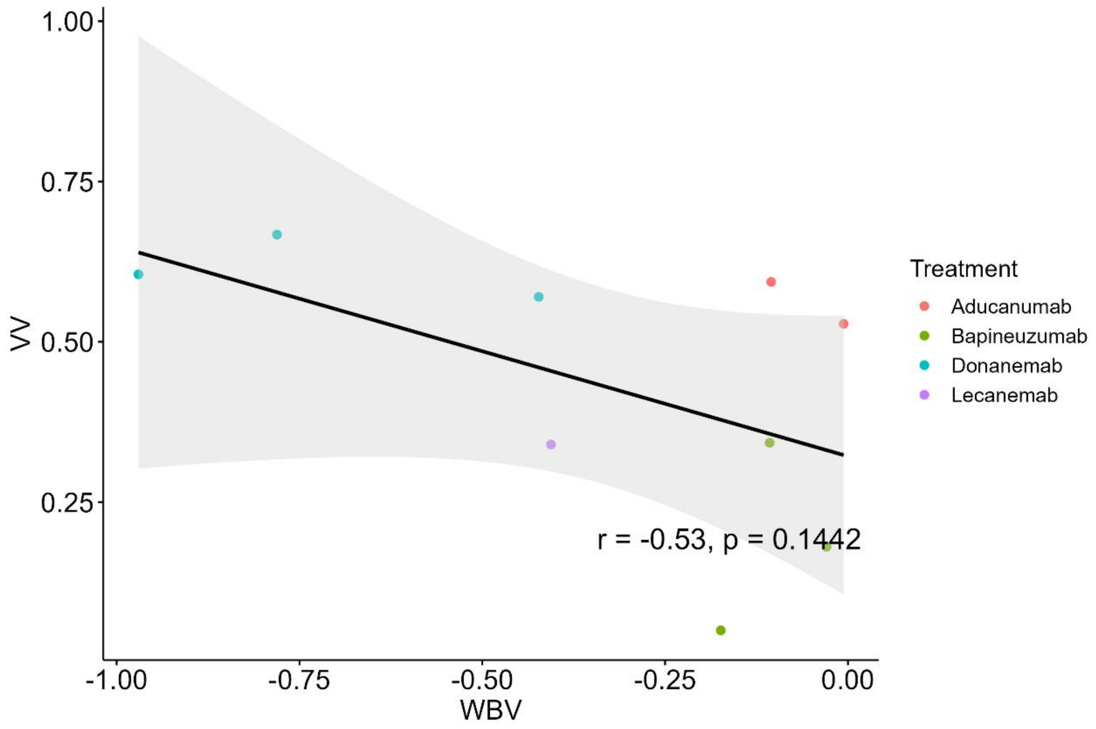
VV
1.00
0.75
0.50
0.25
-1.00
-0.75
-0.50
-0.25
0.00
WBV
r = -0.53, p = 0.1442
Treatment
Aducanumab
Bapineuzumab
Donanemab
Lecanemab
